# Virtual control arms for paediatric myopia trials: external validation of axial-elongation models

**DOI:** 10.64898/2026.08.21.26360973

**Authors:** Ravi C Bakaraju, Praveen K Bandela, Jennifer Sha, Daniel Tilia

## Abstract

**Clinical relevance:** Validated virtual control arms may provide population-level estimates of treatment effect and reduce reliance on untreated control allocations in myopia trials.

**Background:** Untreated single-vision control arms in paediatric myopia efficacy trials are increasingly difficult to justify and retain. Several published models predict untreated childhood axial elongation by region or ethnicity. Here they are implemented unchanged in an open-source tool and validated against an untreated multi-ethnic cohort.

**Methods:** Five published models predicted untreated elongation from baseline age, cycloplegic spherical equivalent, sex, and ethnicity, anchored at baseline axial length (AL) and evaluated at actual follow-up. Predictions were compared with 242 untreated myopic children (Chinese, Vietnamese, Indian) with AL measured at approximately 6 and 12 months, assessing bias, root-mean-square error, and prediction-interval coverage against pre-specified thresholds (bias <0.03 mm; coverage ≥0.90).

**Results:** The regional generalised estimating equation (GEE) and meta-regression models reproduced mean East Asian elongation without meaningful bias at 6 months (GEE bias −0.013 mm; equivalence to ±0.03 mm, p = 0.014) and at 12 months (−0.004 mm), although equivalence was not established at 12 months in an underpowered subgroup (n = 71, all Vietnamese; p = 0.068). Older age-only models under-predicted by 0.07 to 0.12 mm. Published individual prediction intervals were too narrow (coverage 0.77): the means were accurate, the individual uncertainty was not. Indian elongation fell between strata and was matched by no existing model.

**Conclusions:** The models reproduce mean untreated East Asian elongation at 6 months, conditional on cohort independence; South Asian children remain unserved by any existing stratum. The tool is a group-level instrument, not an individual predictor, and a transparent unification of published models in open-source code. Its value for estimating treatment effect awaits back-testing against a trial with a known untreated arm, ideally over 24 to 36 months.

## Introduction

Randomised trials of myopia-control interventions have traditionally used single-vision spectacle lenses as the control arm. ^1–5^ As evidence for effective treatments has accumulated, the ethical justification for prolonged single-vision arms has increasingly been questioned,^6^ and the practical problem is now recruitment and retention: families are reluctant to accept no active treatment when effective options exist, and selective withdrawal of fast progressors can bias the control group.^7^ As myopia control becomes standard practice, assembling a genuinely untreated comparator will only get harder, making validated model-based control a necessity rather than a convenience. Axial length is the preferred outcome, being precise, repeatable, and largely independent of refractive-component change.^8,9^

Longitudinal axial-elongation models and centile curves have been developed to characterise eye growth.^10,11^ A key distinction matters: centile curves describe attained axial length and underestimate elongation in already-myopic children, characterising growth status rather than predicting future elongation, whereas virtual control arms use longitudinal prediction models to estimate untreated elongation from baseline age, refraction, sex, and region.^12^ Several published models are region- or ethnicity-specific. The Naduvilath 2025 generalised estimating equation (GEE) model provides separate East Asian and European strata with distinct myope and non-myope equations;^13^ the Naduvilath 2023 Asian normative model conditions on the number of myopic parents;^14^ the Brennan 2024 meta-regression synthesises 78 studies as a function of age and ethnicity in myopes;^15^ and two older age-only models, the Orinda Longitudinal Study of Myopia (Jones 2005, predominantly White United States children)^16^ and the Singapore Cohort Study of the Risk Factors for Myopia (SCORM; Wong 2010, Singaporean children),^17^ predict elongation from age alone.

Despite models dating from 2005,^16^ their ability to predict untreated progression in independent paediatric populations has not been established, yet the validity of a virtual control arm depends on exactly that. We therefore performed a standardised external validation and direct comparison of five published axial-elongation models in a prospective paediatric myopia cohort and implemented each model in an openly available tool for transparent application and independent re-validation. The tool is a group-level, population-mean instrument by design: it supplies the mean untreated trajectory a control arm requires and makes no claim to individual-level discrimination.

## Methods

### Design and estimand

The primary estimand is a validation quantity: the bias and prediction-interval calibration of each model’s untreated-elongation forecast by region, given a cohort independent of the models’ derivation data. It is a group-level, population-mean trajectory, not individual elongation. The secondary estimand, for a future single-arm trial, is the treatment effect versus the model-predicted counterfactual; only the former is evaluated here.

### Axial-elongation models evaluated

Five published models of untreated axial elongation were implemented and evaluated. The Naduvilath 2025 regional GEE model^13^ was applied with its exact published coefficients for two derivation regions, East Asian (China and Vietnam) and European (United Kingdom, Sweden, and Australia), with separate equations for myopes (baseline SE ≤ −0.50 D) and non-myopes; annual elongation is modelled on the natural-log scale as a function of region, sex, baseline age, a region-by-age interaction, and baseline SE. The Brennan 2024 meta-regression uses age and ethnicity in myopes,^15^ and the Orinda (Jones 2005) and SCORM (Wong 2010) models predict elongation from age alone.^16,17^

Of these, only the regional GEE model covers non-myopes and publishes an individual prediction interval; the remaining three apply to myopes only, as was the entire cohort. Derivation populations and predictors are listed in Table S1 and the equations in Table S2.

No model provides an India-derived stratum, so each candidate equation was applied to Indian children and evaluated: for the GEE model, both East Asian and European strata; for Brennan, both Asian and non-Asian codings. The fifth model (Naduvilath 2023) requires an additional predictor, parental myopia, and was evaluated separately (Supplementary Results S5); it is not part of the released tool. Because elongation decelerates with age, cumulative elongation was integrated as the child aged to the actual elapsed follow-up, then anchored to the observed baseline AL.

### External validation cohort

Three clinical trials (ClinicalTrials.gov NCT06137560, NCT06577948, NCT06692699) at six sites (Appendix) provided de-identified data for untreated myopic children on single-vision correction with no myopia-control treatment; each site had local ethics approval. In total, 242 children (baseline cycloplegic SE ≤ −0.50 D, aged 6 to 14 years) of Chinese, Vietnamese, or Indian ancestry had baseline age, cycloplegic SE, and AL for both eyes, with AL at approximately 6 and 12 months. The right eye was used for the main analysis and the left for a sensitivity analysis. Ancestry denotes ethnic background, not country of residence: Chinese and Vietnamese children fall within the East Asian derivation set, while Indian children, for whom no stratum exists, were evaluated under each candidate equation. Untreated myopic cohort characteristics are given in Table 1.

**Table 1.** Characteristics of the untreated validation cohort (right eyes). Values are means unless stated. SE, spherical equivalent; AL, axial length.

| Group | n | Age, y | Baseline SE, D | Baseline AL, mm | Female, n | 6-mo AL, n | 12-mo AL, n |
| --- | --- | --- | --- | --- | --- | --- | --- |
| Chinese (East Asian, exact) | 53 | 10.1 | −2.22 | 24.45 | 30 | 53 | 0 |
| Vietnamese (East Asian, exact) | 128 | 10.0 | −2.83 | 24.63 | 55 | 117 | 71 |
| Indian (South Asian, provisional) | 61 | 11.2 | −2.42 | 24.00 | 41 | 49 | 0 |
| All | 242 | 10.3 | −2.59 | 24.44 | 126 | 219 | 71 |

### Follow-up visit windows, and outcome metrics

The outcome was observed axial elongation (follow-up minus baseline AL). Predictions were annualised on each child’s actual elapsed follow-up in recorded visit days rather than the nominal month, because visits often ran past schedule: the 6-month visit occurred at 6.6 months on average (202 days) and the 12-month visit at 12.1 months (370 days). A child-visit was analysed only if it had a baseline SE and a baseline and follow-up AL; those missing a baseline were excluded and logged, never on the basis of the outcome.

All metrics are reported per model region and visit, in millimetres, and are defined in Table S2. Bias is predicted minus observed mean elongation (negative denotes under-prediction); the pre-specified threshold is absolute bias <0.03 mm applied to the confidence-interval bounds, a margin about twice biometry repeatability and under a tenth of the annual untreated signal. Root mean squared Error (RMSE) combines bias and scatter. Prediction-interval (PI) coverage is the fraction of eyes whose observed elongation falls within the nominal 95% individual interval, with a pre-specified floor of 0.90.

Uncertainty was quantified by a Wilson interval for coverage, a t-based CI for mean bias, and a child-level cluster bootstrap for residual dispersion; equivalence to zero within ±0.03 mm was tested by two one-sided tests. The recommended treatment-effect estimate, computed by the user and not the tool, is the cumulative absolute reduction in elongation (CARE),^18^ the mean predicted untreated minus the mean observed treated elongation, with its interval combining the treated sampling error and the control’s own confidence interval in quadrature so the control is not treated as error-free.

## Results

### Cohort

The 242 children (mean age 10.3 years; mean baseline SE −2.59 D) were all myopic (241 of 242 at or below −0.50 D; one had a missing baseline refraction). AL at 6 months was available for 219 children and at 12 months for 71. The 12-month follow-up data were entirely from the Vietnamese subgroup, and the Indian and Chinese subgroups had 6-month follow-up only. These imbalances bound the population- and horizon-specific claims below.

### External validation of the models against observed untreated elongation

As the cohort was untreated and entirely myopic, every model was validated directly against observed elongation. For East Asian children, Table 2 reports each model’s mean prediction, observed mean, bias, and RMSE by visit, and Figure 1 relates the fits to each model’s rate-versus-age structure.

**Figure 1.**
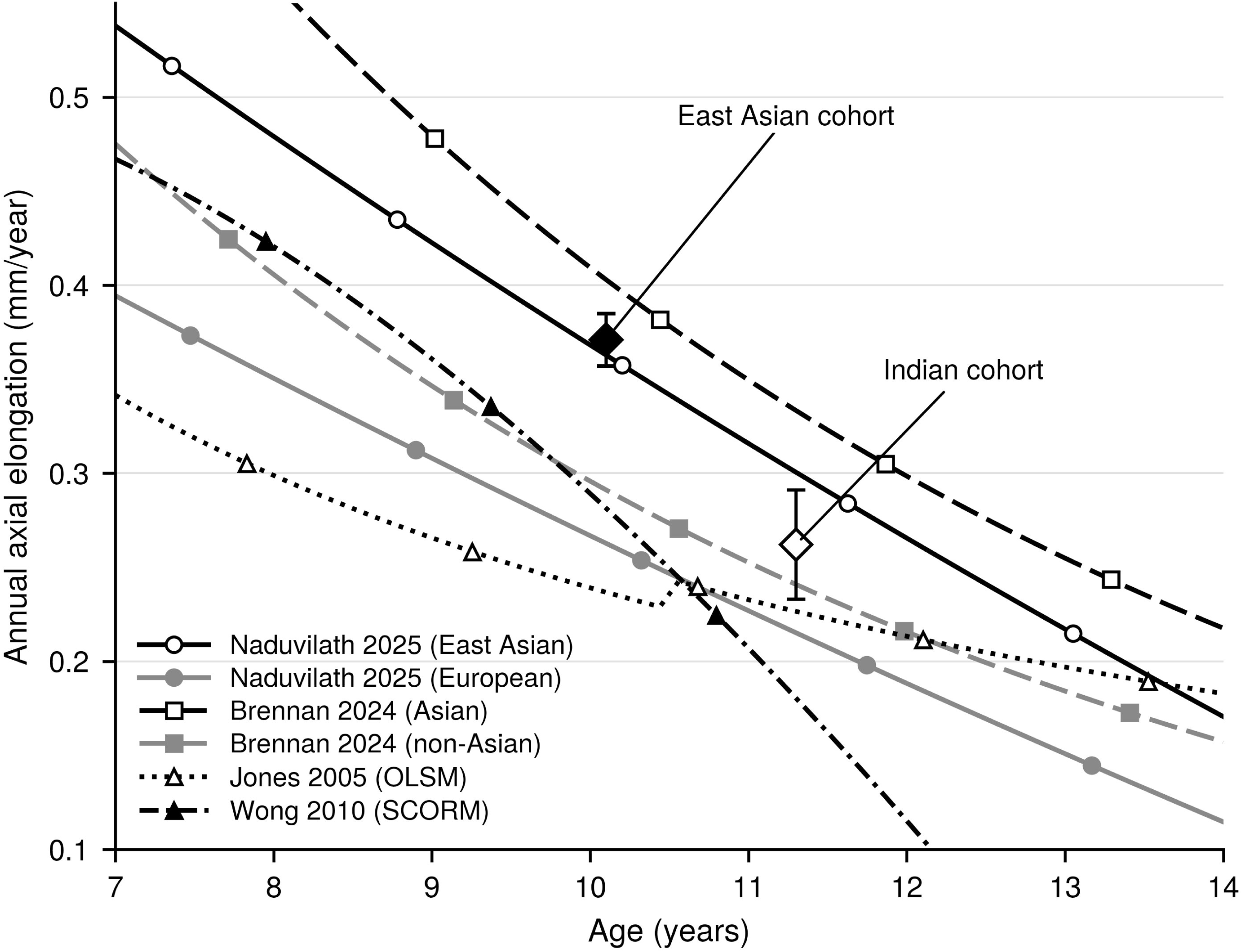
Predicted annual axial-elongation rate against age for each model (myope, baseline SER −2.50 D), with the observed cohort rates overlaid (East Asian, filled circle; Indian, open diamond; mean ± standard error). The East Asian cohort lies on the GEE East Asian curve; the Indian cohort lies between the GEE European and East Asian strata, nearer the East Asian.

**Table 2.** External validation of the four directly comparable models against observed untreated elongation in East Asian children, at actual elapsed follow-up time. Bias is defined as mean predicted minus observed; negative denotes under-prediction. RMSE: Root mean squared error

| Model | Region, visit | n | Mean pred., mm | Mean obs., mm | Bias, mm | RMSE, mm |
| --- | --- | --- | --- | --- | --- | --- |
| Naduvilath 2025 (regional GEE) | East Asian, 6 m | 170 | 0.191 | 0.203 | −0.012 | 0.100 |
| Brennan 2024 (meta-regression) | East Asian, 6 m | 170 | 0.218 | 0.203 | +0.015 | 0.099 |
| Wong 2010 (SCORM) | East Asian, 6 m | 170 | 0.134 | 0.203 | −0.070 | 0.123 |
| Jones 2005 (OLSM) | East Asian, 6 m | 170 | 0.133 | 0.203 | −0.070 | 0.129 |
| Naduvilath 2025 (regional GEE) | East Asian, 12 m | 71 | 0.347 | 0.350 | −0.004 | 0.144 |
| Brennan 2024 (meta-regression) | East Asian, 12 m | 71 | 0.390 | 0.350 | +0.040 | 0.149 |
| Wong 2010 (SCORM) | East Asian, 12 m | 71 | 0.233 | 0.350 | −0.118 | 0.193 |
| Jones 2005 (OLSM) | East Asian, 12 m | 71 | 0.242 | 0.350 | −0.108 | 0.190 |

In East Asian children the Naduvilath’s GEE and Brennan’s meta-regression models were closest to the observed value, within about 0.015 mm at 6 months and 0.040 mm at 12 months, while, age-only models under-predicted by 0.07 to 0.12 mm (Table 2).

The spread across the four models reached 0.04 mm at 12 months, at the edge of the ±0.03 mm margin, so model choice is itself a non-trivial source of uncertainty. For the regional GEE model, the primary comparator, the 6-month East Asian bias was equivalent to zero within ±0.03 mm (−0.013 mm, p = 0.014); at 12 months equivalence was not established (−0.004 mm, p = 0.068, n = 71). Indian children, evaluated under each candidate equation (Table 3, Figure 2), had an observed mean of 0.157 mm at 6 months, falling between the two GEE strata and closest to the East Asian stratum (0.169 mm; +0.012 mm) and the Brennan non-Asian coding (0.142 mm; −0.015 mm); the European stratum under-predicted (0.120 mm; −0.037 mm) and the Brennan Asian coding over-predicted (0.197 mm; +0.040 mm).

**Figure 2.**
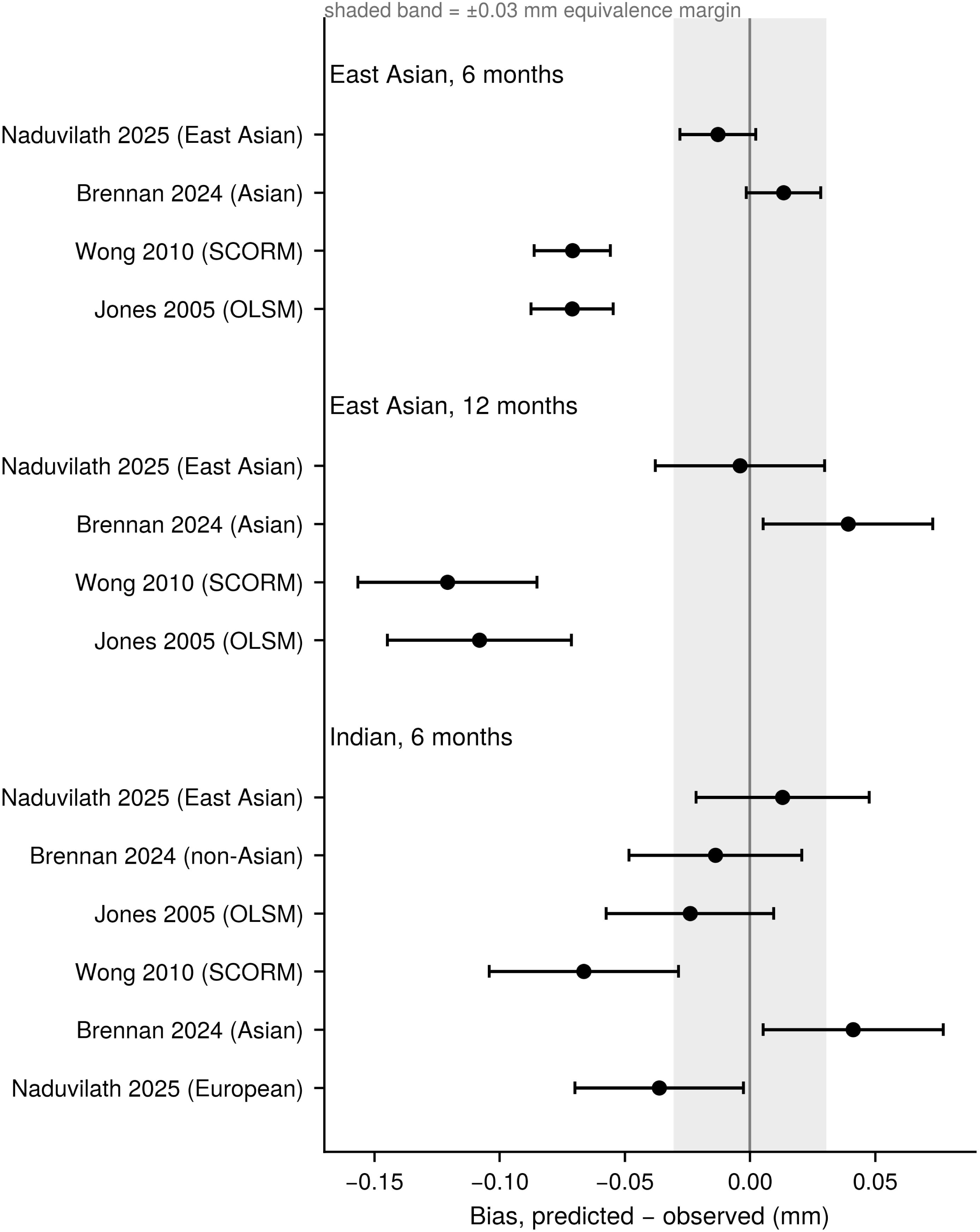
Group-mean bias across all models. Cumulative bias at 6 and 12 months (predicted − observed; mean ± 95% CI) for each model in East Asian children and each candidate equation in Indian children; the shaded band is the ±0.03 mm margin.

**Figure 3.**
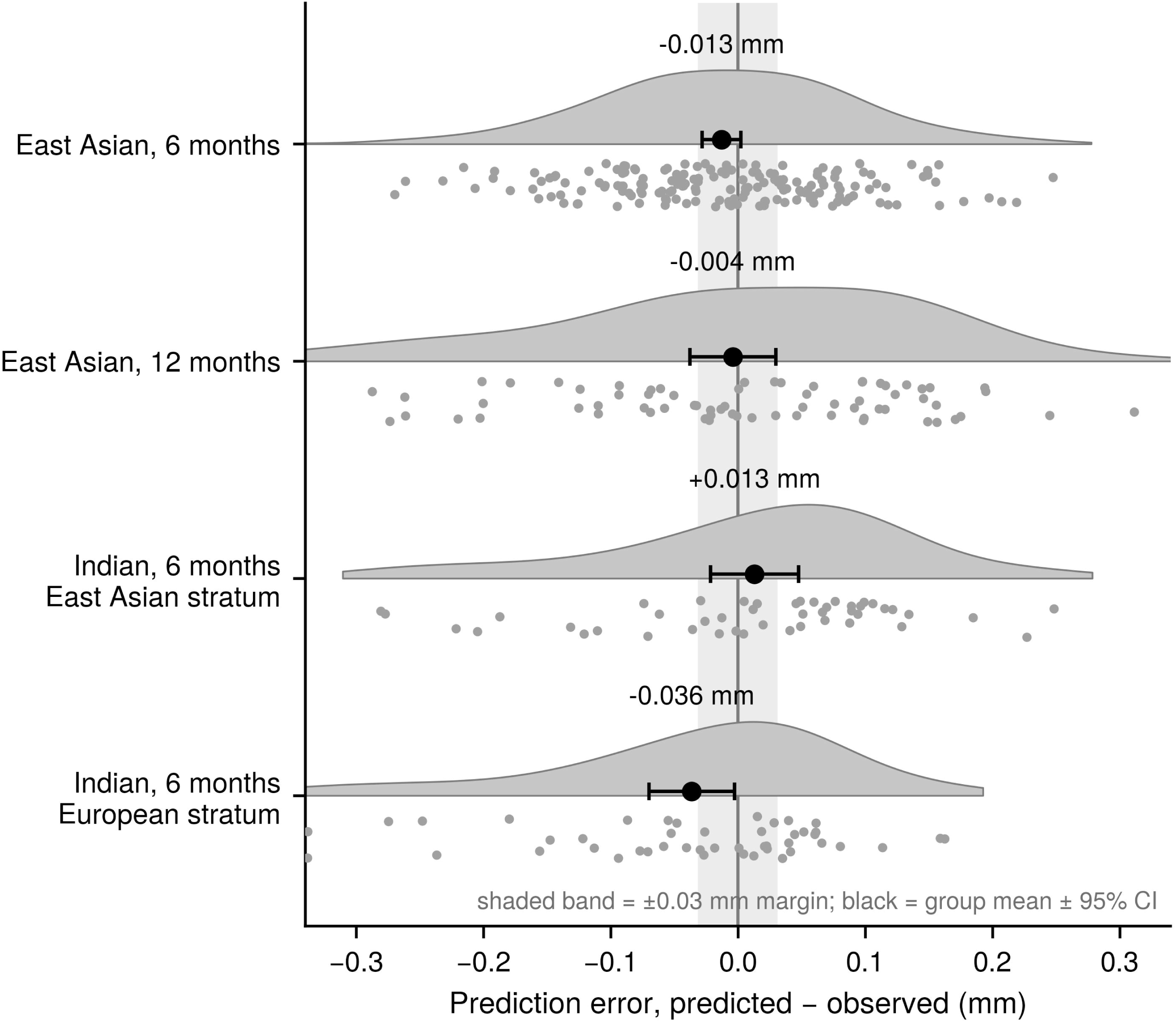
Distribution of prediction error by group (predicted − observed). The half-violin and points show every eye; the black marker is the group mean with 95% CI and the shaded band the ±0.03 mm margin. The group means lie within the margin for East Asian children and for Indian children under the East Asian stratum, while the individual spread is wide of the order of the annual signal; under the European stratum the Indian mean shifts below the margin.

**Table 3.**
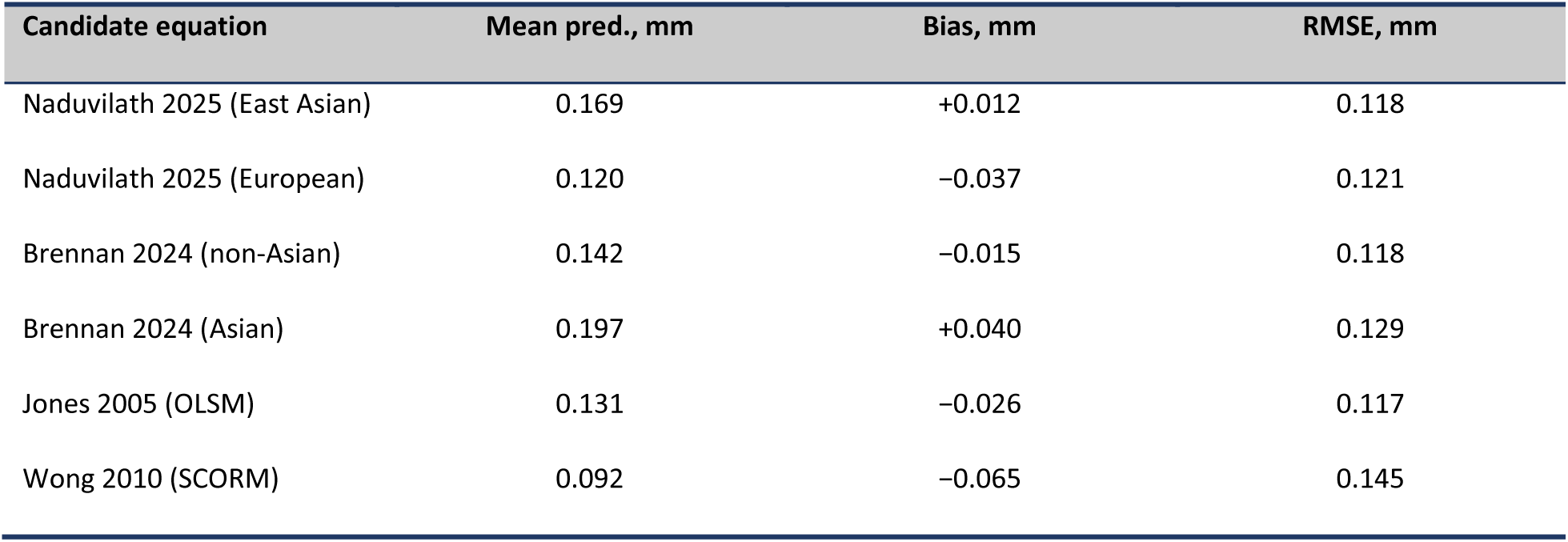
Indian children under each candidate equation (6 months, n = 46; observed mean 0.157 mm). No model has an India-derived stratum, so each candidate is applied and its bias reported. The East Asian stratum of the regional GEE model and the Brennan non-Asian coding reproduce the observed mean most closely. RMSE: Root mean squared error

| Candidate equation | Mean pred., mm | Bias, mm | RMSE, mm |
| --- | --- | --- | --- |
| Naduvilath 2025 (East Asian) | 0.169 | +0.012 | 0.118 |
| Naduvilath 2025 (European) | 0.120 | -0.037 | 0.121 |
| Brennan 2024 (non-Asian) | 0.142 | -0.015 | 0.118 |
| Brennan 2024 (Asian) | 0.197 | +0.040 | 0.129 |
| Jones 2005 (OLSM) | 0.131 | -0.026 | 0.117 |
| Wong 2010 (SCORM) | 0.092 | -0.065 | 0.145 |

The age-only models, under predicted by about 0.026 mm. The Indian East Asian-stratum bias (+0.012 mm) was not equivalent within ±0.03 mm (p = 0.170), consistent with its provisional status.

### Interval coverage and error structure

Interval coverage and calibration could be assessed only for the regional GEE model, the sole bundled model publishing an individual prediction interval (Table 4). Its 95% intervals were too narrow, covering only 0.77 of East Asian observations at 6 months against the 0.90 floor: the mean was accurate, but the stated individual uncertainty was not.

**Table 4.** Interval coverage, calibration slope, and mean accuracy of the regional GEE model with no tuning, at actual elapsed follow-up time.; Pre-specified thresholds: absolute bias <0.03 mm and coverage ≥0.90, applied to the confidence bounds. Coverage is of the nominal 95% prediction interval with a Wilson interval: p = 0.014 (East Asian, 6 months), p = 0.068 (East Asian, 12 months), p = 0.170 (Indian, East Asian stratum).

| Region (routing) | Visit | n | Bias, mm (95% CI) | RMSE | PI coverage (95% CI) | Calib. slope (95% CI) |
| --- | --- | --- | --- | --- | --- | --- |
| East Asian (exact) | 6 m | 170 | -0.013 (-0.028, 0.002) | 0.101 | 0.77 (0.70, 0.83) | 1.24 (0.94, 1.53) |
| East Asian (exact) | 12 m | 71 | -0.004 (-0.038, 0.030) | 0.144 | 0.87 (0.78, 0.93) | 1.06 (0.67, 1.45) |
| S. Asian / Indian (East Asian stratum) | 6 m | 46 | +0.012 (-0.022, +0.046) | 0.118 | 0.74 (0.60, 0.84) | 0.46 (-0.09, 1.02) |

The shortfall was one of scale, not location; standardised residuals were roughly symmetric and centred near zero, yet about 23% fell outside the nominal interval, split across both tails. Visit timing affected the mean, at nominal 6 months the bias was −0.026 mm, but visits occurred at 6.6 months on average, so roughly half of the apparent bias was elapsed growth time rather than model error.

The residual standard deviation of prediction error was 0.142 mm per square-root-year (cluster-bootstrap 95% CI 0.127 to 0.155). With only two horizons this is a two-point fit, the dispersions at 6 and 12 months are consistent with square-root-of-time growth but cannot be distinguished from nearby exponents and do not on their own evidence a random-walk mechanism. Because the published interval scales linearly with time, it understates this dispersion, which is why coverage fell below the floor.

### Sensitivity analyses

The primary East Asian result was tested along five parameters, summarised in Table S3. In every case the group-mean bias remained well within the ±0.03 mm margin, and point estimates moved by amounts at or below biometry repeatability, so no design or analysis choice materially affects the conclusions.

Briefly, the left-eye analysis reproduced the right-eye result (East Asian bias −0.012 mm at 6 months, +0.010 mm at 12 months; between-eye difference ≤0.013 mm), and reversing the numeric sex code changed the bias by less than 0.002 mm; both are detailed in Table S3. Using each child’s actual elapsed follow-up rather than the nominal visit label shifted the 6-month bias from −0.013 to −0.026 mm, confirming that honouring the realised interval matters at the sub-0.02 mm level and is the appropriate primary analysis. Daily rather than monthly integration changed the bias by ≤0.002 mm, and a leave-one-site-out analysis kept the 6-month bias between −0.004 and −0.018 mm around the full-cohort −0.013 mm, so no single site drives the result.

## Discussion

In an independent untreated paediatric myopic cohort, the recent published models predicted mean axial elongation without meaningful bias in East Asian myopes, the population for which they were derived, satisfying a core requirement of a virtual control. Two findings stand out: the published individual prediction intervals were too narrow, covering 0.77 rather than 0.95, so a model can have the right mean, and the wrong spread; and the residual dispersion grew with the square root of follow-up rather than linearly, so the published linear interval understates the spread at these horizons.

### How each model fits the untreated cohort

Applying each model to the same untreated children showed why they differ (Table S2, Figure 1). Three factors separate them: whether the model requires ethnicity, how steeply its rate declines with age, and whether it requires refraction. The Naduvilath 2025 GEE model has an East Asian intercept (+0.147 on the log-rate) and an East Asian age interaction (−0.007 per year),^13^ so at 10 to 11 years its East Asian stratum elongates about 40% faster than its European counterpart, reproducing the East Asian cohort almost exactly (bias −0.013 mm); for Indian children it was the choice of stratum, not the model, that decided the fit. The Brennan meta-regression leaned the same way through a single Asian versus non-Asian term (+0.325 on the log-rate) and a steeper age slope (−0.158 per year), so its Asian coding tracked the East Asian mean at 6 months yet overshot slightly at 12 months.

The older ultrasound-era models use age alone (Orinda: 0.2391/age; SCORM:0.273*Age* − 0.106*Age*\*ln*(*Age*)) and were fitted to historical cohorts that grew more slowly than contemporary myopes, so they sit below the observed rate at every age and under-predict by 0.07 to 0.12 mm. The lesson is not that the older models were wrong in their era, but that virtual control should reach for the most recent, population-matched model and carry the across-model spread, which touched the ±0.03 mm margin at 12 months, as genuine uncertainty.

### Effect of reporting the visit-timing

One methodological point generalises to any use of annualised growth models as counterfactual controls: follow-up must be annualised on actual elapsed time, not the nominal visit label. Here the nominal 6-month visit occurred at 6.6 months on average, and predicting at 6.0 months produced roughly half of the apparent 6-month bias, a 0.02 mm artefact against a 0.03 mm margin. Any external-control analysis that annualises growth should use recorded visit dates and report the realised interval alongside the nominal one.

### External benchmarking against trial control arms

A model-based comparator is credible only if the untreated rates it reproduces match contemporary trial control arms. Against the LAMP placebo arm (0.41 mm/year at ages 4 to 12),^19^ the older East Asian children here elongated about 0.35 mm/year, the expected ordering since younger eyes elongate faster; that rate sits between the single-vision controls of DIMS (0.28 mm/year)^3^ and HAL (0.35 mm/year)^5^ in same-age Chinese children, while the South Asian rate of about 0.28 mm/year sits at the slower end. This supports plausibility rather than providing formal validation: rates must be age-matched, and genuine untreated arms are scarce, since even the ATOM2 0.01% arm is an active treatment rather than an untreated control,^20^ precisely the gap a validated comparator is meant to fill.

### The South Asian gap

The South Asian result marks the boundary of current evidence. No published model has an India-derived stratum, so Indian children were evaluated under each candidate. Their observed elongation (about 0.26 mm/year) falls between the European and East Asian GEE strata and is closest to the East Asian stratum, which reproduces it with least bias alongside the Brennan non-Asian coding, while the European stratum under-predicts. Their progression therefore resembles East Asian more than European myopes, consistent with high axial-progression rates in urban Indian cohorts. The best candidate here nonetheless rests on 46 eyes at a single 6-month visit from one cohort and is provisional; a dedicated South Asian model, or validation against a larger independent cohort, is the clear next step. Centile-curve comparators were rejected because published analyses show AL centiles underestimate myopic eye growth, describing attained magnitude rather than the rate a control arm needs.^10,12^

### Constancy, causal assumptions, and why the tool must be considered supportive

The tool’s strength is not novelty, since none of the models are original here, but transparency: several published models unified in one reproducible place, with multi-model concordance, an integral validation workflow, and a privacy model that removes the data-governance barriers usual to external controls. Constancy, the assumption that a future untreated cohort would progress as the models predict, cannot be verified within a single-arm study that contains no untreated children; random error in the counterfactual mean is bounded by the treated sample size, but systematic error from secular trends, behaviour, or ancestry mix is not.

The tool must therefore be considered supportive evidence. Because the control is a parametric model rather than patient records, patient-level external-control methods such as propensity weighting do not apply; the appropriate inference is a paired comparison of each treated child against the model counterfactual, with uncertainty from the treated sample and, ideally, multiple imputations across the bundled models. The path to stronger evidence is a hybrid design that retains a small concurrent randomised untreated reference, large enough to check constancy locally but far smaller than a full control arm. The tool is a group-level instrument: calibration slopes are consistent with unity but imprecise, and per-eye RMSE of 0.10 to 0.14 mm is of the order of the annual signal, so no individual-level claim should be made. A control arm requires only an unbiased population-mean trajectory, which the validation establishes for East Asian myopes; the wide individual scatter is not part of the control the tool supplies.

### Using the software

The tool is a single self-contained, offline browser application (Supplementary Software version 0.2.1). In a single-arm study it supplies the untreated control while the treatment effect remains the user’s step: model choices (ethnicity routing, a single primary model, timepoints, and acceptance thresholds) are pre-specified; baseline data are entered one row per child; and the tool returns, per child and horizon, the predicted untreated change with its confidence interval of the mean, propagated into the effect (Figure 4). It returns no individual prediction interval, because the published one under-covered against the 0.90 floor. Full operating instructions, input and output formats, the offline and self-test guarantees, and the recommended analysis workflow are given in Supplementary material S4.

**Figure 4.**
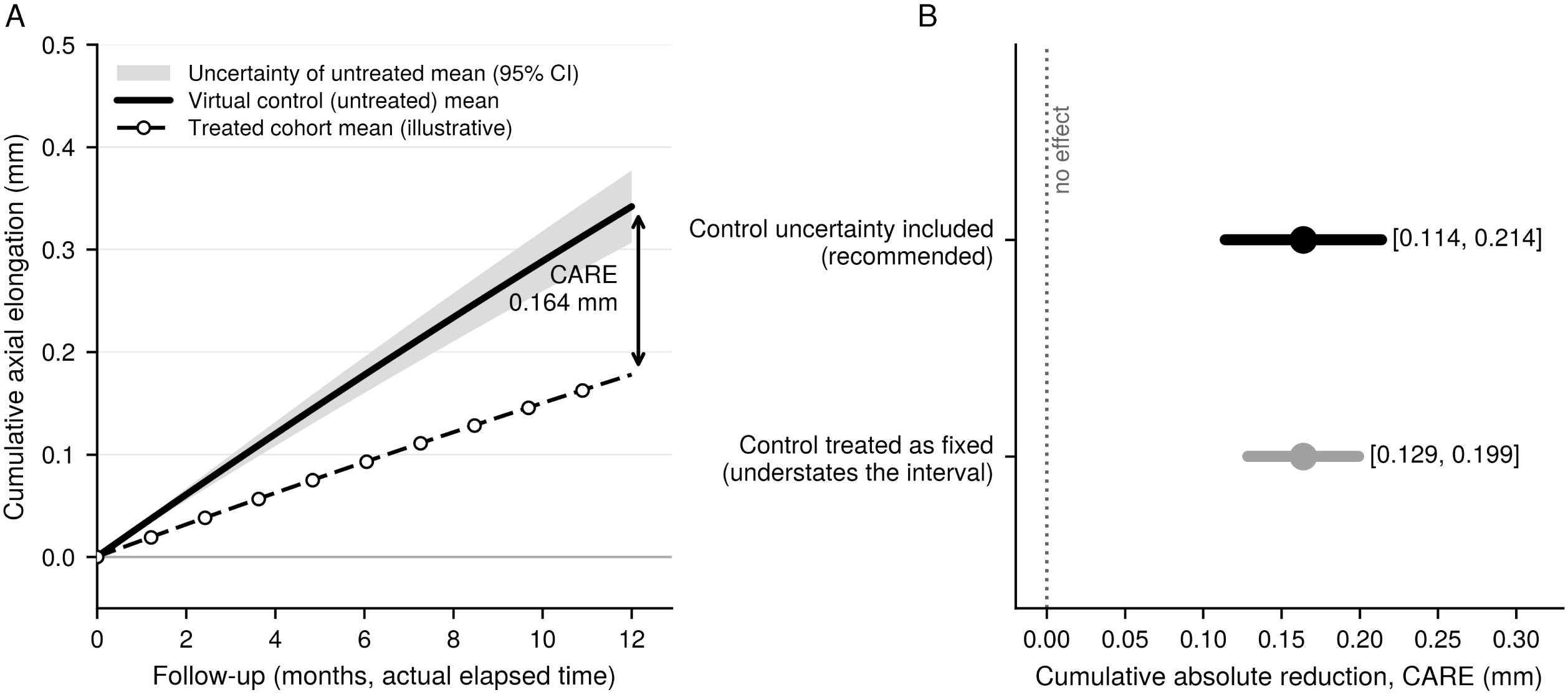
Virtual control under its own uncertainty (illustrative worked example). (A) The mean untreated trajectory with the confidence band of the untreated mean, the only interval the tool supplies; the individual prediction interval is omitted, as it under-covered in validation. The treated cohort falls below, and the cumulative absolute reduction in elongation (CARE) at 12 months is the vertical separation. (B) The same CARE with its 95% interval built two ways: with the control fixed [0.129, 0.199] and with the control’s confidence interval added in quadrature, the recommended [0.114, 0.214]. The point estimate is identical; both clear the no-effect line.

## Limitations

Several limitations bound these conclusions. Most important, external validity requires the validation cohort to be independent of the models’ derivation data, yet the East Asian evidence, and all 12-month evidence, comes from Vietnamese children, and the GEE East Asian stratum was derived partly in Vietnam. The constancy assumption cannot be verified in a treated cohort. Validation is limited to 6 and 12 months, so horizon and ancestry are confounded, and longer intervals are extrapolations of a two-point time law. The largest single analysis includes 170 of 242 children, and the cohort contained no non-myopes, so the non-myopic interval is unvalidated. Calibration slopes are consistent with unity but estimated with little predictor variance, so individual-level discrimination cannot be assessed. Finally, coverage is bounded: only the East Asian stratum was validated directly; South Asian children remain provisional; and the European stratum, though available in the tool, was not tested against an untreated European cohort and remains provisional. Ancestry itself is an imprecise, self-reported proxy under which mixed populations are poorly represented.

## Conclusions

An open-source tool that applies several published axial-elongation models in parallel, with multi-model concordance and a built-in validation workflow, offers myopia researchers a scientifically defensible, privacy-preserving alternative to an untreated randomised arm, suitable as supportive evidence. The predictive models are the published work of their original authors; the contribution here is to implement them faithfully in one place, test them against real untreated data, and release everything so the result can be reproduced. The tool is a group-level, population-mean instrument rather than an individual predictor, and its trustworthiness rests not on any single model but on external validation against real untreated data and concordance across independent published models. The aim is shared scientific infrastructure that makes untreated and single-vision control arms in children the exception rather than the rule.

## Data and code availability

The tool, comprising all bundled models, the validation workflow, the template adapter, and the templates, is released as open-source Python under the GNU General Public License v3.0, with pinned dependency versions, a citation file, and submitted as a supplementary software. The tool runs entirely in the user’s web browser with the analysis executed locally and no data transmitted to any server, and the released source a reader inspects is the exact code that executes. A single command regenerates every table and figure in this paper from the source data under a fixed random seed. Aggregate validation metrics are published in full, and an accompanying spreadsheet provides the per-eye data, formula-driven summary tables, and figures for independent reviews. Individual-level untreated data are available subject to the contributing site’s governance and ethics approvals.

## Supporting information

Supplementary software v0.2.1

## Data Availability

All data produced in the present study are available upon reasonable request to the authors. The relevant software tool is available in supplementary file.

## Acknowledgements

The authors gratefully acknowledge the broader team at nthalmic whose direct and indirect contributions made the underlying clinical trials, data collection, operational execution, software validation, quality review, regulatory support, and study oversight possible. We sincerely thank the following colleagues for their support and contribution: Associate Professor Klaus Ehrmann, Professor Kah Ooi Tan, Dr Cathleen Fedtke, Dr Karen Lahav, Ms Jennie Diec, Mr Darrin Falk, Dr Eon Kim, Ms Kathleen Laarakkers, Dr Fabian Conrad, Ms Claudia Bishop, Ms Aishwarya Porje, and Mr Dicky Yip.

The authors also acknowledge the investigators, site personnel, trial coordinators, and participating children and families involved in the contributing clinical trials, without whom this research would not have been possible. The study utilised data from three nthalmic-sponsored clinical trials conducted across six international sites.

## Artificial Intelligence Use and Disclosure Statement

The authors disclose that Microsoft 365 Copilot was used to assist with portions of the literature review, manuscript editing, software development support, code review, and technical documentation. AI-generated outputs were used solely as an aid to the research and development process and were subject to critical review, verification, modification, and approval by the authors.

The authors accept full responsibility for the accuracy, integrity, interpretation, and originality of the work presented. All scientific conclusions, analyses, software implementations, and manuscript content were reviewed and validated by the authors. This disclosure is provided in the spirit of transparency and to inform readers of the tools used during the conduct of the research and preparation of the manuscript.

