## Supplementary file for "Virtual control arms for paediatric myopia trials: external validation of axial-elongation models"

**Supplementary material S1**

**Table S1** The bundled axial-elongation models, their derivation populations, and their predictors. All coefficients are the exact published values; the tool self-validates each model against its source estimates on start-up. No model is nominated as primary; all are applied in parallel.

| **Model (reference)** | **Derivation population** | **Predictors** | **Applies to** | **Notes** | **Source** |
| --- | --- | --- | --- | --- | --- |
| Naduvilath 2025 (regional GEE)¹ | East Asian (China, Vietnam) and European (UK, Sweden, Australia) | Baseline age, region/ethnicity, sex, region × age, baseline SE | Myopes and non-myopes | Regional GEE; the only bundled model that provides an individual prediction interval | Ophthalmic Physiol Opt 2025; 45:135-151, Table 3 |
| Naduvilath 2023 (parental myopia)² | Asian children | Baseline age, number of myopic parents | Myopes and non-myopes | Adds a parental-myopia input; optional | Ophthalmic Physiol Opt 2023;43(5):1160–1168 |
| Brennan 2024 (meta-regression)³ | Meta-regression of 78 studies; Asian vs. non-Asian | Mean age at evaluation, ethnicity code | Myopes | Meta-regression | Optom Vis Sci 2024;101(8):497–507, Table 1 |
| Jones 2005 (OLSM)⁴ | Predominantly White United States children (Orinda) | Age only | Myopes | Age-only growth curve | Invest Ophthalmol Vis Sci 2005;46:2317–2327 |
| Wong 2010 (SCORM)⁵ | Singaporean children | Age only (axial length model) | Myopes | Age-only growth curve | Invest Ophthalmol Vis Sci 2010;51(3):1341–1347 |

**Notation:** aₜ = age (years) at the evaluation point; SE = baseline cycloplegic spherical equivalent (D); s = 1 if male, else 0; E = 1 if the model region is East Asian, else 0; r = ethnicity code (1 if Asian, else 0); t = elapsed follow-up time (years); z = 1.96 for a 95% interval; λ = population multiplier (fixed at 1 in the released tool). All rates are instantaneous annual axial elongation (mm/year) and are integrated in monthly steps while aging the child.

**Supplementary material S2**

**Table S2** Model and analysis equations exactly as implemented in the released code (virtual_control.py, sensitivity_models.py, and the inference scripts). Coefficients are the exact published values from the respective sources in Table S1.

| **Quantity** | **Formula, as implemented** | **Notes** |
| --- | --- | --- |
| Naduvilath 2025 (regional GEE) rate, myopes (SE ≤ −0.50 D) | $rate\left( a_{t} \right)=\exp\left( \eta\right)- 1,$  $\eta= 0.537 + 0.147E + 0.004s - 0.032a_{t}- 0.007Ea_{t}- 0.007SE$ | Regional GEE model; back transformed from the log scale |
| Naduvilath 2025 (regional GEE) rate, non-myopes (SE > −0.50 D) | $rate\left( a_{t} \right)=\exp\left( \eta\right)- 1,$  $\eta= 0.263 + 0.136 E - 0.003 s - 0.016 a_{t}- 0.007 E a_{t}- 0.002 SE$ | Not used by this cohort (all myopic) |
| Brennan 2024 (meta-regression) rate | $rate\left( a_{t} \right)=\exp\left( 0.362 - 0.158 a_{t}+ 0.325 r \right)$ | Myopes only |
| Jones 2005 (OLSM) rate | $rate\left( a_{t} \right)= \left\{ \begin{aligned} \frac{2.391}{a_{t}}& a_{t}\leq10.5 \\ \frac{2.560}{a_{t}}& a_{t}> 10.5 \end{aligned} \right.$ | Myopes only; age-only |
| Wong 2010 (SCORM) rate | $rate\left( a_{t} \right)=\max\left( 0.273 a_{t}- 0.106a_{t}\ln\left( a_{t} \right), 0 \right)$ | Myopes only; axial length model |
| Cumulative elongation to month m | E(m) = λ · Σ (k = 0 … m−1) rate (a + k/12) / 12 | Monthly integration of the age-varying rate |
| Predicted axial length | AL_pred(m) = AL_baseline + E(m) | Anchored at the child’s baseline AL |
| Observed elongation | ΔAL = AL(t) − AL(baseline) | Annualised on actual elapsed time t, not nominal months |
| Prediction interval (published, linear) | half-width = z · σ_PI · t · λ; σ_PI = 0.11 (myope), 0.08 (non-myope) mm/year | Individual 95% interval; default behaviour |
| Confidence interval of the mean | half-width = z · σ_CI · t · λ; σ_CI = 0.018 (myope), 0.011 (non-myope) mm/year | Interval on the population mean |
| Bias | bias = mean(predicted) − mean(observed) | Threshold abs(bias) < 0.03 mm on the CI bounds |
| RMSE; MAE | RMSE = √mean ((pred − obs)^²); MAE = mean (abs (pred − obs)) | Per-eye error size |
| Prediction-interval coverage | fraction with (pred − half-width) ≤ obs ≤ (pred + half-width) | Ideal 0.95; floor 0.90; Wilson 95% CI |
| Calibration slope, intercept | ordinary least squares of observed on predicted | Ideal slope 1, intercept 0 |
| TOST equivalence vs. ±0.03 mm | equivalent if both one-sided t-tests reject at 0.05 | Confines the bias within the margin |
| CARE (treatment effect) | CARE = mean (predicted untreated) – mean (observed treated) | Positive = benefit; paired child-level bootstrap CI |
| Percentage reduction | percent = CARE / mean (predicted untreated) | Reported but de-emphasised; denominator shrinks with age |

**Supplementary material S3**

**Table S3** Sensitivity analyses of the primary East Asian validation. Group-mean bias (predicted minus observed axial elongation, mm) for the Naduvilath 2025 regional GEE model, East Asian stratum, under each design or analysis choice. The primary analysis is the right eye at each child's actual elapsed follow-up with daily integration (n = 170 at 6 months, n = 71 at 12 months). All variants remain within the ±0.03 mm equivalence margin.

| Sensitivity analysis | EA 6-mo bias (mm) | EA 12-mo bias (mm) | Change vs primary, 6 m (mm) | Change vs primary, 12 m (mm) |
| --- | --- | --- | --- | --- |
| Primary: right eye, actual time, daily integration (n=170/71) | -0.013 | -0.004 | 0.000 | 0.000 |
| 1. Analysis eye: left eye | -0.012 | +0.009 | 0.001 | 0.013 |
| 2. Sex coding reversed | -0.013 | -0.005 | 0.000 | 0.002 |
| 3. Nominal follow-up time (6/12 m labels) | -0.026 | -0.006 | 0.013 | 0.002 |
| 4. Monthly (vs daily) integration | -0.012 | -0.002 | 0.001 | 0.002 |
| 5. Leave-one-site-out (EA 6-m, range) | -0.0179 to -0.0036 | - | 0.009 | - |

**Supplementary material S4**

**Software availability and use:**

The tool is a single self-contained folder [Supplementary Software: Supplementary_Software_v0.2.1]. It runs entirely in a standard web browser, computing on the user's own machine with no account, connection, or server, and it makes no outbound network request, which can be verified from the page's Content-Security-Policy and by running it with the network disconnected. To run it, the user unzips the folder and double-clicks run_windows.bat (Windows) or runs run_mac_linux.sh (macOS or Linux); the launcher serves the local files to the default browser using Python 3, installed once from python.org where absent, the only step needing the internet, after which the tool runs fully offline. Baseline data are uploaded as an Excel workbook (.xlsx; columns id, ethnicity, gender, age, baseline_SER, baseline_AL, stratum), and the predicted untreated control is returned as an Excel workbook (out_predictions.xlsx), one prediction per child and visit with the confidence interval of the mean where the selected model provides one; treated outcome data are never read and never leave the machine. A built-in self-test reproduces the published validation values for all implemented models in the user's own browser, confirming numerical fidelity on the machine in use.

**Using the software in a future single-arm study:**

Used in a single-arm study, the tool supplies the untreated control; the treatment effect is the user's own step. First, pre-specify the model choices before examining data: the routing by ethnicity, the single primary comparator model, the analysis timepoints, and the acceptance thresholds for validity (for example a bias within 0.03 mm, the margin used here, and prediction-interval coverage of at least 0.90). If no validated model is derived for the cohort's ancestry, the best-fitting candidate is selected against those thresholds and marked provisional. Enter one row per child of baseline data only: ethnicity, sex, baseline age, cycloplegic spherical equivalent, and axial length. Ethnicity is the child's ancestry, not country of residence, and is required; a blank row is not predicted, and for an ancestry with no derived model, South Asian in particular, the user must select the stratum, East Asian or European, which flags the result provisional. The tool then returns, per child and horizon, the predicted untreated change and its confidence interval of the mean, the precision of the average untreated trajectory a control arm requires; it returns no individual prediction interval, because the published one under-covered against the 0.90 floor and would overstate the certainty about any single child. Finally, using the pre-specified metric, the user contrasts the observed treated change against the predicted untreated change, propagating the control's own uncertainty (the confidence interval of the mean) into the effect rather than treating the control as fixed (Figure 4), and reports the concordance across the pre-specified model choices as a sensitivity check.

**Supplementary Results S5.**

**Parental myopia as a candidate predictor:**

Parental myopia strongly predicts myopia onset, and one model (Naduvilath 2023, Chinese) conditions on it, so we tested directly whether it improves the untreated prediction. In Chinese children the parental-myopia and regional GEE models were indistinguishable (RMSE 0.092 vs 0.094 mm; bias within 0.001 mm). Regressing the GEE residual on the number of myopic parents gave slopes of +0.010 mm per parent at 6 months (95% CI −0.011 to +0.030) and +0.019 mm at 12 months (−0.025 to +0.063). These intervals include the small effect the 2023 coefficients imply, about 0.015 mm per parent, but the analysis is underpowered for an effect of that size (power approximately 0.31 at 6 months and 0.10 at 12 months), so such an effect can be neither detected nor excluded. This is consistent with the 2023 report that the effect is larger in non-myopes than in myopes, that is, that parental myopia acts more on onset than on progression once myopia is established. Parental myopia is therefore not part of the released model set; it can be revisited in cohorts powered to resolve an effect of this magnitude.
