## Supplementary software v0.2.1 for "Virtual control arms for paediatric myopia trials: external validation of axial-elongation models": app.js

```
// Virtual Control Arm Tool — browser glue.
// Kept in an external file so the page needs no 'unsafe-inline' in its CSP.
// OFFLINE BUILD: the Pyodide runtime and packages are vendored under ./pyodide/,
// so the page makes no external request. See README.md.
const PYODIDE_BASE = "./pyodide/";

const els = {
  dot: document.getElementById('dot'),
  status: document.getElementById('statusText'),
  file: document.getElementById('file'),
  run: document.getElementById('run'),
  selftest: document.getElementById('selftest'),
  model: document.getElementById('model'),
  ethnicity: document.getElementById('ethnicity'),
  downloads: document.getElementById('downloads'),
  preview: document.getElementById('preview'),
  log: document.getElementById('log'),
  ver: document.getElementById('ver'),
};
let pyodide = null;

function setStatus(text, state){
  els.status.innerHTML = text;
  els.dot.className = 'dot' + (state ? ' ' + state : '');
}
function showLog(text, cls){
  els.log.style.display = 'block';
  els.log.innerHTML = cls ? `<span class="${cls}">${text}</span>` : text;
}

// The Python driver: mirrors run_my_trial.main() but takes an in-memory Excel
// file and an optional ethnicity override. It writes the outputs as .xlsx
// workbooks to the in-browser filesystem and returns metadata as JSON; the
// browser reads those workbook bytes back for download.
const DRIVER = `
import pandas as pd, io, json, math
import virtual_control as vc

def _clean(rec):
    # JSON has no NaN; the point-estimate models leave ci_low/ci_high blank.
    return {k: (None if isinstance(v, float) and math.isnan(v) else v) for k, v in rec.items()}

def run_tool(path, model_name, override_ethnicity):
    df = pd.read_excel(path)
    subjects, excl = [], []
    for r in df.itertuples():
        eth = override_ethnicity or getattr(r, "ethnicity", None)
        if pd.isna(eth) or str(eth).strip() == "":
            excl.append({"id": getattr(r,"id","(no id)"),
                         "reason": "ethnicity (ancestry) is required; country of residence is not accepted"})
            continue
        subjects.append({"id": r.id, "ethnicity": eth, "gender": r.gender,
                         "age": r.age, "baseline_SER": r.baseline_SER, "baseline_AL": r.baseline_AL,
                         "stratum": getattr(r,"stratum",None)})
    pred = vc.predict_untreated(subjects, exclusions=excl, model_name=model_name)
    outputs = []
    label = vc.MODEL_LABELS[model_name]
    vc.write_output(pred, "out_predictions.xlsx", model_label=label)
    outputs.append("out_predictions.xlsx")
    if excl:
        vc.write_output(pd.DataFrame(excl), "out_needs_input.xlsx",
                        model_label=label, sheet_name="needs_input")
        outputs.append("out_needs_input.xlsx")
    out = {
        "model_label": vc.MODEL_LABELS[model_name],
        "ships_ci": (model_name == "naduvilath2025"),
        "outputs": outputs,
        "n_pred": int(pred["id"].nunique()) if not pred.empty else 0,
        "n_excluded": len(excl),
        "preview": [_clean(r) for r in pred.head(12).to_dict(orient="records")] if not pred.empty else [],
    }
    return json.dumps(out)
`;

async function boot(){
  try{
    pyodide = await loadPyodide({ indexURL: PYODIDE_BASE });
    setStatus("Loading scientific packages (numpy, pandas)…", "load");
    // numpy + pandas resolve their deps (python-dateutil, pytz, six) from the
    // local lock file; nothing is fetched from the network.
    await pyodide.loadPackage(["numpy","pandas"]);
    // Pure-Python .xlsx support (openpyxl + et_xmlfile), unpacked locally so no
    // micropip / PyPI call is needed. Required: the tool reads and writes .xlsx.
    const zbuf = new Uint8Array(await (await fetch("pyodide/purepy.zip")).arrayBuffer());
    pyodide.unpackArchive(zbuf, "zip");
    pyodide.runPython("import sys\nif '' not in sys.path: sys.path.insert(0, '')");
    pyodide.runPython("import openpyxl");
    setStatus("Loading the virtual-control engine…", "load");
    // Both engine modules are loaded unmodified; sensitivity_models is used by the
    // non-GEE models (Brennan/OLSM/SCORM), which virtual_control imports on demand.
    for(const mod of ["sensitivity_models","virtual_control"]){
      const src = await (await fetch("src/" + mod + ".py")).text();
      pyodide.FS.writeFile(mod + ".py", src);
    }
    await pyodide.runPythonAsync("import virtual_control as vc");
    await pyodide.runPythonAsync(DRIVER);
    // Show the version and release date, read live from the engine (single source),
    // so the header can never disagree with the code that actually runs.
    try{
      const v = pyodide.runPython("vc.__version__ + '|' + vc.__release_date__").split("|");
      if(els.ver) els.ver.textContent = "Version " + v[0] + " · released " + v[1];
    }catch(_){ /* non-fatal: header label only */ }
    setStatus("Ready. Choose a model, then upload your baseline file.", "ok");
    els.file.disabled = false; els.selftest.disabled = false;
    els.file.addEventListener('change', () => { els.run.disabled = !els.file.files.length; });
  }catch(e){
    setStatus("Failed to load the engine: " + e.message, "err");
    console.error(e);
  }
}

els.run.addEventListener('click', async () => {
  if(!pyodide || !els.file.files.length) return;
  els.run.disabled = true; els.downloads.innerHTML = ""; els.preview.innerHTML = "";
  setStatus("Processing your file locally…", "load");
  try{
    const f = els.file.files[0];
    const buf = new Uint8Array(await f.arrayBuffer());
    pyodide.FS.writeFile("input.xlsx", buf);
    pyodide.globals.set("_path", "input.xlsx");
    pyodide.globals.set("_model", els.model.value);
    pyodide.globals.set("_override", els.ethnicity.value || "");
    const res = JSON.parse(await pyodide.runPythonAsync("run_tool(_path, _model, _override)"));
    // read the .xlsx workbook bytes written by the engine and offer them for download
    for(const fn of res.outputs){
      const bytes = pyodide.FS.readFile(fn);   // Uint8Array
      addDownload(fn, bytes);
    }
    if(res.preview.length) renderPreview(res.preview);
    const ciNote = res.ships_ci ? "confidence interval of the mean" : "point estimate; no published confidence interval";
    let msg = `Done. Model: <b>${res.model_label}</b> (${ciNote}). ${res.n_pred} child(ren) predicted`;
    if(res.n_excluded) msg += `, ${res.n_excluded} row(s) need input (see out_needs_input.xlsx)`;
    setStatus(msg + ".", "ok");
  }catch(e){
    setStatus("Error while processing: " + e.message, "err");
    console.error(e);
  }finally{
    els.run.disabled = false;
  }
});

els.selftest.addEventListener('click', async () => {
  setStatus("Running the engine self-test in your browser…", "load");
  try{
    const code = [
      "import io, contextlib, virtual_control as vc",
      "buf = io.StringIO()",
      "with contextlib.redirect_stdout(buf):",
      "    ok = vc.self_test()",
      "str(ok) + chr(10) + buf.getvalue()"
    ].join("\n");
    const out = await pyodide.runPythonAsync(code);
    const passed = out.startsWith("True");
    const body = out.split("\n").slice(1).join("\n")
        .replace(/\[PASS\]/g,'<span class="pass">[PASS]</span>')
        .replace(/\[FAIL\]/g,'<span class="fail">[FAIL]</span>');
    els.log.className = 'log selftest';
    showLog(body || out);
    setStatus(passed
      ? "Self-test PASSED in your browser: the primary model matches the published Naduvilath 2025 Table 4, and all four shipped models reproduce their validated values."
      : "Self-test FAILED — do not use these results; please report this.", passed ? "ok" : "err");
  }catch(e){
    setStatus("Self-test could not run: " + e.message, "err");
  }
});

function addDownload(filename, data){
  // data is a Uint8Array (.xlsx bytes) or a string
  const type = filename.toLowerCase().endsWith('.xlsx')
    ? 'application/vnd.openxmlformats-officedocument.spreadsheetml.sheet'
    : 'application/octet-stream';
  const url = URL.createObjectURL(new Blob([data], {type}));
  const a = document.createElement('a');
  a.href = url; a.download = filename; a.textContent = "Download " + filename;
  els.downloads.appendChild(a);
}
function renderPreview(rows){
  const cols = ["id","region","model_status","model","month","predicted_delta_AL","ci_low","ci_high"]
      .filter(c => c in rows[0]);
  let html = '<table class="prev"><thead><tr>' + cols.map(c=>`<th>${c}</th>`).join('') + '</tr></thead><tbody>';
  for(const r of rows){
    html += '<tr>' + cols.map(c=>{
      let v = r[c];
      if(typeof v === 'number') v = Number.isInteger(v) ? v : v.toFixed(4);
      if(v === null || v === undefined || (typeof v==='number'&&isNaN(v))) v = '—';
      return `<td>${v}</td>`;
    }).join('') + '</tr>';
  }
  html += '</tbody></table><p class="hint">Preview of the first rows. Full result is in the download above.</p>';
  els.preview.innerHTML = html;
}

boot();
```
