## Supplementary software v0.2.1 for "Virtual control arms for paediatric myopia trials: external validation of axial-elongation models": ADAPTATION_NOTES.md

# Adapting the trial export to the virtual-control tool

Source export: `Virtual_Control_data_2026-07-08.xlsx` (sheet "Virtual Control Data", 242 children).
New file: `adapt_template.py`. Modified file: `validate_model.py` (now tolerant of incomplete baselines).

## What the export contained

242 children, one row each, BOTH eyes side by side:
`ID, Ethnicity, Sex, BL_age, BL_SER_OD, BL_SER_OS, BL_AL_OD, BL_AL_OS, 6m_AL_OD, 6m_AL_OS, 12m_AL_OD, 12m_AL_OS`.
Ethnicity: Vietnamese 128, Indian 61, Chinese 53. Follow-up: 219 children at 6 months, 71 at 12 months.

## Mapping decisions (all set in one place, top of `adapt_template.py`)

- **One eye per child.** Primary analysis = RIGHT eye (OD), matching the original template rule and preserving statistical independence. Left eye (OS) exported separately for a sensitivity run. Baseline SER, baseline AL, and every follow-up AL are taken from the same eye; eyes are never mixed within a child.
- **Sex.** Export codes sex 1/2. Mapped 2 = Male, 1 = Female, because in this cohort Sex=2 eyes are ~0.5 mm longer at matched age (the standard male pattern). This choice is immaterial to the result (the model's sex coefficient is ~0.004; flipping it changes 12-month bias from -0.004 to -0.005 mm), but should be confirmed with the trial team for the methods text.
- **Ethnicity** passed through verbatim; the engine routes Chinese/Vietnamese to East Asian (exact) and Indian to European (provisional).

## Exclusions (missing baseline, logged, NOT outcome-based)

Three Indian children were excluded from specific child-visits for missing baselines, recorded in `validation_exclusions.xlsx`:
- W-I-130004: missing baseline SER (cannot be predicted).
- W-I-130094, W-I-130103: missing baseline AL (cannot compute observed elongation).

No child-visit is ever removed on the basis of its outcome.

## How to reproduce

```
python adapt_template.py --eye OD --out untreated_validation_OD.xlsx   # primary
python adapt_template.py --eye OS --out untreated_validation_OS.xlsx   # sensitivity
python validate_model.py            # after pointing it at the converted Excel workbook
```

The validated engine (`virtual_control.py`) was not modified. Only the input adapter was added and the validation runner was made robust to missing baselines.
