## Supplementary software v0.2.1 for "Virtual control arms for paediatric myopia trials: external validation of axial-elongation models": ANALYSIS_PROTOCOL.md

# Recommended analysis: treated arm versus the virtual control

The tool supplies the **virtual untreated control** only (`out_predictions.xlsx`).
It does not compute a treatment effect. You estimate the effect yourself, from
your own treated outcomes, on your own machine. Your outcome data never has to
be shared with anyone. This document specifies how to do that correctly,
including the step most naive analyses get wrong: carrying the control's own
uncertainty into the effect estimate.

## 1. Principle

A virtual control is an **external** control, not a concurrent randomised one.
Two consequences follow, and both shape the statistics below:

- The control has estimation uncertainty of its own (the model and its
  calibration could be off). That uncertainty is **shared across all your
  treated children**, because the same model is applied to each. It therefore
  behaves like a systematic offset and does **not** shrink as you enrol more
  treated children.
- Enrolling more treated children reduces the sampling error of your treated
  mean, but it can never drive the effect's confidence interval to zero, because
  the control-side uncertainty remains. Any method whose interval keeps
  shrinking with treated n alone is anticonservative.

Treat this as **supportive, not pivotal**, evidence.

## 2. Inputs

From the tool, per child *i* and visit month *t* (`out_predictions.xlsx`):

- `predicted_delta_AL` = m(i,t), the point untreated prediction.
- `ci_low`, `ci_high` = the 95% band on the **mean** prediction. Its half-width
  gives the control's estimation standard error:
  se_ctrl(i,t) = (ci_high − ci_low) / (2 × 1.96). This is the only interval the
  tool ships, and it is the one the effect uses.

From you, per child and visit:

- observed treated elongation y(i,t) = treated axial length at visit t minus
  baseline axial length. Same eyes, same measurement convention, same visit
  timing basis as the control's derivation.

## 3. Per-child and arm-level effect

Per child, per visit, the Cumulative Absolute Reduction in Elongation:

    CARE(i,t) = m(i,t) − y(i,t)          (positive = benefit; treated grew less)

Arm mean at visit t, over the n(t) treated children measured at t:

    CARE_bar(t) = mean_i [ m(i,t) − y(i,t) ] = m_bar(t) − y_bar(t)

Report **CARE_mm as the primary metric**. Percent reduction
(CARE_bar / m_bar × 100) may be reported but is cautioned: the denominator
shrinks with age and follow-up, so percentages inflate at older ages and longer
horizons. Analyse each visit month separately, as the tool outputs per visit. If
a child contributes several visits and you want a single pooled estimate, use a
mixed-effects model with a child random effect rather than pooling rows naively.

## 4. Uncertainty (the step that matters)

The standard error of the arm effect at visit t has two independent parts:

    SE_within(t) = SD_i[ CARE(i,t) ] / sqrt(n(t))        treated sampling error
    SE_ctrl(t)   = mean_i[ se_ctrl(i,t) ]                 control estimation error

`SE_within` is the ordinary between-child sampling error; a paired bootstrap of
the per-child CARE values estimates the same quantity. `SE_ctrl` is the control's
own uncertainty. It is the **mean** of the per-child control standard errors, not
their standard error of the mean, precisely because the model error is common to
all children and does not average away over n.

Combine them in quadrature:

    SE_effect(t) = sqrt( SE_within(t)^2 + SE_ctrl(t)^2 )

Then:

    95% CI:  CARE_bar(t)  ±  1.96 × SE_effect(t)
    test:    z = CARE_bar(t) / SE_effect(t),   two-sided p = 2(1 − Phi(|z|))

This is deliberately conservative. A fully rigorous alternative propagates the
model's parameter covariance by Monte Carlo; the `ci` band is the shipped,
sound summary of that covariance and `SE_ctrl` is a defensible proxy. Note the
built-in behaviour: as treated n grows, `SE_within` falls but
`SE_ctrl` does not, so the interval converges to a positive floor set by the
control. That floor is correct, and it is exactly what a within-arm bootstrap
omits.

### Worked example (matches Figure 4)

Figure 4 in the paper is exactly this calculation on illustrative numbers, with
no patient data. At 12 months the virtual control mean is 0.344 mm and the
treated cohort mean is 0.180 mm, so CARE is 0.164 mm. The treated arm has
n = 60 and a per-child SD of 0.140 mm, and the control's confidence interval
gives a control-mean standard error of 0.018 mm.

    SE_within = 0.140 / sqrt(60)          = 0.0181 mm
    SE_ctrl   = 0.018 mm                   (from the tool's ci_low, ci_high)
    SE_effect = sqrt(0.0181^2 + 0.018^2)  = 0.0255 mm

The same effect then carries two different 95% intervals, depending on whether
the control's own uncertainty is included:

    Control treated as fixed (SE_ctrl dropped):  0.164 +/- 1.96 * 0.0181 = [0.129, 0.199]
    Control uncertainty included (recommended):  0.164 +/- 1.96 * 0.0255 = [0.114, 0.214]

The fixed version is narrower only because it discards `SE_ctrl`, which is
exactly the error a virtual control adds. Both intervals clear zero here, so the
effect holds under the wider, correct interval. The point estimate is robust to
model error as well: the modelled untreated mean would have to be over-estimated
by the full 0.164 mm, about 48 percent, to null the effect, whereas the
validated bias is under 0.02 mm.

## 5. What not to do

Do not report a within-arm paired bootstrap of (observed − predicted) as the
effect interval. It treats the virtual control as error-free, drops `SE_ctrl`
entirely, and is therefore anticonservative (too narrow, p too small). The
tool's internal `compare_to_test_arm` uses that bootstrap for internal
validation only and is not exposed by the shipped application; do not adopt it
as your inferential method.

## 6. Design cautions

- **One pre-specified comparator.** Fix the primary control model in advance
  (the validated primary model). Do not run the effect against several candidate
  models and report the most favourable; that inflates type-I error. Other
  models belong only in a pre-declared sensitivity analysis.
- **Applicability.** Validated for East Asian myopes at 6 months (equivalence to
  ±0.03 mm); consistent but not established at 12 months; South Asian routing is
  provisional. Do not extend the claim beyond the validated envelope.
- **Match the derivation.** Use the same biometer class, cycloplegia, ancestry
  coding, and timing basis as the control's derivation. If your population is not
  an exact match, calibrate to your own untreated data first with
  `fit_population_multiplier` (see the README), then re-export the control.
- **Residual confounding.** Secular trends, measurement differences, and
  population mismatch are not removed by a virtual control. State these
  assumptions explicitly alongside any effect estimate.

## 7. Reference implementation (you run this, the tool does not)

Use the tool's LONG output (the default) here, because this analysis merges on
`month`. Given the tool's `out_predictions.xlsx` and your filled
`observed_template.xlsx` (columns: `id, month, observed_elongation`), all held
locally:

```python
import numpy as np, pandas as pd
from scipy.stats import norm

Z = 1.959963984540054
pred = pd.read_excel("out_predictions.xlsx")
obs  = pd.read_excel("observed_template.xlsx")           # YOUR data, kept local

m = obs.merge(pred[["id", "month", "predicted_delta_AL", "ci_low", "ci_high"]],
              on=["id", "month"], how="inner")
m["care"]     = m["predicted_delta_AL"] - m["observed_elongation"]   # per child
m["se_ctrl"]  = (m["ci_high"] - m["ci_low"]) / (2 * Z)                 # per child

rows = []
for t, g in m.groupby("month"):
    n          = len(g)
    care_bar   = g["care"].mean()
    se_within  = g["care"].std(ddof=1) / np.sqrt(n)      # treated sampling error
    se_ctrl    = g["se_ctrl"].mean()                     # shared control error (no /sqrt n)
    se_effect  = np.hypot(se_within, se_ctrl)
    z          = care_bar / se_effect
    rows.append({"month": int(t), "n": n,
                 "CARE_mm": round(care_bar, 4),
                 "CI_low":  round(care_bar - Z * se_effect, 4),
                 "CI_high": round(care_bar + Z * se_effect, 4),
                 "SE_within": round(se_within, 4),
                 "SE_ctrl":   round(se_ctrl, 4),
                 "p_value":   round(2 * (1 - norm.cdf(abs(z))), 4)})
print(pd.DataFrame(rows).to_string(index=False))
```

Report `CARE_mm` with its combined-SE 95% CI as the primary effect, per visit,
with the applicability and design cautions of Section 6.
