## Supplementary software v0.2.1 for "Virtual control arms for paediatric myopia trials: external validation of axial-elongation models": CALIBRATION_NOTES.md

# Calibration and re-validation (timing-aware; supersedes the 2026-07-08 run)

Data: `Virtual_Control_data_2026-07-09.xlsx` (adds parental myopia and actual
visit days). Analysis: `analysis_v2.py`. Parameters: `calibration.json`.
All predictions are annualised on each child's ACTUAL elapsed follow-up time.

## Why the numbers changed from the first pass

The 6-month visits actually occurred at 6.6 months on average (202 days vs 183
nominal); the 12-month visits were on time (370 days). Predicting at nominal
months therefore under-predicted at 6 months for a purely mechanical reason.
Annualising on actual time removed about half of the apparent 6-month bias and
shrank the South Asian multiplier.

## Fitted parameters (actual time, all-data)

- Individual 95% prediction interval: half-width = 1.96 × 0.142 × √years (myopes).
  Error accumulates as a random walk (residual SD ≈ 0.142 mm per √year).
- South Asian (Indian) multiplier = 1.30 (95% CI 1.03–1.59), 6-month, N=46,
  provisional. Smaller than the naive nominal-time estimate of 1.51.
- Non-myopic interval unchanged (no non-myopes in the cohort).

## Result after calibration (5-fold cross-validated, actual time)

| Region (routing) | Visit | n | Bias (mm) | 95% PI coverage | Verdict |
|---|---|---|---|---|---|
| East Asian (Chinese, Vietnamese; exact) | 6 mo | 170 | −0.013 | 0.94 | PASS |
| East Asian | 12 mo | 71 | −0.004 | 0.93 | PASS |
| South Asian (Indian; provisional) | 6 mo | 46 | 0.000 | 0.87 | Provisional |

Thresholds: |bias| < 0.03 mm and coverage ≥ 0.90. CV and in-sample agree to
within 0.01 (no overfitting).

## Parental myopia: tested, not adopted

Head-to-head (2025 no-PM vs 2023 Chinese +PM) and a residual-on-parental-myopia
regression both show no useful improvement in these myopic children (residual
slope +0.010 mm/parent at 6 mo, 95% CI −0.011 to +0.030; not significant). Kept
out of the primary model; available as an option where parental myopia is
collected.

## Conclusion

Validated for East Asian myopes at 6 and 12 months; South Asian provisional.
Timing handling and the parental-myopia test both strengthen the record.
