## Supplementary software v0.2.1 for "Virtual control arms for paediatric myopia trials: external validation of axial-elongation models": INPUT_DICTIONARY.md

# Input dictionary

This tool takes baseline data only and returns the virtual untreated control
(`out_predictions.xlsx`). It does not need, and does not read, treated follow-up
outcomes. The same definitions, including the output columns in section 4, are
also embedded on the "Dictionary" sheet of `input_template.xlsx`, which is the
single Excel template; the tool reads and writes .xlsx only.

## 1. Tool input, baseline only (`input_template.xlsx`)

One row per child. Only these columns are read; any extra columns are ignored.

| Column | Meaning | Values or units | Required |
|--------|---------|-----------------|----------|
| `id` | Child identifier | any unique label | yes |
| `ethnicity` | Ancestry, NOT country of residence. Pick from the dropdown | see accepted values below | yes |
| `gender` | Sex at birth | `F` is Female, `M` is Male | yes |
| `age` | Age at baseline | years, decimal, for example 9.5 | yes |
| `baseline_SER` | Spherical equivalent refraction at baseline | dioptres, negative is myopic, for example -2.50 | yes |
| `baseline_AL` | Axial length at baseline | millimetres, for example 24.10 | yes |
| `stratum` | Model stratum for an ancestry with NO derived model (South Asian/Indian, or unmapped). Ignored when the ancestry already has a model | `east_asian` or `european`; blank otherwise | only for no-model ancestries |

`baseline_SER` sets the refractive group: a child is a myope at a `baseline_SER`
of -0.50 D or more myopic, otherwise a non-myope, which selects the correct model
equation.

`ethnicity` means ancestry, not country of residence: a Chinese child living in
Australia is East Asian, not European. It is required; pick from the dropdown so a
value cannot be misspelled into the wrong group. A row with a blank `ethnicity` is
not predicted and is listed in `out_needs_input.xlsx`.

### Accepted `ethnicity` values

Use exactly these values (the dropdown enforces them). Matching is whole-word and
case insensitive, so "Han Chinese" still resolves to Chinese, but an unrecognised
value is never force-fitted into a bucket: it is flagged and needs a `stratum`
choice.

| Status | Values (aliases accepted) | Model region used |
|--------|---------------------------|-------------------|
| Exact (in the 2025 derivation set) | Chinese (China, Han), Vietnamese (Vietnam), East Asian (Asian), Australian (Australia), European (UK, British, Swedish, White, Caucasian) | its own region |
| Approximate (same region, not sampled) | Taiwanese, Singaporean, Malaysian (Malay) | East Asian |
| Approximate (same region, not sampled) | Portuguese, Spanish, New Zealand (NZ, Kiwi) | European |
| No derived model: YOU choose the stratum | Indian, South Asian (India) | the `stratum` you set (`east_asian` or `european`) |
| Not recognised: no model applies | anything not listed above | none until you set a `stratum` or fix the value |

There is no Indian or South Asian axial-elongation model, and none for an
unrecognised ancestry. The tool does not guess a region for these children. In the
validation the paper applies every candidate model and reports how each stratum
fits; no stratum is correct by derivation. So set `stratum` to `east_asian` or
`european` for these children (guided by the paper and your own population), or
calibrate to your own untreated data with `fit_population_multiplier()`. A no-model
child with a blank `stratum` is not predicted; it appears in `out_needs_input.xlsx`.
When you set a stratum, that child's `model_status` is `user_selected` (provisional).

## 2. Your effect analysis, treated outcomes (`observed_template.xlsx`)

Not read by the tool. This is for your own effect analysis, after you have the
control. One row per child per visit: `id`, `month`, `observed_elongation`
(treated axial length at that visit minus baseline, in mm). Combine it with
`out_predictions.xlsx` following `ANALYSIS_PROTOCOL.md`. Keep it on your own
machine; nothing about this step is shared.

## 3. Model validation, untreated cohort (separate, your paper task)

Validating the models against a real untreated cohort is a separate workflow
(`validate_model.py`), on a different file: an untreated cohort with follow-up
axial length at 6 and 12 months in the same baseline columns plus `AL_m6` and
`AL_m12`. This is the paper's evidence step, not a tool-user step, and it is
documented in the README, not here.

## 3b. Choosing the model (the app dropdown)

You choose which published model to run, one per run (run again to compare). The
tool lays every model out and lets you decide; it does not pick one for you.

| Model (as shown) | `model_name` | Uses ancestry? | Covers | Ships a CI? |
|------------------|--------------|----------------|--------|-------------|
| Naduvilath 2025 (regional GEE) | `naduvilath2025` | yes (East Asian / European) | myopes and non-myopes | yes |
| Brennan 2024 (meta-regression) | `brennan2024` | yes (Asian / non-Asian) | myopes only | no |
| Jones 2005 (OLSM) | `olsm_jones2005` | no (age only) | myopes only | no |
| Wong 2010 (SCORM) | `scorm_wong2010` | no (age only) | myopes only | no |

Only the Naduvilath 2025 GEE ships a confidence interval of the mean, because it
is the only source that publishes the parameter uncertainty needed to derive one.
The other three return a point estimate, with a note saying so; their sources
publish no parameter covariance. This is neutral: a model gets an interval only if
its own publication provides one. See `docs/MODEL_NOTES.md` for the full rationale.

Age-only models (Jones 2005, Wong 2010) do not use ancestry at all: they predict
any child regardless of ethnicity, `region` is `n/a`, and no stratum is needed.
The ethnicity-dependent models still require a stratum for a no-model ancestry.
A non-myope sent to a myope-only model is not predicted; it is listed in
`out_needs_input.xlsx`.

In the app you may also fix the ethnicity for the whole run in a dropdown. If you
do, every row's `ethnicity` column must agree with your choice, or that row is
flagged in `out_needs_input.xlsx` rather than silently overridden (one ethnicity
per run). Leave the dropdown on "(from input file)" to use each row's own value.

## 4. Output columns (`out_predictions.xlsx`)

| Column | Meaning | Units |
|--------|---------|-------|
| `id` | Child identifier, echoed from the input | none |
| `population` | The `ethnicity` value you entered | none |
| `region` | Model region the child was routed to or you selected; `n/a` for age-only models | east_asian, european, or n/a |
| `model` | The model this run used, named as in the paper | e.g. Naduvilath 2025 (regional GEE) |
| `model_status` | How the region was decided | exact, approximate, user_selected, or age_only |
| `mapping_note` | Plain language explanation of the routing, and, for non-GEE models, why there is no confidence interval | none |
| `re_group` | Refractive group used, which sets the equation | myope or non-myope |
| `month` | Follow up horizon | months (6, 12, 18, 24, 36) |
| `predicted_delta_AL` | Predicted untreated change in axial length from baseline, the control value | mm |
| `ci_low`, `ci_high` | 95 percent confidence interval on the mean untreated prediction; blank for models that do not ship one | mm |
| `predicted_AL` | Predicted untreated axial length, which is `baseline_AL` plus `predicted_delta_AL` | mm |

The output can be written in long format (one row per child per month) or wide
format (one row per child, with each column suffixed by the month, for example
`predicted_delta_AL_m12`). Choose in the tool with the "Wide format" checkbox.

### The confidence interval of the mean

`ci_low` and `ci_high` is the 95 percent confidence interval on the mean
untreated change. It says how precisely the model pins down the average
trajectory for a child like this. This is the band a control arm uses, because a
control arm is a group mean comparator; use it for the arm level comparison and
the effect estimate (see `ANALYSIS_PROTOCOL.md`). The tool reports the mean and
this interval only. It does not report an individual prediction interval, because
the tool is a group level instrument and the individual interval was found to
under cover in validation.
