## Supplementary software v0.2.1 for "Virtual control arms for paediatric myopia trials: external validation of axial-elongation models": MODEL_NOTES.md

# How this tool chooses models and reports uncertainty

A neutral, evidence-based note, so you can see the tool takes no side. It
implements published models faithfully, tests them against each other without
favour, and reports each one's uncertainty only as far as its own publication
supports. The science of the models belongs to their original authors; this tool
adds no new growth model.

## The models it bundles

| Model | Conditions on | Applies to | Source |
|-------|---------------|-----------|--------|
| Naduvilath 2025 (regional GEE) | Region/ethnicity, baseline age, sex, refraction, region-by-age | Myopes and non-myopes | Ophthalmic Physiol Opt 2025;45:135-151 |
| Brennan 2024 (meta-regression) | Mean age, Asian vs non-Asian | Myopes | Optom Vis Sci 2024;101(8):497-507 |
| Jones 2005 (OLSM) | Age only | Myopes | Historical ultrasound-era cohort |
| Wong 2010 (SCORM) | Age only | Myopes | Historical ultrasound-era cohort |

## Part 1, the paper: we tested everything against everything

In the validation we applied every bundled model to the same untreated children
and reported each one's bias and error side by side, with no model designated the
primary in advance (see Table 2 for East Asian children and Table 3 for Indian
children). The findings, stated plainly: the Naduvilath 2025 GEE and the Brennan
2024 meta-regression sat closest to the observed untreated growth in East Asian
children; the two age-only ultrasound-era models (OLSM, SCORM) under-predicted
contemporary myopes, because they were fitted to older, slower-growing cohorts;
and Indian children fell between the model strata, matched by no existing model.

## Part 2, the tool: why only the GEE carries a confidence interval

In the tool all four models are offered in a dropdown, named exactly as above, and
you choose which to run (one per run; run again to compare). The tool lays them out
and lets you decide; it does not pick one for you. What differs between them is not
which one we prefer, but which one can carry an interval.

This is the point most likely to look like a preference, so here is the reason in
full. A control arm needs not just a predicted value but a confidence interval on
that value, because the effect analysis must carry the control's own uncertainty
(see ANALYSIS_PROTOCOL.md). Whether a model can supply that interval is decided by
what its own publication reports, not by us:

- Naduvilath 2025 (regional GEE): publishes the parameter uncertainty needed to
  form a confidence interval on the mean prediction. So the tool returns the point
  control and its confidence interval.
- Brennan 2024 (meta-regression): publishes confidence bounds for each coefficient
  but not the covariance between them, and its age term is uncentered, so those
  bounds cannot be combined into a correct mean interval; the only intervals it
  tabulates are wide prediction intervals (roughly minus 40 to plus 60 percent),
  which describe individual and between-study spread, not the precision of the
  mean. We checked the paper and its supplemental appendix directly to confirm
  this. So the tool returns Brennan as a point estimate only.
- Jones 2005 (OLSM) and Wong 2010 (SCORM): age-only equations with no published
  uncertainty of any kind. So the tool returns them as point estimates only.

In short, the tool does not manufacture a confidence interval a source never
published. The Naduvilath 2025 GEE is not a favourite; it is simply the only
bundled model that ships the uncertainty a control arm requires. A point-only
model can still be used for a rough comparison, but its result should not be
carried into a formal effect interval without a separately justified uncertainty.

## What our own data can and cannot support

Our external validation covers East Asian children at 6 and 12 months and Indian
children at 6 months. Predictions outside that envelope, longer horizons or
ancestries we did not validate, are extrapolations and are flagged as such. There
is no Indian or South Asian axial-elongation model in any source, so for those
children the tool does not guess a region; you choose the stratum, and the result
is marked provisional.

## The stance, in one line

Faithful implementation, neutral head-to-head validation, and uncertainty reported
exactly as far as each publication supports, no further and no less.
