## Supplementary software v0.2.1 for "Virtual control arms for paediatric myopia trials: external validation of axial-elongation models": PUBLICATION_READINESS.md

# Publication readiness: making this truthful, objective, and citable

Decision taken: **hold the Indian routing as provisional and report the primary
validation on the untuned published model.** No parameter is tuned to force a
pass. This section is the plan that keeps the paper defensible in peer review and
usable as a citation for internal, external, and regulatory purposes.

## The analysis has two clearly separated layers

**Primary (pre-registered, no tuning).** Externally validate the published
Naduvilath et al. 2025 model on our untreated cohort, exactly as published. This
is the objective core and the default behaviour of the released tool.

- East Asian myopes: mean prediction unbiased (bias −0.025 mm at 6 m, −0.004 mm
  at 12 m).
- Published individual 95% intervals were miscalibrated (coverage 0.75 and 0.87
  vs the 0.90 floor). Reported as a finding, not hidden.
- Indian routing under-predicted (bias −0.053 mm). Reported as a finding.

**Secondary (pre-specified calibration, internally cross-validated).** A
recalibration derived FROM these data and evaluated by 5-fold cross-validation:
sqrt-time interval widening (all myopes) and a provisional Indian multiplier.
Presented as calibration, never as independent validation.

- After calibration, East Asian coverage is nominal (0.95, 0.93): a validated
  improvement in the derivation region.
- Indian mean is corrected but coverage stays 0.87 on N = 46, 6-month, single
  cohort: reported as provisional, requiring an independent Indian cohort.

Writing it this way means every number is true, the tuning is transparent, and no
reader can mistake in-sample calibration for external validation.

## Honest claim wording (safe to publish)

> "In an external cohort of untreated myopic children, the published regional
> model predicted mean axial elongation without significant bias in East Asian
> children at 6 and 12 months. Published individual prediction intervals were too
> narrow; a pre-specified, cross-validated recalibration restored nominal coverage
> in East Asian children. The provisional South Asian (Indian) routing was
> bias-corrected on these data but remained under-covered and is not claimed as
> validated pending an independent cohort."

Do NOT write "the tool is validated for all populations," "Indian routing is
validated," or quote in-sample coverage as if it were out-of-sample.

## Reporting standard: TRIPOD+AI

Structure the manuscript against the TRIPOD checklist for prediction-model
validation studies. The items most likely to draw reviewer fire, and where we
stand:

- Source of data and participants: state provenance and, critically, that the
  untreated validation children are INDEPENDENT of any trial the tool will later
  serve as a control for. Confirm this; overlap is the one thing that invalidates
  the whole use case.
- Outcome definition: axial elongation = follow-up AL − baseline AL, one eye
  (right) per child, optical biometry.
- Predictors and model: published coefficients, cited, unchanged.
- Sample size and missing data: 242 children; exclusions logged in
  validation_exclusions.xlsx (missing baseline only, never outcome-based).
- Model performance: bias, RMSE, limits of agreement, calibration slope/
  intercept, and interval coverage, by region and visit.
- Model updating: the calibration, labelled as such, with cross-validated metrics.

## Pre-registration integrity (do this before submission)

The acceptance thresholds (|bias| < 0.03 mm, coverage ≥ 0.90) live in
validate_model.py. State in the paper whether they were fixed before the data
were seen. If they were, say so and cite the timestamp. If timing is uncertain,
present them as descriptive and let the reader judge; do not backfill a
pre-registration claim. Consider depositing the analysis plan (OSF) now for the
next, independent cohort.

## Citability (so internal, external, and regulatory readers can cite it)

- Archive the exact code version with a DOI (Zenodo, linked to the repository at
  the tagged release). The DOI is what regulators and other papers cite.
- Add CITATION.cff (template provided) so the software is citable independently of
  the paper.
- Licence: GNU General Public License v3.0 (copyleft), in `LICENSE`. No
  freedom-to-operate question arises: the tool is free, non-commercial, and
  re-implements published models, so there is nothing to clear. The one IP
  consequence is deliberate and favourable: publishing the method is defensive
  prior art that prevents a competitor patenting it. nthalmic retains copyright,
  so GPL binds third parties only and does not restrict reuse of the code in our
  own commercial products.
- Data availability: publish the aggregate validation metrics and code; share
  individual-level untreated data under governance if ethics permits, else state
  the restriction.
- Reproducibility: recalibrate.py is deterministic (fixed seed) and the published
  self-validation runs on import; note both.

## Regulatory positioning (unchanged, restated)

A peer-reviewed, DOI-archived, cross-validated tool is strong supporting evidence
and state-of-the-art context. It is not, by itself, regulatory qualification.
Engage TGA or FDA in a pre-submission before relying on the virtual control as a
comparator, and confirm the independence point above.
