## Supplementary software v0.2.1 for "Virtual control arms for paediatric myopia trials: external validation of axial-elongation models": REVIEW_RESPONSE.md

# Response to three expert reviews (statistics, myopia, scientific computing)

Disposition key: [FIXED] changed in the manuscript/code; [AUTHORS] needs a
decision or data only you can provide; [RELEASE] code-release task before
public/JOSS submission.

---

## Open items (last updated 2026-07-13)

Status: manuscript revised after three expert reviews; statistics and code fixes
done; numbers reproduce via `python reproduce.py`; figures regenerated at 600 dpi
plus vector PDF plus an editable Excel workbook; in-text Vancouver citations and
two references (Chamberlain 2024, Gopalakrishnan STEM 2024) added; a TRIPOD+AI
sentence added. Blocked only on the items below.

Blocking submission (only you can close):
- [ ] 1. Confirm the cohort's site/country and NON-OVERLAP with the Naduvilath 2025 (China, Vietnam) and 2023 (China) derivation sets. If overlap, the 12-month East Asian result becomes internal/temporal validation, not external. HIGHEST PRIORITY.
- [ ] 2. Supply cohort methods: biometer make/model, cycloplegia agent/regimen, definition of "untreated", inclusion/exclusion criteria, ethics approval + consent, how ancestry was ascertained.
- [ ] 3. Supply participant flow + attrition: why 219 have a 6-month but only 71 a 12-month visit; completer vs non-completer baseline comparison.
- [ ] 4. Author list (incl. the data contributor) + ORCIDs; corresponding-author details.

Ready when you are (can be done on your word):
- [ ] 5. Draft the participant-flow (CONSORT-style) figure once #3 arrives.
- [ ] 6. [RELEASE] Unify the shipped engine onto one daily integrator + registry so the tool reproduces the tables exactly (currently within 0.002 mm).
- [ ] 7. [RELEASE] Convert self-checks into an asserting pytest suite; add a machine-readable coefficient table citing each source table/page.
- [ ] 8. Complete the TRIPOD+AI checklist and cover letter for the target journal (the manuscript already states TRIPOD+AI adherence).
- [ ] 9. Zenodo archive + DOI wiring in CITATION.cff.

Done this round (was open):
- [x] In-text numbered citations, plus references 12 (Chamberlain 2024) and 13 (STEM 2024).
- [x] `reproduce.py` fixed end to end (`make_figs_v2.py` restored); figures at 600 dpi + vector PDF; editable Excel figure workbook added.
- [x] Nominal/actual calibration timing separated and never mixed; actual time is primary.
- [x] Licence settled: GNU GPL v3.0 (see below); no IP/FTO gate applies.

Target venue (agreed framing): a strong specialist journal (OPO / TVST) or a
Nature-family methods/software outlet; honesty and reproducibility are the pitch.

---

## Statistics review

- [FIXED] Coverage reported as point estimates without CIs → Wilson 95% CIs added to every coverage figure (Tables 2, 3); acceptance rule now applied to confidence bounds.
- [FIXED] Bias treated as a pass without an equivalence test → TOST vs ±0.03 mm added. 6-month East Asian equivalence established (p=0.014); 12-month not established (p=0.068) and downgraded to "consistent, not established" throughout (abstract, results, conclusions).
- [FIXED] South Asian "fail" overstated → its coverage CI (0.77–0.95) includes 0.90, now reported as under-covered but indeterminate.
- [FIXED] Repeated 6/12-month visits pooled as independent → sigma now reported with a child-level cluster bootstrap CI (0.127–0.155); per-cell independence and children counts stated; methods note the repeated-measures structure.
- [FIXED] Parental-myopia null presented as evidence of absence → reframed as an underpowered non-detection (power ~0.31/0.10; equivalence fails).
- [FIXED] Calibration slopes read as "imperfect discrimination" → all slopes shown with CIs, all consistent with 1.0; the discrimination claim removed.
- [FIXED] √time law from two points overstated as a random walk → now an empirical two-point fit consistent with √time (exponents ~0.4–0.6 indistinguishable); extrapolation flagged.
- [FIXED] Multiplicity unmentioned and "best two of four" post-hoc → primary model stated as pre-specified; thresholds called descriptive; model spread reported as an uncertainty component.
- [FIXED] Estimand mixed a trial ATE with a calibration study → split into a validation estimand (this study) and a downstream trial estimand.
- [FIXED] "Agreed within 0.01 on every metric" false for coverage → corrected (bias within 0.01; coverage differs up to ~0.03).
- [FIXED] sigma inconsistency (0.142/0.1417/0.148) → standardised to 0.142 in text.

## Myopia review

- [AUTHORS] Independence of the Vietnamese cohort from the Naduvilath 2025 East Asian derivation (which includes Vietnam) — the single most important item. Manuscript now states this must be confirmed, and that the 12-month result is external only if non-overlap holds. You must confirm the site and non-overlap in writing.
- [FIXED] "European routing" for Indian children reframed as a provisional intermediate proxy; South Asian calibration slope (0.58) reported; static AL vs elongation-rate distinction added; region labels defined as model-region membership, not ethnicity.
- [FIXED] Ethics-of-untreated-arms overclaim softened (single-vision arms remain acceptable; the real issue is recruitment/retention).
- [FIXED] Doubly-robust/TMLE sentence corrected (does not apply to a purely model-based control).
- [FIXED] Coverage label 95% vs acceptance 0.90 clarified; 0.03 mm margin put in biometry/effect context.
- [FIXED] Benchmark paragraph added comparing observed untreated rates to published placebo/single-vision arms.
- [FIXED] "Validated on 242" corrected to analytic n's; 12-month claim marked Vietnamese-only.
- [FIXED] Citations added (IMI clinical trials report; ATOM2, LAMP, DIMS, HAL/Stellest control arms; Bullimore–Brennan; Van Calster calibration; TRIPOD+AI).
- [AUTHORS] Cohort methods to complete: biometer make/model, cycloplegia regimen, definition of untreated, inclusion/exclusion, ethics/consent, ancestry ascertainment, participant-flow diagram and completer-vs-non-completer comparison. A placeholder paragraph marks exactly what is required.

## Scientific-computing review

- [FIXED] `pm_experiment.py` now runs standalone (the OD workbook carries parental myopia).
- [FIXED] Stale `multimodel_validation.xlsx` (nominal time) regenerated at actual time to match Table 4.
- [FIXED] One-command `reproduce.py`, pinned `requirements.txt`, `README.md`, and a `LICENSE` added; the availability statement now matches reality.
- [FIXED] "Within 0.01 on every metric" corrected (see statistics).
- [RELEASE] Unify the validation onto the shipped engine (drive `predict_untreated` + registry; single daily integrator) so the released tool reproduces the tables exactly rather than to within 0.002 mm. Numbers are mean-preserving; this is an integrity/tidiness fix.
- [RELEASE] Convert the `validate*()` print-checks into an asserting `pytest` suite; add a machine-readable coefficient table citing each source table/page.
- [RELEASE] Remove dead `analysis_v2.load()`; document the two data-file provenance; fix the shipped-API sex-coding trap (1/2 vs M/F) and the registry substring fallback.

## The two things only you can close

1. Confirm the cohort's site and its non-overlap with the Naduvilath 2025 (China, Vietnam) and 2023 (China) derivation sets. If overlap exists, the 12-month East Asian result becomes internal/temporal validation.
2. Supply the cohort methods details listed above (biometer, cycloplegia, ethics, inclusion/exclusion, attrition reasons).

Licence settled: GNU GPL v3.0. No IP/FTO gate applies (free, non-commercial,
published-model re-implementation; copyright retained, so reuse in nthalmic's
commercial products stays open).
