## Supplementary software v0.2.1 for "Virtual control arms for paediatric myopia trials: external validation of axial-elongation models": SOURCES.md

# Sources, coverage, and what to download next

## Models wired into virtual_control.py

- **Primary (default):** Naduvilath et al. 2025, regional East Asian / European GEE
  model, exact Table 3 coefficients. Covers Chinese, Vietnamese (East Asian) and
  UK/Sweden/Australia (European) as derivation populations.
- **Sensitivity (sensitivity_models.py):** Brennan 2024, OLSM (Jones 2005), and
  SCORM (Wong 2010).

## Effect reporting (new): CARE

`compare_to_test_arm()` now reports the effect three ways, following Brennan et
al. 2021:
- `delta_effect` = observed - predicted (negative = slowed).
- `CARE_mm` = predicted - observed = **Cumulative Absolute Reduction in
  Elongation** (positive = benefit). This absolute value is the recommended
  primary metric.
- `percent_reduction` = CARE / predicted. Reported but **cautioned**: the
  denominator shrinks with age, so percentages inflate at older ages and longer
  follow-up. Report CARE_mm, not the percentage, as your headline.

**Internal use only.** `compare_to_test_arm()` is for internal validation and is
not exposed by the shipped tool. Its paired within-arm bootstrap treats the
virtual control as error-free: it omits the control's own mean uncertainty
(`SE_ctrl`) and is therefore anticonservative (intervals too narrow, p too
small). Do not adopt it as your inferential method. For the effect interval use
the closed form in `ANALYSIS_PROTOCOL.md` sections 4 and 7, which combines the
treated sampling error and the control's confidence interval in quadrature.

## Multi-model set (primary + sensitivity), all wired in

`sensitivity_models.py` now runs the effect under four published models and
reports concordance. Each is validated against its own source numbers:
- naduvilath2025 (PRIMARY): regional East Asian/European, myope + non-myope.
- brennan2024: meta-regression of 78 studies, Asian vs non-Asian, age only,
  myopes. ln(rate) = 0.362 - 0.158*age + 0.325*race.
- olsm_jones2005: OLSM White US myopes, published axial-length model (Table 4,
  Axial length, Myopes): AL = 18.144 + 2.391*ln(age) (<=10.5y) else
  17.808 + 2.560*ln(age), so rate = dAL/dage = 2.391/age (<=10.5y) else 2.560/age.
- scorm_wong2010: SCORM Singaporean myopes, published axial-length model
  (Table 2, persistent myopia): AL = 20.61 + 0.163*age^2 - 0.053*age^2*ln(age),
  so rate = dAL/dage = 0.273*age - 0.106*age*ln(age).
OLSM and SCORM are ultrasound-era, but each publishes an axial-length growth
model, so the tool uses the published AL curve directly (differentiated to an
annual elongation rate). They and Brennan apply to myopes only.

## Papers already in the folder, and their role

- Naduvilath 2025 (Regional/ethnic, opo.13401) — primary model, wired in.
- Truckenbrod 2021 (German LIFE cohort) — European **percentile curves**, not a
  progression equation. Validates our European numbers (its 50th-centile ~0.115
  mm/yr for 6-16y matches the 2025 European estimates). Not a plug-in model.
- Bullimore, Cheng & Brennan 2025 (centile-curve limitations) — argues centile
  charts underestimate myopic growth and recommends **annualised growth models**
  (exactly our approach). Supports the design; no coefficients.
- McCullough (NICER) 2020 (white UK children) — this cohort is ALREADY inside the
  2025 European region (that model pooled NICER + Sweden + Australia). Confirms
  our European numbers (its myope elongation ~0.30 mm/yr at 6-16y matches). Not a
  separate plug-in.
- Brennan et al. 2021 "Efficacy in myopia control" — defines the CARE metric now
  reported by compare_to_test_arm; warns against percentage-reduction framing.
- Chamberlain et al. 2023 (six-year dual-focus) — worked example of benchmarking
  a treated arm against predicted untreated growth over 6 years.
- Saw 2005 (SCORM), Zadnik 1993 (OLSM), Chua 2016 — background for the
  Chamberlain virtual-cohort precedent; not used for coefficients.

## India (STEM) now embedded

Gopalakrishnan et al. 2024 (STEM, Sankara Nethralaya) is a cross-sectional AL
percentile paper, not a myope progression regression, so it cannot make Indian
"exact" for the progression model on its own. But it delivers two things:
- Its AL percentile table is embedded as `INDIA_STEM_AL_CENTILES` /
  `india_al_centiles()` for centile-based **screening** of Indian children.
- It empirically confirms the Indian -> European routing: STEM median AL growth
  is ~0.136 mm/yr (boys, 6-11y), essentially identical to the European non-myope
  modelled rate (~0.132 mm/yr), and Indian AL is far shorter than Chinese. So
  European is the correct, mildly conservative proxy. The "indian" mapping note
  now records this.

To make Indian myope-exact you still need a STEM-derived progression model or
your own untreated Indian cohort via fit_population_multiplier().

## Coverage by your target populations

| Population | Status now | Route |
|-----------|------------|-------|
| Chinese, Vietnamese | exact | East Asian (in 2025 derivation) |
| Australian | exact | European (in 2025 derivation) |
| Taiwanese, Singaporean, Malaysian | approximate | East Asian region |
| Portuguese, Spanish, New Zealand | approximate | European region |
| Indian | provisional | defaults to European; NOT represented |

## Highest-value downloads to tighten it further

1. **India (fills the real gap).** Gopalakrishnan et al. 2024, "Ocular Biometry
   Percentile Curves and Their Relation to Myopia Development in Indian Children"
   (J Clin Med) — the STEM study (Sankara Nethralaya Tamil Nadu Essilor Myopia
   Study), 4514 Indian children 6-12, longitudinal. This is the India-specific
   dataset. It publishes centile curves rather than a progression regression, so
   to make "indian" exact you either (a) request/derive an annualised progression
   model from the STEM longitudinal data, or (b) fit our
   fit_population_multiplier() to your own Indian untreated cohort.

2. **Efficacy framing (how to report the effect).** Brennan, Toubouti, Cheng &
   Bullimore 2021, "Efficacy in myopia control" (Prog Retin Eye Res) — defines
   the CARE metric (cumulative absolute reduction in axial elongation) and warns
   against percentage-reduction framing, regression to the mean, and rebound.
   This should shape how compare_to_test_arm results are reported.

3. **Real-world virtual-control exemplar.** Chamberlain et al. 2023, "Six-year
   cumulative treatment effect ... dual focus" (Ophthalmic Physiol Opt) — a
   worked example of benchmarking a treated arm against predicted untreated
   growth over 6 years.

4. **Chinese percentiles (cross-check).** He et al. 2023, "Normative data and
   percentile curves for AL and AL/CR in Chinese children 4-18y" (Br J
   Ophthalmol) — for AL/CR-based routing and a percentile cross-check.

5. **European per-country (optional refinement).** McCullough (NICER) 2020, "Axial
   growth and refractive change in white European children" (Sci Rep); Tideman
   et al. 2018 (Acta Ophthalmol). Only needed if you want a Europe model distinct
   from the pooled 2025 European region.

## Practical next step for the Indian gap

Once you have an untreated Indian cohort (even ~50-100 children with baseline SER,
age, and >=1 year AL follow-up):

```python
from virtual_control import fit_population_multiplier, register_population
mult = fit_population_multiplier(indian_df, region="european")  # or "east_asian"
register_population("indian", region="european", status="exact",
                    note="Calibrated to STEM/our cohort 2026", multiplier=mult)
```

That moves Indian from provisional to a data-backed model in one line.
