## Supplementary software v0.2.1 for "Virtual control arms for paediatric myopia trials: external validation of axial-elongation models": index.html

Virtual Control Arm Tool


### Virtual Control Arm Tool

Builds a predicted untreated control arm for single-arm myopia-control studies, from published axial-elongation models. Runs entirely in your browser.

Version 0.2.1 · released 2026-08-09

Your file is read and processed inside this browser tab only. It is never uploaded to any server. The output is generated locally and offered back to you as a download.

#### 1. Choose the model and options

Model

Naduvilath 2025 (regional GEE) — ships a confidence interval
Brennan 2024 (meta-regression) — point estimate
Jones 2005 (OLSM) — point estimate
Wong 2010 (SCORM) — point estimate

One model per run. Only the GEE ships a confidence interval of the mean; the others return a point estimate because their sources publish no parameter covariance.

Ethnicity override (optional)

(use the ethnicity column in the file)
Chinese (East Asian)
Vietnamese (East Asian)
East Asian
European
Australian (European)

Ethnicity is the child's ancestry, not country of residence. If set, it applies to every row. Leave as default to use each row's own value. Ancestries with no derived model need a `stratum` value (`east_asian` or `european`) in the file.

#### 2. Upload your baseline data

Baseline Excel file (.xlsx)


Use the supplied Excel template (`examples/sample_input.xlsx`). Columns: `id, ethnicity, gender, age, baseline_SER, baseline_AL, stratum`. Only baseline columns are used; any treated follow-up columns are ignored. The result is an Excel workbook (`out_predictions.xlsx`). The tool builds the control only; you compute the treatment effect yourself per the analysis protocol, so your treated outcomes never leave your machine.

Run
Run self-test

#### Status

Loading the Python engine (about 21 MB, served from this page itself, no network used)…

Engine: `src/virtual_control.py`, run unmodified. This is the exact source deposited with the paper, so what you read is what executes.
