## Supplementary software v0.2.1 for "Virtual control arms for paediatric myopia trials: external validation of axial-elongation models": pyodide.asm.js

```
"use strict";
var _createPyodideModule = (() => {
  var _scriptName = typeof document != 'undefined' ? document.currentScript?.src : undefined;
  return (
async function(moduleArg = {}) {
  var moduleRtn;

var Module=moduleArg;var readyPromiseResolve,readyPromiseReject;var readyPromise=new Promise((resolve,reject)=>{readyPromiseResolve=resolve;readyPromiseReject=reject});var ENVIRONMENT_IS_WEB=typeof window=="object";var ENVIRONMENT_IS_WORKER=typeof WorkerGlobalScope!="undefined";var ENVIRONMENT_IS_NODE=typeof process=="object"&&process.versions?.node&&process.type!="renderer";var ENVIRONMENT_IS_SHELL=!ENVIRONMENT_IS_WEB&&!ENVIRONMENT_IS_NODE&&!ENVIRONMENT_IS_WORKER;if(ENVIRONMENT_IS_NODE){}var arguments_=[];var thisProgram="./this.program";var quit_=(status,toThrow)=>{throw toThrow};if(typeof __filename!="undefined"){_scriptName=__filename}else if(ENVIRONMENT_IS_WORKER){_scriptName=self.location.href}var scriptDirectory="";function locateFile(path){if(Module["locateFile"]){return Module["locateFile"](path,scriptDirectory)}return scriptDirectory+path}var readAsync,readBinary;if(ENVIRONMENT_IS_NODE){var fs=require("fs");var nodePath=require("path");scriptDirectory=__dirname+"/";readBinary=filename=>{filename=isFileURI(filename)?new URL(filename):filename;var ret=fs.readFileSync(filename);return ret};readAsync=async(filename,binary=true)=>{filename=isFileURI(filename)?new URL(filename):filename;var ret=fs.readFileSync(filename,binary?undefined:"utf8");return ret};if(process.argv.length>1){thisProgram=process.argv[1].replace(/\\/g,"/")}arguments_=process.argv.slice(2);quit_=(status,toThrow)=>{process.exitCode=status;throw toThrow}}else if(ENVIRONMENT_IS_SHELL){readBinary=f=>{if(typeof readbuffer=="function"){return new Uint8Array(readbuffer(f))}let data=read(f,"binary");assert(typeof data=="object");return data};readAsync=async f=>readBinary(f);globalThis.clearTimeout??=id=>{};globalThis.setTimeout??=f=>f();arguments_=globalThis.arguments||globalThis.scriptArgs;if(typeof quit=="function"){quit_=(status,toThrow)=>{setTimeout(()=>{if(!(toThrow instanceof ExitStatus)){let toLog=toThrow;if(toThrow&&typeof toThrow=="object"&&toThrow.stack){toLog=[toThrow,toThrow.stack]}err(`exiting due to exception: ${toLog}`)}quit(status)});throw toThrow}}if(typeof print!="undefined"){globalThis.console??={};console.log=print;console.warn=console.error=globalThis.printErr??print}}else if(ENVIRONMENT_IS_WEB||ENVIRONMENT_IS_WORKER){try{scriptDirectory=new URL(".",_scriptName).href}catch{}{if(ENVIRONMENT_IS_WORKER){readBinary=url=>{var xhr=new XMLHttpRequest;xhr.open("GET",url,false);xhr.responseType="arraybuffer";xhr.send(null);return new Uint8Array(xhr.response)}}readAsync=async url=>{if(isFileURI(url)){return new Promise((resolve,reject)=>{var xhr=new XMLHttpRequest;xhr.open("GET",url,true);xhr.responseType="arraybuffer";xhr.onload=()=>{if(xhr.status==200||xhr.status==0&&xhr.response){resolve(xhr.response);return}reject(xhr.status)};xhr.onerror=reject;xhr.send(null)})}var response=await fetch(url,{credentials:"same-origin"});if(response.ok){return response.arrayBuffer()}throw new Error(response.status+" : "+response.url)}}}else{}var out=console.log.bind(console);var err=console.error.bind(console);var dynamicLibraries=[];var wasmBinary;var wasmMemory;var ABORT=false;var EXITSTATUS;function assert(condition,text){if(!condition){abort(text)}}var HEAP,HEAP8,HEAPU8,HEAP16,HEAPU16,HEAP32,HEAPU32,HEAPF32,HEAP64,HEAPU64,HEAPF64;var runtimeInitialized=false;var runtimeExited=false;var isFileURI=filename=>filename.startsWith("file://");function updateMemoryViews(){var b=wasmMemory.buffer;Module["HEAP8"]=HEAP8=new Int8Array(b);Module["HEAP16"]=HEAP16=new Int16Array(b);Module["HEAPU8"]=HEAPU8=new Uint8Array(b);Module["HEAPU16"]=HEAPU16=new Uint16Array(b);Module["HEAP32"]=HEAP32=new Int32Array(b);Module["HEAPU32"]=HEAPU32=new Uint32Array(b);Module["HEAPF32"]=HEAPF32=new Float32Array(b);Module["HEAPF64"]=HEAPF64=new Float64Array(b);Module["HEAP64"]=HEAP64=new BigInt64Array(b);Module["HEAPU64"]=HEAPU64=new BigUint64Array(b)}function initMemory(){if(Module["wasmMemory"]){wasmMemory=Module["wasmMemory"]}else{var INITIAL_MEMORY=Module["INITIAL_MEMORY"]||20971520;wasmMemory=new WebAssembly.Memory({initial:INITIAL_MEMORY/65536,maximum:65536})}updateMemoryViews()}var __RELOC_FUNCS__=[];function preRun(){if(Module["preRun"]){if(typeof Module["preRun"]=="function")Module["preRun"]=[Module["preRun"]];while(Module["preRun"].length){addOnPreRun(Module["preRun"].shift())}}callRuntimeCallbacks(onPreRuns)}function initRuntime(){runtimeInitialized=true;callRuntimeCallbacks(__RELOC_FUNCS__);callRuntimeCallbacks(onInits);if(!Module["noFSInit"]&&!FS.initialized)FS.init();TTY.init();SOCKFS.root=FS.mount(SOCKFS,{},null);PIPEFS.root=FS.mount(PIPEFS,{},null);wasmExports["__wasm_call_ctors"]();callRuntimeCallbacks(onPostCtors);FS.ignorePermissions=false}function preMain(){callRuntimeCallbacks(onMains)}function exitRuntime(){___funcs_on_exit();callRuntimeCallbacks(onExits);FS.quit();TTY.shutdown();IDBFS.quit();runtimeExited=true}function postRun(){if(Module["postRun"]){if(typeof Module["postRun"]=="function")Module["postRun"]=[Module["postRun"]];while(Module["postRun"].length){addOnPostRun(Module["postRun"].shift())}}callRuntimeCallbacks(onPostRuns)}var runDependencies=0;var dependenciesFulfilled=null;function getUniqueRunDependency(id){return id}function addRunDependency(id){runDependencies++;Module["monitorRunDependencies"]?.(runDependencies)}function removeRunDependency(id){runDependencies--;Module["monitorRunDependencies"]?.(runDependencies);if(runDependencies==0){if(dependenciesFulfilled){var callback=dependenciesFulfilled;dependenciesFulfilled=null;callback()}}}function abort(what){Module["onAbort"]?.(what);what="Aborted("+what+")";err(what);ABORT=true;what+=". Build with -sASSERTIONS for more info.";if(runtimeInitialized){___trap()}var e=new WebAssembly.RuntimeError(what);readyPromiseReject(e);throw e}var wasmBinaryFile;function findWasmBinary(){return locateFile("pyodide.asm.wasm")}function getBinarySync(file){if(file==wasmBinaryFile&&wasmBinary){return new Uint8Array(wasmBinary)}if(readBinary){return readBinary(file)}throw"both async and sync fetching of the wasm failed"}async function getWasmBinary(binaryFile){if(!wasmBinary){try{var response=await readAsync(binaryFile);return new Uint8Array(response)}catch{}}return getBinarySync(binaryFile)}async function instantiateArrayBuffer(binaryFile,imports){try{var binary=await getWasmBinary(binaryFile);var instance=await WebAssembly.instantiate(binary,imports);return instance}catch(reason){err(`failed to asynchronously prepare wasm: ${reason}`);abort(reason)}}async function instantiateAsync(binary,binaryFile,imports){if(!binary&&typeof WebAssembly.instantiateStreaming=="function"&&!isFileURI(binaryFile)&&!ENVIRONMENT_IS_NODE&&!ENVIRONMENT_IS_SHELL){try{var response=fetch(binaryFile,{credentials:"same-origin"});var instantiationResult=await WebAssembly.instantiateStreaming(response,imports);return instantiationResult}catch(reason){err(`wasm streaming compile failed: ${reason}`);err("falling back to ArrayBuffer instantiation")}}return instantiateArrayBuffer(binaryFile,imports)}function getWasmImports(){return{env:wasmImports,wasi_snapshot_preview1:wasmImports,"GOT.mem":new Proxy(wasmImports,GOTHandler),"GOT.func":new Proxy(wasmImports,GOTHandler)}}async function createWasm(){function receiveInstance(instance,module){wasmExports=instance.exports;wasmExports=relocateExports(wasmExports,1024);var metadata=getDylinkMetadata(module);if(metadata.neededDynlibs){dynamicLibraries=metadata.neededDynlibs.concat(dynamicLibraries)}mergeLibSymbols(wasmExports,"main");LDSO.init();loadDylibs();wasmExports=applySignatureConversions(wasmExports);__RELOC_FUNCS__.push(wasmExports["__wasm_apply_data_relocs"]);removeRunDependency("wasm-instantiate");return wasmExports}addRunDependency("wasm-instantiate");function receiveInstantiationResult(result){return receiveInstance(result["instance"],result["module"])}var info=getWasmImports();if(Module["instantiateWasm"]){return new Promise((resolve,reject)=>{Module["instantiateWasm"](info,(mod,inst)=>{resolve(receiveInstance(mod,inst))})})}wasmBinaryFile??=findWasmBinary();try{var result=await instantiateAsync(wasmBinary,wasmBinaryFile,info);var exports=receiveInstantiationResult(result);return exports}catch(e){readyPromiseReject(e);return Promise.reject(e)}}class ExitStatus{name="ExitStatus";constructor(status){this.message=`Program terminated with exit(${status})`;this.status=status}}var GOT={};var currentModuleWeakSymbols=new Set([]);var GOTHandler={get(obj,symName){var rtn=GOT[symName];if(!rtn){rtn=GOT[symName]=new WebAssembly.Global({value:"i32",mutable:true})}if(!currentModuleWeakSymbols.has(symName)){rtn.required=true}return rtn}};var callRuntimeCallbacks=callbacks=>{while(callbacks.length>0){callbacks.shift()(Module)}};var onPostRuns=[];var addOnPostRun=cb=>onPostRuns.push(cb);var onPreRuns=[];var addOnPreRun=cb=>onPreRuns.push(cb);var UTF8Decoder=typeof TextDecoder!="undefined"?new TextDecoder:undefined;var UTF8ArrayToString=(heapOrArray,idx=0,maxBytesToRead=NaN)=>{idx>>>=0;var endIdx=idx+maxBytesToRead;var endPtr=idx;while(heapOrArray[endPtr]&&!(endPtr>=endIdx))++endPtr;if(endPtr-idx>16&&heapOrArray.buffer&&UTF8Decoder){return UTF8Decoder.decode(heapOrArray.subarray(idx,endPtr))}var str="";while(idx<endPtr){var u0=heapOrArray[idx++];if(!(u0&128)){str+=String.fromCharCode(u0);continue}var u1=heapOrArray[idx++]&63;if((u0&224)==192){str+=String.fromCharCode((u0&31)<<6|u1);continue}var u2=heapOrArray[idx++]&63;if((u0&240)==224){u0=(u0&15)<<12|u1<<6|u2}else{u0=(u0&7)<<18|u1<<12|u2<<6|heapOrArray[idx++]&63}if(u0<65536){str+=String.fromCharCode(u0)}else{var ch=u0-65536;str+=String.fromCharCode(55296|ch>>10,56320|ch&1023)}}return str};var getDylinkMetadata=binary=>{var offset=0;var end=0;function getU8(){return binary[offset++]}function getLEB(){var ret=0;var mul=1;while(1){var byte=binary[offset++];ret+=(byte&127)*mul;mul*=128;if(!(byte&128))break}return ret}function getString(){var len=getLEB();offset+=len;return UTF8ArrayToString(binary,offset-len,len)}function getStringList(){var count=getLEB();var rtn=[];while(count--)rtn.push(getString());return rtn}function failIf(condition,message){if(condition)throw new Error(message)}if(binary instanceof WebAssembly.Module){var dylinkSection=WebAssembly.Module.customSections(binary,"dylink.0");failIf(dylinkSection.length===0,"need dylink section");binary=new Uint8Array(dylinkSection[0]);end=binary.length}else{var int32View=new Uint32Array(new Uint8Array(binary.subarray(0,24)).buffer);var magicNumberFound=int32View[0]==1836278016;failIf(!magicNumberFound,"need to see wasm magic number");failIf(binary[8]!==0,"need the dylink section to be first");offset=9;var section_size=getLEB();end=offset+section_size;var name=getString();failIf(name!=="dylink.0")}var customSection={neededDynlibs:[],tlsExports:new Set,weakImports:new Set,runtimePaths:[]};var WASM_DYLINK_MEM_INFO=1;var WASM_DYLINK_NEEDED=2;var WASM_DYLINK_EXPORT_INFO=3;var WASM_DYLINK_IMPORT_INFO=4;var WASM_DYLINK_RUNTIME_PATH=5;var WASM_SYMBOL_TLS=256;var WASM_SYMBOL_BINDING_MASK=3;var WASM_SYMBOL_BINDING_WEAK=1;while(offset<end){var subsectionType=getU8();var subsectionSize=getLEB();if(subsectionType===WASM_DYLINK_MEM_INFO){customSection.memorySize=getLEB();customSection.memoryAlign=getLEB();customSection.tableSize=getLEB();customSection.tableAlign=getLEB()}else if(subsectionType===WASM_DYLINK_NEEDED){customSection.neededDynlibs=getStringList()}else if(subsectionType===WASM_DYLINK_EXPORT_INFO){var count=getLEB();while(count--){var symname=getString();var flags=getLEB();if(flags&WASM_SYMBOL_TLS){customSection.tlsExports.add(symname)}}}else if(subsectionType===WASM_DYLINK_IMPORT_INFO){var count=getLEB();while(count--){var modname=getString();var symname=getString();var flags=getLEB();if((flags&WASM_SYMBOL_BINDING_MASK)==WASM_SYMBOL_BINDING_WEAK){customSection.weakImports.add(symname)}}}else if(subsectionType===WASM_DYLINK_RUNTIME_PATH){customSection.runtimePaths=getStringList()}else{offset+=subsectionSize}}return customSection};function getValue(ptr,type="i8"){if(type.endsWith("*"))type="*";switch(type){case"i1":return HEAP8[ptr>>>0];case"i8":return HEAP8[ptr>>>0];case"i16":return HEAP16[ptr>>>1>>>0];case"i32":return HEAP32[ptr>>>2>>>0];case"i64":return HEAP64[ptr>>>3>>>0];case"float":return HEAPF32[ptr>>>2>>>0];case"double":return HEAPF64[ptr>>>3>>>0];case"*":return HEAPU32[ptr>>>2>>>0];default:abort(`invalid type for getValue: ${type}`)}}var newDSO=(name,handle,syms)=>{var dso={refcount:Infinity,name,exports:syms,global:true};LDSO.loadedLibsByName[name]=dso;if(handle!=undefined){LDSO.loadedLibsByHandle[handle]=dso}return dso};var LDSO={loadedLibsByName:{},loadedLibsByHandle:{},init(){newDSO("__main__",0,wasmImports)}};var ___heap_base=8809120;var alignMemory=(size,alignment)=>Math.ceil(size/alignment)*alignment;var getMemory=size=>{if(runtimeInitialized){return _calloc(size,1)}var ret=___heap_base;var end=ret+alignMemory(size,16);___heap_base=end;GOT["__heap_base"].value=end;return ret};var isInternalSym=symName=>["__cpp_exception","__c_longjmp","__wasm_apply_data_relocs","__dso_handle","__tls_size","__tls_align","__set_stack_limits","_emscripten_tls_init","__wasm_init_tls","__wasm_call_ctors","__start_em_asm","__stop_em_asm","__start_em_js","__stop_em_js"].includes(symName)||symName.startsWith("__em_js__");var uleb128Encode=(n,target)=>{if(n<128){target.push(n)}else{target.push(n%128|128,n>>7)}};var sigToWasmTypes=sig=>{var typeNames={i:"i32",j:"i64",f:"f32",d:"f64",e:"externref",p:"i32"};var type={parameters:[],results:sig[0]=="v"?[]:[typeNames[sig[0]]]};for(var i=1;i<sig.length;++i){type.parameters.push(typeNames[sig[i]])}return type};var generateFuncType=(sig,target)=>{var sigRet=sig.slice(0,1);var sigParam=sig.slice(1);var typeCodes={i:127,p:127,j:126,f:125,d:124,e:111};target.push(96);uleb128Encode(sigParam.length,target);for(var paramType of sigParam){target.push(typeCodes[paramType])}if(sigRet=="v"){target.push(0)}else{target.push(1,typeCodes[sigRet])}};var convertJsFunctionToWasm=(func,sig)=>{if(typeof WebAssembly.Function=="function"){return new WebAssembly.Function(sigToWasmTypes(sig),func)}var typeSectionBody=[1];generateFuncType(sig,typeSectionBody);var bytes=[0,97,115,109,1,0,0,0,1];uleb128Encode(typeSectionBody.length,bytes);bytes.push(...typeSectionBody);bytes.push(2,7,1,1,101,1,102,0,0,7,5,1,1,102,0,0);var module=new WebAssembly.Module(new Uint8Array(bytes));var instance=new WebAssembly.Instance(module,{e:{f:func}});var wrappedFunc=instance.exports["f"];return wrappedFunc};var wasmTableMirror=[];var wasmTable=new WebAssembly.Table({initial:6074,element:"anyfunc"});var getWasmTableEntry=funcPtr=>{var func=wasmTableMirror[funcPtr];if(!func){wasmTableMirror[funcPtr]=func=wasmTable.get(funcPtr)}return func};var updateTableMap=(offset,count)=>{if(functionsInTableMap){for(var i=offset;i<offset+count;i++){var item=getWasmTableEntry(i);if(item){functionsInTableMap.set(item,i)}}}};var functionsInTableMap;var getFunctionAddress=func=>{if(!functionsInTableMap){functionsInTableMap=new WeakMap;updateTableMap(0,wasmTable.length)}return functionsInTableMap.get(func)||0};var freeTableIndexes=[];var getEmptyTableSlot=()=>{if(freeTableIndexes.length){return freeTableIndexes.pop()}try{wasmTable.grow(1)}catch(err){if(!(err instanceof RangeError)){throw err}throw"Unable to grow wasm table. Set ALLOW_TABLE_GROWTH."}return wasmTable.length-1};var setWasmTableEntry=(idx,func)=>{wasmTable.set(idx,func);wasmTableMirror[idx]=wasmTable.get(idx)};var addFunction=(func,sig)=>{var rtn=getFunctionAddress(func);if(rtn){return rtn}var ret=getEmptyTableSlot();try{setWasmTableEntry(ret,func)}catch(err){if(!(err instanceof TypeError)){throw err}var wrapped=convertJsFunctionToWasm(func,sig);setWasmTableEntry(ret,wrapped)}functionsInTableMap.set(func,ret);return ret};var updateGOT=(exports,replace)=>{for(var symName in exports){if(isInternalSym(symName)){continue}var value=exports[symName];GOT[symName]||=new WebAssembly.Global({value:"i32",mutable:true});if(replace||GOT[symName].value==0){if(typeof value=="function"){GOT[symName].value=addFunction(value)}else if(typeof value=="number"){GOT[symName].value=value}else{err(`unhandled export type for '${symName}': ${typeof value}`)}}}};var relocateExports=(exports,memoryBase,replace)=>{var relocated={};for(var e in exports){var value=exports[e];if(typeof value=="object"){value=value.value}if(typeof value=="number"){value+=memoryBase}relocated[e]=value}updateGOT(relocated,replace);return relocated};var isSymbolDefined=symName=>{var existing=wasmImports[symName];if(!existing||existing.stub){return false}return true};var resolveGlobalSymbol=(symName,direct=false)=>{var sym;if(isSymbolDefined(symName)){sym=wasmImports[symName]}return{sym,name:symName}};var onPostCtors=[];var addOnPostCtor=cb=>onPostCtors.push(cb);var UTF8ToString=(ptr,maxBytesToRead)=>{ptr>>>=0;return ptr?UTF8ArrayToString(HEAPU8,ptr,maxBytesToRead):""};var loadWebAssemblyModule=(binary,flags,libName,localScope,handle)=>{var metadata=getDylinkMetadata(binary);currentModuleWeakSymbols=metadata.weakImports;function loadModule(){var memAlign=Math.pow(2,metadata.memoryAlign);var memoryBase=metadata.memorySize?alignMemory(getMemory(metadata.memorySize+memAlign),memAlign):0;var tableBase=metadata.tableSize?wasmTable.length:0;if(handle){HEAP8[handle+8>>>0]=1;HEAPU32[handle+12>>>2>>>0]=memoryBase;HEAP32[handle+16>>>2>>>0]=metadata.memorySize;HEAPU32[handle+20>>>2>>>0]=tableBase;HEAP32[handle+24>>>2>>>0]=metadata.tableSize}if(metadata.tableSize){wasmTable.grow(metadata.tableSize)}var moduleExports;function resolveSymbol(sym){var resolved=resolveGlobalSymbol(sym).sym;if(!resolved&&localScope){resolved=localScope[sym]}if(!resolved){resolved=moduleExports[sym]}return resolved}var proxyHandler={get(stubs,prop){switch(prop){case"__memory_base":return memoryBase;case"__table_base":return tableBase}if(prop in wasmImports&&!wasmImports[prop].stub){var res=wasmImports[prop];return res}if(!(prop in stubs)){var resolved;stubs[prop]=(...args)=>{resolved||=resolveSymbol(prop);if(!resolved){throw new Error(`Dynamic linking error: cannot resolve symbol ${prop}`)}return resolved(...args)}}return stubs[prop]}};var proxy=new Proxy({},proxyHandler);var info={"GOT.mem":new Proxy({},GOTHandler),"GOT.func":new Proxy({},GOTHandler),env:proxy,wasi_snapshot_preview1:proxy};function postInstantiation(module,instance){updateTableMap(tableBase,metadata.tableSize);moduleExports=relocateExports(instance.exports,memoryBase);if(!flags.allowUndefined){reportUndefinedSymbols()}function addEmAsm(addr,body){var args=[];var arity=0;for(;arity<16;arity++){if(body.indexOf("$"+arity)!=-1){args.push("$"+arity)}else{break}}args=args.join(",");var func=`(${args}) => { ${body} };`;ASM_CONSTS[start]=eval(func)}if("__start_em_asm"in moduleExports){var start=moduleExports["__start_em_asm"];var stop=moduleExports["__stop_em_asm"];while(start<stop){var jsString=UTF8ToString(start);addEmAsm(start,jsString);start=HEAPU8.indexOf(0,start)+1}}function addEmJs(name,cSig,body){var jsArgs=[];cSig=cSig.slice(1,-1);if(cSig!="void"){cSig=cSig.split(",");for(var i in cSig){var jsArg=cSig[i].split(" ").pop();jsArgs.push(jsArg.replace("*",""))}}var func=`(${jsArgs}) => ${body};`;moduleExports[name]=eval(func)}for(var name in moduleExports){if(name.startsWith("__em_js__")){var start=moduleExports[name];var jsString=UTF8ToString(start);var parts=jsString.split("<::>");addEmJs(name.replace("__em_js__",""),parts[0],parts[1]);delete moduleExports[name]}}var applyRelocs=moduleExports["__wasm_apply_data_relocs"];if(applyRelocs){if(runtimeInitialized){applyRelocs()}else{__RELOC_FUNCS__.push(applyRelocs)}}var init=moduleExports["__wasm_call_ctors"];if(init){if(runtimeInitialized){init()}else{addOnPostCtor(init)}}return moduleExports}if(flags.loadAsync){return(async()=>{var instance;if(binary instanceof WebAssembly.Module){instance=new WebAssembly.Instance(binary,info)}else{({module:binary,instance}=await WebAssembly.instantiate(binary,info))}return postInstantiation(binary,instance)})()}var module=binary instanceof WebAssembly.Module?binary:new WebAssembly.Module(binary);var instance=new WebAssembly.Instance(module,info);return postInstantiation(module,instance)}flags={...flags,rpath:{parentLibPath:libName,paths:metadata.runtimePaths}};if(flags.loadAsync){return metadata.neededDynlibs.reduce((chain,needed)=>chain.then(()=>{needed=findLibraryFS(needed,flags.rpath)??needed;return loadDynamicLibrary(needed,flags,localScope)}),Promise.resolve()).then(loadModule)}metadata.neededDynlibs.forEach(needed=>{needed=findLibraryFS(needed,flags.rpath)??needed;return loadDynamicLibrary(needed,flags,localScope)});return loadModule()};var mergeLibSymbols=(exports,libName)=>{for(var[sym,exp]of Object.entries(exports)){const setImport=target=>{if(!isSymbolDefined(target)){wasmImports[target]=exp}};setImport(sym);const main_alias="__main_argc_argv";if(sym=="main"){setImport(main_alias)}if(sym==main_alias){setImport("main")}}};var asyncLoad=async url=>{var arrayBuffer=await readAsync(url);return new Uint8Array(arrayBuffer)};var preloadPlugins=[];var registerWasmPlugin=()=>{var wasmPlugin={promiseChainEnd:Promise.resolve(),canHandle:name=>!Module["noWasmDecoding"]&&name.endsWith(".so"),handle:(byteArray,name,onload,onerror)=>{wasmPlugin["promiseChainEnd"]=wasmPlugin["promiseChainEnd"].then(()=>loadWebAssemblyModule(byteArray,{loadAsync:true,nodelete:true},name,{})).then(exports=>{preloadedWasm[name]=exports;onload(byteArray)},error=>{err(`failed to instantiate wasm: ${name}: ${error}`);onerror()})}};preloadPlugins.push(wasmPlugin)};var preloadedWasm={};var PATH={isAbs:path=>path.charAt(0)==="/",splitPath:filename=>{var splitPathRe=/^(\/?|)([\s\S]*?)((?:\.{1,2}|[^\/]+?|)(\.[^.\/]*|))(?:[\/]*)$/;return splitPathRe.exec(filename).slice(1)},normalizeArray:(parts,allowAboveRoot)=>{var up=0;for(var i=parts.length-1;i>=0;i--){var last=parts[i];if(last==="."){parts.splice(i,1)}else if(last===".."){parts.splice(i,1);up++}else if(up){parts.splice(i,1);up--}}if(allowAboveRoot){for(;up;up--){parts.unshift("..")}}return parts},normalize:path=>{var isAbsolute=PATH.isAbs(path),trailingSlash=path.slice(-1)==="/";path=PATH.normalizeArray(path.split("/").filter(p=>!!p),!isAbsolute).join("/");if(!path&&!isAbsolute){path="."}if(path&&trailingSlash){path+="/"}return(isAbsolute?"/":"")+path},dirname:path=>{var result=PATH.splitPath(path),root=result[0],dir=result[1];if(!root&&!dir){return"."}if(dir){dir=dir.slice(0,-1)}return root+dir},basename:path=>path&&path.match(/([^\/]+|\/)\/*$/)[1],join:(...paths)=>PATH.normalize(paths.join("/")),join2:(l,r)=>PATH.normalize(l+"/"+r)};var replaceORIGIN=(parentLibName,rpath)=>{if(rpath.startsWith("$ORIGIN")){var origin=PATH.dirname(parentLibName);return rpath.replace("$ORIGIN",origin)}return rpath};var stackSave=()=>_emscripten_stack_get_current();var stackRestore=val=>__emscripten_stack_restore(val);var withStackSave=f=>{var stack=stackSave();var ret=f();stackRestore(stack);return ret};var stackAlloc=sz=>__emscripten_stack_alloc(sz);var lengthBytesUTF8=str=>{var len=0;for(var i=0;i<str.length;++i){var c=str.charCodeAt(i);if(c<=127){len++}else if(c<=2047){len+=2}else if(c>=55296&&c<=57343){len+=4;++i}else{len+=3}}return len};var stringToUTF8Array=(str,heap,outIdx,maxBytesToWrite)=>{outIdx>>>=0;if(!(maxBytesToWrite>0))return 0;var startIdx=outIdx;var endIdx=outIdx+maxBytesToWrite-1;for(var i=0;i<str.length;++i){var u=str.charCodeAt(i);if(u>=55296&&u<=57343){var u1=str.charCodeAt(++i);u=65536+((u&1023)<<10)|u1&1023}if(u<=127){if(outIdx>=endIdx)break;heap[outIdx++>>>0]=u}else if(u<=2047){if(outIdx+1>=endIdx)break;heap[outIdx++>>>0]=192|u>>6;heap[outIdx++>>>0]=128|u&63}else if(u<=65535){if(outIdx+2>=endIdx)break;heap[outIdx++>>>0]=224|u>>12;heap[outIdx++>>>0]=128|u>>6&63;heap[outIdx++>>>0]=128|u&63}else{if(outIdx+3>=endIdx)break;heap[outIdx++>>>0]=240|u>>18;heap[outIdx++>>>0]=128|u>>12&63;heap[outIdx++>>>0]=128|u>>6&63;heap[outIdx++>>>0]=128|u&63}}heap[outIdx>>>0]=0;return outIdx-startIdx};var stringToUTF8=(str,outPtr,maxBytesToWrite)=>stringToUTF8Array(str,HEAPU8,outPtr,maxBytesToWrite);var stringToUTF8OnStack=str=>{var size=lengthBytesUTF8(str)+1;var ret=stackAlloc(size);stringToUTF8(str,ret,size);return ret};var base64Decode=b64=>{if(ENVIRONMENT_IS_NODE){var buf=Buffer.from(b64,"base64");return new Uint8Array(buf.buffer,buf.byteOffset,buf.length)}var b1,b2,i=0,j=0,bLength=b64.length;var output=new Uint8Array((bLength*3>>2)-(b64[bLength-2]=="=")-(b64[bLength-1]=="="));for(;i<bLength;i+=4,j+=3){b1=base64ReverseLookup[b64.charCodeAt(i+1)];b2=base64ReverseLookup[b64.charCodeAt(i+2)];output[j]=base64ReverseLookup[b64.charCodeAt(i)]<<2|b1>>4;output[j+1]=b1<<4|b2>>2;output[j+2]=b2<<6|base64ReverseLookup[b64.charCodeAt(i+3)]}return output};var initRandomFill=()=>{if(ENVIRONMENT_IS_NODE){var nodeCrypto=require("crypto");return view=>nodeCrypto.randomFillSync(view)}if(ENVIRONMENT_IS_SHELL){return view=>{if(!os.system){throw new Error("randomFill not supported on d8 unless --enable-os-system is passed")}const b64=os.system("sh",["-c",`head -c${view.byteLength} /dev/urandom | base64 --wrap=0`]);view.set(base64Decode(b64))}}return view=>crypto.getRandomValues(view)};var randomFill=view=>{(randomFill=initRandomFill())(view)};var PATH_FS={resolve:(...args)=>{var resolvedPath="",resolvedAbsolute=false;for(var i=args.length-1;i>=-1&&!resolvedAbsolute;i--){var path=i>=0?args[i]:FS.cwd();if(typeof path!="string"){throw new TypeError("Arguments to path.resolve must be strings")}else if(!path){return""}resolvedPath=path+"/"+resolvedPath;resolvedAbsolute=PATH.isAbs(path)}resolvedPath=PATH.normalizeArray(resolvedPath.split("/").filter(p=>!!p),!resolvedAbsolute).join("/");return(resolvedAbsolute?"/":"")+resolvedPath||"."},relative:(from,to)=>{from=PATH_FS.resolve(from).slice(1);to=PATH_FS.resolve(to).slice(1);function trim(arr){var start=0;for(;start<arr.length;start++){if(arr[start]!=="")break}var end=arr.length-1;for(;end>=0;end--){if(arr[end]!=="")break}if(start>end)return[];return arr.slice(start,end-start+1)}var fromParts=trim(from.split("/"));var toParts=trim(to.split("/"));var length=Math.min(fromParts.length,toParts.length);var samePartsLength=length;for(var i=0;i<length;i++){if(fromParts[i]!==toParts[i]){samePartsLength=i;break}}var outputParts=[];for(var i=samePartsLength;i<fromParts.length;i++){outputParts.push("..")}outputParts=outputParts.concat(toParts.slice(samePartsLength));return outputParts.join("/")}};var FS_stdin_getChar_buffer=[];var intArrayFromString=(stringy,dontAddNull,length)=>{var len=length>0?length:lengthBytesUTF8(stringy)+1;var u8array=new Array(len);var numBytesWritten=stringToUTF8Array(stringy,u8array,0,u8array.length);if(dontAddNull)u8array.length=numBytesWritten;return u8array};var FS_stdin_getChar=()=>{if(!FS_stdin_getChar_buffer.length){var result=null;if(ENVIRONMENT_IS_NODE){var BUFSIZE=256;var buf=Buffer.alloc(BUFSIZE);var bytesRead=0;var fd=process.stdin.fd;try{bytesRead=fs.readSync(fd,buf,0,BUFSIZE)}catch(e){if(e.toString().includes("EOF"))bytesRead=0;else throw e}if(bytesRead>0){result=buf.slice(0,bytesRead).toString("utf-8")}}else if(typeof window!="undefined"&&typeof window.prompt=="function"){result=window.prompt("Input: ");if(result!==null){result+="\n"}}else if(typeof readline=="function"){result=readline();if(result){result+="\n"}}else{}if(!result){return null}FS_stdin_getChar_buffer=intArrayFromString(result,true)}return FS_stdin_getChar_buffer.shift()};var TTY={ttys:[],init(){},shutdown(){},register(dev,ops){TTY.ttys[dev]={input:[],output:[],ops};FS.registerDevice(dev,TTY.stream_ops)},stream_ops:{open(stream){var tty=TTY.ttys[stream.node.rdev];if(!tty){throw new FS.ErrnoError(43)}stream.tty=tty;stream.seekable=false},close(stream){stream.tty.ops.fsync(stream.tty)},fsync(stream){stream.tty.ops.fsync(stream.tty)},read(stream,buffer,offset,length,pos){if(!stream.tty||!stream.tty.ops.get_char){throw new FS.ErrnoError(60)}var bytesRead=0;for(var i=0;i<length;i++){var result;try{result=stream.tty.ops.get_char(stream.tty)}catch(e){throw new FS.ErrnoError(29)}if(result===undefined&&bytesRead===0){throw new FS.ErrnoError(6)}if(result===null||result===undefined)break;bytesRead++;buffer[offset+i]=result}if(bytesRead){stream.node.atime=Date.now()}return bytesRead},write(stream,buffer,offset,length,pos){if(!stream.tty||!stream.tty.ops.put_char){throw new FS.ErrnoError(60)}try{for(var i=0;i<length;i++){stream.tty.ops.put_char(stream.tty,buffer[offset+i])}}catch(e){throw new FS.ErrnoError(29)}if(length){stream.node.mtime=stream.node.ctime=Date.now()}return i}},default_tty_ops:{get_char(tty){return FS_stdin_getChar()},put_char(tty,val){if(val===null||val===10){out(UTF8ArrayToString(tty.output));tty.output=[]}else{if(val!=0)tty.output.push(val)}},fsync(tty){if(tty.output?.length>0){out(UTF8ArrayToString(tty.output));tty.output=[]}},ioctl_tcgets(tty){return{c_iflag:25856,c_oflag:5,c_cflag:191,c_lflag:35387,c_cc:[3,28,127,21,4,0,1,0,17,19,26,0,18,15,23,22,0,0,0,0,0,0,0,0,0,0,0,0,0,0,0,0]}},ioctl_tcsets(tty,optional_actions,data){return 0},ioctl_tiocgwinsz(tty){return[24,80]}},default_tty1_ops:{put_char(tty,val){if(val===null||val===10){err(UTF8ArrayToString(tty.output));tty.output=[]}else{if(val!=0)tty.output.push(val)}},fsync(tty){if(tty.output?.length>0){err(UTF8ArrayToString(tty.output));tty.output=[]}}}};var zeroMemory=(ptr,size)=>HEAPU8.fill(0,ptr,ptr+size);var mmapAlloc=size=>{size=alignMemory(size,65536);var ptr=_emscripten_builtin_memalign(65536,size);if(ptr)zeroMemory(ptr,size);return ptr};var MEMFS={ops_table:null,mount(mount){return MEMFS.createNode(null,"/",16895,0)},createNode(parent,name,mode,dev){if(FS.isBlkdev(mode)||FS.isFIFO(mode)){throw new FS.ErrnoError(63)}MEMFS.ops_table||={dir:{node:{getattr:MEMFS.node_ops.getattr,setattr:MEMFS.node_ops.setattr,lookup:MEMFS.node_ops.lookup,mknod:MEMFS.node_ops.mknod,rename:MEMFS.node_ops.rename,unlink:MEMFS.node_ops.unlink,rmdir:MEMFS.node_ops.rmdir,readdir:MEMFS.node_ops.readdir,symlink:MEMFS.node_ops.symlink},stream:{llseek:MEMFS.stream_ops.llseek}},file:{node:{getattr:MEMFS.node_ops.getattr,setattr:MEMFS.node_ops.setattr},stream:{llseek:MEMFS.stream_ops.llseek,read:MEMFS.stream_ops.read,write:MEMFS.stream_ops.write,mmap:MEMFS.stream_ops.mmap,msync:MEMFS.stream_ops.msync}},link:{node:{getattr:MEMFS.node_ops.getattr,setattr:MEMFS.node_ops.setattr,readlink:MEMFS.node_ops.readlink},stream:{}},chrdev:{node:{getattr:MEMFS.node_ops.getattr,setattr:MEMFS.node_ops.setattr},stream:FS.chrdev_stream_ops}};var node=FS.createNode(parent,name,mode,dev);if(FS.isDir(node.mode)){node.node_ops=MEMFS.ops_table.dir.node;node.stream_ops=MEMFS.ops_table.dir.stream;node.contents={}}else if(FS.isFile(node.mode)){node.node_ops=MEMFS.ops_table.file.node;node.stream_ops=MEMFS.ops_table.file.stream;node.usedBytes=0;node.contents=null}else if(FS.isLink(node.mode)){node.node_ops=MEMFS.ops_table.link.node;node.stream_ops=MEMFS.ops_table.link.stream}else if(FS.isChrdev(node.mode)){node.node_ops=MEMFS.ops_table.chrdev.node;node.stream_ops=MEMFS.ops_table.chrdev.stream}node.atime=node.mtime=node.ctime=Date.now();if(parent){parent.contents[name]=node;parent.atime=parent.mtime=parent.ctime=node.atime}return node},getFileDataAsTypedArray(node){if(!node.contents)return new Uint8Array(0);if(node.contents.subarray)return node.contents.subarray(0,node.usedBytes);return new Uint8Array(node.contents)},expandFileStorage(node,newCapacity){var prevCapacity=node.contents?node.contents.length:0;if(prevCapacity>=newCapacity)return;var CAPACITY_DOUBLING_MAX=1024*1024;newCapacity=Math.max(newCapacity,prevCapacity*(prevCapacity<CAPACITY_DOUBLING_MAX?2:1.125)>>>0);if(prevCapacity!=0)newCapacity=Math.max(newCapacity,256);var oldContents=node.contents;node.contents=new Uint8Array(newCapacity);if(node.usedBytes>0)node.contents.set(oldContents.subarray(0,node.usedBytes),0)},resizeFileStorage(node,newSize){if(node.usedBytes==newSize)return;if(newSize==0){node.contents=null;node.usedBytes=0}else{var oldContents=node.contents;node.contents=new Uint8Array(newSize);if(oldContents){node.contents.set(oldContents.subarray(0,Math.min(newSize,node.usedBytes)))}node.usedBytes=newSize}},node_ops:{getattr(node){var attr={};attr.dev=FS.isChrdev(node.mode)?node.id:1;attr.ino=node.id;attr.mode=node.mode;attr.nlink=1;attr.uid=0;attr.gid=0;attr.rdev=node.rdev;if(FS.isDir(node.mode)){attr.size=4096}else if(FS.isFile(node.mode)){attr.size=node.usedBytes}else if(FS.isLink(node.mode)){attr.size=node.link.length}else{attr.size=0}attr.atime=new Date(node.atime);attr.mtime=new Date(node.mtime);attr.ctime=new Date(node.ctime);attr.blksize=4096;attr.blocks=Math.ceil(attr.size/attr.blksize);return attr},setattr(node,attr){for(const key of["mode","atime","mtime","ctime"]){if(attr[key]!=null){node[key]=attr[key]}}if(attr.size!==undefined){MEMFS.resizeFileStorage(node,attr.size)}},lookup(parent,name){throw MEMFS.doesNotExistError},mknod(parent,name,mode,dev){return MEMFS.createNode(parent,name,mode,dev)},rename(old_node,new_dir,new_name){var new_node;try{new_node=FS.lookupNode(new_dir,new_name)}catch(e){}if(new_node){if(FS.isDir(old_node.mode)){for(var i in new_node.contents){throw new FS.ErrnoError(55)}}FS.hashRemoveNode(new_node)}delete old_node.parent.contents[old_node.name];new_dir.contents[new_name]=old_node;old_node.name=new_name;new_dir.ctime=new_dir.mtime=old_node.parent.ctime=old_node.parent.mtime=Date.now()},unlink(parent,name){delete parent.contents[name];parent.ctime=parent.mtime=Date.now()},rmdir(parent,name){var node=FS.lookupNode(parent,name);for(var i in node.contents){throw new FS.ErrnoError(55)}delete parent.contents[name];parent.ctime=parent.mtime=Date.now()},readdir(node){return[".","..",...Object.keys(node.contents)]},symlink(parent,newname,oldpath){var node=MEMFS.createNode(parent,newname,511|40960,0);node.link=oldpath;return node},readlink(node){if(!FS.isLink(node.mode)){throw new FS.ErrnoError(28)}return node.link}},stream_ops:{read(stream,buffer,offset,length,position){var contents=stream.node.contents;if(position>=stream.node.usedBytes)return 0;var size=Math.min(stream.node.usedBytes-position,length);if(size>8&&contents.subarray){buffer.set(contents.subarray(position,position+size),offset)}else{for(var i=0;i<size;i++)buffer[offset+i]=contents[position+i]}return size},write(stream,buffer,offset,length,position,canOwn){if(buffer.buffer===HEAP8.buffer){canOwn=false}if(!length)return 0;var node=stream.node;node.mtime=node.ctime=Date.now();if(buffer.subarray&&(!node.contents||node.contents.subarray)){if(canOwn){node.contents=buffer.subarray(offset,offset+length);node.usedBytes=length;return length}else if(node.usedBytes===0&&position===0){node.contents=buffer.slice(offset,offset+length);node.usedBytes=length;return length}else if(position+length<=node.usedBytes){node.contents.set(buffer.subarray(offset,offset+length),position);return length}}MEMFS.expandFileStorage(node,position+length);if(node.contents.subarray&&buffer.subarray){node.contents.set(buffer.subarray(offset,offset+length),position)}else{for(var i=0;i<length;i++){node.contents[position+i]=buffer[offset+i]}}node.usedBytes=Math.max(node.usedBytes,position+length);return length},llseek(stream,offset,whence){var position=offset;if(whence===1){position+=stream.position}else if(whence===2){if(FS.isFile(stream.node.mode)){position+=stream.node.usedBytes}}if(position<0){throw new FS.ErrnoError(28)}return position},mmap(stream,length,position,prot,flags){if(!FS.isFile(stream.node.mode)){throw new FS.ErrnoError(43)}var ptr;var allocated;var contents=stream.node.contents;if(!(flags&2)&&contents&&contents.buffer===HEAP8.buffer){allocated=false;ptr=contents.byteOffset}else{allocated=true;ptr=mmapAlloc(length);if(!ptr){throw new FS.ErrnoError(48)}if(contents){if(position>0||position+length<contents.length){if(contents.subarray){contents=contents.subarray(position,position+length)}else{contents=Array.prototype.slice.call(contents,position,position+length)}}HEAP8.set(contents,ptr>>>0)}}return{ptr,allocated}},msync(stream,buffer,offset,length,mmapFlags){MEMFS.stream_ops.write(stream,buffer,0,length,offset,false);return 0}}};var FS_createDataFile=(...args)=>FS.createDataFile(...args);var FS_handledByPreloadPlugin=(byteArray,fullname,finish,onerror)=>{if(typeof Browser!="undefined")Browser.init();var handled=false;preloadPlugins.forEach(plugin=>{if(handled)return;if(plugin["canHandle"](fullname)){plugin["handle"](byteArray,fullname,finish,onerror);handled=true}});return handled};var FS_createPreloadedFile=(parent,name,url,canRead,canWrite,onload,onerror,dontCreateFile,canOwn,preFinish)=>{var fullname=name?PATH_FS.resolve(PATH.join2(parent,name)):parent;var dep=getUniqueRunDependency(`cp ${fullname}`);function processData(byteArray){function finish(byteArray){preFinish?.();if(!dontCreateFile){FS_createDataFile(parent,name,byteArray,canRead,canWrite,canOwn)}onload?.();removeRunDependency(dep)}if(FS_handledByPreloadPlugin(byteArray,fullname,finish,()=>{onerror?.();removeRunDependency(dep)})){return}finish(byteArray)}addRunDependency(dep);if(typeof url=="string"){asyncLoad(url).then(processData,onerror)}else{processData(url)}};var FS_modeStringToFlags=str=>{var flagModes={r:0,"r+":2,w:512|64|1,"w+":512|64|2,a:1024|64|1,"a+":1024|64|2};var flags=flagModes[str];if(typeof flags=="undefined"){throw new Error(`Unknown file open mode: ${str}`)}return flags};var FS_getMode=(canRead,canWrite)=>{var mode=0;if(canRead)mode|=292|73;if(canWrite)mode|=146;return mode};var IDBFS={dbs:{},indexedDB:()=>{if(typeof indexedDB!="undefined")return indexedDB;var ret=null;if(typeof window=="object")ret=window.indexedDB||window.mozIndexedDB||window.webkitIndexedDB||window.msIndexedDB;return ret},DB_VERSION:21,DB_STORE_NAME:"FILE_DATA",queuePersist:mount=>{function onPersistComplete(){if(mount.idbPersistState==="again")startPersist();else mount.idbPersistState=0}function startPersist(){mount.idbPersistState="idb";IDBFS.syncfs(mount,false,onPersistComplete)}if(!mount.idbPersistState){mount.idbPersistState=setTimeout(startPersist,0)}else if(mount.idbPersistState==="idb"){mount.idbPersistState="again"}},mount:mount=>{var mnt=MEMFS.mount(mount);if(mount?.opts?.autoPersist){mnt.idbPersistState=0;var memfs_node_ops=mnt.node_ops;mnt.node_ops={...mnt.node_ops};mnt.node_ops.mknod=(parent,name,mode,dev)=>{var node=memfs_node_ops.mknod(parent,name,mode,dev);node.node_ops=mnt.node_ops;node.idbfs_mount=mnt.mount;node.memfs_stream_ops=node.stream_ops;node.stream_ops={...node.stream_ops};node.stream_ops.write=(stream,buffer,offset,length,position,canOwn)=>{stream.node.isModified=true;return node.memfs_stream_ops.write(stream,buffer,offset,length,position,canOwn)};node.stream_ops.close=stream=>{var n=stream.node;if(n.isModified){IDBFS.queuePersist(n.idbfs_mount);n.isModified=false}if(n.memfs_stream_ops.close)return n.memfs_stream_ops.close(stream)};return node};mnt.node_ops.mkdir=(...args)=>(IDBFS.queuePersist(mnt.mount),memfs_node_ops.mkdir(...args));mnt.node_ops.rmdir=(...args)=>(IDBFS.queuePersist(mnt.mount),memfs_node_ops.rmdir(...args));mnt.node_ops.symlink=(...args)=>(IDBFS.queuePersist(mnt.mount),memfs_node_ops.symlink(...args));mnt.node_ops.unlink=(...args)=>(IDBFS.queuePersist(mnt.mount),memfs_node_ops.unlink(...args));mnt.node_ops.rename=(...args)=>(IDBFS.queuePersist(mnt.mount),memfs_node_ops.rename(...args))}return mnt},syncfs:(mount,populate,callback)=>{IDBFS.getLocalSet(mount,(err,local)=>{if(err)return callback(err);IDBFS.getRemoteSet(mount,(err,remote)=>{if(err)return callback(err);var src=populate?remote:local;var dst=populate?local:remote;IDBFS.reconcile(src,dst,callback)})})},quit:()=>{Object.values(IDBFS.dbs).forEach(value=>value.close());IDBFS.dbs={}},getDB:(name,callback)=>{var db=IDBFS.dbs[name];if(db){return callback(null,db)}var req;try{req=IDBFS.indexedDB().open(name,IDBFS.DB_VERSION)}catch(e){return callback(e)}if(!req){return callback("Unable to connect to IndexedDB")}req.onupgradeneeded=e=>{var db=e.target.result;var transaction=e.target.transaction;var fileStore;if(db.objectStoreNames.contains(IDBFS.DB_STORE_NAME)){fileStore=transaction.objectStore(IDBFS.DB_STORE_NAME)}else{fileStore=db.createObjectStore(IDBFS.DB_STORE_NAME)}if(!fileStore.indexNames.contains("timestamp")){fileStore.createIndex("timestamp","timestamp",{unique:false})}};req.onsuccess=()=>{db=req.result;IDBFS.dbs[name]=db;callback(null,db)};req.onerror=e=>{callback(e.target.error);e.preventDefault()}},getLocalSet:(mount,callback)=>{var entries={};function isRealDir(p){return p!=="."&&p!==".."}function toAbsolute(root){return p=>PATH.join2(root,p)}var check=FS.readdir(mount.mountpoint).filter(isRealDir).map(toAbsolute(mount.mountpoint));while(check.length){var path=check.pop();var stat;try{stat=FS.stat(path)}catch(e){return callback(e)}if(FS.isDir(stat.mode)){check.push(...FS.readdir(path).filter(isRealDir).map(toAbsolute(path)))}entries[path]={timestamp:stat.mtime}}return callback(null,{type:"local",entries})},getRemoteSet:(mount,callback)=>{var entries={};IDBFS.getDB(mount.mountpoint,(err,db)=>{if(err)return callback(err);try{var transaction=db.transaction([IDBFS.DB_STORE_NAME],"readonly");transaction.onerror=e=>{callback(e.target.error);e.preventDefault()};var store=transaction.objectStore(IDBFS.DB_STORE_NAME);var index=store.index("timestamp");index.openKeyCursor().onsuccess=event=>{var cursor=event.target.result;if(!cursor){return callback(null,{type:"remote",db,entries})}entries[cursor.primaryKey]={timestamp:cursor.key};cursor.continue()}}catch(e){return callback(e)}})},loadLocalEntry:(path,callback)=>{var stat,node;try{var lookup=FS.lookupPath(path);node=lookup.node;stat=FS.stat(path)}catch(e){return callback(e)}if(FS.isDir(stat.mode)){return callback(null,{timestamp:stat.mtime,mode:stat.mode})}else if(FS.isFile(stat.mode)){node.contents=MEMFS.getFileDataAsTypedArray(node);return callback(null,{timestamp:stat.mtime,mode:stat.mode,contents:node.contents})}else{return callback(new Error("node type not supported"))}},storeLocalEntry:(path,entry,callback)=>{try{if(FS.isDir(entry["mode"])){FS.mkdirTree(path,entry["mode"])}else if(FS.isFile(entry["mode"])){FS.writeFile(path,entry["contents"],{canOwn:true})}else{return callback(new Error("node type not supported"))}FS.chmod(path,entry["mode"]);FS.utime(path,entry["timestamp"],entry["timestamp"])}catch(e){return callback(e)}callback(null)},removeLocalEntry:(path,callback)=>{try{var stat=FS.stat(path);if(FS.isDir(stat.mode)){FS.rmdir(path)}else if(FS.isFile(stat.mode)){FS.unlink(path)}}catch(e){return callback(e)}callback(null)},loadRemoteEntry:(store,path,callback)=>{var req=store.get(path);req.onsuccess=event=>callback(null,event.target.result);req.onerror=e=>{callback(e.target.error);e.preventDefault()}},storeRemoteEntry:(store,path,entry,callback)=>{try{var req=store.put(entry,path)}catch(e){callback(e);return}req.onsuccess=event=>callback();req.onerror=e=>{callback(e.target.error);e.preventDefault()}},removeRemoteEntry:(store,path,callback)=>{var req=store.delete(path);req.onsuccess=event=>callback();req.onerror=e=>{callback(e.target.error);e.preventDefault()}},reconcile:(src,dst,callback)=>{var total=0;var create=[];Object.keys(src.entries).forEach(key=>{var e=src.entries[key];var e2=dst.entries[key];if(!e2||e["timestamp"].getTime()!=e2["timestamp"].getTime()){create.push(key);total++}});var remove=[];Object.keys(dst.entries).forEach(key=>{if(!src.entries[key]){remove.push(key);total++}});if(!total){return callback(null)}var errored=false;var db=src.type==="remote"?src.db:dst.db;var transaction=db.transaction([IDBFS.DB_STORE_NAME],"readwrite");var store=transaction.objectStore(IDBFS.DB_STORE_NAME);function done(err){if(err&&!errored){errored=true;return callback(err)}}transaction.onerror=transaction.onabort=e=>{done(e.target.error);e.preventDefault()};transaction.oncomplete=e=>{if(!errored){callback(null)}};create.sort().forEach(path=>{if(dst.type==="local"){IDBFS.loadRemoteEntry(store,path,(err,entry)=>{if(err)return done(err);IDBFS.storeLocalEntry(path,entry,done)})}else{IDBFS.loadLocalEntry(path,(err,entry)=>{if(err)return done(err);IDBFS.storeRemoteEntry(store,path,entry,done)})}});remove.sort().reverse().forEach(path=>{if(dst.type==="local"){IDBFS.removeLocalEntry(path,done)}else{IDBFS.removeRemoteEntry(store,path,done)}})}};var ERRNO_CODES={EPERM:63,ENOENT:44,ESRCH:71,EINTR:27,EIO:29,ENXIO:60,E2BIG:1,ENOEXEC:45,EBADF:8,ECHILD:12,EAGAIN:6,EWOULDBLOCK:6,ENOMEM:48,EACCES:2,EFAULT:21,ENOTBLK:105,EBUSY:10,EEXIST:20,EXDEV:75,ENODEV:43,ENOTDIR:54,EISDIR:31,EINVAL:28,ENFILE:41,EMFILE:33,ENOTTY:59,ETXTBSY:74,EFBIG:22,ENOSPC:51,ESPIPE:70,EROFS:69,EMLINK:34,EPIPE:64,EDOM:18,ERANGE:68,ENOMSG:49,EIDRM:24,ECHRNG:106,EL2NSYNC:156,EL3HLT:107,EL3RST:108,ELNRNG:109,EUNATCH:110,ENOCSI:111,EL2HLT:112,EDEADLK:16,ENOLCK:46,EBADE:113,EBADR:114,EXFULL:115,ENOANO:104,EBADRQC:103,EBADSLT:102,EDEADLOCK:16,EBFONT:101,ENOSTR:100,ENODATA:116,ETIME:117,ENOSR:118,ENONET:119,ENOPKG:120,EREMOTE:121,ENOLINK:47,EADV:122,ESRMNT:123,ECOMM:124,EPROTO:65,EMULTIHOP:36,EDOTDOT:125,EBADMSG:9,ENOTUNIQ:126,EBADFD:127,EREMCHG:128,ELIBACC:129,ELIBBAD:130,ELIBSCN:131,ELIBMAX:132,ELIBEXEC:133,ENOSYS:52,ENOTEMPTY:55,ENAMETOOLONG:37,ELOOP:32,EOPNOTSUPP:138,EPFNOSUPPORT:139,ECONNRESET:15,ENOBUFS:42,EAFNOSUPPORT:5,EPROTOTYPE:67,ENOTSOCK:57,ENOPROTOOPT:50,ESHUTDOWN:140,ECONNREFUSED:14,EADDRINUSE:3,ECONNABORTED:13,ENETUNREACH:40,ENETDOWN:38,ETIMEDOUT:73,EHOSTDOWN:142,EHOSTUNREACH:23,EINPROGRESS:26,EALREADY:7,EDESTADDRREQ:17,EMSGSIZE:35,EPROTONOSUPPORT:66,ESOCKTNOSUPPORT:137,EADDRNOTAVAIL:4,ENETRESET:39,EISCONN:30,ENOTCONN:53,ETOOMANYREFS:141,EUSERS:136,EDQUOT:19,ESTALE:72,ENOTSUP:138,ENOMEDIUM:148,EILSEQ:25,EOVERFLOW:61,ECANCELED:11,ENOTRECOVERABLE:56,EOWNERDEAD:62,ESTRPIPE:135};var NODEFS={isWindows:false,staticInit(){NODEFS.isWindows=!!process.platform.match(/^win/);var flags=process.binding("constants")["fs"];NODEFS.flagsForNodeMap={1024:flags["O_APPEND"],64:flags["O_CREAT"],128:flags["O_EXCL"],256:flags["O_NOCTTY"],0:flags["O_RDONLY"],2:flags["O_RDWR"],4096:flags["O_SYNC"],512:flags["O_TRUNC"],1:flags["O_WRONLY"],131072:flags["O_NOFOLLOW"]}},convertNodeCode(e){var code=e.code;return ERRNO_CODES[code]},tryFSOperation(f){try{return f()}catch(e){if(!e.code)throw e;if(e.code==="UNKNOWN")throw new FS.ErrnoError(28);throw new FS.ErrnoError(NODEFS.convertNodeCode(e))}},mount(mount){return NODEFS.createNode(null,"/",NODEFS.getMode(mount.opts.root),0)},createNode(parent,name,mode,dev){if(!FS.isDir(mode)&&!FS.isFile(mode)&&!FS.isLink(mode)){throw new FS.ErrnoError(28)}var node=FS.createNode(parent,name,mode);node.node_ops=NODEFS.node_ops;node.stream_ops=NODEFS.stream_ops;return node},getMode(path){return NODEFS.tryFSOperation(()=>{var mode=fs.lstatSync(path).mode;if(NODEFS.isWindows){mode|=(mode&292)>>2}return mode})},realPath(node){var parts=[];while(node.parent!==node){parts.push(node.name);node=node.parent}parts.push(node.mount.opts.root);parts.reverse();return PATH.join(...parts)},flagsForNode(flags){flags&=~2097152;flags&=~2048;flags&=~32768;flags&=~524288;flags&=~65536;var newFlags=0;for(var k in NODEFS.flagsForNodeMap){if(flags&k){newFlags|=NODEFS.flagsForNodeMap[k];flags^=k}}if(flags){throw new FS.ErrnoError(28)}return newFlags},getattr(func,node){var stat=NODEFS.tryFSOperation(func);if(NODEFS.isWindows){if(!stat.blksize){stat.blksize=4096}if(!stat.blocks){stat.blocks=(stat.size+stat.blksize-1)/stat.blksize|0}stat.mode|=(stat.mode&292)>>2}return{dev:stat.dev,ino:node.id,mode:stat.mode,nlink:stat.nlink,uid:stat.uid,gid:stat.gid,rdev:stat.rdev,size:stat.size,atime:stat.atime,mtime:stat.mtime,ctime:stat.ctime,blksize:stat.blksize,blocks:stat.blocks}},setattr(arg,node,attr,chmod,utimes,truncate,stat){NODEFS.tryFSOperation(()=>{if(attr.mode!==undefined){var mode=attr.mode;if(NODEFS.isWindows){mode&=384}chmod(arg,mode);node.mode=attr.mode}if(typeof(attr.atime??attr.mtime)==="number"){var atime=new Date(attr.atime??stat(arg).atime);var mtime=new Date(attr.mtime??stat(arg).mtime);utimes(arg,atime,mtime)}if(attr.size!==undefined){truncate(arg,attr.size)}})},node_ops:{getattr(node){var path=NODEFS.realPath(node);return NODEFS.getattr(()=>fs.lstatSync(path),node)},setattr(node,attr){var path=NODEFS.realPath(node);if(attr.mode!=null&&attr.dontFollow){throw new FS.ErrnoError(52)}NODEFS.setattr(path,node,attr,fs.chmodSync,fs.utimesSync,fs.truncateSync,fs.lstatSync)},lookup(parent,name){var path=PATH.join2(NODEFS.realPath(parent),name);var mode=NODEFS.getMode(path);return NODEFS.createNode(parent,name,mode)},mknod(parent,name,mode,dev){var node=NODEFS.createNode(parent,name,mode,dev);var path=NODEFS.realPath(node);NODEFS.tryFSOperation(()=>{if(FS.isDir(node.mode)){fs.mkdirSync(path,node.mode)}else{fs.writeFileSync(path,"",{mode:node.mode})}});return node},rename(oldNode,newDir,newName){var oldPath=NODEFS.realPath(oldNode);var newPath=PATH.join2(NODEFS.realPath(newDir),newName);try{FS.unlink(newPath)}catch(e){}NODEFS.tryFSOperation(()=>fs.renameSync(oldPath,newPath));oldNode.name=newName},unlink(parent,name){var path=PATH.join2(NODEFS.realPath(parent),name);NODEFS.tryFSOperation(()=>fs.unlinkSync(path))},rmdir(parent,name){var path=PATH.join2(NODEFS.realPath(parent),name);NODEFS.tryFSOperation(()=>fs.rmdirSync(path))},readdir(node){var path=NODEFS.realPath(node);return NODEFS.tryFSOperation(()=>fs.readdirSync(path))},symlink(parent,newName,oldPath){var newPath=PATH.join2(NODEFS.realPath(parent),newName);NODEFS.tryFSOperation(()=>fs.symlinkSync(oldPath,newPath))},readlink(node){var path=NODEFS.realPath(node);return NODEFS.tryFSOperation(()=>fs.readlinkSync(path))},statfs(path){var stats=NODEFS.tryFSOperation(()=>fs.statfsSync(path));stats.frsize=stats.bsize;return stats}},stream_ops:{getattr(stream){return NODEFS.getattr(()=>fs.fstatSync(stream.nfd),stream.node)},setattr(stream,attr){NODEFS.setattr(stream.nfd,stream.node,attr,fs.fchmodSync,fs.futimesSync,fs.ftruncateSync,fs.fstatSync)},open(stream){var path=NODEFS.realPath(stream.node);NODEFS.tryFSOperation(()=>{stream.shared.refcount=1;stream.nfd=fs.openSync(path,NODEFS.flagsForNode(stream.flags))})},close(stream){NODEFS.tryFSOperation(()=>{if(stream.nfd&&--stream.shared.refcount===0){fs.closeSync(stream.nfd)}})},dup(stream){stream.shared.refcount++},read(stream,buffer,offset,length,position){return NODEFS.tryFSOperation(()=>fs.readSync(stream.nfd,new Int8Array(buffer.buffer,offset,length),0,length,position))},write(stream,buffer,offset,length,position){return NODEFS.tryFSOperation(()=>fs.writeSync(stream.nfd,new Int8Array(buffer.buffer,offset,length),0,length,position))},llseek(stream,offset,whence){var position=offset;if(whence===1){position+=stream.position}else if(whence===2){if(FS.isFile(stream.node.mode)){NODEFS.tryFSOperation(()=>{var stat=fs.fstatSync(stream.nfd);position+=stat.size})}}if(position<0){throw new FS.ErrnoError(28)}return position},mmap(stream,length,position,prot,flags){if(!FS.isFile(stream.node.mode)){throw new FS.ErrnoError(43)}var ptr=mmapAlloc(length);NODEFS.stream_ops.read(stream,HEAP8,ptr,length,position);return{ptr,allocated:true}},msync(stream,buffer,offset,length,mmapFlags){NODEFS.stream_ops.write(stream,buffer,0,length,offset,false);return 0}}};var WORKERFS={DIR_MODE:16895,FILE_MODE:33279,reader:null,mount(mount){assert(ENVIRONMENT_IS_WORKER);WORKERFS.reader??=new FileReaderSync;var root=WORKERFS.createNode(null,"/",WORKERFS.DIR_MODE,0);var createdParents={};function ensureParent(path){var parts=path.split("/");var parent=root;for(var i=0;i<parts.length-1;i++){var curr=parts.slice(0,i+1).join("/");createdParents[curr]||=WORKERFS.createNode(parent,parts[i],WORKERFS.DIR_MODE,0);parent=createdParents[curr]}return parent}function base(path){var parts=path.split("/");return parts[parts.length-1]}Array.prototype.forEach.call(mount.opts["files"]||[],function(file){WORKERFS.createNode(ensureParent(file.name),base(file.name),WORKERFS.FILE_MODE,0,file,file.lastModifiedDate)});(mount.opts["blobs"]||[]).forEach(obj=>{WORKERFS.createNode(ensureParent(obj["name"]),base(obj["name"]),WORKERFS.FILE_MODE,0,obj["data"])});(mount.opts["packages"]||[]).forEach(pack=>{pack["metadata"].files.forEach(file=>{var name=file.filename.slice(1);WORKERFS.createNode(ensureParent(name),base(name),WORKERFS.FILE_MODE,0,pack["blob"].slice(file.start,file.end))})});return root},createNode(parent,name,mode,dev,contents,mtime){var node=FS.createNode(parent,name,mode);node.mode=mode;node.node_ops=WORKERFS.node_ops;node.stream_ops=WORKERFS.stream_ops;node.atime=node.mtime=node.ctime=(mtime||new Date).getTime();assert(WORKERFS.FILE_MODE!==WORKERFS.DIR_MODE);if(mode===WORKERFS.FILE_MODE){node.size=contents.size;node.contents=contents}else{node.size=4096;node.contents={}}if(parent){parent.contents[name]=node}return node},node_ops:{getattr(node){return{dev:1,ino:node.id,mode:node.mode,nlink:1,uid:0,gid:0,rdev:0,size:node.size,atime:new Date(node.atime),mtime:new Date(node.mtime),ctime:new Date(node.ctime),blksize:4096,blocks:Math.ceil(node.size/4096)}},setattr(node,attr){for(const key of["mode","atime","mtime","ctime"]){if(attr[key]!=null){node[key]=attr[key]}}},lookup(parent,name){throw new FS.ErrnoError(44)},mknod(parent,name,mode,dev){throw new FS.ErrnoError(63)},rename(oldNode,newDir,newName){throw new FS.ErrnoError(63)},unlink(parent,name){throw new FS.ErrnoError(63)},rmdir(parent,name){throw new FS.ErrnoError(63)},readdir(node){var entries=[".",".."];for(var key of Object.keys(node.contents)){entries.push(key)}return entries},symlink(parent,newName,oldPath){throw new FS.ErrnoError(63)}},stream_ops:{read(stream,buffer,offset,length,position){if(position>=stream.node.size)return 0;var chunk=stream.node.contents.slice(position,position+length);var ab=WORKERFS.reader.readAsArrayBuffer(chunk);buffer.set(new Uint8Array(ab),offset);return chunk.size},write(stream,buffer,offset,length,position){throw new FS.ErrnoError(29)},llseek(stream,offset,whence){var position=offset;if(whence===1){position+=stream.position}else if(whence===2){if(FS.isFile(stream.node.mode)){position+=stream.node.size}}if(position<0){throw new FS.ErrnoError(28)}return position}}};var PROXYFS={mount(mount){return PROXYFS.createNode(null,"/",mount.opts.fs.lstat(mount.opts.root).mode,0)},createNode(parent,name,mode,dev){if(!FS.isDir(mode)&&!FS.isFile(mode)&&!FS.isLink(mode)){throw new FS.ErrnoError(ERRNO_CODES.EINVAL)}var node=FS.createNode(parent,name,mode);node.node_ops=PROXYFS.node_ops;node.stream_ops=PROXYFS.stream_ops;return node},realPath(node){var parts=[];while(node.parent!==node){parts.push(node.name);node=node.parent}parts.push(node.mount.opts.root);parts.reverse();return PATH.join(...parts)},node_ops:{getattr(node){var path=PROXYFS.realPath(node);var stat;try{stat=node.mount.opts.fs.lstat(path)}catch(e){if(!e.code)throw e;throw new FS.ErrnoError(ERRNO_CODES[e.code])}return{dev:stat.dev,ino:stat.ino,mode:stat.mode,nlink:stat.nlink,uid:stat.uid,gid:stat.gid,rdev:stat.rdev,size:stat.size,atime:stat.atime,mtime:stat.mtime,ctime:stat.ctime,blksize:stat.blksize,blocks:stat.blocks}},setattr(node,attr){var path=PROXYFS.realPath(node);try{if(attr.mode!==undefined){node.mount.opts.fs.chmod(path,attr.mode);node.mode=attr.mode}if(attr.atime||attr.mtime){var atime=new Date(attr.atime||attr.mtime);var mtime=new Date(attr.mtime||attr.atime);node.mount.opts.fs.utime(path,atime,mtime)}if(attr.size!==undefined){node.mount.opts.fs.truncate(path,attr.size)}}catch(e){if(!e.code)throw e;throw new FS.ErrnoError(ERRNO_CODES[e.code])}},lookup(parent,name){try{var path=PATH.join2(PROXYFS.realPath(parent),name);var mode=parent.mount.opts.fs.lstat(path).mode;var node=PROXYFS.createNode(parent,name,mode);return node}catch(e){if(!e.code)throw e;throw new FS.ErrnoError(ERRNO_CODES[e.code])}},mknod(parent,name,mode,dev){var node=PROXYFS.createNode(parent,name,mode,dev);var path=PROXYFS.realPath(node);try{if(FS.isDir(node.mode)){node.mount.opts.fs.mkdir(path,node.mode)}else{node.mount.opts.fs.writeFile(path,"",{mode:node.mode})}}catch(e){if(!e.code)throw e;throw new FS.ErrnoError(ERRNO_CODES[e.code])}return node},rename(oldNode,newDir,newName){var oldPath=PROXYFS.realPath(oldNode);var newPath=PATH.join2(PROXYFS.realPath(newDir),newName);try{oldNode.mount.opts.fs.rename(oldPath,newPath);oldNode.name=newName}catch(e){if(!e.code)throw e;throw new FS.ErrnoError(ERRNO_CODES[e.code])}},unlink(parent,name){var path=PATH.join2(PROXYFS.realPath(parent),name);try{parent.mount.opts.fs.unlink(path)}catch(e){if(!e.code)throw e;throw new FS.ErrnoError(ERRNO_CODES[e.code])}},rmdir(parent,name){var path=PATH.join2(PROXYFS.realPath(parent),name);try{parent.mount.opts.fs.rmdir(path)}catch(e){if(!e.code)throw e;throw new FS.ErrnoError(ERRNO_CODES[e.code])}},readdir(node){var path=PROXYFS.realPath(node);try{return node.mount.opts.fs.readdir(path)}catch(e){if(!e.code)throw e;throw new FS.ErrnoError(ERRNO_CODES[e.code])}},symlink(parent,newName,oldPath){var newPath=PATH.join2(PROXYFS.realPath(parent),newName);try{parent.mount.opts.fs.symlink(oldPath,newPath)}catch(e){if(!e.code)throw e;throw new FS.ErrnoError(ERRNO_CODES[e.code])}},readlink(node){var path=PROXYFS.realPath(node);try{return node.mount.opts.fs.readlink(path)}catch(e){if(!e.code)throw e;throw new FS.ErrnoError(ERRNO_CODES[e.code])}}},stream_ops:{open(stream){var path=PROXYFS.realPath(stream.node);try{stream.nfd=stream.node.mount.opts.fs.open(path,stream.flags)}catch(e){if(!e.code)throw e;throw new FS.ErrnoError(ERRNO_CODES[e.code])}},close(stream){try{stream.node.mount.opts.fs.close(stream.nfd)}catch(e){if(!e.code)throw e;throw new FS.ErrnoError(ERRNO_CODES[e.code])}},read(stream,buffer,offset,length,position){try{return stream.node.mount.opts.fs.read(stream.nfd,buffer,offset,length,position)}catch(e){if(!e.code)throw e;throw new FS.ErrnoError(ERRNO_CODES[e.code])}},write(stream,buffer,offset,length,position){try{return stream.node.mount.opts.fs.write(stream.nfd,buffer,offset,length,position)}catch(e){if(!e.code)throw e;throw new FS.ErrnoError(ERRNO_CODES[e.code])}},llseek(stream,offset,whence){var position=offset;if(whence===1){position+=stream.position}else if(whence===2){if(FS.isFile(stream.node.mode)){try{var stat=stream.node.node_ops.getattr(stream.node);position+=stat.size}catch(e){throw new FS.ErrnoError(ERRNO_CODES[e.code])}}}if(position<0){throw new FS.ErrnoError(ERRNO_CODES.EINVAL)}return position}}};var LZ4={DIR_MODE:16895,FILE_MODE:33279,CHUNK_SIZE:-1,codec:null,init(){if(LZ4.codec)return;LZ4.codec=(()=>{var MiniLZ4=function(){var exports={};exports.uncompress=function(input,output,sIdx,eIdx){sIdx=sIdx||0;eIdx=eIdx||input.length-sIdx;for(var i=sIdx,n=eIdx,j=0;i<n;){var token=input[i++];var literals_length=token>>4;if(literals_length>0){var l=literals_length+240;while(l===255){l=input[i++];literals_length+=l}var end=i+literals_length;while(i<end)output[j++]=input[i++];if(i===n)return j}var offset=input[i++]|input[i++]<<8;if(offset===0)return j;if(offset>j)return-(i-2);var match_length=token&15;var l=match_length+240;while(l===255){l=input[i++];match_length+=l}var pos=j-offset;var end=j+match_length+4;while(j<end)output[j++]=output[pos++]}return j};var maxInputSize=2113929216,minMatch=4,hashLog=16,hashShift=minMatch*8-hashLog,hashSize=1<<hashLog,copyLength=8,lastLiterals=5,mfLimit=copyLength+minMatch,skipStrength=6,mlBits=4,mlMask=(1<<mlBits)-1,runBits=8-mlBits,runMask=(1<<runBits)-1,hasher=2654435761;assert(hashShift===16);var hashTable=new Int16Array(1<<16);var empty=new Int16Array(hashTable.length);exports.compressBound=function(isize){return isize>maxInputSize?0:isize+isize/255+16|0};exports.compress=function(src,dst,sIdx,eIdx){hashTable.set(empty);return compressBlock(src,dst,0,sIdx||0,eIdx||dst.length)};function compressBlock(src,dst,pos,sIdx,eIdx){var dpos=sIdx;var dlen=eIdx-sIdx;var anchor=0;if(src.length>=maxInputSize)throw new Error("input too large");if(src.length>mfLimit){var n=exports.compressBound(src.length);if(dlen<n)throw Error("output too small: "+dlen+" < "+n);var step=1,findMatchAttempts=(1<<skipStrength)+3,srcLength=src.length-mfLimit;while(pos+minMatch<srcLength){var sequenceLowBits=src[pos+1]<<8|src[pos];var sequenceHighBits=src[pos+3]<<8|src[pos+2];var hash=Math.imul(sequenceLowBits|sequenceHighBits<<16,hasher)>>>hashShift;var ref=hashTable[hash]-1;hashTable[hash]=pos+1;if(ref<0||pos-ref>>>16>0||((src[ref+3]<<8|src[ref+2])!=sequenceHighBits||(src[ref+1]<<8|src[ref])!=sequenceLowBits)){step=findMatchAttempts++>>skipStrength;pos+=step;continue}findMatchAttempts=(1<<skipStrength)+3;var literals_length=pos-anchor;var offset=pos-ref;pos+=minMatch;ref+=minMatch;var match_length=pos;while(pos<srcLength&&src[pos]==src[ref]){pos++;ref++}match_length=pos-match_length;var token=match_length<mlMask?match_length:mlMask;if(literals_length>=runMask){dst[dpos++]=(runMask<<mlBits)+token;for(var len=literals_length-runMask;len>254;len-=255){dst[dpos++]=255}dst[dpos++]=len}else{dst[dpos++]=(literals_length<<mlBits)+token}for(var i=0;i<literals_length;i++){dst[dpos++]=src[anchor+i]}dst[dpos++]=offset;dst[dpos++]=offset>>8;if(match_length>=mlMask){match_length-=mlMask;while(match_length>=255){match_length-=255;dst[dpos++]=255}dst[dpos++]=match_length}anchor=pos}}if(anchor==0)return 0;literals_length=src.length-anchor;if(literals_length>=runMask){dst[dpos++]=runMask<<mlBits;for(var ln=literals_length-runMask;ln>254;ln-=255){dst[dpos++]=255}dst[dpos++]=ln}else{dst[dpos++]=literals_length<<mlBits}pos=anchor;while(pos<src.length){dst[dpos++]=src[pos++]}return dpos}exports.CHUNK_SIZE=2048;exports.compressPackage=function(data,verify){if(verify){var temp=new Uint8Array(exports.CHUNK_SIZE)}assert(data instanceof ArrayBuffer);data=new Uint8Array(data);console.log("compressing package of size "+data.length);var compressedChunks=[];var successes=[];var offset=0;var total=0;while(offset<data.length){var chunk=data.subarray(offset,offset+exports.CHUNK_SIZE);offset+=exports.CHUNK_SIZE;var bound=exports.compressBound(chunk.length);var compressed=new Uint8Array(bound);var compressedSize=exports.compress(chunk,compressed);if(compressedSize>0){assert(compressedSize<=bound);compressed=compressed.subarray(0,compressedSize);compressedChunks.push(compressed);total+=compressedSize;successes.push(1);if(verify){var back=exports.uncompress(compressed,temp);assert(back===chunk.length,[back,chunk.length]);for(var i=0;i<chunk.length;i++){assert(chunk[i]===temp[i])}}}else{assert(compressedSize===0);compressedChunks.push(chunk);total+=chunk.length;successes.push(0)}}data=null;var compressedData={data:new Uint8Array(total+exports.CHUNK_SIZE*2),cachedOffset:total,cachedIndexes:[-1,-1],cachedChunks:[null,null],offsets:[],sizes:[],successes};offset=0;for(var i=0;i<compressedChunks.length;i++){compressedData["data"].set(compressedChunks[i],offset);compressedData["offsets"][i]=offset;compressedData["sizes"][i]=compressedChunks[i].length;offset+=compressedChunks[i].length}console.log("compressed package into "+[compressedData["data"].length]);assert(offset===total);return compressedData};assert(exports.CHUNK_SIZE<1<<15);return exports}();if(typeof module!="undefined"){module.exports=MiniLZ4}return MiniLZ4})();LZ4.CHUNK_SIZE=LZ4.codec.CHUNK_SIZE},loadPackage(pack,preloadPlugin){LZ4.init();var compressedData=pack["compressedData"]||LZ4.codec.compressPackage(pack["data"]);assert(compressedData["cachedIndexes"].length===compressedData["cachedChunks"].length);for(var i=0;i<compressedData["cachedIndexes"].length;i++){compressedData["cachedIndexes"][i]=-1;compressedData["cachedChunks"][i]=compressedData["data"].subarray(compressedData["cachedOffset"]+i*LZ4.CHUNK_SIZE,compressedData["cachedOffset"]+(i+1)*LZ4.CHUNK_SIZE);assert(compressedData["cachedChunks"][i].length===LZ4.CHUNK_SIZE)}pack["metadata"].files.forEach(file=>{var dir=PATH.dirname(file.filename);var name=PATH.basename(file.filename);FS.createPath("",dir,true,true);var parent=FS.analyzePath(dir).object;LZ4.createNode(parent,name,LZ4.FILE_MODE,0,{compressedData,start:file.start,end:file.end})});if(preloadPlugin){Browser.init();pack["metadata"].files.forEach(file=>{var handled=false;var fullname=file.filename;preloadPlugins.forEach(plugin=>{if(handled)return;if(plugin["canHandle"](fullname)){var dep=getUniqueRunDependency("fp "+fullname);addRunDependency(dep);var finish=()=>removeRunDependency(dep);var byteArray=FS.readFile(fullname);plugin["handle"](byteArray,fullname,finish,finish);handled=true}})})}},createNode(parent,name,mode,dev,contents,mtime){var node=FS.createNode(parent,name,mode);node.mode=mode;node.node_ops=LZ4.node_ops;node.stream_ops=LZ4.stream_ops;this.atime=this.mtime=this.ctime=(mtime||new Date).getTime();assert(LZ4.FILE_MODE!==LZ4.DIR_MODE);if(mode===LZ4.FILE_MODE){node.size=contents.end-contents.start;node.contents=contents}else{node.size=4096;node.contents={}}if(parent){parent.contents[name]=node}return node},node_ops:{getattr(node){return{dev:1,ino:node.id,mode:node.mode,nlink:1,uid:0,gid:0,rdev:0,size:node.size,atime:new Date(node.atime),mtime:new Date(node.mtime),ctime:new Date(node.ctime),blksize:4096,blocks:Math.ceil(node.size/4096)}},setattr(node,attr){for(const key of["mode","atime","mtime","ctime"]){if(attr[key]){node[key]=attr[key]}}},lookup(parent,name){throw new FS.ErrnoError(44)},mknod(parent,name,mode,dev){throw new FS.ErrnoError(63)},rename(oldNode,newDir,newName){throw new FS.ErrnoError(63)},unlink(parent,name){throw new FS.ErrnoError(63)},rmdir(parent,name){throw new FS.ErrnoError(63)},readdir(node){throw new FS.ErrnoError(63)},symlink(parent,newName,oldPath){throw new FS.ErrnoError(63)}},stream_ops:{read(stream,buffer,offset,length,position){length=Math.min(length,stream.node.size-position);if(length<=0)return 0;var contents=stream.node.contents;var compressedData=contents.compressedData;var written=0;while(written<length){var start=contents.start+position+written;var desired=length-written;var chunkIndex=Math.floor(start/LZ4.CHUNK_SIZE);var compressedStart=compressedData["offsets"][chunkIndex];var compressedSize=compressedData["sizes"][chunkIndex];var currChunk;if(compressedData["successes"][chunkIndex]){var found=compressedData["cachedIndexes"].indexOf(chunkIndex);if(found>=0){currChunk=compressedData["cachedChunks"][found]}else{compressedData["cachedIndexes"].pop();compressedData["cachedIndexes"].unshift(chunkIndex);currChunk=compressedData["cachedChunks"].pop();compressedData["cachedChunks"].unshift(currChunk);if(compressedData["debug"]){out("decompressing chunk "+chunkIndex);Module["decompressedChunks"]=(Module["decompressedChunks"]||0)+1}var compressed=compressedData["data"].subarray(compressedStart,compressedStart+compressedSize);var originalSize=LZ4.codec.uncompress(compressed,currChunk);if(chunkIndex<compressedData["successes"].length-1)assert(originalSize===LZ4.CHUNK_SIZE)}}else{currChunk=compressedData["data"].subarray(compressedStart,compressedStart+LZ4.CHUNK_SIZE)}var startInChunk=start%LZ4.CHUNK_SIZE;var endInChunk=Math.min(startInChunk+desired,LZ4.CHUNK_SIZE);buffer.set(currChunk.subarray(startInChunk,endInChunk),offset+written);var currWritten=endInChunk-startInChunk;written+=currWritten}return written},write(stream,buffer,offset,length,position){throw new FS.ErrnoError(29)},llseek(stream,offset,whence){var position=offset;if(whence===1){position+=stream.position}else if(whence===2){if(FS.isFile(stream.node.mode)){position+=stream.node.size}}if(position<0){throw new FS.ErrnoError(28)}return position}}};var FS={root:null,mounts:[],devices:{},streams:[],nextInode:1,nameTable:null,currentPath:"/",initialized:false,ignorePermissions:true,trackingDelegate:{},filesystems:null,syncFSRequests:0,readFiles:{},ErrnoError:class{name="ErrnoError";constructor(errno){this.errno=errno}},FSStream:class{shared={};get object(){return this.node}set object(val){this.node=val}get isRead(){return(this.flags&2097155)!==1}get isWrite(){return(this.flags&2097155)!==0}get isAppend(){return this.flags&1024}get flags(){return this.shared.flags}set flags(val){this.shared.flags=val}get position(){return this.shared.position}set position(val){this.shared.position=val}},FSNode:class{node_ops={};stream_ops={};readMode=292|73;writeMode=146;mounted=null;constructor(parent,name,mode,rdev){if(!parent){parent=this}this.parent=parent;this.mount=parent.mount;this.id=FS.nextInode++;this.name=name;this.mode=mode;this.rdev=rdev;this.atime=this.mtime=this.ctime=Date.now()}get read(){return(this.mode&this.readMode)===this.readMode}set read(val){val?this.mode|=this.readMode:this.mode&=~this.readMode}get write(){return(this.mode&this.writeMode)===this.writeMode}set write(val){val?this.mode|=this.writeMode:this.mode&=~this.writeMode}get isFolder(){return FS.isDir(this.mode)}get isDevice(){return FS.isChrdev(this.mode)}},lookupPath(path,opts={}){if(!path){throw new FS.ErrnoError(44)}opts.follow_mount??=true;if(!PATH.isAbs(path)){path=FS.cwd()+"/"+path}linkloop:for(var nlinks=0;nlinks<40;nlinks++){var parts=path.split("/").filter(p=>!!p);var current=FS.root;var current_path="/";for(var i=0;i<parts.length;i++){var islast=i===parts.length-1;if(islast&&opts.parent){break}if(parts[i]==="."){continue}if(parts[i]===".."){current_path=PATH.dirname(current_path);if(FS.isRoot(current)){path=current_path+"/"+parts.slice(i+1).join("/");continue linkloop}else{current=current.parent}continue}current_path=PATH.join2(current_path,parts[i]);try{current=FS.lookupNode(current,parts[i])}catch(e){if(e?.errno===44&&islast&&opts.noent_okay){return{path:current_path}}throw e}if(FS.isMountpoint(current)&&(!islast||opts.follow_mount)){current=current.mounted.root}if(FS.isLink(current.mode)&&(!islast||opts.follow)){if(!current.node_ops.readlink){throw new FS.ErrnoError(52)}var link=current.node_ops.readlink(current);if(!PATH.isAbs(link)){link=PATH.dirname(current_path)+"/"+link}path=link+"/"+parts.slice(i+1).join("/");continue linkloop}}return{path:current_path,node:current}}throw new FS.ErrnoError(32)},getPath(node){var path;while(true){if(FS.isRoot(node)){var mount=node.mount.mountpoint;if(!path)return mount;return mount[mount.length-1]!=="/"?`${mount}/${path}`:mount+path}path=path?`${node.name}/${path}`:node.name;node=node.parent}},hashName(parentid,name){var hash=0;for(var i=0;i<name.length;i++){hash=(hash<<5)-hash+name.charCodeAt(i)|0}return(parentid+hash>>>0)%FS.nameTable.length},hashAddNode(node){var hash=FS.hashName(node.parent.id,node.name);node.name_next=FS.nameTable[hash];FS.nameTable[hash]=node},hashRemoveNode(node){var hash=FS.hashName(node.parent.id,node.name);if(FS.nameTable[hash]===node){FS.nameTable[hash]=node.name_next}else{var current=FS.nameTable[hash];while(current){if(current.name_next===node){current.name_next=node.name_next;break}current=current.name_next}}},lookupNode(parent,name){var errCode=FS.mayLookup(parent);if(errCode){throw new FS.ErrnoError(errCode)}var hash=FS.hashName(parent.id,name);for(var node=FS.nameTable[hash];node;node=node.name_next){var nodeName=node.name;if(node.parent.id===parent.id&&nodeName===name){return node}}return FS.lookup(parent,name)},createNode(parent,name,mode,rdev){var node=new FS.FSNode(parent,name,mode,rdev);FS.hashAddNode(node);return node},destroyNode(node){FS.hashRemoveNode(node)},isRoot(node){return node===node.parent},isMountpoint(node){return!!node.mounted},isFile(mode){return(mode&61440)===32768},isDir(mode){return(mode&61440)===16384},isLink(mode){return(mode&61440)===40960},isChrdev(mode){return(mode&61440)===8192},isBlkdev(mode){return(mode&61440)===24576},isFIFO(mode){return(mode&61440)===4096},isSocket(mode){return(mode&49152)===49152},flagsToPermissionString(flag){var perms=["r","w","rw"][flag&3];if(flag&512){perms+="w"}return perms},nodePermissions(node,perms){if(FS.ignorePermissions){return 0}if(perms.includes("r")&&!(node.mode&292)){return 2}else if(perms.includes("w")&&!(node.mode&146)){return 2}else if(perms.includes("x")&&!(node.mode&73)){return 2}return 0},mayLookup(dir){if(!FS.isDir(dir.mode))return 54;var errCode=FS.nodePermissions(dir,"x");if(errCode)return errCode;if(!dir.node_ops.lookup)return 2;return 0},mayCreate(dir,name){if(!FS.isDir(dir.mode)){return 54}try{var node=FS.lookupNode(dir,name);return 20}catch(e){}return FS.nodePermissions(dir,"wx")},mayDelete(dir,name,isdir){var node;try{node=FS.lookupNode(dir,name)}catch(e){return e.errno}var errCode=FS.nodePermissions(dir,"wx");if(errCode){return errCode}if(isdir){if(!FS.isDir(node.mode)){return 54}if(FS.isRoot(node)||FS.getPath(node)===FS.cwd()){return 10}}else{if(FS.isDir(node.mode)){return 31}}return 0},mayOpen(node,flags){if(!node){return 44}if(FS.isLink(node.mode)){return 32}else if(FS.isDir(node.mode)){if(FS.flagsToPermissionString(flags)!=="r"||flags&(512|64)){return 31}}return FS.nodePermissions(node,FS.flagsToPermissionString(flags))},checkOpExists(op,err){if(!op){throw new FS.ErrnoError(err)}return op},MAX_OPEN_FDS:4096,nextfd(){for(var fd=0;fd<=FS.MAX_OPEN_FDS;fd++){if(!FS.streams[fd]){return fd}}throw new FS.ErrnoError(33)},getStreamChecked(fd){var stream=FS.getStream(fd);if(!stream){throw new FS.ErrnoError(8)}return stream},getStream:fd=>FS.streams[fd],createStream(stream,fd=-1){stream=Object.assign(new FS.FSStream,stream);if(fd==-1){fd=FS.nextfd()}stream.fd=fd;FS.streams[fd]=stream;return stream},closeStream(fd){FS.streams[fd]=null},dupStream(origStream,fd=-1){var stream=FS.createStream(origStream,fd);stream.stream_ops?.dup?.(stream);return stream},doSetAttr(stream,node,attr){var setattr=stream?.stream_ops.setattr;var arg=setattr?stream:node;setattr??=node.node_ops.setattr;FS.checkOpExists(setattr,63);setattr(arg,attr)},chrdev_stream_ops:{open(stream){var device=FS.getDevice(stream.node.rdev);stream.stream_ops=device.stream_ops;stream.stream_ops.open?.(stream)},llseek(){throw new FS.ErrnoError(70)}},major:dev=>dev>>8,minor:dev=>dev&255,makedev:(ma,mi)=>ma<<8|mi,registerDevice(dev,ops){FS.devices[dev]={stream_ops:ops}},getDevice:dev=>FS.devices[dev],getMounts(mount){var mounts=[];var check=[mount];while(check.length){var m=check.pop();mounts.push(m);check.push(...m.mounts)}return mounts},syncfs(populate,callback){if(typeof populate=="function"){callback=populate;populate=false}FS.syncFSRequests++;if(FS.syncFSRequests>1){err(`warning: ${FS.syncFSRequests} FS.syncfs operations in flight at once, probably just doing extra work`)}var mounts=FS.getMounts(FS.root.mount);var completed=0;function doCallback(errCode){FS.syncFSRequests--;return callback(errCode)}function done(errCode){if(errCode){if(!done.errored){done.errored=true;return doCallback(errCode)}return}if(++completed>=mounts.length){doCallback(null)}}mounts.forEach(mount=>{if(!mount.type.syncfs){return done(null)}mount.type.syncfs(mount,populate,done)})},mount(type,opts,mountpoint){var root=mountpoint==="/";var pseudo=!mountpoint;var node;if(root&&FS.root){throw new FS.ErrnoError(10)}else if(!root&&!pseudo){var lookup=FS.lookupPath(mountpoint,{follow_mount:false});mountpoint=lookup.path;node=lookup.node;if(FS.isMountpoint(node)){throw new FS.ErrnoError(10)}if(!FS.isDir(node.mode)){throw new FS.ErrnoError(54)}}var mount={type,opts,mountpoint,mounts:[]};var mountRoot=type.mount(mount);mountRoot.mount=mount;mount.root=mountRoot;if(root){FS.root=mountRoot}else if(node){node.mounted=mount;if(node.mount){node.mount.mounts.push(mount)}}return mountRoot},unmount(mountpoint){var lookup=FS.lookupPath(mountpoint,{follow_mount:false});if(!FS.isMountpoint(lookup.node)){throw new FS.ErrnoError(28)}var node=lookup.node;var mount=node.mounted;var mounts=FS.getMounts(mount);Object.keys(FS.nameTable).forEach(hash=>{var current=FS.nameTable[hash];while(current){var next=current.name_next;if(mounts.includes(current.mount)){FS.destroyNode(current)}current=next}});node.mounted=null;var idx=node.mount.mounts.indexOf(mount);node.mount.mounts.splice(idx,1)},lookup(parent,name){return parent.node_ops.lookup(parent,name)},mknod(path,mode,dev){var lookup=FS.lookupPath(path,{parent:true});var parent=lookup.node;var name=PATH.basename(path);if(!name){throw new FS.ErrnoError(28)}if(name==="."||name===".."){throw new FS.ErrnoError(20)}var errCode=FS.mayCreate(parent,name);if(errCode){throw new FS.ErrnoError(errCode)}if(!parent.node_ops.mknod){throw new FS.ErrnoError(63)}return parent.node_ops.mknod(parent,name,mode,dev)},statfs(path){return FS.statfsNode(FS.lookupPath(path,{follow:true}).node)},statfsStream(stream){return FS.statfsNode(stream.node)},statfsNode(node){var rtn={bsize:4096,frsize:4096,blocks:1e6,bfree:5e5,bavail:5e5,files:FS.nextInode,ffree:FS.nextInode-1,fsid:42,flags:2,namelen:255};if(node.node_ops.statfs){Object.assign(rtn,node.node_ops.statfs(node.mount.opts.root))}return rtn},create(path,mode=438){mode&=4095;mode|=32768;return FS.mknod(path,mode,0)},mkdir(path,mode=511){mode&=511|512;mode|=16384;if(FS.trackingDelegate["onMakeDirectory"]){FS.trackingDelegate["onMakeDirectory"](path,mode)}return FS.mknod(path,mode,0)},mkdirTree(path,mode){var dirs=path.split("/");var d="";for(var dir of dirs){if(!dir)continue;if(d||PATH.isAbs(path))d+="/";d+=dir;try{FS.mkdir(d,mode)}catch(e){if(e.errno!=20)throw e}}},mkdev(path,mode,dev){if(typeof dev=="undefined"){dev=mode;mode=438}mode|=8192;return FS.mknod(path,mode,dev)},symlink(oldpath,newpath){if(!PATH_FS.resolve(oldpath)){throw new FS.ErrnoError(44)}var lookup=FS.lookupPath(newpath,{parent:true});var parent=lookup.node;if(!parent){throw new FS.ErrnoError(44)}var newname=PATH.basename(newpath);var errCode=FS.mayCreate(parent,newname);if(errCode){throw new FS.ErrnoError(errCode)}if(!parent.node_ops.symlink){throw new FS.ErrnoError(63)}if(FS.trackingDelegate["onMakeSymlink"]){FS.trackingDelegate["onMakeSymlink"](oldpath,newpath)}return parent.node_ops.symlink(parent,newname,oldpath)},rename(old_path,new_path){var old_dirname=PATH.dirname(old_path);var new_dirname=PATH.dirname(new_path);var old_name=PATH.basename(old_path);var new_name=PATH.basename(new_path);var lookup,old_dir,new_dir;lookup=FS.lookupPath(old_path,{parent:true});old_dir=lookup.node;lookup=FS.lookupPath(new_path,{parent:true});new_dir=lookup.node;if(!old_dir||!new_dir)throw new FS.ErrnoError(44);if(old_dir.mount!==new_dir.mount){throw new FS.ErrnoError(75)}var old_node=FS.lookupNode(old_dir,old_name);var relative=PATH_FS.relative(old_path,new_dirname);if(relative.charAt(0)!=="."){throw new FS.ErrnoError(28)}relative=PATH_FS.relative(new_path,old_dirname);if(relative.charAt(0)!=="."){throw new FS.ErrnoError(55)}var new_node;try{new_node=FS.lookupNode(new_dir,new_name)}catch(e){}if(old_node===new_node){return}var isdir=FS.isDir(old_node.mode);var errCode=FS.mayDelete(old_dir,old_name,isdir);if(errCode){throw new FS.ErrnoError(errCode)}errCode=new_node?FS.mayDelete(new_dir,new_name,isdir):FS.mayCreate(new_dir,new_name);if(errCode){throw new FS.ErrnoError(errCode)}if(!old_dir.node_ops.rename){throw new FS.ErrnoError(63)}if(FS.isMountpoint(old_node)||new_node&&FS.isMountpoint(new_node)){throw new FS.ErrnoError(10)}if(new_dir!==old_dir){errCode=FS.nodePermissions(old_dir,"w");if(errCode){throw new FS.ErrnoError(errCode)}}if(FS.trackingDelegate["willMovePath"]){FS.trackingDelegate["willMovePath"](old_path,new_path)}FS.hashRemoveNode(old_node);try{old_dir.node_ops.rename(old_node,new_dir,new_name);old_node.parent=new_dir}catch(e){throw e}finally{FS.hashAddNode(old_node)}if(FS.trackingDelegate["onMovePath"]){FS.trackingDelegate["onMovePath"](old_path,new_path)}},rmdir(path){var lookup=FS.lookupPath(path,{parent:true});var parent=lookup.node;var name=PATH.basename(path);var node=FS.lookupNode(parent,name);var errCode=FS.mayDelete(parent,name,true);if(errCode){throw new FS.ErrnoError(errCode)}if(!parent.node_ops.rmdir){throw new FS.ErrnoError(63)}if(FS.isMountpoint(node)){throw new FS.ErrnoError(10)}if(FS.trackingDelegate["willDeletePath"]){FS.trackingDelegate["willDeletePath"](path)}parent.node_ops.rmdir(parent,name);FS.destroyNode(node);if(FS.trackingDelegate["onDeletePath"]){FS.trackingDelegate["onDeletePath"](path)}},readdir(path){var lookup=FS.lookupPath(path,{follow:true});var node=lookup.node;var readdir=FS.checkOpExists(node.node_ops.readdir,54);return readdir(node)},unlink(path){var lookup=FS.lookupPath(path,{parent:true});var parent=lookup.node;if(!parent){throw new FS.ErrnoError(44)}var name=PATH.basename(path);var node=FS.lookupNode(parent,name);var errCode=FS.mayDelete(parent,name,false);if(errCode){throw new FS.ErrnoError(errCode)}if(!parent.node_ops.unlink){throw new FS.ErrnoError(63)}if(FS.isMountpoint(node)){throw new FS.ErrnoError(10)}if(FS.trackingDelegate["willDeletePath"]){FS.trackingDelegate["willDeletePath"](path)}parent.node_ops.unlink(parent,name);FS.destroyNode(node);if(FS.trackingDelegate["onDeletePath"]){FS.trackingDelegate["onDeletePath"](path)}},readlink(path){var lookup=FS.lookupPath(path);var link=lookup.node;if(!link){throw new FS.ErrnoError(44)}if(!link.node_ops.readlink){throw new FS.ErrnoError(28)}return link.node_ops.readlink(link)},stat(path,dontFollow){var lookup=FS.lookupPath(path,{follow:!dontFollow});var node=lookup.node;var getattr=FS.checkOpExists(node.node_ops.getattr,63);return getattr(node)},fstat(fd){var stream=FS.getStreamChecked(fd);var node=stream.node;var getattr=stream.stream_ops.getattr;var arg=getattr?stream:node;getattr??=node.node_ops.getattr;FS.checkOpExists(getattr,63);return getattr(arg)},lstat(path){return FS.stat(path,true)},doChmod(stream,node,mode,dontFollow){FS.doSetAttr(stream,node,{mode:mode&4095|node.mode&~4095,ctime:Date.now(),dontFollow})},chmod(path,mode,dontFollow){var node;if(typeof path=="string"){var lookup=FS.lookupPath(path,{follow:!dontFollow});node=lookup.node}else{node=path}FS.doChmod(null,node,mode,dontFollow)},lchmod(path,mode){FS.chmod(path,mode,true)},fchmod(fd,mode){var stream=FS.getStreamChecked(fd);FS.doChmod(stream,stream.node,mode,false)},doChown(stream,node,dontFollow){FS.doSetAttr(stream,node,{timestamp:Date.now(),dontFollow})},chown(path,uid,gid,dontFollow){var node;if(typeof path=="string"){var lookup=FS.lookupPath(path,{follow:!dontFollow});node=lookup.node}else{node=path}FS.doChown(null,node,dontFollow)},lchown(path,uid,gid){FS.chown(path,uid,gid,true)},fchown(fd,uid,gid){var stream=FS.getStreamChecked(fd);FS.doChown(stream,stream.node,false)},doTruncate(stream,node,len){if(FS.isDir(node.mode)){throw new FS.ErrnoError(31)}if(!FS.isFile(node.mode)){throw new FS.ErrnoError(28)}var errCode=FS.nodePermissions(node,"w");if(errCode){throw new FS.ErrnoError(errCode)}FS.doSetAttr(stream,node,{size:len,timestamp:Date.now()})},truncate(path,len){if(len<0){throw new FS.ErrnoError(28)}var node;if(typeof path=="string"){var lookup=FS.lookupPath(path,{follow:true});node=lookup.node}else{node=path}FS.doTruncate(null,node,len)},ftruncate(fd,len){var stream=FS.getStreamChecked(fd);if(len<0||(stream.flags&2097155)===0){throw new FS.ErrnoError(28)}FS.doTruncate(stream,stream.node,len)},utime(path,atime,mtime){var lookup=FS.lookupPath(path,{follow:true});var node=lookup.node;var setattr=FS.checkOpExists(node.node_ops.setattr,63);setattr(node,{atime,mtime})},open(path,flags,mode=438){if(path===""){throw new FS.ErrnoError(44)}flags=typeof flags=="string"?FS_modeStringToFlags(flags):flags;if(flags&64){mode=mode&4095|32768}else{mode=0}var node;var isDirPath;if(typeof path=="object"){node=path}else{isDirPath=path.endsWith("/");var lookup=FS.lookupPath(path,{follow:!(flags&131072),noent_okay:true});node=lookup.node;path=lookup.path}var created=false;if(flags&64){if(node){if(flags&128){throw new FS.ErrnoError(20)}}else if(isDirPath){throw new FS.ErrnoError(31)}else{node=FS.mknod(path,mode|511,0);created=true}}if(!node){throw new FS.ErrnoError(44)}if(FS.isChrdev(node.mode)){flags&=~512}if(flags&65536&&!FS.isDir(node.mode)){throw new FS.ErrnoError(54)}if(!created){var errCode=FS.mayOpen(node,flags);if(errCode){throw new FS.ErrnoError(errCode)}}if(flags&512&&!created){FS.truncate(node,0)}var trackingFlags=flags;flags&=~(128|512|131072);var stream=FS.createStream({node,path:FS.getPath(node),flags,seekable:true,position:0,stream_ops:node.stream_ops,ungotten:[],error:false});if(stream.stream_ops.open){stream.stream_ops.open(stream)}if(created){FS.chmod(node,mode&511)}if(Module["logReadFiles"]&&!(flags&1)){if(!(path in FS.readFiles)){FS.readFiles[path]=1;dbg(`FS.trackingDelegate error on read file: ${path}`)}}if(FS.trackingDelegate["onOpenFile"]){FS.trackingDelegate["onOpenFile"](path,trackingFlags)}return stream},close(stream){if(FS.isClosed(stream)){throw new FS.ErrnoError(8)}if(stream.getdents)stream.getdents=null;try{if(stream.stream_ops.close){stream.stream_ops.close(stream)}}catch(e){throw e}finally{FS.closeStream(stream.fd)}stream.fd=null;if(stream.path&&FS.trackingDelegate["onCloseFile"]){FS.trackingDelegate["onCloseFile"](stream.path)}},isClosed(stream){return stream.fd===null},llseek(stream,offset,whence){if(FS.isClosed(stream)){throw new FS.ErrnoError(8)}if(!stream.seekable||!stream.stream_ops.llseek){throw new FS.ErrnoError(70)}if(whence!=0&&whence!=1&&whence!=2){throw new FS.ErrnoError(28)}stream.position=stream.stream_ops.llseek(stream,offset,whence);stream.ungotten=[];if(stream.path&&FS.trackingDelegate["onSeekFile"]){FS.trackingDelegate["onSeekFile"](stream.path,stream.position,whence)}return stream.position},read(stream,buffer,offset,length,position){if(length<0||position<0){throw new FS.ErrnoError(28)}if(FS.isClosed(stream)){throw new FS.ErrnoError(8)}if((stream.flags&2097155)===1){throw new FS.ErrnoError(8)}if(FS.isDir(stream.node.mode)){throw new FS.ErrnoError(31)}if(!stream.stream_ops.read){throw new FS.ErrnoError(28)}var seeking=typeof position!="undefined";if(!seeking){position=stream.position}else if(!stream.seekable){throw new FS.ErrnoError(70)}var bytesRead=stream.stream_ops.read(stream,buffer,offset,length,position);if(!seeking)stream.position+=bytesRead;if(stream.path&&FS.trackingDelegate["onReadFile"]){FS.trackingDelegate["onReadFile"](stream.path,bytesRead)}return bytesRead},write(stream,buffer,offset,length,position,canOwn){if(length<0||position<0){throw new FS.ErrnoError(28)}if(FS.isClosed(stream)){throw new FS.ErrnoError(8)}if((stream.flags&2097155)===0){throw new FS.ErrnoError(8)}if(FS.isDir(stream.node.mode)){throw new FS.ErrnoError(31)}if(!stream.stream_ops.write){throw new FS.ErrnoError(28)}if(stream.seekable&&stream.flags&1024){FS.llseek(stream,0,2)}var seeking=typeof position!="undefined";if(!seeking){position=stream.position}else if(!stream.seekable){throw new FS.ErrnoError(70)}var bytesWritten=stream.stream_ops.write(stream,buffer,offset,length,position,canOwn);if(!seeking)stream.position+=bytesWritten;if(stream.path&&FS.trackingDelegate["onWriteToFile"]){FS.trackingDelegate["onWriteToFile"](stream.path,bytesWritten)}return bytesWritten},mmap(stream,length,position,prot,flags){if((prot&2)!==0&&(flags&2)===0&&(stream.flags&2097155)!==2){throw new FS.ErrnoError(2)}if((stream.flags&2097155)===1){throw new FS.ErrnoError(2)}if(!stream.stream_ops.mmap){throw new FS.ErrnoError(43)}if(!length){throw new FS.ErrnoError(28)}return stream.stream_ops.mmap(stream,length,position,prot,flags)},msync(stream,buffer,offset,length,mmapFlags){if(!stream.stream_ops.msync){return 0}return stream.stream_ops.msync(stream,buffer,offset,length,mmapFlags)},ioctl(stream,cmd,arg){if(!stream.stream_ops.ioctl){throw new FS.ErrnoError(59)}return stream.stream_ops.ioctl(stream,cmd,arg)},readFile(path,opts={}){opts.flags=opts.flags||0;opts.encoding=opts.encoding||"binary";if(opts.encoding!=="utf8"&&opts.encoding!=="binary"){throw new Error(`Invalid encoding type "${opts.encoding}"`)}var ret;var stream=FS.open(path,opts.flags);var stat=FS.stat(path);var length=stat.size;var buf=new Uint8Array(length);FS.read(stream,buf,0,length,0);if(opts.encoding==="utf8"){ret=UTF8ArrayToString(buf)}else if(opts.encoding==="binary"){ret=buf}FS.close(stream);return ret},writeFile(path,data,opts={}){opts.flags=opts.flags||577;var stream=FS.open(path,opts.flags,opts.mode);if(typeof data=="string"){var buf=new Uint8Array(lengthBytesUTF8(data)+1);var actualNumBytes=stringToUTF8Array(data,buf,0,buf.length);FS.write(stream,buf,0,actualNumBytes,undefined,opts.canOwn)}else if(ArrayBuffer.isView(data)){FS.write(stream,data,0,data.byteLength,undefined,opts.canOwn)}else{throw new Error("Unsupported data type")}FS.close(stream)},cwd:()=>FS.currentPath,chdir(path){var lookup=FS.lookupPath(path,{follow:true});if(lookup.node===null){throw new FS.ErrnoError(44)}if(!FS.isDir(lookup.node.mode)){throw new FS.ErrnoError(54)}var errCode=FS.nodePermissions(lookup.node,"x");if(errCode){throw new FS.ErrnoError(errCode)}FS.currentPath=lookup.path},createDefaultDirectories(){FS.mkdir("/tmp");FS.mkdir("/home");FS.mkdir("/home/web_user")},createDefaultDevices(){FS.mkdir("/dev");FS.registerDevice(FS.makedev(1,3),{read:()=>0,write:(stream,buffer,offset,length,pos)=>length,llseek:()=>0});FS.mkdev("/dev/null",FS.makedev(1,3));TTY.register(FS.makedev(5,0),TTY.default_tty_ops);TTY.register(FS.makedev(6,0),TTY.default_tty1_ops);FS.mkdev("/dev/tty",FS.makedev(5,0));FS.mkdev("/dev/tty1",FS.makedev(6,0));var randomBuffer=new Uint8Array(1024),randomLeft=0;var randomByte=()=>{if(randomLeft===0){randomFill(randomBuffer);randomLeft=randomBuffer.byteLength}return randomBuffer[--randomLeft]};FS.createDevice("/dev","random",randomByte);FS.createDevice("/dev","urandom",randomByte);FS.mkdir("/dev/shm");FS.mkdir("/dev/shm/tmp")},createSpecialDirectories(){FS.mkdir("/proc");var proc_self=FS.mkdir("/proc/self");FS.mkdir("/proc/self/fd");FS.mount({mount(){var node=FS.createNode(proc_self,"fd",16895,73);node.stream_ops={llseek:MEMFS.stream_ops.llseek};node.node_ops={lookup(parent,name){var fd=+name;var stream=FS.getStreamChecked(fd);var ret={parent:null,mount:{mountpoint:"fake"},node_ops:{readlink:()=>stream.path},id:fd+1};ret.parent=ret;return ret},readdir(){return Array.from(FS.streams.entries()).filter(([k,v])=>v).map(([k,v])=>k.toString())}};return node}},{},"/proc/self/fd")},createStandardStreams(input,output,error){if(input){FS.createDevice("/dev","stdin",input)}else{FS.symlink("/dev/tty","/dev/stdin")}if(output){FS.createDevice("/dev","stdout",null,output)}else{FS.symlink("/dev/tty","/dev/stdout")}if(error){FS.createDevice("/dev","stderr",null,error)}else{FS.symlink("/dev/tty1","/dev/stderr")}var stdin=FS.open("/dev/stdin",0);var stdout=FS.open("/dev/stdout",1);var stderr=FS.open("/dev/stderr",1)},staticInit(){FS.nameTable=new Array(4096);FS.mount(MEMFS,{},"/");FS.createDefaultDirectories();FS.createDefaultDevices();FS.createSpecialDirectories();FS.filesystems={MEMFS,IDBFS,NODEFS,WORKERFS,PROXYFS}},init(input,output,error){FS.initialized=true;input??=Module["stdin"];output??=Module["stdout"];error??=Module["stderr"];FS.createStandardStreams(input,output,error)},quit(){FS.initialized=false;_fflush(0);for(var stream of FS.streams){if(stream){FS.close(stream)}}},findObject(path,dontResolveLastLink){var ret=FS.analyzePath(path,dontResolveLastLink);if(!ret.exists){return null}return ret.object},analyzePath(path,dontResolveLastLink){try{var lookup=FS.lookupPath(path,{follow:!dontResolveLastLink});path=lookup.path}catch(e){}var ret={isRoot:false,exists:false,error:0,name:null,path:null,object:null,parentExists:false,parentPath:null,parentObject:null};try{var lookup=FS.lookupPath(path,{parent:true});ret.parentExists=true;ret.parentPath=lookup.path;ret.parentObject=lookup.node;ret.name=PATH.basename(path);lookup=FS.lookupPath(path,{follow:!dontResolveLastLink});ret.exists=true;ret.path=lookup.path;ret.object=lookup.node;ret.name=lookup.node.name;ret.isRoot=lookup.path==="/"}catch(e){ret.error=e.errno}return ret},createPath(parent,path,canRead,canWrite){parent=typeof parent=="string"?parent:FS.getPath(parent);var parts=path.split("/").reverse();while(parts.length){var part=parts.pop();if(!part)continue;var current=PATH.join2(parent,part);try{FS.mkdir(current)}catch(e){if(e.errno!=20)throw e}parent=current}return current},createFile(parent,name,properties,canRead,canWrite){var path=PATH.join2(typeof parent=="string"?parent:FS.getPath(parent),name);var mode=FS_getMode(canRead,canWrite);return FS.create(path,mode)},createDataFile(parent,name,data,canRead,canWrite,canOwn){var path=name;if(parent){parent=typeof parent=="string"?parent:FS.getPath(parent);path=name?PATH.join2(parent,name):parent}var mode=FS_getMode(canRead,canWrite);var node=FS.create(path,mode);if(data){if(typeof data=="string"){var arr=new Array(data.length);for(var i=0,len=data.length;i<len;++i)arr[i]=data.charCodeAt(i);data=arr}FS.chmod(node,mode|146);var stream=FS.open(node,577);FS.write(stream,data,0,data.length,0,canOwn);FS.close(stream);FS.chmod(node,mode)}},createDevice(parent,name,input,output){var path=PATH.join2(typeof parent=="string"?parent:FS.getPath(parent),name);var mode=FS_getMode(!!input,!!output);FS.createDevice.major??=64;var dev=FS.makedev(FS.createDevice.major++,0);FS.registerDevice(dev,{open(stream){stream.seekable=false},close(stream){if(output?.buffer?.length){output(10)}},read(stream,buffer,offset,length,pos){var bytesRead=0;for(var i=0;i<length;i++){var result;try{result=input()}catch(e){throw new FS.ErrnoError(29)}if(result===undefined&&bytesRead===0){throw new FS.ErrnoError(6)}if(result===null||result===undefined)break;bytesRead++;buffer[offset+i]=result}if(bytesRead){stream.node.atime=Date.now()}return bytesRead},write(stream,buffer,offset,length,pos){for(var i=0;i<length;i++){try{output(buffer[offset+i])}catch(e){throw new FS.ErrnoError(29)}}if(length){stream.node.mtime=stream.node.ctime=Date.now()}return i}});return FS.mkdev(path,mode,dev)},forceLoadFile(obj){if(obj.isDevice||obj.isFolder||obj.link||obj.contents)return true;dbg(`forceLoadFile: ${obj.url}`);if(typeof XMLHttpRequest!="undefined"){throw new Error("Lazy loading should have been performed (contents set) in createLazyFile, but it was not. Lazy loading only works in web workers. Use --embed-file or --preload-file in emcc on the main thread.")}else{try{obj.contents=readBinary(obj.url);obj.usedBytes=obj.contents.length}catch(e){throw new FS.ErrnoError(29)}}},createLazyFile(parent,name,url,canRead,canWrite){class LazyUint8Array{lengthKnown=false;chunks=[];get(idx){if(idx>this.length-1||idx<0){return undefined}var chunkOffset=idx%this.chunkSize;var chunkNum=idx/this.chunkSize|0;return this.getter(chunkNum)[chunkOffset]}setDataGetter(getter){this.getter=getter}cacheLength(){var xhr=new XMLHttpRequest;xhr.open("HEAD",url,false);xhr.send(null);if(!(xhr.status>=200&&xhr.status<300||xhr.status===304))throw new Error("Couldn't load "+url+". Status: "+xhr.status);var datalength=Number(xhr.getResponseHeader("Content-length"));var header;var hasByteServing=(header=xhr.getResponseHeader("Accept-Ranges"))&&header==="bytes";var usesGzip=(header=xhr.getResponseHeader("Content-Encoding"))&&header==="gzip";var chunkSize=1024*1024;if(!hasByteServing)chunkSize=datalength;var doXHR=(from,to)=>{if(from>to)throw new Error("invalid range ("+from+", "+to+") or no bytes requested!");if(to>datalength-1)throw new Error("only "+datalength+" bytes available! programmer error!");var xhr=new XMLHttpRequest;xhr.open("GET",url,false);if(datalength!==chunkSize)xhr.setRequestHeader("Range","bytes="+from+"-"+to);xhr.responseType="arraybuffer";if(xhr.overrideMimeType){xhr.overrideMimeType("text/plain; charset=x-user-defined")}xhr.send(null);if(!(xhr.status>=200&&xhr.status<300||xhr.status===304))throw new Error("Couldn't load "+url+". Status: "+xhr.status);if(xhr.response!==undefined){return new Uint8Array(xhr.response||[])}return intArrayFromString(xhr.responseText||"",true)};var lazyArray=this;lazyArray.setDataGetter(chunkNum=>{var start=chunkNum*chunkSize;var end=(chunkNum+1)*chunkSize-1;end=Math.min(end,datalength-1);if(typeof lazyArray.chunks[chunkNum]=="undefined"){lazyArray.chunks[chunkNum]=doXHR(start,end)}if(typeof lazyArray.chunks[chunkNum]=="undefined")throw new Error("doXHR failed!");return lazyArray.chunks[chunkNum]});if(usesGzip||!datalength){chunkSize=datalength=1;datalength=this.getter(0).length;chunkSize=datalength;out("LazyFiles on gzip forces download of the whole file when length is accessed")}this._length=datalength;this._chunkSize=chunkSize;this.lengthKnown=true}get length(){if(!this.lengthKnown){this.cacheLength()}return this._length}get chunkSize(){if(!this.lengthKnown){this.cacheLength()}return this._chunkSize}}if(typeof XMLHttpRequest!="undefined"){if(!ENVIRONMENT_IS_WORKER)throw"Cannot do synchronous binary XHRs outside webworkers in modern browsers. Use --embed-file or --preload-file in emcc";var lazyArray=new LazyUint8Array;var properties={isDevice:false,contents:lazyArray}}else{var properties={isDevice:false,url}}var node=FS.createFile(parent,name,properties,canRead,canWrite);if(properties.contents){node.contents=properties.contents}else if(properties.url){node.contents=null;node.url=properties.url}Object.defineProperties(node,{usedBytes:{get:function(){return this.contents.length}}});var stream_ops={};var keys=Object.keys(node.stream_ops);keys.forEach(key=>{var fn=node.stream_ops[key];stream_ops[key]=(...args)=>{FS.forceLoadFile(node);return fn(...args)}});function writeChunks(stream,buffer,offset,length,position){var contents=stream.node.contents;if(position>=contents.length)return 0;var size=Math.min(contents.length-position,length);if(contents.slice){for(var i=0;i<size;i++){buffer[offset+i]=contents[position+i]}}else{for(var i=0;i<size;i++){buffer[offset+i]=contents.get(position+i)}}return size}stream_ops.read=(stream,buffer,offset,length,position)=>{FS.forceLoadFile(node);return writeChunks(stream,buffer,offset,length,position)};stream_ops.mmap=(stream,length,position,prot,flags)=>{FS.forceLoadFile(node);var ptr=mmapAlloc(length);if(!ptr){throw new FS.ErrnoError(48)}writeChunks(stream,HEAP8,ptr,length,position);return{ptr,allocated:true}};node.stream_ops=stream_ops;return node}};var findLibraryFS=(libName,rpath)=>{if(!runtimeInitialized){return undefined}if(PATH.isAbs(libName)){try{FS.lookupPath(libName);return libName}catch(e){return undefined}}var rpathResolved=(rpath?.paths||[]).map(p=>replaceORIGIN(rpath?.parentLibPath,p));return withStackSave(()=>{var bufSize=2*255+2;var buf=stackAlloc(bufSize);var rpathC=stringToUTF8OnStack(rpathResolved.join(":"));var libNameC=stringToUTF8OnStack(libName);var resLibNameC=__emscripten_find_dylib(buf,rpathC,libNameC,bufSize);return resLibNameC?UTF8ToString(resLibNameC):undefined})};function loadDynamicLibrary(libName,flags={global:true,nodelete:true},localScope,handle){var dso=LDSO.loadedLibsByName[libName];if(dso){if(!flags.global){if(localScope){Object.assign(localScope,dso.exports)}}else if(!dso.global){dso.global=true;mergeLibSymbols(dso.exports,libName)}if(flags.nodelete&&dso.refcount!==Infinity){dso.refcount=Infinity}dso.refcount++;if(handle){LDSO.loadedLibsByHandle[handle]=dso}return flags.loadAsync?Promise.resolve(true):true}dso=newDSO(libName,handle,"loading");dso.refcount=flags.nodelete?Infinity:1;dso.global=flags.global;function loadLibData(){if(handle){var data=HEAPU32[handle+28>>>2>>>0];var dataSize=HEAPU32[handle+32>>>2>>>0];if(data&&dataSize){var libData=HEAP8.slice(data,data+dataSize);return flags.loadAsync?Promise.resolve(libData):libData}}var f=findLibraryFS(libName,flags.rpath);if(f){var libData=FS.readFile(f,{encoding:"binary"});return flags.loadAsync?Promise.resolve(libData):libData}var libFile=locateFile(libName);if(flags.loadAsync){return asyncLoad(libFile)}if(!readBinary){throw new Error(`${libFile}: file not found, and synchronous loading of external files is not available`)}return readBinary(libFile)}function getExports(){var preloaded=preloadedWasm[libName];if(preloaded){return flags.loadAsync?Promise.resolve(preloaded):preloaded}if(flags.loadAsync){return loadLibData().then(libData=>loadWebAssemblyModule(libData,flags,libName,localScope,handle))}return loadWebAssemblyModule(loadLibData(),flags,libName,localScope,handle)}function moduleLoaded(exports){if(dso.global){mergeLibSymbols(exports,libName)}else if(localScope){Object.assign(localScope,exports)}dso.exports=exports}if(flags.loadAsync){return getExports().then(exports=>{moduleLoaded(exports);return true})}moduleLoaded(getExports());return true}var reportUndefinedSymbols=()=>{for(var[symName,entry]of Object.entries(GOT)){if(entry.value==0){var value=resolveGlobalSymbol(symName,true).sym;if(!value&&!entry.required){continue}if(typeof value=="function"){entry.value=addFunction(value,value.sig)}else if(typeof value=="number"){entry.value=value}else{throw new Error(`bad export type for '${symName}': ${typeof value}`)}}}};var loadDylibs=()=>{if(!dynamicLibraries.length){reportUndefinedSymbols();return}addRunDependency("loadDylibs");dynamicLibraries.reduce((chain,lib)=>chain.then(()=>loadDynamicLibrary(lib,{loadAsync:true,global:true,nodelete:true,allowUndefined:true})),Promise.resolve()).then(()=>{reportUndefinedSymbols();removeRunDependency("loadDylibs")})};var noExitRuntime=false;function setValue(ptr,value,type="i8"){if(type.endsWith("*"))type="*";switch(type){case"i1":HEAP8[ptr>>>0]=value;break;case"i8":HEAP8[ptr>>>0]=value;break;case"i16":HEAP16[ptr>>>1>>>0]=value;break;case"i32":HEAP32[ptr>>>2>>>0]=value;break;case"i64":HEAP64[ptr>>>3>>>0]=BigInt(value);break;case"float":HEAPF32[ptr>>>2>>>0]=value;break;case"double":HEAPF64[ptr>>>3>>>0]=value;break;case"*":HEAPU32[ptr>>>2>>>0]=value;break;default:abort(`invalid type for setValue: ${type}`)}}var INT53_MAX=9007199254740992;var INT53_MIN=-9007199254740992;var bigintToI53Checked=num=>num<INT53_MIN||num>INT53_MAX?NaN:Number(num);function ___assert_fail(condition,filename,line,func){condition>>>=0;filename>>>=0;func>>>=0;return abort(`Assertion failed: ${UTF8ToString(condition)}, at: `+[filename?UTF8ToString(filename):"unknown filename",line,func?UTF8ToString(func):"unknown function"])}___assert_fail.sig="vppip";var ___c_longjmp=new WebAssembly.Tag({parameters:["i32"]});function ___call_sighandler(fp,sig){fp>>>=0;return getWasmTableEntry(fp)(sig)}___call_sighandler.sig="vpi";var ___cpp_exception=new WebAssembly.Tag({parameters:["i32"]});var ___memory_base=new WebAssembly.Global({value:"i32",mutable:false},1024);var ___stack_high=8809120;var ___stack_low=3566240;var ___stack_pointer=new WebAssembly.Global({value:"i32",mutable:true},8809120);var SYSCALLS={DEFAULT_POLLMASK:5,calculateAt(dirfd,path,allowEmpty){if(PATH.isAbs(path)){return path}var dir;if(dirfd===-100){dir=FS.cwd()}else{var dirstream=SYSCALLS.getStreamFromFD(dirfd);dir=dirstream.path}if(path.length==0){if(!allowEmpty){throw new FS.ErrnoError(44)}return dir}return dir+"/"+path},writeStat(buf,stat){HEAP32[buf>>>2>>>0]=stat.dev;HEAP32[buf+4>>>2>>>0]=stat.mode;HEAPU32[buf+8>>>2>>>0]=stat.nlink;HEAP32[buf+12>>>2>>>0]=stat.uid;HEAP32[buf+16>>>2>>>0]=stat.gid;HEAP32[buf+20>>>2>>>0]=stat.rdev;HEAP64[buf+24>>>3>>>0]=BigInt(stat.size);HEAP32[buf+32>>>2>>>0]=4096;HEAP32[buf+36>>>2>>>0]=stat.blocks;var atime=stat.atime.getTime();var mtime=stat.mtime.getTime();var ctime=stat.ctime.getTime();HEAP64[buf+40>>>3>>>0]=BigInt(Math.floor(atime/1e3));HEAPU32[buf+48>>>2>>>0]=atime%1e3*1e3*1e3;HEAP64[buf+56>>>3>>>0]=BigInt(Math.floor(mtime/1e3));HEAPU32[buf+64>>>2>>>0]=mtime%1e3*1e3*1e3;HEAP64[buf+72>>>3>>>0]=BigInt(Math.floor(ctime/1e3));HEAPU32[buf+80>>>2>>>0]=ctime%1e3*1e3*1e3;HEAP64[buf+88>>>3>>>0]=BigInt(stat.ino);return 0},writeStatFs(buf,stats){HEAP32[buf+4>>>2>>>0]=stats.bsize;HEAP32[buf+40>>>2>>>0]=stats.bsize;HEAP32[buf+8>>>2>>>0]=stats.blocks;HEAP32[buf+12>>>2>>>0]=stats.bfree;HEAP32[buf+16>>>2>>>0]=stats.bavail;HEAP32[buf+20>>>2>>>0]=stats.files;HEAP32[buf+24>>>2>>>0]=stats.ffree;HEAP32[buf+28>>>2>>>0]=stats.fsid;HEAP32[buf+44>>>2>>>0]=stats.flags;HEAP32[buf+36>>>2>>>0]=stats.namelen},doMsync(addr,stream,len,flags,offset){if(!FS.isFile(stream.node.mode)){throw new FS.ErrnoError(43)}if(flags&2){return 0}var buffer=HEAPU8.slice(addr,addr+len);FS.msync(stream,buffer,offset,len,flags)},getStreamFromFD(fd){var stream=FS.getStreamChecked(fd);return stream},varargs:undefined,getStr(ptr){var ret=UTF8ToString(ptr);return ret}};var ___syscall__newselect=function(nfds,readfds,writefds,exceptfds,timeout){readfds>>>=0;writefds>>>=0;exceptfds>>>=0;timeout>>>=0;try{var total=0;var srcReadLow=readfds?HEAP32[readfds>>>2>>>0]:0,srcReadHigh=readfds?HEAP32[readfds+4>>>2>>>0]:0;var srcWriteLow=writefds?HEAP32[writefds>>>2>>>0]:0,srcWriteHigh=writefds?HEAP32[writefds+4>>>2>>>0]:0;var srcExceptLow=exceptfds?HEAP32[exceptfds>>>2>>>0]:0,srcExceptHigh=exceptfds?HEAP32[exceptfds+4>>>2>>>0]:0;var dstReadLow=0,dstReadHigh=0;var dstWriteLow=0,dstWriteHigh=0;var dstExceptLow=0,dstExceptHigh=0;var allLow=(readfds?HEAP32[readfds>>>2>>>0]:0)|(writefds?HEAP32[writefds>>>2>>>0]:0)|(exceptfds?HEAP32[exceptfds>>>2>>>0]:0);var allHigh=(readfds?HEAP32[readfds+4>>>2>>>0]:0)|(writefds?HEAP32[writefds+4>>>2>>>0]:0)|(exceptfds?HEAP32[exceptfds+4>>>2>>>0]:0);var check=(fd,low,high,val)=>fd<32?low&val:high&val;for(var fd=0;fd<nfds;fd++){var mask=1<<fd%32;if(!check(fd,allLow,allHigh,mask)){continue}var stream=SYSCALLS.getStreamFromFD(fd);var flags=SYSCALLS.DEFAULT_POLLMASK;if(stream.stream_ops.poll){var timeoutInMillis=-1;if(timeout){var tv_sec=readfds?HEAP32[timeout>>>2>>>0]:0,tv_usec=readfds?HEAP32[timeout+4>>>2>>>0]:0;timeoutInMillis=(tv_sec+tv_usec/1e6)*1e3}flags=stream.stream_ops.poll(stream,timeoutInMillis)}if(flags&1&&check(fd,srcReadLow,srcReadHigh,mask)){fd<32?dstReadLow=dstReadLow|mask:dstReadHigh=dstReadHigh|mask;total++}if(flags&4&&check(fd,srcWriteLow,srcWriteHigh,mask)){fd<32?dstWriteLow=dstWriteLow|mask:dstWriteHigh=dstWriteHigh|mask;total++}if(flags&2&&check(fd,srcExceptLow,srcExceptHigh,mask)){fd<32?dstExceptLow=dstExceptLow|mask:dstExceptHigh=dstExceptHigh|mask;total++}}if(readfds){HEAP32[readfds>>>2>>>0]=dstReadLow;HEAP32[readfds+4>>>2>>>0]=dstReadHigh}if(writefds){HEAP32[writefds>>>2>>>0]=dstWriteLow;HEAP32[writefds+4>>>2>>>0]=dstWriteHigh}if(exceptfds){HEAP32[exceptfds>>>2>>>0]=dstExceptLow;HEAP32[exceptfds+4>>>2>>>0]=dstExceptHigh}return total}catch(e){if(typeof FS=="undefined"||!(e.name==="ErrnoError"))throw e;return-e.errno}};___syscall__newselect.sig="iipppp";var SOCKFS={websocketArgs:{},callbacks:{},on(event,callback){SOCKFS.callbacks[event]=callback},emit(event,param){SOCKFS.callbacks[event]?.(param)},mount(mount){SOCKFS.websocketArgs=Module["websocket"]||{};(Module["websocket"]??={})["on"]=SOCKFS.on;return FS.createNode(null,"/",16895,0)},createSocket(family,type,protocol){type&=~526336;var streaming=type==1;if(streaming&&protocol&&protocol!=6){throw new FS.ErrnoError(66)}var sock={family,type,protocol,server:null,error:null,peers:{},pending:[],recv_queue:[],sock_ops:SOCKFS.websocket_sock_ops};var name=SOCKFS.nextname();var node=FS.createNode(SOCKFS.root,name,49152,0);node.sock=sock;var stream=FS.createStream({path:name,node,flags:2,seekable:false,stream_ops:SOCKFS.stream_ops});sock.stream=stream;return sock},getSocket(fd){var stream=FS.getStream(fd);if(!stream||!FS.isSocket(stream.node.mode)){return null}return stream.node.sock},stream_ops:{poll(stream){var sock=stream.node.sock;return sock.sock_ops.poll(sock)},ioctl(stream,request,varargs){var sock=stream.node.sock;return sock.sock_ops.ioctl(sock,request,varargs)},read(stream,buffer,offset,length,position){var sock=stream.node.sock;var msg=sock.sock_ops.recvmsg(sock,length);if(!msg){return 0}buffer.set(msg.buffer,offset);return msg.buffer.length},write(stream,buffer,offset,length,position){var sock=stream.node.sock;return sock.sock_ops.sendmsg(sock,buffer,offset,length)},close(stream){var sock=stream.node.sock;sock.sock_ops.close(sock)}},nextname(){if(!SOCKFS.nextname.current){SOCKFS.nextname.current=0}return`socket[${SOCKFS.nextname.current++}]`},websocket_sock_ops:{createPeer(sock,addr,port){var ws;if(typeof addr=="object"){ws=addr;addr=null;port=null}if(ws){if(ws._socket){addr=ws._socket.remoteAddress;port=ws._socket.remotePort}else{var result=/ws[s]?:\/\/([^:]+):(\d+)/.exec(ws.url);if(!result){throw new Error("WebSocket URL must be in the format ws(s)://address:port")}addr=result[1];port=parseInt(result[2],10)}}else{try{var url="ws://".replace("#","//");var subProtocols="binary";var opts=undefined;if(SOCKFS.websocketArgs["url"]){url=SOCKFS.websocketArgs["url"]}if(SOCKFS.websocketArgs["subprotocol"]){subProtocols=SOCKFS.websocketArgs["subprotocol"]}else if(SOCKFS.websocketArgs["subprotocol"]===null){subProtocols="null"}if(url==="ws://"||url==="wss://"){var parts=addr.split("/");url=url+parts[0]+":"+port+"/"+parts.slice(1).join("/")}if(subProtocols!=="null"){subProtocols=subProtocols.replace(/^ +| +$/g,"").split(/ *, */);opts=subProtocols}var WebSocketConstructor;if(ENVIRONMENT_IS_NODE){WebSocketConstructor=require("ws")}else{WebSocketConstructor=WebSocket}ws=new WebSocketConstructor(url,opts);ws.binaryType="arraybuffer"}catch(e){throw new FS.ErrnoError(23)}}var peer={addr,port,socket:ws,msg_send_queue:[]};SOCKFS.websocket_sock_ops.addPeer(sock,peer);SOCKFS.websocket_sock_ops.handlePeerEvents(sock,peer);if(sock.type===2&&typeof sock.sport!="undefined"){peer.msg_send_queue.push(new Uint8Array([255,255,255,255,"p".charCodeAt(0),"o".charCodeAt(0),"r".charCodeAt(0),"t".charCodeAt(0),(sock.sport&65280)>>8,sock.sport&255]))}return peer},getPeer(sock,addr,port){return sock.peers[addr+":"+port]},addPeer(sock,peer){sock.peers[peer.addr+":"+peer.port]=peer},removePeer(sock,peer){delete sock.peers[peer.addr+":"+peer.port]},handlePeerEvents(sock,peer){var first=true;var handleOpen=function(){sock.connecting=false;SOCKFS.emit("open",sock.stream.fd);try{var queued=peer.msg_send_queue.shift();while(queued){peer.socket.send(queued);queued=peer.msg_send_queue.shift()}}catch(e){peer.socket.close()}};function handleMessage(data){if(typeof data=="string"){var encoder=new TextEncoder;data=encoder.encode(data)}else{assert(data.byteLength!==undefined);if(data.byteLength==0){return}data=new Uint8Array(data)}var wasfirst=first;first=false;if(wasfirst&&data.length===10&&data[0]===255&&data[1]===255&&data[2]===255&&data[3]===255&&data[4]==="p".charCodeAt(0)&&data[5]==="o".charCodeAt(0)&&data[6]==="r".charCodeAt(0)&&data[7]==="t".charCodeAt(0)){var newport=data[8]<<8|data[9];SOCKFS.websocket_sock_ops.removePeer(sock,peer);peer.port=newport;SOCKFS.websocket_sock_ops.addPeer(sock,peer);return}sock.recv_queue.push({addr:peer.addr,port:peer.port,data});SOCKFS.emit("message",sock.stream.fd)}if(ENVIRONMENT_IS_NODE){peer.socket.on("open",handleOpen);peer.socket.on("message",function(data,isBinary){if(!isBinary){return}handleMessage(new Uint8Array(data).buffer)});peer.socket.on("close",function(){SOCKFS.emit("close",sock.stream.fd)});peer.socket.on("error",function(error){sock.error=14;SOCKFS.emit("error",[sock.stream.fd,sock.error,"ECONNREFUSED: Connection refused"])})}else{peer.socket.onopen=handleOpen;peer.socket.onclose=function(){SOCKFS.emit("close",sock.stream.fd)};peer.socket.onmessage=function peer_socket_onmessage(event){handleMessage(event.data)};peer.socket.onerror=function(error){sock.error=14;SOCKFS.emit("error",[sock.stream.fd,sock.error,"ECONNREFUSED: Connection refused"])}}},poll(sock){if(sock.type===1&&sock.server){return sock.pending.length?64|1:0}var mask=0;var dest=sock.type===1?SOCKFS.websocket_sock_ops.getPeer(sock,sock.daddr,sock.dport):null;if(sock.recv_queue.length||!dest||dest&&dest.socket.readyState===dest.socket.CLOSING||dest&&dest.socket.readyState===dest.socket.CLOSED){mask|=64|1}if(!dest||dest&&dest.socket.readyState===dest.socket.OPEN){mask|=4}if(dest&&dest.socket.readyState===dest.socket.CLOSING||dest&&dest.socket.readyState===dest.socket.CLOSED){if(sock.connecting){mask|=4}else{mask|=16}}return mask},ioctl(sock,request,arg){switch(request){case 21531:var bytes=0;if(sock.recv_queue.length){bytes=sock.recv_queue[0].data.length}HEAP32[arg>>>2>>>0]=bytes;return 0;default:return 28}},close(sock){if(sock.server){try{sock.server.close()}catch(e){}sock.server=null}for(var peer of Object.values(sock.peers)){try{peer.socket.close()}catch(e){}SOCKFS.websocket_sock_ops.removePeer(sock,peer)}return 0},bind(sock,addr,port){if(typeof sock.saddr!="undefined"||typeof sock.sport!="undefined"){throw new FS.ErrnoError(28)}sock.saddr=addr;sock.sport=port;if(sock.type===2){if(sock.server){sock.server.close();sock.server=null}try{sock.sock_ops.listen(sock,0)}catch(e){if(!(e.name==="ErrnoError"))throw e;if(e.errno!==138)throw e}}},connect(sock,addr,port){if(sock.server){throw new FS.ErrnoError(138)}if(typeof sock.daddr!="undefined"&&typeof sock.dport!="undefined"){var dest=SOCKFS.websocket_sock_ops.getPeer(sock,sock.daddr,sock.dport);if(dest){if(dest.socket.readyState===dest.socket.CONNECTING){throw new FS.ErrnoError(7)}else{throw new FS.ErrnoError(30)}}}var peer=SOCKFS.websocket_sock_ops.createPeer(sock,addr,port);sock.daddr=peer.addr;sock.dport=peer.port;sock.connecting=true},listen(sock,backlog){if(!ENVIRONMENT_IS_NODE){throw new FS.ErrnoError(138)}if(sock.server){throw new FS.ErrnoError(28)}var WebSocketServer=require("ws").Server;var host=sock.saddr;sock.server=new WebSocketServer({host,port:sock.sport});SOCKFS.emit("listen",sock.stream.fd);sock.server.on("connection",function(ws){if(sock.type===1){var newsock=SOCKFS.createSocket(sock.family,sock.type,sock.protocol);var peer=SOCKFS.websocket_sock_ops.createPeer(newsock,ws);newsock.daddr=peer.addr;newsock.dport=peer.port;sock.pending.push(newsock);SOCKFS.emit("connection",newsock.stream.fd)}else{SOCKFS.websocket_sock_ops.createPeer(sock,ws);SOCKFS.emit("connection",sock.stream.fd)}});sock.server.on("close",function(){SOCKFS.emit("close",sock.stream.fd);sock.server=null});sock.server.on("error",function(error){sock.error=23;SOCKFS.emit("error",[sock.stream.fd,sock.error,"EHOSTUNREACH: Host is unreachable"])})},accept(listensock){if(!listensock.server||!listensock.pending.length){throw new FS.ErrnoError(28)}var newsock=listensock.pending.shift();newsock.stream.flags=listensock.stream.flags;return newsock},getname(sock,peer){var addr,port;if(peer){if(sock.daddr===undefined||sock.dport===undefined){throw new FS.ErrnoError(53)}addr=sock.daddr;port=sock.dport}else{addr=sock.saddr||0;port=sock.sport||0}return{addr,port}},sendmsg(sock,buffer,offset,length,addr,port){if(sock.type===2){if(addr===undefined||port===undefined){addr=sock.daddr;port=sock.dport}if(addr===undefined||port===undefined){throw new FS.ErrnoError(17)}}else{addr=sock.daddr;port=sock.dport}var dest=SOCKFS.websocket_sock_ops.getPeer(sock,addr,port);if(sock.type===1){if(!dest||dest.socket.readyState===dest.socket.CLOSING||dest.socket.readyState===dest.socket.CLOSED){throw new FS.ErrnoError(53)}}if(ArrayBuffer.isView(buffer)){offset+=buffer.byteOffset;buffer=buffer.buffer}var data=buffer.slice(offset,offset+length);if(!dest||dest.socket.readyState!==dest.socket.OPEN){if(sock.type===2){if(!dest||dest.socket.readyState===dest.socket.CLOSING||dest.socket.readyState===dest.socket.CLOSED){dest=SOCKFS.websocket_sock_ops.createPeer(sock,addr,port)}}dest.msg_send_queue.push(data);return length}try{dest.socket.send(data);return length}catch(e){throw new FS.ErrnoError(28)}},recvmsg(sock,length){if(sock.type===1&&sock.server){throw new FS.ErrnoError(53)}var queued=sock.recv_queue.shift();if(!queued){if(sock.type===1){var dest=SOCKFS.websocket_sock_ops.getPeer(sock,sock.daddr,sock.dport);if(!dest){throw new FS.ErrnoError(53)}if(dest.socket.readyState===dest.socket.CLOSING||dest.socket.readyState===dest.socket.CLOSED){return null}throw new FS.ErrnoError(6)}throw new FS.ErrnoError(6)}var queuedLength=queued.data.byteLength||queued.data.length;var queuedOffset=queued.data.byteOffset||0;var queuedBuffer=queued.data.buffer||queued.data;var bytesRead=Math.min(length,queuedLength);var res={buffer:new Uint8Array(queuedBuffer,queuedOffset,bytesRead),addr:queued.addr,port:queued.port};if(sock.type===1&&bytesRead<queuedLength){var bytesRemaining=queuedLength-bytesRead;queued.data=new Uint8Array(queuedBuffer,queuedOffset+bytesRead,bytesRemaining);sock.recv_queue.unshift(queued)}return res}}};var getSocketFromFD=fd=>{var socket=SOCKFS.getSocket(fd);if(!socket)throw new FS.ErrnoError(8);return socket};var inetPton4=str=>{var b=str.split(".");for(var i=0;i<4;i++){var tmp=Number(b[i]);if(isNaN(tmp))return null;b[i]=tmp}return(b[0]|b[1]<<8|b[2]<<16|b[3]<<24)>>>0};var inetPton6=str=>{var words;var w,offset,z,i;var valid6regx=/^((?=.*::)(?!.*::.+::)(::)?([\dA-F]{1,4}:(:|\b)|){5}|([\dA-F]{1,4}:){6})((([\dA-F]{1,4}((?!\3)::|:\b|$))|(?!\2\3)){2}|(((2[0-4]|1\d|[1-9])?\d|25[0-5])\.?\b){4})$/i;var parts=[];if(!valid6regx.test(str)){return null}if(str==="::"){return[0,0,0,0,0,0,0,0]}if(str.startsWith("::")){str=str.replace("::","Z:")}else{str=str.replace("::",":Z:")}if(str.indexOf(".")>0){str=str.replace(new RegExp("[.]","g"),":");words=str.split(":");words[words.length-4]=Number(words[words.length-4])+Number(words[words.length-3])*256;words[words.length-3]=Number(words[words.length-2])+Number(words[words.length-1])*256;words=words.slice(0,words.length-2)}else{words=str.split(":")}offset=0;z=0;for(w=0;w<words.length;w++){if(typeof words[w]=="string"){if(words[w]==="Z"){for(z=0;z<8-words.length+1;z++){parts[w+z]=0}offset=z-1}else{parts[w+offset]=_htons(parseInt(words[w],16))}}else{parts[w+offset]=words[w]}}return[parts[1]<<16|parts[0],parts[3]<<16|parts[2],parts[5]<<16|parts[4],parts[7]<<16|parts[6]]};var writeSockaddr=(sa,family,addr,port,addrlen)=>{switch(family){case 2:addr=inetPton4(addr);zeroMemory(sa,16);if(addrlen){HEAP32[addrlen>>>2>>>0]=16}HEAP16[sa>>>1>>>0]=family;HEAP32[sa+4>>>2>>>0]=addr;HEAP16[sa+2>>>1>>>0]=_htons(port);break;case 10:addr=inetPton6(addr);zeroMemory(sa,28);if(addrlen){HEAP32[addrlen>>>2>>>0]=28}HEAP32[sa>>>2>>>0]=family;HEAP32[sa+8>>>2>>>0]=addr[0];HEAP32[sa+12>>>2>>>0]=addr[1];HEAP32[sa+16>>>2>>>0]=addr[2];HEAP32[sa+20>>>2>>>0]=addr[3];HEAP16[sa+2>>>1>>>0]=_htons(port);break;default:return 5}return 0};var DNS={address_map:{id:1,addrs:{},names:{}},lookup_name(name){var res=inetPton4(name);if(res!==null){return name}res=inetPton6(name);if(res!==null){return name}var addr;if(DNS.address_map.addrs[name]){addr=DNS.address_map.addrs[name]}else{var id=DNS.address_map.id++;assert(id<65535,"exceeded max address mappings of 65535");addr="172.29."+(id&255)+"."+(id&65280);DNS.address_map.names[addr]=name;DNS.address_map.addrs[name]=addr}return addr},lookup_addr(addr){if(DNS.address_map.names[addr]){return DNS.address_map.names[addr]}return null}};function ___syscall_accept4(fd,addr,addrlen,flags,d1,d2){addr>>>=0;addrlen>>>=0;try{var sock=getSocketFromFD(fd);var newsock=sock.sock_ops.accept(sock);if(addr){var errno=writeSockaddr(addr,newsock.family,DNS.lookup_name(newsock.daddr),newsock.dport,addrlen)}return newsock.stream.fd}catch(e){if(typeof FS=="undefined"||!(e.name==="ErrnoError"))throw e;return-e.errno}}___syscall_accept4.sig="iippiii";var inetNtop4=addr=>(addr&255)+"."+(addr>>8&255)+"."+(addr>>16&255)+"."+(addr>>24&255);var inetNtop6=ints=>{var str="";var word=0;var longest=0;var lastzero=0;var zstart=0;var len=0;var i=0;var parts=[ints[0]&65535,ints[0]>>16,ints[1]&65535,ints[1]>>16,ints[2]&65535,ints[2]>>16,ints[3]&65535,ints[3]>>16];var hasipv4=true;var v4part="";for(i=0;i<5;i++){if(parts[i]!==0){hasipv4=false;break}}if(hasipv4){v4part=inetNtop4(parts[6]|parts[7]<<16);if(parts[5]===-1){str="::ffff:";str+=v4part;return str}if(parts[5]===0){str="::";if(v4part==="0.0.0.0")v4part="";if(v4part==="0.0.0.1")v4part="1";str+=v4part;return str}}for(word=0;word<8;word++){if(parts[word]===0){if(word-lastzero>1){len=0}lastzero=word;len++}if(len>longest){longest=len;zstart=word-longest+1}}for(word=0;word<8;word++){if(longest>1){if(parts[word]===0&&word>=zstart&&word<zstart+longest){if(word===zstart){str+=":";if(zstart===0)str+=":"}continue}}str+=Number(_ntohs(parts[word]&65535)).toString(16);str+=word<7?":":""}return str};var readSockaddr=(sa,salen)=>{var family=HEAP16[sa>>>1>>>0];var port=_ntohs(HEAPU16[sa+2>>>1>>>0]);var addr;switch(family){case 2:if(salen!==16){return{errno:28}}addr=HEAP32[sa+4>>>2>>>0];addr=inetNtop4(addr);break;case 10:if(salen!==28){return{errno:28}}addr=[HEAP32[sa+8>>>2>>>0],HEAP32[sa+12>>>2>>>0],HEAP32[sa+16>>>2>>>0],HEAP32[sa+20>>>2>>>0]];addr=inetNtop6(addr);break;default:return{errno:5}}return{family,addr,port}};var getSocketAddress=(addrp,addrlen)=>{var info=readSockaddr(addrp,addrlen);if(info.errno)throw new FS.ErrnoError(info.errno);info.addr=DNS.lookup_addr(info.addr)||info.addr;return info};function ___syscall_bind(fd,addr,addrlen,d1,d2,d3){addr>>>=0;addrlen>>>=0;try{var sock=getSocketFromFD(fd);var info=getSocketAddress(addr,addrlen);sock.sock_ops.bind(sock,info.addr,info.port);return 0}catch(e){if(typeof FS=="undefined"||!(e.name==="ErrnoError"))throw e;return-e.errno}}___syscall_bind.sig="iippiii";function ___syscall_chdir(path){path>>>=0;try{path=SYSCALLS.getStr(path);FS.chdir(path);return 0}catch(e){if(typeof FS=="undefined"||!(e.name==="ErrnoError"))throw e;return-e.errno}}___syscall_chdir.sig="ip";function ___syscall_chmod(path,mode){path>>>=0;try{path=SYSCALLS.getStr(path);FS.chmod(path,mode);return 0}catch(e){if(typeof FS=="undefined"||!(e.name==="ErrnoError"))throw e;return-e.errno}}___syscall_chmod.sig="ipi";function ___syscall_connect(fd,addr,addrlen,d1,d2,d3){addr>>>=0;addrlen>>>=0;try{var sock=getSocketFromFD(fd);var info=getSocketAddress(addr,addrlen);sock.sock_ops.connect(sock,info.addr,info.port);return 0}catch(e){if(typeof FS=="undefined"||!(e.name==="ErrnoError"))throw e;return-e.errno}}___syscall_connect.sig="iippiii";function ___syscall_dup(fd){try{var old=SYSCALLS.getStreamFromFD(fd);return FS.dupStream(old).fd}catch(e){if(typeof FS=="undefined"||!(e.name==="ErrnoError"))throw e;return-e.errno}}___syscall_dup.sig="ii";function ___syscall_dup3(fd,newfd,flags){try{var old=SYSCALLS.getStreamFromFD(fd);if(old.fd===newfd)return-28;if(newfd<0||newfd>=FS.MAX_OPEN_FDS)return-8;var existing=FS.getStream(newfd);if(existing)FS.close(existing);return FS.dupStream(old,newfd).fd}catch(e){if(typeof FS=="undefined"||!(e.name==="ErrnoError"))throw e;return-e.errno}}___syscall_dup3.sig="iiii";function ___syscall_faccessat(dirfd,path,amode,flags){path>>>=0;try{path=SYSCALLS.getStr(path);path=SYSCALLS.calculateAt(dirfd,path);if(amode&~7){return-28}var lookup=FS.lookupPath(path,{follow:true});var node=lookup.node;if(!node){return-44}var perms="";if(amode&4)perms+="r";if(amode&2)perms+="w";if(amode&1)perms+="x";if(perms&&FS.nodePermissions(node,perms)){return-2}return 0}catch(e){if(typeof FS=="undefined"||!(e.name==="ErrnoError"))throw e;return-e.errno}}___syscall_faccessat.sig="iipii";var ___syscall_fadvise64=(fd,offset,len,advice)=>0;___syscall_fadvise64.sig="iijji";function ___syscall_fallocate(fd,mode,offset,len){offset=bigintToI53Checked(offset);len=bigintToI53Checked(len);try{if(isNaN(offset)||isNaN(len))return-61;if(mode!=0){return-138}if(offset<0||len<0){return-28}var oldSize=FS.fstat(fd).size;var newSize=offset+len;if(newSize>oldSize){FS.ftruncate(fd,newSize)}return 0}catch(e){if(typeof FS=="undefined"||!(e.name==="ErrnoError"))throw e;return-e.errno}}___syscall_fallocate.sig="iiijj";function ___syscall_fchdir(fd){try{var stream=SYSCALLS.getStreamFromFD(fd);FS.chdir(stream.path);return 0}catch(e){if(typeof FS=="undefined"||!(e.name==="ErrnoError"))throw e;return-e.errno}}___syscall_fchdir.sig="ii";function ___syscall_fchmod(fd,mode){try{FS.fchmod(fd,mode);return 0}catch(e){if(typeof FS=="undefined"||!(e.name==="ErrnoError"))throw e;return-e.errno}}___syscall_fchmod.sig="iii";function ___syscall_fchmodat2(dirfd,path,mode,flags){path>>>=0;try{var nofollow=flags&256;path=SYSCALLS.getStr(path);path=SYSCALLS.calculateAt(dirfd,path);FS.chmod(path,mode,nofollow);return 0}catch(e){if(typeof FS=="undefined"||!(e.name==="ErrnoError"))throw e;return-e.errno}}___syscall_fchmodat2.sig="iipii";function ___syscall_fchown32(fd,owner,group){try{FS.fchown(fd,owner,group);return 0}catch(e){if(typeof FS=="undefined"||!(e.name==="ErrnoError"))throw e;return-e.errno}}___syscall_fchown32.sig="iiii";function ___syscall_fchownat(dirfd,path,owner,group,flags){path>>>=0;try{path=SYSCALLS.getStr(path);var nofollow=flags&256;flags=flags&~256;path=SYSCALLS.calculateAt(dirfd,path);(nofollow?FS.lchown:FS.chown)(path,owner,group);return 0}catch(e){if(typeof FS=="undefined"||!(e.name==="ErrnoError"))throw e;return-e.errno}}___syscall_fchownat.sig="iipiii";var syscallGetVarargI=()=>{var ret=HEAP32[+SYSCALLS.varargs>>>2>>>0];SYSCALLS.varargs+=4;return ret};var syscallGetVarargP=syscallGetVarargI;function ___syscall_fcntl64(fd,cmd,varargs){varargs>>>=0;SYSCALLS.varargs=varargs;try{var stream=SYSCALLS.getStreamFromFD(fd);switch(cmd){case 0:{var arg=syscallGetVarargI();if(arg<0){return-28}while(FS.streams[arg]){arg++}var newStream;newStream=FS.dupStream(stream,arg);return newStream.fd}case 1:case 2:return 0;case 3:return stream.flags;case 4:{var arg=syscallGetVarargI();stream.flags|=arg;return 0}case 12:{var arg=syscallGetVarargP();var offset=0;HEAP16[arg+offset>>>1>>>0]=2;return 0}case 13:case 14:return 0}return-28}catch(e){if(typeof FS=="undefined"||!(e.name==="ErrnoError"))throw e;return-e.errno}}___syscall_fcntl64.sig="iiip";function ___syscall_fdatasync(fd){try{var stream=SYSCALLS.getStreamFromFD(fd);return 0}catch(e){if(typeof FS=="undefined"||!(e.name==="ErrnoError"))throw e;return-e.errno}}___syscall_fdatasync.sig="ii";function ___syscall_fstat64(fd,buf){buf>>>=0;try{return SYSCALLS.writeStat(buf,FS.fstat(fd))}catch(e){if(typeof FS=="undefined"||!(e.name==="ErrnoError"))throw e;return-e.errno}}___syscall_fstat64.sig="iip";function ___syscall_fstatfs64(fd,size,buf){size>>>=0;buf>>>=0;try{var stream=SYSCALLS.getStreamFromFD(fd);SYSCALLS.writeStatFs(buf,FS.statfsStream(stream));return 0}catch(e){if(typeof FS=="undefined"||!(e.name==="ErrnoError"))throw e;return-e.errno}}___syscall_fstatfs64.sig="iipp";function ___syscall_ftruncate64(fd,length){length=bigintToI53Checked(length);try{if(isNaN(length))return-61;FS.ftruncate(fd,length);return 0}catch(e){if(typeof FS=="undefined"||!(e.name==="ErrnoError"))throw e;return-e.errno}}___syscall_ftruncate64.sig="iij";function ___syscall_getcwd(buf,size){buf>>>=0;size>>>=0;try{if(size===0)return-28;var cwd=FS.cwd();var cwdLengthInBytes=lengthBytesUTF8(cwd)+1;if(size<cwdLengthInBytes)return-68;stringToUTF8(cwd,buf,size);return cwdLengthInBytes}catch(e){if(typeof FS=="undefined"||!(e.name==="ErrnoError"))throw e;return-e.errno}}___syscall_getcwd.sig="ipp";function ___syscall_getdents64(fd,dirp,count){dirp>>>=0;count>>>=0;try{var stream=SYSCALLS.getStreamFromFD(fd);stream.getdents||=FS.readdir(stream.path);var struct_size=280;var pos=0;var off=FS.llseek(stream,0,1);var startIdx=Math.floor(off/struct_size);var endIdx=Math.min(stream.getdents.length,startIdx+Math.floor(count/struct_size));for(var idx=startIdx;idx<endIdx;idx++){var id;var type;var name=stream.getdents[idx];if(name==="."){id=stream.node.id;type=4}else if(name===".."){var lookup=FS.lookupPath(stream.path,{parent:true});id=lookup.node.id;type=4}else{var child;try{child=FS.lookupNode(stream.node,name)}catch(e){if(e?.errno===28){continue}throw e}id=child.id;type=FS.isChrdev(child.mode)?2:FS.isDir(child.mode)?4:FS.isLink(child.mode)?10:8}HEAP64[dirp+pos>>>3>>>0]=BigInt(id);HEAP64[dirp+pos+8>>>3>>>0]=BigInt((idx+1)*struct_size);HEAP16[dirp+pos+16>>>1>>>0]=280;HEAP8[dirp+pos+18>>>0]=type;stringToUTF8(name,dirp+pos+19,256);pos+=struct_size}FS.llseek(stream,idx*struct_size,0);return pos}catch(e){if(typeof FS=="undefined"||!(e.name==="ErrnoError"))throw e;return-e.errno}}___syscall_getdents64.sig="iipp";function ___syscall_getpeername(fd,addr,addrlen,d1,d2,d3){addr>>>=0;addrlen>>>=0;try{var sock=getSocketFromFD(fd);if(!sock.daddr){return-53}var errno=writeSockaddr(addr,sock.family,DNS.lookup_name(sock.daddr),sock.dport,addrlen);return 0}catch(e){if(typeof FS=="undefined"||!(e.name==="ErrnoError"))throw e;return-e.errno}}___syscall_getpeername.sig="iippiii";function ___syscall_getsockname(fd,addr,addrlen,d1,d2,d3){addr>>>=0;addrlen>>>=0;try{var sock=getSocketFromFD(fd);var errno=writeSockaddr(addr,sock.family,DNS.lookup_name(sock.saddr||"0.0.0.0"),sock.sport,addrlen);return 0}catch(e){if(typeof FS=="undefined"||!(e.name==="ErrnoError"))throw e;return-e.errno}}___syscall_getsockname.sig="iippiii";function ___syscall_getsockopt(fd,level,optname,optval,optlen,d1){optval>>>=0;optlen>>>=0;try{var sock=getSocketFromFD(fd);if(level===1){if(optname===4){HEAP32[optval>>>2>>>0]=sock.error;HEAP32[optlen>>>2>>>0]=4;sock.error=null;return 0}}return-50}catch(e){if(typeof FS=="undefined"||!(e.name==="ErrnoError"))throw e;return-e.errno}}___syscall_getsockopt.sig="iiiippi";function ___syscall_ioctl(fd,op,varargs){varargs>>>=0;SYSCALLS.varargs=varargs;try{var stream=SYSCALLS.getStreamFromFD(fd);switch(op){case 21509:{if(!stream.tty)return-59;return 0}case 21505:{if(!stream.tty)return-59;if(stream.tty.ops.ioctl_tcgets){var termios=stream.tty.ops.ioctl_tcgets(stream);var argp=syscallGetVarargP();HEAP32[argp>>>2>>>0]=termios.c_iflag||0;HEAP32[argp+4>>>2>>>0]=termios.c_oflag||0;HEAP32[argp+8>>>2>>>0]=termios.c_cflag||0;HEAP32[argp+12>>>2>>>0]=termios.c_lflag||0;for(var i=0;i<32;i++){HEAP8[argp+i+17>>>0]=termios.c_cc[i]||0}return 0}return 0}case 21510:case 21511:case 21512:{if(!stream.tty)return-59;return 0}case 21506:case 21507:case 21508:{if(!stream.tty)return-59;if(stream.tty.ops.ioctl_tcsets){var argp=syscallGetVarargP();var c_iflag=HEAP32[argp>>>2>>>0];var c_oflag=HEAP32[argp+4>>>2>>>0];var c_cflag=HEAP32[argp+8>>>2>>>0];var c_lflag=HEAP32[argp+12>>>2>>>0];var c_cc=[];for(var i=0;i<32;i++){c_cc.push(HEAP8[argp+i+17>>>0])}return stream.tty.ops.ioctl_tcsets(stream.tty,op,{c_iflag,c_oflag,c_cflag,c_lflag,c_cc})}return 0}case 21519:{if(!stream.tty)return-59;var argp=syscallGetVarargP();HEAP32[argp>>>2>>>0]=0;return 0}case 21520:{if(!stream.tty)return-59;return-28}case 21531:{var argp=syscallGetVarargP();return FS.ioctl(stream,op,argp)}case 21523:{if(!stream.tty)return-59;if(stream.tty.ops.ioctl_tiocgwinsz){var winsize=stream.tty.ops.ioctl_tiocgwinsz(stream.tty);var argp=syscallGetVarargP();HEAP16[argp>>>1>>>0]=winsize[0];HEAP16[argp+2>>>1>>>0]=winsize[1]}return 0}case 21524:{if(!stream.tty)return-59;return 0}case 21515:{if(!stream.tty)return-59;return 0}default:return-28}}catch(e){if(typeof FS=="undefined"||!(e.name==="ErrnoError"))throw e;return-e.errno}}___syscall_ioctl.sig="iiip";function ___syscall_listen(fd,backlog){try{var sock=getSocketFromFD(fd);sock.sock_ops.listen(sock,backlog);return 0}catch(e){if(typeof FS=="undefined"||!(e.name==="ErrnoError"))throw e;return-e.errno}}___syscall_listen.sig="iiiiiii";function ___syscall_lstat64(path,buf){path>>>=0;buf>>>=0;try{path=SYSCALLS.getStr(path);return SYSCALLS.writeStat(buf,FS.lstat(path))}catch(e){if(typeof FS=="undefined"||!(e.name==="ErrnoError"))throw e;return-e.errno}}___syscall_lstat64.sig="ipp";function ___syscall_mkdirat(dirfd,path,mode){path>>>=0;try{path=SYSCALLS.getStr(path);path=SYSCALLS.calculateAt(dirfd,path);FS.mkdir(path,mode,0);return 0}catch(e){if(typeof FS=="undefined"||!(e.name==="ErrnoError"))throw e;return-e.errno}}___syscall_mkdirat.sig="iipi";function ___syscall_mknodat(dirfd,path,mode,dev){path>>>=0;try{path=SYSCALLS.getStr(path);path=SYSCALLS.calculateAt(dirfd,path);switch(mode&61440){case 32768:case 8192:case 24576:case 4096:case 49152:break;default:return-28}FS.mknod(path,mode,dev);return 0}catch(e){if(typeof FS=="undefined"||!(e.name==="ErrnoError"))throw e;return-e.errno}}___syscall_mknodat.sig="iipii";function ___syscall_newfstatat(dirfd,path,buf,flags){path>>>=0;buf>>>=0;try{path=SYSCALLS.getStr(path);var nofollow=flags&256;var allowEmpty=flags&4096;flags=flags&~6400;path=SYSCALLS.calculateAt(dirfd,path,allowEmpty);return SYSCALLS.writeStat(buf,nofollow?FS.lstat(path):FS.stat(path))}catch(e){if(typeof FS=="undefined"||!(e.name==="ErrnoError"))throw e;return-e.errno}}___syscall_newfstatat.sig="iippi";function ___syscall_openat(dirfd,path,flags,varargs){path>>>=0;varargs>>>=0;SYSCALLS.varargs=varargs;try{path=SYSCALLS.getStr(path);path=SYSCALLS.calculateAt(dirfd,path);var mode=varargs?syscallGetVarargI():0;return FS.open(path,flags,mode).fd}catch(e){if(typeof FS=="undefined"||!(e.name==="ErrnoError"))throw e;return-e.errno}}___syscall_openat.sig="iipip";var PIPEFS={BUCKET_BUFFER_SIZE:8192,mount(mount){return FS.createNode(null,"/",16384|511,0)},createPipe(){var pipe={buckets:[],refcnt:2,timestamp:new Date};pipe.buckets.push({buffer:new Uint8Array(PIPEFS.BUCKET_BUFFER_SIZE),offset:0,roffset:0});var rName=PIPEFS.nextname();var wName=PIPEFS.nextname();var rNode=FS.createNode(PIPEFS.root,rName,4096,0);var wNode=FS.createNode(PIPEFS.root,wName,4096,0);rNode.pipe=pipe;wNode.pipe=pipe;var readableStream=FS.createStream({path:rName,node:rNode,flags:0,seekable:false,stream_ops:PIPEFS.stream_ops});rNode.stream=readableStream;var writableStream=FS.createStream({path:wName,node:wNode,flags:1,seekable:false,stream_ops:PIPEFS.stream_ops});wNode.stream=writableStream;return{readable_fd:readableStream.fd,writable_fd:writableStream.fd}},stream_ops:{getattr(stream){var node=stream.node;var timestamp=node.pipe.timestamp;return{dev:14,ino:node.id,mode:4480,nlink:1,uid:0,gid:0,rdev:0,size:0,atime:timestamp,mtime:timestamp,ctime:timestamp,blksize:4096,blocks:0}},poll(stream){var pipe=stream.node.pipe;if((stream.flags&2097155)===1){return 256|4}for(var bucket of pipe.buckets){if(bucket.offset-bucket.roffset>0){return 64|1}}return 0},dup(stream){stream.node.pipe.refcnt++},ioctl(stream,request,varargs){return 28},fsync(stream){return 28},read(stream,buffer,offset,length,position){var pipe=stream.node.pipe;var currentLength=0;for(var bucket of pipe.buckets){currentLength+=bucket.offset-bucket.roffset}var data=buffer.subarray(offset,offset+length);if(length<=0){return 0}if(currentLength==0){throw new FS.ErrnoError(6)}var toRead=Math.min(currentLength,length);var totalRead=toRead;var toRemove=0;for(var bucket of pipe.buckets){var bucketSize=bucket.offset-bucket.roffset;if(toRead<=bucketSize){var tmpSlice=bucket.buffer.subarray(bucket.roffset,bucket.offset);if(toRead<bucketSize){tmpSlice=tmpSlice.subarray(0,toRead);bucket.roffset+=toRead}else{toRemove++}data.set(tmpSlice);break}else{var tmpSlice=bucket.buffer.subarray(bucket.roffset,bucket.offset);data.set(tmpSlice);data=data.subarray(tmpSlice.byteLength);toRead-=tmpSlice.byteLength;toRemove++}}if(toRemove&&toRemove==pipe.buckets.length){toRemove--;pipe.buckets[toRemove].offset=0;pipe.buckets[toRemove].roffset=0}pipe.buckets.splice(0,toRemove);return totalRead},write(stream,buffer,offset,length,position){var pipe=stream.node.pipe;var data=buffer.subarray(offset,offset+length);var dataLen=data.byteLength;if(dataLen<=0){return 0}var currBucket=null;if(pipe.buckets.length==0){currBucket={buffer:new Uint8Array(PIPEFS.BUCKET_BUFFER_SIZE),offset:0,roffset:0};pipe.buckets.push(currBucket)}else{currBucket=pipe.buckets[pipe.buckets.length-1]}assert(currBucket.offset<=PIPEFS.BUCKET_BUFFER_SIZE);var freeBytesInCurrBuffer=PIPEFS.BUCKET_BUFFER_SIZE-currBucket.offset;if(freeBytesInCurrBuffer>=dataLen){currBucket.buffer.set(data,currBucket.offset);currBucket.offset+=dataLen;return dataLen}else if(freeBytesInCurrBuffer>0){currBucket.buffer.set(data.subarray(0,freeBytesInCurrBuffer),currBucket.offset);currBucket.offset+=freeBytesInCurrBuffer;data=data.subarray(freeBytesInCurrBuffer,data.byteLength)}var numBuckets=data.byteLength/PIPEFS.BUCKET_BUFFER_SIZE|0;var remElements=data.byteLength%PIPEFS.BUCKET_BUFFER_SIZE;for(var i=0;i<numBuckets;i++){var newBucket={buffer:new Uint8Array(PIPEFS.BUCKET_BUFFER_SIZE),offset:PIPEFS.BUCKET_BUFFER_SIZE,roffset:0};pipe.buckets.push(newBucket);newBucket.buffer.set(data.subarray(0,PIPEFS.BUCKET_BUFFER_SIZE));data=data.subarray(PIPEFS.BUCKET_BUFFER_SIZE,data.byteLength)}if(remElements>0){var newBucket={buffer:new Uint8Array(PIPEFS.BUCKET_BUFFER_SIZE),offset:data.byteLength,roffset:0};pipe.buckets.push(newBucket);newBucket.buffer.set(data)}return dataLen},close(stream){var pipe=stream.node.pipe;pipe.refcnt--;if(pipe.refcnt===0){pipe.buckets=null}}},nextname(){if(!PIPEFS.nextname.current){PIPEFS.nextname.current=0}return"pipe["+PIPEFS.nextname.current+++"]"}};function ___syscall_pipe(fdPtr){fdPtr>>>=0;try{if(fdPtr==0){throw new FS.ErrnoError(21)}var res=PIPEFS.createPipe();HEAP32[fdPtr>>>2>>>0]=res.readable_fd;HEAP32[fdPtr+4>>>2>>>0]=res.writable_fd;return 0}catch(e){if(typeof FS=="undefined"||!(e.name==="ErrnoError"))throw e;return-e.errno}}___syscall_pipe.sig="ip";function ___syscall_poll(fds,nfds,timeout){fds>>>=0;try{var nonzero=0;for(var i=0;i<nfds;i++){var pollfd=fds+8*i;var fd=HEAP32[pollfd>>>2>>>0];var events=HEAP16[pollfd+4>>>1>>>0];var mask=32;var stream=FS.getStream(fd);if(stream){mask=SYSCALLS.DEFAULT_POLLMASK;if(stream.stream_ops.poll){mask=stream.stream_ops.poll(stream,-1)}}mask&=events|8|16;if(mask)nonzero++;HEAP16[pollfd+6>>>1>>>0]=mask}return nonzero}catch(e){if(typeof FS=="undefined"||!(e.name==="ErrnoError"))throw e;return-e.errno}}___syscall_poll.sig="ipii";function ___syscall_readlinkat(dirfd,path,buf,bufsize){path>>>=0;buf>>>=0;bufsize>>>=0;try{path=SYSCALLS.getStr(path);path=SYSCALLS.calculateAt(dirfd,path);if(bufsize<=0)return-28;var ret=FS.readlink(path);var len=Math.min(bufsize,lengthBytesUTF8(ret));var endChar=HEAP8[buf+len>>>0];stringToUTF8(ret,buf,bufsize+1);HEAP8[buf+len>>>0]=endChar;return len}catch(e){if(typeof FS=="undefined"||!(e.name==="ErrnoError"))throw e;return-e.errno}}___syscall_readlinkat.sig="iippp";function ___syscall_recvfrom(fd,buf,len,flags,addr,addrlen){buf>>>=0;len>>>=0;addr>>>=0;addrlen>>>=0;try{var sock=getSocketFromFD(fd);var msg=sock.sock_ops.recvmsg(sock,len);if(!msg)return 0;if(addr){var errno=writeSockaddr(addr,sock.family,DNS.lookup_name(msg.addr),msg.port,addrlen)}HEAPU8.set(msg.buffer,buf>>>0);return msg.buffer.byteLength}catch(e){if(typeof FS=="undefined"||!(e.name==="ErrnoError"))throw e;return-e.errno}}___syscall_recvfrom.sig="iippipp";function ___syscall_recvmsg(fd,message,flags,d1,d2,d3){message>>>=0;try{var sock=getSocketFromFD(fd);var iov=HEAPU32[message+8>>>2>>>0];var num=HEAP32[message+12>>>2>>>0];var total=0;for(var i=0;i<num;i++){total+=HEAP32[iov+(8*i+4)>>>2>>>0]}var msg=sock.sock_ops.recvmsg(sock,total);if(!msg)return 0;var name=HEAPU32[message>>>2>>>0];if(name){var errno=writeSockaddr(name,sock.family,DNS.lookup_name(msg.addr),msg.port)}var bytesRead=0;var bytesRemaining=msg.buffer.byteLength;for(var i=0;bytesRemaining>0&&i<num;i++){var iovbase=HEAPU32[iov+(8*i+0)>>>2>>>0];var iovlen=HEAP32[iov+(8*i+4)>>>2>>>0];if(!iovlen){continue}var length=Math.min(iovlen,bytesRemaining);var buf=msg.buffer.subarray(bytesRead,bytesRead+length);HEAPU8.set(buf,iovbase+bytesRead>>>0);bytesRead+=length;bytesRemaining-=length}return bytesRead}catch(e){if(typeof FS=="undefined"||!(e.name==="ErrnoError"))throw e;return-e.errno}}___syscall_recvmsg.sig="iipiiii";function ___syscall_renameat(olddirfd,oldpath,newdirfd,newpath){oldpath>>>=0;newpath>>>=0;try{oldpath=SYSCALLS.getStr(oldpath);newpath=SYSCALLS.getStr(newpath);oldpath=SYSCALLS.calculateAt(olddirfd,oldpath);newpath=SYSCALLS.calculateAt(newdirfd,newpath);FS.rename(oldpath,newpath);return 0}catch(e){if(typeof FS=="undefined"||!(e.name==="ErrnoError"))throw e;return-e.errno}}___syscall_renameat.sig="iipip";function ___syscall_rmdir(path){path>>>=0;try{path=SYSCALLS.getStr(path);FS.rmdir(path);return 0}catch(e){if(typeof FS=="undefined"||!(e.name==="ErrnoError"))throw e;return-e.errno}}___syscall_rmdir.sig="ip";function ___syscall_sendmsg(fd,message,flags,d1,d2,d3){message>>>=0;d1>>>=0;d2>>>=0;try{var sock=getSocketFromFD(fd);var iov=HEAPU32[message+8>>>2>>>0];var num=HEAP32[message+12>>>2>>>0];var addr,port;var name=HEAPU32[message>>>2>>>0];var namelen=HEAP32[message+4>>>2>>>0];if(name){var info=getSocketAddress(name,namelen);port=info.port;addr=info.addr}var total=0;for(var i=0;i<num;i++){total+=HEAP32[iov+(8*i+4)>>>2>>>0]}var view=new Uint8Array(total);var offset=0;for(var i=0;i<num;i++){var iovbase=HEAPU32[iov+(8*i+0)>>>2>>>0];var iovlen=HEAP32[iov+(8*i+4)>>>2>>>0];for(var j=0;j<iovlen;j++){view[offset++]=HEAP8[iovbase+j>>>0]}}return sock.sock_ops.sendmsg(sock,view,0,total,addr,port)}catch(e){if(typeof FS=="undefined"||!(e.name==="ErrnoError"))throw e;return-e.errno}}___syscall_sendmsg.sig="iipippi";function ___syscall_sendto(fd,message,length,flags,addr,addr_len){message>>>=0;length>>>=0;addr>>>=0;addr_len>>>=0;try{var sock=getSocketFromFD(fd);if(!addr){return FS.write(sock.stream,HEAP8,message,length)}var dest=getSocketAddress(addr,addr_len);return sock.sock_ops.sendmsg(sock,HEAP8,message,length,dest.addr,dest.port)}catch(e){if(typeof FS=="undefined"||!(e.name==="ErrnoError"))throw e;return-e.errno}}___syscall_sendto.sig="iippipp";function ___syscall_socket(domain,type,protocol){try{var sock=SOCKFS.createSocket(domain,type,protocol);return sock.stream.fd}catch(e){if(typeof FS=="undefined"||!(e.name==="ErrnoError"))throw e;return-e.errno}}___syscall_socket.sig="iiiiiii";function ___syscall_stat64(path,buf){path>>>=0;buf>>>=0;try{path=SYSCALLS.getStr(path);return SYSCALLS.writeStat(buf,FS.stat(path))}catch(e){if(typeof FS=="undefined"||!(e.name==="ErrnoError"))throw e;return-e.errno}}___syscall_stat64.sig="ipp";function ___syscall_statfs64(path,size,buf){path>>>=0;size>>>=0;buf>>>=0;try{SYSCALLS.writeStatFs(buf,FS.statfs(SYSCALLS.getStr(path)));return 0}catch(e){if(typeof FS=="undefined"||!(e.name==="ErrnoError"))throw e;return-e.errno}}___syscall_statfs64.sig="ippp";function ___syscall_symlinkat(target,dirfd,linkpath){target>>>=0;linkpath>>>=0;try{target=SYSCALLS.getStr(target);linkpath=SYSCALLS.getStr(linkpath);linkpath=SYSCALLS.calculateAt(dirfd,linkpath);FS.symlink(target,linkpath);return 0}catch(e){if(typeof FS=="undefined"||!(e.name==="ErrnoError"))throw e;return-e.errno}}___syscall_symlinkat.sig="ipip";function ___syscall_truncate64(path,length){path>>>=0;length=bigintToI53Checked(length);try{if(isNaN(length))return-61;path=SYSCALLS.getStr(path);FS.truncate(path,length);return 0}catch(e){if(typeof FS=="undefined"||!(e.name==="ErrnoError"))throw e;return-e.errno}}___syscall_truncate64.sig="ipj";function ___syscall_unlinkat(dirfd,path,flags){path>>>=0;try{path=SYSCALLS.getStr(path);path=SYSCALLS.calculateAt(dirfd,path);if(!flags){FS.unlink(path)}else if(flags===512){FS.rmdir(path)}else{return-28}return 0}catch(e){if(typeof FS=="undefined"||!(e.name==="ErrnoError"))throw e;return-e.errno}}___syscall_unlinkat.sig="iipi";var readI53FromI64=ptr=>HEAPU32[ptr>>>2>>>0]+HEAP32[ptr+4>>>2>>>0]*4294967296;function ___syscall_utimensat(dirfd,path,times,flags){path>>>=0;times>>>=0;try{path=SYSCALLS.getStr(path);path=SYSCALLS.calculateAt(dirfd,path,true);var now=Date.now(),atime,mtime;if(!times){atime=now;mtime=now}else{var seconds=readI53FromI64(times);var nanoseconds=HEAP32[times+8>>>2>>>0];if(nanoseconds==1073741823){atime=now}else if(nanoseconds==1073741822){atime=null}else{atime=seconds*1e3+nanoseconds/(1e3*1e3)}times+=16;seconds=readI53FromI64(times);nanoseconds=HEAP32[times+8>>>2>>>0];if(nanoseconds==1073741823){mtime=now}else if(nanoseconds==1073741822){mtime=null}else{mtime=seconds*1e3+nanoseconds/(1e3*1e3)}}if((mtime??atime)!==null){FS.utime(path,atime,mtime)}return 0}catch(e){if(typeof FS=="undefined"||!(e.name==="ErrnoError"))throw e;return-e.errno}}___syscall_utimensat.sig="iippi";var ___table_base=new WebAssembly.Global({value:"i32",mutable:false},1);var __abort_js=()=>abort("");__abort_js.sig="v";var dlSetError=msg=>{var sp=stackSave();var cmsg=stringToUTF8OnStack(msg);___dl_seterr(cmsg,0);stackRestore(sp)};var dlopenInternal=(handle,jsflags)=>{var filename=UTF8ToString(handle+36);var flags=HEAP32[handle+4>>>2>>>0];filename=PATH.normalize(filename);var searchpaths=[];var global=Boolean(flags&256);var localScope=global?null:{};var combinedFlags={global,nodelete:Boolean(flags&4096),loadAsync:jsflags.loadAsync};if(jsflags.loadAsync){return loadDynamicLibrary(filename,combinedFlags,localScope,handle)}try{return loadDynamicLibrary(filename,combinedFlags,localScope,handle)}catch(e){dlSetError(`Could not load dynamic lib: ${filename}\n${e}`);return 0}};function __dlopen_js(handle){handle>>>=0;return dlopenInternal(handle,{loadAsync:false})}__dlopen_js.sig="pp";function __dlsym_js(handle,symbol,symbolIndex){handle>>>=0;symbol>>>=0;symbolIndex>>>=0;symbol=UTF8ToString(symbol);var result;var newSymIndex;var lib=LDSO.loadedLibsByHandle[handle];if(!lib.exports.hasOwnProperty(symbol)||lib.exports[symbol].stub){dlSetError(`Tried to lookup unknown symbol "${symbol}" in dynamic lib: ${lib.name}`);return 0}newSymIndex=Object.keys(lib.exports).indexOf(symbol);result=lib.exports[symbol];if(typeof result=="function"){var addr=getFunctionAddress(result);if(addr){result=addr}else{result=addFunction(result,result.sig);HEAPU32[symbolIndex>>>2>>>0]=newSymIndex}}return result}__dlsym_js.sig="pppp";var handleException=e=>{if(e instanceof ExitStatus||e=="unwind"){return EXITSTATUS}quit_(1,e)};var runtimeKeepaliveCounter=0;var keepRuntimeAlive=()=>noExitRuntime||runtimeKeepaliveCounter>0;var _proc_exit=code=>{EXITSTATUS=code;if(!keepRuntimeAlive()){Module["onExit"]?.(code);ABORT=true}quit_(code,new ExitStatus(code))};_proc_exit.sig="vi";var exitJS=(status,implicit)=>{EXITSTATUS=status;if(!keepRuntimeAlive()){exitRuntime()}_proc_exit(status)};var _exit=exitJS;_exit.sig="vi";var maybeExit=()=>{if(runtimeExited){return}if(!keepRuntimeAlive()){try{_exit(EXITSTATUS)}catch(e){handleException(e)}}};var callUserCallback=func=>{if(runtimeExited||ABORT){return}try{func();maybeExit()}catch(e){handleException(e)}};var runtimeKeepalivePush=()=>{runtimeKeepaliveCounter+=1};runtimeKeepalivePush.sig="v";var runtimeKeepalivePop=()=>{runtimeKeepaliveCounter-=1};runtimeKeepalivePop.sig="v";function __emscripten_dlopen_js(handle,onsuccess,onerror,user_data){handle>>>=0;onsuccess>>>=0;onerror>>>=0;user_data>>>=0;function errorCallback(e){var filename=UTF8ToString(handle+36);dlSetError(`'Could not load dynamic lib: ${filename}\n${e}`);runtimeKeepalivePop();callUserCallback(()=>getWasmTableEntry(onerror)(handle,user_data))}function successCallback(){runtimeKeepalivePop();callUserCallback(()=>getWasmTableEntry(onsuccess)(handle,user_data))}runtimeKeepalivePush();var promise=dlopenInternal(handle,{loadAsync:true});if(promise){promise.then(successCallback,errorCallback)}else{errorCallback()}}__emscripten_dlopen_js.sig="vpppp";var getExecutableName=()=>thisProgram||"./this.program";function __emscripten_get_progname(str,len){str>>>=0;return stringToUTF8(getExecutableName(),str,len)}__emscripten_get_progname.sig="vpi";function __emscripten_lookup_name(name){name>>>=0;var nameString=UTF8ToString(name);return inetPton4(DNS.lookup_name(nameString))}__emscripten_lookup_name.sig="ip";var __emscripten_runtime_keepalive_clear=()=>{noExitRuntime=false;runtimeKeepaliveCounter=0};__emscripten_runtime_keepalive_clear.sig="v";function __emscripten_system(command){command>>>=0;if(ENVIRONMENT_IS_NODE){if(!command)return 1;var cmdstr=UTF8ToString(command);if(!cmdstr.length)return 0;var cp=require("child_process");var ret=cp.spawnSync(cmdstr,[],{shell:true,stdio:"inherit"});var _W_EXITCODE=(ret,sig)=>ret<<8|sig;if(ret.status===null){var signalToNumber=sig=>{switch(sig){case"SIGHUP":return 1;case"SIGQUIT":return 3;case"SIGFPE":return 8;case"SIGKILL":return 9;case"SIGALRM":return 14;case"SIGTERM":return 15;default:return 2}};return _W_EXITCODE(0,signalToNumber(ret.signal))}return _W_EXITCODE(ret.status,0)}if(!command)return 0;return-52}__emscripten_system.sig="ip";function __gmtime_js(time,tmPtr){time=bigintToI53Checked(time);tmPtr>>>=0;var date=new Date(time*1e3);HEAP32[tmPtr>>>2>>>0]=date.getUTCSeconds();HEAP32[tmPtr+4>>>2>>>0]=date.getUTCMinutes();HEAP32[tmPtr+8>>>2>>>0]=date.getUTCHours();HEAP32[tmPtr+12>>>2>>>0]=date.getUTCDate();HEAP32[tmPtr+16>>>2>>>0]=date.getUTCMonth();HEAP32[tmPtr+20>>>2>>>0]=date.getUTCFullYear()-1900;HEAP32[tmPtr+24>>>2>>>0]=date.getUTCDay();var start=Date.UTC(date.getUTCFullYear(),0,1,0,0,0,0);var yday=(date.getTime()-start)/(1e3*60*60*24)|0;HEAP32[tmPtr+28>>>2>>>0]=yday}__gmtime_js.sig="vjp";var isLeapYear=year=>year%4===0&&(year%100!==0||year%400===0);var MONTH_DAYS_LEAP_CUMULATIVE=[0,31,60,91,121,152,182,213,244,274,305,335];var MONTH_DAYS_REGULAR_CUMULATIVE=[0,31,59,90,120,151,181,212,243,273,304,334];var ydayFromDate=date=>{var leap=isLeapYear(date.getFullYear());var monthDaysCumulative=leap?MONTH_DAYS_LEAP_CUMULATIVE:MONTH_DAYS_REGULAR_CUMULATIVE;var yday=monthDaysCumulative[date.getMonth()]+date.getDate()-1;return yday};function __localtime_js(time,tmPtr){time=bigintToI53Checked(time);tmPtr>>>=0;var date=new Date(time*1e3);HEAP32[tmPtr>>>2>>>0]=date.getSeconds();HEAP32[tmPtr+4>>>2>>>0]=date.getMinutes();HEAP32[tmPtr+8>>>2>>>0]=date.getHours();HEAP32[tmPtr+12>>>2>>>0]=date.getDate();HEAP32[tmPtr+16>>>2>>>0]=date.getMonth();HEAP32[tmPtr+20>>>2>>>0]=date.getFullYear()-1900;HEAP32[tmPtr+24>>>2>>>0]=date.getDay();var yday=ydayFromDate(date)|0;HEAP32[tmPtr+28>>>2>>>0]=yday;HEAP32[tmPtr+36>>>2>>>0]=-(date.getTimezoneOffset()*60);var start=new Date(date.getFullYear(),0,1);var summerOffset=new Date(date.getFullYear(),6,1).getTimezoneOffset();var winterOffset=start.getTimezoneOffset();var dst=(summerOffset!=winterOffset&&date.getTimezoneOffset()==Math.min(winterOffset,summerOffset))|0;HEAP32[tmPtr+32>>>2>>>0]=dst}__localtime_js.sig="vjp";var __mktime_js=function(tmPtr){tmPtr>>>=0;var ret=(()=>{var date=new Date(HEAP32[tmPtr+20>>>2>>>0]+1900,HEAP32[tmPtr+16>>>2>>>0],HEAP32[tmPtr+12>>>2>>>0],HEAP32[tmPtr+8>>>2>>>0],HEAP32[tmPtr+4>>>2>>>0],HEAP32[tmPtr>>>2>>>0],0);var dst=HEAP32[tmPtr+32>>>2>>>0];var guessedOffset=date.getTimezoneOffset();var start=new Date(date.getFullYear(),0,1);var summerOffset=new Date(date.getFullYear(),6,1).getTimezoneOffset();var winterOffset=start.getTimezoneOffset();var dstOffset=Math.min(winterOffset,summerOffset);if(dst<0){HEAP32[tmPtr+32>>>2>>>0]=Number(summerOffset!=winterOffset&&dstOffset==guessedOffset)}else if(dst>0!=(dstOffset==guessedOffset)){var nonDstOffset=Math.max(winterOffset,summerOffset);var trueOffset=dst>0?dstOffset:nonDstOffset;date.setTime(date.getTime()+(trueOffset-guessedOffset)*6e4)}HEAP32[tmPtr+24>>>2>>>0]=date.getDay();var yday=ydayFromDate(date)|0;HEAP32[tmPtr+28>>>2>>>0]=yday;HEAP32[tmPtr>>>2>>>0]=date.getSeconds();HEAP32[tmPtr+4>>>2>>>0]=date.getMinutes();HEAP32[tmPtr+8>>>2>>>0]=date.getHours();HEAP32[tmPtr+12>>>2>>>0]=date.getDate();HEAP32[tmPtr+16>>>2>>>0]=date.getMonth();HEAP32[tmPtr+20>>>2>>>0]=date.getYear();var timeMs=date.getTime();if(isNaN(timeMs)){return-1}return timeMs/1e3})();return BigInt(ret)};__mktime_js.sig="jp";function __mmap_js(len,prot,flags,fd,offset,allocated,addr){len>>>=0;offset=bigintToI53Checked(offset);allocated>>>=0;addr>>>=0;try{var stream=SYSCALLS.getStreamFromFD(fd);var res=FS.mmap(stream,len,offset,prot,flags);var ptr=res.ptr;HEAP32[allocated>>>2>>>0]=res.allocated;HEAPU32[addr>>>2>>>0]=ptr;return 0}catch(e){if(typeof FS=="undefined"||!(e.name==="ErrnoError"))throw e;return-e.errno}}__mmap_js.sig="ipiiijpp";function __msync_js(addr,len,prot,flags,fd,offset){addr>>>=0;len>>>=0;offset=bigintToI53Checked(offset);try{if(isNaN(offset))return-61;SYSCALLS.doMsync(addr,SYSCALLS.getStreamFromFD(fd),len,flags,offset);return 0}catch(e){if(typeof FS=="undefined"||!(e.name==="ErrnoError"))throw e;return-e.errno}}__msync_js.sig="ippiiij";function __munmap_js(addr,len,prot,flags,fd,offset){addr>>>=0;len>>>=0;offset=bigintToI53Checked(offset);try{var stream=SYSCALLS.getStreamFromFD(fd);if(prot&2){SYSCALLS.doMsync(addr,stream,len,flags,offset)}}catch(e){if(typeof FS=="undefined"||!(e.name==="ErrnoError"))throw e;return-e.errno}}__munmap_js.sig="ippiiij";var timers={};var _emscripten_get_now=()=>performance.now();_emscripten_get_now.sig="d";var __setitimer_js=(which,timeout_ms)=>{if(timers[which]){clearTimeout(timers[which].id);delete timers[which]}if(!timeout_ms)return 0;var id=setTimeout(()=>{delete timers[which];callUserCallback(()=>__emscripten_timeout(which,_emscripten_get_now()))},timeout_ms);timers[which]={id,timeout_ms};return 0};__setitimer_js.sig="iid";var __timegm_js=function(tmPtr){tmPtr>>>=0;var ret=(()=>{var time=Date.UTC(HEAP32[tmPtr+20>>>2>>>0]+1900,HEAP32[tmPtr+16>>>2>>>0],HEAP32[tmPtr+12>>>2>>>0],HEAP32[tmPtr+8>>>2>>>0],HEAP32[tmPtr+4>>>2>>>0],HEAP32[tmPtr>>>2>>>0],0);var date=new Date(time);HEAP32[tmPtr+24>>>2>>>0]=date.getUTCDay();var start=Date.UTC(date.getUTCFullYear(),0,1,0,0,0,0);var yday=(date.getTime()-start)/(1e3*60*60*24)|0;HEAP32[tmPtr+28>>>2>>>0]=yday;return date.getTime()/1e3})();return BigInt(ret)};__timegm_js.sig="jp";var __tzset_js=function(timezone,daylight,std_name,dst_name){timezone>>>=0;daylight>>>=0;std_name>>>=0;dst_name>>>=0;var currentYear=(new Date).getFullYear();var winter=new Date(currentYear,0,1);var summer=new Date(currentYear,6,1);var winterOffset=winter.getTimezoneOffset();var summerOffset=summer.getTimezoneOffset();var stdTimezoneOffset=Math.max(winterOffset,summerOffset);HEAPU32[timezone>>>2>>>0]=stdTimezoneOffset*60;HEAP32[daylight>>>2>>>0]=Number(winterOffset!=summerOffset);var extractZone=timezoneOffset=>{var sign=timezoneOffset>=0?"-":"+";var absOffset=Math.abs(timezoneOffset);var hours=String(Math.floor(absOffset/60)).padStart(2,"0");var minutes=String(absOffset%60).padStart(2,"0");return`UTC${sign}${hours}${minutes}`};var winterName=extractZone(winterOffset);var summerName=extractZone(summerOffset);if(summerOffset<winterOffset){stringToUTF8(winterName,std_name,17);stringToUTF8(summerName,dst_name,17)}else{stringToUTF8(winterName,dst_name,17);stringToUTF8(summerName,std_name,17)}};__tzset_js.sig="vpppp";var _emscripten_get_now_res=()=>{if(ENVIRONMENT_IS_NODE){return 1}return 1e3};_emscripten_get_now_res.sig="d";var nowIsMonotonic=1;var checkWasiClock=clock_id=>clock_id>=0&&clock_id<=3;function _clock_res_get(clk_id,pres){pres>>>=0;if(!checkWasiClock(clk_id)){return 28}var nsec;if(clk_id===0){nsec=1e3*1e3}else if(nowIsMonotonic){nsec=_emscripten_get_now_res()}else{return 52}HEAP64[pres>>>3>>>0]=BigInt(nsec);return 0}_clock_res_get.sig="iip";var _emscripten_date_now=()=>Date.now();_emscripten_date_now.sig="d";function _clock_time_get(clk_id,ignored_precision,ptime){ignored_precision=bigintToI53Checked(ignored_precision);ptime>>>=0;if(!checkWasiClock(clk_id)){return 28}var now;if(clk_id===0){now=_emscripten_date_now()}else if(nowIsMonotonic){now=_emscripten_get_now()}else{return 52}var nsec=Math.round(now*1e3*1e3);HEAP64[ptime>>>3>>>0]=BigInt(nsec);return 0}_clock_time_get.sig="iijp";function _create_sentinel(...args){return wasmImports["create_sentinel"](...args)}_create_sentinel.stub=true;var readEmAsmArgsArray=[];var readEmAsmArgs=(sigPtr,buf)=>{readEmAsmArgsArray.length=0;var ch;while(ch=HEAPU8[sigPtr++>>>0]){var wide=ch!=105;wide&=ch!=112;buf+=wide&&buf%8?4:0;readEmAsmArgsArray.push(ch==112?HEAPU32[buf>>>2>>>0]:ch==106?HEAP64[buf>>>3>>>0]:ch==105?HEAP32[buf>>>2>>>0]:HEAPF64[buf>>>3>>>0]);buf+=wide?8:4}return readEmAsmArgsArray};var runEmAsmFunction=(code,sigPtr,argbuf)=>{var args=readEmAsmArgs(sigPtr,argbuf);return ASM_CONSTS[code](...args)};function _emscripten_asm_const_int(code,sigPtr,argbuf){code>>>=0;sigPtr>>>=0;argbuf>>>=0;return runEmAsmFunction(code,sigPtr,argbuf)}_emscripten_asm_const_int.sig="ippp";function _emscripten_console_error(str){str>>>=0;console.error(UTF8ToString(str))}_emscripten_console_error.sig="vp";function _emscripten_console_log(str){str>>>=0;console.log(UTF8ToString(str))}_emscripten_console_log.sig="vp";function _emscripten_console_trace(str){str>>>=0;console.trace(UTF8ToString(str))}_emscripten_console_trace.sig="vp";function _emscripten_console_warn(str){str>>>=0;console.warn(UTF8ToString(str))}_emscripten_console_warn.sig="vp";function _emscripten_err(str){str>>>=0;return err(UTF8ToString(str))}_emscripten_err.sig="vp";var getHeapMax=()=>4294901760;function _emscripten_get_heap_max(){return getHeapMax()}_emscripten_get_heap_max.sig="p";var GLctx;var webgl_enable_ANGLE_instanced_arrays=ctx=>{var ext=ctx.getExtension("ANGLE_instanced_arrays");if(ext){ctx["vertexAttribDivisor"]=(index,divisor)=>ext["vertexAttribDivisorANGLE"](index,divisor);ctx["drawArraysInstanced"]=(mode,first,count,primcount)=>ext["drawArraysInstancedANGLE"](mode,first,count,primcount);ctx["drawElementsInstanced"]=(mode,count,type,indices,primcount)=>ext["drawElementsInstancedANGLE"](mode,count,type,indices,primcount);return 1}};var webgl_enable_OES_vertex_array_object=ctx=>{var ext=ctx.getExtension("OES_vertex_array_object");if(ext){ctx["createVertexArray"]=()=>ext["createVertexArrayOES"]();ctx["deleteVertexArray"]=vao=>ext["deleteVertexArrayOES"](vao);ctx["bindVertexArray"]=vao=>ext["bindVertexArrayOES"](vao);ctx["isVertexArray"]=vao=>ext["isVertexArrayOES"](vao);return 1}};var webgl_enable_WEBGL_draw_buffers=ctx=>{var ext=ctx.getExtension("WEBGL_draw_buffers");if(ext){ctx["drawBuffers"]=(n,bufs)=>ext["drawBuffersWEBGL"](n,bufs);return 1}};var webgl_enable_WEBGL_draw_instanced_base_vertex_base_instance=ctx=>!!(ctx.dibvbi=ctx.getExtension("WEBGL_draw_instanced_base_vertex_base_instance"));var webgl_enable_WEBGL_multi_draw_instanced_base_vertex_base_instance=ctx=>!!(ctx.mdibvbi=ctx.getExtension("WEBGL_multi_draw_instanced_base_vertex_base_instance"));var webgl_enable_EXT_polygon_offset_clamp=ctx=>!!(ctx.extPolygonOffsetClamp=ctx.getExtension("EXT_polygon_offset_clamp"));var webgl_enable_EXT_clip_control=ctx=>!!(ctx.extClipControl=ctx.getExtension("EXT_clip_control"));var webgl_enable_WEBGL_polygon_mode=ctx=>!!(ctx.webglPolygonMode=ctx.getExtension("WEBGL_polygon_mode"));var webgl_enable_WEBGL_multi_draw=ctx=>!!(ctx.multiDrawWebgl=ctx.getExtension("WEBGL_multi_draw"));var getEmscriptenSupportedExtensions=ctx=>{var supportedExtensions=["ANGLE_instanced_arrays","EXT_blend_minmax","EXT_disjoint_timer_query","EXT_frag_depth","EXT_shader_texture_lod","EXT_sRGB","OES_element_index_uint","OES_fbo_render_mipmap","OES_standard_derivatives","OES_texture_float","OES_texture_half_float","OES_texture_half_float_linear","OES_vertex_array_object","WEBGL_color_buffer_float","WEBGL_depth_texture","WEBGL_draw_buffers","EXT_color_buffer_float","EXT_conservative_depth","EXT_disjoint_timer_query_webgl2","EXT_texture_norm16","NV_shader_noperspective_interpolation","WEBGL_clip_cull_distance","EXT_clip_control","EXT_color_buffer_half_float","EXT_depth_clamp","EXT_float_blend","EXT_polygon_offset_clamp","EXT_texture_compression_bptc","EXT_texture_compression_rgtc","EXT_texture_filter_anisotropic","KHR_parallel_shader_compile","OES_texture_float_linear","WEBGL_blend_func_extended","WEBGL_compressed_texture_astc","WEBGL_compressed_texture_etc","WEBGL_compressed_texture_etc1","WEBGL_compressed_texture_s3tc","WEBGL_compressed_texture_s3tc_srgb","WEBGL_debug_renderer_info","WEBGL_debug_shaders","WEBGL_lose_context","WEBGL_multi_draw","WEBGL_polygon_mode"];return(ctx.getSupportedExtensions()||[]).filter(ext=>supportedExtensions.includes(ext))};var GL={counter:1,buffers:[],programs:[],framebuffers:[],renderbuffers:[],textures:[],shaders:[],vaos:[],contexts:[],offscreenCanvases:{},queries:[],samplers:[],transformFeedbacks:[],syncs:[],stringCache:{},stringiCache:{},unpackAlignment:4,unpackRowLength:0,recordError:errorCode=>{if(!GL.lastError){GL.lastError=errorCode}},getNewId:table=>{var ret=GL.counter++;for(var i=table.length;i<ret;i++){table[i]=null}return ret},genObject:(n,buffers,createFunction,objectTable)=>{for(var i=0;i<n;i++){var buffer=GLctx[createFunction]();var id=buffer&&GL.getNewId(objectTable);if(buffer){buffer.name=id;objectTable[id]=buffer}else{GL.recordError(1282)}HEAP32[buffers+i*4>>>2>>>0]=id}},getSource:(shader,count,string,length)=>{var source="";for(var i=0;i<count;++i){var len=length?HEAPU32[length+i*4>>>2>>>0]:undefined;source+=UTF8ToString(HEAPU32[string+i*4>>>2>>>0],len)}return source},createContext:(canvas,webGLContextAttributes)=>{var ctx=webGLContextAttributes.majorVersion>1?canvas.getContext("webgl2",webGLContextAttributes):canvas.getContext("webgl",webGLContextAttributes);if(!ctx)return 0;var handle=GL.registerContext(ctx,webGLContextAttributes);return handle},registerContext:(ctx,webGLContextAttributes)=>{var handle=GL.getNewId(GL.contexts);var context={handle,attributes:webGLContextAttributes,version:webGLContextAttributes.majorVersion,GLctx:ctx};if(ctx.canvas)ctx.canvas.GLctxObject=context;GL.contexts[handle]=context;if(typeof webGLContextAttributes.enableExtensionsByDefault=="undefined"||webGLContextAttributes.enableExtensionsByDefault){GL.initExtensions(context)}return handle},makeContextCurrent:contextHandle=>{GL.currentContext=GL.contexts[contextHandle];Module["ctx"]=GLctx=GL.currentContext?.GLctx;return!(contextHandle&&!GLctx)},getContext:contextHandle=>GL.contexts[contextHandle],deleteContext:contextHandle=>{if(GL.currentContext===GL.contexts[contextHandle]){GL.currentContext=null}if(typeof JSEvents=="object"){JSEvents.removeAllHandlersOnTarget(GL.contexts[contextHandle].GLctx.canvas)}if(GL.contexts[contextHandle]?.GLctx.canvas){GL.contexts[contextHandle].GLctx.canvas.GLctxObject=undefined}GL.contexts[contextHandle]=null},initExtensions:context=>{context||=GL.currentContext;if(context.initExtensionsDone)return;context.initExtensionsDone=true;var GLctx=context.GLctx;webgl_enable_WEBGL_multi_draw(GLctx);webgl_enable_EXT_polygon_offset_clamp(GLctx);webgl_enable_EXT_clip_control(GLctx);webgl_enable_WEBGL_polygon_mode(GLctx);webgl_enable_ANGLE_instanced_arrays(GLctx);webgl_enable_OES_vertex_array_object(GLctx);webgl_enable_WEBGL_draw_buffers(GLctx);webgl_enable_WEBGL_draw_instanced_base_vertex_base_instance(GLctx);webgl_enable_WEBGL_multi_draw_instanced_base_vertex_base_instance(GLctx);if(context.version>=2){GLctx.disjointTimerQueryExt=GLctx.getExtension("EXT_disjoint_timer_query_webgl2")}if(context.version<2||!GLctx.disjointTimerQueryExt){GLctx.disjointTimerQueryExt=GLctx.getExtension("EXT_disjoint_timer_query")}getEmscriptenSupportedExtensions(GLctx).forEach(ext=>{if(!ext.includes("lose_context")&&!ext.includes("debug")){GLctx.getExtension(ext)}})}};var _glActiveTexture=x0=>GLctx.activeTexture(x0);_glActiveTexture.sig="vi";var _emscripten_glActiveTexture=_glActiveTexture;_emscripten_glActiveTexture.sig="vi";var _glAttachShader=(program,shader)=>{GLctx.attachShader(GL.programs[program],GL.shaders[shader])};_glAttachShader.sig="vii";var _emscripten_glAttachShader=_glAttachShader;_emscripten_glAttachShader.sig="vii";var _glBeginQuery=(target,id)=>{GLctx.beginQuery(target,GL.queries[id])};_glBeginQuery.sig="vii";var _emscripten_glBeginQuery=_glBeginQuery;_emscripten_glBeginQuery.sig="vii";var _glBeginQueryEXT=(target,id)=>{GLctx.disjointTimerQueryExt["beginQueryEXT"](target,GL.queries[id])};_glBeginQueryEXT.sig="vii";var _emscripten_glBeginQueryEXT=_glBeginQueryEXT;var _glBeginTransformFeedback=x0=>GLctx.beginTransformFeedback(x0);_glBeginTransformFeedback.sig="vi";var _emscripten_glBeginTransformFeedback=_glBeginTransformFeedback;_emscripten_glBeginTransformFeedback.sig="vi";function _glBindAttribLocation(program,index,name){name>>>=0;GLctx.bindAttribLocation(GL.programs[program],index,UTF8ToString(name))}_glBindAttribLocation.sig="viip";var _emscripten_glBindAttribLocation=_glBindAttribLocation;_emscripten_glBindAttribLocation.sig="viip";var _glBindBuffer=(target,buffer)=>{if(target==35051){GLctx.currentPixelPackBufferBinding=buffer}else if(target==35052){GLctx.currentPixelUnpackBufferBinding=buffer}GLctx.bindBuffer(target,GL.buffers[buffer])};_glBindBuffer.sig="vii";var _emscripten_glBindBuffer=_glBindBuffer;_emscripten_glBindBuffer.sig="vii";var _glBindBufferBase=(target,index,buffer)=>{GLctx.bindBufferBase(target,index,GL.buffers[buffer])};_glBindBufferBase.sig="viii";var _emscripten_glBindBufferBase=_glBindBufferBase;_emscripten_glBindBufferBase.sig="viii";function _glBindBufferRange(target,index,buffer,offset,ptrsize){offset>>>=0;ptrsize>>>=0;GLctx.bindBufferRange(target,index,GL.buffers[buffer],offset,ptrsize)}_glBindBufferRange.sig="viiipp";var _emscripten_glBindBufferRange=_glBindBufferRange;_emscripten_glBindBufferRange.sig="viiipp";var _glBindFramebuffer=(target,framebuffer)=>{GLctx.bindFramebuffer(target,GL.framebuffers[framebuffer])};_glBindFramebuffer.sig="vii";var _emscripten_glBindFramebuffer=_glBindFramebuffer;_emscripten_glBindFramebuffer.sig="vii";var _glBindRenderbuffer=(target,renderbuffer)=>{GLctx.bindRenderbuffer(target,GL.renderbuffers[renderbuffer])};_glBindRenderbuffer.sig="vii";var _emscripten_glBindRenderbuffer=_glBindRenderbuffer;_emscripten_glBindRenderbuffer.sig="vii";var _glBindSampler=(unit,sampler)=>{GLctx.bindSampler(unit,GL.samplers[sampler])};_glBindSampler.sig="vii";var _emscripten_glBindSampler=_glBindSampler;_emscripten_glBindSampler.sig="vii";var _glBindTexture=(target,texture)=>{GLctx.bindTexture(target,GL.textures[texture])};_glBindTexture.sig="vii";var _emscripten_glBindTexture=_glBindTexture;_emscripten_glBindTexture.sig="vii";var _glBindTransformFeedback=(target,id)=>{GLctx.bindTransformFeedback(target,GL.transformFeedbacks[id])};_glBindTransformFeedback.sig="vii";var _emscripten_glBindTransformFeedback=_glBindTransformFeedback;_emscripten_glBindTransformFeedback.sig="vii";var _glBindVertexArray=vao=>{GLctx.bindVertexArray(GL.vaos[vao])};_glBindVertexArray.sig="vi";var _emscripten_glBindVertexArray=_glBindVertexArray;_emscripten_glBindVertexArray.sig="vi";var _glBindVertexArrayOES=_glBindVertexArray;_glBindVertexArrayOES.sig="vi";var _emscripten_glBindVertexArrayOES=_glBindVertexArrayOES;_emscripten_glBindVertexArrayOES.sig="vi";var _glBlendColor=(x0,x1,x2,x3)=>GLctx.blendColor(x0,x1,x2,x3);_glBlendColor.sig="vffff";var _emscripten_glBlendColor=_glBlendColor;_emscripten_glBlendColor.sig="vffff";var _glBlendEquation=x0=>GLctx.blendEquation(x0);_glBlendEquation.sig="vi";var _emscripten_glBlendEquation=_glBlendEquation;_emscripten_glBlendEquation.sig="vi";var _glBlendEquationSeparate=(x0,x1)=>GLctx.blendEquationSeparate(x0,x1);_glBlendEquationSeparate.sig="vii";var _emscripten_glBlendEquationSeparate=_glBlendEquationSeparate;_emscripten_glBlendEquationSeparate.sig="vii";var _glBlendFunc=(x0,x1)=>GLctx.blendFunc(x0,x1);_glBlendFunc.sig="vii";var _emscripten_glBlendFunc=_glBlendFunc;_emscripten_glBlendFunc.sig="vii";var _glBlendFuncSeparate=(x0,x1,x2,x3)=>GLctx.blendFuncSeparate(x0,x1,x2,x3);_glBlendFuncSeparate.sig="viiii";var _emscripten_glBlendFuncSeparate=_glBlendFuncSeparate;_emscripten_glBlendFuncSeparate.sig="viiii";var _glBlitFramebuffer=(x0,x1,x2,x3,x4,x5,x6,x7,x8,x9)=>GLctx.blitFramebuffer(x0,x1,x2,x3,x4,x5,x6,x7,x8,x9);_glBlitFramebuffer.sig="viiiiiiiiii";var _emscripten_glBlitFramebuffer=_glBlitFramebuffer;_emscripten_glBlitFramebuffer.sig="viiiiiiiiii";function _glBufferData(target,size,data,usage){size>>>=0;data>>>=0;GLctx.bufferData(target,data?HEAPU8.subarray(data>>>0,data+size>>>0):size,usage)}_glBufferData.sig="vippi";var _emscripten_glBufferData=_glBufferData;_emscripten_glBufferData.sig="vippi";function _glBufferSubData(target,offset,size,data){offset>>>=0;size>>>=0;data>>>=0;GLctx.bufferSubData(target,offset,HEAPU8.subarray(data>>>0,data+size>>>0))}_glBufferSubData.sig="vippp";var _emscripten_glBufferSubData=_glBufferSubData;_emscripten_glBufferSubData.sig="vippp";var _glCheckFramebufferStatus=x0=>GLctx.checkFramebufferStatus(x0);_glCheckFramebufferStatus.sig="ii";var _emscripten_glCheckFramebufferStatus=_glCheckFramebufferStatus;_emscripten_glCheckFramebufferStatus.sig="ii";var _glClear=x0=>GLctx.clear(x0);_glClear.sig="vi";var _emscripten_glClear=_glClear;_emscripten_glClear.sig="vi";var _glClearBufferfi=(x0,x1,x2,x3)=>GLctx.clearBufferfi(x0,x1,x2,x3);_glClearBufferfi.sig="viifi";var _emscripten_glClearBufferfi=_glClearBufferfi;_emscripten_glClearBufferfi.sig="viifi";function _glClearBufferfv(buffer,drawbuffer,value){value>>>=0;GLctx.clearBufferfv(buffer,drawbuffer,HEAPF32,value>>>2)}_glClearBufferfv.sig="viip";var _emscripten_glClearBufferfv=_glClearBufferfv;_emscripten_glClearBufferfv.sig="viip";function _glClearBufferiv(buffer,drawbuffer,value){value>>>=0;GLctx.clearBufferiv(buffer,drawbuffer,HEAP32,value>>>2)}_glClearBufferiv.sig="viip";var _emscripten_glClearBufferiv=_glClearBufferiv;_emscripten_glClearBufferiv.sig="viip";function _glClearBufferuiv(buffer,drawbuffer,value){value>>>=0;GLctx.clearBufferuiv(buffer,drawbuffer,HEAPU32,value>>>2)}_glClearBufferuiv.sig="viip";var _emscripten_glClearBufferuiv=_glClearBufferuiv;_emscripten_glClearBufferuiv.sig="viip";var _glClearColor=(x0,x1,x2,x3)=>GLctx.clearColor(x0,x1,x2,x3);_glClearColor.sig="vffff";var _emscripten_glClearColor=_glClearColor;_emscripten_glClearColor.sig="vffff";var _glClearDepthf=x0=>GLctx.clearDepth(x0);_glClearDepthf.sig="vf";var _emscripten_glClearDepthf=_glClearDepthf;_emscripten_glClearDepthf.sig="vf";var _glClearStencil=x0=>GLctx.clearStencil(x0);_glClearStencil.sig="vi";var _emscripten_glClearStencil=_glClearStencil;_emscripten_glClearStencil.sig="vi";function _glClientWaitSync(sync,flags,timeout){sync>>>=0;timeout=Number(timeout);return GLctx.clientWaitSync(GL.syncs[sync],flags,timeout)}_glClientWaitSync.sig="ipij";var _emscripten_glClientWaitSync=_glClientWaitSync;_emscripten_glClientWaitSync.sig="ipij";var _glClipControlEXT=(origin,depth)=>{GLctx.extClipControl["clipControlEXT"](origin,depth)};_glClipControlEXT.sig="vii";var _emscripten_glClipControlEXT=_glClipControlEXT;var _glColorMask=(red,green,blue,alpha)=>{GLctx.colorMask(!!red,!!green,!!blue,!!alpha)};_glColorMask.sig="viiii";var _emscripten_glColorMask=_glColorMask;_emscripten_glColorMask.sig="viiii";var _glCompileShader=shader=>{GLctx.compileShader(GL.shaders[shader])};_glCompileShader.sig="vi";var _emscripten_glCompileShader=_glCompileShader;_emscripten_glCompileShader.sig="vi";function _glCompressedTexImage2D(target,level,internalFormat,width,height,border,imageSize,data){data>>>=0;if(GL.currentContext.version>=2){if(GLctx.currentPixelUnpackBufferBinding||!imageSize){GLctx.compressedTexImage2D(target,level,internalFormat,width,height,border,imageSize,data);return}}GLctx.compressedTexImage2D(target,level,internalFormat,width,height,border,HEAPU8.subarray(data>>>0,data+imageSize>>>0))}_glCompressedTexImage2D.sig="viiiiiiip";var _emscripten_glCompressedTexImage2D=_glCompressedTexImage2D;_emscripten_glCompressedTexImage2D.sig="viiiiiiip";function _glCompressedTexImage3D(target,level,internalFormat,width,height,depth,border,imageSize,data){data>>>=0;if(GLctx.currentPixelUnpackBufferBinding){GLctx.compressedTexImage3D(target,level,internalFormat,width,height,depth,border,imageSize,data)}else{GLctx.compressedTexImage3D(target,level,internalFormat,width,height,depth,border,HEAPU8,data,imageSize)}}_glCompressedTexImage3D.sig="viiiiiiiip";var _emscripten_glCompressedTexImage3D=_glCompressedTexImage3D;_emscripten_glCompressedTexImage3D.sig="viiiiiiiip";function _glCompressedTexSubImage2D(target,level,xoffset,yoffset,width,height,format,imageSize,data){data>>>=0;if(GL.currentContext.version>=2){if(GLctx.currentPixelUnpackBufferBinding||!imageSize){GLctx.compressedTexSubImage2D(target,level,xoffset,yoffset,width,height,format,imageSize,data);return}}GLctx.compressedTexSubImage2D(target,level,xoffset,yoffset,width,height,format,HEAPU8.subarray(data>>>0,data+imageSize>>>0))}_glCompressedTexSubImage2D.sig="viiiiiiiip";var _emscripten_glCompressedTexSubImage2D=_glCompressedTexSubImage2D;_emscripten_glCompressedTexSubImage2D.sig="viiiiiiiip";function _glCompressedTexSubImage3D(target,level,xoffset,yoffset,zoffset,width,height,depth,format,imageSize,data){data>>>=0;if(GLctx.currentPixelUnpackBufferBinding){GLctx.compressedTexSubImage3D(target,level,xoffset,yoffset,zoffset,width,height,depth,format,imageSize,data)}else{GLctx.compressedTexSubImage3D(target,level,xoffset,yoffset,zoffset,width,height,depth,format,HEAPU8,data,imageSize)}}_glCompressedTexSubImage3D.sig="viiiiiiiiiip";var _emscripten_glCompressedTexSubImage3D=_glCompressedTexSubImage3D;_emscripten_glCompressedTexSubImage3D.sig="viiiiiiiiiip";function _glCopyBufferSubData(x0,x1,x2,x3,x4){x2>>>=0;x3>>>=0;x4>>>=0;return GLctx.copyBufferSubData(x0,x1,x2,x3,x4)}_glCopyBufferSubData.sig="viippp";var _emscripten_glCopyBufferSubData=_glCopyBufferSubData;_emscripten_glCopyBufferSubData.sig="viippp";var _glCopyTexImage2D=(x0,x1,x2,x3,x4,x5,x6,x7)=>GLctx.copyTexImage2D(x0,x1,x2,x3,x4,x5,x6,x7);_glCopyTexImage2D.sig="viiiiiiii";var _emscripten_glCopyTexImage2D=_glCopyTexImage2D;_emscripten_glCopyTexImage2D.sig="viiiiiiii";var _glCopyTexSubImage2D=(x0,x1,x2,x3,x4,x5,x6,x7)=>GLctx.copyTexSubImage2D(x0,x1,x2,x3,x4,x5,x6,x7);_glCopyTexSubImage2D.sig="viiiiiiii";var _emscripten_glCopyTexSubImage2D=_glCopyTexSubImage2D;_emscripten_glCopyTexSubImage2D.sig="viiiiiiii";var _glCopyTexSubImage3D=(x0,x1,x2,x3,x4,x5,x6,x7,x8)=>GLctx.copyTexSubImage3D(x0,x1,x2,x3,x4,x5,x6,x7,x8);_glCopyTexSubImage3D.sig="viiiiiiiii";var _emscripten_glCopyTexSubImage3D=_glCopyTexSubImage3D;_emscripten_glCopyTexSubImage3D.sig="viiiiiiiii";var _glCreateProgram=()=>{var id=GL.getNewId(GL.programs);var program=GLctx.createProgram();program.name=id;program.maxUniformLength=program.maxAttributeLength=program.maxUniformBlockNameLength=0;program.uniformIdCounter=1;GL.programs[id]=program;return id};_glCreateProgram.sig="i";var _emscripten_glCreateProgram=_glCreateProgram;_emscripten_glCreateProgram.sig="i";var _glCreateShader=shaderType=>{var id=GL.getNewId(GL.shaders);GL.shaders[id]=GLctx.createShader(shaderType);return id};_glCreateShader.sig="ii";var _emscripten_glCreateShader=_glCreateShader;_emscripten_glCreateShader.sig="ii";var _glCullFace=x0=>GLctx.cullFace(x0);_glCullFace.sig="vi";var _emscripten_glCullFace=_glCullFace;_emscripten_glCullFace.sig="vi";function _glDeleteBuffers(n,buffers){buffers>>>=0;for(var i=0;i<n;i++){var id=HEAP32[buffers+i*4>>>2>>>0];var buffer=GL.buffers[id];if(!buffer)continue;GLctx.deleteBuffer(buffer);buffer.name=0;GL.buffers[id]=null;if(id==GLctx.currentPixelPackBufferBinding)GLctx.currentPixelPackBufferBinding=0;if(id==GLctx.currentPixelUnpackBufferBinding)GLctx.currentPixelUnpackBufferBinding=0}}_glDeleteBuffers.sig="vip";var _emscripten_glDeleteBuffers=_glDeleteBuffers;_emscripten_glDeleteBuffers.sig="vip";function _glDeleteFramebuffers(n,framebuffers){framebuffers>>>=0;for(var i=0;i<n;++i){var id=HEAP32[framebuffers+i*4>>>2>>>0];var framebuffer=GL.framebuffers[id];if(!framebuffer)continue;GLctx.deleteFramebuffer(framebuffer);framebuffer.name=0;GL.framebuffers[id]=null}}_glDeleteFramebuffers.sig="vip";var _emscripten_glDeleteFramebuffers=_glDeleteFramebuffers;_emscripten_glDeleteFramebuffers.sig="vip";var _glDeleteProgram=id=>{if(!id)return;var program=GL.programs[id];if(!program){GL.recordError(1281);return}GLctx.deleteProgram(program);program.name=0;GL.programs[id]=null};_glDeleteProgram.sig="vi";var _emscripten_glDeleteProgram=_glDeleteProgram;_emscripten_glDeleteProgram.sig="vi";function _glDeleteQueries(n,ids){ids>>>=0;for(var i=0;i<n;i++){var id=HEAP32[ids+i*4>>>2>>>0];var query=GL.queries[id];if(!query)continue;GLctx.deleteQuery(query);GL.queries[id]=null}}_glDeleteQueries.sig="vip";var _emscripten_glDeleteQueries=_glDeleteQueries;_emscripten_glDeleteQueries.sig="vip";function _glDeleteQueriesEXT(n,ids){ids>>>=0;for(var i=0;i<n;i++){var id=HEAP32[ids+i*4>>>2>>>0];var query=GL.queries[id];if(!query)continue;GLctx.disjointTimerQueryExt["deleteQueryEXT"](query);GL.queries[id]=null}}_glDeleteQueriesEXT.sig="vip";var _emscripten_glDeleteQueriesEXT=_glDeleteQueriesEXT;function _glDeleteRenderbuffers(n,renderbuffers){renderbuffers>>>=0;for(var i=0;i<n;i++){var id=HEAP32[renderbuffers+i*4>>>2>>>0];var renderbuffer=GL.renderbuffers[id];if(!renderbuffer)continue;GLctx.deleteRenderbuffer(renderbuffer);renderbuffer.name=0;GL.renderbuffers[id]=null}}_glDeleteRenderbuffers.sig="vip";var _emscripten_glDeleteRenderbuffers=_glDeleteRenderbuffers;_emscripten_glDeleteRenderbuffers.sig="vip";function _glDeleteSamplers(n,samplers){samplers>>>=0;for(var i=0;i<n;i++){var id=HEAP32[samplers+i*4>>>2>>>0];var sampler=GL.samplers[id];if(!sampler)continue;GLctx.deleteSampler(sampler);sampler.name=0;GL.samplers[id]=null}}_glDeleteSamplers.sig="vip";var _emscripten_glDeleteSamplers=_glDeleteSamplers;_emscripten_glDeleteSamplers.sig="vip";var _glDeleteShader=id=>{if(!id)return;var shader=GL.shaders[id];if(!shader){GL.recordError(1281);return}GLctx.deleteShader(shader);GL.shaders[id]=null};_glDeleteShader.sig="vi";var _emscripten_glDeleteShader=_glDeleteShader;_emscripten_glDeleteShader.sig="vi";function _glDeleteSync(id){id>>>=0;if(!id)return;var sync=GL.syncs[id];if(!sync){GL.recordError(1281);return}GLctx.deleteSync(sync);sync.name=0;GL.syncs[id]=null}_glDeleteSync.sig="vp";var _emscripten_glDeleteSync=_glDeleteSync;_emscripten_glDeleteSync.sig="vp";function _glDeleteTextures(n,textures){textures>>>=0;for(var i=0;i<n;i++){var id=HEAP32[textures+i*4>>>2>>>0];var texture=GL.textures[id];if(!texture)continue;GLctx.deleteTexture(texture);texture.name=0;GL.textures[id]=null}}_glDeleteTextures.sig="vip";var _emscripten_glDeleteTextures=_glDeleteTextures;_emscripten_glDeleteTextures.sig="vip";function _glDeleteTransformFeedbacks(n,ids){ids>>>=0;for(var i=0;i<n;i++){var id=HEAP32[ids+i*4>>>2>>>0];var transformFeedback=GL.transformFeedbacks[id];if(!transformFeedback)continue;GLctx.deleteTransformFeedback(transformFeedback);transformFeedback.name=0;GL.transformFeedbacks[id]=null}}_glDeleteTransformFeedbacks.sig="vip";var _emscripten_glDeleteTransformFeedbacks=_glDeleteTransformFeedbacks;_emscripten_glDeleteTransformFeedbacks.sig="vip";function _glDeleteVertexArrays(n,vaos){vaos>>>=0;for(var i=0;i<n;i++){var id=HEAP32[vaos+i*4>>>2>>>0];GLctx.deleteVertexArray(GL.vaos[id]);GL.vaos[id]=null}}_glDeleteVertexArrays.sig="vip";var _emscripten_glDeleteVertexArrays=_glDeleteVertexArrays;_emscripten_glDeleteVertexArrays.sig="vip";var _glDeleteVertexArraysOES=_glDeleteVertexArrays;_glDeleteVertexArraysOES.sig="vip";var _emscripten_glDeleteVertexArraysOES=_glDeleteVertexArraysOES;_emscripten_glDeleteVertexArraysOES.sig="vip";var _glDepthFunc=x0=>GLctx.depthFunc(x0);_glDepthFunc.sig="vi";var _emscripten_glDepthFunc=_glDepthFunc;_emscripten_glDepthFunc.sig="vi";var _glDepthMask=flag=>{GLctx.depthMask(!!flag)};_glDepthMask.sig="vi";var _emscripten_glDepthMask=_glDepthMask;_emscripten_glDepthMask.sig="vi";var _glDepthRangef=(x0,x1)=>GLctx.depthRange(x0,x1);_glDepthRangef.sig="vff";var _emscripten_glDepthRangef=_glDepthRangef;_emscripten_glDepthRangef.sig="vff";var _glDetachShader=(program,shader)=>{GLctx.detachShader(GL.programs[program],GL.shaders[shader])};_glDetachShader.sig="vii";var _emscripten_glDetachShader=_glDetachShader;_emscripten_glDetachShader.sig="vii";var _glDisable=x0=>GLctx.disable(x0);_glDisable.sig="vi";var _emscripten_glDisable=_glDisable;_emscripten_glDisable.sig="vi";var _glDisableVertexAttribArray=index=>{GLctx.disableVertexAttribArray(index)};_glDisableVertexAttribArray.sig="vi";var _emscripten_glDisableVertexAttribArray=_glDisableVertexAttribArray;_emscripten_glDisableVertexAttribArray.sig="vi";var _glDrawArrays=(mode,first,count)=>{GLctx.drawArrays(mode,first,count)};_glDrawArrays.sig="viii";var _emscripten_glDrawArrays=_glDrawArrays;_emscripten_glDrawArrays.sig="viii";var _glDrawArraysInstanced=(mode,first,count,primcount)=>{GLctx.drawArraysInstanced(mode,first,count,primcount)};_glDrawArraysInstanced.sig="viiii";var _emscripten_glDrawArraysInstanced=_glDrawArraysInstanced;_emscripten_glDrawArraysInstanced.sig="viiii";var _glDrawArraysInstancedANGLE=_glDrawArraysInstanced;var _emscripten_glDrawArraysInstancedANGLE=_glDrawArraysInstancedANGLE;var _glDrawArraysInstancedARB=_glDrawArraysInstanced;var _emscripten_glDrawArraysInstancedARB=_glDrawArraysInstancedARB;var _glDrawArraysInstancedEXT=_glDrawArraysInstanced;var _emscripten_glDrawArraysInstancedEXT=_glDrawArraysInstancedEXT;var _glDrawArraysInstancedNV=_glDrawArraysInstanced;var _emscripten_glDrawArraysInstancedNV=_glDrawArraysInstancedNV;var tempFixedLengthArray=[];function _glDrawBuffers(n,bufs){bufs>>>=0;var bufArray=tempFixedLengthArray[n];for(var i=0;i<n;i++){bufArray[i]=HEAP32[bufs+i*4>>>2>>>0]}GLctx.drawBuffers(bufArray)}_glDrawBuffers.sig="vip";var _emscripten_glDrawBuffers=_glDrawBuffers;_emscripten_glDrawBuffers.sig="vip";var _glDrawBuffersEXT=_glDrawBuffers;var _emscripten_glDrawBuffersEXT=_glDrawBuffersEXT;var _glDrawBuffersWEBGL=_glDrawBuffers;var _emscripten_glDrawBuffersWEBGL=_glDrawBuffersWEBGL;function _glDrawElements(mode,count,type,indices){indices>>>=0;GLctx.drawElements(mode,count,type,indices)}_glDrawElements.sig="viiip";var _emscripten_glDrawElements=_glDrawElements;_emscripten_glDrawElements.sig="viiip";function _glDrawElementsInstanced(mode,count,type,indices,primcount){indices>>>=0;GLctx.drawElementsInstanced(mode,count,type,indices,primcount)}_glDrawElementsInstanced.sig="viiipi";var _emscripten_glDrawElementsInstanced=_glDrawElementsInstanced;_emscripten_glDrawElementsInstanced.sig="viiipi";var _glDrawElementsInstancedANGLE=_glDrawElementsInstanced;var _emscripten_glDrawElementsInstancedANGLE=_glDrawElementsInstancedANGLE;var _glDrawElementsInstancedARB=_glDrawElementsInstanced;var _emscripten_glDrawElementsInstancedARB=_glDrawElementsInstancedARB;var _glDrawElementsInstancedEXT=_glDrawElementsInstanced;var _emscripten_glDrawElementsInstancedEXT=_glDrawElementsInstancedEXT;var _glDrawElementsInstancedNV=_glDrawElementsInstanced;var _emscripten_glDrawElementsInstancedNV=_glDrawElementsInstancedNV;function _glDrawRangeElements(mode,start,end,count,type,indices){indices>>>=0;_glDrawElements(mode,count,type,indices)}_glDrawRangeElements.sig="viiiiip";var _emscripten_glDrawRangeElements=_glDrawRangeElements;_emscripten_glDrawRangeElements.sig="viiiiip";var _glEnable=x0=>GLctx.enable(x0);_glEnable.sig="vi";var _emscripten_glEnable=_glEnable;_emscripten_glEnable.sig="vi";var _glEnableVertexAttribArray=index=>{GLctx.enableVertexAttribArray(index)};_glEnableVertexAttribArray.sig="vi";var _emscripten_glEnableVertexAttribArray=_glEnableVertexAttribArray;_emscripten_glEnableVertexAttribArray.sig="vi";var _glEndQuery=x0=>GLctx.endQuery(x0);_glEndQuery.sig="vi";var _emscripten_glEndQuery=_glEndQuery;_emscripten_glEndQuery.sig="vi";var _glEndQueryEXT=target=>{GLctx.disjointTimerQueryExt["endQueryEXT"](target)};_glEndQueryEXT.sig="vi";var _emscripten_glEndQueryEXT=_glEndQueryEXT;var _glEndTransformFeedback=()=>GLctx.endTransformFeedback();_glEndTransformFeedback.sig="v";var _emscripten_glEndTransformFeedback=_glEndTransformFeedback;_emscripten_glEndTransformFeedback.sig="v";function _glFenceSync(condition,flags){var sync=GLctx.fenceSync(condition,flags);if(sync){var id=GL.getNewId(GL.syncs);sync.name=id;GL.syncs[id]=sync;return id}return 0}_glFenceSync.sig="pii";var _emscripten_glFenceSync=_glFenceSync;_emscripten_glFenceSync.sig="pii";var _glFinish=()=>GLctx.finish();_glFinish.sig="v";var _emscripten_glFinish=_glFinish;_emscripten_glFinish.sig="v";var _glFlush=()=>GLctx.flush();_glFlush.sig="v";var _emscripten_glFlush=_glFlush;_emscripten_glFlush.sig="v";var _glFramebufferRenderbuffer=(target,attachment,renderbuffertarget,renderbuffer)=>{GLctx.framebufferRenderbuffer(target,attachment,renderbuffertarget,GL.renderbuffers[renderbuffer])};_glFramebufferRenderbuffer.sig="viiii";var _emscripten_glFramebufferRenderbuffer=_glFramebufferRenderbuffer;_emscripten_glFramebufferRenderbuffer.sig="viiii";var _glFramebufferTexture2D=(target,attachment,textarget,texture,level)=>{GLctx.framebufferTexture2D(target,attachment,textarget,GL.textures[texture],level)};_glFramebufferTexture2D.sig="viiiii";var _emscripten_glFramebufferTexture2D=_glFramebufferTexture2D;_emscripten_glFramebufferTexture2D.sig="viiiii";var _glFramebufferTextureLayer=(target,attachment,texture,level,layer)=>{GLctx.framebufferTextureLayer(target,attachment,GL.textures[texture],level,layer)};_glFramebufferTextureLayer.sig="viiiii";var _emscripten_glFramebufferTextureLayer=_glFramebufferTextureLayer;_emscripten_glFramebufferTextureLayer.sig="viiiii";var _glFrontFace=x0=>GLctx.frontFace(x0);_glFrontFace.sig="vi";var _emscripten_glFrontFace=_glFrontFace;_emscripten_glFrontFace.sig="vi";function _glGenBuffers(n,buffers){buffers>>>=0;GL.genObject(n,buffers,"createBuffer",GL.buffers)}_glGenBuffers.sig="vip";var _emscripten_glGenBuffers=_glGenBuffers;_emscripten_glGenBuffers.sig="vip";function _glGenFramebuffers(n,ids){ids>>>=0;GL.genObject(n,ids,"createFramebuffer",GL.framebuffers)}_glGenFramebuffers.sig="vip";var _emscripten_glGenFramebuffers=_glGenFramebuffers;_emscripten_glGenFramebuffers.sig="vip";function _glGenQueries(n,ids){ids>>>=0;GL.genObject(n,ids,"createQuery",GL.queries)}_glGenQueries.sig="vip";var _emscripten_glGenQueries=_glGenQueries;_emscripten_glGenQueries.sig="vip";function _glGenQueriesEXT(n,ids){ids>>>=0;for(var i=0;i<n;i++){var query=GLctx.disjointTimerQueryExt["createQueryEXT"]();if(!query){GL.recordError(1282);while(i<n)HEAP32[ids+i++*4>>>2>>>0]=0;return}var id=GL.getNewId(GL.queries);query.name=id;GL.queries[id]=query;HEAP32[ids+i*4>>>2>>>0]=id}}_glGenQueriesEXT.sig="vip";var _emscripten_glGenQueriesEXT=_glGenQueriesEXT;function _glGenRenderbuffers(n,renderbuffers){renderbuffers>>>=0;GL.genObject(n,renderbuffers,"createRenderbuffer",GL.renderbuffers)}_glGenRenderbuffers.sig="vip";var _emscripten_glGenRenderbuffers=_glGenRenderbuffers;_emscripten_glGenRenderbuffers.sig="vip";function _glGenSamplers(n,samplers){samplers>>>=0;GL.genObject(n,samplers,"createSampler",GL.samplers)}_glGenSamplers.sig="vip";var _emscripten_glGenSamplers=_glGenSamplers;_emscripten_glGenSamplers.sig="vip";function _glGenTextures(n,textures){textures>>>=0;GL.genObject(n,textures,"createTexture",GL.textures)}_glGenTextures.sig="vip";var _emscripten_glGenTextures=_glGenTextures;_emscripten_glGenTextures.sig="vip";function _glGenTransformFeedbacks(n,ids){ids>>>=0;GL.genObject(n,ids,"createTransformFeedback",GL.transformFeedbacks)}_glGenTransformFeedbacks.sig="vip";var _emscripten_glGenTransformFeedbacks=_glGenTransformFeedbacks;_emscripten_glGenTransformFeedbacks.sig="vip";function _glGenVertexArrays(n,arrays){arrays>>>=0;GL.genObject(n,arrays,"createVertexArray",GL.vaos)}_glGenVertexArrays.sig="vip";var _emscripten_glGenVertexArrays=_glGenVertexArrays;_emscripten_glGenVertexArrays.sig="vip";var _glGenVertexArraysOES=_glGenVertexArrays;_glGenVertexArraysOES.sig="vip";var _emscripten_glGenVertexArraysOES=_glGenVertexArraysOES;_emscripten_glGenVertexArraysOES.sig="vip";var _glGenerateMipmap=x0=>GLctx.generateMipmap(x0);_glGenerateMipmap.sig="vi";var _emscripten_glGenerateMipmap=_glGenerateMipmap;_emscripten_glGenerateMipmap.sig="vi";var __glGetActiveAttribOrUniform=(funcName,program,index,bufSize,length,size,type,name)=>{program=GL.programs[program];var info=GLctx[funcName](program,index);if(info){var numBytesWrittenExclNull=name&&stringToUTF8(info.name,name,bufSize);if(length)HEAP32[length>>>2>>>0]=numBytesWrittenExclNull;if(size)HEAP32[size>>>2>>>0]=info.size;if(type)HEAP32[type>>>2>>>0]=info.type}};function _glGetActiveAttrib(program,index,bufSize,length,size,type,name){length>>>=0;size>>>=0;type>>>=0;name>>>=0;return __glGetActiveAttribOrUniform("getActiveAttrib",program,index,bufSize,length,size,type,name)}_glGetActiveAttrib.sig="viiipppp";var _emscripten_glGetActiveAttrib=_glGetActiveAttrib;_emscripten_glGetActiveAttrib.sig="viiipppp";function _glGetActiveUniform(program,index,bufSize,length,size,type,name){length>>>=0;size>>>=0;type>>>=0;name>>>=0;return __glGetActiveAttribOrUniform("getActiveUniform",program,index,bufSize,length,size,type,name)}_glGetActiveUniform.sig="viiipppp";var _emscripten_glGetActiveUniform=_glGetActiveUniform;_emscripten_glGetActiveUniform.sig="viiipppp";function _glGetActiveUniformBlockName(program,uniformBlockIndex,bufSize,length,uniformBlockName){length>>>=0;uniformBlockName>>>=0;program=GL.programs[program];var result=GLctx.getActiveUniformBlockName(program,uniformBlockIndex);if(!result)return;if(uniformBlockName&&bufSize>0){var numBytesWrittenExclNull=stringToUTF8(result,uniformBlockName,bufSize);if(length)HEAP32[length>>>2>>>0]=numBytesWrittenExclNull}else{if(length)HEAP32[length>>>2>>>0]=0}}_glGetActiveUniformBlockName.sig="viiipp";var _emscripten_glGetActiveUniformBlockName=_glGetActiveUniformBlockName;_emscripten_glGetActiveUniformBlockName.sig="viiipp";function _glGetActiveUniformBlockiv(program,uniformBlockIndex,pname,params){params>>>=0;if(!params){GL.recordError(1281);return}program=GL.programs[program];if(pname==35393){var name=GLctx.getActiveUniformBlockName(program,uniformBlockIndex);HEAP32[params>>>2>>>0]=name.length+1;return}var result=GLctx.getActiveUniformBlockParameter(program,uniformBlockIndex,pname);if(result===null)return;if(pname==35395){for(var i=0;i<result.length;i++){HEAP32[params+i*4>>>2>>>0]=result[i]}}else{HEAP32[params>>>2>>>0]=result}}_glGetActiveUniformBlockiv.sig="viiip";var _emscripten_glGetActiveUniformBlockiv=_glGetActiveUniformBlockiv;_emscripten_glGetActiveUniformBlockiv.sig="viiip";function _glGetActiveUniformsiv(program,uniformCount,uniformIndices,pname,params){uniformIndices>>>=0;params>>>=0;if(!params){GL.recordError(1281);return}if(uniformCount>0&&uniformIndices==0){GL.recordError(1281);return}program=GL.programs[program];var ids=[];for(var i=0;i<uniformCount;i++){ids.push(HEAP32[uniformIndices+i*4>>>2>>>0])}var result=GLctx.getActiveUniforms(program,ids,pname);if(!result)return;var len=result.length;for(var i=0;i<len;i++){HEAP32[params+i*4>>>2>>>0]=result[i]}}_glGetActiveUniformsiv.sig="viipip";var _emscripten_glGetActiveUniformsiv=_glGetActiveUniformsiv;_emscripten_glGetActiveUniformsiv.sig="viipip";function _glGetAttachedShaders(program,maxCount,count,shaders){count>>>=0;shaders>>>=0;var result=GLctx.getAttachedShaders(GL.programs[program]);var len=result.length;if(len>maxCount){len=maxCount}HEAP32[count>>>2>>>0]=len;for(var i=0;i<len;++i){var id=GL.shaders.indexOf(result[i]);HEAP32[shaders+i*4>>>2>>>0]=id}}_glGetAttachedShaders.sig="viipp";var _emscripten_glGetAttachedShaders=_glGetAttachedShaders;_emscripten_glGetAttachedShaders.sig="viipp";function _glGetAttribLocation(program,name){name>>>=0;return GLctx.getAttribLocation(GL.programs[program],UTF8ToString(name))}_glGetAttribLocation.sig="iip";var _emscripten_glGetAttribLocation=_glGetAttribLocation;_emscripten_glGetAttribLocation.sig="iip";var writeI53ToI64=(ptr,num)=>{HEAPU32[ptr>>>2>>>0]=num;var lower=HEAPU32[ptr>>>2>>>0];HEAPU32[ptr+4>>>2>>>0]=(num-lower)/4294967296};var webglGetExtensions=()=>{var exts=getEmscriptenSupportedExtensions(GLctx);exts=exts.concat(exts.map(e=>"GL_"+e));return exts};var emscriptenWebGLGet=(name_,p,type)=>{if(!p){GL.recordError(1281);return}var ret=undefined;switch(name_){case 36346:ret=1;break;case 36344:if(type!=0&&type!=1){GL.recordError(1280)}return;case 34814:case 36345:ret=0;break;case 34466:var formats=GLctx.getParameter(34467);ret=formats?formats.length:0;break;case 33309:if(GL.currentContext.version<2){GL.recordError(1282);return}ret=webglGetExtensions().length;break;case 33307:case 33308:if(GL.currentContext.version<2){GL.recordError(1280);return}ret=name_==33307?3:0;break}if(ret===undefined){var result=GLctx.getParameter(name_);switch(typeof result){case"number":ret=result;break;case"boolean":ret=result?1:0;break;case"string":GL.recordError(1280);return;case"object":if(result===null){switch(name_){case 34964:case 35725:case 34965:case 36006:case 36007:case 32873:case 34229:case 36662:case 36663:case 35053:case 35055:case 36010:case 35097:case 35869:case 32874:case 36389:case 35983:case 35368:case 34068:{ret=0;break}default:{GL.recordError(1280);return}}}else if(result instanceof Float32Array||result instanceof Uint32Array||result instanceof Int32Array||result instanceof Array){for(var i=0;i<result.length;++i){switch(type){case 0:HEAP32[p+i*4>>>2>>>0]=result[i];break;case 2:HEAPF32[p+i*4>>>2>>>0]=result[i];break;case 4:HEAP8[p+i>>>0]=result[i]?1:0;break}}return}else{try{ret=result.name|0}catch(e){GL.recordError(1280);err(`GL_INVALID_ENUM in glGet${type}v: Unknown object returned from WebGL getParameter(${name_})! (error: ${e})`);return}}break;default:GL.recordError(1280);err(`GL_INVALID_ENUM in glGet${type}v: Native code calling glGet${type}v(${name_}) and it returns ${result} of type ${typeof result}!`);return}}switch(type){case 1:writeI53ToI64(p,ret);break;case 0:HEAP32[p>>>2>>>0]=ret;break;case 2:HEAPF32[p>>>2>>>0]=ret;break;case 4:HEAP8[p>>>0]=ret?1:0;break}};function _glGetBooleanv(name_,p){p>>>=0;return emscriptenWebGLGet(name_,p,4)}_glGetBooleanv.sig="vip";var _emscripten_glGetBooleanv=_glGetBooleanv;_emscripten_glGetBooleanv.sig="vip";function _glGetBufferParameteri64v(target,value,data){data>>>=0;if(!data){GL.recordError(1281);return}writeI53ToI64(data,GLctx.getBufferParameter(target,value))}_glGetBufferParameteri64v.sig="viip";var _emscripten_glGetBufferParameteri64v=_glGetBufferParameteri64v;_emscripten_glGetBufferParameteri64v.sig="viip";function _glGetBufferParameteriv(target,value,data){data>>>=0;if(!data){GL.recordError(1281);return}HEAP32[data>>>2>>>0]=GLctx.getBufferParameter(target,value)}_glGetBufferParameteriv.sig="viip";var _emscripten_glGetBufferParameteriv=_glGetBufferParameteriv;_emscripten_glGetBufferParameteriv.sig="viip";var _glGetError=()=>{var error=GLctx.getError()||GL.lastError;GL.lastError=0;return error};_glGetError.sig="i";var _emscripten_glGetError=_glGetError;_emscripten_glGetError.sig="i";function _glGetFloatv(name_,p){p>>>=0;return emscriptenWebGLGet(name_,p,2)}_glGetFloatv.sig="vip";var _emscripten_glGetFloatv=_glGetFloatv;_emscripten_glGetFloatv.sig="vip";function _glGetFragDataLocation(program,name){name>>>=0;return GLctx.getFragDataLocation(GL.programs[program],UTF8ToString(name))}_glGetFragDataLocation.sig="iip";var _emscripten_glGetFragDataLocation=_glGetFragDataLocation;_emscripten_glGetFragDataLocation.sig="iip";function _glGetFramebufferAttachmentParameteriv(target,attachment,pname,params){params>>>=0;var result=GLctx.getFramebufferAttachmentParameter(target,attachment,pname);if(result instanceof WebGLRenderbuffer||result instanceof WebGLTexture){result=result.name|0}HEAP32[params>>>2>>>0]=result}_glGetFramebufferAttachmentParameteriv.sig="viiip";var _emscripten_glGetFramebufferAttachmentParameteriv=_glGetFramebufferAttachmentParameteriv;_emscripten_glGetFramebufferAttachmentParameteriv.sig="viiip";var emscriptenWebGLGetIndexed=(target,index,data,type)=>{if(!data){GL.recordError(1281);return}var result=GLctx.getIndexedParameter(target,index);var ret;switch(typeof result){case"boolean":ret=result?1:0;break;case"number":ret=result;break;case"object":if(result===null){switch(target){case 35983:case 35368:ret=0;break;default:{GL.recordError(1280);return}}}else if(result instanceof WebGLBuffer){ret=result.name|0}else{GL.recordError(1280);return}break;default:GL.recordError(1280);return}switch(type){case 1:writeI53ToI64(data,ret);break;case 0:HEAP32[data>>>2>>>0]=ret;break;case 2:HEAPF32[data>>>2>>>0]=ret;break;case 4:HEAP8[data>>>0]=ret?1:0;break;default:throw"internal emscriptenWebGLGetIndexed() error, bad type: "+type}};function _glGetInteger64i_v(target,index,data){data>>>=0;return emscriptenWebGLGetIndexed(target,index,data,1)}_glGetInteger64i_v.sig="viip";var _emscripten_glGetInteger64i_v=_glGetInteger64i_v;_emscripten_glGetInteger64i_v.sig="viip";function _glGetInteger64v(name_,p){p>>>=0;emscriptenWebGLGet(name_,p,1)}_glGetInteger64v.sig="vip";var _emscripten_glGetInteger64v=_glGetInteger64v;_emscripten_glGetInteger64v.sig="vip";function _glGetIntegeri_v(target,index,data){data>>>=0;return emscriptenWebGLGetIndexed(target,index,data,0)}_glGetIntegeri_v.sig="viip";var _emscripten_glGetIntegeri_v=_glGetIntegeri_v;_emscripten_glGetIntegeri_v.sig="viip";function _glGetIntegerv(name_,p){p>>>=0;return emscriptenWebGLGet(name_,p,0)}_glGetIntegerv.sig="vip";var _emscripten_glGetIntegerv=_glGetIntegerv;_emscripten_glGetIntegerv.sig="vip";function _glGetInternalformativ(target,internalformat,pname,bufSize,params){params>>>=0;if(bufSize<0){GL.recordError(1281);return}if(!params){GL.recordError(1281);return}var ret=GLctx.getInternalformatParameter(target,internalformat,pname);if(ret===null)return;for(var i=0;i<ret.length&&i<bufSize;++i){HEAP32[params+i*4>>>2>>>0]=ret[i]}}_glGetInternalformativ.sig="viiiip";var _emscripten_glGetInternalformativ=_glGetInternalformativ;_emscripten_glGetInternalformativ.sig="viiiip";function _glGetProgramBinary(program,bufSize,length,binaryFormat,binary){length>>>=0;binaryFormat>>>=0;binary>>>=0;GL.recordError(1282)}_glGetProgramBinary.sig="viippp";var _emscripten_glGetProgramBinary=_glGetProgramBinary;_emscripten_glGetProgramBinary.sig="viippp";function _glGetProgramInfoLog(program,maxLength,length,infoLog){length>>>=0;infoLog>>>=0;var log=GLctx.getProgramInfoLog(GL.programs[program]);if(log===null)log="(unknown error)";var numBytesWrittenExclNull=maxLength>0&&infoLog?stringToUTF8(log,infoLog,maxLength):0;if(length)HEAP32[length>>>2>>>0]=numBytesWrittenExclNull}_glGetProgramInfoLog.sig="viipp";var _emscripten_glGetProgramInfoLog=_glGetProgramInfoLog;_emscripten_glGetProgramInfoLog.sig="viipp";function _glGetProgramiv(program,pname,p){p>>>=0;if(!p){GL.recordError(1281);return}if(program>=GL.counter){GL.recordError(1281);return}program=GL.programs[program];if(pname==35716){var log=GLctx.getProgramInfoLog(program);if(log===null)log="(unknown error)";HEAP32[p>>>2>>>0]=log.length+1}else if(pname==35719){if(!program.maxUniformLength){var numActiveUniforms=GLctx.getProgramParameter(program,35718);for(var i=0;i<numActiveUniforms;++i){program.maxUniformLength=Math.max(program.maxUniformLength,GLctx.getActiveUniform(program,i).name.length+1)}}HEAP32[p>>>2>>>0]=program.maxUniformLength}else if(pname==35722){if(!program.maxAttributeLength){var numActiveAttributes=GLctx.getProgramParameter(program,35721);for(var i=0;i<numActiveAttributes;++i){program.maxAttributeLength=Math.max(program.maxAttributeLength,GLctx.getActiveAttrib(program,i).name.length+1)}}HEAP32[p>>>2>>>0]=program.maxAttributeLength}else if(pname==35381){if(!program.maxUniformBlockNameLength){var numActiveUniformBlocks=GLctx.getProgramParameter(program,35382);for(var i=0;i<numActiveUniformBlocks;++i){program.maxUniformBlockNameLength=Math.max(program.maxUniformBlockNameLength,GLctx.getActiveUniformBlockName(program,i).length+1)}}HEAP32[p>>>2>>>0]=program.maxUniformBlockNameLength}else{HEAP32[p>>>2>>>0]=GLctx.getProgramParameter(program,pname)}}_glGetProgramiv.sig="viip";var _emscripten_glGetProgramiv=_glGetProgramiv;_emscripten_glGetProgramiv.sig="viip";function _glGetQueryObjecti64vEXT(id,pname,params){params>>>=0;if(!params){GL.recordError(1281);return}var query=GL.queries[id];var param;if(GL.currentContext.version<2){param=GLctx.disjointTimerQueryExt["getQueryObjectEXT"](query,pname)}else{param=GLctx.getQueryParameter(query,pname)}var ret;if(typeof param=="boolean"){ret=param?1:0}else{ret=param}writeI53ToI64(params,ret)}_glGetQueryObjecti64vEXT.sig="viip";var _emscripten_glGetQueryObjecti64vEXT=_glGetQueryObjecti64vEXT;function _glGetQueryObjectivEXT(id,pname,params){params>>>=0;if(!params){GL.recordError(1281);return}var query=GL.queries[id];var param=GLctx.disjointTimerQueryExt["getQueryObjectEXT"](query,pname);var ret;if(typeof param=="boolean"){ret=param?1:0}else{ret=param}HEAP32[params>>>2>>>0]=ret}_glGetQueryObjectivEXT.sig="viip";var _emscripten_glGetQueryObjectivEXT=_glGetQueryObjectivEXT;var _glGetQueryObjectui64vEXT=_glGetQueryObjecti64vEXT;var _emscripten_glGetQueryObjectui64vEXT=_glGetQueryObjectui64vEXT;function _glGetQueryObjectuiv(id,pname,params){params>>>=0;if(!params){GL.recordError(1281);return}var query=GL.queries[id];var param=GLctx.getQueryParameter(query,pname);var ret;if(typeof param=="boolean"){ret=param?1:0}else{ret=param}HEAP32[params>>>2>>>0]=ret}_glGetQueryObjectuiv.sig="viip";var _emscripten_glGetQueryObjectuiv=_glGetQueryObjectuiv;_emscripten_glGetQueryObjectuiv.sig="viip";var _glGetQueryObjectuivEXT=_glGetQueryObjectivEXT;var _emscripten_glGetQueryObjectuivEXT=_glGetQueryObjectuivEXT;function _glGetQueryiv(target,pname,params){params>>>=0;if(!params){GL.recordError(1281);return}HEAP32[params>>>2>>>0]=GLctx.getQuery(target,pname)}_glGetQueryiv.sig="viip";var _emscripten_glGetQueryiv=_glGetQueryiv;_emscripten_glGetQueryiv.sig="viip";function _glGetQueryivEXT(target,pname,params){params>>>=0;if(!params){GL.recordError(1281);return}HEAP32[params>>>2>>>0]=GLctx.disjointTimerQueryExt["getQueryEXT"](target,pname)}_glGetQueryivEXT.sig="viip";var _emscripten_glGetQueryivEXT=_glGetQueryivEXT;function _glGetRenderbufferParameteriv(target,pname,params){params>>>=0;if(!params){GL.recordError(1281);return}HEAP32[params>>>2>>>0]=GLctx.getRenderbufferParameter(target,pname)}_glGetRenderbufferParameteriv.sig="viip";var _emscripten_glGetRenderbufferParameteriv=_glGetRenderbufferParameteriv;_emscripten_glGetRenderbufferParameteriv.sig="viip";function _glGetSamplerParameterfv(sampler,pname,params){params>>>=0;if(!params){GL.recordError(1281);return}HEAPF32[params>>>2>>>0]=GLctx.getSamplerParameter(GL.samplers[sampler],pname)}_glGetSamplerParameterfv.sig="viip";var _emscripten_glGetSamplerParameterfv=_glGetSamplerParameterfv;_emscripten_glGetSamplerParameterfv.sig="viip";function _glGetSamplerParameteriv(sampler,pname,params){params>>>=0;if(!params){GL.recordError(1281);return}HEAP32[params>>>2>>>0]=GLctx.getSamplerParameter(GL.samplers[sampler],pname)}_glGetSamplerParameteriv.sig="viip";var _emscripten_glGetSamplerParameteriv=_glGetSamplerParameteriv;_emscripten_glGetSamplerParameteriv.sig="viip";function _glGetShaderInfoLog(shader,maxLength,length,infoLog){length>>>=0;infoLog>>>=0;var log=GLctx.getShaderInfoLog(GL.shaders[shader]);if(log===null)log="(unknown error)";var numBytesWrittenExclNull=maxLength>0&&infoLog?stringToUTF8(log,infoLog,maxLength):0;if(length)HEAP32[length>>>2>>>0]=numBytesWrittenExclNull}_glGetShaderInfoLog.sig="viipp";var _emscripten_glGetShaderInfoLog=_glGetShaderInfoLog;_emscripten_glGetShaderInfoLog.sig="viipp";function _glGetShaderPrecisionFormat(shaderType,precisionType,range,precision){range>>>=0;precision>>>=0;var result=GLctx.getShaderPrecisionFormat(shaderType,precisionType);HEAP32[range>>>2>>>0]=result.rangeMin;HEAP32[range+4>>>2>>>0]=result.rangeMax;HEAP32[precision>>>2>>>0]=result.precision}_glGetShaderPrecisionFormat.sig="viipp";var _emscripten_glGetShaderPrecisionFormat=_glGetShaderPrecisionFormat;_emscripten_glGetShaderPrecisionFormat.sig="viipp";function _glGetShaderSource(shader,bufSize,length,source){length>>>=0;source>>>=0;var result=GLctx.getShaderSource(GL.shaders[shader]);if(!result)return;var numBytesWrittenExclNull=bufSize>0&&source?stringToUTF8(result,source,bufSize):0;if(length)HEAP32[length>>>2>>>0]=numBytesWrittenExclNull}_glGetShaderSource.sig="viipp";var _emscripten_glGetShaderSource=_glGetShaderSource;_emscripten_glGetShaderSource.sig="viipp";function _glGetShaderiv(shader,pname,p){p>>>=0;if(!p){GL.recordError(1281);return}if(pname==35716){var log=GLctx.getShaderInfoLog(GL.shaders[shader]);if(log===null)log="(unknown error)";var logLength=log?log.length+1:0;HEAP32[p>>>2>>>0]=logLength}else if(pname==35720){var source=GLctx.getShaderSource(GL.shaders[shader]);var sourceLength=source?source.length+1:0;HEAP32[p>>>2>>>0]=sourceLength}else{HEAP32[p>>>2>>>0]=GLctx.getShaderParameter(GL.shaders[shader],pname)}}_glGetShaderiv.sig="viip";var _emscripten_glGetShaderiv=_glGetShaderiv;_emscripten_glGetShaderiv.sig="viip";var stringToNewUTF8=str=>{var size=lengthBytesUTF8(str)+1;var ret=_malloc(size);if(ret)stringToUTF8(str,ret,size);return ret};function _glGetString(name_){var ret=GL.stringCache[name_];if(!ret){switch(name_){case 7939:ret=stringToNewUTF8(webglGetExtensions().join(" "));break;case 7936:case 7937:case 37445:case 37446:var s=GLctx.getParameter(name_);if(!s){GL.recordError(1280)}ret=s?stringToNewUTF8(s):0;break;case 7938:var webGLVersion=GLctx.getParameter(7938);var glVersion=`OpenGL ES 2.0 (${webGLVersion})`;if(GL.currentContext.version>=2)glVersion=`OpenGL ES 3.0 (${webGLVersion})`;ret=stringToNewUTF8(glVersion);break;case 35724:var glslVersion=GLctx.getParameter(35724);var ver_re=/^WebGL GLSL ES ([0-9]\.[0-9][0-9]?)(?:$| .*)/;var ver_num=glslVersion.match(ver_re);if(ver_num!==null){if(ver_num[1].length==3)ver_num[1]=ver_num[1]+"0";glslVersion=`OpenGL ES GLSL ES ${ver_num[1]} (${glslVersion})`}ret=stringToNewUTF8(glslVersion);break;default:GL.recordError(1280)}GL.stringCache[name_]=ret}return ret}_glGetString.sig="pi";var _emscripten_glGetString=_glGetString;_emscripten_glGetString.sig="pi";function _glGetStringi(name,index){if(GL.currentContext.version<2){GL.recordError(1282);return 0}var stringiCache=GL.stringiCache[name];if(stringiCache){if(index<0||index>=stringiCache.length){GL.recordError(1281);return 0}return stringiCache[index]}switch(name){case 7939:var exts=webglGetExtensions().map(stringToNewUTF8);stringiCache=GL.stringiCache[name]=exts;if(index<0||index>=stringiCache.length){GL.recordError(1281);return 0}return stringiCache[index];default:GL.recordError(1280);return 0}}_glGetStringi.sig="pii";var _emscripten_glGetStringi=_glGetStringi;_emscripten_glGetStringi.sig="pii";function _glGetSynciv(sync,pname,bufSize,length,values){sync>>>=0;length>>>=0;values>>>=0;if(bufSize<0){GL.recordError(1281);return}if(!values){GL.recordError(1281);return}var ret=GLctx.getSyncParameter(GL.syncs[sync],pname);if(ret!==null){HEAP32[values>>>2>>>0]=ret;if(length)HEAP32[length>>>2>>>0]=1}}_glGetSynciv.sig="vpiipp";var _emscripten_glGetSynciv=_glGetSynciv;_emscripten_glGetSynciv.sig="vpiipp";function _glGetTexParameterfv(target,pname,params){params>>>=0;if(!params){GL.recordError(1281);return}HEAPF32[params>>>2>>>0]=GLctx.getTexParameter(target,pname)}_glGetTexParameterfv.sig="viip";var _emscripten_glGetTexParameterfv=_glGetTexParameterfv;_emscripten_glGetTexParameterfv.sig="viip";function _glGetTexParameteriv(target,pname,params){params>>>=0;if(!params){GL.recordError(1281);return}HEAP32[params>>>2>>>0]=GLctx.getTexParameter(target,pname)}_glGetTexParameteriv.sig="viip";var _emscripten_glGetTexParameteriv=_glGetTexParameteriv;_emscripten_glGetTexParameteriv.sig="viip";function _glGetTransformFeedbackVarying(program,index,bufSize,length,size,type,name){length>>>=0;size>>>=0;type>>>=0;name>>>=0;program=GL.programs[program];var info=GLctx.getTransformFeedbackVarying(program,index);if(!info)return;if(name&&bufSize>0){var numBytesWrittenExclNull=stringToUTF8(info.name,name,bufSize);if(length)HEAP32[length>>>2>>>0]=numBytesWrittenExclNull}else{if(length)HEAP32[length>>>2>>>0]=0}if(size)HEAP32[size>>>2>>>0]=info.size;if(type)HEAP32[type>>>2>>>0]=info.type}_glGetTransformFeedbackVarying.sig="viiipppp";var _emscripten_glGetTransformFeedbackVarying=_glGetTransformFeedbackVarying;_emscripten_glGetTransformFeedbackVarying.sig="viiipppp";function _glGetUniformBlockIndex(program,uniformBlockName){uniformBlockName>>>=0;return GLctx.getUniformBlockIndex(GL.programs[program],UTF8ToString(uniformBlockName))}_glGetUniformBlockIndex.sig="iip";var _emscripten_glGetUniformBlockIndex=_glGetUniformBlockIndex;_emscripten_glGetUniformBlockIndex.sig="iip";function _glGetUniformIndices(program,uniformCount,uniformNames,uniformIndices){uniformNames>>>=0;uniformIndices>>>=0;if(!uniformIndices){GL.recordError(1281);return}if(uniformCount>0&&(uniformNames==0||uniformIndices==0)){GL.recordError(1281);return}program=GL.programs[program];var names=[];for(var i=0;i<uniformCount;i++)names.push(UTF8ToString(HEAP32[uniformNames+i*4>>>2>>>0]));var result=GLctx.getUniformIndices(program,names);if(!result)return;var len=result.length;for(var i=0;i<len;i++){HEAP32[uniformIndices+i*4>>>2>>>0]=result[i]}}_glGetUniformIndices.sig="viipp";var _emscripten_glGetUniformIndices=_glGetUniformIndices;_emscripten_glGetUniformIndices.sig="viipp";var jstoi_q=str=>parseInt(str);var webglGetLeftBracePos=name=>name.slice(-1)=="]"&&name.lastIndexOf("[");var webglPrepareUniformLocationsBeforeFirstUse=program=>{var uniformLocsById=program.uniformLocsById,uniformSizeAndIdsByName=program.uniformSizeAndIdsByName,i,j;if(!uniformLocsById){program.uniformLocsById=uniformLocsById={};program.uniformArrayNamesById={};var numActiveUniforms=GLctx.getProgramParameter(program,35718);for(i=0;i<numActiveUniforms;++i){var u=GLctx.getActiveUniform(program,i);var nm=u.name;var sz=u.size;var lb=webglGetLeftBracePos(nm);var arrayName=lb>0?nm.slice(0,lb):nm;var id=program.uniformIdCounter;program.uniformIdCounter+=sz;uniformSizeAndIdsByName[arrayName]=[sz,id];for(j=0;j<sz;++j){uniformLocsById[id]=j;program.uniformArrayNamesById[id++]=arrayName}}}};function _glGetUniformLocation(program,name){name>>>=0;name=UTF8ToString(name);if(program=GL.programs[program]){webglPrepareUniformLocationsBeforeFirstUse(program);var uniformLocsById=program.uniformLocsById;var arrayIndex=0;var uniformBaseName=name;var leftBrace=webglGetLeftBracePos(name);if(leftBrace>0){arrayIndex=jstoi_q(name.slice(leftBrace+1))>>>0;uniformBaseName=name.slice(0,leftBrace)}var sizeAndId=program.uniformSizeAndIdsByName[uniformBaseName];if(sizeAndId&&arrayIndex<sizeAndId[0]){arrayIndex+=sizeAndId[1];if(uniformLocsById[arrayIndex]=uniformLocsById[arrayIndex]||GLctx.getUniformLocation(program,name)){return arrayIndex}}}else{GL.recordError(1281)}return-1}_glGetUniformLocation.sig="iip";var _emscripten_glGetUniformLocation=_glGetUniformLocation;_emscripten_glGetUniformLocation.sig="iip";var webglGetUniformLocation=location=>{var p=GLctx.currentProgram;if(p){var webglLoc=p.uniformLocsById[location];if(typeof webglLoc=="number"){p.uniformLocsById[location]=webglLoc=GLctx.getUniformLocation(p,p.uniformArrayNamesById[location]+(webglLoc>0?`[${webglLoc}]`:""))}return webglLoc}else{GL.recordError(1282)}};var emscriptenWebGLGetUniform=(program,location,params,type)=>{if(!params){GL.recordError(1281);return}program=GL.programs[program];webglPrepareUniformLocationsBeforeFirstUse(program);var data=GLctx.getUniform(program,webglGetUniformLocation(location));if(typeof data=="number"||typeof data=="boolean"){switch(type){case 0:HEAP32[params>>>2>>>0]=data;break;case 2:HEAPF32[params>>>2>>>0]=data;break}}else{for(var i=0;i<data.length;i++){switch(type){case 0:HEAP32[params+i*4>>>2>>>0]=data[i];break;case 2:HEAPF32[params+i*4>>>2>>>0]=data[i];break}}}};function _glGetUniformfv(program,location,params){params>>>=0;emscriptenWebGLGetUniform(program,location,params,2)}_glGetUniformfv.sig="viip";var _emscripten_glGetUniformfv=_glGetUniformfv;_emscripten_glGetUniformfv.sig="viip";function _glGetUniformiv(program,location,params){params>>>=0;emscriptenWebGLGetUniform(program,location,params,0)}_glGetUniformiv.sig="viip";var _emscripten_glGetUniformiv=_glGetUniformiv;_emscripten_glGetUniformiv.sig="viip";function _glGetUniformuiv(program,location,params){params>>>=0;return emscriptenWebGLGetUniform(program,location,params,0)}_glGetUniformuiv.sig="viip";var _emscripten_glGetUniformuiv=_glGetUniformuiv;_emscripten_glGetUniformuiv.sig="viip";var emscriptenWebGLGetVertexAttrib=(index,pname,params,type)=>{if(!params){GL.recordError(1281);return}var data=GLctx.getVertexAttrib(index,pname);if(pname==34975){HEAP32[params>>>2>>>0]=data&&data["name"]}else if(typeof data=="number"||typeof data=="boolean"){switch(type){case 0:HEAP32[params>>>2>>>0]=data;break;case 2:HEAPF32[params>>>2>>>0]=data;break;case 5:HEAP32[params>>>2>>>0]=Math.fround(data);break}}else{for(var i=0;i<data.length;i++){switch(type){case 0:HEAP32[params+i*4>>>2>>>0]=data[i];break;case 2:HEAPF32[params+i*4>>>2>>>0]=data[i];break;case 5:HEAP32[params+i*4>>>2>>>0]=Math.fround(data[i]);break}}}};function _glGetVertexAttribIiv(index,pname,params){params>>>=0;emscriptenWebGLGetVertexAttrib(index,pname,params,0)}_glGetVertexAttribIiv.sig="viip";var _emscripten_glGetVertexAttribIiv=_glGetVertexAttribIiv;_emscripten_glGetVertexAttribIiv.sig="viip";var _glGetVertexAttribIuiv=_glGetVertexAttribIiv;_glGetVertexAttribIuiv.sig="viip";var _emscripten_glGetVertexAttribIuiv=_glGetVertexAttribIuiv;_emscripten_glGetVertexAttribIuiv.sig="viip";function _glGetVertexAttribPointerv(index,pname,pointer){pointer>>>=0;if(!pointer){GL.recordError(1281);return}HEAP32[pointer>>>2>>>0]=GLctx.getVertexAttribOffset(index,pname)}_glGetVertexAttribPointerv.sig="viip";var _emscripten_glGetVertexAttribPointerv=_glGetVertexAttribPointerv;_emscripten_glGetVertexAttribPointerv.sig="viip";function _glGetVertexAttribfv(index,pname,params){params>>>=0;emscriptenWebGLGetVertexAttrib(index,pname,params,2)}_glGetVertexAttribfv.sig="viip";var _emscripten_glGetVertexAttribfv=_glGetVertexAttribfv;_emscripten_glGetVertexAttribfv.sig="viip";function _glGetVertexAttribiv(index,pname,params){params>>>=0;emscriptenWebGLGetVertexAttrib(index,pname,params,5)}_glGetVertexAttribiv.sig="viip";var _emscripten_glGetVertexAttribiv=_glGetVertexAttribiv;_emscripten_glGetVertexAttribiv.sig="viip";var _glHint=(x0,x1)=>GLctx.hint(x0,x1);_glHint.sig="vii";var _emscripten_glHint=_glHint;_emscripten_glHint.sig="vii";function _glInvalidateFramebuffer(target,numAttachments,attachments){attachments>>>=0;var list=tempFixedLengthArray[numAttachments];for(var i=0;i<numAttachments;i++){list[i]=HEAP32[attachments+i*4>>>2>>>0]}GLctx.invalidateFramebuffer(target,list)}_glInvalidateFramebuffer.sig="viip";var _emscripten_glInvalidateFramebuffer=_glInvalidateFramebuffer;_emscripten_glInvalidateFramebuffer.sig="viip";function _glInvalidateSubFramebuffer(target,numAttachments,attachments,x,y,width,height){attachments>>>=0;var list=tempFixedLengthArray[numAttachments];for(var i=0;i<numAttachments;i++){list[i]=HEAP32[attachments+i*4>>>2>>>0]}GLctx.invalidateSubFramebuffer(target,list,x,y,width,height)}_glInvalidateSubFramebuffer.sig="viipiiii";var _emscripten_glInvalidateSubFramebuffer=_glInvalidateSubFramebuffer;_emscripten_glInvalidateSubFramebuffer.sig="viipiiii";var _glIsBuffer=buffer=>{var b=GL.buffers[buffer];if(!b)return 0;return GLctx.isBuffer(b)};_glIsBuffer.sig="ii";var _emscripten_glIsBuffer=_glIsBuffer;_emscripten_glIsBuffer.sig="ii";var _glIsEnabled=x0=>GLctx.isEnabled(x0);_glIsEnabled.sig="ii";var _emscripten_glIsEnabled=_glIsEnabled;_emscripten_glIsEnabled.sig="ii";var _glIsFramebuffer=framebuffer=>{var fb=GL.framebuffers[framebuffer];if(!fb)return 0;return GLctx.isFramebuffer(fb)};_glIsFramebuffer.sig="ii";var _emscripten_glIsFramebuffer=_glIsFramebuffer;_emscripten_glIsFramebuffer.sig="ii";var _glIsProgram=program=>{program=GL.programs[program];if(!program)return 0;return GLctx.isProgram(program)};_glIsProgram.sig="ii";var _emscripten_glIsProgram=_glIsProgram;_emscripten_glIsProgram.sig="ii";var _glIsQuery=id=>{var query=GL.queries[id];if(!query)return 0;return GLctx.isQuery(query)};_glIsQuery.sig="ii";var _emscripten_glIsQuery=_glIsQuery;_emscripten_glIsQuery.sig="ii";var _glIsQueryEXT=id=>{var query=GL.queries[id];if(!query)return 0;return GLctx.disjointTimerQueryExt["isQueryEXT"](query)};_glIsQueryEXT.sig="ii";var _emscripten_glIsQueryEXT=_glIsQueryEXT;var _glIsRenderbuffer=renderbuffer=>{var rb=GL.renderbuffers[renderbuffer];if(!rb)return 0;return GLctx.isRenderbuffer(rb)};_glIsRenderbuffer.sig="ii";var _emscripten_glIsRenderbuffer=_glIsRenderbuffer;_emscripten_glIsRenderbuffer.sig="ii";var _glIsSampler=id=>{var sampler=GL.samplers[id];if(!sampler)return 0;return GLctx.isSampler(sampler)};_glIsSampler.sig="ii";var _emscripten_glIsSampler=_glIsSampler;_emscripten_glIsSampler.sig="ii";var _glIsShader=shader=>{var s=GL.shaders[shader];if(!s)return 0;return GLctx.isShader(s)};_glIsShader.sig="ii";var _emscripten_glIsShader=_glIsShader;_emscripten_glIsShader.sig="ii";function _glIsSync(sync){sync>>>=0;return GLctx.isSync(GL.syncs[sync])}_glIsSync.sig="ip";var _emscripten_glIsSync=_glIsSync;_emscripten_glIsSync.sig="ip";var _glIsTexture=id=>{var texture=GL.textures[id];if(!texture)return 0;return GLctx.isTexture(texture)};_glIsTexture.sig="ii";var _emscripten_glIsTexture=_glIsTexture;_emscripten_glIsTexture.sig="ii";var _glIsTransformFeedback=id=>GLctx.isTransformFeedback(GL.transformFeedbacks[id]);_glIsTransformFeedback.sig="ii";var _emscripten_glIsTransformFeedback=_glIsTransformFeedback;_emscripten_glIsTransformFeedback.sig="ii";var _glIsVertexArray=array=>{var vao=GL.vaos[array];if(!vao)return 0;return GLctx.isVertexArray(vao)};_glIsVertexArray.sig="ii";var _emscripten_glIsVertexArray=_glIsVertexArray;_emscripten_glIsVertexArray.sig="ii";var _glIsVertexArrayOES=_glIsVertexArray;_glIsVertexArrayOES.sig="ii";var _emscripten_glIsVertexArrayOES=_glIsVertexArrayOES;_emscripten_glIsVertexArrayOES.sig="ii";var _glLineWidth=x0=>GLctx.lineWidth(x0);_glLineWidth.sig="vf";var _emscripten_glLineWidth=_glLineWidth;_emscripten_glLineWidth.sig="vf";var _glLinkProgram=program=>{program=GL.programs[program];GLctx.linkProgram(program);program.uniformLocsById=0;program.uniformSizeAndIdsByName={}};_glLinkProgram.sig="vi";var _emscripten_glLinkProgram=_glLinkProgram;_emscripten_glLinkProgram.sig="vi";var _glPauseTransformFeedback=()=>GLctx.pauseTransformFeedback();_glPauseTransformFeedback.sig="v";var _emscripten_glPauseTransformFeedback=_glPauseTransformFeedback;_emscripten_glPauseTransformFeedback.sig="v";var _glPixelStorei=(pname,param)=>{if(pname==3317){GL.unpackAlignment=param}else if(pname==3314){GL.unpackRowLength=param}GLctx.pixelStorei(pname,param)};_glPixelStorei.sig="vii";var _emscripten_glPixelStorei=_glPixelStorei;_emscripten_glPixelStorei.sig="vii";var _glPolygonModeWEBGL=(face,mode)=>{GLctx.webglPolygonMode["polygonModeWEBGL"](face,mode)};_glPolygonModeWEBGL.sig="vii";var _emscripten_glPolygonModeWEBGL=_glPolygonModeWEBGL;var _glPolygonOffset=(x0,x1)=>GLctx.polygonOffset(x0,x1);_glPolygonOffset.sig="vff";var _emscripten_glPolygonOffset=_glPolygonOffset;_emscripten_glPolygonOffset.sig="vff";var _glPolygonOffsetClampEXT=(factor,units,clamp)=>{GLctx.extPolygonOffsetClamp["polygonOffsetClampEXT"](factor,units,clamp)};_glPolygonOffsetClampEXT.sig="vfff";var _emscripten_glPolygonOffsetClampEXT=_glPolygonOffsetClampEXT;function _glProgramBinary(program,binaryFormat,binary,length){binary>>>=0;GL.recordError(1280)}_glProgramBinary.sig="viipi";var _emscripten_glProgramBinary=_glProgramBinary;_emscripten_glProgramBinary.sig="viipi";var _glProgramParameteri=(program,pname,value)=>{GL.recordError(1280)};_glProgramParameteri.sig="viii";var _emscripten_glProgramParameteri=_glProgramParameteri;_emscripten_glProgramParameteri.sig="viii";var _glQueryCounterEXT=(id,target)=>{GLctx.disjointTimerQueryExt["queryCounterEXT"](GL.queries[id],target)};_glQueryCounterEXT.sig="vii";var _emscripten_glQueryCounterEXT=_glQueryCounterEXT;var _glReadBuffer=x0=>GLctx.readBuffer(x0);_glReadBuffer.sig="vi";var _emscripten_glReadBuffer=_glReadBuffer;_emscripten_glReadBuffer.sig="vi";var computeUnpackAlignedImageSize=(width,height,sizePerPixel)=>{function roundedToNextMultipleOf(x,y){return x+y-1&-y}var plainRowSize=(GL.unpackRowLength||width)*sizePerPixel;var alignedRowSize=roundedToNextMultipleOf(plainRowSize,GL.unpackAlignment);return height*alignedRowSize};var colorChannelsInGlTextureFormat=format=>{var colorChannels={5:3,6:4,8:2,29502:3,29504:4,26917:2,26918:2,29846:3,29847:4};return colorChannels[format-6402]||1};var heapObjectForWebGLType=type=>{type-=5120;if(type==0)return HEAP8;if(type==1)return HEAPU8;if(type==2)return HEAP16;if(type==4)return HEAP32;if(type==6)return HEAPF32;if(type==5||type==28922||type==28520||type==30779||type==30782)return HEAPU32;return HEAPU16};var toTypedArrayIndex=(pointer,heap)=>pointer>>>31-Math.clz32(heap.BYTES_PER_ELEMENT);var emscriptenWebGLGetTexPixelData=(type,format,width,height,pixels,internalFormat)=>{var heap=heapObjectForWebGLType(type);var sizePerPixel=colorChannelsInGlTextureFormat(format)*heap.BYTES_PER_ELEMENT;var bytes=computeUnpackAlignedImageSize(width,height,sizePerPixel);return heap.subarray(toTypedArrayIndex(pixels,heap)>>>0,toTypedArrayIndex(pixels+bytes,heap)>>>0)};function _glReadPixels(x,y,width,height,format,type,pixels){pixels>>>=0;if(GL.currentContext.version>=2){if(GLctx.currentPixelPackBufferBinding){GLctx.readPixels(x,y,width,height,format,type,pixels);return}}var pixelData=emscriptenWebGLGetTexPixelData(type,format,width,height,pixels,format);if(!pixelData){GL.recordError(1280);return}GLctx.readPixels(x,y,width,height,format,type,pixelData)}_glReadPixels.sig="viiiiiip";var _emscripten_glReadPixels=_glReadPixels;_emscripten_glReadPixels.sig="viiiiiip";var _glReleaseShaderCompiler=()=>{};_glReleaseShaderCompiler.sig="v";var _emscripten_glReleaseShaderCompiler=_glReleaseShaderCompiler;_emscripten_glReleaseShaderCompiler.sig="v";var _glRenderbufferStorage=(x0,x1,x2,x3)=>GLctx.renderbufferStorage(x0,x1,x2,x3);_glRenderbufferStorage.sig="viiii";var _emscripten_glRenderbufferStorage=_glRenderbufferStorage;_emscripten_glRenderbufferStorage.sig="viiii";var _glRenderbufferStorageMultisample=(x0,x1,x2,x3,x4)=>GLctx.renderbufferStorageMultisample(x0,x1,x2,x3,x4);_glRenderbufferStorageMultisample.sig="viiiii";var _emscripten_glRenderbufferStorageMultisample=_glRenderbufferStorageMultisample;_emscripten_glRenderbufferStorageMultisample.sig="viiiii";var _glResumeTransformFeedback=()=>GLctx.resumeTransformFeedback();_glResumeTransformFeedback.sig="v";var _emscripten_glResumeTransformFeedback=_glResumeTransformFeedback;_emscripten_glResumeTransformFeedback.sig="v";var _glSampleCoverage=(value,invert)=>{GLctx.sampleCoverage(value,!!invert)};_glSampleCoverage.sig="vfi";var _emscripten_glSampleCoverage=_glSampleCoverage;_emscripten_glSampleCoverage.sig="vfi";var _glSamplerParameterf=(sampler,pname,param)=>{GLctx.samplerParameterf(GL.samplers[sampler],pname,param)};_glSamplerParameterf.sig="viif";var _emscripten_glSamplerParameterf=_glSamplerParameterf;_emscripten_glSamplerParameterf.sig="viif";function _glSamplerParameterfv(sampler,pname,params){params>>>=0;var param=HEAPF32[params>>>2>>>0];GLctx.samplerParameterf(GL.samplers[sampler],pname,param)}_glSamplerParameterfv.sig="viip";var _emscripten_glSamplerParameterfv=_glSamplerParameterfv;_emscripten_glSamplerParameterfv.sig="viip";var _glSamplerParameteri=(sampler,pname,param)=>{GLctx.samplerParameteri(GL.samplers[sampler],pname,param)};_glSamplerParameteri.sig="viii";var _emscripten_glSamplerParameteri=_glSamplerParameteri;_emscripten_glSamplerParameteri.sig="viii";function _glSamplerParameteriv(sampler,pname,params){params>>>=0;var param=HEAP32[params>>>2>>>0];GLctx.samplerParameteri(GL.samplers[sampler],pname,param)}_glSamplerParameteriv.sig="viip";var _emscripten_glSamplerParameteriv=_glSamplerParameteriv;_emscripten_glSamplerParameteriv.sig="viip";var _glScissor=(x0,x1,x2,x3)=>GLctx.scissor(x0,x1,x2,x3);_glScissor.sig="viiii";var _emscripten_glScissor=_glScissor;_emscripten_glScissor.sig="viiii";function _glShaderBinary(count,shaders,binaryformat,binary,length){shaders>>>=0;binary>>>=0;GL.recordError(1280)}_glShaderBinary.sig="vipipi";var _emscripten_glShaderBinary=_glShaderBinary;_emscripten_glShaderBinary.sig="vipipi";function _glShaderSource(shader,count,string,length){string>>>=0;length>>>=0;var source=GL.getSource(shader,count,string,length);GLctx.shaderSource(GL.shaders[shader],source)}_glShaderSource.sig="viipp";var _emscripten_glShaderSource=_glShaderSource;_emscripten_glShaderSource.sig="viipp";var _glStencilFunc=(x0,x1,x2)=>GLctx.stencilFunc(x0,x1,x2);_glStencilFunc.sig="viii";var _emscripten_glStencilFunc=_glStencilFunc;_emscripten_glStencilFunc.sig="viii";var _glStencilFuncSeparate=(x0,x1,x2,x3)=>GLctx.stencilFuncSeparate(x0,x1,x2,x3);_glStencilFuncSeparate.sig="viiii";var _emscripten_glStencilFuncSeparate=_glStencilFuncSeparate;_emscripten_glStencilFuncSeparate.sig="viiii";var _glStencilMask=x0=>GLctx.stencilMask(x0);_glStencilMask.sig="vi";var _emscripten_glStencilMask=_glStencilMask;_emscripten_glStencilMask.sig="vi";var _glStencilMaskSeparate=(x0,x1)=>GLctx.stencilMaskSeparate(x0,x1);_glStencilMaskSeparate.sig="vii";var _emscripten_glStencilMaskSeparate=_glStencilMaskSeparate;_emscripten_glStencilMaskSeparate.sig="vii";var _glStencilOp=(x0,x1,x2)=>GLctx.stencilOp(x0,x1,x2);_glStencilOp.sig="viii";var _emscripten_glStencilOp=_glStencilOp;_emscripten_glStencilOp.sig="viii";var _glStencilOpSeparate=(x0,x1,x2,x3)=>GLctx.stencilOpSeparate(x0,x1,x2,x3);_glStencilOpSeparate.sig="viiii";var _emscripten_glStencilOpSeparate=_glStencilOpSeparate;_emscripten_glStencilOpSeparate.sig="viiii";function _glTexImage2D(target,level,internalFormat,width,height,border,format,type,pixels){pixels>>>=0;if(GL.currentContext.version>=2){if(GLctx.currentPixelUnpackBufferBinding){GLctx.texImage2D(target,level,internalFormat,width,height,border,format,type,pixels);return}}var pixelData=pixels?emscriptenWebGLGetTexPixelData(type,format,width,height,pixels,internalFormat):null;GLctx.texImage2D(target,level,internalFormat,width,height,border,format,type,pixelData)}_glTexImage2D.sig="viiiiiiiip";var _emscripten_glTexImage2D=_glTexImage2D;_emscripten_glTexImage2D.sig="viiiiiiiip";function _glTexImage3D(target,level,internalFormat,width,height,depth,border,format,type,pixels){pixels>>>=0;if(GLctx.currentPixelUnpackBufferBinding){GLctx.texImage3D(target,level,internalFormat,width,height,depth,border,format,type,pixels)}else if(pixels){var heap=heapObjectForWebGLType(type);var pixelData=emscriptenWebGLGetTexPixelData(type,format,width,height*depth,pixels,internalFormat);GLctx.texImage3D(target,level,internalFormat,width,height,depth,border,format,type,pixelData)}else{GLctx.texImage3D(target,level,internalFormat,width,height,depth,border,format,type,null)}}_glTexImage3D.sig="viiiiiiiiip";var _emscripten_glTexImage3D=_glTexImage3D;_emscripten_glTexImage3D.sig="viiiiiiiiip";var _glTexParameterf=(x0,x1,x2)=>GLctx.texParameterf(x0,x1,x2);_glTexParameterf.sig="viif";var _emscripten_glTexParameterf=_glTexParameterf;_emscripten_glTexParameterf.sig="viif";function _glTexParameterfv(target,pname,params){params>>>=0;var param=HEAPF32[params>>>2>>>0];GLctx.texParameterf(target,pname,param)}_glTexParameterfv.sig="viip";var _emscripten_glTexParameterfv=_glTexParameterfv;_emscripten_glTexParameterfv.sig="viip";var _glTexParameteri=(x0,x1,x2)=>GLctx.texParameteri(x0,x1,x2);_glTexParameteri.sig="viii";var _emscripten_glTexParameteri=_glTexParameteri;_emscripten_glTexParameteri.sig="viii";function _glTexParameteriv(target,pname,params){params>>>=0;var param=HEAP32[params>>>2>>>0];GLctx.texParameteri(target,pname,param)}_glTexParameteriv.sig="viip";var _emscripten_glTexParameteriv=_glTexParameteriv;_emscripten_glTexParameteriv.sig="viip";var _glTexStorage2D=(x0,x1,x2,x3,x4)=>GLctx.texStorage2D(x0,x1,x2,x3,x4);_glTexStorage2D.sig="viiiii";var _emscripten_glTexStorage2D=_glTexStorage2D;_emscripten_glTexStorage2D.sig="viiiii";var _glTexStorage3D=(x0,x1,x2,x3,x4,x5)=>GLctx.texStorage3D(x0,x1,x2,x3,x4,x5);_glTexStorage3D.sig="viiiiii";var _emscripten_glTexStorage3D=_glTexStorage3D;_emscripten_glTexStorage3D.sig="viiiiii";function _glTexSubImage2D(target,level,xoffset,yoffset,width,height,format,type,pixels){pixels>>>=0;if(GL.currentContext.version>=2){if(GLctx.currentPixelUnpackBufferBinding){GLctx.texSubImage2D(target,level,xoffset,yoffset,width,height,format,type,pixels);return}}var pixelData=pixels?emscriptenWebGLGetTexPixelData(type,format,width,height,pixels,0):null;GLctx.texSubImage2D(target,level,xoffset,yoffset,width,height,format,type,pixelData)}_glTexSubImage2D.sig="viiiiiiiip";var _emscripten_glTexSubImage2D=_glTexSubImage2D;_emscripten_glTexSubImage2D.sig="viiiiiiiip";function _glTexSubImage3D(target,level,xoffset,yoffset,zoffset,width,height,depth,format,type,pixels){pixels>>>=0;if(GLctx.currentPixelUnpackBufferBinding){GLctx.texSubImage3D(target,level,xoffset,yoffset,zoffset,width,height,depth,format,type,pixels)}else if(pixels){var heap=heapObjectForWebGLType(type);GLctx.texSubImage3D(target,level,xoffset,yoffset,zoffset,width,height,depth,format,type,heap,toTypedArrayIndex(pixels,heap))}else{GLctx.texSubImage3D(target,level,xoffset,yoffset,zoffset,width,height,depth,format,type,null)}}_glTexSubImage3D.sig="viiiiiiiiiip";var _emscripten_glTexSubImage3D=_glTexSubImage3D;_emscripten_glTexSubImage3D.sig="viiiiiiiiiip";function _glTransformFeedbackVaryings(program,count,varyings,bufferMode){varyings>>>=0;program=GL.programs[program];var vars=[];for(var i=0;i<count;i++)vars.push(UTF8ToString(HEAP32[varyings+i*4>>>2>>>0]));GLctx.transformFeedbackVaryings(program,vars,bufferMode)}_glTransformFeedbackVaryings.sig="viipi";var _emscripten_glTransformFeedbackVaryings=_glTransformFeedbackVaryings;_emscripten_glTransformFeedbackVaryings.sig="viipi";var _glUniform1f=(location,v0)=>{GLctx.uniform1f(webglGetUniformLocation(location),v0)};_glUniform1f.sig="vif";var _emscripten_glUniform1f=_glUniform1f;_emscripten_glUniform1f.sig="vif";var miniTempWebGLFloatBuffers=[];function _glUniform1fv(location,count,value){value>>>=0;if(count<=288){var view=miniTempWebGLFloatBuffers[count];for(var i=0;i<count;++i){view[i]=HEAPF32[value+4*i>>>2>>>0]}}else{var view=HEAPF32.subarray(value>>>2>>>0,value+count*4>>>2>>>0)}GLctx.uniform1fv(webglGetUniformLocation(location),view)}_glUniform1fv.sig="viip";var _emscripten_glUniform1fv=_glUniform1fv;_emscripten_glUniform1fv.sig="viip";var _glUniform1i=(location,v0)=>{GLctx.uniform1i(webglGetUniformLocation(location),v0)};_glUniform1i.sig="vii";var _emscripten_glUniform1i=_glUniform1i;_emscripten_glUniform1i.sig="vii";var miniTempWebGLIntBuffers=[];function _glUniform1iv(location,count,value){value>>>=0;if(count<=288){var view=miniTempWebGLIntBuffers[count];for(var i=0;i<count;++i){view[i]=HEAP32[value+4*i>>>2>>>0]}}else{var view=HEAP32.subarray(value>>>2>>>0,value+count*4>>>2>>>0)}GLctx.uniform1iv(webglGetUniformLocation(location),view)}_glUniform1iv.sig="viip";var _emscripten_glUniform1iv=_glUniform1iv;_emscripten_glUniform1iv.sig="viip";var _glUniform1ui=(location,v0)=>{GLctx.uniform1ui(webglGetUniformLocation(location),v0)};_glUniform1ui.sig="vii";var _emscripten_glUniform1ui=_glUniform1ui;_emscripten_glUniform1ui.sig="vii";function _glUniform1uiv(location,count,value){value>>>=0;count&&GLctx.uniform1uiv(webglGetUniformLocation(location),HEAPU32,value>>>2,count)}_glUniform1uiv.sig="viip";var _emscripten_glUniform1uiv=_glUniform1uiv;_emscripten_glUniform1uiv.sig="viip";var _glUniform2f=(location,v0,v1)=>{GLctx.uniform2f(webglGetUniformLocation(location),v0,v1)};_glUniform2f.sig="viff";var _emscripten_glUniform2f=_glUniform2f;_emscripten_glUniform2f.sig="viff";function _glUniform2fv(location,count,value){value>>>=0;if(count<=144){count*=2;var view=miniTempWebGLFloatBuffers[count];for(var i=0;i<count;i+=2){view[i]=HEAPF32[value+4*i>>>2>>>0];view[i+1]=HEAPF32[value+(4*i+4)>>>2>>>0]}}else{var view=HEAPF32.subarray(value>>>2>>>0,value+count*8>>>2>>>0)}GLctx.uniform2fv(webglGetUniformLocation(location),view)}_glUniform2fv.sig="viip";var _emscripten_glUniform2fv=_glUniform2fv;_emscripten_glUniform2fv.sig="viip";var _glUniform2i=(location,v0,v1)=>{GLctx.uniform2i(webglGetUniformLocation(location),v0,v1)};_glUniform2i.sig="viii";var _emscripten_glUniform2i=_glUniform2i;_emscripten_glUniform2i.sig="viii";function _glUniform2iv(location,count,value){value>>>=0;if(count<=144){count*=2;var view=miniTempWebGLIntBuffers[count];for(var i=0;i<count;i+=2){view[i]=HEAP32[value+4*i>>>2>>>0];view[i+1]=HEAP32[value+(4*i+4)>>>2>>>0]}}else{var view=HEAP32.subarray(value>>>2>>>0,value+count*8>>>2>>>0)}GLctx.uniform2iv(webglGetUniformLocation(location),view)}_glUniform2iv.sig="viip";var _emscripten_glUniform2iv=_glUniform2iv;_emscripten_glUniform2iv.sig="viip";var _glUniform2ui=(location,v0,v1)=>{GLctx.uniform2ui(webglGetUniformLocation(location),v0,v1)};_glUniform2ui.sig="viii";var _emscripten_glUniform2ui=_glUniform2ui;_emscripten_glUniform2ui.sig="viii";function _glUniform2uiv(location,count,value){value>>>=0;count&&GLctx.uniform2uiv(webglGetUniformLocation(location),HEAPU32,value>>>2,count*2)}_glUniform2uiv.sig="viip";var _emscripten_glUniform2uiv=_glUniform2uiv;_emscripten_glUniform2uiv.sig="viip";var _glUniform3f=(location,v0,v1,v2)=>{GLctx.uniform3f(webglGetUniformLocation(location),v0,v1,v2)};_glUniform3f.sig="vifff";var _emscripten_glUniform3f=_glUniform3f;_emscripten_glUniform3f.sig="vifff";function _glUniform3fv(location,count,value){value>>>=0;if(count<=96){count*=3;var view=miniTempWebGLFloatBuffers[count];for(var i=0;i<count;i+=3){view[i]=HEAPF32[value+4*i>>>2>>>0];view[i+1]=HEAPF32[value+(4*i+4)>>>2>>>0];view[i+2]=HEAPF32[value+(4*i+8)>>>2>>>0]}}else{var view=HEAPF32.subarray(value>>>2>>>0,value+count*12>>>2>>>0)}GLctx.uniform3fv(webglGetUniformLocation(location),view)}_glUniform3fv.sig="viip";var _emscripten_glUniform3fv=_glUniform3fv;_emscripten_glUniform3fv.sig="viip";var _glUniform3i=(location,v0,v1,v2)=>{GLctx.uniform3i(webglGetUniformLocation(location),v0,v1,v2)};_glUniform3i.sig="viiii";var _emscripten_glUniform3i=_glUniform3i;_emscripten_glUniform3i.sig="viiii";function _glUniform3iv(location,count,value){value>>>=0;if(count<=96){count*=3;var view=miniTempWebGLIntBuffers[count];for(var i=0;i<count;i+=3){view[i]=HEAP32[value+4*i>>>2>>>0];view[i+1]=HEAP32[value+(4*i+4)>>>2>>>0];view[i+2]=HEAP32[value+(4*i+8)>>>2>>>0]}}else{var view=HEAP32.subarray(value>>>2>>>0,value+count*12>>>2>>>0)}GLctx.uniform3iv(webglGetUniformLocation(location),view)}_glUniform3iv.sig="viip";var _emscripten_glUniform3iv=_glUniform3iv;_emscripten_glUniform3iv.sig="viip";var _glUniform3ui=(location,v0,v1,v2)=>{GLctx.uniform3ui(webglGetUniformLocation(location),v0,v1,v2)};_glUniform3ui.sig="viiii";var _emscripten_glUniform3ui=_glUniform3ui;_emscripten_glUniform3ui.sig="viiii";function _glUniform3uiv(location,count,value){value>>>=0;count&&GLctx.uniform3uiv(webglGetUniformLocation(location),HEAPU32,value>>>2,count*3)}_glUniform3uiv.sig="viip";var _emscripten_glUniform3uiv=_glUniform3uiv;_emscripten_glUniform3uiv.sig="viip";var _glUniform4f=(location,v0,v1,v2,v3)=>{GLctx.uniform4f(webglGetUniformLocation(location),v0,v1,v2,v3)};_glUniform4f.sig="viffff";var _emscripten_glUniform4f=_glUniform4f;_emscripten_glUniform4f.sig="viffff";function _glUniform4fv(location,count,value){value>>>=0;if(count<=72){var view=miniTempWebGLFloatBuffers[4*count];var heap=HEAPF32;value=value>>>2;count*=4;for(var i=0;i<count;i+=4){var dst=value+i;view[i]=heap[dst>>>0];view[i+1]=heap[dst+1>>>0];view[i+2]=heap[dst+2>>>0];view[i+3]=heap[dst+3>>>0]}}else{var view=HEAPF32.subarray(value>>>2>>>0,value+count*16>>>2>>>0)}GLctx.uniform4fv(webglGetUniformLocation(location),view)}_glUniform4fv.sig="viip";var _emscripten_glUniform4fv=_glUniform4fv;_emscripten_glUniform4fv.sig="viip";var _glUniform4i=(location,v0,v1,v2,v3)=>{GLctx.uniform4i(webglGetUniformLocation(location),v0,v1,v2,v3)};_glUniform4i.sig="viiiii";var _emscripten_glUniform4i=_glUniform4i;_emscripten_glUniform4i.sig="viiiii";function _glUniform4iv(location,count,value){value>>>=0;if(count<=72){count*=4;var view=miniTempWebGLIntBuffers[count];for(var i=0;i<count;i+=4){view[i]=HEAP32[value+4*i>>>2>>>0];view[i+1]=HEAP32[value+(4*i+4)>>>2>>>0];view[i+2]=HEAP32[value+(4*i+8)>>>2>>>0];view[i+3]=HEAP32[value+(4*i+12)>>>2>>>0]}}else{var view=HEAP32.subarray(value>>>2>>>0,value+count*16>>>2>>>0)}GLctx.uniform4iv(webglGetUniformLocation(location),view)}_glUniform4iv.sig="viip";var _emscripten_glUniform4iv=_glUniform4iv;_emscripten_glUniform4iv.sig="viip";var _glUniform4ui=(location,v0,v1,v2,v3)=>{GLctx.uniform4ui(webglGetUniformLocation(location),v0,v1,v2,v3)};_glUniform4ui.sig="viiiii";var _emscripten_glUniform4ui=_glUniform4ui;_emscripten_glUniform4ui.sig="viiiii";function _glUniform4uiv(location,count,value){value>>>=0;count&&GLctx.uniform4uiv(webglGetUniformLocation(location),HEAPU32,value>>>2,count*4)}_glUniform4uiv.sig="viip";var _emscripten_glUniform4uiv=_glUniform4uiv;_emscripten_glUniform4uiv.sig="viip";var _glUniformBlockBinding=(program,uniformBlockIndex,uniformBlockBinding)=>{program=GL.programs[program];GLctx.uniformBlockBinding(program,uniformBlockIndex,uniformBlockBinding)};_glUniformBlockBinding.sig="viii";var _emscripten_glUniformBlockBinding=_glUniformBlockBinding;_emscripten_glUniformBlockBinding.sig="viii";function _glUniformMatrix2fv(location,count,transpose,value){value>>>=0;if(count<=72){count*=4;var view=miniTempWebGLFloatBuffers[count];for(var i=0;i<count;i+=4){view[i]=HEAPF32[value+4*i>>>2>>>0];view[i+1]=HEAPF32[value+(4*i+4)>>>2>>>0];view[i+2]=HEAPF32[value+(4*i+8)>>>2>>>0];view[i+3]=HEAPF32[value+(4*i+12)>>>2>>>0]}}else{var view=HEAPF32.subarray(value>>>2>>>0,value+count*16>>>2>>>0)}GLctx.uniformMatrix2fv(webglGetUniformLocation(location),!!transpose,view)}_glUniformMatrix2fv.sig="viiip";var _emscripten_glUniformMatrix2fv=_glUniformMatrix2fv;_emscripten_glUniformMatrix2fv.sig="viiip";function _glUniformMatrix2x3fv(location,count,transpose,value){value>>>=0;count&&GLctx.uniformMatrix2x3fv(webglGetUniformLocation(location),!!transpose,HEAPF32,value>>>2,count*6)}_glUniformMatrix2x3fv.sig="viiip";var _emscripten_glUniformMatrix2x3fv=_glUniformMatrix2x3fv;_emscripten_glUniformMatrix2x3fv.sig="viiip";function _glUniformMatrix2x4fv(location,count,transpose,value){value>>>=0;count&&GLctx.uniformMatrix2x4fv(webglGetUniformLocation(location),!!transpose,HEAPF32,value>>>2,count*8)}_glUniformMatrix2x4fv.sig="viiip";var _emscripten_glUniformMatrix2x4fv=_glUniformMatrix2x4fv;_emscripten_glUniformMatrix2x4fv.sig="viiip";function _glUniformMatrix3fv(location,count,transpose,value){value>>>=0;if(count<=32){count*=9;var view=miniTempWebGLFloatBuffers[count];for(var i=0;i<count;i+=9){view[i]=HEAPF32[value+4*i>>>2>>>0];view[i+1]=HEAPF32[value+(4*i+4)>>>2>>>0];view[i+2]=HEAPF32[value+(4*i+8)>>>2>>>0];view[i+3]=HEAPF32[value+(4*i+12)>>>2>>>0];view[i+4]=HEAPF32[value+(4*i+16)>>>2>>>0];view[i+5]=HEAPF32[value+(4*i+20)>>>2>>>0];view[i+6]=HEAPF32[value+(4*i+24)>>>2>>>0];view[i+7]=HEAPF32[value+(4*i+28)>>>2>>>0];view[i+8]=HEAPF32[value+(4*i+32)>>>2>>>0]}}else{var view=HEAPF32.subarray(value>>>2>>>0,value+count*36>>>2>>>0)}GLctx.uniformMatrix3fv(webglGetUniformLocation(location),!!transpose,view)}_glUniformMatrix3fv.sig="viiip";var _emscripten_glUniformMatrix3fv=_glUniformMatrix3fv;_emscripten_glUniformMatrix3fv.sig="viiip";function _glUniformMatrix3x2fv(location,count,transpose,value){value>>>=0;count&&GLctx.uniformMatrix3x2fv(webglGetUniformLocation(location),!!transpose,HEAPF32,value>>>2,count*6)}_glUniformMatrix3x2fv.sig="viiip";var _emscripten_glUniformMatrix3x2fv=_glUniformMatrix3x2fv;_emscripten_glUniformMatrix3x2fv.sig="viiip";function _glUniformMatrix3x4fv(location,count,transpose,value){value>>>=0;count&&GLctx.uniformMatrix3x4fv(webglGetUniformLocation(location),!!transpose,HEAPF32,value>>>2,count*12)}_glUniformMatrix3x4fv.sig="viiip";var _emscripten_glUniformMatrix3x4fv=_glUniformMatrix3x4fv;_emscripten_glUniformMatrix3x4fv.sig="viiip";function _glUniformMatrix4fv(location,count,transpose,value){value>>>=0;if(count<=18){var view=miniTempWebGLFloatBuffers[16*count];var heap=HEAPF32;value=value>>>2;count*=16;for(var i=0;i<count;i+=16){var dst=value+i;view[i]=heap[dst>>>0];view[i+1]=heap[dst+1>>>0];view[i+2]=heap[dst+2>>>0];view[i+3]=heap[dst+3>>>0];view[i+4]=heap[dst+4>>>0];view[i+5]=heap[dst+5>>>0];view[i+6]=heap[dst+6>>>0];view[i+7]=heap[dst+7>>>0];view[i+8]=heap[dst+8>>>0];view[i+9]=heap[dst+9>>>0];view[i+10]=heap[dst+10>>>0];view[i+11]=heap[dst+11>>>0];view[i+12]=heap[dst+12>>>0];view[i+13]=heap[dst+13>>>0];view[i+14]=heap[dst+14>>>0];view[i+15]=heap[dst+15>>>0]}}else{var view=HEAPF32.subarray(value>>>2>>>0,value+count*64>>>2>>>0)}GLctx.uniformMatrix4fv(webglGetUniformLocation(location),!!transpose,view)}_glUniformMatrix4fv.sig="viiip";var _emscripten_glUniformMatrix4fv=_glUniformMatrix4fv;_emscripten_glUniformMatrix4fv.sig="viiip";function _glUniformMatrix4x2fv(location,count,transpose,value){value>>>=0;count&&GLctx.uniformMatrix4x2fv(webglGetUniformLocation(location),!!transpose,HEAPF32,value>>>2,count*8)}_glUniformMatrix4x2fv.sig="viiip";var _emscripten_glUniformMatrix4x2fv=_glUniformMatrix4x2fv;_emscripten_glUniformMatrix4x2fv.sig="viiip";function _glUniformMatrix4x3fv(location,count,transpose,value){value>>>=0;count&&GLctx.uniformMatrix4x3fv(webglGetUniformLocation(location),!!transpose,HEAPF32,value>>>2,count*12)}_glUniformMatrix4x3fv.sig="viiip";var _emscripten_glUniformMatrix4x3fv=_glUniformMatrix4x3fv;_emscripten_glUniformMatrix4x3fv.sig="viiip";var _glUseProgram=program=>{program=GL.programs[program];GLctx.useProgram(program);GLctx.currentProgram=program};_glUseProgram.sig="vi";var _emscripten_glUseProgram=_glUseProgram;_emscripten_glUseProgram.sig="vi";var _glValidateProgram=program=>{GLctx.validateProgram(GL.programs[program])};_glValidateProgram.sig="vi";var _emscripten_glValidateProgram=_glValidateProgram;_emscripten_glValidateProgram.sig="vi";var _glVertexAttrib1f=(x0,x1)=>GLctx.vertexAttrib1f(x0,x1);_glVertexAttrib1f.sig="vif";var _emscripten_glVertexAttrib1f=_glVertexAttrib1f;_emscripten_glVertexAttrib1f.sig="vif";function _glVertexAttrib1fv(index,v){v>>>=0;GLctx.vertexAttrib1f(index,HEAPF32[v>>>2])}_glVertexAttrib1fv.sig="vip";var _emscripten_glVertexAttrib1fv=_glVertexAttrib1fv;_emscripten_glVertexAttrib1fv.sig="vip";var _glVertexAttrib2f=(x0,x1,x2)=>GLctx.vertexAttrib2f(x0,x1,x2);_glVertexAttrib2f.sig="viff";var _emscripten_glVertexAttrib2f=_glVertexAttrib2f;_emscripten_glVertexAttrib2f.sig="viff";function _glVertexAttrib2fv(index,v){v>>>=0;GLctx.vertexAttrib2f(index,HEAPF32[v>>>2],HEAPF32[v+4>>>2])}_glVertexAttrib2fv.sig="vip";var _emscripten_glVertexAttrib2fv=_glVertexAttrib2fv;_emscripten_glVertexAttrib2fv.sig="vip";var _glVertexAttrib3f=(x0,x1,x2,x3)=>GLctx.vertexAttrib3f(x0,x1,x2,x3);_glVertexAttrib3f.sig="vifff";var _emscripten_glVertexAttrib3f=_glVertexAttrib3f;_emscripten_glVertexAttrib3f.sig="vifff";function _glVertexAttrib3fv(index,v){v>>>=0;GLctx.vertexAttrib3f(index,HEAPF32[v>>>2],HEAPF32[v+4>>>2],HEAPF32[v+8>>>2])}_glVertexAttrib3fv.sig="vip";var _emscripten_glVertexAttrib3fv=_glVertexAttrib3fv;_emscripten_glVertexAttrib3fv.sig="vip";var _glVertexAttrib4f=(x0,x1,x2,x3,x4)=>GLctx.vertexAttrib4f(x0,x1,x2,x3,x4);_glVertexAttrib4f.sig="viffff";var _emscripten_glVertexAttrib4f=_glVertexAttrib4f;_emscripten_glVertexAttrib4f.sig="viffff";function _glVertexAttrib4fv(index,v){v>>>=0;GLctx.vertexAttrib4f(index,HEAPF32[v>>>2],HEAPF32[v+4>>>2],HEAPF32[v+8>>>2],HEAPF32[v+12>>>2])}_glVertexAttrib4fv.sig="vip";var _emscripten_glVertexAttrib4fv=_glVertexAttrib4fv;_emscripten_glVertexAttrib4fv.sig="vip";var _glVertexAttribDivisor=(index,divisor)=>{GLctx.vertexAttribDivisor(index,divisor)};_glVertexAttribDivisor.sig="vii";var _emscripten_glVertexAttribDivisor=_glVertexAttribDivisor;_emscripten_glVertexAttribDivisor.sig="vii";var _glVertexAttribDivisorANGLE=_glVertexAttribDivisor;var _emscripten_glVertexAttribDivisorANGLE=_glVertexAttribDivisorANGLE;var _glVertexAttribDivisorARB=_glVertexAttribDivisor;var _emscripten_glVertexAttribDivisorARB=_glVertexAttribDivisorARB;var _glVertexAttribDivisorEXT=_glVertexAttribDivisor;var _emscripten_glVertexAttribDivisorEXT=_glVertexAttribDivisorEXT;var _glVertexAttribDivisorNV=_glVertexAttribDivisor;var _emscripten_glVertexAttribDivisorNV=_glVertexAttribDivisorNV;var _glVertexAttribI4i=(x0,x1,x2,x3,x4)=>GLctx.vertexAttribI4i(x0,x1,x2,x3,x4);_glVertexAttribI4i.sig="viiiii";var _emscripten_glVertexAttribI4i=_glVertexAttribI4i;_emscripten_glVertexAttribI4i.sig="viiiii";function _glVertexAttribI4iv(index,v){v>>>=0;GLctx.vertexAttribI4i(index,HEAP32[v>>>2],HEAP32[v+4>>>2],HEAP32[v+8>>>2],HEAP32[v+12>>>2])}_glVertexAttribI4iv.sig="vip";var _emscripten_glVertexAttribI4iv=_glVertexAttribI4iv;_emscripten_glVertexAttribI4iv.sig="vip";var _glVertexAttribI4ui=(x0,x1,x2,x3,x4)=>GLctx.vertexAttribI4ui(x0,x1,x2,x3,x4);_glVertexAttribI4ui.sig="viiiii";var _emscripten_glVertexAttribI4ui=_glVertexAttribI4ui;_emscripten_glVertexAttribI4ui.sig="viiiii";function _glVertexAttribI4uiv(index,v){v>>>=0;GLctx.vertexAttribI4ui(index,HEAPU32[v>>>2],HEAPU32[v+4>>>2],HEAPU32[v+8>>>2],HEAPU32[v+12>>>2])}_glVertexAttribI4uiv.sig="vip";var _emscripten_glVertexAttribI4uiv=_glVertexAttribI4uiv;_emscripten_glVertexAttribI4uiv.sig="vip";function _glVertexAttribIPointer(index,size,type,stride,ptr){ptr>>>=0;GLctx.vertexAttribIPointer(index,size,type,stride,ptr)}_glVertexAttribIPointer.sig="viiiip";var _emscripten_glVertexAttribIPointer=_glVertexAttribIPointer;_emscripten_glVertexAttribIPointer.sig="viiiip";function _glVertexAttribPointer(index,size,type,normalized,stride,ptr){ptr>>>=0;GLctx.vertexAttribPointer(index,size,type,!!normalized,stride,ptr)}_glVertexAttribPointer.sig="viiiiip";var _emscripten_glVertexAttribPointer=_glVertexAttribPointer;_emscripten_glVertexAttribPointer.sig="viiiiip";var _glViewport=(x0,x1,x2,x3)=>GLctx.viewport(x0,x1,x2,x3);_glViewport.sig="viiii";var _emscripten_glViewport=_glViewport;_emscripten_glViewport.sig="viiii";function _glWaitSync(sync,flags,timeout){sync>>>=0;timeout=Number(timeout);GLctx.waitSync(GL.syncs[sync],flags,timeout)}_glWaitSync.sig="vpij";var _emscripten_glWaitSync=_glWaitSync;_emscripten_glWaitSync.sig="vpij";function _emscripten_out(str){str>>>=0;return out(UTF8ToString(str))}_emscripten_out.sig="vp";class HandleAllocator{allocated=[undefined];freelist=[];get(id){return this.allocated[id]}has(id){return this.allocated[id]!==undefined}allocate(handle){var id=this.freelist.pop()||this.allocated.length;this.allocated[id]=handle;return id}free(id){this.allocated[id]=undefined;this.freelist.push(id)}}var promiseMap=new HandleAllocator;var makePromise=()=>{var promiseInfo={};promiseInfo.promise=new Promise((resolve,reject)=>{promiseInfo.reject=reject;promiseInfo.resolve=resolve});promiseInfo.id=promiseMap.allocate(promiseInfo);return promiseInfo};function _emscripten_promise_create(){return makePromise().id}_emscripten_promise_create.sig="p";function _emscripten_promise_destroy(id){id>>>=0;promiseMap.free(id)}_emscripten_promise_destroy.sig="vp";var getPromise=id=>promiseMap.get(id).promise;function _emscripten_promise_resolve(id,result,value){id>>>=0;value>>>=0;var info=promiseMap.get(id);switch(result){case 0:info.resolve(value);return;case 1:info.resolve(getPromise(value));return;case 2:info.resolve(getPromise(value));_emscripten_promise_destroy(value);return;case 3:info.reject(value);return}}_emscripten_promise_resolve.sig="vpip";var growMemory=size=>{var b=wasmMemory.buffer;var pages=(size-b.byteLength+65535)/65536|0;try{wasmMemory.grow(pages);updateMemoryViews();return 1}catch(e){}};function _emscripten_resize_heap(requestedSize){requestedSize>>>=0;var oldSize=HEAPU8.length;var maxHeapSize=getHeapMax();if(requestedSize>maxHeapSize){return false}for(var cutDown=1;cutDown<=4;cutDown*=2){var overGrownHeapSize=oldSize*(1+.2/cutDown);overGrownHeapSize=Math.min(overGrownHeapSize,requestedSize+100663296);var newSize=Math.min(maxHeapSize,alignMemory(Math.max(requestedSize,overGrownHeapSize),65536));var replacement=growMemory(newSize);if(replacement){return true}}return false}_emscripten_resize_heap.sig="ip";var _emscripten_runtime_keepalive_pop=runtimeKeepalivePop;_emscripten_runtime_keepalive_pop.sig="v";var _emscripten_runtime_keepalive_push=runtimeKeepalivePush;_emscripten_runtime_keepalive_push.sig="v";var ENV={};var getEnvStrings=()=>{if(!getEnvStrings.strings){var lang=(typeof navigator=="object"&&navigator.languages&&navigator.languages[0]||"C").replace("-","_")+".UTF-8";var env={USER:"web_user",LOGNAME:"web_user",PATH:"/",PWD:"/",HOME:"/home/web_user",LANG:lang,_:getExecutableName()};for(var x in ENV){if(ENV[x]===undefined)delete env[x];else env[x]=ENV[x]}var strings=[];for(var x in env){strings.push(`${x}=${env[x]}`)}getEnvStrings.strings=strings}return getEnvStrings.strings};function _environ_get(__environ,environ_buf){__environ>>>=0;environ_buf>>>=0;var bufSize=0;var envp=0;for(var string of getEnvStrings()){var ptr=environ_buf+bufSize;HEAPU32[__environ+envp>>>2>>>0]=ptr;bufSize+=stringToUTF8(string,ptr,Infinity)+1;envp+=4}return 0}_environ_get.sig="ipp";function _environ_sizes_get(penviron_count,penviron_buf_size){penviron_count>>>=0;penviron_buf_size>>>=0;var strings=getEnvStrings();HEAPU32[penviron_count>>>2>>>0]=strings.length;var bufSize=0;for(var string of strings){bufSize+=lengthBytesUTF8(string)+1}HEAPU32[penviron_buf_size>>>2>>>0]=bufSize;return 0}_environ_sizes_get.sig="ipp";function _fd_close(fd){try{var stream=SYSCALLS.getStreamFromFD(fd);FS.close(stream);return 0}catch(e){if(typeof FS=="undefined"||!(e.name==="ErrnoError"))throw e;return e.errno}}_fd_close.sig="ii";function _fd_fdstat_get(fd,pbuf){pbuf>>>=0;try{var rightsBase=0;var rightsInheriting=0;var flags=0;{var stream=SYSCALLS.getStreamFromFD(fd);var type=stream.tty?2:FS.isDir(stream.mode)?3:FS.isLink(stream.mode)?7:4}HEAP8[pbuf>>>0]=type;HEAP16[pbuf+2>>>1>>>0]=flags;HEAP64[pbuf+8>>>3>>>0]=BigInt(rightsBase);HEAP64[pbuf+16>>>3>>>0]=BigInt(rightsInheriting);return 0}catch(e){if(typeof FS=="undefined"||!(e.name==="ErrnoError"))throw e;return e.errno}}_fd_fdstat_get.sig="iip";var doReadv=(stream,iov,iovcnt,offset)=>{var ret=0;for(var i=0;i<iovcnt;i++){var ptr=HEAPU32[iov>>>2>>>0];var len=HEAPU32[iov+4>>>2>>>0];iov+=8;var curr=FS.read(stream,HEAP8,ptr,len,offset);if(curr<0)return-1;ret+=curr;if(curr<len)break;if(typeof offset!="undefined"){offset+=curr}}return ret};function _fd_pread(fd,iov,iovcnt,offset,pnum){iov>>>=0;iovcnt>>>=0;offset=bigintToI53Checked(offset);pnum>>>=0;try{if(isNaN(offset))return 61;var stream=SYSCALLS.getStreamFromFD(fd);var num=doReadv(stream,iov,iovcnt,offset);HEAPU32[pnum>>>2>>>0]=num;return 0}catch(e){if(typeof FS=="undefined"||!(e.name==="ErrnoError"))throw e;return e.errno}}_fd_pread.sig="iippjp";var doWritev=(stream,iov,iovcnt,offset)=>{var ret=0;for(var i=0;i<iovcnt;i++){var ptr=HEAPU32[iov>>>2>>>0];var len=HEAPU32[iov+4>>>2>>>0];iov+=8;var curr=FS.write(stream,HEAP8,ptr,len,offset);if(curr<0)return-1;ret+=curr;if(curr<len){break}if(typeof offset!="undefined"){offset+=curr}}return ret};function _fd_pwrite(fd,iov,iovcnt,offset,pnum){iov>>>=0;iovcnt>>>=0;offset=bigintToI53Checked(offset);pnum>>>=0;try{if(isNaN(offset))return 61;var stream=SYSCALLS.getStreamFromFD(fd);var num=doWritev(stream,iov,iovcnt,offset);HEAPU32[pnum>>>2>>>0]=num;return 0}catch(e){if(typeof FS=="undefined"||!(e.name==="ErrnoError"))throw e;return e.errno}}_fd_pwrite.sig="iippjp";function _fd_read(fd,iov,iovcnt,pnum){iov>>>=0;iovcnt>>>=0;pnum>>>=0;try{var stream=SYSCALLS.getStreamFromFD(fd);var num=doReadv(stream,iov,iovcnt);HEAPU32[pnum>>>2>>>0]=num;return 0}catch(e){if(typeof FS=="undefined"||!(e.name==="ErrnoError"))throw e;return e.errno}}_fd_read.sig="iippp";function _fd_seek(fd,offset,whence,newOffset){offset=bigintToI53Checked(offset);newOffset>>>=0;try{if(isNaN(offset))return 61;var stream=SYSCALLS.getStreamFromFD(fd);FS.llseek(stream,offset,whence);HEAP64[newOffset>>>3>>>0]=BigInt(stream.position);if(stream.getdents&&offset===0&&whence===0)stream.getdents=null;return 0}catch(e){if(typeof FS=="undefined"||!(e.name==="ErrnoError"))throw e;return e.errno}}_fd_seek.sig="iijip";function _fd_sync(fd){try{var stream=SYSCALLS.getStreamFromFD(fd);if(stream.stream_ops?.fsync){return stream.stream_ops.fsync(stream)}return 0}catch(e){if(typeof FS=="undefined"||!(e.name==="ErrnoError"))throw e;return e.errno}}_fd_sync.sig="ii";function _fd_write(fd,iov,iovcnt,pnum){iov>>>=0;iovcnt>>>=0;pnum>>>=0;try{var stream=SYSCALLS.getStreamFromFD(fd);var num=doWritev(stream,iov,iovcnt);HEAPU32[pnum>>>2>>>0]=num;return 0}catch(e){if(typeof FS=="undefined"||!(e.name==="ErrnoError"))throw e;return e.errno}}_fd_write.sig="iippp";function _getaddrinfo(node,service,hint,out){node>>>=0;service>>>=0;hint>>>=0;out>>>=0;var addrs=[];var canon=null;var addr=0;var port=0;var flags=0;var family=0;var type=0;var proto=0;var ai,last;function allocaddrinfo(family,type,proto,canon,addr,port){var sa,salen,ai;var errno;salen=family===10?28:16;addr=family===10?inetNtop6(addr):inetNtop4(addr);sa=_malloc(salen);errno=writeSockaddr(sa,family,addr,port);assert(!errno);ai=_malloc(32);HEAP32[ai+4>>>2>>>0]=family;HEAP32[ai+8>>>2>>>0]=type;HEAP32[ai+12>>>2>>>0]=proto;HEAPU32[ai+24>>>2>>>0]=canon;HEAPU32[ai+20>>>2>>>0]=sa;if(family===10){HEAP32[ai+16>>>2>>>0]=28}else{HEAP32[ai+16>>>2>>>0]=16}HEAP32[ai+28>>>2>>>0]=0;return ai}if(hint){flags=HEAP32[hint>>>2>>>0];family=HEAP32[hint+4>>>2>>>0];type=HEAP32[hint+8>>>2>>>0];proto=HEAP32[hint+12>>>2>>>0]}if(type&&!proto){proto=type===2?17:6}if(!type&&proto){type=proto===17?2:1}if(proto===0){proto=6}if(type===0){type=1}if(!node&&!service){return-2}if(flags&~(1|2|4|1024|8|16|32)){return-1}if(hint!==0&&HEAP32[hint>>>2>>>0]&2&&!node){return-1}if(flags&32){return-2}if(type!==0&&type!==1&&type!==2){return-7}if(family!==0&&family!==2&&family!==10){return-6}if(service){service=UTF8ToString(service);port=parseInt(service,10);if(isNaN(port)){if(flags&1024){return-2}return-8}}if(!node){if(family===0){family=2}if((flags&1)===0){if(family===2){addr=_htonl(2130706433)}else{addr=[0,0,0,_htonl(1)]}}ai=allocaddrinfo(family,type,proto,null,addr,port);HEAPU32[out>>>2>>>0]=ai;return 0}node=UTF8ToString(node);addr=inetPton4(node);if(addr!==null){if(family===0||family===2){family=2}else if(family===10&&flags&8){addr=[0,0,_htonl(65535),addr];family=10}else{return-2}}else{addr=inetPton6(node);if(addr!==null){if(family===0||family===10){family=10}else{return-2}}}if(addr!=null){ai=allocaddrinfo(family,type,proto,node,addr,port);HEAPU32[out>>>2>>>0]=ai;return 0}if(flags&4){return-2}node=DNS.lookup_name(node);addr=inetPton4(node);if(family===0){family=2}else if(family===10){addr=[0,0,_htonl(65535),addr]}ai=allocaddrinfo(family,type,proto,null,addr,port);HEAPU32[out>>>2>>>0]=ai;return 0}_getaddrinfo.sig="ipppp";function _getnameinfo(sa,salen,node,nodelen,serv,servlen,flags){sa>>>=0;node>>>=0;serv>>>=0;var info=readSockaddr(sa,salen);if(info.errno){return-6}var port=info.port;var addr=info.addr;var overflowed=false;if(node&&nodelen){var lookup;if(flags&1||!(lookup=DNS.lookup_addr(addr))){if(flags&8){return-2}}else{addr=lookup}var numBytesWrittenExclNull=stringToUTF8(addr,node,nodelen);if(numBytesWrittenExclNull+1>=nodelen){overflowed=true}}if(serv&&servlen){port=""+port;var numBytesWrittenExclNull=stringToUTF8(port,serv,servlen);if(numBytesWrittenExclNull+1>=servlen){overflowed=true}}if(overflowed){return-12}return 0}_getnameinfo.sig="ipipipii";var Protocols={list:[],map:{}};var stringToAscii=(str,buffer)=>{for(var i=0;i<str.length;++i){HEAP8[buffer++>>>0]=str.charCodeAt(i)}HEAP8[buffer>>>0]=0};var _setprotoent=stayopen=>{function allocprotoent(name,proto,aliases){var nameBuf=_malloc(name.length+1);stringToAscii(name,nameBuf);var j=0;var length=aliases.length;var aliasListBuf=_malloc((length+1)*4);for(var i=0;i<length;i++,j+=4){var alias=aliases[i];var aliasBuf=_malloc(alias.length+1);stringToAscii(alias,aliasBuf);HEAPU32[aliasListBuf+j>>>2>>>0]=aliasBuf}HEAPU32[aliasListBuf+j>>>2>>>0]=0;var pe=_malloc(12);HEAPU32[pe>>>2>>>0]=nameBuf;HEAPU32[pe+4>>>2>>>0]=aliasListBuf;HEAP32[pe+8>>>2>>>0]=proto;return pe}var list=Protocols.list;var map=Protocols.map;if(list.length===0){var entry=allocprotoent("tcp",6,["TCP"]);list.push(entry);map["tcp"]=map["6"]=entry;entry=allocprotoent("udp",17,["UDP"]);list.push(entry);map["udp"]=map["17"]=entry}_setprotoent.index=0};_setprotoent.sig="vi";function _getprotobyname(name){name>>>=0;name=UTF8ToString(name);_setprotoent(true);var result=Protocols.map[name];return result}_getprotobyname.sig="pp";function _is_sentinel(...args){return wasmImports["is_sentinel"](...args)}_is_sentinel.stub=true;function _random_get(buffer,size){buffer>>>=0;size>>>=0;try{randomFill(HEAPU8.subarray(buffer>>>0,buffer+size>>>0));return 0}catch(e){if(typeof FS=="undefined"||!(e.name==="ErrnoError"))throw e;return e.errno}}_random_get.sig="ipp";var _stackAlloc=stackAlloc;var _stackRestore=stackSave;var _stackSave=stackSave;var FS_createPath=(...args)=>FS.createPath(...args);var FS_unlink=(...args)=>FS.unlink(...args);var FS_createLazyFile=(...args)=>FS.createLazyFile(...args);var FS_createDevice=(...args)=>FS.createDevice(...args);var writeI53ToI64Clamped=(ptr,num)=>{if(num>0x8000000000000000){HEAPU32[ptr>>>2>>>0]=4294967295;HEAPU32[ptr+4>>>2>>>0]=2147483647}else if(num<-0x8000000000000000){HEAPU32[ptr>>>2>>>0]=0;HEAPU32[ptr+4>>>2>>>0]=2147483648}else{writeI53ToI64(ptr,num)}};var writeI53ToI64Signaling=(ptr,num)=>{if(num>0x8000000000000000||num<-0x8000000000000000){throw`RangeError: ${num}`}writeI53ToI64(ptr,num)};var writeI53ToU64Clamped=(ptr,num)=>{if(num>0x10000000000000000){HEAPU32[ptr>>>2>>>0]=4294967295;HEAPU32[ptr+4>>>2>>>0]=4294967295}else if(num<0){HEAPU32[ptr>>>2>>>0]=0;HEAPU32[ptr+4>>>2>>>0]=0}else{writeI53ToI64(ptr,num)}};var writeI53ToU64Signaling=(ptr,num)=>{if(num<0||num>0x10000000000000000){throw`RangeError: ${num}`}writeI53ToI64(ptr,num)};var readI53FromU64=ptr=>HEAPU32[ptr>>>2>>>0]+HEAPU32[ptr+4>>>2>>>0]*4294967296;var convertI32PairToI53=(lo,hi)=>(lo>>>0)+hi*4294967296;var convertI32PairToI53Checked=(lo,hi)=>hi+2097152>>>0<4194305-!!lo?(lo>>>0)+hi*4294967296:NaN;var convertU32PairToI53=(lo,hi)=>(lo>>>0)+(hi>>>0)*4294967296;var getTempRet0=val=>__emscripten_tempret_get();var setTempRet0=val=>__emscripten_tempret_set(val);var ptrToString=ptr=>"0x"+ptr.toString(16).padStart(8,"0");function _emscripten_notify_memory_growth(memoryIndex){memoryIndex>>>=0;updateMemoryViews()}_emscripten_notify_memory_growth.sig="vp";var strError=errno=>UTF8ToString(_strerror(errno));var _endprotoent=()=>{};_endprotoent.sig="v";function _getprotoent(number){if(_setprotoent.index===Protocols.list.length){return 0}var result=Protocols.list[_setprotoent.index++];return result}_getprotoent.sig="p";function _getprotobynumber(number){_setprotoent(true);var result=Protocols.map[number];return result}_getprotobynumber.sig="pi";var Sockets={BUFFER_SIZE:10240,MAX_BUFFER_SIZE:10485760,nextFd:1,fds:{},nextport:1,maxport:65535,peer:null,connections:{},portmap:{},localAddr:4261412874,addrPool:[33554442,50331658,67108874,83886090,100663306,117440522,134217738,150994954,167772170,184549386,201326602,218103818,234881034]};function _emscripten_run_script(ptr){ptr>>>=0;eval(UTF8ToString(ptr))}_emscripten_run_script.sig="vp";function _emscripten_run_script_int(ptr){ptr>>>=0;return eval(UTF8ToString(ptr))|0}_emscripten_run_script_int.sig="ip";function _emscripten_run_script_string(ptr){ptr>>>=0;var s=eval(UTF8ToString(ptr));if(s==null){return 0}s+="";var me=_emscripten_run_script_string;var len=lengthBytesUTF8(s);if(!me.bufferSize||me.bufferSize<len+1){if(me.bufferSize)_free(me.buffer);me.bufferSize=len+1;me.buffer=_malloc(me.bufferSize)}stringToUTF8(s,me.buffer,me.bufferSize);return me.buffer}_emscripten_run_script_string.sig="pp";var _emscripten_random=()=>Math.random();_emscripten_random.sig="f";var _emscripten_performance_now=()=>performance.now();_emscripten_performance_now.sig="d";var __emscripten_get_now_is_monotonic=()=>nowIsMonotonic;__emscripten_get_now_is_monotonic.sig="i";var warnOnce=text=>{warnOnce.shown||={};if(!warnOnce.shown[text]){warnOnce.shown[text]=1;if(ENVIRONMENT_IS_NODE)text="warning: "+text;err(text)}};var jsStackTrace=()=>(new Error).stack.toString();var getCallstack=flags=>{var callstack=jsStackTrace();var lines=callstack.split("\n");callstack="";var firefoxRe=new RegExp("\\s*(.*?)@(.*?):([0-9]+):([0-9]+)");var chromeRe=new RegExp("\\s*at (.*?) \\((.*):(.*):(.*)\\)");for(var line of lines){var symbolName="";var file="";var lineno=0;var column=0;var parts=chromeRe.exec(line);if(parts?.length==5){symbolName=parts[1];file=parts[2];lineno=parts[3];column=parts[4]}else{parts=firefoxRe.exec(line);if(parts?.length>=4){symbolName=parts[1];file=parts[2];lineno=parts[3];column=parts[4]|0}else{callstack+=line+"\n";continue}}if(symbolName=="_emscripten_log"||symbolName=="_emscripten_get_callstack"){callstack="";continue}if(flags&24){if(flags&64){file=file.substring(file.replace(/\\/g,"/").lastIndexOf("/")+1)}callstack+=`    at ${symbolName} (${file}:${lineno}:${column})\n`}}callstack=callstack.replace(/\s+$/,"");return callstack};var emscriptenLog=(flags,str)=>{if(flags&24){str=str.replace(/\s+$/,"");str+=(str.length>0?"\n":"")+getCallstack(flags)}if(flags&1){if(flags&4){console.error(str)}else if(flags&2){console.warn(str)}else if(flags&512){console.info(str)}else if(flags&256){console.debug(str)}else{console.log(str)}}else if(flags&6){err(str)}else{out(str)}};var reallyNegative=x=>x<0||x===0&&1/x===-Infinity;var reSign=(value,bits)=>{if(value<=0){return value}var half=bits<=32?Math.abs(1<<bits-1):Math.pow(2,bits-1);if(value>=half&&(bits<=32||value>half)){value=-2*half+value}return value};var unSign=(value,bits)=>{if(value>=0){return value}return bits<=32?2*Math.abs(1<<bits-1)+value:Math.pow(2,bits)+value};var strLen=ptr=>{var end=ptr;while(HEAPU8[end>>>0])++end;return end-ptr};var formatString=(format,varargs)=>{var textIndex=format;var argIndex=varargs;function prepVararg(ptr,type){if(type==="double"||type==="i64"){if(ptr&7){ptr+=4}}else{}return ptr}function getNextArg(type){var ret;argIndex=prepVararg(argIndex,type);if(type==="double"){ret=HEAPF64[argIndex>>>3>>>0];argIndex+=8}else if(type=="i64"){ret=[HEAP32[argIndex>>>2>>>0],HEAP32[argIndex+4>>>2>>>0]];argIndex+=8}else{type="i32";ret=HEAP32[argIndex>>>2>>>0];argIndex+=4}return ret}var ret=[];var curr,next,currArg;while(1){var startTextIndex=textIndex;curr=HEAP8[textIndex>>>0];if(curr===0)break;next=HEAP8[textIndex+1>>>0];if(curr==37){var flagAlwaysSigned=false;var flagLeftAlign=false;var flagAlternative=false;var flagZeroPad=false;var flagPadSign=false;flagsLoop:while(1){switch(next){case 43:flagAlwaysSigned=true;break;case 45:flagLeftAlign=true;break;case 35:flagAlternative=true;break;case 48:if(flagZeroPad){break flagsLoop}else{flagZeroPad=true;break}case 32:flagPadSign=true;break;default:break flagsLoop}textIndex++;next=HEAP8[textIndex+1>>>0]}var width=0;if(next==42){width=getNextArg("i32");textIndex++;next=HEAP8[textIndex+1>>>0]}else{while(next>=48&&next<=57){width=width*10+(next-48);textIndex++;next=HEAP8[textIndex+1>>>0]}}var precisionSet=false,precision=-1;if(next==46){precision=0;precisionSet=true;textIndex++;next=HEAP8[textIndex+1>>>0];if(next==42){precision=getNextArg("i32");textIndex++}else{while(1){var precisionChr=HEAP8[textIndex+1>>>0];if(precisionChr<48||precisionChr>57)break;precision=precision*10+(precisionChr-48);textIndex++}}next=HEAP8[textIndex+1>>>0]}if(precision<0){precision=6;precisionSet=false}var argSize;switch(String.fromCharCode(next)){case"h":var nextNext=HEAP8[textIndex+2>>>0];if(nextNext==104){textIndex++;argSize=1}else{argSize=2}break;case"l":var nextNext=HEAP8[textIndex+2>>>0];if(nextNext==108){textIndex++;argSize=8}else{argSize=4}break;case"L":case"q":case"j":argSize=8;break;case"z":case"t":case"I":argSize=4;break;default:argSize=null}if(argSize)textIndex++;next=HEAP8[textIndex+1>>>0];switch(String.fromCharCode(next)){case"d":case"i":case"u":case"o":case"x":case"X":case"p":{var signed=next==100||next==105;argSize=argSize||4;currArg=getNextArg("i"+argSize*8);var argText;if(argSize==8){currArg=next==117?convertU32PairToI53(currArg[0],currArg[1]):convertI32PairToI53(currArg[0],currArg[1])}if(argSize<=4){var limit=Math.pow(256,argSize)-1;currArg=(signed?reSign:unSign)(currArg&limit,argSize*8)}var currAbsArg=Math.abs(currArg);var prefix="";if(next==100||next==105){argText=reSign(currArg,8*argSize).toString(10)}else if(next==117){argText=unSign(currArg,8*argSize).toString(10);currArg=Math.abs(currArg)}else if(next==111){argText=(flagAlternative?"0":"")+currAbsArg.toString(8)}else if(next==120||next==88){prefix=flagAlternative&&currArg!=0?"0x":"";if(currArg<0){currArg=-currArg;argText=(currAbsArg-1).toString(16);var buffer=[];for(var i=0;i<argText.length;i++){buffer.push((15-parseInt(argText[i],16)).toString(16))}argText=buffer.join("");while(argText.length<argSize*2)argText="f"+argText}else{argText=currAbsArg.toString(16)}if(next==88){prefix=prefix.toUpperCase();argText=argText.toUpperCase()}}else if(next==112){if(currAbsArg===0){argText="(nil)"}else{prefix="0x";argText=currAbsArg.toString(16)}}if(precisionSet){while(argText.length<precision){argText="0"+argText}}if(currArg>=0){if(flagAlwaysSigned){prefix="+"+prefix}else if(flagPadSign){prefix=" "+prefix}}if(argText.charAt(0)=="-"){prefix="-"+prefix;argText=argText.slice(1)}while(prefix.length+argText.length<width){if(flagLeftAlign){argText+=" "}else{if(flagZeroPad){argText="0"+argText}else{prefix=" "+prefix}}}argText=prefix+argText;argText.split("").forEach(chr=>ret.push(chr.charCodeAt(0)));break}case"f":case"F":case"e":case"E":case"g":case"G":{currArg=getNextArg("double");var argText;if(isNaN(currArg)){argText="nan";flagZeroPad=false}else if(!isFinite(currArg)){argText=(currArg<0?"-":"")+"inf";flagZeroPad=false}else{var isGeneral=false;var effectivePrecision=Math.min(precision,20);if(next==103||next==71){isGeneral=true;precision=precision||1;var exponent=parseInt(currArg.toExponential(effectivePrecision).split("e")[1],10);if(precision>exponent&&exponent>=-4){next=(next==103?"f":"F").charCodeAt(0);precision-=exponent+1}else{next=(next==103?"e":"E").charCodeAt(0);precision--}effectivePrecision=Math.min(precision,20)}if(next==101||next==69){argText=currArg.toExponential(effectivePrecision);if(/[eE][-+]\d$/.test(argText)){argText=argText.slice(0,-1)+"0"+argText.slice(-1)}}else if(next==102||next==70){argText=currArg.toFixed(effectivePrecision);if(currArg===0&&reallyNegative(currArg)){argText="-"+argText}}var parts=argText.split("e");if(isGeneral&&!flagAlternative){while(parts[0].length>1&&parts[0].includes(".")&&(parts[0].slice(-1)=="0"||parts[0].slice(-1)==".")){parts[0]=parts[0].slice(0,-1)}}else{if(flagAlternative&&argText.indexOf(".")==-1)parts[0]+=".";while(precision>effectivePrecision++)parts[0]+="0"}argText=parts[0]+(parts.length>1?"e"+parts[1]:"");if(next==69)argText=argText.toUpperCase();if(currArg>=0){if(flagAlwaysSigned){argText="+"+argText}else if(flagPadSign){argText=" "+argText}}}while(argText.length<width){if(flagLeftAlign){argText+=" "}else{if(flagZeroPad&&(argText[0]=="-"||argText[0]=="+")){argText=argText[0]+"0"+argText.slice(1)}else{argText=(flagZeroPad?"0":" ")+argText}}}if(next<97)argText=argText.toUpperCase();argText.split("").forEach(chr=>ret.push(chr.charCodeAt(0)));break}case"s":{var arg=getNextArg("i8*");var argLength=arg?strLen(arg):"(null)".length;if(precisionSet)argLength=Math.min(argLength,precision);if(!flagLeftAlign){while(argLength<width--){ret.push(32)}}if(arg){for(var i=0;i<argLength;i++){ret.push(HEAPU8[arg++>>>0])}}else{ret=ret.concat(intArrayFromString("(null)".slice(0,argLength),true))}if(flagLeftAlign){while(argLength<width--){ret.push(32)}}break}case"c":{if(flagLeftAlign)ret.push(getNextArg("i8"));while(--width>0){ret.push(32)}if(!flagLeftAlign)ret.push(getNextArg("i8"));break}case"n":{var ptr=getNextArg("i32*");HEAP32[ptr>>>2>>>0]=ret.length;break}case"%":{ret.push(curr);break}default:{for(var i=startTextIndex;i<textIndex+2;i++){ret.push(HEAP8[i>>>0])}}}textIndex+=2}else{ret.push(curr);textIndex+=1}}return ret};function _emscripten_log(flags,format,varargs){format>>>=0;varargs>>>=0;var result=formatString(format,varargs);var str=UTF8ArrayToString(result);emscriptenLog(flags,str)}_emscripten_log.sig="vipp";function _emscripten_get_compiler_setting(name){name>>>=0;throw"You must build with -sRETAIN_COMPILER_SETTINGS for getCompilerSetting or emscripten_get_compiler_setting to work"}_emscripten_get_compiler_setting.sig="pp";var _emscripten_has_asyncify=()=>0;_emscripten_has_asyncify.sig="i";var _emscripten_debugger=()=>{debugger};_emscripten_debugger.sig="v";function _emscripten_print_double(x,to,max){to>>>=0;var str=x+"";if(to)return stringToUTF8(str,to,max);else return lengthBytesUTF8(str)}_emscripten_print_double.sig="idpi";function _emscripten_asm_const_double(code,sigPtr,argbuf){code>>>=0;sigPtr>>>=0;argbuf>>>=0;return runEmAsmFunction(code,sigPtr,argbuf)}_emscripten_asm_const_double.sig="dppp";function _emscripten_asm_const_ptr(code,sigPtr,argbuf){code>>>=0;sigPtr>>>=0;argbuf>>>=0;return runEmAsmFunction(code,sigPtr,argbuf)}_emscripten_asm_const_ptr.sig="pppp";var runMainThreadEmAsm=(emAsmAddr,sigPtr,argbuf,sync)=>{var args=readEmAsmArgs(sigPtr,argbuf);return ASM_CONSTS[emAsmAddr](...args)};function _emscripten_asm_const_int_sync_on_main_thread(emAsmAddr,sigPtr,argbuf){emAsmAddr>>>=0;sigPtr>>>=0;argbuf>>>=0;return runMainThreadEmAsm(emAsmAddr,sigPtr,argbuf,1)}_emscripten_asm_const_int_sync_on_main_thread.sig="ippp";function _emscripten_asm_const_ptr_sync_on_main_thread(emAsmAddr,sigPtr,argbuf){emAsmAddr>>>=0;sigPtr>>>=0;argbuf>>>=0;return runMainThreadEmAsm(emAsmAddr,sigPtr,argbuf,1)}_emscripten_asm_const_ptr_sync_on_main_thread.sig="pppp";var _emscripten_asm_const_double_sync_on_main_thread=_emscripten_asm_const_int_sync_on_main_thread;_emscripten_asm_const_double_sync_on_main_thread.sig="dppp";function _emscripten_asm_const_async_on_main_thread(emAsmAddr,sigPtr,argbuf){emAsmAddr>>>=0;sigPtr>>>=0;argbuf>>>=0;return runMainThreadEmAsm(emAsmAddr,sigPtr,argbuf,0)}_emscripten_asm_const_async_on_main_thread.sig="vppp";function __Unwind_Backtrace(func,arg){func>>>=0;arg>>>=0;var trace=getCallstack();var parts=trace.split("\n");for(var i=0;i<parts.length;i++){var ret=getWasmTableEntry(func)(0,arg);if(ret!==0)return}}__Unwind_Backtrace.sig="ipp";function __Unwind_GetIPInfo(context,ipBefore){context>>>=0;ipBefore>>>=0;return abort("Unwind_GetIPInfo")}__Unwind_GetIPInfo.sig="ppp";function __Unwind_FindEnclosingFunction(ip){ip>>>=0;return 0}__Unwind_FindEnclosingFunction.sig="pp";var listenOnce=(object,event,func)=>object.addEventListener(event,func,{once:true});var autoResumeAudioContext=(ctx,elements)=>{if(!elements){elements=[document,document.getElementById("canvas")]}["keydown","mousedown","touchstart"].forEach(event=>{elements.forEach(element=>{if(element){listenOnce(element,event,()=>{if(ctx.state==="suspended")ctx.resume()})}})})};var dynCall=(sig,ptr,args=[],promising=false)=>{var func=getWasmTableEntry(ptr);var rtn=func(...args);function convert(rtn){return sig[0]=="p"?rtn>>>0:rtn}return convert(rtn)};var getDynCaller=(sig,ptr,promising=false)=>(...args)=>dynCall(sig,ptr,args,promising);var _emscripten_exit_with_live_runtime=()=>{runtimeKeepalivePush();throw"unwind"};_emscripten_exit_with_live_runtime.sig="v";var _emscripten_force_exit=status=>{__emscripten_runtime_keepalive_clear();_exit(status)};_emscripten_force_exit.sig="vi";function _emscripten_outn(str,len){str>>>=0;len>>>=0;return out(UTF8ToString(str,len))}_emscripten_outn.sig="vpp";function _emscripten_errn(str,len){str>>>=0;len>>>=0;return err(UTF8ToString(str,len))}_emscripten_errn.sig="vpp";var _emscripten_throw_number=number=>{throw number};_emscripten_throw_number.sig="vd";function _emscripten_throw_string(str){str>>>=0;throw UTF8ToString(str)}_emscripten_throw_string.sig="vp";var _emscripten_runtime_keepalive_check=keepRuntimeAlive;_emscripten_runtime_keepalive_check.sig="i";var asmjsMangle=x=>{if(x=="__main_argc_argv"){x="main"}return x.startsWith("dynCall_")?x:"_"+x};var ___global_base=1024;function __emscripten_fs_load_embedded_files(ptr){ptr>>>=0;do{var name_addr=HEAPU32[ptr>>>2>>>0];ptr+=4;var len=HEAPU32[ptr>>>2>>>0];ptr+=4;var content=HEAPU32[ptr>>>2>>>0];ptr+=4;var name=UTF8ToString(name_addr);FS.createPath("/",PATH.dirname(name),true,true);FS.createDataFile(name,null,HEAP8.subarray(content>>>0,content+len>>>0),true,true,true)}while(HEAPU32[ptr>>>2>>>0])}__emscripten_fs_load_embedded_files.sig="vp";var POINTER_SIZE=4;function getNativeTypeSize(type){switch(type){case"i1":case"i8":case"u8":return 1;case"i16":case"u16":return 2;case"i32":case"u32":return 4;case"i64":case"u64":return 8;case"float":return 4;case"double":return 8;default:{if(type.endsWith("*")){return POINTER_SIZE}if(type[0]==="i"){const bits=Number(type.slice(1));assert(bits%8===0,`getNativeTypeSize invalid bits ${bits}, ${type} type`);return bits/8}return 0}}}var onInits=[];var addOnInit=cb=>onInits.push(cb);var onMains=[];var addOnPreMain=cb=>onMains.push(cb);var onExits=[];var addOnExit=cb=>onExits.push(cb);var STACK_SIZE=5242880;var STACK_ALIGN=16;var ASSERTIONS=0;var getCFunc=ident=>{var func=Module["_"+ident];return func};var writeArrayToMemory=(array,buffer)=>{HEAP8.set(array,buffer>>>0)};var ccall=(ident,returnType,argTypes,args,opts)=>{var toC={string:str=>{var ret=0;if(str!==null&&str!==undefined&&str!==0){ret=stringToUTF8OnStack(str)}return ret},array:arr=>{var ret=stackAlloc(arr.length);writeArrayToMemory(arr,ret);return ret}};function convertReturnValue(ret){if(returnType==="string"){return UTF8ToString(ret)}if(returnType==="boolean")return Boolean(ret);return ret}var func=getCFunc(ident);var cArgs=[];var stack=0;if(args){for(var i=0;i<args.length;i++){var converter=toC[argTypes[i]];if(converter){if(stack===0)stack=stackSave();cArgs[i]=converter(args[i])}else{cArgs[i]=args[i]}}}var ret=func(...cArgs);function onDone(ret){if(stack!==0)stackRestore(stack);return convertReturnValue(ret)}ret=onDone(ret);return ret};var cwrap=(ident,returnType,argTypes,opts)=>{var numericArgs=!argTypes||argTypes.every(type=>type==="number"||type==="boolean");var numericRet=returnType!=="string";if(numericRet&&numericArgs&&!opts){return getCFunc(ident)}return(...args)=>ccall(ident,returnType,argTypes,args,opts)};var removeFunction=index=>{functionsInTableMap.delete(getWasmTableEntry(index));setWasmTableEntry(index,null);freeTableIndexes.push(index)};var _emscripten_math_cbrt=Math.cbrt;_emscripten_math_cbrt.sig="dd";var _emscripten_math_pow=Math.pow;_emscripten_math_pow.sig="ddd";var _emscripten_math_random=Math.random;_emscripten_math_random.sig="d";var _emscripten_math_sign=Math.sign;_emscripten_math_sign.sig="dd";var _emscripten_math_sqrt=Math.sqrt;_emscripten_math_sqrt.sig="dd";var _emscripten_math_exp=Math.exp;_emscripten_math_exp.sig="dd";var _emscripten_math_expm1=Math.expm1;_emscripten_math_expm1.sig="dd";var _emscripten_math_fmod=(x,y)=>x%y;_emscripten_math_fmod.sig="ddd";var _emscripten_math_log=Math.log;_emscripten_math_log.sig="dd";var _emscripten_math_log1p=Math.log1p;_emscripten_math_log1p.sig="dd";var _emscripten_math_log10=Math.log10;_emscripten_math_log10.sig="dd";var _emscripten_math_log2=Math.log2;_emscripten_math_log2.sig="dd";var _emscripten_math_round=Math.round;_emscripten_math_round.sig="dd";var _emscripten_math_acos=Math.acos;_emscripten_math_acos.sig="dd";var _emscripten_math_acosh=Math.acosh;_emscripten_math_acosh.sig="dd";var _emscripten_math_asin=Math.asin;_emscripten_math_asin.sig="dd";var _emscripten_math_asinh=Math.asinh;_emscripten_math_asinh.sig="dd";var _emscripten_math_atan=Math.atan;_emscripten_math_atan.sig="dd";var _emscripten_math_atanh=Math.atanh;_emscripten_math_atanh.sig="dd";var _emscripten_math_atan2=Math.atan2;_emscripten_math_atan2.sig="ddd";var _emscripten_math_cos=Math.cos;_emscripten_math_cos.sig="dd";var _emscripten_math_cosh=Math.cosh;_emscripten_math_cosh.sig="dd";function _emscripten_math_hypot(count,varargs){varargs>>>=0;var args=[];for(var i=0;i<count;++i){args.push(HEAPF64[varargs+i*8>>>3>>>0])}return Math.hypot(...args)}_emscripten_math_hypot.sig="dip";var _emscripten_math_sin=Math.sin;_emscripten_math_sin.sig="dd";var _emscripten_math_sinh=Math.sinh;_emscripten_math_sinh.sig="dd";var _emscripten_math_tan=Math.tan;_emscripten_math_tan.sig="dd";var _emscripten_math_tanh=Math.tanh;_emscripten_math_tanh.sig="dd";var intArrayToString=array=>{var ret=[];for(var i=0;i<array.length;i++){var chr=array[i];if(chr>255){chr&=255}ret.push(String.fromCharCode(chr))}return ret.join("")};var AsciiToString=ptr=>{ptr>>>=0;var str="";while(1){var ch=HEAPU8[ptr++>>>0];if(!ch)return str;str+=String.fromCharCode(ch)}};var UTF16Decoder=typeof TextDecoder!="undefined"?new TextDecoder("utf-16le"):undefined;var UTF16ToString=(ptr,maxBytesToRead)=>{var idx=ptr>>>1;var maxIdx=idx+maxBytesToRead/2;var endIdx=idx;while(!(endIdx>=maxIdx)&&HEAPU16[endIdx>>>0])++endIdx;if(endIdx-idx>16&&UTF16Decoder)return UTF16Decoder.decode(HEAPU16.subarray(idx>>>0,endIdx>>>0));var str="";for(var i=idx;!(i>=maxIdx);++i){var codeUnit=HEAPU16[i>>>0];if(codeUnit==0)break;str+=String.fromCharCode(codeUnit)}return str};var stringToUTF16=(str,outPtr,maxBytesToWrite)=>{maxBytesToWrite??=2147483647;if(maxBytesToWrite<2)return 0;maxBytesToWrite-=2;var startPtr=outPtr;var numCharsToWrite=maxBytesToWrite<str.length*2?maxBytesToWrite/2:str.length;for(var i=0;i<numCharsToWrite;++i){var codeUnit=str.charCodeAt(i);HEAP16[outPtr>>>1>>>0]=codeUnit;outPtr+=2}HEAP16[outPtr>>>1>>>0]=0;return outPtr-startPtr};var lengthBytesUTF16=str=>str.length*2;var UTF32ToString=(ptr,maxBytesToRead)=>{var i=0;var str="";while(!(i>=maxBytesToRead/4)){var utf32=HEAP32[ptr+i*4>>>2>>>0];if(utf32==0)break;++i;if(utf32>=65536){var ch=utf32-65536;str+=String.fromCharCode(55296|ch>>10,56320|ch&1023)}else{str+=String.fromCharCode(utf32)}}return str};var stringToUTF32=(str,outPtr,maxBytesToWrite)=>{outPtr>>>=0;maxBytesToWrite??=2147483647;if(maxBytesToWrite<4)return 0;var startPtr=outPtr;var endPtr=startPtr+maxBytesToWrite-4;for(var i=0;i<str.length;++i){var codeUnit=str.charCodeAt(i);if(codeUnit>=55296&&codeUnit<=57343){var trailSurrogate=str.charCodeAt(++i);codeUnit=65536+((codeUnit&1023)<<10)|trailSurrogate&1023}HEAP32[outPtr>>>2>>>0]=codeUnit;outPtr+=4;if(outPtr+4>endPtr)break}HEAP32[outPtr>>>2>>>0]=0;return outPtr-startPtr};var lengthBytesUTF32=str=>{var len=0;for(var i=0;i<str.length;++i){var codeUnit=str.charCodeAt(i);if(codeUnit>=55296&&codeUnit<=57343)++i;len+=4}return len};var JSEvents={memcpy(target,src,size){HEAP8.set(HEAP8.subarray(src>>>0,src+size>>>0),target>>>0)},removeAllEventListeners(){while(JSEvents.eventHandlers.length){JSEvents._removeHandler(JSEvents.eventHandlers.length-1)}JSEvents.deferredCalls=[]},registerRemoveEventListeners(){if(!JSEvents.removeEventListenersRegistered){addOnExit(JSEvents.removeAllEventListeners);JSEvents.removeEventListenersRegistered=true}},inEventHandler:0,deferredCalls:[],deferCall(targetFunction,precedence,argsList){function arraysHaveEqualContent(arrA,arrB){if(arrA.length!=arrB.length)return false;for(var i in arrA){if(arrA[i]!=arrB[i])return false}return true}for(var call of JSEvents.deferredCalls){if(call.targetFunction==targetFunction&&arraysHaveEqualContent(call.argsList,argsList)){return}}JSEvents.deferredCalls.push({targetFunction,precedence,argsList});JSEvents.deferredCalls.sort((x,y)=>x.precedence<y.precedence)},removeDeferredCalls(targetFunction){JSEvents.deferredCalls=JSEvents.deferredCalls.filter(call=>call.targetFunction!=targetFunction)},canPerformEventHandlerRequests(){if(navigator.userActivation){return navigator.userActivation.isActive}return JSEvents.inEventHandler&&JSEvents.currentEventHandler.allowsDeferredCalls},runDeferredCalls(){if(!JSEvents.canPerformEventHandlerRequests()){return}var deferredCalls=JSEvents.deferredCalls;JSEvents.deferredCalls=[];for(var call of deferredCalls){call.targetFunction(...call.argsList)}},eventHandlers:[],removeAllHandlersOnTarget:(target,eventTypeString)=>{for(var i=0;i<JSEvents.eventHandlers.length;++i){if(JSEvents.eventHandlers[i].target==target&&(!eventTypeString||eventTypeString==JSEvents.eventHandlers[i].eventTypeString)){JSEvents._removeHandler(i--)}}},_removeHandler(i){var h=JSEvents.eventHandlers[i];h.target.removeEventListener(h.eventTypeString,h.eventListenerFunc,h.useCapture);JSEvents.eventHandlers.splice(i,1)},registerOrRemoveHandler(eventHandler){if(!eventHandler.target){return-4}if(eventHandler.callbackfunc){eventHandler.eventListenerFunc=function(event){++JSEvents.inEventHandler;JSEvents.currentEventHandler=eventHandler;JSEvents.runDeferredCalls();eventHandler.handlerFunc(event);JSEvents.runDeferredCalls();--JSEvents.inEventHandler};eventHandler.target.addEventListener(eventHandler.eventTypeString,eventHandler.eventListenerFunc,eventHandler.useCapture);JSEvents.eventHandlers.push(eventHandler);JSEvents.registerRemoveEventListeners()}else{for(var i=0;i<JSEvents.eventHandlers.length;++i){if(JSEvents.eventHandlers[i].target==eventHandler.target&&JSEvents.eventHandlers[i].eventTypeString==eventHandler.eventTypeString){JSEvents._removeHandler(i--)}}}return 0},getNodeNameForTarget(target){if(!target)return"";if(target==window)return"#window";if(target==screen)return"#screen";return target?.nodeName||""},fullscreenEnabled(){return document.fullscreenEnabled||document.webkitFullscreenEnabled}};var maybeCStringToJsString=cString=>cString>2?UTF8ToString(cString):cString;var specialHTMLTargets=[0,typeof document!="undefined"?document:0,typeof window!="undefined"?window:0];var findEventTarget=target=>{target=maybeCStringToJsString(target);var domElement=specialHTMLTargets[target]||(typeof document!="undefined"?document.querySelector(target):null);return domElement};var registerKeyEventCallback=(target,userData,useCapture,callbackfunc,eventTypeId,eventTypeString,targetThread)=>{JSEvents.keyEvent||=_malloc(160);var keyEventHandlerFunc=e=>{var keyEventData=JSEvents.keyEvent;HEAPF64[keyEventData>>>3>>>0]=e.timeStamp;var idx=keyEventData>>>2;HEAP32[idx+2>>>0]=e.location;HEAP8[keyEventData+12>>>0]=e.ctrlKey;HEAP8[keyEventData+13>>>0]=e.shiftKey;HEAP8[keyEventData+14>>>0]=e.altKey;HEAP8[keyEventData+15>>>0]=e.metaKey;HEAP8[keyEventData+16>>>0]=e.repeat;HEAP32[idx+5>>>0]=e.charCode;HEAP32[idx+6>>>0]=e.keyCode;HEAP32[idx+7>>>0]=e.which;stringToUTF8(e.key||"",keyEventData+32,32);stringToUTF8(e.code||"",keyEventData+64,32);stringToUTF8(e.char||"",keyEventData+96,32);stringToUTF8(e.locale||"",keyEventData+128,32);if(getWasmTableEntry(callbackfunc)(eventTypeId,keyEventData,userData))e.preventDefault()};var eventHandler={target:findEventTarget(target),eventTypeString,callbackfunc,handlerFunc:keyEventHandlerFunc,useCapture};return JSEvents.registerOrRemoveHandler(eventHandler)};var findCanvasEventTarget=findEventTarget;function _emscripten_set_keypress_callback_on_thread(target,userData,useCapture,callbackfunc,targetThread){target>>>=0;userData>>>=0;callbackfunc>>>=0;targetThread>>>=0;return registerKeyEventCallback(target,userData,useCapture,callbackfunc,1,"keypress",targetThread)}_emscripten_set_keypress_callback_on_thread.sig="ippipp";function _emscripten_set_keydown_callback_on_thread(target,userData,useCapture,callbackfunc,targetThread){target>>>=0;userData>>>=0;callbackfunc>>>=0;targetThread>>>=0;return registerKeyEventCallback(target,userData,useCapture,callbackfunc,2,"keydown",targetThread)}_emscripten_set_keydown_callback_on_thread.sig="ippipp";function _emscripten_set_keyup_callback_on_thread(target,userData,useCapture,callbackfunc,targetThread){target>>>=0;userData>>>=0;callbackfunc>>>=0;targetThread>>>=0;return registerKeyEventCallback(target,userData,useCapture,callbackfunc,3,"keyup",targetThread)}_emscripten_set_keyup_callback_on_thread.sig="ippipp";var getBoundingClientRect=e=>specialHTMLTargets.indexOf(e)<0?e.getBoundingClientRect():{left:0,top:0};var fillMouseEventData=(eventStruct,e,target)=>{HEAPF64[eventStruct>>>3>>>0]=e.timeStamp;var idx=eventStruct>>>2;HEAP32[idx+2>>>0]=e.screenX;HEAP32[idx+3>>>0]=e.screenY;HEAP32[idx+4>>>0]=e.clientX;HEAP32[idx+5>>>0]=e.clientY;HEAP8[eventStruct+24>>>0]=e.ctrlKey;HEAP8[eventStruct+25>>>0]=e.shiftKey;HEAP8[eventStruct+26>>>0]=e.altKey;HEAP8[eventStruct+27>>>0]=e.metaKey;HEAP16[idx*2+14>>>0]=e.button;HEAP16[idx*2+15>>>0]=e.buttons;HEAP32[idx+8>>>0]=e["movementX"];HEAP32[idx+9>>>0]=e["movementY"];var rect=getBoundingClientRect(target);HEAP32[idx+10>>>0]=e.clientX-(rect.left|0);HEAP32[idx+11>>>0]=e.clientY-(rect.top|0)};var registerMouseEventCallback=(target,userData,useCapture,callbackfunc,eventTypeId,eventTypeString,targetThread)=>{JSEvents.mouseEvent||=_malloc(64);target=findEventTarget(target);var mouseEventHandlerFunc=(e=event)=>{fillMouseEventData(JSEvents.mouseEvent,e,target);if(getWasmTableEntry(callbackfunc)(eventTypeId,JSEvents.mouseEvent,userData))e.preventDefault()};var eventHandler={target,allowsDeferredCalls:eventTypeString!="mousemove"&&eventTypeString!="mouseenter"&&eventTypeString!="mouseleave",eventTypeString,callbackfunc,handlerFunc:mouseEventHandlerFunc,useCapture};return JSEvents.registerOrRemoveHandler(eventHandler)};function _emscripten_set_click_callback_on_thread(target,userData,useCapture,callbackfunc,targetThread){target>>>=0;userData>>>=0;callbackfunc>>>=0;targetThread>>>=0;return registerMouseEventCallback(target,userData,useCapture,callbackfunc,4,"click",targetThread)}_emscripten_set_click_callback_on_thread.sig="ippipp";function _emscripten_set_mousedown_callback_on_thread(target,userData,useCapture,callbackfunc,targetThread){target>>>=0;userData>>>=0;callbackfunc>>>=0;targetThread>>>=0;return registerMouseEventCallback(target,userData,useCapture,callbackfunc,5,"mousedown",targetThread)}_emscripten_set_mousedown_callback_on_thread.sig="ippipp";function _emscripten_set_mouseup_callback_on_thread(target,userData,useCapture,callbackfunc,targetThread){target>>>=0;userData>>>=0;callbackfunc>>>=0;targetThread>>>=0;return registerMouseEventCallback(target,userData,useCapture,callbackfunc,6,"mouseup",targetThread)}_emscripten_set_mouseup_callback_on_thread.sig="ippipp";function _emscripten_set_dblclick_callback_on_thread(target,userData,useCapture,callbackfunc,targetThread){target>>>=0;userData>>>=0;callbackfunc>>>=0;targetThread>>>=0;return registerMouseEventCallback(target,userData,useCapture,callbackfunc,7,"dblclick",targetThread)}_emscripten_set_dblclick_callback_on_thread.sig="ippipp";function _emscripten_set_mousemove_callback_on_thread(target,userData,useCapture,callbackfunc,targetThread){target>>>=0;userData>>>=0;callbackfunc>>>=0;targetThread>>>=0;return registerMouseEventCallback(target,userData,useCapture,callbackfunc,8,"mousemove",targetThread)}_emscripten_set_mousemove_callback_on_thread.sig="ippipp";function _emscripten_set_mouseenter_callback_on_thread(target,userData,useCapture,callbackfunc,targetThread){target>>>=0;userData>>>=0;callbackfunc>>>=0;targetThread>>>=0;return registerMouseEventCallback(target,userData,useCapture,callbackfunc,33,"mouseenter",targetThread)}_emscripten_set_mouseenter_callback_on_thread.sig="ippipp";function _emscripten_set_mouseleave_callback_on_thread(target,userData,useCapture,callbackfunc,targetThread){target>>>=0;userData>>>=0;callbackfunc>>>=0;targetThread>>>=0;return registerMouseEventCallback(target,userData,useCapture,callbackfunc,34,"mouseleave",targetThread)}_emscripten_set_mouseleave_callback_on_thread.sig="ippipp";function _emscripten_set_mouseover_callback_on_thread(target,userData,useCapture,callbackfunc,targetThread){target>>>=0;userData>>>=0;callbackfunc>>>=0;targetThread>>>=0;return registerMouseEventCallback(target,userData,useCapture,callbackfunc,35,"mouseover",targetThread)}_emscripten_set_mouseover_callback_on_thread.sig="ippipp";function _emscripten_set_mouseout_callback_on_thread(target,userData,useCapture,callbackfunc,targetThread){target>>>=0;userData>>>=0;callbackfunc>>>=0;targetThread>>>=0;return registerMouseEventCallback(target,userData,useCapture,callbackfunc,36,"mouseout",targetThread)}_emscripten_set_mouseout_callback_on_thread.sig="ippipp";function _emscripten_get_mouse_status(mouseState){mouseState>>>=0;if(!JSEvents.mouseEvent)return-7;JSEvents.memcpy(mouseState,JSEvents.mouseEvent,64);return 0}_emscripten_get_mouse_status.sig="ip";var registerWheelEventCallback=(target,userData,useCapture,callbackfunc,eventTypeId,eventTypeString,targetThread)=>{JSEvents.wheelEvent||=_malloc(96);var wheelHandlerFunc=(e=event)=>{var wheelEvent=JSEvents.wheelEvent;fillMouseEventData(wheelEvent,e,target);HEAPF64[wheelEvent+64>>>3>>>0]=e["deltaX"];HEAPF64[wheelEvent+72>>>3>>>0]=e["deltaY"];HEAPF64[wheelEvent+80>>>3>>>0]=e["deltaZ"];HEAP32[wheelEvent+88>>>2>>>0]=e["deltaMode"];if(getWasmTableEntry(callbackfunc)(eventTypeId,wheelEvent,userData))e.preventDefault()};var eventHandler={target,allowsDeferredCalls:true,eventTypeString,callbackfunc,handlerFunc:wheelHandlerFunc,useCapture};return JSEvents.registerOrRemoveHandler(eventHandler)};function _emscripten_set_wheel_callback_on_thread(target,userData,useCapture,callbackfunc,targetThread){target>>>=0;userData>>>=0;callbackfunc>>>=0;targetThread>>>=0;target=findEventTarget(target);if(!target)return-4;if(typeof target.onwheel!="undefined"){return registerWheelEventCallback(target,userData,useCapture,callbackfunc,9,"wheel",targetThread)}else{return-1}}_emscripten_set_wheel_callback_on_thread.sig="ippipp";var registerUiEventCallback=(target,userData,useCapture,callbackfunc,eventTypeId,eventTypeString,targetThread)=>{JSEvents.uiEvent||=_malloc(36);target=findEventTarget(target);var uiEventHandlerFunc=(e=event)=>{if(e.target!=target){return}var b=document.body;if(!b){return}var uiEvent=JSEvents.uiEvent;HEAP32[uiEvent>>>2>>>0]=0;HEAP32[uiEvent+4>>>2>>>0]=b.clientWidth;HEAP32[uiEvent+8>>>2>>>0]=b.clientHeight;HEAP32[uiEvent+12>>>2>>>0]=innerWidth;HEAP32[uiEvent+16>>>2>>>0]=innerHeight;HEAP32[uiEvent+20>>>2>>>0]=outerWidth;HEAP32[uiEvent+24>>>2>>>0]=outerHeight;HEAP32[uiEvent+28>>>2>>>0]=pageXOffset|0;HEAP32[uiEvent+32>>>2>>>0]=pageYOffset|0;if(getWasmTableEntry(callbackfunc)(eventTypeId,uiEvent,userData))e.preventDefault()};var eventHandler={target,eventTypeString,callbackfunc,handlerFunc:uiEventHandlerFunc,useCapture};return JSEvents.registerOrRemoveHandler(eventHandler)};function _emscripten_set_resize_callback_on_thread(target,userData,useCapture,callbackfunc,targetThread){target>>>=0;userData>>>=0;callbackfunc>>>=0;targetThread>>>=0;return registerUiEventCallback(target,userData,useCapture,callbackfunc,10,"resize",targetThread)}_emscripten_set_resize_callback_on_thread.sig="ippipp";function _emscripten_set_scroll_callback_on_thread(target,userData,useCapture,callbackfunc,targetThread){target>>>=0;userData>>>=0;callbackfunc>>>=0;targetThread>>>=0;return registerUiEventCallback(target,userData,useCapture,callbackfunc,11,"scroll",targetThread)}_emscripten_set_scroll_callback_on_thread.sig="ippipp";var registerFocusEventCallback=(target,userData,useCapture,callbackfunc,eventTypeId,eventTypeString,targetThread)=>{JSEvents.focusEvent||=_malloc(256);var focusEventHandlerFunc=(e=event)=>{var nodeName=JSEvents.getNodeNameForTarget(e.target);var id=e.target.id?e.target.id:"";var focusEvent=JSEvents.focusEvent;stringToUTF8(nodeName,focusEvent+0,128);stringToUTF8(id,focusEvent+128,128);if(getWasmTableEntry(callbackfunc)(eventTypeId,focusEvent,userData))e.preventDefault()};var eventHandler={target:findEventTarget(target),eventTypeString,callbackfunc,handlerFunc:focusEventHandlerFunc,useCapture};return JSEvents.registerOrRemoveHandler(eventHandler)};function _emscripten_set_blur_callback_on_thread(target,userData,useCapture,callbackfunc,targetThread){target>>>=0;userData>>>=0;callbackfunc>>>=0;targetThread>>>=0;return registerFocusEventCallback(target,userData,useCapture,callbackfunc,12,"blur",targetThread)}_emscripten_set_blur_callback_on_thread.sig="ippipp";function _emscripten_set_focus_callback_on_thread(target,userData,useCapture,callbackfunc,targetThread){target>>>=0;userData>>>=0;callbackfunc>>>=0;targetThread>>>=0;return registerFocusEventCallback(target,userData,useCapture,callbackfunc,13,"focus",targetThread)}_emscripten_set_focus_callback_on_thread.sig="ippipp";function _emscripten_set_focusin_callback_on_thread(target,userData,useCapture,callbackfunc,targetThread){target>>>=0;userData>>>=0;callbackfunc>>>=0;targetThread>>>=0;return registerFocusEventCallback(target,userData,useCapture,callbackfunc,14,"focusin",targetThread)}_emscripten_set_focusin_callback_on_thread.sig="ippipp";function _emscripten_set_focusout_callback_on_thread(target,userData,useCapture,callbackfunc,targetThread){target>>>=0;userData>>>=0;callbackfunc>>>=0;targetThread>>>=0;return registerFocusEventCallback(target,userData,useCapture,callbackfunc,15,"focusout",targetThread)}_emscripten_set_focusout_callback_on_thread.sig="ippipp";var fillDeviceOrientationEventData=(eventStruct,e,target)=>{HEAPF64[eventStruct>>>3>>>0]=e.alpha;HEAPF64[eventStruct+8>>>3>>>0]=e.beta;HEAPF64[eventStruct+16>>>3>>>0]=e.gamma;HEAP8[eventStruct+24>>>0]=e.absolute};var registerDeviceOrientationEventCallback=(target,userData,useCapture,callbackfunc,eventTypeId,eventTypeString,targetThread)=>{JSEvents.deviceOrientationEvent||=_malloc(32);var deviceOrientationEventHandlerFunc=(e=event)=>{fillDeviceOrientationEventData(JSEvents.deviceOrientationEvent,e,target);if(getWasmTableEntry(callbackfunc)(eventTypeId,JSEvents.deviceOrientationEvent,userData))e.preventDefault()};var eventHandler={target:findEventTarget(target),eventTypeString,callbackfunc,handlerFunc:deviceOrientationEventHandlerFunc,useCapture};return JSEvents.registerOrRemoveHandler(eventHandler)};function _emscripten_set_deviceorientation_callback_on_thread(userData,useCapture,callbackfunc,targetThread){userData>>>=0;callbackfunc>>>=0;targetThread>>>=0;return registerDeviceOrientationEventCallback(2,userData,useCapture,callbackfunc,16,"deviceorientation",targetThread)}_emscripten_set_deviceorientation_callback_on_thread.sig="ipipp";function _emscripten_get_deviceorientation_status(orientationState){orientationState>>>=0;if(!JSEvents.deviceOrientationEvent)return-7;JSEvents.memcpy(orientationState,JSEvents.deviceOrientationEvent,32);return 0}_emscripten_get_deviceorientation_status.sig="ip";var fillDeviceMotionEventData=(eventStruct,e,target)=>{var supportedFields=0;var a=e["acceleration"];supportedFields|=a&&1;var ag=e["accelerationIncludingGravity"];supportedFields|=ag&&2;var rr=e["rotationRate"];supportedFields|=rr&&4;a=a||{};ag=ag||{};rr=rr||{};HEAPF64[eventStruct>>>3>>>0]=a["x"];HEAPF64[eventStruct+8>>>3>>>0]=a["y"];HEAPF64[eventStruct+16>>>3>>>0]=a["z"];HEAPF64[eventStruct+24>>>3>>>0]=ag["x"];HEAPF64[eventStruct+32>>>3>>>0]=ag["y"];HEAPF64[eventStruct+40>>>3>>>0]=ag["z"];HEAPF64[eventStruct+48>>>3>>>0]=rr["alpha"];HEAPF64[eventStruct+56>>>3>>>0]=rr["beta"];HEAPF64[eventStruct+64>>>3>>>0]=rr["gamma"]};var registerDeviceMotionEventCallback=(target,userData,useCapture,callbackfunc,eventTypeId,eventTypeString,targetThread)=>{JSEvents.deviceMotionEvent||=_malloc(80);var deviceMotionEventHandlerFunc=(e=event)=>{fillDeviceMotionEventData(JSEvents.deviceMotionEvent,e,target);if(getWasmTableEntry(callbackfunc)(eventTypeId,JSEvents.deviceMotionEvent,userData))e.preventDefault()};var eventHandler={target:findEventTarget(target),eventTypeString,callbackfunc,handlerFunc:deviceMotionEventHandlerFunc,useCapture};return JSEvents.registerOrRemoveHandler(eventHandler)};function _emscripten_set_devicemotion_callback_on_thread(userData,useCapture,callbackfunc,targetThread){userData>>>=0;callbackfunc>>>=0;targetThread>>>=0;return registerDeviceMotionEventCallback(2,userData,useCapture,callbackfunc,17,"devicemotion",targetThread)}_emscripten_set_devicemotion_callback_on_thread.sig="ipipp";function _emscripten_get_devicemotion_status(motionState){motionState>>>=0;if(!JSEvents.deviceMotionEvent)return-7;JSEvents.memcpy(motionState,JSEvents.deviceMotionEvent,80);return 0}_emscripten_get_devicemotion_status.sig="ip";var screenOrientation=()=>{if(!window.screen)return undefined;return screen.orientation||screen["mozOrientation"]||screen["webkitOrientation"]};var fillOrientationChangeEventData=eventStruct=>{var orientationsType1=["portrait-primary","portrait-secondary","landscape-primary","landscape-secondary"];var orientationsType2=["portrait","portrait","landscape","landscape"];var orientationIndex=0;var orientationAngle=0;var screenOrientObj=screenOrientation();if(typeof screenOrientObj==="object"){orientationIndex=orientationsType1.indexOf(screenOrientObj.type);if(orientationIndex<0){orientationIndex=orientationsType2.indexOf(screenOrientObj.type)}if(orientationIndex>=0){orientationIndex=1<<orientationIndex}orientationAngle=screenOrientObj.angle}else{orientationAngle=window.orientation}HEAP32[eventStruct>>>2>>>0]=orientationIndex;HEAP32[eventStruct+4>>>2>>>0]=orientationAngle};var registerOrientationChangeEventCallback=(target,userData,useCapture,callbackfunc,eventTypeId,eventTypeString,targetThread)=>{JSEvents.orientationChangeEvent||=_malloc(8);var orientationChangeEventHandlerFunc=(e=event)=>{var orientationChangeEvent=JSEvents.orientationChangeEvent;fillOrientationChangeEventData(orientationChangeEvent);if(getWasmTableEntry(callbackfunc)(eventTypeId,orientationChangeEvent,userData))e.preventDefault()};var eventHandler={target,eventTypeString,callbackfunc,handlerFunc:orientationChangeEventHandlerFunc,useCapture};return JSEvents.registerOrRemoveHandler(eventHandler)};function _emscripten_set_orientationchange_callback_on_thread(userData,useCapture,callbackfunc,targetThread){userData>>>=0;callbackfunc>>>=0;targetThread>>>=0;if(!window.screen||!screen.orientation)return-1;return registerOrientationChangeEventCallback(screen.orientation,userData,useCapture,callbackfunc,18,"change",targetThread)}_emscripten_set_orientationchange_callback_on_thread.sig="ipipp";function _emscripten_get_orientation_status(orientationChangeEvent){orientationChangeEvent>>>=0;if(!screenOrientation()&&typeof orientation=="undefined")return-1;fillOrientationChangeEventData(orientationChangeEvent);return 0}_emscripten_get_orientation_status.sig="ip";var _emscripten_lock_orientation=allowedOrientations=>{var orientations=[];if(allowedOrientations&1)orientations.push("portrait-primary");if(allowedOrientations&2)orientations.push("portrait-secondary");if(allowedOrientations&4)orientations.push("landscape-primary");if(allowedOrientations&8)orientations.push("landscape-secondary");var succeeded;if(screen.lockOrientation){succeeded=screen.lockOrientation(orientations)}else if(screen.mozLockOrientation){succeeded=screen.mozLockOrientation(orientations)}else if(screen.webkitLockOrientation){succeeded=screen.webkitLockOrientation(orientations)}else{return-1}if(succeeded){return 0}return-6};_emscripten_lock_orientation.sig="ii";var _emscripten_unlock_orientation=()=>{if(screen.unlockOrientation){screen.unlockOrientation()}else if(screen.mozUnlockOrientation){screen.mozUnlockOrientation()}else if(screen.webkitUnlockOrientation){screen.webkitUnlockOrientation()}else{return-1}return 0};_emscripten_unlock_orientation.sig="i";var fillFullscreenChangeEventData=eventStruct=>{var fullscreenElement=document.fullscreenElement||document.mozFullScreenElement||document.webkitFullscreenElement||document.msFullscreenElement;var isFullscreen=!!fullscreenElement;HEAP8[eventStruct>>>0]=isFullscreen;HEAP8[eventStruct+1>>>0]=JSEvents.fullscreenEnabled();var reportedElement=isFullscreen?fullscreenElement:JSEvents.previousFullscreenElement;var nodeName=JSEvents.getNodeNameForTarget(reportedElement);var id=reportedElement?.id||"";stringToUTF8(nodeName,eventStruct+2,128);stringToUTF8(id,eventStruct+130,128);HEAP32[eventStruct+260>>>2>>>0]=reportedElement?reportedElement.clientWidth:0;HEAP32[eventStruct+264>>>2>>>0]=reportedElement?reportedElement.clientHeight:0;HEAP32[eventStruct+268>>>2>>>0]=screen.width;HEAP32[eventStruct+272>>>2>>>0]=screen.height;if(isFullscreen){JSEvents.previousFullscreenElement=fullscreenElement}};var registerFullscreenChangeEventCallback=(target,userData,useCapture,callbackfunc,eventTypeId,eventTypeString,targetThread)=>{JSEvents.fullscreenChangeEvent||=_malloc(276);var fullscreenChangeEventhandlerFunc=(e=event)=>{var fullscreenChangeEvent=JSEvents.fullscreenChangeEvent;fillFullscreenChangeEventData(fullscreenChangeEvent);if(getWasmTableEntry(callbackfunc)(eventTypeId,fullscreenChangeEvent,userData))e.preventDefault()};var eventHandler={target,eventTypeString,callbackfunc,handlerFunc:fullscreenChangeEventhandlerFunc,useCapture};return JSEvents.registerOrRemoveHandler(eventHandler)};function _emscripten_set_fullscreenchange_callback_on_thread(target,userData,useCapture,callbackfunc,targetThread){target>>>=0;userData>>>=0;callbackfunc>>>=0;targetThread>>>=0;if(!JSEvents.fullscreenEnabled())return-1;target=findEventTarget(target);if(!target)return-4;registerFullscreenChangeEventCallback(target,userData,useCapture,callbackfunc,19,"webkitfullscreenchange",targetThread);return registerFullscreenChangeEventCallback(target,userData,useCapture,callbackfunc,19,"fullscreenchange",targetThread)}_emscripten_set_fullscreenchange_callback_on_thread.sig="ippipp";function _emscripten_get_fullscreen_status(fullscreenStatus){fullscreenStatus>>>=0;if(!JSEvents.fullscreenEnabled())return-1;fillFullscreenChangeEventData(fullscreenStatus);return 0}_emscripten_get_fullscreen_status.sig="ip";function _emscripten_get_canvas_element_size(target,width,height){target>>>=0;width>>>=0;height>>>=0;var canvas=findCanvasEventTarget(target);if(!canvas)return-4;HEAP32[width>>>2>>>0]=canvas.width;HEAP32[height>>>2>>>0]=canvas.height}_emscripten_get_canvas_element_size.sig="ippp";var getCanvasElementSize=target=>{var sp=stackSave();var w=stackAlloc(8);var h=w+4;var targetInt=stringToUTF8OnStack(target.id);var ret=_emscripten_get_canvas_element_size(targetInt,w,h);var size=[HEAP32[w>>>2>>>0],HEAP32[h>>>2>>>0]];stackRestore(sp);return size};function _emscripten_set_canvas_element_size(target,width,height){target>>>=0;var canvas=findCanvasEventTarget(target);if(!canvas)return-4;canvas.width=width;canvas.height=height;return 0}_emscripten_set_canvas_element_size.sig="ipii";var setCanvasElementSize=(target,width,height)=>{if(!target.controlTransferredOffscreen){target.width=width;target.height=height}else{var sp=stackSave();var targetInt=stringToUTF8OnStack(target.id);_emscripten_set_canvas_element_size(targetInt,width,height);stackRestore(sp)}};var currentFullscreenStrategy={};var registerRestoreOldStyle=canvas=>{var canvasSize=getCanvasElementSize(canvas);var oldWidth=canvasSize[0];var oldHeight=canvasSize[1];var oldCssWidth=canvas.style.width;var oldCssHeight=canvas.style.height;var oldBackgroundColor=canvas.style.backgroundColor;var oldDocumentBackgroundColor=document.body.style.backgroundColor;var oldPaddingLeft=canvas.style.paddingLeft;var oldPaddingRight=canvas.style.paddingRight;var oldPaddingTop=canvas.style.paddingTop;var oldPaddingBottom=canvas.style.paddingBottom;var oldMarginLeft=canvas.style.marginLeft;var oldMarginRight=canvas.style.marginRight;var oldMarginTop=canvas.style.marginTop;var oldMarginBottom=canvas.style.marginBottom;var oldDocumentBodyMargin=document.body.style.margin;var oldDocumentOverflow=document.documentElement.style.overflow;var oldDocumentScroll=document.body.scroll;var oldImageRendering=canvas.style.imageRendering;function restoreOldStyle(){var fullscreenElement=document.fullscreenElement||document.webkitFullscreenElement;if(!fullscreenElement){document.removeEventListener("fullscreenchange",restoreOldStyle);document.removeEventListener("webkitfullscreenchange",restoreOldStyle);setCanvasElementSize(canvas,oldWidth,oldHeight);canvas.style.width=oldCssWidth;canvas.style.height=oldCssHeight;canvas.style.backgroundColor=oldBackgroundColor;if(!oldDocumentBackgroundColor)document.body.style.backgroundColor="white";document.body.style.backgroundColor=oldDocumentBackgroundColor;canvas.style.paddingLeft=oldPaddingLeft;canvas.style.paddingRight=oldPaddingRight;canvas.style.paddingTop=oldPaddingTop;canvas.style.paddingBottom=oldPaddingBottom;canvas.style.marginLeft=oldMarginLeft;canvas.style.marginRight=oldMarginRight;canvas.style.marginTop=oldMarginTop;canvas.style.marginBottom=oldMarginBottom;document.body.style.margin=oldDocumentBodyMargin;document.documentElement.style.overflow=oldDocumentOverflow;document.body.scroll=oldDocumentScroll;canvas.style.imageRendering=oldImageRendering;if(canvas.GLctxObject)canvas.GLctxObject.GLctx.viewport(0,0,oldWidth,oldHeight);if(currentFullscreenStrategy.canvasResizedCallback){getWasmTableEntry(currentFullscreenStrategy.canvasResizedCallback)(37,0,currentFullscreenStrategy.canvasResizedCallbackUserData)}}}document.addEventListener("fullscreenchange",restoreOldStyle);document.addEventListener("webkitfullscreenchange",restoreOldStyle);return restoreOldStyle};var setLetterbox=(element,topBottom,leftRight)=>{element.style.paddingLeft=element.style.paddingRight=leftRight+"px";element.style.paddingTop=element.style.paddingBottom=topBottom+"px"};var JSEvents_resizeCanvasForFullscreen=(target,strategy)=>{var restoreOldStyle=registerRestoreOldStyle(target);var cssWidth=strategy.softFullscreen?innerWidth:screen.width;var cssHeight=strategy.softFullscreen?innerHeight:screen.height;var rect=getBoundingClientRect(target);var windowedCssWidth=rect.width;var windowedCssHeight=rect.height;var canvasSize=getCanvasElementSize(target);var windowedRttWidth=canvasSize[0];var windowedRttHeight=canvasSize[1];if(strategy.scaleMode==3){setLetterbox(target,(cssHeight-windowedCssHeight)/2,(cssWidth-windowedCssWidth)/2);cssWidth=windowedCssWidth;cssHeight=windowedCssHeight}else if(strategy.scaleMode==2){if(cssWidth*windowedRttHeight<windowedRttWidth*cssHeight){var desiredCssHeight=windowedRttHeight*cssWidth/windowedRttWidth;setLetterbox(target,(cssHeight-desiredCssHeight)/2,0);cssHeight=desiredCssHeight}else{var desiredCssWidth=windowedRttWidth*cssHeight/windowedRttHeight;setLetterbox(target,0,(cssWidth-desiredCssWidth)/2);cssWidth=desiredCssWidth}}target.style.backgroundColor||="black";document.body.style.backgroundColor||="black";target.style.width=cssWidth+"px";target.style.height=cssHeight+"px";if(strategy.filteringMode==1){target.style.imageRendering="optimizeSpeed";target.style.imageRendering="-moz-crisp-edges";target.style.imageRendering="-o-crisp-edges";target.style.imageRendering="-webkit-optimize-contrast";target.style.imageRendering="optimize-contrast";target.style.imageRendering="crisp-edges";target.style.imageRendering="pixelated"}var dpiScale=strategy.canvasResolutionScaleMode==2?devicePixelRatio:1;if(strategy.canvasResolutionScaleMode!=0){var newWidth=cssWidth*dpiScale|0;var newHeight=cssHeight*dpiScale|0;setCanvasElementSize(target,newWidth,newHeight);if(target.GLctxObject)target.GLctxObject.GLctx.viewport(0,0,newWidth,newHeight)}return restoreOldStyle};var JSEvents_requestFullscreen=(target,strategy)=>{if(strategy.scaleMode!=0||strategy.canvasResolutionScaleMode!=0){JSEvents_resizeCanvasForFullscreen(target,strategy)}if(target.requestFullscreen){target.requestFullscreen()}else if(target.webkitRequestFullscreen){target.webkitRequestFullscreen(Element.ALLOW_KEYBOARD_INPUT)}else{return JSEvents.fullscreenEnabled()?-3:-1}currentFullscreenStrategy=strategy;if(strategy.canvasResizedCallback){getWasmTableEntry(strategy.canvasResizedCallback)(37,0,strategy.canvasResizedCallbackUserData)}return 0};var hideEverythingExceptGivenElement=onlyVisibleElement=>{var child=onlyVisibleElement;var parent=child.parentNode;var hiddenElements=[];while(child!=document.body){var children=parent.children;for(var currChild of children){if(currChild!=child){hiddenElements.push({node:currChild,displayState:currChild.style.display});currChild.style.display="none"}}child=parent;parent=parent.parentNode}return hiddenElements};var restoreHiddenElements=hiddenElements=>{for(var elem of hiddenElements){elem.node.style.display=elem.displayState}};var restoreOldWindowedStyle=null;var softFullscreenResizeWebGLRenderTarget=()=>{var dpr=devicePixelRatio;var inHiDPIFullscreenMode=currentFullscreenStrategy.canvasResolutionScaleMode==2;var inAspectRatioFixedFullscreenMode=currentFullscreenStrategy.scaleMode==2;var inPixelPerfectFullscreenMode=currentFullscreenStrategy.canvasResolutionScaleMode!=0;var inCenteredWithoutScalingFullscreenMode=currentFullscreenStrategy.scaleMode==3;var screenWidth=inHiDPIFullscreenMode?Math.round(innerWidth*dpr):innerWidth;var screenHeight=inHiDPIFullscreenMode?Math.round(innerHeight*dpr):innerHeight;var w=screenWidth;var h=screenHeight;var canvas=currentFullscreenStrategy.target;var canvasSize=getCanvasElementSize(canvas);var x=canvasSize[0];var y=canvasSize[1];var topMargin;if(inAspectRatioFixedFullscreenMode){if(w*y<x*h)h=w*y/x|0;else if(w*y>x*h)w=h*x/y|0;topMargin=(screenHeight-h)/2|0}if(inPixelPerfectFullscreenMode){setCanvasElementSize(canvas,w,h);if(canvas.GLctxObject)canvas.GLctxObject.GLctx.viewport(0,0,w,h)}if(inHiDPIFullscreenMode){topMargin/=dpr;w/=dpr;h/=dpr;w=Math.round(w*1e4)/1e4;h=Math.round(h*1e4)/1e4;topMargin=Math.round(topMargin*1e4)/1e4}if(inCenteredWithoutScalingFullscreenMode){var t=(innerHeight-jstoi_q(canvas.style.height))/2;var b=(innerWidth-jstoi_q(canvas.style.width))/2;setLetterbox(canvas,t,b)}else{canvas.style.width=w+"px";canvas.style.height=h+"px";var b=(innerWidth-w)/2;setLetterbox(canvas,topMargin,b)}if(!inCenteredWithoutScalingFullscreenMode&&currentFullscreenStrategy.canvasResizedCallback){getWasmTableEntry(currentFullscreenStrategy.canvasResizedCallback)(37,0,currentFullscreenStrategy.canvasResizedCallbackUserData)}};var doRequestFullscreen=(target,strategy)=>{if(!JSEvents.fullscreenEnabled())return-1;target=findEventTarget(target);if(!target)return-4;if(!target.requestFullscreen&&!target.webkitRequestFullscreen){return-3}if(!JSEvents.canPerformEventHandlerRequests()){if(strategy.deferUntilInEventHandler){JSEvents.deferCall(JSEvents_requestFullscreen,1,[target,strategy]);return 1}return-2}return JSEvents_requestFullscreen(target,strategy)};function _emscripten_request_fullscreen(target,deferUntilInEventHandler){target>>>=0;var strategy={scaleMode:0,canvasResolutionScaleMode:0,filteringMode:0,deferUntilInEventHandler,canvasResizedCallbackTargetThread:2};return doRequestFullscreen(target,strategy)}_emscripten_request_fullscreen.sig="ipi";function _emscripten_request_fullscreen_strategy(target,deferUntilInEventHandler,fullscreenStrategy){target>>>=0;fullscreenStrategy>>>=0;var strategy={scaleMode:HEAP32[fullscreenStrategy>>>2>>>0],canvasResolutionScaleMode:HEAP32[fullscreenStrategy+4>>>2>>>0],filteringMode:HEAP32[fullscreenStrategy+8>>>2>>>0],deferUntilInEventHandler,canvasResizedCallback:HEAP32[fullscreenStrategy+12>>>2>>>0],canvasResizedCallbackUserData:HEAP32[fullscreenStrategy+16>>>2>>>0]};return doRequestFullscreen(target,strategy)}_emscripten_request_fullscreen_strategy.sig="ipip";function _emscripten_enter_soft_fullscreen(target,fullscreenStrategy){target>>>=0;fullscreenStrategy>>>=0;target=findEventTarget(target);if(!target)return-4;var strategy={scaleMode:HEAP32[fullscreenStrategy>>>2>>>0],canvasResolutionScaleMode:HEAP32[fullscreenStrategy+4>>>2>>>0],filteringMode:HEAP32[fullscreenStrategy+8>>>2>>>0],canvasResizedCallback:HEAP32[fullscreenStrategy+12>>>2>>>0],canvasResizedCallbackUserData:HEAP32[fullscreenStrategy+16>>>2>>>0],target,softFullscreen:true};var restoreOldStyle=JSEvents_resizeCanvasForFullscreen(target,strategy);document.documentElement.style.overflow="hidden";document.body.scroll="no";document.body.style.margin="0px";var hiddenElements=hideEverythingExceptGivenElement(target);function restoreWindowedState(){restoreOldStyle();restoreHiddenElements(hiddenElements);removeEventListener("resize",softFullscreenResizeWebGLRenderTarget);if(strategy.canvasResizedCallback){getWasmTableEntry(strategy.canvasResizedCallback)(37,0,strategy.canvasResizedCallbackUserData)}currentFullscreenStrategy=0}restoreOldWindowedStyle=restoreWindowedState;currentFullscreenStrategy=strategy;addEventListener("resize",softFullscreenResizeWebGLRenderTarget);if(strategy.canvasResizedCallback){getWasmTableEntry(strategy.canvasResizedCallback)(37,0,strategy.canvasResizedCallbackUserData)}return 0}_emscripten_enter_soft_fullscreen.sig="ipp";var _emscripten_exit_soft_fullscreen=()=>{restoreOldWindowedStyle?.();restoreOldWindowedStyle=null;return 0};_emscripten_exit_soft_fullscreen.sig="i";var _emscripten_exit_fullscreen=()=>{if(!JSEvents.fullscreenEnabled())return-1;JSEvents.removeDeferredCalls(JSEvents_requestFullscreen);var d=specialHTMLTargets[1];if(d.exitFullscreen){d.fullscreenElement&&d.exitFullscreen()}else if(d.webkitExitFullscreen){d.webkitFullscreenElement&&d.webkitExitFullscreen()}else{return-1}return 0};_emscripten_exit_fullscreen.sig="i";var fillPointerlockChangeEventData=eventStruct=>{var pointerLockElement=document.pointerLockElement||document.mozPointerLockElement||document.webkitPointerLockElement||document.msPointerLockElement;var isPointerlocked=!!pointerLockElement;HEAP8[eventStruct>>>0]=isPointerlocked;var nodeName=JSEvents.getNodeNameForTarget(pointerLockElement);var id=pointerLockElement?.id||"";stringToUTF8(nodeName,eventStruct+1,128);stringToUTF8(id,eventStruct+129,128)};var registerPointerlockChangeEventCallback=(target,userData,useCapture,callbackfunc,eventTypeId,eventTypeString,targetThread)=>{JSEvents.pointerlockChangeEvent||=_malloc(257);var pointerlockChangeEventHandlerFunc=(e=event)=>{var pointerlockChangeEvent=JSEvents.pointerlockChangeEvent;fillPointerlockChangeEventData(pointerlockChangeEvent);if(getWasmTableEntry(callbackfunc)(eventTypeId,pointerlockChangeEvent,userData))e.preventDefault()};var eventHandler={target,eventTypeString,callbackfunc,handlerFunc:pointerlockChangeEventHandlerFunc,useCapture};return JSEvents.registerOrRemoveHandler(eventHandler)};function _emscripten_set_pointerlockchange_callback_on_thread(target,userData,useCapture,callbackfunc,targetThread){target>>>=0;userData>>>=0;callbackfunc>>>=0;targetThread>>>=0;if(!document||!document.body||!document.body.requestPointerLock&&!document.body.mozRequestPointerLock&&!document.body.webkitRequestPointerLock&&!document.body.msRequestPointerLock){return-1}target=findEventTarget(target);if(!target)return-4;registerPointerlockChangeEventCallback(target,userData,useCapture,callbackfunc,20,"mozpointerlockchange",targetThread);registerPointerlockChangeEventCallback(target,userData,useCapture,callbackfunc,20,"webkitpointerlockchange",targetThread);registerPointerlockChangeEventCallback(target,userData,useCapture,callbackfunc,20,"mspointerlockchange",targetThread);return registerPointerlockChangeEventCallback(target,userData,useCapture,callbackfunc,20,"pointerlockchange",targetThread)}_emscripten_set_pointerlockchange_callback_on_thread.sig="ippipp";var registerPointerlockErrorEventCallback=(target,userData,useCapture,callbackfunc,eventTypeId,eventTypeString,targetThread)=>{var pointerlockErrorEventHandlerFunc=(e=event)=>{if(getWasmTableEntry(callbackfunc)(eventTypeId,0,userData))e.preventDefault()};var eventHandler={target,eventTypeString,callbackfunc,handlerFunc:pointerlockErrorEventHandlerFunc,useCapture};return JSEvents.registerOrRemoveHandler(eventHandler)};function _emscripten_set_pointerlockerror_callback_on_thread(target,userData,useCapture,callbackfunc,targetThread){target>>>=0;userData>>>=0;callbackfunc>>>=0;targetThread>>>=0;if(!document||!document.body.requestPointerLock&&!document.body.mozRequestPointerLock&&!document.body.webkitRequestPointerLock&&!document.body.msRequestPointerLock){return-1}target=findEventTarget(target);if(!target)return-4;registerPointerlockErrorEventCallback(target,userData,useCapture,callbackfunc,38,"mozpointerlockerror",targetThread);registerPointerlockErrorEventCallback(target,userData,useCapture,callbackfunc,38,"webkitpointerlockerror",targetThread);registerPointerlockErrorEventCallback(target,userData,useCapture,callbackfunc,38,"mspointerlockerror",targetThread);return registerPointerlockErrorEventCallback(target,userData,useCapture,callbackfunc,38,"pointerlockerror",targetThread)}_emscripten_set_pointerlockerror_callback_on_thread.sig="ippipp";function _emscripten_get_pointerlock_status(pointerlockStatus){pointerlockStatus>>>=0;if(pointerlockStatus)fillPointerlockChangeEventData(pointerlockStatus);if(!document.body||!document.body.requestPointerLock&&!document.body.mozRequestPointerLock&&!document.body.webkitRequestPointerLock&&!document.body.msRequestPointerLock){return-1}return 0}_emscripten_get_pointerlock_status.sig="ip";var requestPointerLock=target=>{if(target.requestPointerLock){target.requestPointerLock()}else{if(document.body.requestPointerLock){return-3}return-1}return 0};function _emscripten_request_pointerlock(target,deferUntilInEventHandler){target>>>=0;target=findEventTarget(target);if(!target)return-4;if(!target.requestPointerLock){return-1}if(!JSEvents.canPerformEventHandlerRequests()){if(deferUntilInEventHandler){JSEvents.deferCall(requestPointerLock,2,[target]);return 1}return-2}return requestPointerLock(target)}_emscripten_request_pointerlock.sig="ipi";var _emscripten_exit_pointerlock=()=>{JSEvents.removeDeferredCalls(requestPointerLock);if(document.exitPointerLock){document.exitPointerLock()}else{return-1}return 0};_emscripten_exit_pointerlock.sig="i";var _emscripten_vibrate=msecs=>{if(!navigator.vibrate)return-1;navigator.vibrate(msecs);return 0};_emscripten_vibrate.sig="ii";function _emscripten_vibrate_pattern(msecsArray,numEntries){msecsArray>>>=0;if(!navigator.vibrate)return-1;var vibrateList=[];for(var i=0;i<numEntries;++i){var msecs=HEAP32[msecsArray+i*4>>>2>>>0];vibrateList.push(msecs)}navigator.vibrate(vibrateList);return 0}_emscripten_vibrate_pattern.sig="ipi";var fillVisibilityChangeEventData=eventStruct=>{var visibilityStates=["hidden","visible","prerender","unloaded"];var visibilityState=visibilityStates.indexOf(document.visibilityState);HEAP8[eventStruct>>>0]=document.hidden;HEAP32[eventStruct+4>>>2>>>0]=visibilityState};var registerVisibilityChangeEventCallback=(target,userData,useCapture,callbackfunc,eventTypeId,eventTypeString,targetThread)=>{JSEvents.visibilityChangeEvent||=_malloc(8);var visibilityChangeEventHandlerFunc=(e=event)=>{var visibilityChangeEvent=JSEvents.visibilityChangeEvent;fillVisibilityChangeEventData(visibilityChangeEvent);if(getWasmTableEntry(callbackfunc)(eventTypeId,visibilityChangeEvent,userData))e.preventDefault()};var eventHandler={target,eventTypeString,callbackfunc,handlerFunc:visibilityChangeEventHandlerFunc,useCapture};return JSEvents.registerOrRemoveHandler(eventHandler)};function _emscripten_set_visibilitychange_callback_on_thread(userData,useCapture,callbackfunc,targetThread){userData>>>=0;callbackfunc>>>=0;targetThread>>>=0;if(!specialHTMLTargets[1]){return-4}return registerVisibilityChangeEventCallback(specialHTMLTargets[1],userData,useCapture,callbackfunc,21,"visibilitychange",targetThread)}_emscripten_set_visibilitychange_callback_on_thread.sig="ipipp";function _emscripten_get_visibility_status(visibilityStatus){visibilityStatus>>>=0;if(typeof document.visibilityState=="undefined"&&typeof document.hidden=="undefined"){return-1}fillVisibilityChangeEventData(visibilityStatus);return 0}_emscripten_get_visibility_status.sig="ip";var registerTouchEventCallback=(target,userData,useCapture,callbackfunc,eventTypeId,eventTypeString,targetThread)=>{JSEvents.touchEvent||=_malloc(1552);target=findEventTarget(target);var touchEventHandlerFunc=e=>{var t,touches={},et=e.touches;for(let t of et){t.isChanged=t.onTarget=0;touches[t.identifier]=t}for(let t of e.changedTouches){t.isChanged=1;touches[t.identifier]=t}for(let t of e.targetTouches){touches[t.identifier].onTarget=1}var touchEvent=JSEvents.touchEvent;HEAPF64[touchEvent>>>3>>>0]=e.timeStamp;HEAP8[touchEvent+12>>>0]=e.ctrlKey;HEAP8[touchEvent+13>>>0]=e.shiftKey;HEAP8[touchEvent+14>>>0]=e.altKey;HEAP8[touchEvent+15>>>0]=e.metaKey;var idx=touchEvent+16;var targetRect=getBoundingClientRect(target);var numTouches=0;for(let t of Object.values(touches)){var idx32=idx>>>2;HEAP32[idx32+0>>>0]=t.identifier;HEAP32[idx32+1>>>0]=t.screenX;HEAP32[idx32+2>>>0]=t.screenY;HEAP32[idx32+3>>>0]=t.clientX;HEAP32[idx32+4>>>0]=t.clientY;HEAP32[idx32+5>>>0]=t.pageX;HEAP32[idx32+6>>>0]=t.pageY;HEAP8[idx+28>>>0]=t.isChanged;HEAP8[idx+29>>>0]=t.onTarget;HEAP32[idx32+8>>>0]=t.clientX-(targetRect.left|0);HEAP32[idx32+9>>>0]=t.clientY-(targetRect.top|0);idx+=48;if(++numTouches>31){break}}HEAP32[touchEvent+8>>>2>>>0]=numTouches;if(getWasmTableEntry(callbackfunc)(eventTypeId,touchEvent,userData))e.preventDefault()};var eventHandler={target,allowsDeferredCalls:eventTypeString=="touchstart"||eventTypeString=="touchend",eventTypeString,callbackfunc,handlerFunc:touchEventHandlerFunc,useCapture};return JSEvents.registerOrRemoveHandler(eventHandler)};function _emscripten_set_touchstart_callback_on_thread(target,userData,useCapture,callbackfunc,targetThread){target>>>=0;userData>>>=0;callbackfunc>>>=0;targetThread>>>=0;return registerTouchEventCallback(target,userData,useCapture,callbackfunc,22,"touchstart",targetThread)}_emscripten_set_touchstart_callback_on_thread.sig="ippipp";function _emscripten_set_touchend_callback_on_thread(target,userData,useCapture,callbackfunc,targetThread){target>>>=0;userData>>>=0;callbackfunc>>>=0;targetThread>>>=0;return registerTouchEventCallback(target,userData,useCapture,callbackfunc,23,"touchend",targetThread)}_emscripten_set_touchend_callback_on_thread.sig="ippipp";function _emscripten_set_touchmove_callback_on_thread(target,userData,useCapture,callbackfunc,targetThread){target>>>=0;userData>>>=0;callbackfunc>>>=0;targetThread>>>=0;return registerTouchEventCallback(target,userData,useCapture,callbackfunc,24,"touchmove",targetThread)}_emscripten_set_touchmove_callback_on_thread.sig="ippipp";function _emscripten_set_touchcancel_callback_on_thread(target,userData,useCapture,callbackfunc,targetThread){target>>>=0;userData>>>=0;callbackfunc>>>=0;targetThread>>>=0;return registerTouchEventCallback(target,userData,useCapture,callbackfunc,25,"touchcancel",targetThread)}_emscripten_set_touchcancel_callback_on_thread.sig="ippipp";var fillGamepadEventData=(eventStruct,e)=>{HEAPF64[eventStruct>>>3>>>0]=e.timestamp;for(var i=0;i<e.axes.length;++i){HEAPF64[eventStruct+i*8+16>>>3>>>0]=e.axes[i]}for(var i=0;i<e.buttons.length;++i){if(typeof e.buttons[i]=="object"){HEAPF64[eventStruct+i*8+528>>>3>>>0]=e.buttons[i].value}else{HEAPF64[eventStruct+i*8+528>>>3>>>0]=e.buttons[i]}}for(var i=0;i<e.buttons.length;++i){if(typeof e.buttons[i]=="object"){HEAP8[eventStruct+i+1040>>>0]=e.buttons[i].pressed}else{HEAP8[eventStruct+i+1040>>>0]=e.buttons[i]==1}}HEAP8[eventStruct+1104>>>0]=e.connected;HEAP32[eventStruct+1108>>>2>>>0]=e.index;HEAP32[eventStruct+8>>>2>>>0]=e.axes.length;HEAP32[eventStruct+12>>>2>>>0]=e.buttons.length;stringToUTF8(e.id,eventStruct+1112,64);stringToUTF8(e.mapping,eventStruct+1176,64)};var registerGamepadEventCallback=(target,userData,useCapture,callbackfunc,eventTypeId,eventTypeString,targetThread)=>{JSEvents.gamepadEvent||=_malloc(1240);var gamepadEventHandlerFunc=(e=event)=>{var gamepadEvent=JSEvents.gamepadEvent;fillGamepadEventData(gamepadEvent,e["gamepad"]);if(getWasmTableEntry(callbackfunc)(eventTypeId,gamepadEvent,userData))e.preventDefault()};var eventHandler={target:findEventTarget(target),allowsDeferredCalls:true,eventTypeString,callbackfunc,handlerFunc:gamepadEventHandlerFunc,useCapture};return JSEvents.registerOrRemoveHandler(eventHandler)};var _emscripten_sample_gamepad_data=()=>{try{if(navigator.getGamepads)return(JSEvents.lastGamepadState=navigator.getGamepads())?0:-1}catch(e){navigator.getGamepads=null}return-1};_emscripten_sample_gamepad_data.sig="i";function _emscripten_set_gamepadconnected_callback_on_thread(userData,useCapture,callbackfunc,targetThread){userData>>>=0;callbackfunc>>>=0;targetThread>>>=0;if(_emscripten_sample_gamepad_data())return-1;return registerGamepadEventCallback(2,userData,useCapture,callbackfunc,26,"gamepadconnected",targetThread)}_emscripten_set_gamepadconnected_callback_on_thread.sig="ipipp";function _emscripten_set_gamepaddisconnected_callback_on_thread(userData,useCapture,callbackfunc,targetThread){userData>>>=0;callbackfunc>>>=0;targetThread>>>=0;if(_emscripten_sample_gamepad_data())return-1;return registerGamepadEventCallback(2,userData,useCapture,callbackfunc,27,"gamepaddisconnected",targetThread)}_emscripten_set_gamepaddisconnected_callback_on_thread.sig="ipipp";var _emscripten_get_num_gamepads=()=>JSEvents.lastGamepadState.length;_emscripten_get_num_gamepads.sig="i";function _emscripten_get_gamepad_status(index,gamepadState){gamepadState>>>=0;if(index<0||index>=JSEvents.lastGamepadState.length)return-5;if(!JSEvents.lastGamepadState[index])return-7;fillGamepadEventData(gamepadState,JSEvents.lastGamepadState[index]);return 0}_emscripten_get_gamepad_status.sig="iip";var registerBeforeUnloadEventCallback=(target,userData,useCapture,callbackfunc,eventTypeId,eventTypeString)=>{var beforeUnloadEventHandlerFunc=(e=event)=>{var confirmationMessage=getWasmTableEntry(callbackfunc)(eventTypeId,0,userData);if(confirmationMessage){confirmationMessage=UTF8ToString(confirmationMessage)}if(confirmationMessage){e.preventDefault();e.returnValue=confirmationMessage;return confirmationMessage}};var eventHandler={target:findEventTarget(target),eventTypeString,callbackfunc,handlerFunc:beforeUnloadEventHandlerFunc,useCapture};return JSEvents.registerOrRemoveHandler(eventHandler)};function _emscripten_set_beforeunload_callback_on_thread(userData,callbackfunc,targetThread){userData>>>=0;callbackfunc>>>=0;targetThread>>>=0;if(typeof onbeforeunload=="undefined")return-1;if(targetThread!==1)return-5;return registerBeforeUnloadEventCallback(2,userData,true,callbackfunc,28,"beforeunload")}_emscripten_set_beforeunload_callback_on_thread.sig="ippp";var fillBatteryEventData=(eventStruct,e)=>{HEAPF64[eventStruct>>>3>>>0]=e.chargingTime;HEAPF64[eventStruct+8>>>3>>>0]=e.dischargingTime;HEAPF64[eventStruct+16>>>3>>>0]=e.level;HEAP8[eventStruct+24>>>0]=e.charging};var battery=()=>navigator.battery||navigator.mozBattery||navigator.webkitBattery;var registerBatteryEventCallback=(target,userData,useCapture,callbackfunc,eventTypeId,eventTypeString,targetThread)=>{JSEvents.batteryEvent||=_malloc(32);var batteryEventHandlerFunc=(e=event)=>{var batteryEvent=JSEvents.batteryEvent;fillBatteryEventData(batteryEvent,battery());if(getWasmTableEntry(callbackfunc)(eventTypeId,batteryEvent,userData))e.preventDefault()};var eventHandler={target:findEventTarget(target),eventTypeString,callbackfunc,handlerFunc:batteryEventHandlerFunc,useCapture};return JSEvents.registerOrRemoveHandler(eventHandler)};function _emscripten_set_batterychargingchange_callback_on_thread(userData,callbackfunc,targetThread){userData>>>=0;callbackfunc>>>=0;targetThread>>>=0;if(!battery())return-1;return registerBatteryEventCallback(battery(),userData,true,callbackfunc,29,"chargingchange",targetThread)}_emscripten_set_batterychargingchange_callback_on_thread.sig="ippp";function _emscripten_set_batterylevelchange_callback_on_thread(userData,callbackfunc,targetThread){userData>>>=0;callbackfunc>>>=0;targetThread>>>=0;if(!battery())return-1;return registerBatteryEventCallback(battery(),userData,true,callbackfunc,30,"levelchange",targetThread)}_emscripten_set_batterylevelchange_callback_on_thread.sig="ippp";function _emscripten_get_battery_status(batteryState){batteryState>>>=0;if(!battery())return-1;fillBatteryEventData(batteryState,battery());return 0}_emscripten_get_battery_status.sig="ip";function _emscripten_set_element_css_size(target,width,height){target>>>=0;target=findEventTarget(target);if(!target)return-4;target.style.width=width+"px";target.style.height=height+"px";return 0}_emscripten_set_element_css_size.sig="ipdd";function _emscripten_get_element_css_size(target,width,height){target>>>=0;width>>>=0;height>>>=0;target=findEventTarget(target);if(!target)return-4;var rect=getBoundingClientRect(target);HEAPF64[width>>>3>>>0]=rect.width;HEAPF64[height>>>3>>>0]=rect.height;return 0}_emscripten_get_element_css_size.sig="ippp";var _emscripten_html5_remove_all_event_listeners=()=>JSEvents.removeAllEventListeners();_emscripten_html5_remove_all_event_listeners.sig="v";var _emscripten_request_animation_frame=function(cb,userData){cb>>>=0;userData>>>=0;return requestAnimationFrame(timeStamp=>getWasmTableEntry(cb)(timeStamp,userData))};_emscripten_request_animation_frame.sig="ipp";var _emscripten_cancel_animation_frame=id=>cancelAnimationFrame(id);_emscripten_cancel_animation_frame.sig="vi";function _emscripten_request_animation_frame_loop(cb,userData){cb>>>=0;userData>>>=0;function tick(timeStamp){if(getWasmTableEntry(cb)(timeStamp,userData)){requestAnimationFrame(tick)}}return requestAnimationFrame(tick)}_emscripten_request_animation_frame_loop.sig="vpp";var _emscripten_get_device_pixel_ratio=()=>typeof devicePixelRatio=="number"&&devicePixelRatio||1;_emscripten_get_device_pixel_ratio.sig="d";function _emscripten_get_callstack(flags,str,maxbytes){str>>>=0;var callstack=getCallstack(flags);if(!str||maxbytes<=0){return lengthBytesUTF8(callstack)+1}var bytesWrittenExcludingNull=stringToUTF8(callstack,str,maxbytes);return bytesWrittenExcludingNull+1}_emscripten_get_callstack.sig="iipi";var convertFrameToPC=frame=>{abort("Cannot use convertFrameToPC (needed by __builtin_return_address) without -sUSE_OFFSET_CONVERTER");return 0};function _emscripten_return_address(level){var callstack=jsStackTrace().split("\n");if(callstack[0]=="Error"){callstack.shift()}var caller=callstack[level+3];return convertFrameToPC(caller)}_emscripten_return_address.sig="pi";var UNWIND_CACHE={};var saveInUnwindCache=callstack=>{callstack.forEach(frame=>{var pc=convertFrameToPC(frame);if(pc){UNWIND_CACHE[pc]=frame}})};function _emscripten_stack_snapshot(){var callstack=jsStackTrace().split("\n");if(callstack[0]=="Error"){callstack.shift()}saveInUnwindCache(callstack);UNWIND_CACHE.last_addr=convertFrameToPC(callstack[3]);UNWIND_CACHE.last_stack=callstack;return UNWIND_CACHE.last_addr}_emscripten_stack_snapshot.sig="p";function _emscripten_stack_unwind_buffer(addr,buffer,count){addr>>>=0;buffer>>>=0;var stack;if(UNWIND_CACHE.last_addr==addr){stack=UNWIND_CACHE.last_stack}else{stack=jsStackTrace().split("\n");if(stack[0]=="Error"){stack.shift()}saveInUnwindCache(stack)}var offset=3;while(stack[offset]&&convertFrameToPC(stack[offset])!=addr){++offset}for(var i=0;i<count&&stack[i+offset];++i){HEAP32[buffer+i*4>>>2>>>0]=convertFrameToPC(stack[i+offset])}return i}_emscripten_stack_unwind_buffer.sig="ippi";function _emscripten_pc_get_function(pc){pc>>>=0;abort("Cannot use emscripten_pc_get_function without -sUSE_OFFSET_CONVERTER");return 0}_emscripten_pc_get_function.sig="pp";var convertPCtoSourceLocation=pc=>{if(UNWIND_CACHE.last_get_source_pc==pc)return UNWIND_CACHE.last_source;var match;var source;if(!source){var frame=UNWIND_CACHE[pc];if(!frame)return null;if(match=/\((.*):(\d+):(\d+)\)$/.exec(frame)){source={file:match[1],line:match[2],column:match[3]}}else if(match=/@(.*):(\d+):(\d+)/.exec(frame)){source={file:match[1],line:match[2],column:match[3]}}}UNWIND_CACHE.last_get_source_pc=pc;UNWIND_CACHE.last_source=source;return source};function _emscripten_pc_get_file(pc){pc>>>=0;var result=convertPCtoSourceLocation(pc);if(!result)return 0;if(_emscripten_pc_get_file.ret)_free(_emscripten_pc_get_file.ret);_emscripten_pc_get_file.ret=stringToNewUTF8(result.file);return _emscripten_pc_get_file.ret}_emscripten_pc_get_file.sig="pp";function _emscripten_pc_get_line(pc){pc>>>=0;var result=convertPCtoSourceLocation(pc);return result?result.line:0}_emscripten_pc_get_line.sig="ip";function _emscripten_pc_get_column(pc){pc>>>=0;var result=convertPCtoSourceLocation(pc);return result?result.column||0:0}_emscripten_pc_get_column.sig="ip";var wasiRightsToMuslOFlags=rights=>{if(rights&2&&rights&64){return 2}if(rights&2){return 0}if(rights&64){return 1}throw new FS.ErrnoError(28)};var wasiOFlagsToMuslOFlags=oflags=>{var musl_oflags=0;if(oflags&1){musl_oflags|=64}if(oflags&8){musl_oflags|=512}if(oflags&2){musl_oflags|=65536}if(oflags&4){musl_oflags|=128}return musl_oflags};var _emscripten_unwind_to_js_event_loop=()=>{throw"unwind"};_emscripten_unwind_to_js_event_loop.sig="v";var safeSetTimeout=(func,timeout)=>{runtimeKeepalivePush();return setTimeout(()=>{runtimeKeepalivePop();callUserCallback(func)},timeout)};var setImmediateWrapped=func=>{setImmediateWrapped.mapping||=[];var id=setImmediateWrapped.mapping.length;setImmediateWrapped.mapping[id]=setImmediate(()=>{setImmediateWrapped.mapping[id]=undefined;func()});return id};var _emscripten_set_main_loop_timing=(mode,value)=>{MainLoop.timingMode=mode;MainLoop.timingValue=value;if(!MainLoop.func){return 1}if(!MainLoop.running){runtimeKeepalivePush();MainLoop.running=true}if(mode==0){MainLoop.scheduler=function MainLoop_scheduler_setTimeout(){var timeUntilNextTick=Math.max(0,MainLoop.tickStartTime+value-_emscripten_get_now())|0;setTimeout(MainLoop.runner,timeUntilNextTick)};MainLoop.method="timeout"}else if(mode==1){MainLoop.scheduler=function MainLoop_scheduler_rAF(){MainLoop.requestAnimationFrame(MainLoop.runner)};MainLoop.method="rAF"}else if(mode==2){if(typeof MainLoop.setImmediate=="undefined"){if(typeof setImmediate=="undefined"){var setImmediates=[];var emscriptenMainLoopMessageId="setimmediate";var MainLoop_setImmediate_messageHandler=event=>{if(event.data===emscriptenMainLoopMessageId||event.data.target===emscriptenMainLoopMessageId){event.stopPropagation();setImmediates.shift()()}};addEventListener("message",MainLoop_setImmediate_messageHandler,true);MainLoop.setImmediate=func=>{setImmediates.push(func);if(ENVIRONMENT_IS_WORKER){Module["setImmediates"]??=[];Module["setImmediates"].push(func);postMessage({target:emscriptenMainLoopMessageId})}else postMessage(emscriptenMainLoopMessageId,"*")}}else{MainLoop.setImmediate=setImmediate}}MainLoop.scheduler=function MainLoop_scheduler_setImmediate(){MainLoop.setImmediate(MainLoop.runner)};MainLoop.method="immediate"}return 0};_emscripten_set_main_loop_timing.sig="iii";var setMainLoop=(iterFunc,fps,simulateInfiniteLoop,arg,noSetTiming)=>{MainLoop.func=iterFunc;MainLoop.arg=arg;var thisMainLoopId=MainLoop.currentlyRunningMainloop;function checkIsRunning(){if(thisMainLoopId<MainLoop.currentlyRunningMainloop){runtimeKeepalivePop();maybeExit();return false}return true}MainLoop.running=false;MainLoop.runner=function MainLoop_runner(){if(ABORT)return;if(MainLoop.queue.length>0){var start=Date.now();var blocker=MainLoop.queue.shift();blocker.func(blocker.arg);if(MainLoop.remainingBlockers){var remaining=MainLoop.remainingBlockers;var next=remaining%1==0?remaining-1:Math.floor(remaining);if(blocker.counted){MainLoop.remainingBlockers=next}else{next=next+.5;MainLoop.remainingBlockers=(8*remaining+next)/9}}MainLoop.updateStatus();if(!checkIsRunning())return;setTimeout(MainLoop.runner,0);return}if(!checkIsRunning())return;MainLoop.currentFrameNumber=MainLoop.currentFrameNumber+1|0;if(MainLoop.timingMode==1&&MainLoop.timingValue>1&&MainLoop.currentFrameNumber%MainLoop.timingValue!=0){MainLoop.scheduler();return}else if(MainLoop.timingMode==0){MainLoop.tickStartTime=_emscripten_get_now()}MainLoop.runIter(iterFunc);if(!checkIsRunning())return;MainLoop.scheduler()};if(!noSetTiming){if(fps>0){_emscripten_set_main_loop_timing(0,1e3/fps)}else{_emscripten_set_main_loop_timing(1,1)}MainLoop.scheduler()}if(simulateInfiniteLoop){throw"unwind"}};var MainLoop={running:false,scheduler:null,method:"",currentlyRunningMainloop:0,func:null,arg:0,timingMode:0,timingValue:0,currentFrameNumber:0,queue:[],preMainLoop:[],postMainLoop:[],pause(){MainLoop.scheduler=null;MainLoop.currentlyRunningMainloop++},resume(){MainLoop.currentlyRunningMainloop++;var timingMode=MainLoop.timingMode;var timingValue=MainLoop.timingValue;var func=MainLoop.func;MainLoop.func=null;setMainLoop(func,0,false,MainLoop.arg,true);_emscripten_set_main_loop_timing(timingMode,timingValue);MainLoop.scheduler()},updateStatus(){if(Module["setStatus"]){var message=Module["statusMessage"]||"Please wait...";var remaining=MainLoop.remainingBlockers??0;var expected=MainLoop.expectedBlockers??0;if(remaining){if(remaining<expected){Module["setStatus"](`{message} ({expected - remaining}/{expected})`)}else{Module["setStatus"](message)}}else{Module["setStatus"]("")}}},init(){Module["preMainLoop"]&&MainLoop.preMainLoop.push(Module["preMainLoop"]);Module["postMainLoop"]&&MainLoop.postMainLoop.push(Module["postMainLoop"])},runIter(func){if(ABORT)return;for(var pre of MainLoop.preMainLoop){if(pre()===false){return}}callUserCallback(func);for(var post of MainLoop.postMainLoop){post()}},nextRAF:0,fakeRequestAnimationFrame(func){var now=Date.now();if(MainLoop.nextRAF===0){MainLoop.nextRAF=now+1e3/60}else{while(now+2>=MainLoop.nextRAF){MainLoop.nextRAF+=1e3/60}}var delay=Math.max(MainLoop.nextRAF-now,0);setTimeout(func,delay)},requestAnimationFrame(func){if(typeof requestAnimationFrame=="function"){requestAnimationFrame(func);return}var RAF=MainLoop.fakeRequestAnimationFrame;RAF(func)}};var safeRequestAnimationFrame=func=>{runtimeKeepalivePush();return MainLoop.requestAnimationFrame(()=>{runtimeKeepalivePop();callUserCallback(func)})};var clearImmediateWrapped=id=>{clearImmediate(setImmediateWrapped.mapping[id]);setImmediateWrapped.mapping[id]=undefined};var emClearImmediate;var emSetImmediate;var emClearImmediate_deps=["$emSetImmediate"];var _emscripten_set_immediate=function(cb,userData){cb>>>=0;userData>>>=0;runtimeKeepalivePush();return emSetImmediate(()=>{runtimeKeepalivePop();callUserCallback(()=>getWasmTableEntry(cb)(userData))})};_emscripten_set_immediate.sig="ipp";var _emscripten_clear_immediate=id=>{runtimeKeepalivePop();emClearImmediate(id)};_emscripten_clear_immediate.sig="vi";var _emscripten_set_immediate_loop=function(cb,userData){cb>>>=0;userData>>>=0;function tick(){callUserCallback(()=>{if(getWasmTableEntry(cb)(userData)){emSetImmediate(tick)}else{runtimeKeepalivePop()}})}runtimeKeepalivePush();emSetImmediate(tick)};_emscripten_set_immediate_loop.sig="vpp";var _emscripten_set_timeout=function(cb,msecs,userData){cb>>>=0;userData>>>=0;return safeSetTimeout(()=>getWasmTableEntry(cb)(userData),msecs)};_emscripten_set_timeout.sig="ipdp";var _emscripten_clear_timeout=clearTimeout;_emscripten_clear_timeout.sig="vi";var _emscripten_set_timeout_loop=function(cb,msecs,userData){cb>>>=0;userData>>>=0;function tick(){var t=_emscripten_get_now();var n=t+msecs;runtimeKeepalivePop();callUserCallback(()=>{if(getWasmTableEntry(cb)(t,userData)){runtimeKeepalivePush();var remaining=n-_emscripten_get_now();remaining=Math.max(0,remaining);setTimeout(tick,remaining)}})}runtimeKeepalivePush();return setTimeout(tick,0)};_emscripten_set_timeout_loop.sig="vpdp";var _emscripten_set_interval=function(cb,msecs,userData){cb>>>=0;userData>>>=0;runtimeKeepalivePush();return setInterval(()=>{callUserCallback(()=>getWasmTableEntry(cb)(userData))},msecs)};_emscripten_set_interval.sig="ipdp";var _emscripten_clear_interval=id=>{runtimeKeepalivePop();clearInterval(id)};_emscripten_clear_interval.sig="vi";var _emscripten_async_call=function(func,arg,millis){func>>>=0;arg>>>=0;var wrapper=()=>getWasmTableEntry(func)(arg);if(millis>=0||ENVIRONMENT_IS_NODE){safeSetTimeout(wrapper,millis)}else{safeRequestAnimationFrame(wrapper)}};_emscripten_async_call.sig="vppi";var registerPostMainLoop=f=>{typeof MainLoop!="undefined"&&MainLoop.postMainLoop.push(f)};var registerPreMainLoop=f=>{typeof MainLoop!="undefined"&&MainLoop.preMainLoop.push(f)};function _emscripten_get_main_loop_timing(mode,value){mode>>>=0;value>>>=0;if(mode)HEAP32[mode>>>2>>>0]=MainLoop.timingMode;if(value)HEAP32[value>>>2>>>0]=MainLoop.timingValue}_emscripten_get_main_loop_timing.sig="vpp";function _emscripten_set_main_loop(func,fps,simulateInfiniteLoop){func>>>=0;var iterFunc=getWasmTableEntry(func);setMainLoop(iterFunc,fps,simulateInfiniteLoop)}_emscripten_set_main_loop.sig="vpii";var _emscripten_set_main_loop_arg=function(func,arg,fps,simulateInfiniteLoop){func>>>=0;arg>>>=0;var iterFunc=()=>getWasmTableEntry(func)(arg);setMainLoop(iterFunc,fps,simulateInfiniteLoop,arg)};_emscripten_set_main_loop_arg.sig="vppii";var _emscripten_cancel_main_loop=()=>{MainLoop.pause();MainLoop.func=null};_emscripten_cancel_main_loop.sig="v";var _emscripten_pause_main_loop=()=>MainLoop.pause();_emscripten_pause_main_loop.sig="v";var _emscripten_resume_main_loop=()=>MainLoop.resume();_emscripten_resume_main_loop.sig="v";var __emscripten_push_main_loop_blocker=function(func,arg,name){func>>>=0;arg>>>=0;name>>>=0;MainLoop.queue.push({func:()=>{getWasmTableEntry(func)(arg)},name:UTF8ToString(name),counted:true});MainLoop.updateStatus()};__emscripten_push_main_loop_blocker.sig="vppp";var __emscripten_push_uncounted_main_loop_blocker=function(func,arg,name){func>>>=0;arg>>>=0;name>>>=0;MainLoop.queue.push({func:()=>{getWasmTableEntry(func)(arg)},name:UTF8ToString(name),counted:false});MainLoop.updateStatus()};__emscripten_push_uncounted_main_loop_blocker.sig="vppp";var _emscripten_set_main_loop_expected_blockers=num=>{MainLoop.expectedBlockers=num;MainLoop.remainingBlockers=num;MainLoop.updateStatus()};_emscripten_set_main_loop_expected_blockers.sig="vi";var idsToPromises=(idBuf,size)=>{var promises=[];for(var i=0;i<size;i++){var id=HEAP32[idBuf+i*4>>>2>>>0];promises[i]=getPromise(id)}return promises};var makePromiseCallback=(callback,userData)=>value=>{runtimeKeepalivePop();var stack=stackSave();var resultPtr=stackAlloc(POINTER_SIZE);HEAPU32[resultPtr>>>2>>>0]=0;try{var result=getWasmTableEntry(callback)(resultPtr,userData,value);var resultVal=HEAPU32[resultPtr>>>2>>>0]}catch(e){if(typeof e!="number"){throw 0}throw e}finally{stackRestore(stack)}switch(result){case 0:return resultVal;case 1:return getPromise(resultVal);case 2:var ret=getPromise(resultVal);_emscripten_promise_destroy(resultVal);return ret;case 3:throw resultVal}};function _emscripten_promise_then(id,onFulfilled,onRejected,userData){id>>>=0;onFulfilled>>>=0;onRejected>>>=0;userData>>>=0;runtimeKeepalivePush();var promise=getPromise(id);var newId=promiseMap.allocate({promise:promise.then(makePromiseCallback(onFulfilled,userData),makePromiseCallback(onRejected,userData))});return newId}_emscripten_promise_then.sig="ppppp";var _emscripten_promise_all=function(idBuf,resultBuf,size){idBuf>>>=0;resultBuf>>>=0;size>>>=0;var promises=idsToPromises(idBuf,size);var id=promiseMap.allocate({promise:Promise.all(promises).then(results=>{if(resultBuf){for(var i=0;i<size;i++){var result=results[i];HEAPU32[resultBuf+i*4>>>2>>>0]=result}}return resultBuf})});return id};_emscripten_promise_all.sig="pppp";var setPromiseResult=(ptr,fulfill,value)=>{var result=fulfill?0:3;HEAP32[ptr>>>2>>>0]=result;HEAPU32[ptr+4>>>2>>>0]=value};var _emscripten_promise_all_settled=function(idBuf,resultBuf,size){idBuf>>>=0;resultBuf>>>=0;size>>>=0;var promises=idsToPromises(idBuf,size);var id=promiseMap.allocate({promise:Promise.allSettled(promises).then(results=>{if(resultBuf){var offset=resultBuf;for(var i=0;i<size;i++,offset+=8){if(results[i].status==="fulfilled"){setPromiseResult(offset,true,results[i].value)}else{setPromiseResult(offset,false,results[i].reason)}}}return resultBuf})});return id};_emscripten_promise_all_settled.sig="pppp";var _emscripten_promise_any=function(idBuf,errorBuf,size){idBuf>>>=0;errorBuf>>>=0;size>>>=0;var promises=idsToPromises(idBuf,size);var id=promiseMap.allocate({promise:Promise.any(promises).catch(err=>{if(errorBuf){for(var i=0;i<size;i++){HEAPU32[errorBuf+i*4>>>2>>>0]=err.errors[i]}}throw errorBuf})});return id};_emscripten_promise_any.sig="pppp";function _emscripten_promise_race(idBuf,size){idBuf>>>=0;size>>>=0;var promises=idsToPromises(idBuf,size);var id=promiseMap.allocate({promise:Promise.race(promises)});return id}_emscripten_promise_race.sig="ppp";function _emscripten_promise_await(returnValuePtr,id){returnValuePtr>>>=0;id>>>=0;abort("emscripten_promise_await is only available with ASYNCIFY")}_emscripten_promise_await.sig="vpp";var getExceptionMessageCommon=ptr=>{var sp=stackSave();var type_addr_addr=stackAlloc(4);var message_addr_addr=stackAlloc(4);___get_exception_message(ptr,type_addr_addr,message_addr_addr);var type_addr=HEAPU32[type_addr_addr>>>2>>>0];var message_addr=HEAPU32[message_addr_addr>>>2>>>0];var type=UTF8ToString(type_addr);_free(type_addr);var message;if(message_addr){message=UTF8ToString(message_addr);_free(message_addr)}stackRestore(sp);return[type,message]};var getCppExceptionTag=()=>___cpp_exception;var getCppExceptionThrownObjectFromWebAssemblyException=ex=>{var unwind_header=ex.getArg(getCppExceptionTag(),0);return ___thrown_object_from_unwind_exception(unwind_header)};var incrementExceptionRefcount=ex=>{var ptr=getCppExceptionThrownObjectFromWebAssemblyException(ex);___cxa_increment_exception_refcount(ptr)};var decrementExceptionRefcount=ex=>{var ptr=getCppExceptionThrownObjectFromWebAssemblyException(ex);___cxa_decrement_exception_refcount(ptr)};var getExceptionMessage=ex=>{var ptr=getCppExceptionThrownObjectFromWebAssemblyException(ex);return getExceptionMessageCommon(ptr)};var Browser={useWebGL:false,isFullscreen:false,pointerLock:false,moduleContextCreatedCallbacks:[],workers:[],preloadedImages:{},preloadedAudios:{},getCanvas:()=>Module["canvas"],init(){if(Browser.initted)return;Browser.initted=true;var imagePlugin={};imagePlugin["canHandle"]=function imagePlugin_canHandle(name){return!Module["noImageDecoding"]&&/\.(jpg|jpeg|png|bmp|webp)$/i.test(name)};imagePlugin["handle"]=function imagePlugin_handle(byteArray,name,onload,onerror){var b=new Blob([byteArray],{type:Browser.getMimetype(name)});if(b.size!==byteArray.length){b=new Blob([new Uint8Array(byteArray).buffer],{type:Browser.getMimetype(name)})}var url=URL.createObjectURL(b);var img=new Image;img.onload=()=>{var canvas=document.createElement("canvas");canvas.width=img.width;canvas.height=img.height;var ctx=canvas.getContext("2d");ctx.drawImage(img,0,0);Browser.preloadedImages[name]=canvas;URL.revokeObjectURL(url);onload?.(byteArray)};img.onerror=event=>{err(`Image ${url} could not be decoded`);onerror?.()};img.src=url};preloadPlugins.push(imagePlugin);var audioPlugin={};audioPlugin["canHandle"]=function audioPlugin_canHandle(name){return!Module["noAudioDecoding"]&&name.slice(-4)in{".ogg":1,".wav":1,".mp3":1}};audioPlugin["handle"]=function audioPlugin_handle(byteArray,name,onload,onerror){var done=false;function finish(audio){if(done)return;done=true;Browser.preloadedAudios[name]=audio;onload?.(byteArray)}function fail(){if(done)return;done=true;Browser.preloadedAudios[name]=new Audio;onerror?.()}var b=new Blob([byteArray],{type:Browser.getMimetype(name)});var url=URL.createObjectURL(b);var audio=new Audio;audio.addEventListener("canplaythrough",()=>finish(audio),false);audio.onerror=function audio_onerror(event){if(done)return;err(`warning: browser could not fully decode audio ${name}, trying slower base64 approach`);function encode64(data){var BASE="ABCDEFGHIJKLMNOPQRSTUVWXYZabcdefghijklmnopqrstuvwxyz0123456789+/";var PAD="=";var ret="";var leftchar=0;var leftbits=0;for(var i=0;i<data.length;i++){leftchar=leftchar<<8|data[i];leftbits+=8;while(leftbits>=6){var curr=leftchar>>leftbits-6&63;leftbits-=6;ret+=BASE[curr]}}if(leftbits==2){ret+=BASE[(leftchar&3)<<4];ret+=PAD+PAD}else if(leftbits==4){ret+=BASE[(leftchar&15)<<2];ret+=PAD}return ret}audio.src="data:audio/x-"+name.slice(-3)+";base64,"+encode64(byteArray);finish(audio)};audio.src=url;safeSetTimeout(()=>{finish(audio)},1e4)};preloadPlugins.push(audioPlugin);function pointerLockChange(){var canvas=Browser.getCanvas();Browser.pointerLock=document["pointerLockElement"]===canvas||document["mozPointerLockElement"]===canvas||document["webkitPointerLockElement"]===canvas||document["msPointerLockElement"]===canvas}var canvas=Browser.getCanvas();if(canvas){canvas.requestPointerLock=canvas["requestPointerLock"]||canvas["mozRequestPointerLock"]||canvas["webkitRequestPointerLock"]||canvas["msRequestPointerLock"]||(()=>{});canvas.exitPointerLock=document["exitPointerLock"]||document["mozExitPointerLock"]||document["webkitExitPointerLock"]||document["msExitPointerLock"]||(()=>{});canvas.exitPointerLock=canvas.exitPointerLock.bind(document);document.addEventListener("pointerlockchange",pointerLockChange,false);document.addEventListener("mozpointerlockchange",pointerLockChange,false);document.addEventListener("webkitpointerlockchange",pointerLockChange,false);document.addEventListener("mspointerlockchange",pointerLockChange,false);if(Module["elementPointerLock"]){canvas.addEventListener("click",ev=>{if(!Browser.pointerLock&&Browser.getCanvas().requestPointerLock){Browser.getCanvas().requestPointerLock();ev.preventDefault()}},false)}}},createContext(canvas,useWebGL,setInModule,webGLContextAttributes){if(useWebGL&&Module["ctx"]&&canvas==Browser.getCanvas())return Module["ctx"];var ctx;var contextHandle;if(useWebGL){var contextAttributes={antialias:false,alpha:false,majorVersion:typeof WebGL2RenderingContext!="undefined"?2:1};if(webGLContextAttributes){for(var attribute in webGLContextAttributes){contextAttributes[attribute]=webGLContextAttributes[attribute]}}if(typeof GL!="undefined"){contextHandle=GL.createContext(canvas,contextAttributes);if(contextHandle){ctx=GL.getContext(contextHandle).GLctx}}}else{ctx=canvas.getContext("2d")}if(!ctx)return null;if(setInModule){Module["ctx"]=ctx;if(useWebGL)GL.makeContextCurrent(contextHandle);Browser.useWebGL=useWebGL;Browser.moduleContextCreatedCallbacks.forEach(callback=>callback());Browser.init()}return ctx},fullscreenHandlersInstalled:false,lockPointer:undefined,resizeCanvas:undefined,requestFullscreen(lockPointer,resizeCanvas){Browser.lockPointer=lockPointer;Browser.resizeCanvas=resizeCanvas;if(typeof Browser.lockPointer=="undefined")Browser.lockPointer=true;if(typeof Browser.resizeCanvas=="undefined")Browser.resizeCanvas=false;var canvas=Browser.getCanvas();function fullscreenChange(){Browser.isFullscreen=false;var canvasContainer=canvas.parentNode;if((document["fullscreenElement"]||document["mozFullScreenElement"]||document["msFullscreenElement"]||document["webkitFullscreenElement"]||document["webkitCurrentFullScreenElement"])===canvasContainer){canvas.exitFullscreen=Browser.exitFullscreen;if(Browser.lockPointer)canvas.requestPointerLock();Browser.isFullscreen=true;if(Browser.resizeCanvas){Browser.setFullscreenCanvasSize()}else{Browser.updateCanvasDimensions(canvas)}}else{canvasContainer.parentNode.insertBefore(canvas,canvasContainer);canvasContainer.parentNode.removeChild(canvasContainer);if(Browser.resizeCanvas){Browser.setWindowedCanvasSize()}else{Browser.updateCanvasDimensions(canvas)}}Module["onFullScreen"]?.(Browser.isFullscreen);Module["onFullscreen"]?.(Browser.isFullscreen)}if(!Browser.fullscreenHandlersInstalled){Browser.fullscreenHandlersInstalled=true;document.addEventListener("fullscreenchange",fullscreenChange,false);document.addEventListener("mozfullscreenchange",fullscreenChange,false);document.addEventListener("webkitfullscreenchange",fullscreenChange,false);document.addEventListener("MSFullscreenChange",fullscreenChange,false)}var canvasContainer=document.createElement("div");canvas.parentNode.insertBefore(canvasContainer,canvas);canvasContainer.appendChild(canvas);canvasContainer.requestFullscreen=canvasContainer["requestFullscreen"]||canvasContainer["mozRequestFullScreen"]||canvasContainer["msRequestFullscreen"]||(canvasContainer["webkitRequestFullscreen"]?()=>canvasContainer["webkitRequestFullscreen"](Element["ALLOW_KEYBOARD_INPUT"]):null)||(canvasContainer["webkitRequestFullScreen"]?()=>canvasContainer["webkitRequestFullScreen"](Element["ALLOW_KEYBOARD_INPUT"]):null);canvasContainer.requestFullscreen()},exitFullscreen(){if(!Browser.isFullscreen){return false}var CFS=document["exitFullscreen"]||document["cancelFullScreen"]||document["mozCancelFullScreen"]||document["msExitFullscreen"]||document["webkitCancelFullScreen"]||(()=>{});CFS.apply(document,[]);return true},safeSetTimeout(func,timeout){return safeSetTimeout(func,timeout)},getMimetype(name){return{jpg:"image/jpeg",jpeg:"image/jpeg",png:"image/png",bmp:"image/bmp",ogg:"audio/ogg",wav:"audio/wav",mp3:"audio/mpeg"}[name.slice(name.lastIndexOf(".")+1)]},getUserMedia(func){window.getUserMedia||=navigator["getUserMedia"]||navigator["mozGetUserMedia"];window.getUserMedia(func)},getMovementX(event){return event["movementX"]||event["mozMovementX"]||event["webkitMovementX"]||0},getMovementY(event){return event["movementY"]||event["mozMovementY"]||event["webkitMovementY"]||0},getMouseWheelDelta(event){var delta=0;switch(event.type){case"DOMMouseScroll":delta=event.detail/3;break;case"mousewheel":delta=event.wheelDelta/120;break;case"wheel":delta=event.deltaY;switch(event.deltaMode){case 0:delta/=100;break;case 1:delta/=3;break;case 2:delta*=80;break;default:throw"unrecognized mouse wheel delta mode: "+event.deltaMode}break;default:throw"unrecognized mouse wheel event: "+event.type}return delta},mouseX:0,mouseY:0,mouseMovementX:0,mouseMovementY:0,touches:{},lastTouches:{},calculateMouseCoords(pageX,pageY){var canvas=Browser.getCanvas();var rect=canvas.getBoundingClientRect();var scrollX=typeof window.scrollX!="undefined"?window.scrollX:window.pageXOffset;var scrollY=typeof window.scrollY!="undefined"?window.scrollY:window.pageYOffset;var adjustedX=pageX-(scrollX+rect.left);var adjustedY=pageY-(scrollY+rect.top);adjustedX=adjustedX*(canvas.width/rect.width);adjustedY=adjustedY*(canvas.height/rect.height);return{x:adjustedX,y:adjustedY}},setMouseCoords(pageX,pageY){const{x,y}=Browser.calculateMouseCoords(pageX,pageY);Browser.mouseMovementX=x-Browser.mouseX;Browser.mouseMovementY=y-Browser.mouseY;Browser.mouseX=x;Browser.mouseY=y},calculateMouseEvent(event){if(Browser.pointerLock){if(event.type!="mousemove"&&"mozMovementX"in event){Browser.mouseMovementX=Browser.mouseMovementY=0}else{Browser.mouseMovementX=Browser.getMovementX(event);Browser.mouseMovementY=Browser.getMovementY(event)}Browser.mouseX+=Browser.mouseMovementX;Browser.mouseY+=Browser.mouseMovementY}else{if(event.type==="touchstart"||event.type==="touchend"||event.type==="touchmove"){var touch=event.touch;if(touch===undefined){return}var coords=Browser.calculateMouseCoords(touch.pageX,touch.pageY);if(event.type==="touchstart"){Browser.lastTouches[touch.identifier]=coords;Browser.touches[touch.identifier]=coords}else if(event.type==="touchend"||event.type==="touchmove"){var last=Browser.touches[touch.identifier];last||=coords;Browser.lastTouches[touch.identifier]=last;Browser.touches[touch.identifier]=coords}return}Browser.setMouseCoords(event.pageX,event.pageY)}},resizeListeners:[],updateResizeListeners(){var canvas=Browser.getCanvas();Browser.resizeListeners.forEach(listener=>listener(canvas.width,canvas.height))},setCanvasSize(width,height,noUpdates){var canvas=Browser.getCanvas();Browser.updateCanvasDimensions(canvas,width,height);if(!noUpdates)Browser.updateResizeListeners()},windowedWidth:0,windowedHeight:0,setFullscreenCanvasSize(){if(typeof SDL!="undefined"){var flags=HEAPU32[SDL.screen>>>2>>>0];flags=flags|8388608;HEAP32[SDL.screen>>>2>>>0]=flags}Browser.updateCanvasDimensions(Browser.getCanvas());Browser.updateResizeListeners()},setWindowedCanvasSize(){if(typeof SDL!="undefined"){var flags=HEAPU32[SDL.screen>>>2>>>0];flags=flags&~8388608;HEAP32[SDL.screen>>>2>>>0]=flags}Browser.updateCanvasDimensions(Browser.getCanvas());Browser.updateResizeListeners()},updateCanvasDimensions(canvas,wNative,hNative){if(wNative&&hNative){canvas.widthNative=wNative;canvas.heightNative=hNative}else{wNative=canvas.widthNative;hNative=canvas.heightNative}var w=wNative;var h=hNative;if(Module["forcedAspectRatio"]>0){if(w/h<Module["forcedAspectRatio"]){w=Math.round(h*Module["forcedAspectRatio"])}else{h=Math.round(w/Module["forcedAspectRatio"])}}if((document["fullscreenElement"]||document["mozFullScreenElement"]||document["msFullscreenElement"]||document["webkitFullscreenElement"]||document["webkitCurrentFullScreenElement"])===canvas.parentNode&&typeof screen!="undefined"){var factor=Math.min(screen.width/w,screen.height/h);w=Math.round(w*factor);h=Math.round(h*factor)}if(Browser.resizeCanvas){if(canvas.width!=w)canvas.width=w;if(canvas.height!=h)canvas.height=h;if(typeof canvas.style!="undefined"){canvas.style.removeProperty("width");canvas.style.removeProperty("height")}}else{if(canvas.width!=wNative)canvas.width=wNative;if(canvas.height!=hNative)canvas.height=hNative;if(typeof canvas.style!="undefined"){if(w!=wNative||h!=hNative){canvas.style.setProperty("width",w+"px","important");canvas.style.setProperty("height",h+"px","important")}else{canvas.style.removeProperty("width");canvas.style.removeProperty("height")}}}}};var requestFullscreen=Browser.requestFullscreen;var setCanvasSize=Browser.setCanvasSize;var getUserMedia=Browser.getUserMedia;var createContext=Browser.createContext;var _emscripten_run_preload_plugins=function(file,onload,onerror){file>>>=0;onload>>>=0;onerror>>>=0;runtimeKeepalivePush();var _file=UTF8ToString(file);var data=FS.analyzePath(_file);if(!data.exists)return-1;FS.createPreloadedFile(PATH.dirname(_file),PATH.basename(_file),new Uint8Array(data.object.contents),true,true,()=>{runtimeKeepalivePop();if(onload)getWasmTableEntry(onload)(file)},()=>{runtimeKeepalivePop();if(onerror)getWasmTableEntry(onerror)(file)},true);return 0};_emscripten_run_preload_plugins.sig="ippp";var Browser_asyncPrepareDataCounter=0;var _emscripten_run_preload_plugins_data=function(data,size,suffix,arg,onload,onerror){data>>>=0;suffix>>>=0;arg>>>=0;onload>>>=0;onerror>>>=0;runtimeKeepalivePush();var _suffix=UTF8ToString(suffix);var name="prepare_data_"+Browser_asyncPrepareDataCounter+++"."+_suffix;var cname=stringToNewUTF8(name);FS.createPreloadedFile("/",name,HEAPU8.subarray(data>>>0,data+size>>>0),true,true,()=>{runtimeKeepalivePop();if(onload)getWasmTableEntry(onload)(arg,cname)},()=>{runtimeKeepalivePop();if(onerror)getWasmTableEntry(onerror)(arg)},true)};_emscripten_run_preload_plugins_data.sig="vpipppp";var _emscripten_async_run_script=function(script,millis){script>>>=0;safeSetTimeout(()=>_emscripten_run_script(script),millis)};_emscripten_async_run_script.sig="vpi";var _emscripten_async_load_script=async function(url,onload,onerror){url>>>=0;onload>>>=0;onerror>>>=0;url=UTF8ToString(url);runtimeKeepalivePush();var loadDone=()=>{runtimeKeepalivePop();if(onload){var onloadCallback=()=>callUserCallback(getWasmTableEntry(onload));if(runDependencies>0){dependenciesFulfilled=onloadCallback}else{onloadCallback()}}};var loadError=()=>{runtimeKeepalivePop();if(onerror){callUserCallback(getWasmTableEntry(onerror))}};if(ENVIRONMENT_IS_NODE){try{var data=await readAsync(url,false);eval(data);loadDone()}catch(e){err(e);loadError()}return}var script=document.createElement("script");script.onload=loadDone;script.onerror=loadError;script.src=url;document.body.appendChild(script)};_emscripten_async_load_script.sig="vppp";function _emscripten_get_window_title(){var buflen=256;if(!_emscripten_get_window_title.buffer){_emscripten_get_window_title.buffer=_malloc(buflen)}stringToUTF8(document.title,_emscripten_get_window_title.buffer,buflen);return _emscripten_get_window_title.buffer}_emscripten_get_window_title.sig="p";function _emscripten_set_window_title(title){title>>>=0;return document.title=UTF8ToString(title)}_emscripten_set_window_title.sig="vp";function _emscripten_get_screen_size(width,height){width>>>=0;height>>>=0;HEAP32[width>>>2>>>0]=screen.width;HEAP32[height>>>2>>>0]=screen.height}_emscripten_get_screen_size.sig="vpp";var _emscripten_hide_mouse=()=>{var styleSheet=document.styleSheets[0];var rules=styleSheet.cssRules;for(var i=0;i<rules.length;i++){if(rules[i].cssText.startsWith("canvas")){styleSheet.deleteRule(i);i--}}styleSheet.insertRule("canvas.emscripten { border: 1px solid black; cursor: none; }",0)};_emscripten_hide_mouse.sig="v";var _emscripten_set_canvas_size=(width,height)=>Browser.setCanvasSize(width,height);_emscripten_set_canvas_size.sig="vii";function _emscripten_get_canvas_size(width,height,isFullscreen){width>>>=0;height>>>=0;isFullscreen>>>=0;var canvas=Browser.getCanvas();HEAP32[width>>>2>>>0]=canvas.width;HEAP32[height>>>2>>>0]=canvas.height;HEAP32[isFullscreen>>>2>>>0]=Browser.isFullscreen?1:0}_emscripten_get_canvas_size.sig="vppp";function _emscripten_create_worker(url){url>>>=0;url=UTF8ToString(url);var id=Browser.workers.length;var info={worker:new Worker(url),callbacks:[],awaited:0,buffer:0,bufferSize:0};info.worker.onmessage=function info_worker_onmessage(msg){if(ABORT)return;var info=Browser.workers[id];if(!info)return;var callbackId=msg.data["callbackId"];var callbackInfo=info.callbacks[callbackId];if(!callbackInfo)return;if(msg.data["finalResponse"]){info.awaited--;info.callbacks[callbackId]=null;runtimeKeepalivePop()}var data=msg.data["data"];if(data){if(!data.byteLength)data=new Uint8Array(data);if(!info.buffer||info.bufferSize<data.length){if(info.buffer)_free(info.buffer);info.bufferSize=data.length;info.buffer=_malloc(data.length)}HEAPU8.set(data,info.buffer>>>0);callbackInfo.func(info.buffer,data.length,callbackInfo.arg)}else{callbackInfo.func(0,0,callbackInfo.arg)}};Browser.workers.push(info);return id}_emscripten_create_worker.sig="ip";var _emscripten_destroy_worker=id=>{var info=Browser.workers[id];info.worker.terminate();if(info.buffer)_free(info.buffer);Browser.workers[id]=null};_emscripten_destroy_worker.sig="vi";function _emscripten_call_worker(id,funcName,data,size,callback,arg){funcName>>>=0;data>>>=0;callback>>>=0;arg>>>=0;funcName=UTF8ToString(funcName);var info=Browser.workers[id];var callbackId=-1;if(callback){runtimeKeepalivePush();callbackId=info.callbacks.length;info.callbacks.push({func:getWasmTableEntry(callback),arg});info.awaited++}var transferObject={funcName,callbackId,data:data?new Uint8Array(HEAPU8.subarray(data>>>0,data+size>>>0)):0};if(data){info.worker.postMessage(transferObject,[transferObject.data.buffer])}else{info.worker.postMessage(transferObject)}}_emscripten_call_worker.sig="vippipp";var _emscripten_get_worker_queue_size=id=>{var info=Browser.workers[id];if(!info)return-1;return info.awaited};_emscripten_get_worker_queue_size.sig="ii";var getPreloadedImageData=(path,w,h)=>{path=PATH_FS.resolve(path);var canvas=Browser.preloadedImages[path];if(!canvas)return 0;var ctx=canvas.getContext("2d");var image=ctx.getImageData(0,0,canvas.width,canvas.height);var buf=_malloc(canvas.width*canvas.height*4);HEAPU8.set(image.data,buf>>>0);HEAP32[w>>>2>>>0]=canvas.width;HEAP32[h>>>2>>>0]=canvas.height;return buf};function _emscripten_get_preloaded_image_data(path,w,h){path>>>=0;w>>>=0;h>>>=0;return getPreloadedImageData(UTF8ToString(path),w,h)}_emscripten_get_preloaded_image_data.sig="pppp";var getPreloadedImageData__data=["$PATH_FS","malloc"];function _emscripten_get_preloaded_image_data_from_FILE(file,w,h){file>>>=0;w>>>=0;h>>>=0;var fd=_fileno(file);var stream=FS.getStream(fd);if(stream){return getPreloadedImageData(stream.path,w,h)}return 0}_emscripten_get_preloaded_image_data_from_FILE.sig="pppp";var wget={wgetRequests:{},nextWgetRequestHandle:0,getNextWgetRequestHandle(){var handle=wget.nextWgetRequestHandle;wget.nextWgetRequestHandle++;return handle}};var FS_mkdirTree=(path,mode)=>FS.mkdirTree(path,mode);var _emscripten_async_wget=function(url,file,onload,onerror){url>>>=0;file>>>=0;onload>>>=0;onerror>>>=0;runtimeKeepalivePush();var _url=UTF8ToString(url);var _file=UTF8ToString(file);_file=PATH_FS.resolve(_file);function doCallback(callback){if(callback){runtimeKeepalivePop();callUserCallback(()=>{var sp=stackSave();getWasmTableEntry(callback)(stringToUTF8OnStack(_file));stackRestore(sp)})}}var destinationDirectory=PATH.dirname(_file);FS_createPreloadedFile(destinationDirectory,PATH.basename(_file),_url,true,true,()=>doCallback(onload),()=>doCallback(onerror),false,false,()=>{try{FS_unlink(_file)}catch(e){}FS_mkdirTree(destinationDirectory)})};_emscripten_async_wget.sig="vpppp";var _emscripten_async_wget_data=async function(url,userdata,onload,onerror){url>>>=0;userdata>>>=0;onload>>>=0;onerror>>>=0;runtimeKeepalivePush();try{var byteArray=await asyncLoad(UTF8ToString(url));runtimeKeepalivePop();callUserCallback(()=>{var buffer=_malloc(byteArray.length);HEAPU8.set(byteArray,buffer>>>0);getWasmTableEntry(onload)(userdata,buffer,byteArray.length);_free(buffer)})}catch(e){if(onerror){runtimeKeepalivePop();callUserCallback(()=>{getWasmTableEntry(onerror)(userdata)})}}};_emscripten_async_wget_data.sig="vpppp";var _emscripten_async_wget2=function(url,file,request,param,userdata,onload,onerror,onprogress){url>>>=0;file>>>=0;request>>>=0;param>>>=0;userdata>>>=0;onload>>>=0;onerror>>>=0;onprogress>>>=0;runtimeKeepalivePush();var _url=UTF8ToString(url);var _file=UTF8ToString(file);_file=PATH_FS.resolve(_file);var _request=UTF8ToString(request);var _param=UTF8ToString(param);var index=_file.lastIndexOf("/");var http=new XMLHttpRequest;http.open(_request,_url,true);http.responseType="arraybuffer";var handle=wget.getNextWgetRequestHandle();var destinationDirectory=PATH.dirname(_file);http.onload=e=>{runtimeKeepalivePop();if(http.status>=200&&http.status<300){try{FS.unlink(_file)}catch(e){}FS.mkdirTree(destinationDirectory);FS.createDataFile(_file.slice(0,index),_file.slice(index+1),new Uint8Array(http.response),true,true,false);if(onload){var sp=stackSave();getWasmTableEntry(onload)(handle,userdata,stringToUTF8OnStack(_file));stackRestore(sp)}}else{if(onerror)getWasmTableEntry(onerror)(handle,userdata,http.status)}delete wget.wgetRequests[handle]};http.onerror=e=>{runtimeKeepalivePop();if(onerror)getWasmTableEntry(onerror)(handle,userdata,http.status);delete wget.wgetRequests[handle]};http.onprogress=e=>{if(e.lengthComputable||e.lengthComputable===undefined&&e.total!=0){var percentComplete=e.loaded/e.total*100;if(onprogress)getWasmTableEntry(onprogress)(handle,userdata,percentComplete)}};http.onabort=e=>{runtimeKeepalivePop();delete wget.wgetRequests[handle]};if(_request=="POST"){http.setRequestHeader("Content-type","application/x-www-form-urlencoded");http.send(_param)}else{http.send(null)}wget.wgetRequests[handle]=http;return handle};_emscripten_async_wget2.sig="ipppppppp";function _emscripten_async_wget2_data(url,request,param,userdata,free,onload,onerror,onprogress){url>>>=0;request>>>=0;param>>>=0;userdata>>>=0;onload>>>=0;onerror>>>=0;onprogress>>>=0;var _url=UTF8ToString(url);var _request=UTF8ToString(request);var _param=UTF8ToString(param);var http=new XMLHttpRequest;http.open(_request,_url,true);http.responseType="arraybuffer";var handle=wget.getNextWgetRequestHandle();function onerrorjs(){if(onerror){var sp=stackSave();var statusText=0;if(http.statusText){statusText=stringToUTF8OnStack(http.statusText)}getWasmTableEntry(onerror)(handle,userdata,http.status,statusText);stackRestore(sp)}}http.onload=e=>{if(http.status>=200&&http.status<300||http.status===0&&_url.slice(0,4).toLowerCase()!="http"){var byteArray=new Uint8Array(http.response);var buffer=_malloc(byteArray.length);HEAPU8.set(byteArray,buffer>>>0);if(onload)getWasmTableEntry(onload)(handle,userdata,buffer,byteArray.length);if(free)_free(buffer)}else{onerrorjs()}delete wget.wgetRequests[handle]};http.onerror=e=>{onerrorjs();delete wget.wgetRequests[handle]};http.onprogress=e=>{if(onprogress)getWasmTableEntry(onprogress)(handle,userdata,e.loaded,e.lengthComputable||e.lengthComputable===undefined?e.total:0)};http.onabort=e=>{delete wget.wgetRequests[handle]};if(_request=="POST"){http.setRequestHeader("Content-type","application/x-www-form-urlencoded");http.send(_param)}else{http.send(null)}wget.wgetRequests[handle]=http;return handle}_emscripten_async_wget2_data.sig="ippppippp";var _emscripten_async_wget2_abort=handle=>{var http=wget.wgetRequests[handle];http?.abort()};_emscripten_async_wget2_abort.sig="vi";function ___asctime_r(tmPtr,buf){tmPtr>>>=0;buf>>>=0;var date={tm_sec:HEAP32[tmPtr>>>2>>>0],tm_min:HEAP32[tmPtr+4>>>2>>>0],tm_hour:HEAP32[tmPtr+8>>>2>>>0],tm_mday:HEAP32[tmPtr+12>>>2>>>0],tm_mon:HEAP32[tmPtr+16>>>2>>>0],tm_year:HEAP32[tmPtr+20>>>2>>>0],tm_wday:HEAP32[tmPtr+24>>>2>>>0]};var days=["Sun","Mon","Tue","Wed","Thu","Fri","Sat"];var months=["Jan","Feb","Mar","Apr","May","Jun","Jul","Aug","Sep","Oct","Nov","Dec"];var s=days[date.tm_wday]+" "+months[date.tm_mon]+(date.tm_mday<10?"  ":" ")+date.tm_mday+(date.tm_hour<10?" 0":" ")+date.tm_hour+(date.tm_min<10?":0":":")+date.tm_min+(date.tm_sec<10?":0":":")+date.tm_sec+" "+(1900+date.tm_year)+"\n";stringToUTF8(s,buf,26);return buf}___asctime_r.sig="ppp";var MONTH_DAYS_REGULAR=[31,28,31,30,31,30,31,31,30,31,30,31];var MONTH_DAYS_LEAP=[31,29,31,30,31,30,31,31,30,31,30,31];var arraySum=(array,index)=>{var sum=0;for(var i=0;i<=index;sum+=array[i++]){}return sum};var addDays=(date,days)=>{var newDate=new Date(date.getTime());while(days>0){var leap=isLeapYear(newDate.getFullYear());var currentMonth=newDate.getMonth();var daysInCurrentMonth=(leap?MONTH_DAYS_LEAP:MONTH_DAYS_REGULAR)[currentMonth];if(days>daysInCurrentMonth-newDate.getDate()){days-=daysInCurrentMonth-newDate.getDate()+1;newDate.setDate(1);if(currentMonth<11){newDate.setMonth(currentMonth+1)}else{newDate.setMonth(0);newDate.setFullYear(newDate.getFullYear()+1)}}else{newDate.setDate(newDate.getDate()+days);return newDate}}return newDate};function _strptime(buf,format,tm){buf>>>=0;format>>>=0;tm>>>=0;var pattern=UTF8ToString(format);var SPECIAL_CHARS="\\!@#$^&*()+=-[]/{}|:<>?,.";for(var i=0,ii=SPECIAL_CHARS.length;i<ii;++i){pattern=pattern.replace(new RegExp("\\"+SPECIAL_CHARS[i],"g"),"\\"+SPECIAL_CHARS[i])}var EQUIVALENT_MATCHERS={A:"%a",B:"%b",c:"%a %b %d %H:%M:%S %Y",D:"%m\\/%d\\/%y",e:"%d",F:"%Y-%m-%d",h:"%b",R:"%H\\:%M",r:"%I\\:%M\\:%S\\s%p",T:"%H\\:%M\\:%S",x:"%m\\/%d\\/(?:%y|%Y)",X:"%H\\:%M\\:%S"};var DATE_PATTERNS={a:"(?:Sun(?:day)?)|(?:Mon(?:day)?)|(?:Tue(?:sday)?)|(?:Wed(?:nesday)?)|(?:Thu(?:rsday)?)|(?:Fri(?:day)?)|(?:Sat(?:urday)?)",b:"(?:Jan(?:uary)?)|(?:Feb(?:ruary)?)|(?:Mar(?:ch)?)|(?:Apr(?:il)?)|May|(?:Jun(?:e)?)|(?:Jul(?:y)?)|(?:Aug(?:ust)?)|(?:Sep(?:tember)?)|(?:Oct(?:ober)?)|(?:Nov(?:ember)?)|(?:Dec(?:ember)?)",C:"\\d\\d",d:"0[1-9]|[1-9](?!\\d)|1\\d|2\\d|30|31",H:"\\d(?!\\d)|[0,1]\\d|20|21|22|23",I:"\\d(?!\\d)|0\\d|10|11|12",j:"00[1-9]|0?[1-9](?!\\d)|0?[1-9]\\d(?!\\d)|[1,2]\\d\\d|3[0-6]\\d",m:"0[1-9]|[1-9](?!\\d)|10|11|12",M:"0\\d|\\d(?!\\d)|[1-5]\\d",n:" ",p:"AM|am|PM|pm|A\\.M\\.|a\\.m\\.|P\\.M\\.|p\\.m\\.",S:"0\\d|\\d(?!\\d)|[1-5]\\d|60",U:"0\\d|\\d(?!\\d)|[1-4]\\d|50|51|52|53",W:"0\\d|\\d(?!\\d)|[1-4]\\d|50|51|52|53",w:"[0-6]",y:"\\d\\d",Y:"\\d\\d\\d\\d",t:" ",z:"Z|(?:[\\+\\-]\\d\\d:?(?:\\d\\d)?)"};var MONTH_NUMBERS={JAN:0,FEB:1,MAR:2,APR:3,MAY:4,JUN:5,JUL:6,AUG:7,SEP:8,OCT:9,NOV:10,DEC:11};var DAY_NUMBERS_SUN_FIRST={SUN:0,MON:1,TUE:2,WED:3,THU:4,FRI:5,SAT:6};var DAY_NUMBERS_MON_FIRST={MON:0,TUE:1,WED:2,THU:3,FRI:4,SAT:5,SUN:6};var capture=[];var pattern_out=pattern.replace(/%(.)/g,(m,c)=>EQUIVALENT_MATCHERS[c]||m).replace(/%(.)/g,(_,c)=>{let pat=DATE_PATTERNS[c];if(pat){capture.push(c);return`(${pat})`}else{return c}}).replace(/\s+/g,"\\s*");var matches=new RegExp("^"+pattern_out,"i").exec(UTF8ToString(buf));function initDate(){function fixup(value,min,max){return typeof value!="number"||isNaN(value)?min:value>=min?value<=max?value:max:min}return{year:fixup(HEAP32[tm+20>>>2>>>0]+1900,1970,9999),month:fixup(HEAP32[tm+16>>>2>>>0],0,11),day:fixup(HEAP32[tm+12>>>2>>>0],1,31),hour:fixup(HEAP32[tm+8>>>2>>>0],0,23),min:fixup(HEAP32[tm+4>>>2>>>0],0,59),sec:fixup(HEAP32[tm>>>2>>>0],0,59),gmtoff:0}}if(matches){var date=initDate();var value;var getMatch=symbol=>{var pos=capture.indexOf(symbol);if(pos>=0){return matches[pos+1]}return};if(value=getMatch("S")){date.sec=Number(value)}if(value=getMatch("M")){date.min=Number(value)}if(value=getMatch("H")){date.hour=Number(value)}else if(value=getMatch("I")){var hour=Number(value);if(value=getMatch("p")){hour+=value.toUpperCase()[0]==="P"?12:0}date.hour=hour}if(value=getMatch("Y")){date.year=Number(value)}else if(value=getMatch("y")){var year=Number(value);if(value=getMatch("C")){year+=Number(value)*100}else{year+=year<69?2e3:1900}date.year=year}if(value=getMatch("m")){date.month=Number(value)-1}else if(value=getMatch("b")){date.month=MONTH_NUMBERS[value.substring(0,3).toUpperCase()]||0}if(value=getMatch("d")){date.day=Number(value)}else if(value=getMatch("j")){var day=Number(value);var leapYear=isLeapYear(date.year);for(var month=0;month<12;++month){var daysUntilMonth=arraySum(leapYear?MONTH_DAYS_LEAP:MONTH_DAYS_REGULAR,month-1);if(day<=daysUntilMonth+(leapYear?MONTH_DAYS_LEAP:MONTH_DAYS_REGULAR)[month]){date.day=day-daysUntilMonth}}}else if(value=getMatch("a")){var weekDay=value.substring(0,3).toUpperCase();if(value=getMatch("U")){var weekDayNumber=DAY_NUMBERS_SUN_FIRST[weekDay];var weekNumber=Number(value);var janFirst=new Date(date.year,0,1);var endDate;if(janFirst.getDay()===0){endDate=addDays(janFirst,weekDayNumber+7*(weekNumber-1))}else{endDate=addDays(janFirst,7-janFirst.getDay()+weekDayNumber+7*(weekNumber-1))}date.day=endDate.getDate();date.month=endDate.getMonth()}else if(value=getMatch("W")){var weekDayNumber=DAY_NUMBERS_MON_FIRST[weekDay];var weekNumber=Number(value);var janFirst=new Date(date.year,0,1);var endDate;if(janFirst.getDay()===1){endDate=addDays(janFirst,weekDayNumber+7*(weekNumber-1))}else{endDate=addDays(janFirst,7-janFirst.getDay()+1+weekDayNumber+7*(weekNumber-1))}date.day=endDate.getDate();date.month=endDate.getMonth()}}if(value=getMatch("z")){if(value.toLowerCase()==="z"){date.gmtoff=0}else{var match=value.match(/^((?:\-|\+)\d\d):?(\d\d)?/);date.gmtoff=match[1]*3600;if(match[2]){date.gmtoff+=date.gmtoff>0?match[2]*60:-match[2]*60}}}var fullDate=new Date(date.year,date.month,date.day,date.hour,date.min,date.sec,0);HEAP32[tm>>>2>>>0]=fullDate.getSeconds();HEAP32[tm+4>>>2>>>0]=fullDate.getMinutes();HEAP32[tm+8>>>2>>>0]=fullDate.getHours();HEAP32[tm+12>>>2>>>0]=fullDate.getDate();HEAP32[tm+16>>>2>>>0]=fullDate.getMonth();HEAP32[tm+20>>>2>>>0]=fullDate.getFullYear()-1900;HEAP32[tm+24>>>2>>>0]=fullDate.getDay();HEAP32[tm+28>>>2>>>0]=arraySum(isLeapYear(fullDate.getFullYear())?MONTH_DAYS_LEAP:MONTH_DAYS_REGULAR,fullDate.getMonth()-1)+fullDate.getDate()-1;HEAP32[tm+32>>>2>>>0]=0;HEAP32[tm+36>>>2>>>0]=date.gmtoff;return buf+intArrayFromString(matches[0]).length-1}return 0}_strptime.sig="pppp";function _strptime_l(buf,format,tm,locale){buf>>>=0;format>>>=0;tm>>>=0;locale>>>=0;return _strptime(buf,format,tm)}_strptime_l.sig="ppppp";function __dlsym_catchup_js(handle,symbolIndex){handle>>>=0;var lib=LDSO.loadedLibsByHandle[handle];var symDict=lib.exports;var symName=Object.keys(symDict)[symbolIndex];var sym=symDict[symName];var result=addFunction(sym,sym.sig);return result}__dlsym_catchup_js.sig="ppi";var FS_readFile=(...args)=>FS.readFile(...args);var FS_root=(...args)=>FS.root(...args);var FS_mounts=(...args)=>FS.mounts(...args);var FS_devices=(...args)=>FS.devices(...args);var FS_streams=(...args)=>FS.streams(...args);var FS_nextInode=(...args)=>FS.nextInode(...args);var FS_nameTable=(...args)=>FS.nameTable(...args);var FS_currentPath=(...args)=>FS.currentPath(...args);var FS_initialized=(...args)=>FS.initialized(...args);var FS_ignorePermissions=(...args)=>FS.ignorePermissions(...args);var FS_trackingDelegate=(...args)=>FS.trackingDelegate(...args);var FS_filesystems=(...args)=>FS.filesystems(...args);var FS_syncFSRequests=(...args)=>FS.syncFSRequests(...args);var FS_readFiles=(...args)=>FS.readFiles(...args);var FS_lookupPath=(...args)=>FS.lookupPath(...args);var FS_getPath=(...args)=>FS.getPath(...args);var FS_hashName=(...args)=>FS.hashName(...args);var FS_hashAddNode=(...args)=>FS.hashAddNode(...args);var FS_hashRemoveNode=(...args)=>FS.hashRemoveNode(...args);var FS_lookupNode=(...args)=>FS.lookupNode(...args);var FS_createNode=(...args)=>FS.createNode(...args);var FS_destroyNode=(...args)=>FS.destroyNode(...args);var FS_isRoot=(...args)=>FS.isRoot(...args);var FS_isMountpoint=(...args)=>FS.isMountpoint(...args);var FS_isFile=(...args)=>FS.isFile(...args);var FS_isDir=(...args)=>FS.isDir(...args);var FS_isLink=(...args)=>FS.isLink(...args);var FS_isChrdev=(...args)=>FS.isChrdev(...args);var FS_isBlkdev=(...args)=>FS.isBlkdev(...args);var FS_isFIFO=(...args)=>FS.isFIFO(...args);var FS_isSocket=(...args)=>FS.isSocket(...args);var FS_flagsToPermissionString=(...args)=>FS.flagsToPermissionString(...args);var FS_nodePermissions=(...args)=>FS.nodePermissions(...args);var FS_mayLookup=(...args)=>FS.mayLookup(...args);var FS_mayCreate=(...args)=>FS.mayCreate(...args);var FS_mayDelete=(...args)=>FS.mayDelete(...args);var FS_mayOpen=(...args)=>FS.mayOpen(...args);var FS_checkOpExists=(...args)=>FS.checkOpExists(...args);var FS_nextfd=(...args)=>FS.nextfd(...args);var FS_getStreamChecked=(...args)=>FS.getStreamChecked(...args);var FS_getStream=(...args)=>FS.getStream(...args);var FS_createStream=(...args)=>FS.createStream(...args);var FS_closeStream=(...args)=>FS.closeStream(...args);var FS_dupStream=(...args)=>FS.dupStream(...args);var FS_doSetAttr=(...args)=>FS.doSetAttr(...args);var FS_chrdev_stream_ops=(...args)=>FS.chrdev_stream_ops(...args);var FS_major=(...args)=>FS.major(...args);var FS_minor=(...args)=>FS.minor(...args);var FS_makedev=(...args)=>FS.makedev(...args);var FS_registerDevice=(...args)=>FS.registerDevice(...args);var FS_getDevice=(...args)=>FS.getDevice(...args);var FS_getMounts=(...args)=>FS.getMounts(...args);var FS_syncfs=(...args)=>FS.syncfs(...args);var FS_mount=(...args)=>FS.mount(...args);var FS_unmount=(...args)=>FS.unmount(...args);var FS_lookup=(...args)=>FS.lookup(...args);var FS_mknod=(...args)=>FS.mknod(...args);var FS_statfs=(...args)=>FS.statfs(...args);var FS_statfsStream=(...args)=>FS.statfsStream(...args);var FS_statfsNode=(...args)=>FS.statfsNode(...args);var FS_create=(...args)=>FS.create(...args);var FS_mkdir=(...args)=>FS.mkdir(...args);var FS_mkdev=(...args)=>FS.mkdev(...args);var FS_symlink=(...args)=>FS.symlink(...args);var FS_rename=(...args)=>FS.rename(...args);var FS_rmdir=(...args)=>FS.rmdir(...args);var FS_readdir=(...args)=>FS.readdir(...args);var FS_readlink=(...args)=>FS.readlink(...args);var FS_stat=(...args)=>FS.stat(...args);var FS_fstat=(...args)=>FS.fstat(...args);var FS_lstat=(...args)=>FS.lstat(...args);var FS_doChmod=(...args)=>FS.doChmod(...args);var FS_chmod=(...args)=>FS.chmod(...args);var FS_lchmod=(...args)=>FS.lchmod(...args);var FS_fchmod=(...args)=>FS.fchmod(...args);var FS_doChown=(...args)=>FS.doChown(...args);var FS_chown=(...args)=>FS.chown(...args);var FS_lchown=(...args)=>FS.lchown(...args);var FS_fchown=(...args)=>FS.fchown(...args);var FS_doTruncate=(...args)=>FS.doTruncate(...args);var FS_truncate=(...args)=>FS.truncate(...args);var FS_ftruncate=(...args)=>FS.ftruncate(...args);var FS_utime=(...args)=>FS.utime(...args);var FS_open=(...args)=>FS.open(...args);var FS_close=(...args)=>FS.close(...args);var FS_isClosed=(...args)=>FS.isClosed(...args);var FS_llseek=(...args)=>FS.llseek(...args);var FS_read=(...args)=>FS.read(...args);var FS_write=(...args)=>FS.write(...args);var FS_mmap=(...args)=>FS.mmap(...args);var FS_msync=(...args)=>FS.msync(...args);var FS_ioctl=(...args)=>FS.ioctl(...args);var FS_writeFile=(...args)=>FS.writeFile(...args);var FS_cwd=(...args)=>FS.cwd(...args);var FS_chdir=(...args)=>FS.chdir(...args);var FS_createDefaultDirectories=(...args)=>FS.createDefaultDirectories(...args);var FS_createDefaultDevices=(...args)=>FS.createDefaultDevices(...args);var FS_createSpecialDirectories=(...args)=>FS.createSpecialDirectories(...args);var FS_createStandardStreams=(...args)=>FS.createStandardStreams(...args);var FS_staticInit=(...args)=>FS.staticInit(...args);var FS_init=(...args)=>FS.init(...args);var FS_quit=(...args)=>FS.quit(...args);var FS_findObject=(...args)=>FS.findObject(...args);var FS_analyzePath=(...args)=>FS.analyzePath(...args);var FS_createFile=(...args)=>FS.createFile(...args);var FS_forceLoadFile=(...args)=>FS.forceLoadFile(...args);var _setNetworkCallback=(event,userData,callback)=>{function _callback(data){callUserCallback(()=>{if(event==="error"){withStackSave(()=>{var msg=stringToUTF8OnStack(data[2]);getWasmTableEntry(callback)(data[0],data[1],msg,userData)})}else{getWasmTableEntry(callback)(data,userData)}})}runtimeKeepalivePush();SOCKFS.on(event,callback?_callback:null)};function _emscripten_set_socket_error_callback(userData,callback){userData>>>=0;callback>>>=0;return _setNetworkCallback("error",userData,callback)}_emscripten_set_socket_error_callback.sig="vpp";function _emscripten_set_socket_open_callback(userData,callback){userData>>>=0;callback>>>=0;return _setNetworkCallback("open",userData,callback)}_emscripten_set_socket_open_callback.sig="vpp";function _emscripten_set_socket_listen_callback(userData,callback){userData>>>=0;callback>>>=0;return _setNetworkCallback("listen",userData,callback)}_emscripten_set_socket_listen_callback.sig="vpp";function _emscripten_set_socket_connection_callback(userData,callback){userData>>>=0;callback>>>=0;return _setNetworkCallback("connection",userData,callback)}_emscripten_set_socket_connection_callback.sig="vpp";function _emscripten_set_socket_message_callback(userData,callback){userData>>>=0;callback>>>=0;return _setNetworkCallback("message",userData,callback)}_emscripten_set_socket_message_callback.sig="vpp";function _emscripten_set_socket_close_callback(userData,callback){userData>>>=0;callback>>>=0;return _setNetworkCallback("close",userData,callback)}_emscripten_set_socket_close_callback.sig="vpp";function _emscripten_webgl_enable_ANGLE_instanced_arrays(ctx){ctx>>>=0;return webgl_enable_ANGLE_instanced_arrays(GL.contexts[ctx].GLctx)}_emscripten_webgl_enable_ANGLE_instanced_arrays.sig="ip";function _emscripten_webgl_enable_OES_vertex_array_object(ctx){ctx>>>=0;return webgl_enable_OES_vertex_array_object(GL.contexts[ctx].GLctx)}_emscripten_webgl_enable_OES_vertex_array_object.sig="ip";function _emscripten_webgl_enable_WEBGL_draw_buffers(ctx){ctx>>>=0;return webgl_enable_WEBGL_draw_buffers(GL.contexts[ctx].GLctx)}_emscripten_webgl_enable_WEBGL_draw_buffers.sig="ip";function _emscripten_webgl_enable_WEBGL_multi_draw(ctx){ctx>>>=0;return webgl_enable_WEBGL_multi_draw(GL.contexts[ctx].GLctx)}_emscripten_webgl_enable_WEBGL_multi_draw.sig="ip";function _emscripten_webgl_enable_EXT_polygon_offset_clamp(ctx){ctx>>>=0;return webgl_enable_EXT_polygon_offset_clamp(GL.contexts[ctx].GLctx)}_emscripten_webgl_enable_EXT_polygon_offset_clamp.sig="ip";function _emscripten_webgl_enable_EXT_clip_control(ctx){ctx>>>=0;return webgl_enable_EXT_clip_control(GL.contexts[ctx].GLctx)}_emscripten_webgl_enable_EXT_clip_control.sig="ip";function _emscripten_webgl_enable_WEBGL_polygon_mode(ctx){ctx>>>=0;return webgl_enable_WEBGL_polygon_mode(GL.contexts[ctx].GLctx)}_emscripten_webgl_enable_WEBGL_polygon_mode.sig="ip";function _glVertexPointer(size,type,stride,ptr){ptr>>>=0;throw"Legacy GL function (glVertexPointer) called. If you want legacy GL emulation, you need to compile with -sLEGACY_GL_EMULATION to enable legacy GL emulation."}_glVertexPointer.sig="viiip";var _glMatrixMode=()=>{throw"Legacy GL function (glMatrixMode) called. If you want legacy GL emulation, you need to compile with -sLEGACY_GL_EMULATION to enable legacy GL emulation."};_glMatrixMode.sig="vi";var _glBegin=()=>{throw"Legacy GL function (glBegin) called. If you want legacy GL emulation, you need to compile with -sLEGACY_GL_EMULATION to enable legacy GL emulation."};_glBegin.sig="vi";var _glLoadIdentity=()=>{throw"Legacy GL function (glLoadIdentity) called. If you want legacy GL emulation, you need to compile with -sLEGACY_GL_EMULATION to enable legacy GL emulation."};_glLoadIdentity.sig="v";function _glMultiDrawArraysWEBGL(mode,firsts,counts,drawcount){firsts>>>=0;counts>>>=0;GLctx.multiDrawWebgl["multiDrawArraysWEBGL"](mode,HEAP32,firsts>>>2,HEAP32,counts>>>2,drawcount)}_glMultiDrawArraysWEBGL.sig="vippi";var _glMultiDrawArrays=_glMultiDrawArraysWEBGL;_glMultiDrawArrays.sig="vippi";var _glMultiDrawArraysANGLE=_glMultiDrawArraysWEBGL;function _glMultiDrawArraysInstancedWEBGL(mode,firsts,counts,instanceCounts,drawcount){firsts>>>=0;counts>>>=0;instanceCounts>>>=0;GLctx.multiDrawWebgl["multiDrawArraysInstancedWEBGL"](mode,HEAP32,firsts>>>2,HEAP32,counts>>>2,HEAP32,instanceCounts>>>2,drawcount)}_glMultiDrawArraysInstancedWEBGL.sig="vipppi";var _glMultiDrawArraysInstancedANGLE=_glMultiDrawArraysInstancedWEBGL;function _glMultiDrawElementsWEBGL(mode,counts,type,offsets,drawcount){counts>>>=0;offsets>>>=0;GLctx.multiDrawWebgl["multiDrawElementsWEBGL"](mode,HEAP32,counts>>>2,type,HEAP32,offsets>>>2,drawcount)}_glMultiDrawElementsWEBGL.sig="vipipi";var _glMultiDrawElements=_glMultiDrawElementsWEBGL;_glMultiDrawElements.sig="vipipi";var _glMultiDrawElementsANGLE=_glMultiDrawElementsWEBGL;function _glMultiDrawElementsInstancedWEBGL(mode,counts,type,offsets,instanceCounts,drawcount){counts>>>=0;offsets>>>=0;instanceCounts>>>=0;GLctx.multiDrawWebgl["multiDrawElementsInstancedWEBGL"](mode,HEAP32,counts>>>2,type,HEAP32,offsets>>>2,HEAP32,instanceCounts>>>2,drawcount)}_glMultiDrawElementsInstancedWEBGL.sig="vipippi";var _glMultiDrawElementsInstancedANGLE=_glMultiDrawElementsInstancedWEBGL;var _glClearDepth=x0=>GLctx.clearDepth(x0);_glClearDepth.sig="vd";var _glDepthRange=(x0,x1)=>GLctx.depthRange(x0,x1);_glDepthRange.sig="vdd";var _emscripten_glVertexPointer=_glVertexPointer;_emscripten_glVertexPointer.sig="viiip";var _emscripten_glMatrixMode=_glMatrixMode;_emscripten_glMatrixMode.sig="vi";var _emscripten_glBegin=_glBegin;_emscripten_glBegin.sig="vi";var _emscripten_glLoadIdentity=_glLoadIdentity;_emscripten_glLoadIdentity.sig="v";var _emscripten_glMultiDrawArrays=_glMultiDrawArrays;_emscripten_glMultiDrawArrays.sig="vippi";var _emscripten_glMultiDrawArraysANGLE=_glMultiDrawArraysANGLE;var _emscripten_glMultiDrawArraysWEBGL=_glMultiDrawArraysWEBGL;var _emscripten_glMultiDrawArraysInstancedANGLE=_glMultiDrawArraysInstancedANGLE;var _emscripten_glMultiDrawArraysInstancedWEBGL=_glMultiDrawArraysInstancedWEBGL;var _emscripten_glMultiDrawElements=_glMultiDrawElements;_emscripten_glMultiDrawElements.sig="vipipi";var _emscripten_glMultiDrawElementsANGLE=_glMultiDrawElementsANGLE;var _emscripten_glMultiDrawElementsWEBGL=_glMultiDrawElementsWEBGL;var _emscripten_glMultiDrawElementsInstancedANGLE=_glMultiDrawElementsInstancedANGLE;var _emscripten_glMultiDrawElementsInstancedWEBGL=_glMultiDrawElementsInstancedWEBGL;var _emscripten_glClearDepth=_glClearDepth;_emscripten_glClearDepth.sig="vd";var _emscripten_glDepthRange=_glDepthRange;_emscripten_glDepthRange.sig="vdd";function _glGetBufferSubData(target,offset,size,data){offset>>>=0;size>>>=0;data>>>=0;if(!data){GL.recordError(1281);return}size&&GLctx.getBufferSubData(target,offset,HEAPU8.subarray(data>>>0,data+size>>>0))}_glGetBufferSubData.sig="vippp";var _glDrawArraysInstancedBaseInstanceWEBGL=(mode,first,count,instanceCount,baseInstance)=>{GLctx.dibvbi["drawArraysInstancedBaseInstanceWEBGL"](mode,first,count,instanceCount,baseInstance)};_glDrawArraysInstancedBaseInstanceWEBGL.sig="viiiii";var _glDrawArraysInstancedBaseInstance=_glDrawArraysInstancedBaseInstanceWEBGL;_glDrawArraysInstancedBaseInstance.sig="viiiii";var _glDrawArraysInstancedBaseInstanceANGLE=_glDrawArraysInstancedBaseInstanceWEBGL;var _glDrawElementsInstancedBaseVertexBaseInstanceWEBGL=(mode,count,type,offset,instanceCount,baseVertex,baseinstance)=>{GLctx.dibvbi["drawElementsInstancedBaseVertexBaseInstanceWEBGL"](mode,count,type,offset,instanceCount,baseVertex,baseinstance)};_glDrawElementsInstancedBaseVertexBaseInstanceWEBGL.sig="viiiiiii";var _glDrawElementsInstancedBaseVertexBaseInstanceANGLE=_glDrawElementsInstancedBaseVertexBaseInstanceWEBGL;function _emscripten_webgl_enable_WEBGL_draw_instanced_base_vertex_base_instance(ctx){ctx>>>=0;return webgl_enable_WEBGL_draw_instanced_base_vertex_base_instance(GL.contexts[ctx].GLctx)}_emscripten_webgl_enable_WEBGL_draw_instanced_base_vertex_base_instance.sig="ip";var _glMultiDrawArraysInstancedBaseInstanceWEBGL=(mode,firsts,counts,instanceCounts,baseInstances,drawCount)=>{GLctx.mdibvbi["multiDrawArraysInstancedBaseInstanceWEBGL"](mode,HEAP32,firsts>>>2,HEAP32,counts>>>2,HEAP32,instanceCounts>>>2,HEAPU32,baseInstances>>>2,drawCount)};_glMultiDrawArraysInstancedBaseInstanceWEBGL.sig="viiiiii";var _glMultiDrawArraysInstancedBaseInstanceANGLE=_glMultiDrawArraysInstancedBaseInstanceWEBGL;var _glMultiDrawElementsInstancedBaseVertexBaseInstanceWEBGL=(mode,counts,type,offsets,instanceCounts,baseVertices,baseInstances,drawCount)=>{GLctx.mdibvbi["multiDrawElementsInstancedBaseVertexBaseInstanceWEBGL"](mode,HEAP32,counts>>>2,type,HEAP32,offsets>>>2,HEAP32,instanceCounts>>>2,HEAP32,baseVertices>>>2,HEAPU32,baseInstances>>>2,drawCount)};_glMultiDrawElementsInstancedBaseVertexBaseInstanceWEBGL.sig="viiiiiiii";var _glMultiDrawElementsInstancedBaseVertexBaseInstanceANGLE=_glMultiDrawElementsInstancedBaseVertexBaseInstanceWEBGL;function _emscripten_webgl_enable_WEBGL_multi_draw_instanced_base_vertex_base_instance(ctx){ctx>>>=0;return webgl_enable_WEBGL_multi_draw_instanced_base_vertex_base_instance(GL.contexts[ctx].GLctx)}_emscripten_webgl_enable_WEBGL_multi_draw_instanced_base_vertex_base_instance.sig="ip";var _emscripten_glGetBufferSubData=_glGetBufferSubData;_emscripten_glGetBufferSubData.sig="vippp";var _emscripten_glDrawArraysInstancedBaseInstanceWEBGL=_glDrawArraysInstancedBaseInstanceWEBGL;var _emscripten_glDrawArraysInstancedBaseInstance=_glDrawArraysInstancedBaseInstance;_emscripten_glDrawArraysInstancedBaseInstance.sig="viiiii";var _emscripten_glDrawArraysInstancedBaseInstanceANGLE=_glDrawArraysInstancedBaseInstanceANGLE;var _emscripten_glDrawElementsInstancedBaseVertexBaseInstanceWEBGL=_glDrawElementsInstancedBaseVertexBaseInstanceWEBGL;var _emscripten_glDrawElementsInstancedBaseVertexBaseInstanceANGLE=_glDrawElementsInstancedBaseVertexBaseInstanceANGLE;var _emscripten_glMultiDrawArraysInstancedBaseInstanceWEBGL=_glMultiDrawArraysInstancedBaseInstanceWEBGL;var _emscripten_glMultiDrawArraysInstancedBaseInstanceANGLE=_glMultiDrawArraysInstancedBaseInstanceANGLE;var _emscripten_glMultiDrawElementsInstancedBaseVertexBaseInstanceWEBGL=_glMultiDrawElementsInstancedBaseVertexBaseInstanceWEBGL;var _emscripten_glMultiDrawElementsInstancedBaseVertexBaseInstanceANGLE=_glMultiDrawElementsInstancedBaseVertexBaseInstanceANGLE;var ALLOC_NORMAL=0;var ALLOC_STACK=1;var allocate=(slab,allocator)=>{var ret;if(allocator==ALLOC_STACK){ret=stackAlloc(slab.length)}else{ret=_malloc(slab.length)}if(!slab.subarray&&!slab.slice){slab=new Uint8Array(slab)}HEAPU8.set(slab,ret>>>0);return ret};var writeStringToMemory=(string,buffer,dontAddNull)=>{warnOnce("writeStringToMemory is deprecated and should not be called! Use stringToUTF8() instead!");var lastChar,end;if(dontAddNull){end=buffer+lengthBytesUTF8(string);lastChar=HEAP8[end>>>0]}stringToUTF8(string,buffer,Infinity);if(dontAddNull)HEAP8[end>>>0]=lastChar};var writeAsciiToMemory=(str,buffer,dontAddNull)=>{for(var i=0;i<str.length;++i){HEAP8[buffer++>>>0]=str.charCodeAt(i)}if(!dontAddNull)HEAP8[buffer>>>0]=0};var allocateUTF8=stringToNewUTF8;var allocateUTF8OnStack=stringToUTF8OnStack;var demangle=func=>{demangle.recursionGuard=(demangle.recursionGuard|0)+1;if(demangle.recursionGuard>1)return func;return withStackSave(()=>{try{var s=func;if(s.startsWith("__Z"))s=s.slice(1);var buf=stringToUTF8OnStack(s);var status=stackAlloc(4);var ret=___cxa_demangle(buf,0,0,status);if(HEAP32[status>>>2>>>0]===0&&ret){return UTF8ToString(ret)}}catch(e){}finally{_free(ret);if(demangle.recursionGuard<2)--demangle.recursionGuard}return func})};var stackTrace=()=>{var js=jsStackTrace();if(Module["extraStackTrace"])js+="\n"+Module["extraStackTrace"]();return js};var print=out;var printErr=err;var jstoi_s=Number;var _emscripten_is_main_browser_thread=()=>!ENVIRONMENT_IS_WORKER;var webSockets=new HandleAllocator;var WS={socketEvent:null,getSocket(socketId){if(!webSockets.has(socketId)){return 0}return webSockets.get(socketId)},getSocketEvent(socketId){this.socketEvent||=_malloc(520);HEAPU32[this.socketEvent>>>2>>>0]=socketId;return this.socketEvent}};function _emscripten_websocket_get_ready_state(socketId,readyState){readyState>>>=0;var socket=WS.getSocket(socketId);if(!socket){return-3}HEAP16[readyState>>>1>>>0]=socket.readyState;return 0}_emscripten_websocket_get_ready_state.sig="iip";function _emscripten_websocket_get_buffered_amount(socketId,bufferedAmount){bufferedAmount>>>=0;var socket=WS.getSocket(socketId);if(!socket){return-3}HEAPU32[bufferedAmount>>>2>>>0]=socket.bufferedAmount;return 0}_emscripten_websocket_get_buffered_amount.sig="iip";function _emscripten_websocket_get_extensions(socketId,extensions,extensionsLength){extensions>>>=0;var socket=WS.getSocket(socketId);if(!socket){return-3}if(!extensions)return-5;stringToUTF8(socket.extensions,extensions,extensionsLength);return 0}_emscripten_websocket_get_extensions.sig="iipi";function _emscripten_websocket_get_extensions_length(socketId,extensionsLength){extensionsLength>>>=0;var socket=WS.getSocket(socketId);if(!socket){return-3}if(!extensionsLength)return-5;HEAP32[extensionsLength>>>2>>>0]=lengthBytesUTF8(socket.extensions)+1;return 0}_emscripten_websocket_get_extensions_length.sig="iip";function _emscripten_websocket_get_protocol(socketId,protocol,protocolLength){protocol>>>=0;var socket=WS.getSocket(socketId);if(!socket){return-3}if(!protocol)return-5;stringToUTF8(socket.protocol,protocol,protocolLength);return 0}_emscripten_websocket_get_protocol.sig="iipi";function _emscripten_websocket_get_protocol_length(socketId,protocolLength){protocolLength>>>=0;var socket=WS.getSocket(socketId);if(!socket){return-3}if(!protocolLength)return-5;HEAP32[protocolLength>>>2>>>0]=lengthBytesUTF8(socket.protocol)+1;return 0}_emscripten_websocket_get_protocol_length.sig="iip";function _emscripten_websocket_get_url(socketId,url,urlLength){url>>>=0;var socket=WS.getSocket(socketId);if(!socket){return-3}if(!url)return-5;stringToUTF8(socket.url,url,urlLength);return 0}_emscripten_websocket_get_url.sig="iipi";function _emscripten_websocket_get_url_length(socketId,urlLength){urlLength>>>=0;var socket=WS.getSocket(socketId);if(!socket){return-3}if(!urlLength)return-5;HEAP32[urlLength>>>2>>>0]=lengthBytesUTF8(socket.url)+1;return 0}_emscripten_websocket_get_url_length.sig="iip";function _emscripten_websocket_set_onopen_callback_on_thread(socketId,userData,callbackFunc,thread){userData>>>=0;callbackFunc>>>=0;thread>>>=0;var socket=WS.getSocket(socketId);if(!socket){return-3}socket.onopen=function(e){var eventPtr=WS.getSocketEvent(socketId);getWasmTableEntry(callbackFunc)(0,eventPtr,userData)};return 0}_emscripten_websocket_set_onopen_callback_on_thread.sig="iippp";function _emscripten_websocket_set_onerror_callback_on_thread(socketId,userData,callbackFunc,thread){userData>>>=0;callbackFunc>>>=0;thread>>>=0;var socket=WS.getSocket(socketId);if(!socket){return-3}socket.onerror=function(e){var eventPtr=WS.getSocketEvent(socketId);getWasmTableEntry(callbackFunc)(0,eventPtr,userData)};return 0}_emscripten_websocket_set_onerror_callback_on_thread.sig="iippp";function _emscripten_websocket_set_onclose_callback_on_thread(socketId,userData,callbackFunc,thread){userData>>>=0;callbackFunc>>>=0;thread>>>=0;var socket=WS.getSocket(socketId);if(!socket){return-3}socket.onclose=function(e){var eventPtr=WS.getSocketEvent(socketId);HEAP8[eventPtr+4>>>0]=e.wasClean,HEAP16[eventPtr+6>>>1>>>0]=e.code,stringToUTF8(e.reason,eventPtr+8,512);getWasmTableEntry(callbackFunc)(0,eventPtr,userData)};return 0}_emscripten_websocket_set_onclose_callback_on_thread.sig="iippp";function _emscripten_websocket_set_onmessage_callback_on_thread(socketId,userData,callbackFunc,thread){userData>>>=0;callbackFunc>>>=0;thread>>>=0;var socket=WS.getSocket(socketId);if(!socket){return-3}socket.onmessage=function(e){var isText=typeof e.data=="string";if(isText){var buf=stringToNewUTF8(e.data);var len=lengthBytesUTF8(e.data)+1}else{var len=e.data.byteLength;var buf=_malloc(len);HEAP8.set(new Uint8Array(e.data),buf>>>0)}var eventPtr=WS.getSocketEvent(socketId);HEAPU32[eventPtr+4>>>2>>>0]=buf,HEAP32[eventPtr+8>>>2>>>0]=len,HEAP8[eventPtr+12>>>0]=isText,getWasmTableEntry(callbackFunc)(0,eventPtr,userData);_free(buf)};return 0}_emscripten_websocket_set_onmessage_callback_on_thread.sig="iippp";function _emscripten_websocket_new(createAttributes){createAttributes>>>=0;if(typeof WebSocket=="undefined"){return-1}if(!createAttributes){return-5}var url=UTF8ToString(HEAPU32[createAttributes>>>2>>>0]);var protocols=HEAPU32[createAttributes+4>>>2>>>0];var socket=protocols?new WebSocket(url,UTF8ToString(protocols).split(",")):new WebSocket(url);socket.binaryType="arraybuffer";var socketId=webSockets.allocate(socket);return socketId}_emscripten_websocket_new.sig="ip";function _emscripten_websocket_send_utf8_text(socketId,textData){textData>>>=0;var socket=WS.getSocket(socketId);if(!socket){return-3}var str=UTF8ToString(textData);socket.send(str);return 0}_emscripten_websocket_send_utf8_text.sig="iip";function _emscripten_websocket_send_binary(socketId,binaryData,dataLength){binaryData>>>=0;var socket=WS.getSocket(socketId);if(!socket){return-3}socket.send(HEAPU8.subarray(binaryData>>>0,binaryData+dataLength>>>0));return 0}_emscripten_websocket_send_binary.sig="iipi";function _emscripten_websocket_close(socketId,code,reason){reason>>>=0;var socket=WS.getSocket(socketId);if(!socket){return-3}var reasonStr=reason?UTF8ToString(reason):undefined;if(reason)socket.close(code||undefined,UTF8ToString(reason));else if(code)socket.close(code);else socket.close();return 0}_emscripten_websocket_close.sig="iiip";var _emscripten_websocket_delete=socketId=>{var socket=WS.getSocket(socketId);if(!socket){return-3}socket.onopen=socket.onerror=socket.onclose=socket.onmessage=null;webSockets.free(socketId);return 0};_emscripten_websocket_delete.sig="ii";var _emscripten_websocket_is_supported=()=>typeof WebSocket!="undefined";_emscripten_websocket_is_supported.sig="i";var _emscripten_websocket_deinitialize=()=>{for(var i in WS.sockets){var socket=WS.sockets[i];if(socket){socket.close();_emscripten_websocket_delete(i)}}WS.sockets=[]};_emscripten_websocket_deinitialize.sig="v";var writeGLArray=(arr,dst,dstLength,heapType)=>{var len=arr.length;var writeLength=dstLength<len?dstLength:len;var heap=heapType?HEAPF32:HEAP32;dst=dst>>>2;for(var i=0;i<writeLength;++i){heap[dst+i>>>0]=arr[i]}return len};var webglPowerPreferences=["default","low-power","high-performance"];function _emscripten_webgl_do_create_context(target,attributes){target>>>=0;attributes>>>=0;var attr32=attributes>>>2;var powerPreference=HEAP32[attr32+(8>>2)>>>0];var contextAttributes={alpha:!!HEAP8[attributes+0>>>0],depth:!!HEAP8[attributes+1>>>0],stencil:!!HEAP8[attributes+2>>>0],antialias:!!HEAP8[attributes+3>>>0],premultipliedAlpha:!!HEAP8[attributes+4>>>0],preserveDrawingBuffer:!!HEAP8[attributes+5>>>0],powerPreference:webglPowerPreferences[powerPreference],failIfMajorPerformanceCaveat:!!HEAP8[attributes+12>>>0],majorVersion:HEAP32[attr32+(16>>2)>>>0],minorVersion:HEAP32[attr32+(20>>2)>>>0],enableExtensionsByDefault:HEAP8[attributes+24>>>0],explicitSwapControl:HEAP8[attributes+25>>>0],proxyContextToMainThread:HEAP32[attr32+(28>>2)>>>0],renderViaOffscreenBackBuffer:HEAP8[attributes+32>>>0]};var canvas=findCanvasEventTarget(target);if(!canvas){return 0}if(contextAttributes.explicitSwapControl){return 0}var contextHandle=GL.createContext(canvas,contextAttributes);return contextHandle}_emscripten_webgl_do_create_context.sig="ppp";var _emscripten_webgl_create_context=_emscripten_webgl_do_create_context;_emscripten_webgl_create_context.sig="ppp";function _emscripten_webgl_do_get_current_context(){return GL.currentContext?GL.currentContext.handle:0}_emscripten_webgl_do_get_current_context.sig="p";var _emscripten_webgl_get_current_context=_emscripten_webgl_do_get_current_context;_emscripten_webgl_get_current_context.sig="p";var _emscripten_webgl_do_commit_frame=()=>{if(!GL.currentContext||!GL.currentContext.GLctx){return-3}if(!GL.currentContext.attributes.explicitSwapControl){return-3}return 0};_emscripten_webgl_do_commit_frame.sig="i";var _emscripten_webgl_commit_frame=_emscripten_webgl_do_commit_frame;_emscripten_webgl_commit_frame.sig="i";function _emscripten_webgl_make_context_current(contextHandle){contextHandle>>>=0;var success=GL.makeContextCurrent(contextHandle);return success?0:-5}_emscripten_webgl_make_context_current.sig="ip";function _emscripten_webgl_get_drawing_buffer_size(contextHandle,width,height){contextHandle>>>=0;width>>>=0;height>>>=0;var GLContext=GL.getContext(contextHandle);if(!GLContext||!GLContext.GLctx||!width||!height){return-5}HEAP32[width>>>2>>>0]=GLContext.GLctx.drawingBufferWidth;HEAP32[height>>>2>>>0]=GLContext.GLctx.drawingBufferHeight;return 0}_emscripten_webgl_get_drawing_buffer_size.sig="ippp";function _emscripten_webgl_get_context_attributes(c,a){c>>>=
```
