## Supplementary software v0.2.1 for "Virtual control arms for paediatric myopia trials: external validation of axial-elongation models": pyodide.js

```
"use strict";var loadPyodide=(()=>{var ce=Object.create;var A=Object.defineProperty;var de=Object.getOwnPropertyDescriptor;var ue=Object.getOwnPropertyNames;var fe=Object.getPrototypeOf,me=Object.prototype.hasOwnProperty;var o=(e,t)=>A(e,"name",{value:t,configurable:!0}),m=(e=>typeof require<"u"?require:typeof Proxy<"u"?new Proxy(e,{get:(t,r)=>(typeof require<"u"?require:t)[r]}):e)(function(e){if(typeof require<"u")return require.apply(this,arguments);throw Error('Dynamic require of "'+e+'" is not supported')});var pe=(e,t)=>{for(var r in t)A(e,r,{get:t[r],enumerable:!0})},H=(e,t,r,s)=>{if(t&&typeof t=="object"||typeof t=="function")for(let a of ue(t))!me.call(e,a)&&a!==r&&A(e,a,{get:()=>t[a],enumerable:!(s=de(t,a))||s.enumerable});return e};var b=(e,t,r)=>(r=e!=null?ce(fe(e)):{},H(t||!e||!e.__esModule?A(r,"default",{value:e,enumerable:!0}):r,e)),ge=e=>H(A({},"__esModule",{value:!0}),e);var V=(()=>{for(var e=new Uint8Array(128),t=0;t<64;t++)e[t<26?t+65:t<52?t+71:t<62?t-4:t*4-205]=t;return r=>{for(var s=r.length,a=new Uint8Array((s-(r[s-1]=="=")-(r[s-2]=="="))*3/4|0),n=0,i=0;n<s;){var l=e[r.charCodeAt(n++)],c=e[r.charCodeAt(n++)],d=e[r.charCodeAt(n++)],u=e[r.charCodeAt(n++)];a[i++]=l<<2|c>>4,a[i++]=c<<4|d>>2,a[i++]=d<<6|u}return a}})();var Me={};pe(Me,{loadPyodide:()=>j,version:()=>L});function ye(e){return!isNaN(parseFloat(e))&&isFinite(e)}o(ye,"_isNumber");function E(e){return e.charAt(0).toUpperCase()+e.substring(1)}o(E,"_capitalize");function U(e){return function(){return this[e]}}o(U,"_getter");var k=["isConstructor","isEval","isNative","isToplevel"],I=["columnNumber","lineNumber"],N=["fileName","functionName","source"],be=["args"],ve=["evalOrigin"],T=k.concat(I,N,be,ve);function g(e){if(e)for(var t=0;t<T.length;t++)e[T[t]]!==void 0&&this["set"+E(T[t])](e[T[t]])}o(g,"StackFrame");g.prototype={getArgs:o(function(){return this.args},"getArgs"),setArgs:o(function(e){if(Object.prototype.toString.call(e)!=="[object Array]")throw new TypeError("Args must be an Array");this.args=e},"setArgs"),getEvalOrigin:o(function(){return this.evalOrigin},"getEvalOrigin"),setEvalOrigin:o(function(e){if(e instanceof g)this.evalOrigin=e;else if(e instanceof Object)this.evalOrigin=new g(e);else throw new TypeError("Eval Origin must be an Object or StackFrame")},"setEvalOrigin"),toString:o(function(){var e=this.getFileName()||"",t=this.getLineNumber()||"",r=this.getColumnNumber()||"",s=this.getFunctionName()||"";return this.getIsEval()?e?"[eval] ("+e+":"+t+":"+r+")":"[eval]:"+t+":"+r:s?s+" ("+e+":"+t+":"+r+")":e+":"+t+":"+r},"toString")};g.fromString=o(function(t){var r=t.indexOf("("),s=t.lastIndexOf(")"),a=t.substring(0,r),n=t.substring(r+1,s).split(","),i=t.substring(s+1);if(i.indexOf("@")===0)var l=/@(.+?)(?::(\d+))?(?::(\d+))?$/.exec(i,""),c=l[1],d=l[2],u=l[3];return new g({functionName:a,args:n||void 0,fileName:c,lineNumber:d||void 0,columnNumber:u||void 0})},"StackFrame$$fromString");for(h=0;h<k.length;h++)g.prototype["get"+E(k[h])]=U(k[h]),g.prototype["set"+E(k[h])]=function(e){return function(t){this[e]=!!t}}(k[h]);var h;for(S=0;S<I.length;S++)g.prototype["get"+E(I[S])]=U(I[S]),g.prototype["set"+E(I[S])]=function(e){return function(t){if(!ye(t))throw new TypeError(e+" must be a Number");this[e]=Number(t)}}(I[S]);var S;for(w=0;w<N.length;w++)g.prototype["get"+E(N[w])]=U(N[w]),g.prototype["set"+E(N[w])]=function(e){return function(t){this[e]=String(t)}}(N[w]);var w,D=g;function he(){var e=/^\s*at .*(\S+:\d+|\(native\))/m,t=/^(eval@)?(\[native code])?$/;return{parse:o(function(s){if(s.stack&&s.stack.match(e))return this.parseV8OrIE(s);if(s.stack)return this.parseFFOrSafari(s);throw new Error("Cannot parse given Error object")},"ErrorStackParser$$parse"),extractLocation:o(function(s){if(s.indexOf(":")===-1)return[s];var a=/(.+?)(?::(\d+))?(?::(\d+))?$/,n=a.exec(s.replace(/[()]/g,""));return[n[1],n[2]||void 0,n[3]||void 0]},"ErrorStackParser$$extractLocation"),parseV8OrIE:o(function(s){var a=s.stack.split(`
`).filter(function(n){return!!n.match(e)},this);return a.map(function(n){n.indexOf("(eval ")>-1&&(n=n.replace(/eval code/g,"eval").replace(/(\(eval at [^()]*)|(,.*$)/g,""));var i=n.replace(/^\s+/,"").replace(/\(eval code/g,"(").replace(/^.*?\s+/,""),l=i.match(/ (\(.+\)$)/);i=l?i.replace(l[0],""):i;var c=this.extractLocation(l?l[1]:i),d=l&&i||void 0,u=["eval","<anonymous>"].indexOf(c[0])>-1?void 0:c[0];return new D({functionName:d,fileName:u,lineNumber:c[1],columnNumber:c[2],source:n})},this)},"ErrorStackParser$$parseV8OrIE"),parseFFOrSafari:o(function(s){var a=s.stack.split(`
`).filter(function(n){return!n.match(t)},this);return a.map(function(n){if(n.indexOf(" > eval")>-1&&(n=n.replace(/ line (\d+)(?: > eval line \d+)* > eval:\d+:\d+/g,":$1")),n.indexOf("@")===-1&&n.indexOf(":")===-1)return new D({functionName:n});var i=/((.*".+"[^@]*)?[^@]*)(?:@)/,l=n.match(i),c=l&&l[1]?l[1]:void 0,d=this.extractLocation(n.replace(i,""));return new D({functionName:c,fileName:d[0],lineNumber:d[1],columnNumber:d[2],source:n})},this)},"ErrorStackParser$$parseFFOrSafari")}}o(he,"ErrorStackParser");var Se=new he;var z=Se;var y=typeof process=="object"&&typeof process.versions=="object"&&typeof process.versions.node=="string"&&!process.browser,C=y&&typeof module<"u"&&typeof module.exports<"u"&&typeof m<"u"&&typeof __dirname<"u",q=y&&!C,ze=typeof globalThis.Bun<"u",we=typeof Deno<"u",J=!y&&!we,Y=J&&typeof window=="object"&&typeof document=="object"&&typeof document.createElement=="function"&&"sessionStorage"in window&&typeof importScripts!="function",K=J&&typeof importScripts=="function"&&typeof self=="object",qe=typeof navigator=="object"&&typeof navigator.userAgent=="string"&&navigator.userAgent.indexOf("Chrome")==-1&&navigator.userAgent.indexOf("Safari")>-1,R=typeof read=="function"&&typeof load=="function";var X,M,Q,G,B;async function W(){if(!y||(X=(await import(/* webpackIgnore */"node:url")).default,G=await import(/* webpackIgnore */"node:fs"),B=await import(/* webpackIgnore */"node:fs/promises"),Q=(await import(/* webpackIgnore */"node:vm")).default,M=await import(/* webpackIgnore */"node:path"),$=M.sep,typeof m<"u"))return;let e=G,t=await import(/* webpackIgnore */"node:crypto"),r=await import(/* webpackIgnore */"ws"),s=await import(/* webpackIgnore */"node:child_process"),a={fs:e,crypto:t,ws:r,child_process:s};globalThis.require=function(n){return a[n]}}o(W,"initNodeModules");function Ee(e,t){return M.resolve(t||".",e)}o(Ee,"node_resolvePath");function Pe(e,t){return t===void 0&&(t=location),new URL(e,t).toString()}o(Pe,"browser_resolvePath");var x;y?x=Ee:R?x=o(e=>e,"resolvePath"):x=Pe;var $;y||($="/");function ke(e,t){return e.startsWith("file://")&&(e=e.slice(7)),e.includes("://")?{response:fetch(e)}:{binary:B.readFile(e).then(r=>new Uint8Array(r.buffer,r.byteOffset,r.byteLength))}}o(ke,"node_getBinaryResponse");function Ie(e,t){if(e.startsWith("file://")&&(e=e.slice(7)),e.includes("://"))throw new Error("Shell cannot fetch urls");return{binary:Promise.resolve(new Uint8Array(readbuffer(e)))}}o(Ie,"shell_getBinaryResponse");function Ne(e,t){let r=new URL(e,location);return{response:fetch(r,t?{integrity:t}:{})}}o(Ne,"browser_getBinaryResponse");var _;y?_=ke:R?_=Ie:_=Ne;async function Z(e,t){let{response:r,binary:s}=_(e,t);if(s)return s;let a=await r;if(!a.ok)throw new Error(`Failed to load '${e}': request failed.`);return new Uint8Array(await a.arrayBuffer())}o(Z,"loadBinaryFile");var F;if(Y)F=o(async e=>await import(/* webpackIgnore */e),"loadScript");else if(K)F=o(async e=>{try{globalThis.importScripts(e)}catch(t){if(t instanceof TypeError)await import(/* webpackIgnore */e);else throw t}},"loadScript");else if(y)F=Fe;else if(R)F=load;else throw new Error("Cannot determine runtime environment");async function Fe(e){e.startsWith("file://")&&(e=e.slice(7)),e.includes("://")?Q.runInThisContext(await(await fetch(e)).text()):await import(/* webpackIgnore */X.pathToFileURL(e).href)}o(Fe,"nodeLoadScript");async function ee(e){if(y){await W();let t=await B.readFile(e,{encoding:"utf8"});return JSON.parse(t)}else if(R){let t=read(e);return JSON.parse(t)}else return await(await fetch(e)).json()}o(ee,"loadLockFile");async function te(){if(C)return __dirname;let e;try{throw new Error}catch(s){e=s}let t=z.parse(e)[0].fileName;if(y&&!t.startsWith("file://")&&(t=`file://${t}`),q){let s=await import(/* webpackIgnore */"node:path");return(await import(/* webpackIgnore */"node:url")).fileURLToPath(s.dirname(t))}let r=t.lastIndexOf($);if(r===-1)throw new Error("Could not extract indexURL path from pyodide module location");return t.slice(0,r)}o(te,"calculateDirname");function ne(e){return e.substring(0,e.lastIndexOf("/")+1)||globalThis.location?.toString()||"."}o(ne,"calculateInstallBaseUrl");function re(e){let t=e.FS,r=e.FS.filesystems.MEMFS,s=e.PATH,a={DIR_MODE:16895,FILE_MODE:33279,mount:o(function(n){if(!n.opts.fileSystemHandle)throw new Error("opts.fileSystemHandle is required");return r.mount.apply(null,arguments)},"mount"),syncfs:o(async(n,i,l)=>{try{let c=a.getLocalSet(n),d=await a.getRemoteSet(n),u=i?d:c,f=i?c:d;await a.reconcile(n,u,f),l(null)}catch(c){l(c)}},"syncfs"),getLocalSet:o(n=>{let i=Object.create(null);function l(u){return u!=="."&&u!==".."}o(l,"isRealDir");function c(u){return f=>s.join2(u,f)}o(c,"toAbsolute");let d=t.readdir(n.mountpoint).filter(l).map(c(n.mountpoint));for(;d.length;){let u=d.pop(),f=t.stat(u);t.isDir(f.mode)&&d.push.apply(d,t.readdir(u).filter(l).map(c(u))),i[u]={timestamp:f.mtime,mode:f.mode}}return{type:"local",entries:i}},"getLocalSet"),getRemoteSet:o(async n=>{let i=Object.create(null),l=await xe(n.opts.fileSystemHandle);for(let[c,d]of l)c!=="."&&(i[s.join2(n.mountpoint,c)]={timestamp:d.kind==="file"?new Date((await d.getFile()).lastModified):new Date,mode:d.kind==="file"?a.FILE_MODE:a.DIR_MODE});return{type:"remote",entries:i,handles:l}},"getRemoteSet"),loadLocalEntry:o(n=>{let l=t.lookupPath(n).node,c=t.stat(n);if(t.isDir(c.mode))return{timestamp:c.mtime,mode:c.mode};if(t.isFile(c.mode))return l.contents=r.getFileDataAsTypedArray(l),{timestamp:c.mtime,mode:c.mode,contents:l.contents};throw new Error("node type not supported")},"loadLocalEntry"),storeLocalEntry:o((n,i)=>{if(t.isDir(i.mode))t.mkdirTree(n,i.mode);else if(t.isFile(i.mode))t.writeFile(n,i.contents,{canOwn:!0});else throw new Error("node type not supported");t.chmod(n,i.mode),t.utime(n,i.timestamp,i.timestamp)},"storeLocalEntry"),removeLocalEntry:o(n=>{var i=t.stat(n);t.isDir(i.mode)?t.rmdir(n):t.isFile(i.mode)&&t.unlink(n)},"removeLocalEntry"),loadRemoteEntry:o(async n=>{if(n.kind==="file"){let i=await n.getFile();return{contents:new Uint8Array(await i.arrayBuffer()),mode:a.FILE_MODE,timestamp:new Date(i.lastModified)}}else{if(n.kind==="directory")return{mode:a.DIR_MODE,timestamp:new Date};throw new Error("unknown kind: "+n.kind)}},"loadRemoteEntry"),storeRemoteEntry:o(async(n,i,l)=>{let c=n.get(s.dirname(i)),d=t.isFile(l.mode)?await c.getFileHandle(s.basename(i),{create:!0}):await c.getDirectoryHandle(s.basename(i),{create:!0});if(d.kind==="file"){let u=await d.createWritable();await u.write(l.contents),await u.close()}n.set(i,d)},"storeRemoteEntry"),removeRemoteEntry:o(async(n,i)=>{await n.get(s.dirname(i)).removeEntry(s.basename(i)),n.delete(i)},"removeRemoteEntry"),reconcile:o(async(n,i,l)=>{let c=0,d=[];Object.keys(i.entries).forEach(function(p){let v=i.entries[p],P=l.entries[p];(!P||t.isFile(v.mode)&&v.timestamp.getTime()>P.timestamp.getTime())&&(d.push(p),c++)}),d.sort();let u=[];if(Object.keys(l.entries).forEach(function(p){i.entries[p]||(u.push(p),c++)}),u.sort().reverse(),!c)return;let f=i.type==="remote"?i.handles:l.handles;for(let p of d){let v=s.normalize(p.replace(n.mountpoint,"/")).substring(1);if(l.type==="local"){let P=f.get(v),le=await a.loadRemoteEntry(P);a.storeLocalEntry(p,le)}else{let P=a.loadLocalEntry(p);await a.storeRemoteEntry(f,v,P)}}for(let p of u)if(l.type==="local")a.removeLocalEntry(p);else{let v=s.normalize(p.replace(n.mountpoint,"/")).substring(1);await a.removeRemoteEntry(f,v)}},"reconcile")};e.FS.filesystems.NATIVEFS_ASYNC=a}o(re,"initializeNativeFS");var xe=o(async e=>{let t=[];async function r(a){for await(let n of a.values())t.push(n),n.kind==="directory"&&await r(n)}o(r,"collect"),await r(e);let s=new Map;s.set(".",e);for(let a of t){let n=(await e.resolve(a)).join("/");s.set(n,a)}return s},"getFsHandles");var ie=V("AGFzbQEAAAABDANfAGAAAW9gAW8BfwMDAgECByECD2NyZWF0ZV9zZW50aW5lbAAAC2lzX3NlbnRpbmVsAAEKEwIHAPsBAPsbCwkAIAD7GvsUAAs=");var Ae=async function(){if(!(globalThis.navigator&&(/iPad|iPhone|iPod/.test(navigator.userAgent)||navigator.platform==="MacIntel"&&typeof navigator.maxTouchPoints<"u"&&navigator.maxTouchPoints>1)))try{let t=await WebAssembly.compile(ie);return await WebAssembly.instantiate(t)}catch(t){if(t instanceof WebAssembly.CompileError)return;throw t}}();async function oe(){let e=await Ae;if(e)return e.exports;let t=Symbol("error marker");return{create_sentinel:o(()=>t,"create_sentinel"),is_sentinel:o(r=>r===t,"is_sentinel")}}o(oe,"getSentinelImport");function se(e){let t={noImageDecoding:!0,noAudioDecoding:!0,noWasmDecoding:!1,preRun:Ue(e),print:e.stdout,printErr:e.stderr,onExit(r){t.exitCode=r},thisProgram:e._sysExecutable,arguments:e.args,API:{config:e},locateFile:o(r=>e.indexURL+r,"locateFile"),instantiateWasm:Ce(e.indexURL)};return t}o(se,"createSettings");function Re(e){return function(t){let r="/";try{t.FS.mkdirTree(e)}catch(s){console.error(`Error occurred while making a home directory '${e}':`),console.error(s),console.error(`Using '${r}' for a home directory instead`),e=r}t.FS.chdir(e)}}o(Re,"createHomeDirectory");function Oe(e){return function(t){Object.assign(t.ENV,e)}}o(Oe,"setEnvironment");function Le(e){return e?[async t=>{t.addRunDependency("fsInitHook");try{await e(t.FS,{sitePackages:t.API.sitePackages})}finally{t.removeRunDependency("fsInitHook")}}]:[]}o(Le,"callFsInitHook");function Te(e){let t=e.HEAPU32[e._Py_Version>>>2],r=t>>>24&255,s=t>>>16&255,a=t>>>8&255;return[r,s,a]}o(Te,"computeVersionTuple");function De(e){let t=Z(e);return async r=>{r.API.pyVersionTuple=Te(r);let[s,a]=r.API.pyVersionTuple;r.FS.mkdirTree("/lib"),r.API.sitePackages=`/lib/python${s}.${a}/site-packages`,r.FS.mkdirTree(r.API.sitePackages),r.addRunDependency("install-stdlib");try{let n=await t;r.FS.writeFile(`/lib/python${s}${a}.zip`,n)}catch(n){console.error("Error occurred while installing the standard library:"),console.error(n)}finally{r.removeRunDependency("install-stdlib")}}}o(De,"installStdlib");function Ue(e){let t;return e.stdLibURL!=null?t=e.stdLibURL:t=e.indexURL+"python_stdlib.zip",[De(t),Re(e.env.HOME),Oe(e.env),re,...Le(e.fsInit)]}o(Ue,"getFileSystemInitializationFuncs");function Ce(e){if(typeof WasmOffsetConverter<"u")return;let{binary:t,response:r}=_(e+"pyodide.asm.wasm"),s=oe();return function(a,n){return async function(){a.sentinel=await s;try{let i;r?i=await WebAssembly.instantiateStreaming(r,a):i=await WebAssembly.instantiate(await t,a);let{instance:l,module:c}=i;n(l,c)}catch(i){console.warn("wasm instantiation failed!"),console.warn(i)}}(),{}}}o(Ce,"getInstantiateWasmFunc");var ae="0.28.3";function O(e){return e===void 0||e.endsWith("/")?e:e+"/"}o(O,"withTrailingSlash");var L=ae;async function j(e={}){if(e.lockFileContents&&e.lockFileURL)throw new Error("Can't pass both lockFileContents and lockFileURL");await W();let t=e.indexURL||await te();t=O(x(t));let r=e;if(r.packageBaseUrl=O(r.packageBaseUrl),r.cdnUrl=O(r.packageBaseUrl??`https://cdn.jsdelivr.net/pyodide/v${L}/full/`),!e.lockFileContents){let f=e.lockFileURL??t+"pyodide-lock.json";r.lockFileContents=ee(f),r.packageBaseUrl??=ne(f)}r.indexURL=t,r.packageCacheDir&&(r.packageCacheDir=O(x(r.packageCacheDir)));let s={fullStdLib:!1,jsglobals:globalThis,stdin:globalThis.prompt?globalThis.prompt:void 0,args:[],env:{},packages:[],packageCacheDir:r.packageBaseUrl,enableRunUntilComplete:!0,checkAPIVersion:!0,BUILD_ID:"cc7a4bb4c6f36f12592fef0934292b5fddb0e313ca5dc4a5a9519bb7610c67e3"},a=Object.assign(s,r);a.env.HOME??="/home/pyodide",a.env.PYTHONINSPECT??="1";let n=se(a),i=n.API;if(i.lockFilePromise=Promise.resolve(r.lockFileContents),typeof _createPyodideModule!="function"){let f=`${a.indexURL}pyodide.asm.js`;await F(f)}let l;if(e._loadSnapshot){let f=await e._loadSnapshot;ArrayBuffer.isView(f)?l=f:l=new Uint8Array(f),n.noInitialRun=!0,n.INITIAL_MEMORY=l.length}let c=await _createPyodideModule(n);if(n.exitCode!==void 0)throw new c.ExitStatus(n.exitCode);if(e.pyproxyToStringRepr&&i.setPyProxyToStringMethod(!0),e.convertNullToNone&&i.setCompatNullToNone(!0),i.version!==L&&a.checkAPIVersion)throw new Error(`Pyodide version does not match: '${L}' <==> '${i.version}'. If you updated the Pyodide version, make sure you also updated the 'indexURL' parameter passed to loadPyodide.`);c.locateFile=f=>{throw f.endsWith(".so")?new Error(`Failed to find dynamic library "${f}"`):new Error(`Unexpected call to locateFile("${f}")`)};let d;l&&(d=i.restoreSnapshot(l));let u=i.finalizeBootstrap(d,e._snapshotDeserializer);return i.sys.path.insert(0,""),i._pyodide.set_excepthook(),await i.packageIndexReady,i.initializeStreams(a.stdin,a.stdout,a.stderr),u}o(j,"loadPyodide");globalThis.loadPyodide=j;return ge(Me);})();
try{Object.assign(exports,loadPyodide)}catch(_){}
globalThis.loadPyodide=loadPyodide.loadPyodide;
//# sourceMappingURL=pyodide.js.map
```
