## Supplementary software v0.2.1 for "Virtual control arms for paediatric myopia trials: external validation of axial-elongation models": pyodide.mjs

```
var oe=Object.defineProperty;var o=(e,t)=>oe(e,"name",{value:t,configurable:!0}),A=(e=>typeof require<"u"?require:typeof Proxy<"u"?new Proxy(e,{get:(t,r)=>(typeof require<"u"?require:t)[r]}):e)(function(e){if(typeof require<"u")return require.apply(this,arguments);throw Error('Dynamic require of "'+e+'" is not supported')});var W=(()=>{for(var e=new Uint8Array(128),t=0;t<64;t++)e[t<26?t+65:t<52?t+71:t<62?t-4:t*4-205]=t;return r=>{for(var s=r.length,a=new Uint8Array((s-(r[s-1]=="=")-(r[s-2]=="="))*3/4|0),n=0,i=0;n<s;){var c=e[r.charCodeAt(n++)],l=e[r.charCodeAt(n++)],d=e[r.charCodeAt(n++)],u=e[r.charCodeAt(n++)];a[i++]=c<<2|l>>4,a[i++]=l<<4|d>>2,a[i++]=d<<6|u}return a}})();function se(e){return!isNaN(parseFloat(e))&&isFinite(e)}o(se,"_isNumber");function S(e){return e.charAt(0).toUpperCase()+e.substring(1)}o(S,"_capitalize");function L(e){return function(){return this[e]}}o(L,"_getter");var E=["isConstructor","isEval","isNative","isToplevel"],P=["columnNumber","lineNumber"],k=["fileName","functionName","source"],ae=["args"],ce=["evalOrigin"],R=E.concat(P,k,ae,ce);function p(e){if(e)for(var t=0;t<R.length;t++)e[R[t]]!==void 0&&this["set"+S(R[t])](e[R[t]])}o(p,"StackFrame");p.prototype={getArgs:o(function(){return this.args},"getArgs"),setArgs:o(function(e){if(Object.prototype.toString.call(e)!=="[object Array]")throw new TypeError("Args must be an Array");this.args=e},"setArgs"),getEvalOrigin:o(function(){return this.evalOrigin},"getEvalOrigin"),setEvalOrigin:o(function(e){if(e instanceof p)this.evalOrigin=e;else if(e instanceof Object)this.evalOrigin=new p(e);else throw new TypeError("Eval Origin must be an Object or StackFrame")},"setEvalOrigin"),toString:o(function(){var e=this.getFileName()||"",t=this.getLineNumber()||"",r=this.getColumnNumber()||"",s=this.getFunctionName()||"";return this.getIsEval()?e?"[eval] ("+e+":"+t+":"+r+")":"[eval]:"+t+":"+r:s?s+" ("+e+":"+t+":"+r+")":e+":"+t+":"+r},"toString")};p.fromString=o(function(t){var r=t.indexOf("("),s=t.lastIndexOf(")"),a=t.substring(0,r),n=t.substring(r+1,s).split(","),i=t.substring(s+1);if(i.indexOf("@")===0)var c=/@(.+?)(?::(\d+))?(?::(\d+))?$/.exec(i,""),l=c[1],d=c[2],u=c[3];return new p({functionName:a,args:n||void 0,fileName:l,lineNumber:d||void 0,columnNumber:u||void 0})},"StackFrame$$fromString");for(b=0;b<E.length;b++)p.prototype["get"+S(E[b])]=L(E[b]),p.prototype["set"+S(E[b])]=function(e){return function(t){this[e]=!!t}}(E[b]);var b;for(v=0;v<P.length;v++)p.prototype["get"+S(P[v])]=L(P[v]),p.prototype["set"+S(P[v])]=function(e){return function(t){if(!se(t))throw new TypeError(e+" must be a Number");this[e]=Number(t)}}(P[v]);var v;for(h=0;h<k.length;h++)p.prototype["get"+S(k[h])]=L(k[h]),p.prototype["set"+S(k[h])]=function(e){return function(t){this[e]=String(t)}}(k[h]);var h,O=p;function le(){var e=/^\s*at .*(\S+:\d+|\(native\))/m,t=/^(eval@)?(\[native code])?$/;return{parse:o(function(s){if(s.stack&&s.stack.match(e))return this.parseV8OrIE(s);if(s.stack)return this.parseFFOrSafari(s);throw new Error("Cannot parse given Error object")},"ErrorStackParser$$parse"),extractLocation:o(function(s){if(s.indexOf(":")===-1)return[s];var a=/(.+?)(?::(\d+))?(?::(\d+))?$/,n=a.exec(s.replace(/[()]/g,""));return[n[1],n[2]||void 0,n[3]||void 0]},"ErrorStackParser$$extractLocation"),parseV8OrIE:o(function(s){var a=s.stack.split(`
`).filter(function(n){return!!n.match(e)},this);return a.map(function(n){n.indexOf("(eval ")>-1&&(n=n.replace(/eval code/g,"eval").replace(/(\(eval at [^()]*)|(,.*$)/g,""));var i=n.replace(/^\s+/,"").replace(/\(eval code/g,"(").replace(/^.*?\s+/,""),c=i.match(/ (\(.+\)$)/);i=c?i.replace(c[0],""):i;var l=this.extractLocation(c?c[1]:i),d=c&&i||void 0,u=["eval","<anonymous>"].indexOf(l[0])>-1?void 0:l[0];return new O({functionName:d,fileName:u,lineNumber:l[1],columnNumber:l[2],source:n})},this)},"ErrorStackParser$$parseV8OrIE"),parseFFOrSafari:o(function(s){var a=s.stack.split(`
`).filter(function(n){return!n.match(t)},this);return a.map(function(n){if(n.indexOf(" > eval")>-1&&(n=n.replace(/ line (\d+)(?: > eval line \d+)* > eval:\d+:\d+/g,":$1")),n.indexOf("@")===-1&&n.indexOf(":")===-1)return new O({functionName:n});var i=/((.*".+"[^@]*)?[^@]*)(?:@)/,c=n.match(i),l=c&&c[1]?c[1]:void 0,d=this.extractLocation(n.replace(i,""));return new O({functionName:l,fileName:d[0],lineNumber:d[1],columnNumber:d[2],source:n})},this)},"ErrorStackParser$$parseFFOrSafari")}}o(le,"ErrorStackParser");var de=new le;var $=de;var g=typeof process=="object"&&typeof process.versions=="object"&&typeof process.versions.node=="string"&&!process.browser,T=g&&typeof module<"u"&&typeof module.exports<"u"&&typeof A<"u"&&typeof __dirname<"u",j=g&&!T,Te=typeof globalThis.Bun<"u",ue=typeof Deno<"u",H=!g&&!ue,V=H&&typeof window=="object"&&typeof document=="object"&&typeof document.createElement=="function"&&"sessionStorage"in window&&typeof importScripts!="function",z=H&&typeof importScripts=="function"&&typeof self=="object",De=typeof navigator=="object"&&typeof navigator.userAgent=="string"&&navigator.userAgent.indexOf("Chrome")==-1&&navigator.userAgent.indexOf("Safari")>-1,x=typeof read=="function"&&typeof load=="function";var J,D,Y,q,U;async function C(){if(!g||(J=(await import("node:url")).default,q=await import("node:fs"),U=await import("node:fs/promises"),Y=(await import("node:vm")).default,D=await import("node:path"),M=D.sep,typeof A<"u"))return;let e=q,t=await import("node:crypto"),r=await import("ws"),s=await import("node:child_process"),a={fs:e,crypto:t,ws:r,child_process:s};globalThis.require=function(n){return a[n]}}o(C,"initNodeModules");function fe(e,t){return D.resolve(t||".",e)}o(fe,"node_resolvePath");function me(e,t){return t===void 0&&(t=location),new URL(e,t).toString()}o(me,"browser_resolvePath");var N;g?N=fe:x?N=o(e=>e,"resolvePath"):N=me;var M;g||(M="/");function pe(e,t){return e.startsWith("file://")&&(e=e.slice(7)),e.includes("://")?{response:fetch(e)}:{binary:U.readFile(e).then(r=>new Uint8Array(r.buffer,r.byteOffset,r.byteLength))}}o(pe,"node_getBinaryResponse");function ge(e,t){if(e.startsWith("file://")&&(e=e.slice(7)),e.includes("://"))throw new Error("Shell cannot fetch urls");return{binary:Promise.resolve(new Uint8Array(readbuffer(e)))}}o(ge,"shell_getBinaryResponse");function ye(e,t){let r=new URL(e,location);return{response:fetch(r,t?{integrity:t}:{})}}o(ye,"browser_getBinaryResponse");var F;g?F=pe:x?F=ge:F=ye;async function K(e,t){let{response:r,binary:s}=F(e,t);if(s)return s;let a=await r;if(!a.ok)throw new Error(`Failed to load '${e}': request failed.`);return new Uint8Array(await a.arrayBuffer())}o(K,"loadBinaryFile");var I;if(V)I=o(async e=>await import(e),"loadScript");else if(z)I=o(async e=>{try{globalThis.importScripts(e)}catch(t){if(t instanceof TypeError)await import(e);else throw t}},"loadScript");else if(g)I=be;else if(x)I=load;else throw new Error("Cannot determine runtime environment");async function be(e){e.startsWith("file://")&&(e=e.slice(7)),e.includes("://")?Y.runInThisContext(await(await fetch(e)).text()):await import(J.pathToFileURL(e).href)}o(be,"nodeLoadScript");async function G(e){if(g){await C();let t=await U.readFile(e,{encoding:"utf8"});return JSON.parse(t)}else if(x){let t=read(e);return JSON.parse(t)}else return await(await fetch(e)).json()}o(G,"loadLockFile");async function X(){if(T)return __dirname;let e;try{throw new Error}catch(s){e=s}let t=$.parse(e)[0].fileName;if(g&&!t.startsWith("file://")&&(t=`file://${t}`),j){let s=await import("node:path");return(await import("node:url")).fileURLToPath(s.dirname(t))}let r=t.lastIndexOf(M);if(r===-1)throw new Error("Could not extract indexURL path from pyodide module location");return t.slice(0,r)}o(X,"calculateDirname");function Q(e){return e.substring(0,e.lastIndexOf("/")+1)||globalThis.location?.toString()||"."}o(Q,"calculateInstallBaseUrl");function Z(e){let t=e.FS,r=e.FS.filesystems.MEMFS,s=e.PATH,a={DIR_MODE:16895,FILE_MODE:33279,mount:o(function(n){if(!n.opts.fileSystemHandle)throw new Error("opts.fileSystemHandle is required");return r.mount.apply(null,arguments)},"mount"),syncfs:o(async(n,i,c)=>{try{let l=a.getLocalSet(n),d=await a.getRemoteSet(n),u=i?d:l,f=i?l:d;await a.reconcile(n,u,f),c(null)}catch(l){c(l)}},"syncfs"),getLocalSet:o(n=>{let i=Object.create(null);function c(u){return u!=="."&&u!==".."}o(c,"isRealDir");function l(u){return f=>s.join2(u,f)}o(l,"toAbsolute");let d=t.readdir(n.mountpoint).filter(c).map(l(n.mountpoint));for(;d.length;){let u=d.pop(),f=t.stat(u);t.isDir(f.mode)&&d.push.apply(d,t.readdir(u).filter(c).map(l(u))),i[u]={timestamp:f.mtime,mode:f.mode}}return{type:"local",entries:i}},"getLocalSet"),getRemoteSet:o(async n=>{let i=Object.create(null),c=await ve(n.opts.fileSystemHandle);for(let[l,d]of c)l!=="."&&(i[s.join2(n.mountpoint,l)]={timestamp:d.kind==="file"?new Date((await d.getFile()).lastModified):new Date,mode:d.kind==="file"?a.FILE_MODE:a.DIR_MODE});return{type:"remote",entries:i,handles:c}},"getRemoteSet"),loadLocalEntry:o(n=>{let c=t.lookupPath(n).node,l=t.stat(n);if(t.isDir(l.mode))return{timestamp:l.mtime,mode:l.mode};if(t.isFile(l.mode))return c.contents=r.getFileDataAsTypedArray(c),{timestamp:l.mtime,mode:l.mode,contents:c.contents};throw new Error("node type not supported")},"loadLocalEntry"),storeLocalEntry:o((n,i)=>{if(t.isDir(i.mode))t.mkdirTree(n,i.mode);else if(t.isFile(i.mode))t.writeFile(n,i.contents,{canOwn:!0});else throw new Error("node type not supported");t.chmod(n,i.mode),t.utime(n,i.timestamp,i.timestamp)},"storeLocalEntry"),removeLocalEntry:o(n=>{var i=t.stat(n);t.isDir(i.mode)?t.rmdir(n):t.isFile(i.mode)&&t.unlink(n)},"removeLocalEntry"),loadRemoteEntry:o(async n=>{if(n.kind==="file"){let i=await n.getFile();return{contents:new Uint8Array(await i.arrayBuffer()),mode:a.FILE_MODE,timestamp:new Date(i.lastModified)}}else{if(n.kind==="directory")return{mode:a.DIR_MODE,timestamp:new Date};throw new Error("unknown kind: "+n.kind)}},"loadRemoteEntry"),storeRemoteEntry:o(async(n,i,c)=>{let l=n.get(s.dirname(i)),d=t.isFile(c.mode)?await l.getFileHandle(s.basename(i),{create:!0}):await l.getDirectoryHandle(s.basename(i),{create:!0});if(d.kind==="file"){let u=await d.createWritable();await u.write(c.contents),await u.close()}n.set(i,d)},"storeRemoteEntry"),removeRemoteEntry:o(async(n,i)=>{await n.get(s.dirname(i)).removeEntry(s.basename(i)),n.delete(i)},"removeRemoteEntry"),reconcile:o(async(n,i,c)=>{let l=0,d=[];Object.keys(i.entries).forEach(function(m){let y=i.entries[m],w=c.entries[m];(!w||t.isFile(y.mode)&&y.timestamp.getTime()>w.timestamp.getTime())&&(d.push(m),l++)}),d.sort();let u=[];if(Object.keys(c.entries).forEach(function(m){i.entries[m]||(u.push(m),l++)}),u.sort().reverse(),!l)return;let f=i.type==="remote"?i.handles:c.handles;for(let m of d){let y=s.normalize(m.replace(n.mountpoint,"/")).substring(1);if(c.type==="local"){let w=f.get(y),ie=await a.loadRemoteEntry(w);a.storeLocalEntry(m,ie)}else{let w=a.loadLocalEntry(m);await a.storeRemoteEntry(f,y,w)}}for(let m of u)if(c.type==="local")a.removeLocalEntry(m);else{let y=s.normalize(m.replace(n.mountpoint,"/")).substring(1);await a.removeRemoteEntry(f,y)}},"reconcile")};e.FS.filesystems.NATIVEFS_ASYNC=a}o(Z,"initializeNativeFS");var ve=o(async e=>{let t=[];async function r(a){for await(let n of a.values())t.push(n),n.kind==="directory"&&await r(n)}o(r,"collect"),await r(e);let s=new Map;s.set(".",e);for(let a of t){let n=(await e.resolve(a)).join("/");s.set(n,a)}return s},"getFsHandles");var ee=W("AGFzbQEAAAABDANfAGAAAW9gAW8BfwMDAgECByECD2NyZWF0ZV9zZW50aW5lbAAAC2lzX3NlbnRpbmVsAAEKEwIHAPsBAPsbCwkAIAD7GvsUAAs=");var Se=async function(){if(!(globalThis.navigator&&(/iPad|iPhone|iPod/.test(navigator.userAgent)||navigator.platform==="MacIntel"&&typeof navigator.maxTouchPoints<"u"&&navigator.maxTouchPoints>1)))try{let t=await WebAssembly.compile(ee);return await WebAssembly.instantiate(t)}catch(t){if(t instanceof WebAssembly.CompileError)return;throw t}}();async function te(){let e=await Se;if(e)return e.exports;let t=Symbol("error marker");return{create_sentinel:o(()=>t,"create_sentinel"),is_sentinel:o(r=>r===t,"is_sentinel")}}o(te,"getSentinelImport");function ne(e){let t={noImageDecoding:!0,noAudioDecoding:!0,noWasmDecoding:!1,preRun:Ne(e),print:e.stdout,printErr:e.stderr,onExit(r){t.exitCode=r},thisProgram:e._sysExecutable,arguments:e.args,API:{config:e},locateFile:o(r=>e.indexURL+r,"locateFile"),instantiateWasm:Fe(e.indexURL)};return t}o(ne,"createSettings");function we(e){return function(t){let r="/";try{t.FS.mkdirTree(e)}catch(s){console.error(`Error occurred while making a home directory '${e}':`),console.error(s),console.error(`Using '${r}' for a home directory instead`),e=r}t.FS.chdir(e)}}o(we,"createHomeDirectory");function Ee(e){return function(t){Object.assign(t.ENV,e)}}o(Ee,"setEnvironment");function Pe(e){return e?[async t=>{t.addRunDependency("fsInitHook");try{await e(t.FS,{sitePackages:t.API.sitePackages})}finally{t.removeRunDependency("fsInitHook")}}]:[]}o(Pe,"callFsInitHook");function ke(e){let t=e.HEAPU32[e._Py_Version>>>2],r=t>>>24&255,s=t>>>16&255,a=t>>>8&255;return[r,s,a]}o(ke,"computeVersionTuple");function Ie(e){let t=K(e);return async r=>{r.API.pyVersionTuple=ke(r);let[s,a]=r.API.pyVersionTuple;r.FS.mkdirTree("/lib"),r.API.sitePackages=`/lib/python${s}.${a}/site-packages`,r.FS.mkdirTree(r.API.sitePackages),r.addRunDependency("install-stdlib");try{let n=await t;r.FS.writeFile(`/lib/python${s}${a}.zip`,n)}catch(n){console.error("Error occurred while installing the standard library:"),console.error(n)}finally{r.removeRunDependency("install-stdlib")}}}o(Ie,"installStdlib");function Ne(e){let t;return e.stdLibURL!=null?t=e.stdLibURL:t=e.indexURL+"python_stdlib.zip",[Ie(t),we(e.env.HOME),Ee(e.env),Z,...Pe(e.fsInit)]}o(Ne,"getFileSystemInitializationFuncs");function Fe(e){if(typeof WasmOffsetConverter<"u")return;let{binary:t,response:r}=F(e+"pyodide.asm.wasm"),s=te();return function(a,n){return async function(){a.sentinel=await s;try{let i;r?i=await WebAssembly.instantiateStreaming(r,a):i=await WebAssembly.instantiate(await t,a);let{instance:c,module:l}=i;n(c,l)}catch(i){console.warn("wasm instantiation failed!"),console.warn(i)}}(),{}}}o(Fe,"getInstantiateWasmFunc");var re="0.28.3";function _(e){return e===void 0||e.endsWith("/")?e:e+"/"}o(_,"withTrailingSlash");var B=re;async function at(e={}){if(e.lockFileContents&&e.lockFileURL)throw new Error("Can't pass both lockFileContents and lockFileURL");await C();let t=e.indexURL||await X();t=_(N(t));let r=e;if(r.packageBaseUrl=_(r.packageBaseUrl),r.cdnUrl=_(r.packageBaseUrl??`https://cdn.jsdelivr.net/pyodide/v${B}/full/`),!e.lockFileContents){let f=e.lockFileURL??t+"pyodide-lock.json";r.lockFileContents=G(f),r.packageBaseUrl??=Q(f)}r.indexURL=t,r.packageCacheDir&&(r.packageCacheDir=_(N(r.packageCacheDir)));let s={fullStdLib:!1,jsglobals:globalThis,stdin:globalThis.prompt?globalThis.prompt:void 0,args:[],env:{},packages:[],packageCacheDir:r.packageBaseUrl,enableRunUntilComplete:!0,checkAPIVersion:!0,BUILD_ID:"cc7a4bb4c6f36f12592fef0934292b5fddb0e313ca5dc4a5a9519bb7610c67e3"},a=Object.assign(s,r);a.env.HOME??="/home/pyodide",a.env.PYTHONINSPECT??="1";let n=ne(a),i=n.API;if(i.lockFilePromise=Promise.resolve(r.lockFileContents),typeof _createPyodideModule!="function"){let f=`${a.indexURL}pyodide.asm.js`;await I(f)}let c;if(e._loadSnapshot){let f=await e._loadSnapshot;ArrayBuffer.isView(f)?c=f:c=new Uint8Array(f),n.noInitialRun=!0,n.INITIAL_MEMORY=c.length}let l=await _createPyodideModule(n);if(n.exitCode!==void 0)throw new l.ExitStatus(n.exitCode);if(e.pyproxyToStringRepr&&i.setPyProxyToStringMethod(!0),e.convertNullToNone&&i.setCompatNullToNone(!0),i.version!==B&&a.checkAPIVersion)throw new Error(`Pyodide version does not match: '${B}' <==> '${i.version}'. If you updated the Pyodide version, make sure you also updated the 'indexURL' parameter passed to loadPyodide.`);l.locateFile=f=>{throw f.endsWith(".so")?new Error(`Failed to find dynamic library "${f}"`):new Error(`Unexpected call to locateFile("${f}")`)};let d;c&&(d=i.restoreSnapshot(c));let u=i.finalizeBootstrap(d,e._snapshotDeserializer);return i.sys.path.insert(0,""),i._pyodide.set_excepthook(),await i.packageIndexReady,i.initializeStreams(a.stdin,a.stdout,a.stderr),u}o(at,"loadPyodide");export{at as loadPyodide,B as version};
//# sourceMappingURL=pyodide.mjs.map
```
