## Supplementary software v0.2.1 for "Virtual control arms for paediatric myopia trials: external validation of axial-elongation models": README.md

# Virtual Control Arm Tool

An open, transparent tool that builds a predicted untreated ("virtual") control
arm for single-arm myopia-control studies from published axial-elongation models.
It adds no new model; it applies published models in parallel, externally
validated against an untreated multi-ethnic cohort.

This folder is the complete, citable deposit. It contains both:

- **the runnable tool**, which runs entirely in your web browser, and
- **the full source and reproduction code**, so every table and figure in the
  paper can be regenerated.

> **About this release.** The tool is delivered as a self-contained, offline
> web application (no installation, nothing leaves your machine). It replaces the
> earlier desktop executable, which has been retired. Input and output are Excel
> workbooks (`.xlsx`) throughout, in both the tool and the reproduction scripts.

**Your data never leaves your machine. The browser tool makes no network request
of any kind:** the Python runtime and all packages are vendored under `pyodide/`
and served locally. There is no server, no login, and nothing to install beyond a
way to open the page.

## Quick start (using the tool)

1. **Windows:** double-click `run_windows.bat`.
   **macOS / Linux:** run `./run_mac_linux.sh`.
2. Your browser opens at `http://localhost:8000`. Choose a model, upload your
   baseline file, and download the predicted control.

If you prefer, from this folder run `python -m http.server 8000` and open
`http://localhost:8000`. Any static file server works. Opening `index.html`
directly from the file system will not work, because the page loads its own
assets over http.

**If you do not have Python:** the launcher uses Python only to open the page in
your browser. Install Python 3 once (Windows and macOS: https://www.python.org/downloads/,
ticking "Add python.exe to PATH" on Windows; Linux: your package manager). That
one-time download is the only time the internet is used. After Python is
installed, the tool runs fully offline and never sends your data anywhere; the
tool itself makes no network request, which you can verify from the
Content-Security-Policy in `index.html`.

Click **Run self-test** in the page to confirm, in your own browser, that the
predictions match the published Naduvilath 2025 Table 4 values.

## The no-network guarantee is machine-verifiable

`index.html` sets a Content-Security-Policy of `connect-src 'self'`: the browser
blocks any attempt by the page to send data anywhere. Confirm it by reading that
one line, by watching the browser Network tab produce no request while you use the
tool, or by disconnecting from the internet and running it anyway.

## Reproducing the paper's tables and figures

The analysis code is standard Python (numpy, pandas, scipy, matplotlib, openpyxl;
see `src/`). With the governed cohort data in place, the validation and figure
scripts regenerate the paper's tables and figures under a fixed seed. Individual-
level untreated data are available subject to the contributing site's governance
and ethics approvals; the code is released in full so the analysis can be
inspected and re-run.

```
pip install numpy pandas scipy matplotlib openpyxl
cd src
python virtual_control.py      # engine self-validation (six checks vs Naduvilath 2025 Table 4)
python demo.py                 # full pipeline on synthetic data (no governed data needed)
# validate_model.py, reproduce.py, make_figs_v2.py, make_fig4.py regenerate paper artefacts
```

## Folder layout

```
index.html, app.js            the browser tool (app.js keeps the CSP strict; loads src/ unmodified)
run_windows.bat / .sh         one-click local launchers
pyodide/                      vendored Pyodide 0.28.3 runtime + packages (~21 MB)
src/                          full engine and analysis code
  virtual_control.py            the engine (predict_untreated is the core)
  sensitivity_models.py         the four-model dispatch
  validate_model.py, reproduce.py, make_figs_v2.py, make_fig4.py, ...  reproduction scripts
  gui_app.py                    optional local desktop interface (same engine; not required)
tests/test_engine.py          engine test suite (pytest)
docs/                         analysis protocol, input dictionary, model notes, sources, and more
examples/                     sample_input.xlsx and an example output workbook
LICENSE, CITATION.cff         GPL-3.0-or-later; how to cite
```

## Depositing with the paper

A Zenodo DOI is sufficient and is the recommended citation; a public code host is
optional and not required. Upload this folder (zipped) to Zenodo as a new record;
it issues a DOI on publish. Because Pyodide is vendored, the archive is complete
and self-contained, and both the tool and the reproduction code will still run
years from now with no dependency on any CDN or package index. Add the DOI to
`CITATION.cff` once minted.

## Input format

One row per child, in an Excel workbook (`.xlsx`); use the supplied
`examples/sample_input.xlsx` as the template. Columns:
`id, ethnicity, gender, age, baseline_SER, baseline_AL, stratum`.
Only baseline columns are used. Ethnicity is the child's ancestry, not country of
residence, and is required. An ancestry with no derived model (South Asian in
particular) is not guessed: put a `stratum` value (`east_asian` or `european`), or
the child is not predicted and is listed in `out_needs_input.xlsx`. See
`docs/INPUT_DICTIONARY.md`.

## What the tool does and does not do

It builds the **control only** (`out_predictions.xlsx`: predicted untreated
elongation per visit, with the confidence interval of the mean where the model
publishes one). It does **not** compute a treatment effect. Compare your treated
outcomes against this control yourself, following `docs/ANALYSIS_PROTOCOL.md`, so
your treated outcome data never has to leave your machine.

## Model choice

One model per run. Only the Naduvilath 2025 regional GEE ships a confidence
interval of the mean; Brennan 2024, Jones 2005 (OLSM), and Wong 2010 (SCORM)
return a point estimate because their sources publish no parameter covariance. The
age-only models ignore ethnicity; the myope-only models exclude non-myopes. See
`docs/MODEL_NOTES.md`.

## Reproducibility note

The browser build pins Pyodide 0.28.3 (numpy 2.2.5, pandas 2.3.1, Python 3.13).
The shipped computation uses only elementary operations and a fixed-coefficient
inverse-normal polynomial, so results are independent of the numpy build to far
below the paper's reporting precision; the in-browser self-test confirms this for
each reader.
