## Supplementary software v0.2.1 for "Virtual control arms for paediatric myopia trials: external validation of axial-elongation models": run_mac_linux.sh

```
#!/usr/bin/env bash
# Virtual Control Arm Tool - local launcher (macOS / Linux)
# Serves this folder on a FREE port and opens your browser when it is ready.
# Nothing leaves your machine. Press Ctrl+C to stop.
cd "$(dirname "$0")" || exit 1

if ! command -v python3 >/dev/null 2>&1; then
  echo
  echo "  Python 3 was not found on this computer."
  echo
  echo "  This tool uses Python only to open the page in your browser."
  echo "  macOS: run 'python3' once and accept the Command Line Tools prompt,"
  echo "         or install from https://www.python.org/downloads/"
  echo "  Linux: install with your package manager, e.g. 'sudo apt install python3'."
  echo
  echo "  That one-time install is the ONLY time the internet is used. After that"
  echo "  the tool runs fully offline and never sends your data anywhere."
  echo
  exit 1
fi

exec python3 serve.py
```
