## Supplementary software v0.2.1 for "Virtual control arms for paediatric myopia trials: external validation of axial-elongation models": run_windows.bat

```
@echo off
REM Virtual Control Arm Tool - local launcher (Windows)
REM Serves this folder on a FREE port and opens your browser when it is ready.
REM Nothing leaves your machine. Close this window to stop.
cd /d "%~dp0"

where py >nul 2>nul && ( py serve.py & goto :eof )
where python >nul 2>nul && ( python serve.py & goto :eof )
goto :nopython

:nopython
echo(
echo   Python 3 was not found on this computer.
echo(
echo   This tool uses Python only to open the page in your browser. Install
echo   Python 3 once from:  https://www.python.org/downloads/
echo   and tick "Add python.exe to PATH" during setup.
echo(
echo   That one-time download is the ONLY time the internet is used. After
echo   Python is installed, this tool runs fully offline and never sends your
echo   data anywhere.
echo(
echo   When Python is installed, double-click this file again.
echo(
pause
```
