## Supplementary software v0.2.1 for "Virtual control arms for paediatric myopia trials: external validation of axial-elongation models": serve.py

```
#!/usr/bin/env python3
"""
Virtual Control Arm Tool -- local launcher.

Serves this folder to your own browser and nothing else. It:
  * chooses a FREE port automatically, so it never clashes with a server that is
    already running (the old cause of a stale page on a fixed port);
  * opens the browser only AFTER the server is actually listening;
  * sends no-cache headers, so the browser can never show you an out-of-date
    page after the tool is updated.

Everything stays on your machine. Python is used only to serve these local files;
no data leaves the computer. Close this window (or press Ctrl+C) to stop.
"""
import functools
import http.server
import os
import sys
import threading
import webbrowser

HERE = os.path.dirname(os.path.abspath(__file__))
os.chdir(HERE)


class _NoCacheHandler(http.server.SimpleHTTPRequestHandler):
    """Serve the local files, but tell the browser never to cache them, so an
    updated build is always the one you see."""

    def end_headers(self):
        self.send_header("Cache-Control", "no-store, no-cache, must-revalidate, max-age=0")
        self.send_header("Pragma", "no-cache")
        self.send_header("Expires", "0")
        super().end_headers()

    def log_message(self, *args):
        pass  # keep the console quiet


def main():
    handler = functools.partial(_NoCacheHandler, directory=HERE)
    # Port 0 asks the OS for any free port; 127.0.0.1 keeps it to this machine.
    # ThreadingHTTPServer (not a single-threaded server) is essential: the Pyodide
    # runtime loads over several overlapping keep-alive connections, and a
    # single-threaded server would stall on the first one and never finish
    # ("Loading the Python engine..." hangs forever).
    with http.server.ThreadingHTTPServer(("127.0.0.1", 0), handler) as httpd:
        httpd.daemon_threads = True
        port = httpd.server_address[1]
        url = "http://localhost:%d/index.html" % port
        print("Virtual Control Arm Tool is running at: %s" % url)
        print("Leave this window open while you use the tool. Close it to stop.")
        # The server is already listening at this point, so opening the browser
        # now cannot race ahead of it.
        threading.Thread(target=webbrowser.open, args=(url,), daemon=True).start()
        try:
            httpd.serve_forever()
        except KeyboardInterrupt:
            print("\nStopped.")
            return 0
    return 0


if __name__ == "__main__":
    sys.exit(main())
```
