## Supplementary software v0.2.1 for "Virtual control arms for paediatric myopia trials: external validation of axial-elongation models": adapt_template.py

```
"""
adapt_template.py  --  ingest the trial-team data export and emit the canonical
template the virtual-control tool expects.

WHY THIS EXISTS
---------------
The validated engine (virtual_control.py) and both runners (validate_model.py,
run_my_trial.py) expect ONE row per child with these columns:

    id, ethnicity, gender, age, baseline_SER, baseline_AL, AL_m6, AL_m12
    (the trial runner also allows AL_m18, AL_m24, AL_m36)

The trial team's export (e.g. Virtual_Control_data_2026-07-08.xlsx) is laid out
differently: one row per child but BOTH eyes side by side, sex coded 1/2, and
different headers:

    ID, Ethnicity, Sex, BL_age,
    BL_SER_OD, BL_SER_OS, BL_AL_OD, BL_AL_OS,
    6m_AL_OD, 6m_AL_OS, 12m_AL_OD, 12m_AL_OS

This module maps that export onto the canonical template WITHOUT touching the
validated engine. Every design decision is explicit and logged so it can be
defended in the paper's methods.

DECISIONS (change here, in one place, if the trial team says otherwise)
----------------------------------------------------------------------
1. EYE:  one eye per child is used for statistical independence (the validation
   design reduces each child to observed - predicted and tests N differences).
   Primary eye = RIGHT (OD). Set eye="OS" for the left-eye sensitivity run.
   baseline_SER and baseline_AL, and every follow-up AL, are all taken from the
   SAME eye. We do NOT mix eyes within a child.

2. SEX:  the export codes sex as 1/2. The convention is set by SEX_MAP below.
   Default: {2: "M", 1: "F"} -- data-driven, because in this cohort the Sex=2
   eyes are ~0.5 mm longer at matched age, the standard male pattern. If the
   trial team confirms the opposite convention, flip SEX_MAP and re-run; nothing
   else changes.

3. ETHNICITY:  passed through verbatim; the engine's population registry routes
   Chinese/Vietnamese -> East Asian (exact) and Indian -> European (provisional).

4. UNITS/CLEANING:  AL in mm, SER in dioptres, age in years. Rows are not
   dropped or edited on the basis of outcome; only genuinely empty cells become
   blanks (NaN) so a child with a missing visit still contributes where it has
   data. The one QC guard is a physiologic-range flag (reported, not silently
   removed).

USAGE
-----
    python adapt_template.py                      # OD, writes converted Excel workbook
    python adapt_template.py --eye OS             # left-eye sensitivity export
    python adapt_template.py --in FILE --out FILE

    # or from code:
    from adapt_template import adapt
    df = adapt("Virtual_Control_data_2026-07-08.xlsx", eye="OD")

DEPENDENCIES: pandas, numpy, openpyxl (for .xlsx).
"""
from __future__ import annotations
import argparse
import os
import numpy as np
import pandas as pd

# ---- the two conventions the trial team owns; override here if confirmed ----
SEX_MAP = {2: "M", 1: "F"}          # data-driven default (see decision 2)
PRIMARY_EYE = "OD"                   # right eye is primary (see decision 1)

# canonical output columns the engine expects (validation subset)
CANONICAL = ["id", "ethnicity", "gender", "age",
             "baseline_SER", "baseline_AL", "AL_m6", "AL_m12"]

# how each export column maps to (canonical_stub, eye) so we can pick one eye
#   canonical_stub in {SER, AL, AL_m6, AL_m12}; baseline handled by stub SER/AL
_EXPORT_EYE_COLS = {
    "BL_SER_OD": ("baseline_SER", "OD"), "BL_SER_OS": ("baseline_SER", "OS"),
    "BL_AL_OD":  ("baseline_AL",  "OD"), "BL_AL_OS":  ("baseline_AL",  "OS"),
    "6m_AL_OD":  ("AL_m6",  "OD"),       "6m_AL_OS":  ("AL_m6",  "OS"),
    "12m_AL_OD": ("AL_m12", "OD"),       "12m_AL_OS": ("AL_m12", "OS"),
}
# fixed (non-eye) columns: export header -> canonical header
_FIXED = {"ID": "id", "Ethnicity": "ethnicity", "BL_age": "age"}
# physiologic sanity ranges (flag only; never auto-drop)
_RANGES = {
    "age": (4.0, 20.0),
    "baseline_SER": (-15.0, 3.0),
    "baseline_AL": (18.0, 30.0),
    "AL_m6": (18.0, 32.0),
    "AL_m12": (18.0, 32.0),
}


def _read_any(path: str) -> pd.DataFrame:
    return pd.read_excel(path)


def _is_new_layout(df: pd.DataFrame) -> bool:
    return "BL_AL_OD" in df.columns or "6m_AL_OD" in df.columns


def adapt(path: str, eye: str = PRIMARY_EYE,
          sex_map: dict | None = None) -> pd.DataFrame:
    """Return a canonical one-row-per-child DataFrame for the chosen eye.

    If `path` is already in canonical form it is returned as-is (so the old
    template keeps working). New-layout files are mapped per the decisions above.
    """
    eye = eye.upper()
    if eye not in ("OD", "OS"):
        raise ValueError("eye must be 'OD' (right) or 'OS' (left)")
    sex_map = SEX_MAP if sex_map is None else sex_map
    raw = _read_any(path)

    # already canonical? pass through untouched.
    if not _is_new_layout(raw):
        missing = [c for c in ["id", "ethnicity", "gender", "age",
                               "baseline_SER", "baseline_AL"] if c not in raw.columns]
        if missing:
            raise ValueError("Unrecognised template; missing columns: %s" % missing)
        return raw.copy()

    out = pd.DataFrame()
    # fixed columns
    for src, dst in _FIXED.items():
        out[dst] = raw[src]
    # sex 1/2 -> M/F (unmapped codes -> NaN, flagged later)
    out["gender"] = raw["Sex"].map(sex_map)
    # per-eye columns for the selected eye only
    for src, (dst, e) in _EXPORT_EYE_COLS.items():
        if e == eye and src in raw.columns:
            out[dst] = raw[src]

    # order canonical columns first, keep only those we produced
    cols = [c for c in CANONICAL if c in out.columns]
    out = out[cols]
    out.attrs["eye"] = eye
    out.attrs["sex_map"] = sex_map
    out.attrs["n_children"] = len(out)
    return out


def qc_report(df: pd.DataFrame) -> pd.DataFrame:
    """Row-level QC flags (does not modify data). Columns: id, issue."""
    rows = []
    for r in df.itertuples():
        rid = getattr(r, "id", None)
        if pd.isna(getattr(r, "gender", np.nan)):
            rows.append({"id": rid, "issue": "sex code not in SEX_MAP"})
        for col, (lo, hi) in _RANGES.items():
            v = getattr(r, col, None)
            if v is not None and pd.notna(v) and not (lo <= float(v) <= hi):
                rows.append({"id": rid, "issue": f"{col}={v} outside [{lo},{hi}]"})
        # non-positive elongation is biologically possible (measurement noise) but
        # worth counting; flag clearly negative shrinkage beyond noise.
        b = getattr(r, "baseline_AL", np.nan)
        for col in ("AL_m6", "AL_m12"):
            v = getattr(r, col, np.nan)
            if pd.notna(b) and pd.notna(v) and (float(v) - float(b)) < -0.10:
                rows.append({"id": rid, "issue": f"{col} shrinks >0.10mm vs baseline"})
    return pd.DataFrame(rows)


def summarise(df: pd.DataFrame) -> str:
    lines = []
    lines.append(f"children: {len(df)}   eye: {df.attrs.get('eye', 'n/a')}   "
                 f"sex_map: {df.attrs.get('sex_map')}")
    if "ethnicity" in df:
        lines.append("ethnicity: " + ", ".join(
            f"{k}={v}" for k, v in df['ethnicity'].value_counts().items()))
    if "gender" in df:
        lines.append("gender: " + ", ".join(
            f"{k}={v}" for k, v in df['gender'].value_counts(dropna=False).items()))
    for col in ("baseline_AL", "AL_m6", "AL_m12"):
        if col in df:
            lines.append(f"{col}: n={df[col].notna().sum()}  "
                         f"mean={df[col].mean():.3f}" if df[col].notna().any()
                         else f"{col}: n=0")
    return "\n".join(lines)


def main():
    ap = argparse.ArgumentParser(description="Adapt trial export to canonical template")
    ap.add_argument("--in", dest="inp", default="Virtual_Control_data_2026-07-08.xlsx")
    ap.add_argument("--out", dest="out", default=None,
                    help="output Excel workbook (default: untreated_validation_<eye>.xlsx)")
    ap.add_argument("--eye", default=PRIMARY_EYE, choices=["OD", "OS", "od", "os"])
    args = ap.parse_args()
    eye = args.eye.upper()
    out = args.out or f"untreated_validation_{eye}.xlsx"

    df = adapt(args.inp, eye=eye)
    df.to_excel(out, index=False)
    print("=== ADAPTED:", args.inp, "->", out, "===")
    print(summarise(df))
    qc = qc_report(df)
    print(f"\nQC flags: {len(qc)}")
    if len(qc):
        print(qc.to_string(index=False))
    qc_path = os.path.splitext(out)[0] + "_qc.xlsx"
    qc.to_excel(qc_path, index=False)
    print(f"\nWrote {out} and {qc_path}")


if __name__ == "__main__":
    main()
```
