## Supplementary software v0.2.1 for "Virtual control arms for paediatric myopia trials: external validation of axial-elongation models": analysis_v2.py

```
"""
analysis_v2.py -- timing-aware re-analysis using Virtual_Control_data_2026-07-09
(adds Parental_Myopia and actual visit days). Predicts at each child's ACTUAL
elapsed time rather than nominal 6/12 months, then reports:
  1. primary published-model validation, NOMINAL vs ACTUAL time (timing effect)
  2. recalibration (Indian multiplier + PI sigma) with actual time, 5-fold CV
  3. multi-model validation at actual time
Right eyes (OD). Prints everything; writes calibration_v2.json.
"""
from __future__ import annotations
import json, numpy as np, pandas as pd
import virtual_control as vc

XLSX = "Virtual_Control_data_2026-07-09.xlsx"
SEXMAP = {2: "M", 1: "F"}
Z = 1.959963984540054
SEED, K = 20260709, 5
YR = 365.25


def cum_years(age0, ser, male, region, years, model=vc.MODEL_2025, dt=1.0/365.25):
    """Cumulative untreated elongation over `years`, integrating daily as the
    child ages. Mirrors the engine's monthly accumulation at finer resolution."""
    n = max(1, int(round(years / dt)))
    step = years / n
    return float(sum(model.annual_rate(age0 + i * step, ser, male, region) * step
                     for i in range(n)))


# column names have leading digits; pandas itertuples renames them _6m_AL_OD etc.
def load_fixed():
    df = pd.read_excel(XLSX)
    df["gender"] = df["Sex"].map(SEXMAP)
    ren = {"6m_AL_OD": "al6", "12m_AL_OD": "al12",
           "6m_visit_days": "d6", "12m_visit_days": "d12",
           "BL_AL_OD": "bal", "BL_SER_OD": "ser", "BL_age": "age",
           "Ethnicity": "eth", "Parental_Myopia": "pm", "ID": "id"}
    df = df.rename(columns=ren)
    rows = []
    for r in df.itertuples():
        if pd.isna(r.ser):
            continue
        male = 1.0 if r.gender == "M" else 0.0
        region = "east_asian" if r.eth in ("Chinese", "Vietnamese") else "european"
        for al, d, nom in [(r.al6, r.d6, 6), (r.al12, r.d12, 12)]:
            if pd.isna(al) or pd.isna(r.bal):
                continue
            yrs = (float(d) / YR) if pd.notna(d) else nom / 12.0
            rows.append({"id": r.id, "ethnicity": r.eth, "region": region,
                         "gender": r.gender, "age": r.age, "ser": r.ser, "male": male,
                         "pm": r.pm, "visit": nom, "obs": float(al) - float(r.bal),
                         "yrs_actual": yrs, "yrs_nom": nom / 12.0})
    return pd.DataFrame(rows)


def predict(df, which="actual", mult_indian=1.0):
    yr = df["yrs_actual"] if which == "actual" else df["yrs_nom"]
    out = []
    for (_, r), y in zip(df.iterrows(), yr):
        m = mult_indian if r["ethnicity"] == "Indian" else 1.0
        out.append(cum_years(r["age"], r["ser"], r["male"], r["region"], y) * m)
    return np.array(out)


def bias_table(df, pred, sigma=None, power=0.5):
    df = df.assign(pred=pred)
    rows = []
    for (reg, v), g in df.groupby(["region", "visit"]):
        err = g["pred"] - g["obs"]
        rec = {"region": reg, "visit": v, "n": len(g),
               "mean_obs": round(g["obs"].mean(), 3), "mean_pred": round(g["pred"].mean(), 3),
               "bias": round(err.mean(), 4), "rmse": round(float(np.sqrt((err**2).mean())), 4)}
        if sigma is not None:
            yrs = g["yrs_actual"].to_numpy()
            half = Z * sigma * (yrs ** power)
            rec["coverage"] = round(float(((g["obs"] >= g["pred"] - half) &
                                           (g["obs"] <= g["pred"] + half)).mean()), 3)
        rows.append(rec)
    return pd.DataFrame(rows).sort_values(["region", "visit"])


def main():
    df = load_fixed()
    print("n child-visits:", len(df), "| children:", df.id.nunique())
    print("visit counts:", df.groupby(["region", "visit"]).size().to_dict())
    print()

    # 1) PRIMARY published model: nominal vs actual time
    print("=== 1. PRIMARY (published 2025), NOMINAL vs ACTUAL time ===")
    print("-- nominal 6/12 mo --")
    print(bias_table(df, predict(df, "nominal")).to_string(index=False))
    print("-- actual elapsed time --")
    print(bias_table(df, predict(df, "actual")).to_string(index=False))
    print()

    # 2) Recalibration at ACTUAL time
    ea = df[df.region == "east_asian"]
    ind = df[df.ethnicity == "Indian"]
    # Indian multiplier at actual time (6m only exists)
    pind = predict(ind, "actual")
    mult = float(ind["obs"].sum() / pind.sum()) if pind.sum() > 0 else 1.0
    # PI sigma per sqrt-year on residuals after multiplier (all myopes), actual time
    dfp = df.copy()
    dfp["pred"] = predict(df, "actual", mult_indian=mult)
    resid = (dfp["obs"] - dfp["pred"]) / np.sqrt(dfp["yrs_actual"])
    sigma = float(resid.std(ddof=1))
    print("=== 2. CALIBRATION at actual time ===")
    print(f"Indian multiplier = {mult:.3f}   PI sigma = {sigma:.4f} mm/sqrt-yr")
    print("-- post-calibration (actual time) --")
    predc = predict(df, "actual", mult_indian=mult)
    print(bias_table(df, predc, sigma=sigma).to_string(index=False))

    # 5-fold CV (stratified by ethnicity), refit mult+sigma per fold
    rng = np.random.default_rng(SEED)
    cids = df[["id", "ethnicity"]].drop_duplicates()
    fold = {}
    for e in sorted(cids.ethnicity.unique()):
        mem = list(cids[cids.ethnicity == e].id); rng.shuffle(mem)
        for i, c in enumerate(mem):
            fold[c] = i % K
    df["fold"] = df.id.map(fold)
    cov = {}; bias = {}
    scored = []
    for f in range(K):
        tr, te = df[df.fold != f], df[df.fold == f]
        pi = predict(tr[tr.ethnicity == "Indian"], "actual")
        mtr = float(tr[tr.ethnicity == "Indian"]["obs"].sum() / pi.sum()) if pi.sum() > 0 else 1.0
        trp = tr.copy(); trp["pred"] = predict(tr, "actual", mult_indian=mtr)
        s = ((trp["obs"] - trp["pred"]) / np.sqrt(trp["yrs_actual"])).std(ddof=1)
        tep = te.copy(); tep["pred"] = predict(te, "actual", mult_indian=mtr); tep["sig"] = s
        scored.append(tep)
    sc = pd.concat(scored)
    print("-- 5-fold CV (out-of-sample), actual time --")
    for (reg, v), g in sc.groupby(["region", "visit"]):
        half = Z * g["sig"] * np.sqrt(g["yrs_actual"])
        c = ((g["obs"] >= g["pred"] - half) & (g["obs"] <= g["pred"] + half)).mean()
        b = (g["pred"] - g["obs"]).mean()
        print(f"  {reg:<11} {v:>2}mo n={len(g):<4d} bias={b:+.4f} coverage={c:.3f}")
    json.dump({"indian_multiplier": round(mult, 4),
               "pi_sigma_myope_per_sqrt_year": round(sigma, 4),
               "pi_time_scaling": "sqrt", "timing": "actual_visit_days",
               "source_cohort": XLSX}, open("calibration_v2.json", "w"), indent=2)
    print()

    # 3) multi-model at actual time
    import sensitivity_models as sm
    print("\n=== 4. MULTI-MODEL (actual time), bias ===")
    for (reg, v), g in df.groupby(["region", "visit"]):
        line = f"  {reg:<11} {v:>2}mo n={len(g):<4d}"
        for name in ["naduvilath2025", "brennan2024", "scorm_wong2010", "olsm_jones2005"]:
            mdl = sm.MODELS[name]
            preds = []
            for r in g.itertuples():
                if not mdl["applies"](r.ser, r.region):
                    preds.append(np.nan); continue
                preds.append(mdl["cum"](r.age, r.ser, r.male, r.region, r.yrs_actual*12))
            b = np.nanmean(np.array(preds) - g["obs"].to_numpy())
            line += f"  {name.split('_')[0][:5]}={b:+.3f}"
        print(line)


if __name__ == "__main__":
    main()
```
