## Supplementary software v0.2.1 for "Virtual control arms for paediatric myopia trials: external validation of axial-elongation models": build_excel_figures.py

```
"""
build_excel_figures.py -- write figures/virtual_control_figures.xlsx: the source
data for Figures 1-3 plus NATIVE, EDITABLE Excel charts, for journals that require
editable figures or a source-data file. Deterministic; imports analysis_v2 so the
numbers match the manuscript and the PNG/PDF figures exactly.
"""
import os, json
import numpy as np
import analysis_v2 as A
from openpyxl import Workbook
from openpyxl.chart import ScatterChart, BarChart, Reference, Series
from openpyxl.chart.marker import Marker
import sensitivity_models as sm

OUTDIR = "figures" if os.path.isdir("figures") else "."
OUT = os.path.join(OUTDIR, "virtual_control_figures.xlsx")

df = A.load_fixed()
calfile = "calibration_v2.json" if os.path.exists("calibration_v2.json") else "calibration.json"
mult = float(json.load(open(calfile))["indian_multiplier"])
df = df.copy()
df["pred_cal"] = A.predict(df, "actual", mult_indian=mult)
df["diff"] = df["pred_cal"] - df["obs"]
df["mean_po"] = (df["pred_cal"] + df["obs"]) / 2.0

GROUPS = [("east_asian", 6, "East Asian 6mo"),
          ("east_asian", 12, "East Asian 12mo"),
          ("european", 6, "South Asian 6mo")]

wb = Workbook()

# ---- Fig 1: calibration scatter (observed x, predicted y) ----
ws1 = wb.active; ws1.title = "Fig1_calibration"
col = 1
series_cols = []
for reg, v, lab in GROUPS:
    g = df[(df.region == reg) & (df.visit == v)]
    ws1.cell(1, col, lab + " observed"); ws1.cell(1, col + 1, lab + " predicted")
    for i, (_, r) in enumerate(g.iterrows(), start=2):
        ws1.cell(i, col, round(float(r["obs"]), 4))
        ws1.cell(i, col + 1, round(float(r["pred_cal"]), 4))
    series_cols.append((col, col + 1, len(g), lab))
    col += 3
# identity line data
idc = col
ws1.cell(1, idc, "identity x"); ws1.cell(1, idc + 1, "identity y")
ws1.cell(2, idc, 0); ws1.cell(2, idc + 1, 0)
ws1.cell(3, idc, 0.8); ws1.cell(3, idc + 1, 0.8)

ch1 = ScatterChart(); ch1.title = "Figure 1. Observed vs predicted (after calibration)"
ch1.x_axis.title = "Observed untreated elongation (mm)"
ch1.y_axis.title = "Predicted untreated elongation (mm)"
ch1.height = 12; ch1.width = 12
ch1.x_axis.scaling.min = 0; ch1.x_axis.scaling.max = 0.8
ch1.y_axis.scaling.min = 0; ch1.y_axis.scaling.max = 0.8
for xc, yc, n, lab in series_cols:
    xref = Reference(ws1, min_col=xc, min_row=2, max_row=n + 1)
    yref = Reference(ws1, min_col=yc, min_row=2, max_row=n + 1)
    s = Series(yref, xref, title=lab)
    s.marker = Marker(symbol="circle", size=5); s.graphicalProperties.line.noFill = True
    ch1.series.append(s)
# identity line series (line, no markers)
xref = Reference(ws1, min_col=idc, min_row=2, max_row=3)
yref = Reference(ws1, min_col=idc + 1, min_row=2, max_row=3)
sid = Series(yref, xref, title="line of identity")
sid.marker = Marker(symbol="none")
ch1.series.append(sid)
ws1.add_chart(ch1, "M2")

# ---- Fig 2: Bland-Altman (mean x, diff y) ----
ws2 = wb.create_sheet("Fig2_bland_altman")
col = 1; series_cols = []
for reg, v, lab in GROUPS:
    g = df[(df.region == reg) & (df.visit == v)]
    ws2.cell(1, col, lab + " mean"); ws2.cell(1, col + 1, lab + " diff")
    for i, (_, r) in enumerate(g.iterrows(), start=2):
        ws2.cell(i, col, round(float(r["mean_po"]), 4))
        ws2.cell(i, col + 1, round(float(r["diff"]), 4))
    series_cols.append((col, col + 1, len(g), lab))
    col += 3
mean_d = float(df["diff"].mean()); sd = float(df["diff"].std(ddof=1))
lo, hi = mean_d - 1.96 * sd, mean_d + 1.96 * sd
# reference lines across x 0..0.7
lc = col
for j, (name, yval) in enumerate([("mean", mean_d), ("+1.96SD", hi), ("-1.96SD", lo)]):
    ws2.cell(1, lc + 2 * j, name + " x"); ws2.cell(1, lc + 2 * j + 1, name + " y")
    ws2.cell(2, lc + 2 * j, 0); ws2.cell(2, lc + 2 * j + 1, round(yval, 4))
    ws2.cell(3, lc + 2 * j, 0.7); ws2.cell(3, lc + 2 * j + 1, round(yval, 4))

ch2 = ScatterChart(); ch2.title = "Figure 2. Bland-Altman (predicted - observed)"
ch2.x_axis.title = "Mean of predicted and observed (mm)"
ch2.y_axis.title = "Predicted minus observed (mm)"
ch2.height = 12; ch2.width = 14
for xc, yc, n, lab in series_cols:
    xref = Reference(ws2, min_col=xc, min_row=2, max_row=n + 1)
    yref = Reference(ws2, min_col=yc, min_row=2, max_row=n + 1)
    s = Series(yref, xref, title=lab)
    s.marker = Marker(symbol="circle", size=5); s.graphicalProperties.line.noFill = True
    ch2.series.append(s)
for j, name in enumerate(["mean", "+1.96SD", "-1.96SD"]):
    xref = Reference(ws2, min_col=lc + 2 * j, min_row=2, max_row=3)
    yref = Reference(ws2, min_col=lc + 2 * j + 1, min_row=2, max_row=3)
    s = Series(yref, xref, title=name); s.marker = Marker(symbol="none")
    ch2.series.append(s)
ws2.add_chart(ch2, "N2")

# ---- Fig 3: multi-model bias, clustered bars ----
ws3 = wb.create_sheet("Fig3_multimodel")
models = [("naduvilath2025", "Naduvilath 2025 (primary)"),
          ("brennan2024", "Brennan 2024"),
          ("scorm_wong2010", "SCORM (Wong 2010)"),
          ("olsm_jones2005", "OLSM (Jones 2005)")]
ws3.cell(1, 1, "Cell")
for j, (_, lab) in enumerate(models, start=2):
    ws3.cell(1, j, lab)
for i, (reg, v, lab) in enumerate(GROUPS, start=2):
    ws3.cell(i, 1, lab)
    g = df[(df.region == reg) & (df.visit == v)]
    for j, (name, _) in enumerate(models, start=2):
        mdl = sm.MODELS[name]; preds = []
        for r in g.itertuples():
            if not mdl["applies"](r.ser, r.region):
                preds.append(np.nan); continue
            preds.append(mdl["cum"](r.age, r.ser, r.male, r.region, r.yrs_actual * 12))
        ws3.cell(i, j, round(float(np.nanmean(np.array(preds) - g["obs"].to_numpy())), 4))
ch3 = BarChart(); ch3.type = "col"; ch3.grouping = "clustered"
ch3.title = "Figure 3. Bias (predicted - observed) by model"
ch3.y_axis.title = "Bias (mm)"; ch3.x_axis.title = "Region and visit"
ch3.height = 10; ch3.width = 18
data = Reference(ws3, min_col=2, max_col=1 + len(models), min_row=1, max_row=1 + len(GROUPS))
cats = Reference(ws3, min_col=1, min_row=2, max_row=1 + len(GROUPS))
ch3.add_data(data, titles_from_data=True); ch3.set_categories(cats)
ws3.add_chart(ch3, "H2")

# ---- notes ----
wsn = wb.create_sheet("README")
notes = [
    "Source data and native Excel charts for Figures 1-3.",
    f"Timing basis: actual visit days (primary). Indian multiplier {mult} from {calfile}.",
    "Numbers match the manuscript and the PNG/PDF figures exactly.",
    "Charts are native and editable; edit the data cells and the charts update.",
    "Fig1: observed vs predicted, after calibration, by region x visit, with identity line.",
    "Fig2: Bland-Altman, predicted - observed, with mean and +/-1.96 SD lines.",
    "Fig3: multi-model bias (predicted - observed) with the +/-0.03 mm target.",
]
for i, t in enumerate(notes, start=1):
    wsn.cell(i, 1, t)

wb.save(OUT)
print("wrote", OUT)
```
