## Supplementary software v0.2.1 for "Virtual control arms for paediatric myopia trials: external validation of axial-elongation models": compute_inference.py

```
"""Inferential statistics the reviewers asked for: coverage Wilson CIs, bias CIs
and TOST vs +/-0.03, calibration-slope CIs, cluster-bootstrap sigma CI, PM
equivalence. Actual visit time throughout.

TIMING BASIS: this is the PRIMARY analysis, at each child's ACTUAL elapsed
follow-up time (see the paper's visit-timing section). The nominal 6/12-month
analysis in recalibrate.py is reported separately as the visit-timing CONTRAST,
not as the primary. To avoid mixing conventions, this script reads the actual-
time calibration (calibration_v2.json, written by analysis_v2.py) and NOT the
nominal calibration.json. Both bases are reported; they are never mixed."""
import os, json, numpy as np, pandas as pd
from scipy import stats
import analysis_v2 as A
Z = 1.959963984540054
# Actual-time calibration to match the actual-time predictions used below.
_CALFILE = "calibration_v2.json" if os.path.exists("calibration_v2.json") else "calibration.json"
cal = json.load(open(_CALFILE))
MULT, SIG = cal["indian_multiplier"], cal["pi_sigma_myope_per_sqrt_year"]
print(f"[timing=actual visit days; calibration source={_CALFILE}; "
      f"Indian multiplier={MULT}, PI sigma={SIG}]")

df = A.load_fixed()

def wilson(k, n, z=Z):
    if n == 0: return (np.nan, np.nan)
    p = k / n; d = 1 + z*z/n
    c = (p + z*z/(2*n)) / d
    h = z*np.sqrt(p*(1-p)/n + z*z/(4*n*n)) / d
    return (c-h, c+h)

def cell_stats(g, sigma, mult):
    n = len(g); nch = g["id"].nunique()
    pred = A.predict(g, "actual", mult_indian=mult)
    err = pred - g["obs"].to_numpy()
    bias = err.mean(); sd = err.std(ddof=1); se = sd/np.sqrt(n)
    ci = (bias - stats.t.ppf(.975, n-1)*se, bias + stats.t.ppf(.975, n-1)*se)
    # TOST vs +/-0.03
    t_lo = (bias-(-0.03))/se; t_hi = (bias-0.03)/se
    p_tost = max(1-stats.t.cdf(t_lo, n-1), stats.t.cdf(t_hi, n-1))
    yrs = g["yrs_actual"].to_numpy(); half = Z*sigma*np.sqrt(yrs)
    cov_in = ((g["obs"]>=pred-half)&(g["obs"]<=pred+half))
    k = int(cov_in.sum()); cov = k/n; wl, wh = wilson(k, n)
    # calibration slope + CI
    x = pred; y = g["obs"].to_numpy()
    if np.std(x) > 0:
        sl, ic, r, p, sse = stats.linregress(x, y)
        slci = (sl - Z*sse, sl + Z*sse)
    else:
        sl, slci = np.nan, (np.nan, np.nan)
    return dict(n=n, nch=nch, bias=bias, biasci=ci, p_tost=p_tost,
                cov=cov, covci=(wl, wh), slope=sl, slci=slci)

print("=== Per-cell inference (calibrated, actual time) ===")
for (reg, v), g in df.groupby(["region", "visit"]):
    s = cell_stats(g, SIG, MULT)
    print(f"{reg} {v}mo: n={s['n']} children={s['nch']}")
    print(f"   bias={s['bias']:+.4f} 95%CI[{s['biasci'][0]:+.4f},{s['biasci'][1]:+.4f}] TOST(±0.03)p={s['p_tost']:.3f} "
          f"{'EQUIV' if s['p_tost']<0.05 else 'not-equiv'}")
    print(f"   coverage={s['cov']:.3f} Wilson95%[{s['covci'][0]:.3f},{s['covci'][1]:.3f}]")
    print(f"   calib_slope={s['slope']:.2f} 95%CI[{s['slci'][0]:.2f},{s['slci'][1]:.2f}]")

# published-PI coverage Wilson CIs (myope pi_se linear, mult 1)
import virtual_control as vc
print("\n=== Published-PI coverage Wilson CIs (no calibration) ===")
for (reg, v), g in df.groupby(["region", "visit"]):
    pred = A.predict(g, "actual", mult_indian=1.0)
    yrs = g["yrs_actual"].to_numpy(); half = Z*vc.MODEL_2025.my_pi_se*yrs
    k = int(((g["obs"]>=pred-half)&(g["obs"]<=pred+half)).sum()); n=len(g)
    wl, wh = wilson(k, n)
    print(f"{reg} {v}mo: coverage={k/n:.3f} Wilson95%[{wl:.3f},{wh:.3f}]")

# cluster (by child) bootstrap sigma CI
print("\n=== sigma: cluster bootstrap by child ===")
dfp = df.copy(); dfp["pred"] = A.predict(df, "actual", mult_indian=MULT)
dfp["z"] = (dfp["obs"]-dfp["pred"])/np.sqrt(dfp["yrs_actual"])
ids = dfp["id"].unique(); rng = np.random.default_rng(20260709)
by = {i: dfp[dfp.id==i]["z"].values for i in ids}
boot = []
for _ in range(5000):
    pick = rng.choice(ids, len(ids), replace=True)
    boot.append(np.concatenate([by[i] for i in pick]).std(ddof=1))
print(f"sigma={dfp['z'].std(ddof=1):.4f} cluster95%[{np.percentile(boot,2.5):.4f},{np.percentile(boot,97.5):.4f}]  "
      f"n_children={dfp['id'].nunique()} n_rows={len(dfp)}")

# in-sample vs cross-validated coverage: see analysis_v2.py output for the
# per-fold numbers (in-sample and 5-fold CV are printed there side by side).
print("\n=== in-sample vs CV coverage: see analysis_v2.py (actual-time) for per-fold values ===")
```
