## Supplementary software v0.2.1 for "Virtual control arms for paediatric myopia trials: external validation of axial-elongation models": demo.py

```
"""
demo.py -- run every user-facing workflow on SYNTHETIC data, so a reader without
the governed cohort can see the tool work end to end. Generates its own synthetic
inputs if they are missing.

    python demo.py

It writes everything to ./demo_out/ and does three things:
  1. engine self-validation against Naduvilath 2025 (needs no data at all)
  2. validation on a synthetic untreated cohort (bias, coverage, plots)
  3. a trial prediction (the virtual untreated control) on input_template.xlsx

It never runs the calibration-fitting scripts, so the shipped
calibration_v2.json / calibration.json are never overwritten. The exact paper
tables come from reproduce.py on the GOVERNED cohort and are not reproduced here.

THE SYNTHETIC DATA ARE NOT REAL PATIENTS. They are drawn from the published model
plus random-walk noise, purely so the code executes and its outputs have the right
shape. Numbers from them are illustrative of the mechanics only, never a result.

Deterministic (fixed seed, no network, no wall-clock). Depends on numpy, pandas,
matplotlib, openpyxl, and the tool modules.
"""
from __future__ import annotations
import os
import sys
import numpy as np
import pandas as pd
import virtual_control as vc

SEED = 20260720
YR = 365.25
N_PER_GROUP = {"Chinese": 40, "Vietnamese": 30, "Indian": 30}
INDIAN_TRUTH_MULT = 1.30          # exercises the provisional multiplier path
SIGMA_SQRT_YR = 0.15              # residual SD per sqrt-year (random walk)

SRC = os.path.dirname(os.path.abspath(__file__))
OUT = os.path.join(SRC, "demo_out")
SYNTH_UNTREATED = os.path.join(OUT, "untreated_validation_SYNTHETIC.xlsx")
INPUT_TEMPLATE = os.path.join(SRC, "input_template.xlsx")


def _synth_followup(rng, age, ser, male, eth, years):
    region = "east_asian" if eth in ("Chinese", "Vietnamese") else "european"
    point = vc.MODEL_2025.cumulative(age, ser, male, region, int(round(years * 12)))
    if eth == "Indian":
        point *= INDIAN_TRUTH_MULT
    return max(0.0, point + rng.normal(0.0, SIGMA_SQRT_YR * np.sqrt(years)))


def make_synthetic():
    """Write a synthetic canonical validation file to demo_out/. Deterministic."""
    rng = np.random.default_rng(SEED)
    rows = []
    cid = 1
    for eth, n in N_PER_GROUP.items():
        for _ in range(n):
            male = int(rng.integers(0, 2))
            age = float(np.round(rng.uniform(7.0, 12.0), 1))
            ser = float(np.round(rng.uniform(-5.0, -0.5), 2))
            bal = float(np.round(rng.uniform(23.5, 25.5), 2))
            d6 = float(np.round(rng.normal(183, 18)))
            has12 = eth != "Indian" and rng.random() < 0.55
            d12 = float(np.round(rng.normal(365, 25))) if has12 else np.nan
            al6 = float(np.round(bal + _synth_followup(rng, age, ser, float(male), eth, d6 / YR), 2))
            al12 = (float(np.round(bal + _synth_followup(rng, age, ser, float(male), eth, d12 / YR), 2))
                    if has12 else np.nan)
            rows.append({"id": f"S{cid:03d}", "ethnicity": eth,
                         "gender": "M" if male else "F", "age": age,
                         "baseline_SER": ser, "baseline_AL": bal,
                         "AL_m6": al6, "AL_m12": al12})
            cid += 1
    df = pd.DataFrame(rows)
    df.to_excel(SYNTH_UNTREATED, index=False)
    return df


def main():
    os.makedirs(OUT, exist_ok=True)
    if not os.path.exists(SYNTH_UNTREATED):
        make_synthetic()
    os.chdir(OUT)                    # every artefact lands in demo_out/

    import validate_model as vm
    import run_my_trial as rmt

    print("=" * 64, "\n1. ENGINE SELF-VALIDATION (no data needed)\n", "=" * 64, sep="")
    ok = vc.validate_against_paper()
    print("engine self-validation:", "PASS" if ok else "FAIL")

    print("\n" + "=" * 64, "\n2. VALIDATION on SYNTHETIC untreated cohort\n", "=" * 64, sep="")
    vm.run_validation(path=SYNTH_UNTREATED)

    print("\n" + "=" * 64, "\n3. TRIAL prediction (virtual control) on input_template.xlsx\n", "=" * 64, sep="")
    rmt.WIDE = INPUT_TEMPLATE
    rmt.main()

    print("\n" + "=" * 64)
    print("demo.py complete. Artefacts in src/demo_out/ :")
    for f in sorted(os.listdir(OUT)):
        print("   ", f)
    print("\nSYNTHETIC inputs -> illustrative outputs only. Shipped calibrations were")
    print("not modified. Paper tables require the governed cohort (see README).")


if __name__ == "__main__":
    main()
```
