## Supplementary software v0.2.1 for "Virtual control arms for paediatric myopia trials: external validation of axial-elongation models": gui_app.py

```
"""
Virtual Control Arm Tool -- desktop application (no Python knowledge required).

Double-click the built executable, choose your filled-in template, choose an
output folder, and press Run. One result file is written:

  out_predictions.xlsx  -- per patient, per visit: the predicted untreated axial
                          elongation with its mean confidence band (for the
                          Naduvilath 2025 GEE model; the other models return a
                          point estimate, as their sources publish no interval).

You choose the model and the ethnicity at the top of the window. One model and
one ethnicity are run per pass; run again to compare a different model.

This tool builds the VIRTUAL UNTREATED CONTROL only. It deliberately does NOT
compute a treatment effect (CARE, delta, p-value, or percent reduction). You
compare your own treated outcomes against this control yourself, keeping your
outcome data private. The recommended analysis, including how to carry the
control's uncertainty into the effect estimate, is in docs/ANALYSIS_PROTOCOL.md.

All computation is local. Nothing is uploaded.

Copyright (C) 2026 nthalmic Pty Ltd. GNU General Public License v3.0 or later.
"""
import os
import sys
import traceback

import pandas as pd

import virtual_control as vc

# Version is defined once, in the engine (virtual_control.py); never hard-code it
# here, so the GUI can never disagree with the engine, outputs, or CITATION.
__version__ = vc.__version__
__release_date__ = vc.__release_date__

VISIT_COLS = {"AL_m6": 6, "AL_m12": 12, "AL_m18": 18, "AL_m24": 24, "AL_m36": 36}
REQUIRED_BASE = ["id", "age", "baseline_SER", "baseline_AL"]


def resource_path(name):
    """Path to a bundled data file, whether run from source or a PyInstaller exe."""
    base = getattr(sys, "_MEIPASS", os.path.dirname(os.path.abspath(__file__)))
    return os.path.join(base, name)


def _read_table(path):
    """Read an Excel (.xlsx) input into a DataFrame."""
    return pd.read_excel(path)


def _to_wide(df, value_cols):
    """Reshape a long per-child-per-month table to one row per child, with each
    value column suffixed by the month, for example predicted_delta_AL_m12."""
    months = sorted(int(m) for m in df["month"].unique())
    id_cols = [c for c in df.columns if c not in value_cols and c != "month"]
    out = df.drop_duplicates("id").set_index("id")[[c for c in id_cols if c != "id"]].copy()
    for m in months:
        sub = df[df["month"] == m].set_index("id")
        for v in value_cols:
            out[f"{v}_m{m}"] = sub[v]
    return out.reset_index()


PRED_VALUE_COLS = ["predicted_delta_AL", "ci_low", "ci_high", "predicted_AL"]

# The tool-facing model menu: first-author + model labels, exactly as in the paper.
# Order is neutral (publication year); no model is presented as "primary" here.
# label -> internal model_name understood by vc.predict_untreated.
MODEL_MENU = [
    ("Naduvilath 2025 (regional GEE)", "naduvilath2025"),
    ("Brennan 2024 (meta-regression)", "brennan2024"),
    ("Jones 2005 (OLSM)", "olsm_jones2005"),
    ("Wong 2010 (SCORM)", "scorm_wong2010"),
]
MODEL_LABEL_TO_NAME = {lbl: name for lbl, name in MODEL_MENU}

# Ethnicity dropdown. The app selection is authoritative for the run; the file's
# ethnicity column must agree or the row is flagged (the user chose one ethnicity
# per run). "(from input file)" defers to the per-row file column instead.
ETH_FROM_FILE = "(from input file)"
ETH_CHOICES = [
    ETH_FROM_FILE,
    "Chinese", "Vietnamese", "Taiwanese", "Singaporean", "Malaysian",
    "Australian", "European", "British", "Swedish", "Portuguese", "Spanish",
    "New Zealand", "Indian", "South Asian",
]


def run_analysis(input_path, out_dir, layout="long",
                 model_name="naduvilath2025", gui_ethnicity=None):
    """Headless core, shared by the GUI and by tests.

    Builds the virtual untreated control from baseline data and writes
    out_predictions.xlsx. It does NOT compute a treatment effect: the tool's job
    is to supply the control arm, and the effect analysis is the analyst's own
    step (see docs/ANALYSIS_PROTOCOL.md), run on outcome data that never has to
    leave the analyst's machine.

    One model is run per call (`model_name`, the user's choice; run again to
    compare). Only the Naduvilath 2025 GEE ships a confidence interval of the
    mean; the other models return a point estimate with a note, because their
    sources publish no parameter covariance.

    `gui_ethnicity`, when given and not "(from input file)", is authoritative for
    the whole run: any file row whose ethnicity disagrees is flagged (not
    predicted) rather than silently overridden, and a blank file ethnicity takes
    the app selection.

    Returns (predictions_df, summary_text). Raises ValueError with a
    plain-language message on bad input.
    """
    model_name = (model_name or "naduvilath2025").strip().lower()
    if model_name not in {n for _, n in MODEL_MENU}:
        raise ValueError("Unknown model selection: %r" % model_name)
    gui_eth = (gui_ethnicity or "").strip()
    use_gui_eth = bool(gui_eth) and gui_eth != ETH_FROM_FILE

    df = _read_table(input_path)

    missing = [c for c in REQUIRED_BASE if c not in df.columns]
    if "ethnicity" not in df.columns:
        missing.append("ethnicity")
    if "gender" not in df.columns:
        missing.append("gender")
    if missing:
        raise ValueError(
            "The input is missing these required columns: "
            + ", ".join(missing)
            + ".\nEthnicity means ancestry, not country of residence."
            + "\nUse the bundled template (File > Load example template) as a guide."
        )

    subjects, excl = [], []
    for r in df.itertuples():
        eth = getattr(r, "ethnicity", None)
        file_eth = "" if (pd.isna(eth) or str(eth).strip() == "") else str(eth).strip()
        if use_gui_eth:
            # The app selection is authoritative for the run. A file ethnicity
            # that disagrees is a user error: flag it, do not silently override.
            if file_eth and file_eth.lower() != gui_eth.lower():
                excl.append({"id": getattr(r, "id", "(no id)"),
                             "reason": ("ethnicity in the file ('%s') does not match the "
                                        "ethnicity selected in the app ('%s'); correct "
                                        "one of them (one ethnicity per run)"
                                        % (file_eth, gui_eth))})
                continue
            row_eth = gui_eth                      # blank file ethnicity takes the app choice
        else:
            if not file_eth:
                excl.append({"id": getattr(r, "id", "(no id)"),
                             "reason": "ethnicity (ancestry) is required; country of "
                                       "residence is not accepted"})
                continue
            row_eth = file_eth
        subjects.append(
            {"id": r.id, "ethnicity": row_eth, "gender": r.gender,
             "age": r.age, "baseline_SER": r.baseline_SER, "baseline_AL": r.baseline_AL,
             "stratum": getattr(r, "stratum", None)})

    # Detect whether the analyst also supplied treated follow-up axial lengths.
    # We do NOT use them; we only note their presence so the analyst is pointed
    # to the correct effect-analysis procedure.
    treated_visits = 0
    for r in df.itertuples():
        for col in VISIT_COLS:
            al = getattr(r, col, None)
            if al is not None and pd.notna(al):
                treated_visits += 1

    model_label = {n: lbl for lbl, n in MODEL_MENU}.get(model_name, model_name)
    ships_ci = (model_name == "naduvilath2025")

    pred = vc.predict_untreated(subjects, exclusions=excl, model_name=model_name)
    os.makedirs(out_dir, exist_ok=True)
    pred_out = _to_wide(pred, PRED_VALUE_COLS) if (layout == "wide" and not pred.empty) else pred
    pred_path = os.path.join(out_dir, "out_predictions.xlsx")
    vc.write_output(pred_out, pred_path, model_label=model_label)

    needs_path = None
    if excl:
        needs_path = os.path.join(out_dir, "out_needs_input.xlsx")
        vc.write_output(pd.DataFrame(excl), needs_path,
                        model_label=model_label, sheet_name="needs_input")

    fmt = ("wide (one row per child, columns suffixed by month)" if layout == "wide"
           else "long (one row per child per month)")
    eth_line = ("    Ethnicity (app): " + gui_eth) if use_gui_eth else "    Ethnicity: from input file"
    lines = [
        f"Virtual untreated control written to:\n  {pred_path}",
        f"\nModel: {model_label}{eth_line}",
        f"Patients: {df['id'].nunique()}    Output format: {fmt}",
    ]
    if ships_ci:
        lines.append(
            "\nEach child has a predicted untreated change in axial length "
            "(predicted_delta_AL) with its mean confidence band (ci_low, ci_high).")
    else:
        lines.append(
            "\nEach child has a predicted untreated change in axial length "
            "(predicted_delta_AL). This model's source publishes no parameter "
            "covariance, so ci_low/ci_high are blank: it is a point estimate. "
            "For a control arm with a confidence interval of the mean, run the "
            "Naduvilath 2025 (regional GEE) model. This model applies to myopes "
            "only (baseline SER ≤ −0.50 D); non-myopes are listed in "
            "out_needs_input.xlsx.")

    if treated_visits:
        lines.append(
            f"\nNote: {treated_visits} treated follow-up value(s) were present in "
            "the input. This tool does NOT compute the treatment effect. Compare "
            "your treated outcomes against this control using the procedure in "
            "docs/ANALYSIS_PROTOCOL.md, which shows how to carry the control's "
            "uncertainty into the effect estimate. Your outcome data stay with you."
        )
    else:
        lines.append(
            "\nNext step: analyse your treated outcomes against this control using "
            "docs/ANALYSIS_PROTOCOL.md. The tool intentionally stops at the control."
        )

    if needs_path:
        lines.append(
            f"\n{len(excl)} row(s) were NOT predicted (see {needs_path}): a missing "
            "ethnicity, or an ancestry with no derived model (e.g. South Asian) that "
            "needs a 'stratum' choice of east_asian or european in the input.")

    lines.append(
        "\nReminder: this tool is supportive evidence, validated for East Asian "
        "myopes. There is no South Asian model; those children are predicted only "
        "under a stratum you choose, and are provisional. See the paper.")
    return pred, "\n".join(lines)


# ---------------------------------------------------------------------------
# GUI (tkinter is only imported here, so the headless core stays dependency-light)
# ---------------------------------------------------------------------------
def launch_gui():
    import tkinter as tk
    from tkinter import filedialog, messagebox, scrolledtext, ttk

    root = tk.Tk()
    root.title(f"Virtual Control Arm Tool  v{__version__}")
    root.geometry("760x600")
    root.minsize(640, 500)

    state = {"input": tk.StringVar(), "outdir": tk.StringVar(),
             "wide": tk.BooleanVar(value=False),
             "model": tk.StringVar(value=MODEL_MENU[0][0]),
             "ethnicity": tk.StringVar(value=ETH_FROM_FILE)}

    def pick_input():
        p = filedialog.askopenfilename(
            title="Choose your filled-in template",
            filetypes=[("Data files", "*.xlsx"), ("All files", "*.*")])
        if p:
            state["input"].set(p)
            if not state["outdir"].get():
                state["outdir"].set(os.path.dirname(p))

    def pick_outdir():
        p = filedialog.askdirectory(title="Choose an output folder")
        if p:
            state["outdir"].set(p)

    def load_example():
        import shutil
        # The template is a single Excel workbook: an 'Enter data' sheet with
        # dropdowns plus a 'Dictionary' sheet defining every input and output.
        src = resource_path("input_template.xlsx")
        dest = filedialog.asksaveasfilename(
            title="Save the template to fill in",
            defaultextension=".xlsx", initialfile="input_template.xlsx",
            filetypes=[("Excel", "*.xlsx")])
        if dest:
            try:
                shutil.copyfile(src, dest)
            except Exception as e:  # noqa: BLE001
                messagebox.showerror("Template", f"Could not save the template.\n{e}")
                return
            state["input"].set(dest)
            log(f"Template saved to:\n  {dest}\nOpen it in Excel, read the "
                "'Dictionary' tab, fill the 'Enter data' tab, save, then press Run.\n")

    def log(msg):
        out.configure(state="normal")
        out.insert("end", msg + "\n")
        out.see("end")
        out.configure(state="disabled")

    def run():
        inp, od = state["input"].get(), state["outdir"].get()
        if not inp:
            messagebox.showwarning("Run", "Choose an input file first.")
            return
        if not od:
            state["outdir"].set(os.path.dirname(inp))
            od = state["outdir"].get()
        try:
            log("Running ...")
            layout = "wide" if state["wide"].get() else "long"
            model_name = MODEL_LABEL_TO_NAME.get(state["model"].get(), "naduvilath2025")
            gui_eth = state["ethnicity"].get()
            _, summary = run_analysis(inp, od, layout=layout,
                                      model_name=model_name, gui_ethnicity=gui_eth)
            log(summary + "\n" + "-" * 60)
            messagebox.showinfo(
                "Done",
                "Virtual control written to the output folder.\n"
                "Analyse your treated outcomes against it using "
                "docs/ANALYSIS_PROTOCOL.md.")
        except ValueError as e:
            messagebox.showerror("Input problem", str(e))
        except Exception as e:  # noqa: BLE001
            messagebox.showerror("Error", f"{e}\n\n{traceback.format_exc()}")

    pad = {"padx": 8, "pady": 4}
    top = tk.Frame(root)
    top.pack(fill="x", **pad)

    tk.Button(top, text="1. Choose input file", width=20, command=pick_input).grid(row=0, column=0, **pad)
    tk.Entry(top, textvariable=state["input"]).grid(row=0, column=1, sticky="we", **pad)
    tk.Button(top, text="2. Output folder", width=20, command=pick_outdir).grid(row=1, column=0, **pad)
    tk.Entry(top, textvariable=state["outdir"]).grid(row=1, column=1, sticky="we", **pad)
    top.columnconfigure(1, weight=1)

    sel = tk.Frame(root)
    sel.pack(fill="x", **pad)
    tk.Label(sel, text="3. Model").grid(row=0, column=0, sticky="w", **pad)
    ttk.Combobox(sel, textvariable=state["model"], state="readonly",
                 values=[lbl for lbl, _ in MODEL_MENU], width=32
                 ).grid(row=0, column=1, sticky="w", **pad)
    tk.Label(sel, text="Ethnicity (ancestry)").grid(row=0, column=2, sticky="w", **pad)
    ttk.Combobox(sel, textvariable=state["ethnicity"], state="readonly",
                 values=ETH_CHOICES, width=20).grid(row=0, column=3, sticky="w", **pad)
    tk.Label(sel, text="One model and one ethnicity per run. Only Naduvilath 2025 "
                       "(GEE) returns a confidence interval; the others give a point "
                       "estimate.\nEthnicity is the child's ancestry, not country of "
                       "residence. '(from input file)' uses each row's ethnicity "
                       "column; any other choice must match the file or the row is "
                       "flagged.",
             justify="left", fg="#444", wraplength=720
             ).grid(row=1, column=0, columnspan=4, sticky="w", padx=8)

    mid = tk.Frame(root)
    mid.pack(fill="x", **pad)
    tk.Button(mid, text="Load example template", command=load_example).pack(side="left", **pad)
    tk.Checkbutton(mid, text="Wide format (one row per child)",
                   variable=state["wide"]).pack(side="left", **pad)
    tk.Button(mid, text="Run", width=14, bg="#1a7f4b", fg="white",
              command=run).pack(side="right", **pad)

    out = scrolledtext.ScrolledText(root, height=20, state="disabled", wrap="word")
    out.pack(fill="both", expand=True, **pad)

    log("Virtual Control Arm Tool for myopia-control studies.\n"
        "Step 1: choose your filled-in Excel template (.xlsx).\n"
        "Step 2: choose where to save the results.\n"
        "Then press Run. New to the format? Press 'Load example template'.\n"
        "This tool outputs the virtual untreated control only; you run the\n"
        "effect analysis yourself (see docs/ANALYSIS_PROTOCOL.md).\n"
        "All computation is local; nothing is uploaded.\n" + "-" * 60)

    root.mainloop()


if __name__ == "__main__":
    # If launched with a file argument (drag-and-drop onto the exe), run headless.
    if len(sys.argv) > 1 and os.path.isfile(sys.argv[1]):
        inp = sys.argv[1]
        outdir = sys.argv[2] if len(sys.argv) > 2 else os.path.dirname(inp)
        _, summary = run_analysis(inp, outdir)
        print(summary)
    else:
        launch_gui()
```
