## Supplementary software v0.2.1 for "Virtual control arms for paediatric myopia trials: external validation of axial-elongation models": make_fig4.py

```
"""
make_fig4.py -- Figure 4 for the virtual-control manuscript.

"Using the model under its own uncertainty." A worked, ILLUSTRATIVE example
(representative East Asian myopes, no real patient data) that shows how the
virtual-control output enters the trial statistics:
  Panel A -- the virtual-control mean untreated trajectory with the
             uncertainty of the mean (the confidence interval, the only band
             the tool ships), the treated cohort mean below it, and CARE at 12
             mo as the vertical gap, shown with the recommended effect interval.
  Panel B -- the SAME effect's 95% interval built two ways: with the control
             treated as fixed (too narrow), and with the control's own
             uncertainty included (recommended).

The effect interval uses ONLY what the tool ships (the control confidence
interval of the mean) plus the user's treated summary (SD and n). No Monte
Carlo. It is the closed form of docs/ANALYSIS_PROTOCOL.md section 4:
    SE_within = SD_treated / sqrt(n)                treated sampling error
    SE_ctrl   = (ci_high - ci_low) / (2 * 1.96)     from the tool output
    SE_effect = sqrt(SE_within**2 + SE_ctrl**2)
    CI        = CARE  +/-  1.96 * SE_effect
"Control treated as fixed" drops SE_ctrl; that omission is why it is too narrow.

Deterministic. Matches the palette of Figures 1-3.
"""
import numpy as np
import matplotlib
matplotlib.use("Agg")
import matplotlib.pyplot as plt
import virtual_control as vc

NAVY, SKY, RED, GREY = "#1F3864", "#5B9BD5", "#B00020", "#7F7F7F"
BANDMEAN = "#BBD0EA"
Z = 1.959963984540054
plt.rcParams.update({"savefig.dpi": 600, "pdf.fonttype": 42, "ps.fonttype": 42,
                     "font.size": 11, "axes.grid": True,
                     "grid.color": "#DDDDDD", "grid.linewidth": 0.6})

m = vc.MODEL_2025
AGE, SER, MALE = 10.0, -2.5, 0.5           # representative EA myope cohort
CI_SE = m.se(SER, "ci")                     # mean-prediction SE, mm per year (from the tool ci)
N = 60                                      # treated completers at 12 mo (illustrative)
TREATED_12 = 0.18                           # treated cohort mean elongation at 12 mo (illustrative)
SD_TREATED = 0.14                           # individual treated residual SD (illustrative)

# ----- trajectories -----
months = np.arange(0, 13)
yrs = months / 12.0
vc_mean = np.array([m.cumulative(AGE, SER, MALE, "east_asian", int(k)) if k > 0 else 0.0
                    for k in months])
vc12 = vc_mean[-1]                                        # 0.344 mm
mean_half = Z * CI_SE * yrs                               # uncertainty of the mean trajectory
treated_mean = vc_mean * (TREATED_12 / vc12)             # illustrative treated shape
CARE = vc12 - TREATED_12                                  # 0.164 mm

# ----- closed-form effect interval (ANALYSIS_PROTOCOL section 4) -----
se_within = SD_TREATED / np.sqrt(N)                      # treated sampling error
se_ctrl   = CI_SE                                        # control-mean SE at 12 mo (yrs = 1)
se_eff    = np.hypot(se_within, se_ctrl)                 # combined, in quadrature
fixed = (CARE - Z * se_within, CARE + Z * se_within)    # control treated as fixed (too narrow)
reco  = (CARE - Z * se_eff,    CARE + Z * se_eff)       # control uncertainty included (recommended)

fig, (axA, axB) = plt.subplots(1, 2, figsize=(13.2, 5.3), gridspec_kw={"width_ratios": [1.05, 1.0]})

# ===== Panel A =====
axA.fill_between(months, vc_mean - mean_half, vc_mean + mean_half, color=BANDMEAN,
                 label="Uncertainty of the untreated mean (confidence interval)")
axA.plot(months, vc_mean, color=NAVY, lw=2.4, label="Virtual control (untreated) mean")
axA.plot(months, treated_mean, color=SKY, lw=2.4, ls="--", marker="o", ms=4,
         label="Treated cohort mean (illustrative)")
axA.annotate("", xy=(12, vc12), xytext=(12, TREATED_12),
             arrowprops=dict(arrowstyle="<->", color=RED, lw=1.8))
axA.text(9.1, (vc12 + TREATED_12) / 2, "CARE\n%.3f mm" % CARE, color=RED, fontsize=10,
         ha="right", va="center", fontweight="bold")
axA.set_xlim(0, 12.9); axA.set_ylim(-0.05, 0.5)
axA.set_xticks([0, 3, 6, 9, 12])
axA.set_xlabel("Follow-up (months, actual elapsed time)")
axA.set_ylabel("Cumulative axial elongation (mm)")
axA.set_title("A. Reading the effect against the control's uncertainty", fontsize=11, loc="left")
axA.axhline(0, color="#999999", lw=0.8)
axA.legend(loc="upper left", fontsize=7.8, framealpha=0.95)

# ===== Panel B: the SAME effect's interval, built two ways =====
rows = [("Control uncertainty included\n(recommended)", reco, SKY),
        ("Control treated as fixed\n(understates the interval)", fixed, GREY)]
axB.axvline(0, color=RED, ls=":", lw=1.4)
axB.text(0.004, 1.6, "no effect", color=RED, fontsize=8.5, rotation=90, va="top")
for i, (lab, (lo, hi), col) in enumerate(rows):
    y = 1 - i
    axB.plot([lo, hi], [y, y], color=col, lw=4.2, solid_capstyle="round", zorder=3)
    axB.plot(CARE, y, "o", color=col, ms=10, zorder=4)
    axB.text(hi + 0.007, y, f"[{lo:.3f}, {hi:.3f}]", va="center", fontsize=9.2, color="#333333")
axB.set_yticks([1, 0]); axB.set_yticklabels([r[0] for r in rows], fontsize=9.2)
axB.set_xlim(-0.02, 0.30); axB.set_ylim(-0.7, 1.9)
axB.set_xlabel("Cumulative absolute reduction in elongation, CARE (mm)")
axB.set_title("B. The effect interval: control fixed versus its uncertainty included",
              fontsize=11, loc="left")
axB.grid(axis="y", visible=False)

fig.tight_layout()
fig.savefig("/tmp/fig4_using_the_model.png", dpi=600)
print("CARE=%.4f  vc12=%.4f  treated=%.3f" % (CARE, vc12, TREATED_12))
print("fixed", np.round(fixed, 3), "recommended", np.round(reco, 3))
```
