## Supplementary software v0.2.1 for "Virtual control arms for paediatric myopia trials: external validation of axial-elongation models": make_figs_v2.py

```
"""
make_figs_v2.py -- regenerate the three manuscript figures from the source data,
deterministically, at ACTUAL follow-up time with the pre-specified calibration.

Writes publication-quality figures (600-dpi PNG and vector PDF) into figures/ if
that folder exists, else the current directory. reproduce.py then collects the
PNGs into repro_out/:
    fig1_calibration  -- observed vs predicted untreated elongation, after
                         calibration, by region x visit, with the line of identity.
    fig2_bland_altman -- Bland-Altman agreement (predicted - observed), after
                         calibration, pooled, with mean and +/-1.96 SD.
    fig3_multimodel   -- bias (predicted - observed) per model, by region x visit,
                         with the +/-0.03 mm pre-specified acceptance band.

Timing: actual visit days (the PRIMARY analysis; nominal time is reported only as
the visit-timing contrast). Calibration: the actual-time Indian multiplier from
calibration_v2.json is applied to South Asian eyes; East Asian is not retuned.
Point predictions for East Asian are the published model, unchanged.

DEPENDENCIES: numpy, pandas, matplotlib. Imports analysis_v2 and sensitivity_models.
Self-contained and side-effect-free apart from writing the figures.
"""
from __future__ import annotations
import json
import numpy as np
import pandas as pd
import matplotlib
matplotlib.use("Agg")
import matplotlib.pyplot as plt

import analysis_v2 as A
import sensitivity_models as sm

# ---- deterministic style; colours match the shipped figures ----
NAVY, SKY, RED, GREY = "#1F3864", "#5B9BD5", "#B00020", "#7F7F7F"
BAND = "#E6E6E6"
import os
def _save(fig, stem):
    outdir = "figures" if os.path.isdir("figures") else "."
    fig.savefig(os.path.join(outdir, stem + ".png"), dpi=600)
    fig.savefig(os.path.join(outdir, stem + ".pdf"))
    # keep a cwd PNG too so reproduce.py can collect it into repro_out/
    if outdir != ".":
        fig.savefig(stem + ".png", dpi=600)
    plt.close(fig)

plt.rcParams.update({"figure.dpi": 150, "savefig.dpi": 600, "pdf.fonttype": 42, "ps.fonttype": 42, "font.size": 12,
                     "axes.grid": True, "grid.color": "#DDDDDD", "grid.linewidth": 0.6})

# region x visit series, in the fixed order used by the manuscript legend
SERIES = [
    ("east_asian", 6,  "East Asian, 6 mo",  NAVY, "o", "none"),
    ("east_asian", 12, "East Asian, 12 mo", SKY,  "^", "none"),
    ("european",   6,  "South Asian, 6 mo", RED,  "s", "none"),
]


def _calibrated_frame():
    """Actual-time frame with a calibrated prediction column.
    South Asian (European-routed Indian) eyes get the Indian multiplier from the
    ACTUAL-time calibration (calibration_v2.json), because these figures are the
    actual-follow-up-time view; East Asian eyes are the published model, unchanged.
    Falls back to calibration.json if the v2 file is absent."""
    df = A.load_fixed()
    import os
    calfile = "calibration_v2.json" if os.path.exists("calibration_v2.json") else "calibration.json"
    mult = float(json.load(open(calfile))["indian_multiplier"])
    df = df.copy()
    df["pred_cal"] = A.predict(df, "actual", mult_indian=mult)
    df["diff"] = df["pred_cal"] - df["obs"]   # predicted - observed
    return df, mult


def fig1_calibration(df):
    fig, ax = plt.subplots(figsize=(7.6, 7.0))
    for reg, v, label, col, mk, _ in SERIES:
        g = df[(df.region == reg) & (df.visit == v)]
        ax.scatter(g["obs"], g["pred_cal"], s=34, marker=mk, facecolors="none",
                   edgecolors=col, linewidths=1.2, alpha=0.9,
                   label=f"{label} (n={len(g)})")
    lim = 0.8
    ax.plot([0, lim], [0, lim], ls="--", color=GREY, lw=1.3, label="line of identity")
    ax.set_xlim(0, lim); ax.set_ylim(0, lim)
    ax.set_xlabel("Observed untreated elongation (mm)")
    ax.set_ylabel("Predicted untreated elongation (mm)")
    ax.set_aspect("equal")
    ax.legend(loc="upper left", frameon=False, fontsize=11)
    fig.tight_layout(); _save(fig, "fig1_calibration")


def fig2_bland_altman(df):
    mean_d = float(df["diff"].mean())
    sd = float(df["diff"].std(ddof=1))
    lo, hi = mean_d - 1.96 * sd, mean_d + 1.96 * sd
    fig, ax = plt.subplots(figsize=(8.2, 6.2))
    for reg, v, label, col, mk, _ in SERIES:
        g = df[(df.region == reg) & (df.visit == v)]
        avg = (g["pred_cal"] + g["obs"]) / 2.0
        ax.scatter(avg, g["diff"], s=34, marker=mk, facecolors="none",
                   edgecolors=col, linewidths=1.2, alpha=0.9,
                   label=f"{label} (n={len(g)})")
    ax.axhline(mean_d, color=GREY, lw=1.6)
    ax.axhline(hi, color=GREY, lw=1.2, ls="--")
    ax.axhline(lo, color=GREY, lw=1.2, ls="--")
    xr = ax.get_xlim()[1]
    ax.text(xr, mean_d, f" mean ({mean_d:+.3f})", va="center", ha="right", color=GREY, fontsize=10)
    ax.text(xr, hi, f" +1.96 SD ({hi:+.3f})", va="bottom", ha="right", color=GREY, fontsize=10)
    ax.text(xr, lo, f" -1.96 SD ({lo:+.3f})", va="top", ha="right", color=GREY, fontsize=10)
    ax.set_xlabel("Mean of predicted and observed (mm)")
    ax.set_ylabel("Predicted minus observed (mm)")
    ax.legend(loc="lower left", frameon=False, fontsize=11)
    fig.tight_layout(); _save(fig, "fig2_bland_altman")


def fig3_multimodel(df):
    order = [("naduvilath2025", "Naduvilath 2025 (primary)", NAVY),
             ("brennan2024", "Brennan 2024", SKY),
             ("scorm_wong2010", "SCORM (Wong 2010)", GREY),
             ("olsm_jones2005", "OLSM (Jones 2005)", RED)]
    cells = [("east_asian", 6), ("east_asian", 12), ("european", 6)]
    cell_lab = ["East Asian\n6 mo", "East Asian\n12 mo", "South Asian\n6 mo"]
    bias = {name: [] for name, _, _ in order}
    for reg, v in cells:
        g = df[(df.region == reg) & (df.visit == v)]
        for name, _, _ in order:
            mdl = sm.MODELS[name]
            preds = []
            for r in g.itertuples():
                if not mdl["applies"](r.ser, r.region):
                    preds.append(np.nan); continue
                preds.append(mdl["cum"](r.age, r.ser, r.male, r.region,
                                        round(r.yrs_actual * 12)))
            bias[name].append(float(np.nanmean(np.array(preds) - g["obs"].to_numpy())))
    fig, ax = plt.subplots(figsize=(10.8, 6.4))
    x = np.arange(len(cells)); w = 0.2
    ax.axhspan(-0.03, 0.03, color=BAND, label="+/- 0.03 mm target")
    for i, (name, label, col) in enumerate(order):
        ax.bar(x + (i - 1.5) * w, bias[name], width=w, color=col,
               edgecolor="white", linewidth=0.6, label=label)
    ax.axhline(0, color="black", lw=1.0)
    ax.set_xticks(x); ax.set_xticklabels(cell_lab)
    ax.set_ylabel("Bias, predicted minus observed (mm)")
    ax.grid(axis="x", visible=False)
    handles, labels = ax.get_legend_handles_labels()
    ax.legend(handles, labels, loc="lower left", frameon=False, fontsize=11)
    fig.tight_layout(); _save(fig, "fig3_multimodel")


def main():
    df, mult = _calibrated_frame()
    fig1_calibration(df)
    fig2_bland_altman(df)
    fig3_multimodel(df)
    md = df["diff"].mean(); sd = df["diff"].std(ddof=1)
    print(f"make_figs_v2: wrote fig1_calibration.png, fig2_bland_altman.png, "
          f"fig3_multimodel.png (Indian multiplier={mult:.4f}, "
          f"Bland-Altman mean={md:+.3f}, +/-1.96SD=[{md-1.96*sd:+.3f}, {md+1.96*sd:+.3f}])")


main()
```
