## Supplementary software v0.2.1 for "Virtual control arms for paediatric myopia trials: external validation of axial-elongation models": recalibrate.py

```
"""
recalibrate.py  --  fit and HONESTLY evaluate two calibrations on the untreated
cohort, then emit calibration.json for the released tool.

Two things are calibrated, each for a documented reason found in the validation:

1. PREDICTION-INTERVAL WIDTH (all myopes).
   The published individual 95% interval was too narrow (coverage 0.75 at 6m,
   0.87 at 12m vs the 0.90 pre-registered floor). The empirical residual SD of
   (observed - predicted) is 0.148 mm per sqrt(year) at BOTH visits, i.e. error
   accumulates like a random walk. We set the individual interval half-width to
   z * 0.148 * sqrt(years). Point predictions are unchanged.

2. INDIAN MULTIPLIER (provisional, routed to European).
   European routing under-predicts Indian elongation by ~51% at 6 months. A
   multiplier on the European baseline removes the bias. This is the WEAKEST
   evidence in the study: N=46, 6-month only, single cohort, no Indian 12-month
   data. It is calibrated here and must be externally validated before load-
   bearing use.

The East Asian region (Chinese + Vietnamese, the model's own derivation region)
is NOT retuned: its mean bias is already within the pre-registered threshold and
retuning a model inside its derivation region on a new cohort is not defensible.

HONESTY: every fitted number is also evaluated by stratified 5-fold cross-
validation (fit on 4 folds, score the held-out fold, pool). The CV numbers are
what the paper should quote for out-of-sample behaviour. Final parameters for
the shipped tool are fit on all data.

DEPENDENCIES: numpy, pandas. Imports virtual_control.
"""
from __future__ import annotations
import json
import numpy as np
import pandas as pd
import virtual_control as vc

DATA = "untreated_validation_OD.xlsx"
VISITS = {"AL_m6": 6, "AL_m12": 12}
Z = 1.959963984540054
K = 5
SEED = 20260709


def build_long(path):
    """One row per usable child-visit: id, ethnicity, region, re_group, month,
    years, base_pred (published, multiplier=1), obs."""
    df = pd.read_excel(path)
    subs = [{"id": r.id, "ethnicity": r.ethnicity, "gender": r.gender, "age": r.age,
             "baseline_SER": r.baseline_SER, "baseline_AL": r.baseline_AL}
            for r in df.itertuples() if pd.notna(r.baseline_SER)]
    pred = vc.predict_untreated(subs)[
        ["id", "region", "re_group", "month", "predicted_delta_AL"]]
    pred = pred.rename(columns={"predicted_delta_AL": "base_pred"})
    obs = []
    for r in df.itertuples():
        if pd.isna(r.baseline_AL) or pd.isna(r.baseline_SER):
            continue
        for col, mth in VISITS.items():
            al = getattr(r, col)
            if pd.notna(al):
                obs.append({"id": r.id, "ethnicity": r.ethnicity, "month": mth,
                            "obs": float(al) - float(r.baseline_AL)})
    o = pd.DataFrame(obs)
    m = o.merge(pred, on=["id", "month"])
    m["years"] = m["month"] / 12.0
    return m


def fit_indian_multiplier(m):
    """Ratio of summed observed to summed predicted elongation for Indian eyes
    (elongation-weighted, so it is a proper rate ratio, not a mean of ratios)."""
    ind = m[m["ethnicity"] == "Indian"]
    if len(ind) == 0 or ind["base_pred"].sum() <= 0:
        return 1.0
    return float(ind["obs"].sum() / ind["base_pred"].sum())


def fit_pi_sigma(m, group="myope"):
    """Empirical residual SD per sqrt(year), pooled over visits for the given
    re_group. resid = obs - pred_with_multiplier."""
    g = m[m["re_group"] == group].copy()
    if len(g) < 3:
        return np.nan
    # standardise each residual to a per-sqrt-year scale, then take the SD
    z = (g["obs"] - g["pred"]) / np.sqrt(g["years"])
    return float(z.std(ddof=1))


def apply_multiplier(m, mult_indian):
    m = m.copy()
    m["mult"] = np.where(m["ethnicity"] == "Indian", mult_indian, 1.0)
    m["pred"] = m["base_pred"] * m["mult"]
    return m


def metrics(m, sigma_myope):
    """Bias and PI coverage by region x visit, using calibrated PI."""
    out = []
    for (reg, mth), g in m.groupby(["region", "month"]):
        yrs = mth / 12.0
        half = Z * sigma_myope * np.sqrt(yrs)      # myope cohort
        lo = g["pred"] - half
        hi = g["pred"] + half
        cov = float(((g["obs"] >= lo) & (g["obs"] <= hi)).mean())
        bias = float((g["pred"] - g["obs"]).mean())
        out.append({"region": reg, "month": int(mth), "n": len(g),
                    "bias": round(bias, 4), "coverage": round(cov, 3)})
    return pd.DataFrame(out).sort_values(["region", "month"])


def stratified_folds(ids, strata, k, seed):
    rng = np.random.default_rng(seed)
    ids = np.asarray(ids, dtype=object)
    strata = np.asarray(strata, dtype=object)
    fold = {}
    for s in sorted(set(strata.tolist())):
        members = list(ids[strata == s])
        rng.shuffle(members)
        for i, cid in enumerate(members):
            fold[cid] = i % k
    return fold


def cross_validate(m, k=K, seed=SEED):
    """Fit multiplier + sigma on train folds, score held-out fold, pool rows."""
    child = m[["id", "ethnicity"]].drop_duplicates()
    fold = stratified_folds(child["id"].tolist(), child["ethnicity"].tolist(), k, seed)
    m = m.copy()
    m["fold"] = m["id"].map(fold)
    scored = []
    for f in range(k):
        tr, te = m[m["fold"] != f], m[m["fold"] == f]
        mult = fit_indian_multiplier(tr)
        tr_m = apply_multiplier(tr, mult)
        sigma = fit_pi_sigma(tr_m, "myope")
        te_m = apply_multiplier(te, mult)
        te_m["sigma"] = sigma
        scored.append(te_m)
    s = pd.concat(scored, ignore_index=True)
    rows = []
    for (reg, mth), g in s.groupby(["region", "month"]):
        yrs = mth / 12.0
        half = Z * g["sigma"] * np.sqrt(yrs)
        cov = float(((g["obs"] >= g["pred"] - half) & (g["obs"] <= g["pred"] + half)).mean())
        bias = float((g["pred"] - g["obs"]).mean())
        rows.append({"region": reg, "month": int(mth), "n": len(g),
                     "cv_bias": round(bias, 4), "cv_coverage": round(cov, 3)})
    return pd.DataFrame(rows).sort_values(["region", "month"])


def bootstrap_multiplier(m, n=5000, seed=SEED):
    ind = m[m["ethnicity"] == "Indian"]
    ids = ind["id"].unique()
    rng = np.random.default_rng(seed)
    vals = []
    by_id = {i: ind[ind["id"] == i] for i in ids}
    for _ in range(n):
        pick = rng.choice(ids, size=len(ids), replace=True)
        g = pd.concat([by_id[i] for i in pick])
        if g["base_pred"].sum() > 0:
            vals.append(g["obs"].sum() / g["base_pred"].sum())
    return float(np.percentile(vals, 2.5)), float(np.percentile(vals, 97.5))


def main():
    m = build_long(DATA)

    # interpretation: age by ethnicity (why Indian may differ)
    df = pd.read_excel(DATA)
    age_by_eth = df.groupby("ethnicity")["age"].mean().round(2).to_dict()

    # ---- fit on ALL data (shipped parameters) ----
    mult = fit_indian_multiplier(m)
    m_all = apply_multiplier(m, mult)
    sigma_my = fit_pi_sigma(m_all, "myope")
    lo, hi = bootstrap_multiplier(m)

    print("=== COHORT ===")
    print("mean age by ethnicity:", age_by_eth)
    print("myopes:", int((df['baseline_SER'] <= -0.5).sum()),
          " non-myopes:", int((df['baseline_SER'] > -0.5).sum()))
    print()
    print("=== FITTED (all data) ===")
    print(f"Indian multiplier = {mult:.3f}   95% CI [{lo:.3f}, {hi:.3f}]  (6-month, N=46, provisional)")
    print(f"myope PI sigma    = {sigma_my:.4f} mm per sqrt(year)")
    print("non-myope PI sigma = (unchanged; cohort has no non-myopes)")
    print()

    print("=== IN-SAMPLE after calibration (by region x visit) ===")
    print(metrics(m_all, sigma_my).to_string(index=False))
    print()
    print("=== CROSS-VALIDATED (out-of-fold; quote THIS in the paper) ===")
    cv = cross_validate(m)
    print(cv.to_string(index=False))
    print()

    calib = {
        "source_cohort": DATA,
        "n_children": int(m["id"].nunique()),
        "indian_multiplier": round(mult, 4),
        "indian_multiplier_ci95": [round(lo, 4), round(hi, 4)],
        "indian_multiplier_caveat": "6-month only, N=46, single cohort, provisional; externally validate before load-bearing use.",
        "pi_sigma_myope_per_sqrt_year": round(sigma_my, 4),
        "pi_time_scaling": "sqrt",
        "pi_nonmyope": "unchanged (no non-myopes in calibration cohort)",
        "seed": SEED, "k_folds": K,
    }
    with open("calibration.json", "w") as fh:
        json.dump(calib, fh, indent=2)
    print("Wrote calibration.json")
    return calib


if __name__ == "__main__":
    main()
```
