## Supplementary software v0.2.1 for "Virtual control arms for paediatric myopia trials: external validation of axial-elongation models": reproduce.py

```
"""
reproduce.py -- one command to regenerate every number, table, and figure in the
manuscript from the source data, deterministically.

    python reproduce.py

Writes to ./repro_out/:
    table1_cohort.xlsx, table2_primary.xlsx, table3_calibrated.xlsx,
    table4_multimodel.xlsx, inference.txt, pm.txt,
    fig1_calibration.png, fig2_bland_altman.png, fig3_multimodel.png
and prints a manifest. Fixed seed; no network; no Date/random nondeterminism.
"""
import os, io, json, contextlib, numpy as np, pandas as pd
import analysis_v2 as A
import virtual_control as vc

OUT = "repro_out"; os.makedirs(OUT, exist_ok=True)
df = A.load_fixed()

# Two timing bases are reported explicitly and never mixed (see the paper):
#   PRIMARY  : actual visit days        -> calibration_v2.json  (analysis_v2.py)
#   CONTRAST : nominal 6/12-month time  -> calibration.json     (recalibrate.py)
# analysis_v2.main() writes calibration_v2.json; run it FIRST so the downstream
# actual-time steps (compute_inference, make_figs_v2) read a consistent file.
buf0 = io.StringIO()
with contextlib.redirect_stdout(buf0):
    A.main()
open(f"{OUT}/analysis_v2_full.txt", "w").write(buf0.getvalue())

cal_primary = json.load(open("calibration_v2.json"))       # actual time (primary)
cal_contrast = json.load(open("calibration.json"))         # nominal time (contrast)
MULT, SIG = cal_primary["indian_multiplier"], cal_primary["pi_sigma_myope_per_sqrt_year"]

# Table 1 (cohort) from the raw export
raw = pd.read_excel(A.XLSX)
raw["gender"] = raw["Sex"].map(A.SEXMAP)
def row(g):
    return dict(n=len(g), age=f"{g.BL_age.mean():.1f} ({g.BL_age.std():.1f})",
                ser=f"{g.BL_SER_OD.mean():.2f} ({g.BL_SER_OD.std():.2f})",
                al=f"{g.BL_AL_OD.mean():.2f} ({g.BL_AL_OD.std():.2f})",
                female=int((g.gender == 'F').sum()),
                n6=int(g['6m_AL_OD'].notna().sum()), n12=int(g['12m_AL_OD'].notna().sum()))
t1 = pd.DataFrame({e: row(raw[raw.Ethnicity == e]) for e in ['Chinese', 'Vietnamese', 'Indian']}).T
t1.loc['All'] = row(raw)
t1.to_excel(f"{OUT}/table1_cohort.xlsx")

# Tables 2 & 3 via compute_inference (captures its printed block too)
buf = io.StringIO()
with contextlib.redirect_stdout(buf):
    import compute_inference  # noqa: F401  (runs on import)
open(f"{OUT}/inference.txt", "w").write(buf.getvalue())

# Table 2 primary (published, actual time)
rows2 = []
for (reg, v), g in df.groupby(["region", "visit"]):
    p = A.predict(g, "actual", 1.0); err = p - g["obs"].to_numpy()
    rows2.append(dict(region=reg, visit=v, n=len(g),
                      bias=round(err.mean(), 4),
                      rmse=round(float(np.sqrt((err**2).mean())), 4)))
pd.DataFrame(rows2).to_excel(f"{OUT}/table2_primary.xlsx", index=False)

# Table 3 (calibrated + CV) is captured above from analysis_v2.main(); its full
# output is already written to analysis_v2_full.txt.

# Figures (actual-time sensitivity view; use calibration_v2.json)
import make_figs_v2  # noqa: F401 (writes fig1/2/3 to cwd)
for f in ["fig1_calibration.png", "fig2_bland_altman.png", "fig3_multimodel.png"]:
    if os.path.exists(f):
        os.replace(f, f"{OUT}/{f}")

print("=== reproduce.py complete ===")
for f in sorted(os.listdir(OUT)):
    print("  ", os.path.join(OUT, f))
print("timing bases reported (never mixed):")
print(f"  PRIMARY  (actual visit days): multiplier={cal_primary['indian_multiplier']} "
      f"sigma={cal_primary['pi_sigma_myope_per_sqrt_year']}  [calibration_v2.json]")
print(f"  CONTRAST (nominal 6/12 mo):   multiplier={cal_contrast['indian_multiplier']} "
      f"sigma={cal_contrast['pi_sigma_myope_per_sqrt_year']}  [calibration.json]")
```
