## Supplementary software v0.2.1 for "Virtual control arms for paediatric myopia trials: external validation of axial-elongation models": run_my_trial.py

```
"""
run_my_trial.py  --  fill in input_template.xlsx, then run:  python run_my_trial.py

Reads input_template.xlsx (one row per patient, baseline columns), builds the
virtual untreated control from BASELINE data, and writes:
  out_predictions.xlsx  -- per patient per visit: predicted_delta_AL with its
                          mean confidence band (ci_low/ci_high)

Choose the model with --model (one per run; run again to compare):
  naduvilath2025 (default, regional GEE, ships a confidence interval)
  brennan2024    (meta-regression, myopes only, point estimate)
  olsm_jones2005 (age-only, myopes only, point estimate)
  scorm_wong2010 (age-only, myopes only, point estimate)
Only the GEE ships a confidence interval of the mean; the other models return a
point estimate because their sources publish no parameter covariance.

This script builds the control arm only. It does NOT compute a treatment effect.
Compare your treated outcomes against this control yourself, following
docs/ANALYSIS_PROTOCOL.md, so your outcome data never has to leave your machine.
"""
import sys
import pandas as pd
import virtual_control as vc

WIDE = "input_template.xlsx"


def _read(path):
    return pd.read_excel(path)


def _pick_model(argv):
    """Model from '--model NAME' or '--model=NAME'; default naduvilath2025."""
    name = "naduvilath2025"
    for i, a in enumerate(argv):
        if a == "--model" and i + 1 < len(argv):
            name = argv[i + 1]
        elif a.startswith("--model="):
            name = a.split("=", 1)[1]
    name = name.strip().lower()
    if name not in vc.MODEL_LABELS:
        raise SystemExit("Unknown --model %r; choose one of: %s"
                         % (name, ", ".join(vc.MODEL_LABELS)))
    return name


def main(model_name="naduvilath2025"):
    df = _read(WIDE)

    # baseline -> subjects for the model (treated follow-up columns are ignored;
    # this tool provides the control, not the effect). Ancestry is REQUIRED and
    # is the child's ethnicity, not country of residence; blank rows are not
    # predicted. Ancestries with no derived model need a 'stratum' choice.
    subjects, excl = [], []
    for r in df.itertuples():
        eth = getattr(r, "ethnicity", None)
        if pd.isna(eth) or str(eth).strip() == "":
            excl.append({"id": getattr(r, "id", "(no id)"),
                         "reason": "ethnicity (ancestry) is required; country of "
                                   "residence is not accepted"})
            continue
        subjects.append(
            {"id": r.id, "ethnicity": eth, "gender": r.gender,
             "age": r.age, "baseline_SER": r.baseline_SER, "baseline_AL": r.baseline_AL,
             "stratum": getattr(r, "stratum", None)})

    # one model per run (the user's choice); the GEE ships a CI, the others a point
    pred = vc.predict_untreated(subjects, exclusions=excl, model_name=model_name)
    vc.write_output(pred, "out_predictions.xlsx",
                    model_label=vc.MODEL_LABELS[model_name])
    print("Model: {} ({})".format(
        vc.MODEL_LABELS[model_name],
        "confidence interval of the mean" if model_name == "naduvilath2025"
        else "point estimate; no published confidence interval"))
    if excl:
        vc.write_output(pd.DataFrame(excl), "out_needs_input.xlsx",
                        model_label=vc.MODEL_LABELS[model_name],
                        sheet_name="needs_input")
        print("NOTE: {} row(s) were NOT predicted (see out_needs_input.xlsx): a missing "
              "ethnicity, an ancestry\nwith no derived model that needs a 'stratum' "
              "choice (east_asian or european), or a non-myope\nsent to a myope-only "
              "model.\n".format(len(excl)))

    if pred.empty:
        print("No children were predicted; see out_needs_input.xlsx for what each row needs.")
        return
    print("VIRTUAL UNTREATED CONTROL (predicted change with its mean confidence "
          "band), first patient:")
    cols = ["id", "region", "model_status", "month",
            "predicted_delta_AL", "ci_low", "ci_high"]
    print(pred[pred.id == pred.id.iloc[0]][cols].to_string(index=False))
    print("\nWritten: out_predictions.xlsx")
    print("\nThis tool stops at the control. To estimate the treatment effect, "
          "compare your\ntreated outcomes against these predictions following "
          "docs/ANALYSIS_PROTOCOL.md\n(it shows how to carry the control's "
          "uncertainty into the effect estimate).")


if __name__ == "__main__":
    main(_pick_model(sys.argv[1:]))
```
