## Supplementary software v0.2.1 for "Virtual control arms for paediatric myopia trials: external validation of axial-elongation models": sensitivity_models.py

```
"""
sensitivity_models.py  --  multi-model virtual control (primary + sensitivity).

An externally-controlled design is only convincing if the result holds across
several independent normative models. This module runs the primary model AND
published sensitivity models, then reports concordance of the treatment effect
across them. Self-contained (numpy + pandas only); does not import
virtual_control, so it can be tested independently.

Models (all exact published equations; each validated against its source):

  naduvilath2025  PRIMARY. Regional GEE, East Asian / European, myope + non-myope.
                  Naduvilath et al. Ophthalmic Physiol Opt 2025;45:135-151, Table 3.
  brennan2024     Meta-regression of 78 studies; Asian vs non-Asian, age only,
                  myopes. ln(rate)=0.362-0.158*age+0.325*race (race=1 Asian).
                  Brennan et al. Optom Vis Sci 2024;101:497-507, Table 1.
  olsm_jones2005  OLSM AXIAL LENGTH model, predominantly White US myopes (age
                  only). AL=18.144+2.391*ln(age) (<=10.5y) else 17.808+2.560*ln(age),
                  so rate=dAL/dage=2.391/age (<=10.5y) else 2.560/age.
                  Jones et al. IOVS 2005;46:2317-2327, Table 4 (Axial length, Myopes).
  scorm_wong2010  SCORM AXIAL LENGTH model, Singaporean myopes (age only).
                  AL=20.61+0.163*age^2-0.053*age^2*ln(age), so
                  rate=dAL/dage=0.273*age-0.106*age*ln(age).
                  Wong et al. IOVS 2010;51:1341-1347, Table 2 (AL, persistent myopia).

Notes on use as SENSITIVITY models:
- OLSM and SCORM are older and ultrasound-era, but each source publishes an
  axial-length growth model, so the tool uses the published AL curve directly
  (differentiated to an annual elongation rate); no VCD proxy is used.
- OLSM/SCORM/Brennan are MYOPE models: they return NaN for non-myopes
  (baseline SER > -0.50 D); the primary Naduvilath 2025 covers non-myopes.
- Concordance across models is the point: if the effect is similar under all
  applicable models, the finding is robust to model choice.
"""
import numpy as np
import pandas as pd

MONTHS = (6, 12, 18, 24, 36)

# ---- minimal country/ethnicity -> region map (east_asian | european) ----
_REGION = {
    "chinese": "east_asian", "china": "east_asian", "han": "east_asian",
    "vietnamese": "east_asian", "vietnam": "east_asian", "taiwanese": "east_asian",
    "singaporean": "east_asian", "malaysian": "east_asian", "east asian": "east_asian",
    "asian": "east_asian",
    "australian": "european", "european": "european", "white": "european",
    "caucasian": "european", "uk": "european", "british": "european",
    "swedish": "european", "portuguese": "european", "spanish": "european",
    "new zealand": "european", "indian": "european", "south asian": "european",
}
def region_of(country):
    return _REGION.get(str(country).strip().lower(), "european")  # default european


# ============================ model rate functions ============================
def _cum(rate_fn, age0, month):
    """Cumulative elongation from age0 over `month` months. Whole months are
    integrated monthly (identical to before); a fractional remainder is added as
    a partial month, so actual elapsed follow-up time is honoured, not rounded."""
    whole = int(month)
    total = sum(rate_fn(age0 + m / 12.0) / 12.0 for m in range(whole))
    frac = float(month) - whole
    if frac > 0:
        total += rate_fn(age0 + whole / 12.0) / 12.0 * frac
    return float(total)


def _naduvilath2025_rate(age, ser, male, region):
    ea = 1.0 if region == "east_asian" else 0.0
    if ser <= -0.50:
        y = 0.537 + 0.147 * ea + 0.004 * male - 0.032 * age - 0.007 * ea * age - 0.007 * ser
    else:
        y = 0.263 + 0.136 * ea - 0.003 * male - 0.016 * age - 0.007 * ea * age - 0.002 * ser
    return np.exp(y) - 1.0


def _brennan2024_rate(age, region):
    race = 1.0 if region == "east_asian" else 0.0
    return float(np.exp(0.362 - 0.158 * age + 0.325 * race))


def _olsm_rate(age):
    return (2.391 if age <= 10.5 else 2.560) / age


def _scorm_rate(age):
    # Wong 2010 SCORM, Table 2, persistent-myopia AXIAL LENGTH model:
    #   AL(age) = 20.61 + 0.163*age^2 - 0.053*age^2*ln(age)
    # Annual axial elongation is the derivative dAL/d(age):
    #   0.326*age - 0.053*(2*age*ln(age) + age) = 0.273*age - 0.106*age*ln(age)
    return max(0.273 * age - 0.106 * age * np.log(age), 0.0)


# model registry: name -> dict(cumulative, applies, meta)
MODELS = {
    "naduvilath2025": {
        "role": "primary",
        "source": "Naduvilath 2025 (regional EA/EU)",
        "applies": lambda ser, region: True,
        "cum": lambda age0, ser, male, region, M: _cum(
            lambda a: _naduvilath2025_rate(a, ser, male, region), age0, M),
    },
    "brennan2024": {
        "role": "sensitivity",
        "source": "Brennan 2024 meta-regression (Asian/non-Asian, myopes)",
        "applies": lambda ser, region: ser <= -0.50,
        "cum": lambda age0, ser, male, region, M: _cum(
            lambda a: _brennan2024_rate(a, region), age0, M),
    },
    "olsm_jones2005": {
        "role": "sensitivity",
        "source": "OLSM / Jones 2005 (White US myopes)",
        "applies": lambda ser, region: ser <= -0.50,
        "cum": lambda age0, ser, male, region, M: _cum(_olsm_rate, age0, M),
    },
    "scorm_wong2010": {
        "role": "sensitivity",
        "source": "SCORM / Wong 2010 (Singaporean myopes)",
        "applies": lambda ser, region: ser <= -0.50,
        "cum": lambda age0, ser, male, region, M: _cum(_scorm_rate, age0, M),
    },
}


# ================================ public API =================================
def predict_all_models(subjects, months=MONTHS):
    """Per subject, per visit, per applicable model: predicted untreated elongation.
    subjects: list of dicts with id, age, gender, baseline_SER, country."""
    rows = []
    for s in subjects:
        sid = s.get("id")
        age = float(s["age"])
        ser = float(s.get("baseline_SER", -2.0))
        male = 1.0 if str(s.get("gender", "F")).upper().startswith("M") else 0.0
        region = region_of(s.get("country") or s.get("ethnicity") or "")
        for name, mdl in MODELS.items():
            if not mdl["applies"](ser, region):
                continue
            for M in months:
                rows.append({"id": sid, "region": region, "month": M,
                             "model": name, "role": mdl["role"],
                             "predicted": round(mdl["cum"](age, ser, male, region, M), 4)})
    return pd.DataFrame(rows)


def compare_models(subjects, observed, months=MONTHS):
    """Group-level treatment effect (CARE) under each model, plus a concordance
    table showing how much the effect varies across models per visit.
    observed: DataFrame id, month, observed_elongation."""
    pred = predict_all_models(subjects, months)
    obs_mean = observed.groupby("month")["observed_elongation"].mean()

    rows = []
    for (name, M), g in pred.groupby(["model", "month"]):
        if M not in obs_mean.index:
            continue
        mp = g["predicted"].mean()
        mo = float(obs_mean.loc[M])
        rows.append({"model": name, "role": MODELS[name]["role"], "month": int(M),
                     "n": g["id"].nunique(),
                     "mean_predicted_untreated": round(mp, 4),
                     "mean_observed": round(mo, 4),
                     "CARE_mm": round(mp - mo, 4)})
    per_model = pd.DataFrame(rows).sort_values(["month", "role", "model"]).reset_index(drop=True)

    # concordance: spread of CARE across models at each visit
    conc = []
    for M, g in per_model.groupby("month"):
        care = g["CARE_mm"]
        prim = g.loc[g["role"] == "primary", "CARE_mm"]
        conc.append({"month": int(M),
                     "CARE_primary": round(float(prim.iloc[0]), 4) if len(prim) else np.nan,
                     "CARE_min": round(float(care.min()), 4),
                     "CARE_max": round(float(care.max()), 4),
                     "CARE_range": round(float(care.max() - care.min()), 4),
                     "n_models": len(g)})
    concordance = pd.DataFrame(conc).sort_values("month").reset_index(drop=True)
    return per_model, concordance


def validate():
    """Self-check each model against its published source numbers."""
    ok = True
    checks = [
        ("Brennan 8y Asian cum->15", _cum(lambda a: _brennan2024_rate(a, "east_asian"), 8, 7 * 12), 2.38, 0.03),
        ("Brennan annual @10.5 Asian", _brennan2024_rate(10.5, "east_asian"), 0.38, 0.02),
        ("OLSM annual @8", _olsm_rate(8), 0.30, 0.02),
        ("OLSM annual @12", _olsm_rate(12), 0.21, 0.02),
        ("SCORM(AL) annual @9", _scorm_rate(9), 0.36, 0.02),
        ("Naduvilath2025 EA myope @10 -2D", _naduvilath2025_rate(10, -2, 0.5, "east_asian"), 0.36, 0.02),
    ]
    print("Sensitivity-model validation:")
    for label, got, want, tol in checks:
        good = abs(got - want) <= tol
        ok = ok and good
        print("  [{}] {}: {:.3f} (source ~{:.2f})".format("PASS" if good else "FAIL", label, got, want))
    return ok


if __name__ == "__main__":
    validate()
    subjects = [
        {"id": "S1", "age": 9.0, "gender": "F", "baseline_SER": -2.0, "country": "Chinese"},
        {"id": "S2", "age": 10.5, "gender": "M", "baseline_SER": -3.0, "country": "Australian"},
        {"id": "S3", "age": 8.0, "gender": "F", "baseline_SER": -1.5, "country": "Indian"},
    ]
    observed = pd.DataFrame([
        {"id": "S1", "month": 12, "observed_elongation": 0.20},
        {"id": "S2", "month": 12, "observed_elongation": 0.16},
        {"id": "S3", "month": 12, "observed_elongation": 0.14},
    ])
    per_model, concordance = compare_models(subjects, observed, months=(12,))
    print("\nEffect under each model (12 months):")
    print(per_model.to_string(index=False))
    print("\nConcordance across models:")
    print(concordance.to_string(index=False))
```
