## Supplementary software v0.2.1 for "Virtual control arms for paediatric myopia trials: external validation of axial-elongation models": validate_model.py

```
"""
validate_model.py  --  validate the virtual-control model against YOUR untreated data.

This is the credibility anchor for the paper. Feed it real UNTREATED children
(children who wore single-vision / no myopia-control), and it checks how well the
model's predicted untreated elongation matches what actually happened, per visit
and per population/region.

    Fill untreated_validation_template.xlsx, then run:
        python validate_model.py

It writes:
    validation_by_visit.xlsx        -- metrics per visit (all children)
    validation_by_region_visit.xlsx -- metrics per region x visit (catches population bias)
    validation_calibration.png     -- observed vs predicted, with the line of identity
and prints a PASS/FAIL against pre-specified thresholds.

Metrics reported
----------------
  n, mean_pred, mean_obs
  bias      = mean(predicted - observed)      -> systematic error; ideal ~0
  RMSE, MAE = typical size of the per-child error
  LoA_low/high = 95% limits of agreement (Bland-Altman)
  PI_coverage  = fraction of observed values inside the model's 95% prediction
                 interval; ideal ~0.95, threshold >= 0.90
  calib_slope, calib_intercept = regression of observed on predicted;
                 ideal slope 1, intercept 0

DEPENDENCIES: numpy, pandas, matplotlib.  Imports virtual_control.py.
"""
import numpy as np
import pandas as pd
import matplotlib
matplotlib.use("Agg")
import matplotlib.pyplot as plt

import os
import virtual_control as vc

# The filled validation file lives in the sibling "data to fill in" folder.
# Look there first, then in the current folder, then the working directory.
_HERE = os.path.dirname(os.path.abspath(__file__))
def _find_untreated_data():
    candidates = [
        os.path.join(_HERE, "..", "data to fill in", "untreated_validation_template.xlsx"),
        os.path.join(_HERE, "untreated_validation_template.xlsx"),
        "untreated_validation_template.xlsx",
    ]
    for c in candidates:
        if os.path.exists(c):
            return c
    return candidates[0]  # default path even if missing (clear error on read)

UNTREATED_DATA = _find_untreated_data()
# This validation is limited to 6- and 12-month untreated data (the follow-up we
# have). 18/24/36-month validation is left as a stated limitation for the paper.
VISIT_COLS = {"AL_m6": 6, "AL_m12": 12}
Z = 1.959963984540054

# pre-specified acceptance thresholds (edit and PRE-REGISTER before you look at data)
THRESHOLDS = {"bias_abs_max": 0.03, "coverage_min": 0.90}   # applied at each visit


def _wilson(k, n, z=Z):
    """Wilson score interval for a binomial proportion k/n."""
    if n == 0:
        return (0.0, 0.0)
    p = k / n
    d = 1.0 + z * z / n
    centre = p + z * z / (2 * n)
    half = z * ((p * (1 - p) / n + z * z / (4 * n * n)) ** 0.5)
    return ((centre - half) / d, (centre + half) / d)


def load(path, exclusions=None):
    """Return a long dataframe: id, region, model_status, month, predicted, pi_low,
    pi_high, observed  (observed elongation = follow-up AL - baseline AL).

    Real cohorts have incomplete baselines. A child-visit can only enter the
    validation if it has (a) a baseline SER so the model can PREDICT, and (b) a
    baseline AL plus that follow-up AL so we can OBSERVE the elongation. Child-
    visits missing either are excluded here with a logged reason (missing
    baseline, NOT an outcome-based drop). Pass a list as `exclusions` to collect
    the manifest; it is also written to validation_exclusions.xlsx by the caller.
    """
    df = pd.read_excel(path)
    log = exclusions if exclusions is not None else []

    def _eth(r):
        return getattr(r, "ethnicity", None) or getattr(r, "country", None)

    # PREDICT: only subjects with a usable baseline SER (needed by the model).
    subjects = []
    for r in df.itertuples():
        if pd.isna(getattr(r, "baseline_SER", np.nan)):
            log.append({"id": r.id, "reason": "missing baseline_SER (cannot predict)"})
            continue
        subjects.append({"id": r.id, "ethnicity": _eth(r), "gender": r.gender,
                         "age": r.age, "baseline_SER": r.baseline_SER,
                         "baseline_AL": r.baseline_AL})
    pred = vc.predict_untreated(subjects, include_pi=True, require_stratum=False)  # coverage needs the PI; validation uses the registry region

    # OBSERVE: only child-visits with baseline AL and this follow-up AL present.
    obs_rows = []
    for r in df.itertuples():
        base = getattr(r, "baseline_AL", np.nan)
        for col, mth in VISIT_COLS.items():
            al = getattr(r, col, None)
            if pd.isna(al):
                continue
            if pd.isna(base):
                log.append({"id": r.id, "reason": f"missing baseline_AL "
                            f"(cannot observe {col})"})
                continue
            obs_rows.append({"id": r.id, "month": mth,
                             "observed": float(al) - float(base)})
    observed = pd.DataFrame(obs_rows)

    m = pred.merge(observed, on=["id", "month"], how="inner")
    m = m.rename(columns={"predicted_delta_AL": "predicted"})
    # Any residual non-finite (defensive) is dropped and logged.
    bad = m[m["predicted"].isna() | m["observed"].isna()]
    for r in bad.itertuples():
        log.append({"id": r.id, "reason": "non-finite predicted/observed"})
    m = m[m["predicted"].notna() & m["observed"].notna()].copy()
    m["err"] = m["predicted"] - m["observed"]
    m["in_PI"] = (m["observed"] >= m["pi_low"]) & (m["observed"] <= m["pi_high"])
    return m


def _metrics(g):
    err = g["err"].to_numpy(float)
    n = len(g)
    if n >= 2 and g["predicted"].std() > 0:
        slope, inter = np.polyfit(g["predicted"].to_numpy(float), g["observed"].to_numpy(float), 1)
        sd = err.std(ddof=1)
    else:
        slope = inter = sd = np.nan
    return {
        "n": n,
        "mean_pred": round(float(g["predicted"].mean()), 3),
        "mean_obs": round(float(g["observed"].mean()), 3),
        "bias": round(float(err.mean()), 3),
        "RMSE": round(float(np.sqrt((err ** 2).mean())), 3),
        "MAE": round(float(np.abs(err).mean()), 3),
        "LoA_low": round(float(err.mean() - 1.96 * sd), 3) if np.isfinite(sd) else np.nan,
        "LoA_high": round(float(err.mean() + 1.96 * sd), 3) if np.isfinite(sd) else np.nan,
        "PI_coverage": round(float(g["in_PI"].mean()), 2),
        "calib_slope": round(float(slope), 2) if np.isfinite(slope) else np.nan,
        "calib_intercept": round(float(inter), 3) if np.isfinite(inter) else np.nan,
    }


def _grouped(m, keys):
    rows = []
    for key, g in m.groupby(keys):
        rec = dict(zip(keys if isinstance(keys, list) else [keys],
                       key if isinstance(key, tuple) else [key]))
        rec.update(_metrics(g))
        rows.append(rec)
    return pd.DataFrame(rows)


def plots(m):
    fig, ax = plt.subplots(figsize=(5.6, 4.8))
    colors = {"east_asian": "#1F3864", "european": "#B23A48"}
    for reg, g in m.groupby("region"):
        c = colors.get(reg, "#4C9F70")
        ax.scatter(g["predicted"], g["observed"], s=12, alpha=.55, color=c, label=reg)
    lim = [0, float(m["predicted"].max()) * 1.1]
    ax.plot(lim, lim, "k--", lw=1, label="identity")
    ax.set_title("Calibration: observed vs predicted untreated elongation")
    ax.set_xlabel("predicted untreated elongation (mm)")
    ax.set_ylabel("observed untreated elongation (mm)")
    ax.legend(fontsize=8)
    plt.tight_layout()
    fig.savefig("validation_calibration.png", dpi=140)
    plt.close(fig)


def run_validation(path=UNTREATED_DATA):
    exclusions = []
    m = load(path, exclusions=exclusions)
    excl_df = pd.DataFrame(exclusions)
    excl_df.to_excel("validation_exclusions.xlsx", index=False)
    if len(excl_df):
        print("=== EXCLUDED child-visits (missing baseline; logged, not outcome-based) ===")
        print(excl_df.to_string(index=False))
        print()
    by_visit = _grouped(m, "month").sort_values("month")
    by_region = _grouped(m, ["region", "month"]).sort_values(["region", "month"])
    by_visit.to_excel("validation_by_visit.xlsx", index=False)
    by_region.to_excel("validation_by_region_visit.xlsx", index=False)
    plots(m)

    print("=== VALIDATION: overall by visit ===")
    print(by_visit.to_string(index=False))
    print("\n=== VALIDATION: by region x visit (this catches population-specific bias) ===")
    print(by_region.to_string(index=False))

    # Acceptance is judged on the CONFIDENCE BOUNDS, not the point estimates,
    # matching the paper's rule: bias equivalence needs the whole 95% bias CI
    # inside +/- the margin (TOST logic), and coverage is judged against the
    # Wilson interval so a value whose CI still spans the 0.90 floor is
    # INDETERMINATE (more data needed), not a hard fail.
    print("\n=== PASS / FAIL by region at each validated visit "
          "(bias 95% CI within +/-{:.02g} and coverage Wilson CI >= {:.0%}) ===".format(
              THRESHOLDS["bias_abs_max"], THRESHOLDS["coverage_min"]))
    for (reg, mth), g in m.groupby(["region", "month"]):
        e = g["err"].to_numpy(dtype=float)
        n = len(e)
        mean_b = float(e.mean())
        se = e.std(ddof=1) / (n ** 0.5) if n > 1 else float("nan")
        blo, bhi = mean_b - Z * se, mean_b + Z * se
        bias_ok = (blo > -THRESHOLDS["bias_abs_max"]) and (bhi < THRESHOLDS["bias_abs_max"])
        k = int(g["in_PI"].sum())
        cov = k / n
        wlo, whi = _wilson(k, n)
        cov_ok = wlo >= THRESHOLDS["coverage_min"]
        cov_bad = whi < THRESHOLDS["coverage_min"]
        if bias_ok and cov_ok:
            verdict = "PASS"
        elif (not bias_ok) or cov_bad:
            verdict = "FAIL -> recalibrate this region with fit_population_multiplier()"
        else:
            verdict = "INDETERMINATE (CI spans the threshold; more data needed)"
        print("  {:<12} {:>3d}m  n={:<4d} bias={:+.3f} [{:+.3f},{:+.3f}]  "
              "coverage={:.2f} [{:.2f},{:.2f}]  -> {}".format(
                  reg, int(mth), n, mean_b, blo, bhi, cov, wlo, whi, verdict))
    print("\nArtefacts: validation_by_visit.xlsx, validation_by_region_visit.xlsx, "
          "validation_calibration.png")
    return by_visit, by_region


if __name__ == "__main__":
    run_validation()
```
