## Supplementary software v0.2.1 for "Virtual control arms for paediatric myopia trials: external validation of axial-elongation models": virtual_control.py

```
"""
virtual_control.py  --  one-file, multi-population virtual (untreated) control
for a myopia trial.

WHAT IT DOES
------------
1. You pass in an array of subjects (baseline details only), each tagged with a
   country or ethnicity.
2. The code routes each subject to the correct region in a real published model,
   then returns the PREDICTED UNTREATED axial elongation (mm) at 6, 12, 18, 24,
   36 months: one point number per timepoint, plus an interval, plus which model
   region was used and how good that mapping is.
3. You pass in your treated arm's observed elongation and it runs the stats
   (treated minus predicted-untreated) with a 95% CI, a p-value, and the CARE
   metric.

THE MODEL (real, published, multi-region)
-----------------------------------------
Default: Naduvilath et al. 2025 GEE model of axial elongation, fitted jointly on
East Asian (China, Vietnam) and European (UK, Sweden, Australia) children, with
EXACT coefficients for both regions (Ophthalmic Physiol Opt 45:135-151, Table 3).
Separate coefficient sets for myopes (baseline SER <= -0.50 D) and non-myopes.
Self-validates against the paper's Table 4.

MULTI-POPULATION ROUTING
------------------------
country/ethnicity -> region + status flag:
    'exact'       -> the population is in the model's derivation set
    'approximate' -> same region, but that exact population was not sampled
    'provisional' -> not represented in either region; use own data if possible
    'unmapped'    -> no mapping; defaulted; must review
Call population_support() for the table. Add/override with register_population().
Calibrate to your own data with fit_population_multiplier().

DEPENDENCIES: numpy and pandas only. No scipy, no statsmodels, no R.
"""
from __future__ import annotations
from dataclasses import dataclass, field
import numpy as np
import pandas as pd

MONTHS = (6, 12, 18, 24, 36)

# =============================================================================
# SOFTWARE VERSION -- single source of truth for the whole tool.
# Every other component (GUI title, web-app header, output workbooks, CITATION)
# derives its version from here so releases can never drift apart. Bump both on
# each controlled release.
# =============================================================================
__version__ = "0.2.1"
__release_date__ = "2026-08-09"   # ISO date this version was released


def tool_info_frame(model_label=None, generated_on=None):
    """Provenance block written into every output workbook so a result file
    always identifies the tool build that produced it (configuration control).
    Version and release date are fixed by the build; model_label and
    generated_on describe the individual run."""
    import datetime as _dt
    rows = [
        ("Tool", "Virtual Control Arm Tool"),
        ("Version", __version__),
        ("Release date", __release_date__),
    ]
    if model_label is not None:
        rows.append(("Model used", str(model_label)))
    on = generated_on if generated_on is not None else _dt.date.today().isoformat()
    rows.append(("Generated on", on))
    return pd.DataFrame(rows, columns=["field", "value"])


def write_output(df, path, model_label=None, sheet_name="predictions"):
    """Write a result DataFrame to `path`, then append a separate 'tool_info'
    sheet carrying the version stamp. The data sheet is written exactly as
    passed (same columns, same order, first sheet), so any existing reader that
    calls pd.read_excel(path) is unaffected; only an extra sheet is added."""
    with pd.ExcelWriter(path, engine="openpyxl") as _xl:
        df.to_excel(_xl, sheet_name=sheet_name, index=False)
        tool_info_frame(model_label).to_excel(_xl, sheet_name="tool_info", index=False)


# =============================================================================
# 1. The published model: Naduvilath et al. 2025, Table 3 (exact coefficients)
# =============================================================================
@dataclass
class RegionalModel2025:
    name: str = "Naduvilath 2025 (East Asian / European regional)"
    source: str = "Naduvilath et al. Ophthalmic Physiol Opt 2025;45:135-151, Table 3"
    # --- myopes at baseline (SER <= -0.50 D) ---
    my_intercept: float = 0.537
    my_east_asian: float = 0.147      # European = 0 (reference)
    my_male: float = 0.004            # Female = 0 (reference)
    my_age: float = -0.032
    my_east_asian_age: float = -0.007  # European:age = 0
    my_ser: float = -0.007
    my_pi_se: float = 0.11            # individual prediction-interval SE (mm/yr)
    my_ci_se: float = 0.018           # SE of the mean for CI (mm/yr)
    # --- non-myopes (SER > -0.50 D) ---
    nm_intercept: float = 0.263
    nm_east_asian: float = 0.136
    nm_male: float = -0.003
    nm_age: float = -0.016
    nm_east_asian_age: float = -0.007
    nm_ser: float = -0.002
    nm_pi_se: float = 0.08
    nm_ci_se: float = 0.011

    def is_myope(self, ser):
        return ser <= -0.50

    def annual_rate(self, age, ser, male, region):
        ea = 1.0 if region == "east_asian" else 0.0
        if self.is_myope(ser):
            y = (self.my_intercept + self.my_east_asian * ea + self.my_male * male
                 + self.my_age * age + self.my_east_asian_age * ea * age
                 + self.my_ser * ser)
        else:
            y = (self.nm_intercept + self.nm_east_asian * ea + self.nm_male * male
                 + self.nm_age * age + self.nm_east_asian_age * ea * age
                 + self.nm_ser * ser)
        return float(np.exp(y) - 1.0)

    def se(self, ser, which):
        if self.is_myope(ser):
            return self.my_pi_se if which == "pi" else self.my_ci_se
        return self.nm_pi_se if which == "pi" else self.nm_ci_se

    def cumulative(self, base_age, ser, male, region, month):
        total = 0.0
        for m in range(month):
            total += self.annual_rate(base_age + m / 12.0, ser, male, region) / 12.0
        return total


MODEL_2025 = RegionalModel2025()


# =============================================================================
# 1a. Optional EMPIRICAL prediction-interval calibration (opt-in; default OFF)
# =============================================================================
# The published individual prediction interval (my_pi_se / nm_pi_se) scales
# LINEARLY with follow-up time. External validation on an untreated cohort showed
# it was too narrow, and that the residual SD of (observed - predicted) scales
# with SQRT(time) (a random walk), constant at ~0.15 mm per sqrt-year. When a
# calibration is set here, predict_untreated() uses:
#       PI half-width = z * sigma_group * (years ** power)
# with sigma an ABSOLUTE residual SD (NOT scaled by the population multiplier,
# because the multiplier corrects the MEAN, not the spread). Point predictions,
# the CI-of-the-mean band, and the paper self-validation are all unchanged.
# Set to None (clear_pi_calibration) to restore exact published behaviour.
_PI_CALIB = None   # e.g. {"myope": 0.1492, "nonmyope": None, "power": 0.5, "source": "..."}


def set_pi_calibration(myope=None, nonmyope=None, power=0.5, source=""):
    """Turn on empirical PI calibration. sigma in mm per (year**power).
    A None sigma for a group falls back to the published linear PI for that
    group (used e.g. for non-myopes when the calibration cohort had none)."""
    global _PI_CALIB
    _PI_CALIB = {"myope": myope, "nonmyope": nonmyope,
                 "power": float(power), "source": source}
    return _PI_CALIB


def clear_pi_calibration():
    """Restore exact published (linear) prediction intervals."""
    global _PI_CALIB
    _PI_CALIB = None


# =============================================================================
# 1b. India reference: STEM cross-sectional AL percentiles (Gopalakrishnan 2024)
# =============================================================================
# Axial length (mm) cut-offs by sex, age (grade 1/4/6 ~ ages 6/9/11) and centile,
# from the Sankara Nethralaya Tamil Nadu Essilor Myopia (STEM) study, Table 1.
# Cross-sectional, non-cycloplegic (myopia defined as SER <= -0.75 D). Use for
# centile-based SCREENING / tracking of Indian children, NOT as a myope
# progression model (STEM does not publish one). It confirms Indian AL tracks
# European children (slightly shorter/slower by age 9-11); this is context only.
# The tool does NOT route Indian to a region automatically: South Asian ancestry
# has no derived progression model, so it is predicted only under a stratum the
# user explicitly selects (east_asian or european), and is flagged provisional.
INDIA_STEM_AL_CENTILES = {
    "M": {  # age -> {centile: AL mm}
        6:  {2: 21.37, 5: 21.61, 10: 21.88, 25: 22.29, 50: 22.67, 75: 23.11, 90: 23.55, 95: 23.79, 98: 24.12, 99: 24.42},
        9:  {2: 21.54, 5: 21.97, 10: 22.17, 25: 22.65, 50: 23.05, 75: 23.50, 90: 23.92, 95: 24.23, 98: 24.66, 99: 25.08},
        11: {2: 22.01, 5: 22.23, 10: 22.48, 25: 22.92, 50: 23.35, 75: 23.79, 90: 24.29, 95: 24.65, 98: 25.11, 99: 25.45},
    },
    "F": {
        6:  {2: 20.99, 5: 21.22, 10: 21.40, 25: 21.76, 50: 22.18, 75: 22.54, 90: 22.93, 95: 23.16, 98: 23.41, 99: 23.76},
        9:  {2: 21.27, 5: 21.57, 10: 21.83, 25: 22.17, 50: 22.58, 75: 22.99, 90: 23.42, 95: 23.73, 98: 24.07, 99: 24.34},
        11: {2: 21.51, 5: 21.80, 10: 21.97, 25: 22.37, 50: 22.79, 75: 23.19, 90: 23.73, 95: 24.04, 98: 24.51, 99: 24.77},
    },
}


def india_al_centiles() -> pd.DataFrame:
    """STEM Indian axial-length percentile table as a tidy DataFrame
    (sex, age, centile, AL_mm). For screening / centile-tracking only."""
    rows = []
    for sex, ages in INDIA_STEM_AL_CENTILES.items():
        for age, cents in ages.items():
            for c, al in cents.items():
                rows.append({"sex": sex, "age": age, "centile": c, "AL_mm": al})
    return pd.DataFrame(rows)


# =============================================================================
# 2. Population registry: country / ethnicity -> region + status (+ multiplier)
# =============================================================================
_REGISTRY = {}   # canonical_key -> {"region","status","note","multiplier"}
_ALIASES = {}    # raw string -> canonical_key


def _norm(s):
    return str(s).strip().lower().replace("-", " ").replace("_", " ").replace("  ", " ")


def register_population(key, region, status, note="", multiplier=1.0, aliases=None):
    """Add or replace a population mapping.
    region: 'east_asian' or 'european'. multiplier scales the region baseline
    (use for a provisional population calibrated to your own data)."""
    k = _norm(key)
    _REGISTRY[k] = {"region": region, "status": status, "note": note,
                    "multiplier": float(multiplier)}
    _ALIASES[k] = k
    for a in (aliases or []):
        _ALIASES[_norm(a)] = k
    return k


def resolve_population(country_or_ethnicity, default_key="default"):
    raw = _norm(country_or_ethnicity)
    if raw in _ALIASES:
        k = _ALIASES[raw]
        return k, _REGISTRY[k]
    # Fallback: whole-word (token) matching, longest alias first, so a stray
    # substring cannot mis-route (e.g. "han" inside "Ghana", "nz" inside
    # "Tanzanian"). Multi-word aliases like "south asian" match by the exact
    # lookup above; this only catches partials such as "Han Chinese".
    tokens = set(raw.split())
    for alias in sorted(_ALIASES, key=lambda a: -len(a.split())):
        atoks = alias.split()
        if atoks and set(atoks).issubset(tokens):
            k = _ALIASES[alias]
            return k, _REGISTRY[k]
    return default_key, _REGISTRY[default_key]


# ---- default shipped mappings (based on the 2025 derivation set) ----
register_population("chinese", "east_asian", "exact",
                    "In the 2025 East Asian derivation cohort (China).",
                    aliases=["china", "han", "mainland chinese"])
register_population("vietnamese", "east_asian", "exact",
                    "In the 2025 East Asian derivation cohort (Vietnam).",
                    aliases=["vietnam"])
register_population("east asian", "east_asian", "exact",
                    "Model region.", aliases=["asian"])
register_population("taiwanese", "east_asian", "approximate",
                    "East Asian region; Taiwan not specifically sampled. Verify.",
                    aliases=["taiwan"])
register_population("singaporean", "east_asian", "approximate",
                    "East Asian region; Singapore not in the 2025 set. Approximation.",
                    aliases=["singapore"])
register_population("malaysian", "east_asian", "approximate",
                    "Mixed Malay/Chinese/Indian; routed to East Asian as nearest. Verify or supply local data.",
                    aliases=["malaysia", "malay"])
register_population("australian", "european", "exact",
                    "In the 2025 European-ancestry derivation cohort (Australia).",
                    aliases=["australia", "aussie"])
register_population("european", "european", "exact",
                    "Model region (UK, Sweden, Australia European ancestry).",
                    aliases=["white", "caucasian", "uk", "british", "swedish", "sweden"])
register_population("portuguese", "european", "approximate",
                    "European ancestry; Portugal not specifically sampled. Reasonable within-region approximation.",
                    aliases=["portugal"])
register_population("spanish", "european", "approximate",
                    "European ancestry; Spain not specifically sampled. Reasonable within-region approximation.",
                    aliases=["spain"])
register_population("new zealand", "european", "approximate",
                    "European ancestry assumed; Maori/Pasifika not represented. Verify.",
                    aliases=["nz", "new zealander", "kiwi"])
register_population("indian", "european", "no_model",
                    "No Indian or South Asian axial-elongation model exists; this ancestry "
                    "is in no bundled model's derivation set. The tool does not guess a "
                    "stratum. Choose east_asian or european in the input 'stratum' column "
                    "(the paper validates both and reports the fit), or calibrate to your "
                    "own untreated data with fit_population_multiplier(). Without a choice "
                    "the child is not predicted and is listed for review. The 'european' "
                    "field here is only the internal validation candidate, not a default.",
                    aliases=["india", "south asian"])
register_population("default", "european", "unmapped",
                    "Ancestry not recognised, so no model applies. The tool does not guess: "
                    "set 'stratum' to east_asian or european in the input to predict this "
                    "child, or correct the ethnicity. The child is otherwise listed for review.")


# =============================================================================
# 3. Predict untreated elongation for an array of subjects
# =============================================================================
def _male_flag(gender):
    return 1.0 if str(gender).strip().upper() in ("M", "MALE", "1") else 0.0


def _population_of(subject):
    # Ancestry label. Prefer 'ethnicity'; 'population' and 'country' are legacy
    # aliases some internal callers use for the SAME thing (ancestry). This is
    # ancestry, never country of residence (a Chinese child in Australia is East
    # Asian); the shipped input layer accepts only 'ethnicity'.
    return (subject.get("ethnicity") or subject.get("population")
            or subject.get("country") or "")


# Model registry for the tool-facing dropdown. First-author + model labels, as in
# the paper. Only the GEE ships a confidence interval (it is the only source that
# publishes parameter uncertainty); the other three return a point estimate with a
# note. This is neutral: a model gets a CI only if ITS OWN publication provides one.
MODEL_LABELS = {
    "naduvilath2025": "Naduvilath 2025 (regional GEE)",
    "brennan2024":    "Brennan 2024 (meta-regression)",
    "olsm_jones2005": "Jones 2005 (OLSM)",
    "scorm_wong2010": "Wong 2010 (SCORM)",
}
# Which models use ancestry/region (vs age only), and which cover non-myopes.
_ETHNICITY_DEPENDENT = {"naduvilath2025", "brennan2024"}
_COVERS_NONMYOPES = {"naduvilath2025"}   # only the GEE; the rest are myope-only
_NO_CI_NOTE = ("Point estimate; this model's source publishes no parameter "
               "covariance, so no confidence interval of the mean is available.")


# --- URS-05 input plausibility guardrails (v0.2.0) --------------------------
# Flag gross data-entry or unit errors so an implausible baseline is never used
# to compute an implausible absolute axial length. These are entry-sanity bounds,
# deliberately wider than any model applicability range; predicted delta-AL is
# unaffected. A flagged row is not predicted and is reported to the caller.
PLAUSIBILITY = {
    "age_years": (3.0, 18.0),
    "baseline_SER_D": (-25.0, 15.0),
    "baseline_AL_mm": (15.0, 35.0),
}


def _implausibility_reason(age, ser, base_al):
    """Return a human reason if any baseline value is grossly implausible, else None."""
    lo, hi = PLAUSIBILITY["age_years"]
    if not (lo <= age <= hi):
        return ("baseline age {:g} years is outside the plausible range {:g}-{:g}; "
                "check the entry".format(age, lo, hi))
    lo, hi = PLAUSIBILITY["baseline_SER_D"]
    if not (lo <= ser <= hi):
        return ("baseline SER {:g} D is outside the plausible range {:g} to {:g} D; "
                "check the entry".format(ser, lo, hi))
    lo, hi = PLAUSIBILITY["baseline_AL_mm"]
    if (base_al is not None) and (not pd.isna(base_al)) and not (lo <= base_al <= hi):
        return ("baseline axial length {:g} mm is outside the plausible range "
                "{:g}-{:g} mm; check the entry (units?)".format(base_al, lo, hi))
    return None


def predict_untreated(subjects, model: RegionalModel2025 = MODEL_2025,
                      months=MONTHS, ci=0.95, include_pi=False,
                      exclusions=None, require_stratum=True,
                      model_name="naduvilath2025") -> pd.DataFrame:
    """Ingest an array of subject dicts; return predicted untreated elongation,
    routing each subject to the model region for its population.

    Recognised keys per subject (extras ignored):
        id, age, gender, baseline_SER, baseline_AL, ethnicity, stratum

    Output (one row per subject per month):
        id, population, region, model, model_status, mapping_note, re_group,
        month, predicted_delta_AL, ci_low, ci_high, predicted_AL

    model_name selects which published model to run (one per call, the user's
    choice; run again to compare):
        'naduvilath2025' (default) -- regional GEE, ethnicity-dependent, covers
            myopes and non-myopes, the only model that ships a confidence interval
            of the mean (ci_low, ci_high) because it is the only source that
            publishes parameter uncertainty.
        'brennan2024'   -- meta-regression, ethnicity-dependent, MYOPE-ONLY, point
            estimate only (ci_low/ci_high = NaN).
        'olsm_jones2005', 'scorm_wong2010' -- AGE-ONLY (ethnicity not used,
            region='n/a'), MYOPE-ONLY, point estimate only.
    Non-GEE models leave ci_low/ci_high NaN and append a note explaining why (the
    source publishes no parameter covariance). See MODEL_LABELS for display names.

    Ancestry with no derived model (Indian/South Asian, or anything unmapped) is
    NOT guessed by the ethnicity-dependent models. If the subject supplies
    `stratum` = east_asian or european it is predicted under that stratum
    (model_status='user_selected'); otherwise, when require_stratum is True (the
    shipped default), the subject is skipped and appended to `exclusions` (a list)
    for the caller to report. Internal callers (e.g. the validation) pass
    require_stratum=False to use the registry region. Age-only models ignore
    ancestry entirely and predict regardless of stratum.

    A non-myope passed to a myope-only model is skipped and appended to
    `exclusions`. The confidence interval of the mean (ci_low, ci_high) is what a
    control arm needs. The individual prediction interval (pi_low, pi_high) is
    added only when include_pi=True (GEE only), for the validation coverage
    analysis; it is not part of the shipped output.
    """
    model_name = (model_name or "naduvilath2025").strip().lower()
    if model_name not in MODEL_LABELS:
        raise ValueError("Unknown model_name %r; choose one of: %s"
                         % (model_name, ", ".join(MODEL_LABELS)))
    is_gee = (model_name == "naduvilath2025")
    ethnicity_dependent = model_name in _ETHNICITY_DEPENDENT
    covers_nonmyopes = model_name in _COVERS_NONMYOPES
    label = MODEL_LABELS[model_name]
    if not is_gee:
        import sensitivity_models as _sens   # numpy+pandas only; no circular import
        _cum = _sens.MODELS[model_name]["cum"]

    z = 1.959963984540054 if abs(ci - 0.95) < 1e-9 else _z(ci)
    rows = []
    for s in subjects:
        sid = s.get("id", s.get("subject_id"))
        age = float(s["age"])
        ser = float(s.get("baseline_SER", -2.0))
        male = _male_flag(s.get("gender", "F"))
        base_al = s.get("baseline_AL", np.nan)
        base_al = float(base_al) if base_al is not None and not pd.isna(base_al) else np.nan
        _reason = _implausibility_reason(age, ser, base_al)
        if _reason is not None:
            if exclusions is not None:
                exclusions.append({"id": sid,
                                   "population": _population_of(s) or "(none given)",
                                   "reason": _reason})
            continue
        key, entry = resolve_population(_population_of(s))
        region, mult = entry["region"], entry["multiplier"]
        status, note = entry["status"], entry["note"]
        # normalise to the canonical region tokens (accept "East Asian", "east-asian", etc.)
        stratum = str(s.get("stratum") or "").strip().lower().replace(" ", "_").replace("-", "_")

        if ethnicity_dependent:
            # Ancestry with no derived model is not guessed: needs a user stratum.
            if status in ("no_model", "unmapped"):
                if stratum in ("east_asian", "european"):
                    region, mult, status = stratum, 1.0, "user_selected"
                    note = ("No ancestry-matched model; predicted under the user-selected "
                            "%s stratum." % stratum)
                elif require_stratum:
                    if exclusions is not None:
                        exclusions.append({
                            "id": sid,
                            "population": _population_of(s) or "(none given)",
                            "reason": ("no derived model for this ancestry; set 'stratum' to "
                                       "east_asian or european in the input, or correct the "
                                       "ethnicity")})
                    continue
        else:
            # Age-only model (OLSM/SCORM): ethnicity is not used for prediction.
            region, mult, status = "n/a", 1.0, "age_only"
            note = ("Age-only model; ethnicity/region is not used by this model. "
                    "The child's ethnicity was still recorded on input.")

        # Non-GEE models are myope-only; the GEE covers non-myopes too.
        if not covers_nonmyopes and ser > -0.50:
            if exclusions is not None:
                exclusions.append({
                    "id": sid,
                    "population": _population_of(s) or "(none given)",
                    "reason": ("%s applies to myopes only (baseline SER <= -0.50 D); "
                               "this child is not a myope at baseline" % label)})
            continue

        grp = "myope" if (model.is_myope(ser) if is_gee else ser <= -0.50) else "non-myope"
        # region token the non-GEE cum() expects (age-only ignores it anyway)
        cum_region = region if ethnicity_dependent else "european"
        for mth in months:
            if is_gee:
                point = model.cumulative(age, ser, male, region, mth) * mult
            else:
                point = _cum(age, ser, male, cum_region, mth)
            yrs = mth / 12.0
            row = {
                "id": sid, "population": _population_of(s) or "(none given)",
                "region": region, "model": label, "model_status": status,
                "mapping_note": note, "re_group": grp, "month": mth,
                "predicted_delta_AL": round(point, 4),
            }
            if is_gee:
                # Individual prediction interval: empirical calibration if set,
                # else the published linear PI. Empirical sigma is absolute (not
                # scaled by the mean-correcting multiplier). The tool ships the
                # mean CI only; the PI is included only when include_pi=True, for
                # the validation coverage analysis (it under-covers), never in the
                # end-user output.
                if _PI_CALIB is not None:
                    sig = _PI_CALIB["myope"] if model.is_myope(ser) else _PI_CALIB["nonmyope"]
                    if sig is not None and np.isfinite(sig):
                        pi = z * float(sig) * (yrs ** _PI_CALIB["power"])
                    else:
                        pi = z * model.se(ser, "pi") * yrs * mult
                else:
                    pi = z * model.se(ser, "pi") * yrs * mult
                cise = z * model.se(ser, "ci") * yrs * mult
                if include_pi:
                    row["pi_low"] = round(point - pi, 4)
                    row["pi_high"] = round(point + pi, 4)
                row["ci_low"] = round(point - cise, 4)
                row["ci_high"] = round(point + cise, 4)
            else:
                # No published parameter covariance -> point estimate only.
                if include_pi:
                    row["pi_low"] = np.nan
                    row["pi_high"] = np.nan
                row["ci_low"] = np.nan
                row["ci_high"] = np.nan
                row["mapping_note"] = (note + " " + _NO_CI_NOTE).strip()
            row["predicted_AL"] = round(base_al + point, 4) if not np.isnan(base_al) else np.nan
            rows.append(row)
    return pd.DataFrame(rows)


# =============================================================================
# 4. Compare treated arm against predicted untreated
# =============================================================================
def compare_to_test_arm(predictions, observed, n_boot=10000, seed=20260706, ci=0.95):
    """Treatment effect = observed treated elongation - predicted untreated, per
    month, with a paired-bootstrap 95% CI and p-value. observed columns:
    id, month, observed_elongation.

    INTERNAL / VALIDATION USE ONLY. This is not exposed by the shipped tool
    (gui_app.py / run_my_trial.py output the control only). Its paired bootstrap
    resamples the observed-minus-predicted differences and therefore treats the
    virtual control as error-free: it does NOT propagate the control's own mean
    uncertainty (ci_low/ci_high) or calibration uncertainty, so its CI and
    p-value are anticonservative. For the recommended end-user analysis, which
    carries the control's uncertainty into the effect, see
    docs/ANALYSIS_PROTOCOL.md. Retained here only for internal reproducibility
    and the deterministic-signing test.

    Reported effect measures (Brennan et al. 2021, "Efficacy in myopia control"):
      - delta_effect      : observed - predicted (negative = treatment slowed growth)
      - CARE_mm           : Cumulative Absolute Reduction in Elongation
                            = predicted_untreated - observed (positive = benefit).
                            This absolute measure is the recommended primary metric.
      - percent_reduction : CARE / predicted_untreated. Reported but CAUTIONED:
                            the denominator shrinks with age, so percentages inflate
                            at older ages and longer follow-up. Prefer CARE_mm.
    """
    rng = np.random.default_rng(seed)
    alpha = (1 - ci) / 2 * 100
    pred = predictions[["id", "month", "predicted_delta_AL"]]
    merged = observed.merge(pred, on=["id", "month"], how="inner")
    merged["paired_diff"] = merged["observed_elongation"] - merged["predicted_delta_AL"]
    out = []
    for mth, g in merged.groupby("month"):
        d = g["paired_diff"].to_numpy(dtype=float)
        d = d[np.isfinite(d)]
        n = d.size
        if n == 0:
            continue
        boot = rng.choice(d, size=(n_boot, n), replace=True).mean(axis=1)
        lo, hi = np.percentile(boot, [alpha, 100 - alpha])
        p_boot = 2 * min((boot >= 0).mean(), (boot <= 0).mean())
        mean_obs = float(g["observed_elongation"].mean())
        mean_pred = float(g["predicted_delta_AL"].mean())
        care = mean_pred - mean_obs                       # positive = benefit
        pct = (care / mean_pred * 100.0) if mean_pred > 0 else np.nan
        out.append({
            "month": int(mth), "n": n,
            "mean_observed": round(mean_obs, 4),
            "mean_predicted_untreated": round(mean_pred, 4),
            "delta_effect": round(float(d.mean()), 4),
            "ci_low": round(float(lo), 4), "ci_high": round(float(hi), 4),
            "p_value_bootstrap": round(min(1.0, float(p_boot)), 4),
            "CARE_mm": round(care, 4),                     # primary absolute metric
            "percent_reduction": round(pct, 1) if np.isfinite(pct) else np.nan,  # cautioned
        })
    return pd.DataFrame(out).sort_values("month").reset_index(drop=True)


# =============================================================================
# 5. Calibrate a population to your own untreated data (returns a multiplier)
# =============================================================================
def fit_population_multiplier(df, region="european", model: RegionalModel2025 = MODEL_2025):
    """For a population not in the model (e.g. Indian), estimate a scale factor so
    the chosen region baseline matches YOUR untreated cohort's mean annual
    elongation. Register it: register_population('indian', region, 'exact',
    note='calibrated', multiplier=<returned value>).

    df columns: id, age_years, AL, baseline_SER, gender  (one row per child per visit)."""
    d = df.sort_values(["id", "age_years"])
    g = d.groupby("id")
    first, last = g.first(), g.last()
    dyr = (last["age_years"] - first["age_years"]).replace(0, np.nan)
    obs_rate = (last["AL"] - first["AL"]) / dyr
    mean_age = (first["age_years"] + last["age_years"]) / 2
    ser = first["baseline_SER"] if "baseline_SER" in first else pd.Series(-2.0, index=first.index)
    male = (first["gender"].astype(str).str.upper().str[0] == "M").astype(float) if "gender" in first else 0.5
    pred = [model.annual_rate(a, s, mm, region)
            for a, s, mm in zip(mean_age, ser, male)]
    ratio = float(np.nanmean(obs_rate) / np.nanmean(pred))
    return round(ratio, 4) if np.isfinite(ratio) and ratio > 0 else 1.0


# =============================================================================
# 6. Coverage view and validation
# =============================================================================
def population_support() -> pd.DataFrame:
    seen = {}
    for alias, key in _ALIASES.items():
        seen.setdefault(key, []).append(alias)
    rows = []
    for key, entry in _REGISTRY.items():
        rows.append({
            "population_key": key,
            "aliases": ", ".join(sorted(a for a in seen.get(key, []) if a != key)),
            "region": entry["region"], "status": entry["status"],
            "multiplier": entry["multiplier"], "note": entry["note"],
        })
    order = {"exact": 0, "approximate": 1, "provisional": 2, "unmapped": 3}
    return pd.DataFrame(rows).sort_values(
        ["status", "population_key"],
        key=lambda c: c.map(order).fillna(9) if c.name == "status" else c
    ).reset_index(drop=True)


def validate_against_paper():
    m = MODEL_2025
    checks = [
        ("EA myope age10 -2D ~0.36", m.annual_rate(10, -2, 0.5, "east_asian"), 0.36),
        ("EU myope age10 -2D ~0.26", m.annual_rate(10, -2, 0.5, "european"), 0.26),
        ("EA myope age6 -1D ~0.58", m.annual_rate(6, -1, 0.5, "east_asian"), 0.58),
        ("EU myope age6 -1D ~0.43", m.annual_rate(6, -1, 0.5, "european"), 0.43),
        ("EA nonmyope age10 0D ~0.18", m.annual_rate(10, 0.0, 0.5, "east_asian"), 0.18),
        ("EU nonmyope age10 0D ~0.11", m.annual_rate(10, 0.0, 0.5, "european"), 0.11),
    ]
    print("Validation against Naduvilath et al. 2025 (Table 4):")
    ok = True
    for label, got, want in checks:
        good = abs(got - want) <= 0.02
        ok = ok and good
        print("  [{}] {}: got {:.3f}".format("PASS" if good else "FAIL", label, got))
    return ok


# Golden predicted_delta_AL (mm) pinned from the validated engine, so the
# self-test can confirm ALL FOUR shipped models reproduce, not only the GEE.
# Subjects are myopes, age 9, baseline SER -2.0 D, male: EA = East Asian
# (Chinese), EU = European (Australian). Regenerate these numbers only when the
# engine is intentionally changed and re-validated.
_SELFTEST_SUBJECTS = {
    "EA": {"id": "EA", "age": 9.0, "gender": "M", "baseline_SER": -2.0, "ethnicity": "Chinese"},
    "EU": {"id": "EU", "age": 9.0, "gender": "M", "baseline_SER": -2.0, "ethnicity": "Australian"},
}
_GOLDEN = [
    # (model_name, subject_key, month, expected_delta_AL_mm)
    ("naduvilath2025", "EA", 6, 0.2045), ("naduvilath2025", "EA", 12, 0.3954),
    ("naduvilath2025", "EU", 6, 0.1487), ("naduvilath2025", "EU", 12, 0.2871),
    ("brennan2024",    "EA", 6, 0.2320), ("brennan2024",    "EA", 12, 0.4465),
    ("brennan2024",    "EU", 6, 0.1677), ("brennan2024",    "EU", 12, 0.3226),
    ("olsm_jones2005", "EA", 6, 0.1299), ("olsm_jones2005", "EA", 12, 0.2530),
    ("scorm_wong2010", "EA", 6, 0.1734), ("scorm_wong2010", "EA", 12, 0.3290),
]


def validate_models_golden(tol=1e-3):
    """Confirm every shipped model reproduces its pinned value: build integrity
    for all four models, not only the GEE. A mismatch means the deployed build
    has drifted from the validated engine."""
    print("Model reproduction (all four shipped models):")
    ok = True
    for model_name, skey, month, want in _GOLDEN:
        got = float(predict_untreated([_SELFTEST_SUBJECTS[skey]], months=(month,),
                                      model_name=model_name)["predicted_delta_AL"].iloc[0])
        good = abs(got - want) <= tol
        ok = ok and good
        print("  [{}] {}, {} {}mo: got {:.4f} (expect {:.4f})".format(
            "PASS" if good else "FAIL", MODEL_LABELS[model_name], skey, month, got, want))
    return ok


def self_test():
    """Full self-test run by the tool. Two parts: the primary model against the
    published Naduvilath 2025 Table 4, then all four shipped models against
    pinned golden values. Returns True only if every check passes."""
    a = validate_against_paper()
    print()
    b = validate_models_golden()
    return bool(a and b)


def _z(ci):
    p = 0.5 + ci / 2
    a = [-39.6968302866538, 220.946098424521, -275.928510446969, 138.357751867269, -30.6647980661472, 2.50662827745924]
    b = [-54.4760987982241, 161.585836858041, -155.698979859887, 66.8013118877197, -13.2806815528857]
    c = [-0.00778489400243029, -0.322396458041136, -2.40075827716184, -2.54973253934373, 4.37466414146497, 2.93816398269878]
    d = [0.00778469570904146, 0.32246712907004, 2.445134137143, 3.75440866190742]
    pl, ph = 0.02425, 1 - 0.02425
    if p < pl:
        q = np.sqrt(-2 * np.log(p)); return (((((c[0]*q+c[1])*q+c[2])*q+c[3])*q+c[4])*q+c[5]) / ((((d[0]*q+d[1])*q+d[2])*q+d[3])*q+1)
    if p > ph:
        q = np.sqrt(-2 * np.log(1 - p)); return -(((((c[0]*q+c[1])*q+c[2])*q+c[3])*q+c[4])*q+c[5]) / ((((d[0]*q+d[1])*q+d[2])*q+d[3])*q+1)
    q = p - 0.5; r = q*q
    return (((((a[0]*r+a[1])*r+a[2])*r+a[3])*r+a[4])*r+a[5])*q / (((((b[0]*r+b[1])*r+b[2])*r+b[3])*r+b[4])*r+1)


# =============================================================================
# 7. Demo
# =============================================================================
if __name__ == "__main__":
    self_test()

    print("\nPOPULATION SUPPORT:")
    print(population_support().drop(columns="note").to_string(index=False))

    subjects = [
        {"id": "CN01", "age": 9.0,  "gender": "F", "baseline_SER": -2.00, "country": "Chinese"},
        {"id": "VN01", "age": 9.0,  "gender": "F", "baseline_SER": -2.00, "country": "Vietnamese"},
        {"id": "AU01", "age": 9.0,  "gender": "F", "baseline_SER": -2.00, "country": "Australian"},
        # South Asian has no derived model; the user must choose a stratum (here
        # east_asian) or the child is not predicted.
        {"id": "IN01", "age": 9.0,  "gender": "F", "baseline_SER": -2.00,
         "country": "Indian", "stratum": "east_asian"},
    ]
    pred = predict_untreated(subjects, months=(12, 24, 36))
    cols = ["id", "population", "region", "model_status", "month", "predicted_delta_AL", "ci_low", "ci_high"]
    print("\nPREDICTED UNTREATED ELONGATION (mm), routed by population:")
    print(pred[cols].to_string(index=False))

    print("\nINDIA STEM AL percentile reference (boys, 50th centile):")
    ic = india_al_centiles()
    print(ic[(ic.sex == "M") & (ic.centile == 50)].to_string(index=False))
```
