## Supplementary software v0.2.1 for "Virtual control arms for paediatric myopia trials: external validation of axial-elongation models": test_engine.py

```
"""
Minimal but load-bearing tests for the virtual-control engine.

    pip install pytest
    pytest -q                      # run from the archive root

These assert (1) the engine still matches the published model, (2) population
routing is correct, (3) predictions are numerically stable (golden values), and
(4) the treatment-effect stats are deterministic and correctly signed. They need
no patient data. Keep them green before every release.
"""
import os
import sys

import numpy as np
import pandas as pd
import pytest

# the code lives in ../src relative to this tests/ folder
SRC = os.path.join(os.path.dirname(os.path.dirname(os.path.abspath(__file__))), "src")
sys.path.insert(0, SRC)

import virtual_control as vc          # noqa: E402
import gui_app                        # noqa: E402

TEMPLATE = os.path.join(SRC, "input_template.xlsx")


def test_self_validation_passes():
    assert vc.validate_against_paper() is True


def test_full_self_test_covers_all_four_models():
    """The self-test exercised by the tool covers every shipped model, not just
    the GEE: the golden set names all four, and the combined self_test() passes."""
    assert vc.validate_models_golden() is True
    assert vc.self_test() is True
    covered = {m for (m, _k, _mo, _v) in vc._GOLDEN}
    assert covered == {"naduvilath2025", "brennan2024", "olsm_jones2005", "scorm_wong2010"}


@pytest.mark.parametrize("population,region,status", [
    ("Chinese", "east_asian", "exact"),
    ("Vietnamese", "east_asian", "exact"),
    ("Australian", "european", "exact"),
    ("Indian", "european", "no_model"),       # South Asian: no derived model
    ("Klingon", "european", "unmapped"),      # unknown -> not guessed
])
def test_population_routing(population, region, status):
    key, entry = vc.resolve_population(population)
    assert entry["region"] == region
    assert entry["status"] == status


def test_no_model_ancestry_refused_without_stratum():
    """An ancestry with no derived model (South Asian) is NOT guessed: with no
    stratum it is excluded and reported; with a stratum it is predicted and
    flagged user_selected."""
    subj = [{"id": "SA", "age": 9.0, "gender": "F", "baseline_SER": -2.0,
             "ethnicity": "South Asian"}]
    excl = []
    pred = vc.predict_untreated(subj, months=(12,), exclusions=excl)
    assert pred.empty and len(excl) == 1 and excl[0]["id"] == "SA"

    subj2 = [{**subj[0], "stratum": "east_asian"}]
    pred2 = vc.predict_untreated(subj2, months=(12,))
    assert not pred2.empty
    assert (pred2["region"] == "east_asian").all()
    assert (pred2["model_status"] == "user_selected").all()


def test_blank_ethnicity_not_predicted(tmp_path):
    """A row with blank ethnicity is not predicted and is listed in
    out_needs_input.xlsx; country of residence is not used as ancestry."""
    df = pd.read_excel(TEMPLATE)
    df.loc[0, "ethnicity"] = None                      # blank the first child's ancestry
    p = os.path.join(str(tmp_path), "in.xlsx")
    df.to_excel(p, index=False)
    pred, summary = gui_app.run_analysis(p, str(tmp_path))
    assert os.path.exists(os.path.join(str(tmp_path), "out_needs_input.xlsx"))
    assert (pred["id"] != df.loc[0, "id"]).all()


def test_golden_prediction_values():
    """Point predictions must not drift. Values pinned from the validated engine."""
    subjects = [
        {"id": "CN", "age": 9.0, "gender": "F", "baseline_SER": -2.0, "country": "Chinese"},
        {"id": "AU", "age": 9.0, "gender": "F", "baseline_SER": -2.0, "country": "Australian"},
    ]
    pred = vc.predict_untreated(subjects, months=(12,))
    cn = float(pred[pred.id == "CN"].predicted_delta_AL.iloc[0])
    au = float(pred[pred.id == "AU"].predicted_delta_AL.iloc[0])
    assert cn == pytest.approx(0.3898, abs=1e-4)
    assert au == pytest.approx(0.2819, abs=1e-4)
    assert cn > au                              # East Asian myopes elongate faster


def test_tool_outputs_control_only(tmp_path):
    """The shipped tool writes the virtual control ONLY: out_predictions.xlsx is
    produced, no effect file, and run_analysis returns a 2-tuple. The effect
    calculation is the analyst's own step (docs/ANALYSIS_PROTOCOL.md)."""
    result = gui_app.run_analysis(TEMPLATE, str(tmp_path))
    assert len(result) == 2                                       # (pred, summary)
    pred, summary = result
    assert os.path.exists(os.path.join(str(tmp_path), "out_predictions.xlsx"))
    assert not os.path.exists(os.path.join(str(tmp_path), "out_effect.xlsx"))
    # the tool no longer produces a parental-myopia output
    assert not os.path.exists(os.path.join(str(tmp_path), "out_predictions_parental_myopia.xlsx"))
    assert not pred.empty
    assert "ANALYSIS_PROTOCOL" in summary


def test_wide_layout(tmp_path):
    """Wide layout writes one row per child with month-suffixed columns; the
    default long layout writes one row per child per month."""
    gui_app.run_analysis(TEMPLATE, str(tmp_path), layout="wide")
    w = pd.read_excel(os.path.join(str(tmp_path), "out_predictions.xlsx"))
    assert len(w) == 3
    assert "predicted_delta_AL_m12" in w.columns and "ci_low_m6" in w.columns
    assert "month" not in w.columns


def test_effect_engine_is_deterministic_and_signed():
    """The internal-only effect engine (vc.compare_to_test_arm, not exposed by the
    shipped tool) stays reproducible and CARE is signed so positive = benefit.
    Uses a synthetic treated arm at 60% of the predicted control."""
    df = pd.read_excel(TEMPLATE)
    subjects = [{"id": r.id, "ethnicity": r.ethnicity, "gender": r.gender,
                 "age": r.age, "baseline_SER": r.baseline_SER,
                 "baseline_AL": r.baseline_AL,
                 "stratum": getattr(r, "stratum", None)} for r in df.itertuples()]
    pred = vc.predict_untreated(subjects)
    obs = pd.DataFrame({"id": pred["id"], "month": pred["month"],
                        "observed_elongation": pred["predicted_delta_AL"] * 0.6})
    eff1 = vc.compare_to_test_arm(pred, obs)
    eff2 = vc.compare_to_test_arm(pred, obs)
    pd.testing.assert_frame_equal(eff1, eff2)                     # fixed bootstrap seed
    assert (eff1["CARE_mm"] > 0).all()
    assert np.allclose(eff1["CARE_mm"], -eff1["delta_effect"], atol=1e-6)


def test_implausible_baseline_flagged(tmp_path):
    """URS-05: grossly implausible baselines (AL 50 mm, age 40 y) are flagged and
    not predicted, instead of producing an implausible absolute AL."""
    subj = [{"id": "BAD", "age": 9.0, "gender": "F", "baseline_SER": -2.0,
             "baseline_AL": 50.0, "ethnicity": "Chinese"},
            {"id": "OK", "age": 9.0, "gender": "F", "baseline_SER": -2.0,
             "baseline_AL": 24.0, "ethnicity": "Chinese"}]
    excl = []
    pred = vc.predict_untreated(subj, months=(12,), exclusions=excl)
    ids = set(pred["id"])
    assert "OK" in ids and "BAD" not in ids
    assert any(e["id"] == "BAD" and "axial length" in e["reason"] for e in excl)
    excl2 = []
    vc.predict_untreated([{"id": "AGE", "age": 40.0, "gender": "F",
                           "baseline_SER": -2.0, "ethnicity": "Chinese"}],
                         months=(12,), exclusions=excl2)
    assert excl2 and "age" in excl2[0]["reason"]


def test_output_has_ci_not_pi():
    """The shipped output carries the mean confidence interval and NOT the
    individual prediction interval; the CI widens with time. The PI is available
    only on request (include_pi=True), for the validation coverage analysis."""
    subjects = [{"id": "X", "age": 9.0, "gender": "F", "baseline_SER": -2.0,
                 "country": "Chinese"}]
    pred = vc.predict_untreated(subjects, months=(6, 12, 24))
    assert "ci_low" in pred.columns and "ci_high" in pred.columns
    assert "pi_low" not in pred.columns and "pi_high" not in pred.columns
    widths = (pred["ci_high"] - pred["ci_low"]).tolist()
    assert widths == sorted(widths)                    # monotonically non-decreasing
    pi = vc.predict_untreated(subjects, months=(6,), include_pi=True)
    assert "pi_low" in pi.columns and "pi_high" in pi.columns


# ---------------------------------------------------------------------------
# Model-selection dropdown: one model per run, neutral. Only the GEE ships a CI.
# ---------------------------------------------------------------------------
def test_only_gee_ships_a_confidence_interval():
    """The GEE returns a finite mean CI; every other model returns a point
    estimate with NaN ci_low/ci_high and a note saying why."""
    subj = [{"id": "CN", "age": 9.0, "gender": "M", "baseline_SER": -2.0,
             "ethnicity": "Chinese"}]
    gee = vc.predict_untreated(subj, months=(12,), model_name="naduvilath2025")
    assert np.isfinite(gee["ci_low"]).all() and np.isfinite(gee["ci_high"]).all()
    assert (gee["model"] == "Naduvilath 2025 (regional GEE)").all()
    for name in ("brennan2024", "olsm_jones2005", "scorm_wong2010"):
        p = vc.predict_untreated(subj, months=(12,), model_name=name)
        assert not p.empty
        assert p["ci_low"].isna().all() and p["ci_high"].isna().all()
        assert p["mapping_note"].str.contains("no").any()
        assert (p["model"] == vc.MODEL_LABELS[name]).all()


def test_age_only_models_ignore_ethnicity():
    """OLSM/SCORM are age-only: they predict a no-model ancestry (Indian) without
    a stratum, with region 'n/a', and give the same number regardless of ancestry.
    Ethnicity-dependent models still refuse the same child."""
    indian = [{"id": "IN", "age": 9.0, "gender": "M", "baseline_SER": -2.0,
               "ethnicity": "Indian"}]
    for name in ("olsm_jones2005", "scorm_wong2010"):
        excl = []
        p = vc.predict_untreated(indian, months=(12,), exclusions=excl, model_name=name)
        assert not p.empty and not excl
        assert (p["region"] == "n/a").all()
    # same age-only number for a different ancestry
    chinese = [{**indian[0], "id": "CN", "ethnicity": "Chinese"}]
    a = vc.predict_untreated(indian, months=(12,), model_name="olsm_jones2005")
    b = vc.predict_untreated(chinese, months=(12,), model_name="olsm_jones2005")
    assert a["predicted_delta_AL"].iloc[0] == pytest.approx(
        b["predicted_delta_AL"].iloc[0], abs=1e-9)
    # ethnicity-dependent model still refuses Indian without a stratum
    excl = []
    r = vc.predict_untreated(indian, months=(12,), exclusions=excl, model_name="brennan2024")
    assert r.empty and len(excl) == 1


def test_non_gee_models_are_myope_only():
    """A non-myope is predicted by the GEE but excluded (flagged) by every
    myope-only model."""
    hyperope = [{"id": "H", "age": 9.0, "gender": "F", "baseline_SER": +0.5,
                 "ethnicity": "Chinese"}]
    assert not vc.predict_untreated(hyperope, months=(12,),
                                    model_name="naduvilath2025").empty
    for name in ("brennan2024", "olsm_jones2005", "scorm_wong2010"):
        excl = []
        p = vc.predict_untreated(hyperope, months=(12,), exclusions=excl, model_name=name)
        assert p.empty and len(excl) == 1
        assert "myope" in excl[0]["reason"]


def test_unknown_model_name_raises():
    subj = [{"id": "X", "age": 9.0, "gender": "F", "baseline_SER": -2.0,
             "ethnicity": "Chinese"}]
    with pytest.raises(ValueError):
        vc.predict_untreated(subj, model_name="not_a_model")


def test_gui_ethnicity_overrides_and_flags_mismatch(tmp_path):
    """When an ethnicity is chosen in the app, a file row that disagrees is
    flagged (not predicted); a blank file ethnicity takes the app choice."""
    df = pd.DataFrame([
        {"id": "P1", "ethnicity": "Chinese", "gender": "M", "age": 9,
         "baseline_SER": -2.0, "baseline_AL": 24.0},
        {"id": "P2", "ethnicity": "European", "gender": "F", "age": 10,
         "baseline_SER": -1.0, "baseline_AL": 23.5},
    ])
    p = os.path.join(str(tmp_path), "in.xlsx")
    df.to_excel(p, index=False)
    pred, _ = gui_app.run_analysis(p, str(tmp_path),
                                   model_name="naduvilath2025", gui_ethnicity="Chinese")
    assert set(pred["id"]) == {"P1"}                       # P2 mismatched -> flagged
    needs = pd.read_excel(os.path.join(str(tmp_path), "out_needs_input.xlsx"))
    assert (needs["id"] == "P2").any()
    assert needs["reason"].str.contains("does not match").any()
```
